# Systematic approach to outcome assessment from coded electronic healthcare records in the DaRe2THINK NHS-embedded randomised trial

**DOI:** 10.1101/2022.05.24.22275434

**Authors:** Xiaoxia Wang, Alastair R Mobley, Otilia Tica, Kelvin Okoth, Rebecca E Ghosh, Puja Myles, Tim Williams, Sandra Haynes, Krishnarajah Nirantharakumar, David Shukla, Dipak Kotecha, the DaRe2THINK Trial Committees

## Abstract

**Background:** Improving the efficiency of clinical trials is key to their continued importance in directing evidence-based patient care. Digital innovations, in particular the use of electronic healthcare records (EHR), allow for large-scale screening and follow-up of participants. However, it is critical these developments are accompanied by robust and transparent methods that can support high quality and high clinical value research.

**Methods:** The DaRe2THINK trial includes a series of novel processes, including nationwide pseudonymised pre-screening of the primary care EHR across England, digital enrolment, remote e-consent, and ‘no-visit’ follow-up by linking all primary and secondary care health data with patient-reported outcomes.

**Findings:** DaRe2THINK is a pragmatic, healthcare-embedded randomised trial testing whether earlier use of direct oral anticoagulants in patients with prior or current atrial fibrillation can prevent thromboembolic events and cognitive decline (www.birmingham.ac.uk/dare2think). This paper outlines the systematic approach and methodology employed to define patient information and outcome events. This includes transparency on all medical code lists and phenotypes used in the trial across a variety of national data sources, including Clinical Practice Research Datalink Aurum (primary care), Hospital Episode Statistics (secondary care) and the Office for National Statistics (mortality).

**Interpretation:** Co-designed by a patient and public involvement team, DaRe2THINK presents an opportunity to transform the approach to randomised trials in the setting of routine healthcare, providing high-quality evidence generation in populations representative of the community at-risk.

## INTRODUCTION

Rapid uptake of electronic healthcare record (EHR) systems across the world have led to the opportunity for large-scale, real-world, longitudinal clinical research.[1] In countries with national health systems such as the United Kingdom, there is also the potential to link all EHRs for an individual patient, across both primary and secondary care, and from birth to death. Standardised coding systems provide the basis for harnessing medical information from different healthcare providers, with linkage allowing for a complete picture of each patient’s history, healthcare utilisation and adverse events.[2] This is an attractive (and cost-efficient) prospect for clinical research, particular as the quality of EHR data is typically high due to the need for accurate billing and reimbursement purposes. [3-5]

Observational clinical research has benefited for many decades from EHR data. Evolving technology and coding systems, coupled with more complete EHR coverage, has provided the opportunity for randomised controlled trials (RCT) to be embedded alongside EHR systems, taking advantage of existing clinical data and follow-up. The coronavirus pandemic has epitomised how innovations in the design and running of RCTs are critical to address unmet clinical need. However, RCTs remain too costly, tend to recruit selective patients within high-performing centres, and frequently do not match the real population at-risk in our communities.

The DaRe2 approach (healthcare Data for pragmatic clinical Research in the NHS – primary 2 secondary) was designed to incorporate recent innovations in EHR and clinical research, combined with advances in mobile technology, to provide an end-to-end framework for RCTs embedded in Primary Care in England. The DaRe2THINK trial provides an exemplar of a “remote-RCT” to address a key public health concern, requiring no physical patient visits despite being a clinical trial of an investigational medicinal product (CTIMP).[6] In this paper, we outline the design features of DaRe2THINK, and the systematic approach used to define patient information and outcome events that will be accrued during the trial. All medical code lists and phenotypes are presented for full transparency, and for future use by other researchers using EHR data.

## THE DaRe2THINK TRIAL

Using the digital potential of the EHR, DaRe2THINK is transformational project that will underpin a new trajectory for NHS-based research for patient benefit. DaRe2THINK is led by the University of Birmingham Institute of Cardiovascular Sciences, in collaboration with the Clinical Practice Research Datalink (CPRD; part of the Medicines and Healthcare products Regulatory Agency). The trial is supported by the University Hospitals Birmingham NHS Foundation Trust, University of Birmingham Institute of Applied Health Research and Centre for Patient Reported Outcomes Research, Aston University, London School of Economics, University of Oxford Nuffield Department of Population Health, NIHR Clinical Research Network (West Midlands Primary Care), and Health Data Research UK Midlands.

### DaRe2 platform features

1. Automated secure screening of inclusion and exclusion criteria across more than 13 million NHS patients, including around 1 in 4 primary care sites in England that are part of CPRD. This provides rapid and cost-efficient screening of a diverse and representative proportion of the UK population[7], yet maintains patient privacy using pseudonymised records that can only be re-identified by local NHS staff.
2. Targeted enrolment of primary care sites identified as having potentially-eligible patients, streamlining recruitment by focusing resources on high-value sites.
3. Dynamic adaptive screening of the population at each site, with updates on a weekly basis of newly-eligible patients, reducing burden on local NHS staff and simplifying recruitment.
4. Easy enrolment of patients, with remote e-consent using their own smartphone/tablet/computer, one-click randomisation by primary care staff, and drug prescription via usual clinical systems.
5. ‘No-visit’ follow-up, utilising NHS records linked across national primary and secondary care for capture of endpoints without the patient attending the healthcare site, and no need for NHS investigators to complete case report forms.
6. Regular scheduled patient-reported outcomes, with requests sent by automated text message to each participant’s telephone and email, and secure data acquisition through online completion.
7. Automated collation of safety outcomes from the primary care NHS record, avoiding the possibility of missed events unknown to the local investigator.
8. Co-design and management of all processes by a Patient and Public Involvement (PPI) team [8], supporting a patient-centric approach with sustainable and valued output.

### DaRe2THINK design

DaRe2THINK will be the first exemplar of this system, and is focused on the intersection of key national priorities for health and social care services – the common heart rhythm disorder called atrial fibrillation (AF) and the impact this condition has on thromboembolic events, including long-term cognitive decline and vascular dementia. Patients with AF suffer from a high rate of morbidity, with one in four stroke patients having AF as a potential cause, half of AF patients developing heart failure that responds poorly to treatment, and the rate of death doubled at all ages.[9-11] Most patients with AF develop progressive subclinical cerebral damage over time [12, 13], with high rates of cognitive impairment and a 40% increased risk of dementia compared to normal sinus rhythm.[14, 15] As the prevalence of AF is expected to double in the coming decades [16], this will place an unsustainable burden on healthcare and social services.

DaRe2THINK is a pragmatic, NHS-embedded, open-label, event-driven, parallel-group RCT. It will test whether earlier use of anticoagulants is effective and cost-effective at preventing thromboembolic events (primary outcome) and cognitive decline (key secondary outcome) in patients with AF at low or intermediate risk of stroke. Around 3,000 patients will be randomised to either starting a direct oral anticoagulant (DOAC; used due to their excellent safety and efficacy profile [17, 18]), or continuing standard-of-care where anticoagulation is instituted when the patient is older with clear risk factors for stroke.[19] A summary of the DaRe2THINK trial is presented in **Figure 1**. Due to the ability to access pseudonymised NHS records, DaRe2THINK will be able to adapt inclusion and exclusion criteria to reflect current changes in practice and extraneous factors (e.g. the coronavirus pandemic). Recruitment has commenced across England, and the current DaRe2THINK protocol is available at https://www.birmingham.ac.uk/dare2think. The trial has received ethical/Health Research Authority approval (REC number: 21/NE/0021; IRAS project ID: 290420), regulatory approval (MHRA CTA 21761/0364) and is registered at Clinicaltrials.gov (NCT04700826), ISRCTN (21157803) and EudraCT (2020-005774-10). A plain English summary written by the PPI team is provided in **Table 1**.

**Table 1:** Plain English summary of DaRe2THINK.

|  |  |
| --- | --- |
| **Challenges and opportunities** | The National Health Service (NHS) is unlike any other in the world, caring for people throughout their lives both in the community and in hospitals. At the heart of DaRe2THINK is that health data collected within these services can be used for the benefit of patients. Clinical trials are an important way to understand how new treatments can be used in the NHS, but many trials struggle to find the right patients, or be relevant to their needs. The DaRe2 approach (healthcare <u>D</u>ata for pragmatic clinical <u>R</u>esearch in the NHS – primary <u>2</u> secondary) will test a new way of running trials based at General Practitioner (GP) surgeries using routine NHS information. We will include patients that don't normally take part in clinical trials and follow them up without the need to revisit their GP or attend hospital. This approach could improve the health and well-being of those treated by the NHS, whilst reducing the time needed from staff and patients to engage in important research. |
| **Addressing a gap in current healthcare** | As an example of this new system, DaRe2THINK will target an issue of huge importance to patients, our NHS and the social care system. Atrial fibrillation (AF) is a common heart rhythm condition that leads to a high chance of stroke, frequent hospital admissions and poor quality of life. Patients also have a much higher risk of cognitive decline (trouble remembering, concentrating or making every-day decisions) and dementia. This may be due to silent 'micro-strokes' that gradually damage the brain over time. Blood thinning tablets (anticoagulants) greatly reduce the number of patients with AF that will suffer a stroke, but are usually only given to older patients or those with other health issues. This may be too late to avoid dementia. It also leaves those younger than 65 years, and some patients aged 65-75 without treatment that could prevent these devastating complications. |
| **Aim of DaRe2THINK** | A new class of blood thinning tablets are now widely used in the NHS which are more convenient for patients to take, and have a lower risk of bleeding than older treatments. These drugs could provide an effective way to prevent strokes, brain damage and dementia in later life for a broader group of patients, but this needs to be tested in a clinical trial. |
| **How the trial works** | With the support of a Patient and Public Involvement Team and a national network of research nurses and GPs, the trial will include 3,000 patients from up to 600 GP surgeries across England. Each patient will either continue their current treatment or start an additional blood thinning tablet on a random basis. Patients will be followed up automatically within the NHS to look at the difference in those who suffer from strokes, blood clots, heart attacks, other problems with the blood vessels and dementia. Patients will self-report their memory, reaction times and quality of life using simple questionnaires through their mobile phone or the internet, again without needing to revisit their doctor. |
| **Results and impact** | DaRe2THINK will answer important questions for a growing number of patients with AF. The combination of information from the community as well as hospitals across the NHS will allow us to see whether these blood thinning tablets should be prescribed more widely. DaRe2THINK will allow us to develop and improve this new clinical trial system so that future research in the NHS will continue to benefit those patients most in need. |

**Figure 1:**
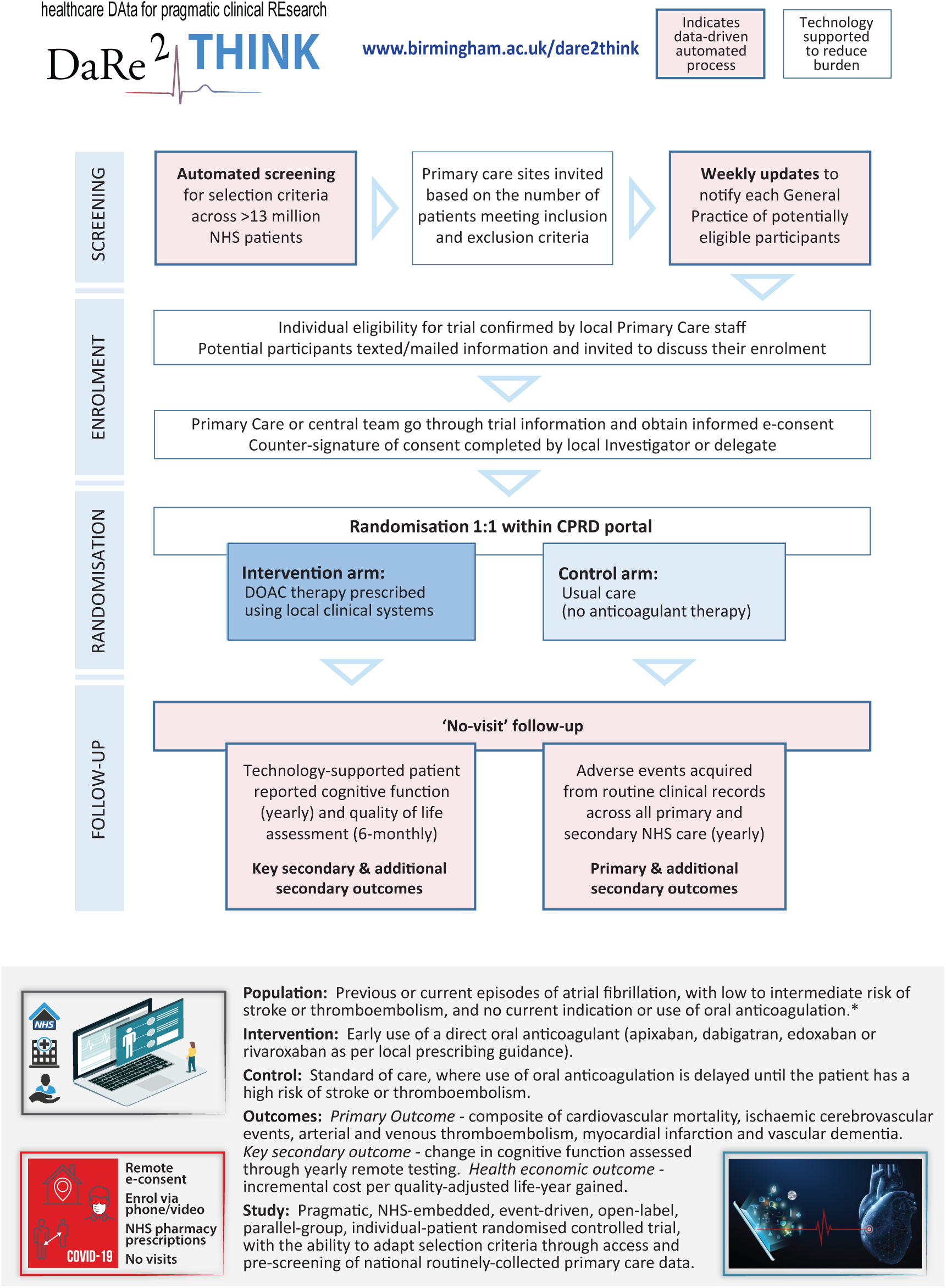
DaRe2THINK key innovations. Flowchart of the DaRe2 approach (healthcare Data for pragmatic clinical Research in the NHS – primary 2 secondary) and summary of the DaRe2THINK clinical trial. * Please see current protocol at www.birmingham.ac.uk/dare2think for full details of inclusion and exclusion criteria. CPRD = Clinical Practice Research Datalink; DOAC = direct oral anticoagulant; NHS = National Health Service.

## SYSTEMATIC CODING METHODOLOGY

The DaRe2 approach links and combines NHS data from primary care, secondary care and other national databases in order to provide a full and complete picture of each participant’s health and healthcare utilisation. The UK public health system is free at the point of delivery to all citizens, with indirect reimbursement via local health authorities based on coded data for diseases and procedures performed. In this section, we detail the processes employed in the DaRe2THINK trial to systematically catalogue and incorporate all relevant codes, including for selection criteria, baseline variables, outcome endpoints. In addition, DaRe2THINK includes monthly automated searches of the primary care record to assess for safety events. For transparency, all codes used are presented in **Appendix 1** for other researchers to see, comment, update and re-use. The DaRe2THINK trial adheres to the CODE-EHR best practice framework for the use of structured electronic healthcare records in clinical research.[2] This study meets all five of the CODE-EHR minimum standards, and will in addition meet all 5 standards for preferred criteria once completed; further details are presented in **Appendix 2**. The DaRe2THINK investigators are committed to the FAIR principles (Findable, Accessible, Interoperable and Reusable).[20]

### Data sources

Primary care data are obtained through CPRD Aurum, a prospectively-collected, population-based, pseudonymised medical record database that collects daily information directly from NHS primary care sites that are part of the CPRD network and use the Egton Medical Information Systems (EMIS) software system. As of March 2021 when DaRe2THINK was initiated, CPRD Aurum included 39,555,354 research acceptable patients of which 13,299,826 were actively registered (19.9% of the UK population) across 1,375 active primary care sites (15.3% of UK general practices). All baseline data for DaRe2THINK trial participants is extracted from CPRD Aurum, obviating the need for investigators to fill in case report forms. CPRD Aurum includes codes for diagnosis and non-prescription data (medcodeid), which is cross-mapped to SNOMED CT (UK edition) [21] and Read Version 2 [22], with prescriptions by product code (prodcodeid) mapped to the Dictionary of Medicines and Devices.[23]

Secondary care Hospital Episode Statistics (HES) Admitted Patient Care data, which contains information on all admissions to NHS hospitals in England, as well as NHS-funded care at independent providers, are linked at the patient level to CPRD Aurum by NHS Digital. With around 99% of hospital activity in England funded by the NHS, this provides almost total coverage of healthcare utilisation for the DaRe2THINK trial, including admission and discharge date, admission type (pre-planned or emergency), primary diagnosis (reason for hospitalisation), secondary diagnoses, and any procedures performed during the hospital stay. HES uses the International Classification of Diseases (ICD) version 10 for diagnosis codes [24], and the Office of Population Censuses and Surveys Classification of Surgical Operation and Procedures (OPCS) version 4 for procedures.[25]

Death and cause of death are obtained via linkage with the Office for National Statistics (ONS) mortality database [26], which includes all deaths in England occurring both within and outside any healthcare setting. Data in ONS is coded using ICD-10.

### Development of the medical code lists

To achieve a comprehensive set of contemporary codes which are suitable for defining outcome and safety events in a clinical trial, we designed and employed a four-phase systematic framework to develop code lists for data extraction from the listed data sources. The approach is summarised in **Figure 2**. Throughout the four phases we used DExtER, an automated platform for clinical epidemiology that was developed to support clinical code list development and validation of phenotypes.[27]

**Figure 2:**
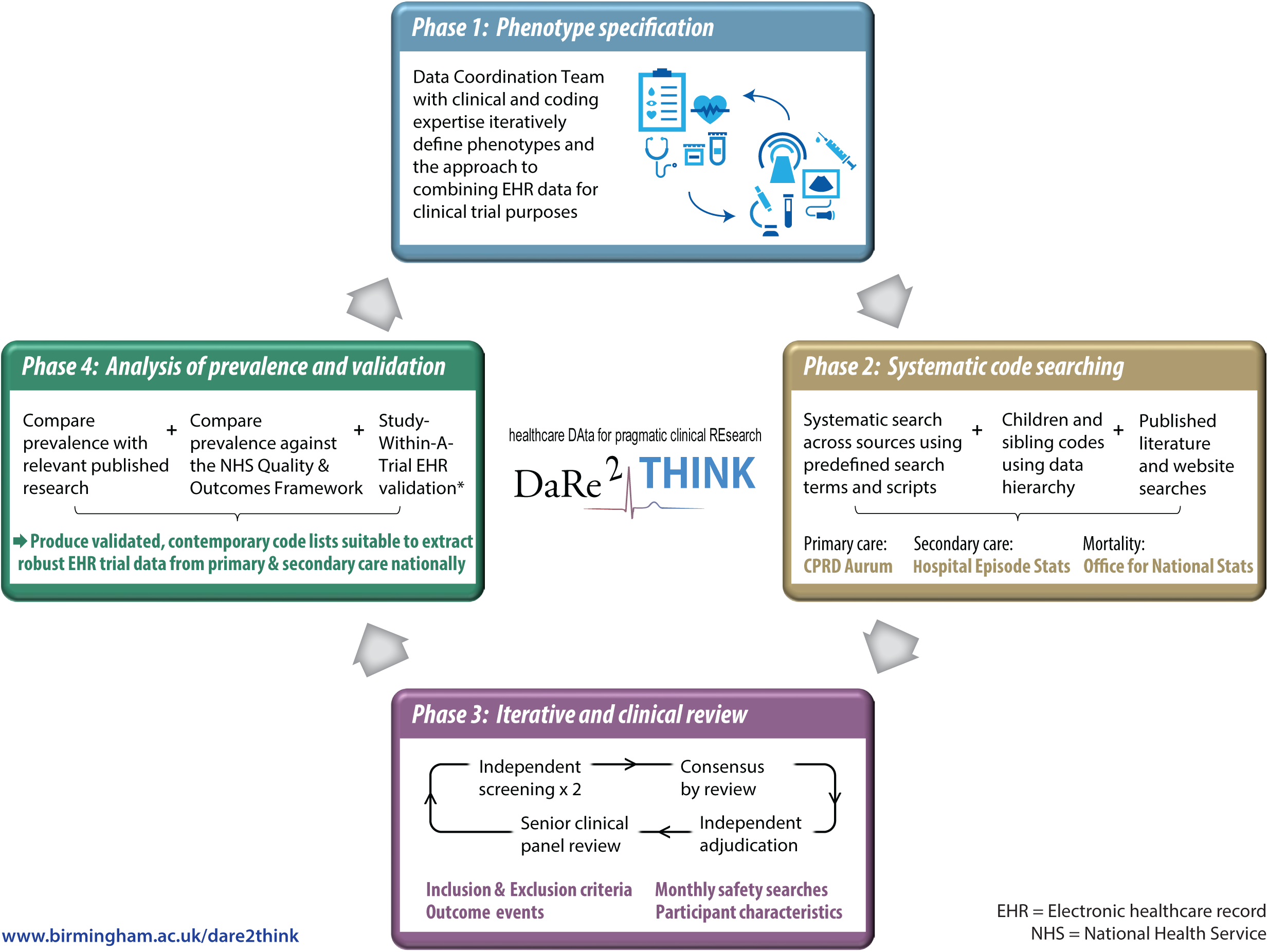
DaRe2THINK coding framework. Systematic, iterative approach to defining code lists from national healthcare data sources. * Study-Within-A-Trial to be performed in a subset of patients with clinical events identified using primary and secondary healthcare records.[28]

#### Phase 1: Phenotype specification

Prior to searching for relevant medical codes, the DaRe2THINK trial selection criteria and outcome measures were first transformed into relevant phenotypes for each data source. This was an iterative process that accounted for the limitations of the data source and, where needed, combined phenotypes to derive a particular outcome. For example, ‘major bleeding’ is not recorded as such in any primary care dataset, hence we combined a phenotype list for bleeding events with a list generated to identify concurrent hospitalisation. This provided a code list for the safety outcome of ‘bleeding resulting in hospitalisation’ based solely on CPRD Aurum data. This process was facilitated by the Data Coordination Team, a specific DaRe2THINK committee which includes clinical expertise in primary and secondary care, data scientists with knowledge and experience in using medical codes, and representatives of CPRD.

#### Phase 2: Systematic code searching

A systematic search was carried out against the agreed phenotypes using the medical code dictionaries (SNOMED CT and Read codes for CPRD Aurum, and ICD-10 codes for HES and ONS). When searching medical code dictionaries, search terms and acronyms were agreed with clinical input. The hierarchy structure of each dictionary was used to find additional relevant children or sibling codes, and these were collated into a dynamic code list. The list was supplemented by any codes identified by other researchers after screening of previously-published and relevant research. To facilitate future use, or if there are updates to coding systems during the trial follow-up period, all search terms and searching scripts are saved as meta data. Medication codes required in the phenotyping algorithms were extracted from the CPRD Aurum product lists based on both British National Formulary (BNF) codes and drug substance names. CPRD provide code lists for demographics and measurements of ethnicity, height, weight, blood pressure, etc.

#### Phase 3: Iterative and clinical review

The codes found in the systematic search went through a multi-stage review process. This included review by data and clinical scientists. Code lists were reviewed independently by two individuals, marking codes for inclusion, exclusion or further review. Discrepancies between the reviewers and any queries were resolved by consensus agreement after adjudication by a senior investigator. All resultant code lists were then further reviewed by senior clinical experts to ensure that code lists were correct and consistent.

#### Phase 4: Analysis of prevalence and validation

For validation purposes, the prevalence of conditions based on the final code lists was then cross-checked with prevalence estimates from relevant publications and the Quality & Outcomes Framework (a national system designed to remunerate general practices for providing good quality care to their patients). Results were discussed with the DaRe2THINK Data Coordination Team, and then suggested changes iteratively fed back to phases 1 through 3 as appropriate. This process will be updated regularly through implementation so that phenotype definitions can be adjusted accordingly, for example if there is a change to coding systems. A Study-Within-A-Trial (SWAT) is also planned in a subset of participants to compare outcome events obtained from coded primary and secondary EHR data with in-depth and granular information extracted from one of the UK’s largest hospital trusts (University Hospitals Birmingham NHS Foundation Trust). Further details are available in the prospectively-registered Medical Research Council SWAT repository [28]; in brief, this will involve extraction of text using machine learning methods from letters, clinical notes, imaging and time-series data to determine accuracy and missingness compared to primary and secondary EHR data.

## RESULTS & FINAL CODE LISTS

Using the systematic approach discussed above, we searched a total of 1,176,611 codes, including 1,159,849 from primary care, and 16,762 from secondary care and national databases. This led to the creation of a total of 77 code lists across the different sections of the trial.

### Participant selection criteria

The DaRe2THINK trial uses automated pre-screening of the primary care EHR to determine inclusion and exclusion according to the defined selection criteria. **Table 2** lists the main components of the selection criteria and associated CPRD Aurum code lists. A total of 3,494 codes were included in these code lists using the systematic approach, including contribution from 14 published code lists.[29, 30] Individual codes for each list can be found in **Appendix 1 Tables S1 to S23**.

**Table 2:** Summary of code lists for selection criteria.

| Selection criterion (CPRD Aurum) | No. of codes | Appendix Table |
| --- | --- | --- |
| Diagnosis of AF (previous, current or chronic) | 43 | S1 |
| Existing use of an anticoagulant |  |  |
| Oral anticoagulants | 54 | S2 |
| Low weight molecular heparin | 80 | S3 |
| Stroke | 120 | S4 |
| Transient ischaemic attack | 11 | S5 |
| Arterial thromboembolism | 81 | S6 |
| Stroke risk factors |  |  |
| Myocardial infarction | 80 | S7 |
| Peripheral arterial disease | 16 | S8 |
| Aortic disease | 4 | S9 |
| Diabetes mellitus | 455 | S10 |
| Antidiabetic drug/insulin | 433 | S11 |
| Hypertension | 85 | S12 |
| Antihypertensive drug | 724 | S13 |
| Heart failure | 54 | S14 |
| Loop diuretic therapy | 59 | S15 |
| Prior major bleeding<br>(intracranial bleed, or requiring hospitalisation) | 161 | S16 |
| High bleeding risk |  |  |
| Gastrointestinal tract ulcer | 234 | S17 |
| Brain injury | 568 | S18 |
| Spinal injury | 1 | S19 |
| Eye injury | 11 | S20 |
| Estimated glomerular filtration rate <30 mL/min | 6 | S21 |
| Azole-antimycotic treatment | 20 | S22 |
| Current diagnosis of dementia | 194 | S23 |

### Outcome events and safety search

Outcome events in the DaRe2THINK trial are collated from primary care, secondary care and national mortality data. Automated safety searches are also performed on a monthly basis in the primary care EHR. **Table 3** provides a summary of the code lists used for outcome events, including 4,434 codes from CPRD Aurum and 610 codes from HES and ONS. For safety events, 3,081 CPRD Aurum codes were used to determine ischaemic stroke, intracranial bleeding, gastrointestinal bleeding and other bleeding, with 483 codes to define associated hospitalisation. Code lists were supplemented after review of 27 code lists from existing published literature [31-44], with the final list of individual codes detailed in **Appendix 1 Tables S24 to S44**.

**Table 3:** Summary of code lists for outcome events.

| Outcome event | CPRD Aurum |  |  | Hospital Episode Statistics |  |  |
| --- | --- | --- | --- | --- | --- | --- |
|  | Number of codes reviewed | Final number of codes | Appendix table | Number of codes reviewed | Final number of codes | Appendix table |
| Cardiovascular mortality | □- | -□ | -□ | 423 | 353 | S24 |
| Ischaemic stroke | 677 | 315 | S25 | 41 | 10 | S26 |
| Transient ischaemic attack | 62 | 44 | S27 | 41 | 30 | S28 |
| Pulmonary and venous thromboembolism | 1759 | 212 | S29 | □ |  | □ |
| Arterial thromboembolism | 1759 | 306 | S30 |  |  | □ |
| Thromboembolic event | □ | □ | □ | 144 | 67 | S31 |
| Myocardial infarction | 215 | 150 | S32 | 22 | 19 | S33 |
| Heart failure | 430 | 100 | S43 | 22 | 10 | S44 |
| Vascular dementia | 252 | 58 | S34 | 6 | 6 | S35 |
| Gastrointestinal bleeding | 902 | 558 | S36 | 42 | 25 | S37 |
| Other bleeding | 4088 | 1642 | S38 | 235 | 59 | S39 |
| Hospitalisation event | 3701 | 483 | S40 | -□ | □- | □- |
| Intracranial bleeding | 1405 | 566 | S41 | 36 | 31 | S42 |

### Baseline characteristics

To remove the need for case report forms, baseline characteristics in the DaRe2THINK trial are extracted from the primary care EHR. **Table 4** provides a summary of these code lists, with a total of 6,802 codes from CPRD Aurum. Measurements and blood results are extracted as the latest value in the past 12 months, medications during the last 12 months, and health conditions throughout the participant’s primary care EHR. For heart failure, hypertension and diabetes, diagnosis is classified in two ways; (1) use of one or more of the relevant codes identified in the EHR, and (2) a ‘secure’ diagnosis based on the code identified *in addition* to an associated medication in the last 12 months (loop diuretic therapy, antihypertensive medications or oral antidiabetic/insulin, respectively). Codes are provided in **Appendix 1 Tables S1 to S77**.

**Table 4:** Summary of code lists for baseline characteristics.

| Characteristic (CPRD Aurum) | Number of codes | Appendix table |
| --- | --- | --- |
| Ethnicity | 327 | S45 |
| Smoking status | 104 | S46 |
| Measurements (height, weight, body mass index, blood pressure, heart rate) | 58 | S47 |
| Blood tests (creatinine, eGFR, haemoglobin, HbA1c, lipids) | 117 | S48 |
| Atrial fibrillation | 43 | S1 |
| Prior atrial fibrillation ablation | 42 | S49 |
| Heart failure | 100 | S43 |
| Hypertension | 85 | S12 |
| Myocardial infarction | 80 | S32 |
| Stroke | 120 | S4 |
| Venous thromboembolism | 212 | S29 |
| Arterial thromboembolism | 306 | S30 |
| Peripheral artery disease & aortic plaque/atherosclerosis | 20 | S8, S9 |
| Diabetes mellitus | 455 | S10 |
| Hyperthyroidism | 73 | S50 |
| Hypothyroidism | 90 | S51 |
| Chronic obstructive pulmonary disease | 191 | S52 |
| Eye-related diseases & procedure | 293 | S53-S60 |
| GI bleeding | 558 | S36 |
| Intracranial bleeding | 566 | S41 |
| Other bleeding | 1642 | S38 |
| Hospitalisation | 483 | S40 |
| Antiplatelet therapy | 79 | S61 |
| Diuretics | 243 | S15, S62-S64 |
| Calcium channel blockers | 267 | S65, S66 |
| ACE inhibitors | 54 | S67 |
| Angiotensin II receptor antagonists | 116 | S68 |
| Beta-blockers | 115 | S69 |
| Alpha-blockers | 58 | S70 |
| Aldosterone receptor antagonist | 32 | S71 |
| Other antihypertensive drugs | 76 | S72 |
| Antiarrhythmic drugs (Class 1 and 3) | 94 | S73, S74 |
| Digoxin | 14 | S75 |
| Sodium-glucose transport protein 2 inhibitors | 34 | S76 |
| Non-steroidal anti-inflammatory drugs | 86 | S77 |

## DISCUSSION

The DaRe2 framework was designed to link together a series of innovations for a digital clinical trial, creating a patient-centred approach with high quality output, yet low burden on healthcare staff. The ability to pre-screen from over 13 million NHS patient records allows for targeted and efficient recruitment, and simplifies enrolment of patients that are representative of the real-world population. Combining all health data from primary and secondary care sources nationally enables a ‘remote’ RCT, with no requirement for baseline or follow-up visits. These processes are supported by advances in digital technology, including remote e-consent and patient reported outcomes. The DaRe2THINK trial is using this sequence of innovations to test a hypothesis of critical importance to public health, with the aim of preventing the long-term consequences of atrial fibrillation, including cognitive decline and vascular dementia.

This paper outlines our approach to defining phenotypes that support all aspects of the trial, from understanding the characteristics of recruited participants, through to determination of clinical endpoints. Unlike conventional trials with case report forms, DaRe2THINK extracts all relevant information from structured and coded healthcare data sources. Hence, it is critical for dissemination and transparency that all coding schema for this trial are pre-published and available for evaluation (plus re-use) by other researchers and clinicians. In a published review of 450 EHR-based studies, only 19 (5.1%) were accompanied by a full set of clinical codes [45], severely limiting the value of those studies and their implementation to routine care. The emergence of EHR systems across the world provides an important opportunity for healthcare-embedded clinical research [1], but only when this is accompanied by a clear pathway from data collection to interpretation.[2]

The broad scope of coded healthcare data has been a limiting factor in prior attempts at EHR-embedded clinical trials. As evidenced by the large number of codes screened for this trial (over a million), a robust and systematic approach is required to ensure that patient events are not missed. The UK NHS is ideally suited to operationalise this sort of innovative trial, as structured healthcare data is used for quality and reimbursement purposes [4] and there is an ability to link across different healthcare sources.[46] Linkage is critical to understanding the full healthcare utilisation of each participant.[3] In this case allowing for the combination of historical and future primary care data, with that pertaining to hospitalisation and procedures across the NHS nationally, as well as information on cause of death from the national register. EHR-based trials may have an advantage in this regard compared to traditional trials, where investigators are relied on to document safety and outcome events. In the modern era of multiple heath providers for a multitude of different health conditions, sourcing safety and outcome events from national data has the potential to avoid missing endpoints, and potentially eliminate ascertainment bias in unblinded research. Although the value of trials based on routine EHR data is likely to vary depending on the disease being researched [47], it is clear that the direction of travel is to enhance existing clinical trial infrastructure with digital innovations.[48] In this regard, DaRe2THINK is the vanguard for cost-efficient RCTs embedded in clinical care. Previous trials have shown that although endpoints based on structured healthcare data may not replicate the traditional adjudication committee output, the estimated intervention effect is almost identical for adjudicated compared to EHR follow-up.[49, 50].

There remain substantial limitations to effective use of EHR data for clinical trials, including ongoing changes to coding systems. For example, ICD-10 includes 68,000 codes (a number of countries have their own editions, with ICD-11 launched this year), and SNOMED CT has over 300,000 clinical concepts. New codes can also be created within primary healthcare databases, highlighting the importance of iterative review and updates as exemplified by our coding framework. Universal limitations, such as missing data, variation in data quality, reliability of coding and imprecision in the choice of code used, are common across real-world data sources. These and specific areas of bias such as lack of representativeness and loss to follow-up are substantially minimised in DaRe2THINK due to the NHS being the de facto provider of almost all healthcare in the UK, and CPRD providing broad and representative coverage. In this setting, concordance between primary and secondary care health data is high. For example, in a random sample of 50,000 patients in CPRD Aurum, 94% of the 1,260 patients with a code for myocardial infarction had corroborating evidence in the secondary care HES records.[3] Similar to our transparent approach to coding, the process of integrating primary and secondary healthcare sources in DaRe2THINK will be documented in an open-access, prospectively-published Statical Analysis Plan.

The DaRe2 approach was only possible with the support and guidance of our Patient and Public Involvement (PPI) team. We are indebted to the group for their constructive criticism throughout the design process, and their ongoing contribution to trial management to produce sustainable systems focused on the needs and wants of patients. Achieving a social licence for data-related clinical research is critical, and only achievable with high-level involvement by the public, for example using the PPI-POSITIVE approach.[8]

## CONCLUSION

The DaRe2THINK trial is using a series of digital innovations to reshape the deployment of an adaptive randomised clinical trial embedded in routine healthcare, with minimal burden for staff and patients. Integrating national healthcare data from primary and secondary sources, the system will provide evidence for a key public health concern, the prevention of cognitive decline and dementia in the rapidly growing number of patients with atrial fibrillation. Systematic and transparent methodology in the use of structured healthcare data, together with a patient and public mandate, are critical for evolution of these novel approaches in order to improve healthcare in our communities.

## Supporting information

ICMJE

## Data Availability

No data is presented in the manuscript - this paper provides all codes used in the trial (see appendix)

https://www.birmingham.ac.uk/d2t

## Authors’ contributions

The coding framework was designed and implemented by XW, OT, KO, KN, DS and DK. The manuscript was drafted by XW, ARM and DK, with all other authors editing the manuscript for intellectual content. DK provided supervision and was responsible for the decision to submit the manuscript.

## Data Sharing

No additional data are available at this time.

## Acknowledgements

In memory of Jaqueline Jones, member of the Patient and Public Involvement team. We would like to acknowledge all members of the DaRe2THINK team and trial committees:

**Trial Management Group:** Susan Beatty (Clinical Practice Research Datalink, MHRA), Samir Mehta (Birmingham Clinical Trials Unit), Sophie Breeze (Clinical Practice Research Datalink, MHRA), Karen Lancaster, (Clinical Practice Research Datalink, MHRA), Stuart Fordyce (Clinical Practice Research Datalink, MHRA).

**Data Coordination Team:** Naomi Allen (University of Oxford, UK Biobank), Melanie Calvert (Centre for Patient Reported Outcomes Research & Birmingham Health Partners Centre for Regulatory Science and Innovation), Alastair Denniston (University Hospitals Birmingham NHS Foundation Trust & University of Birmingham), George Gkoutos (University of Birmingham & University Hospitals Birmingham NHS Foundation Trust).

**Health Economics Team:** Sahan Jayawardana (London School of Economics and Political Science), Elias Mossialos (London School of Economics and Political Science).

**Expert Advisory Group:** Simon Ball (University Hospitals Birmingham NHS Foundation Trust & Health Data Research UK Midlands), Colin Baigent (University of Oxford), Peter Brocklehurst (Birmingham Clinical Trials Unit), Will Lester (University Hospitals Birmingham NHS Foundation Trust), Richard McManus (University of Oxford), Stefano Seri (Aston University), Janet Valentine (Innovate UK, UK Research & Innovation).

**Independent Trial Steering Committee:** A John Camm (St. George’s University of London), Sandra Haynes MBE (Patient and Public Involvement team lead), Dame Julie Moore (University of Warwick), Amy Rogers (University of Dundee), Mary Stanbury (Patient & Public Involvement).

**Independent Data Monitoring Committee:** Marcus Flather (University of East Anglia), Suzy Walker (General Practitioner, Fortrose Medical Practice, Highland), Duolao Wang (Liverpool School of Tropical Medicine).

