## Appendix 1 for "Systematic approach to outcome assessment from coded electronic healthcare records in the DaRe2THINK NHS-embedded randomised trial"

healthcare DAta for pragmatic clinical REsearch

DaRe2THINK

### Appendix 1: DaRe2THINK code lists


**Chapter 1. Code lists for selection criteria** ... **3**

Table S1. Atrial fibrillation Aurum codes ... 3

Table S2. Anticoagulant Aurum product codes ... 5

Table S3. Low molecular weight heparin Aurum product codes ... 7

Table S4. Stroke Aurum codes ... 10

Table S5. Transient ischaemic attack selection Aurum codes ... 13

Table S6. Arterial thromboembolism Aurum codes ... 14

Table S7. Myocadial infarction Aurum codes ... 17

Table S8. Periperal arterial disease Aurum codes ... 20

Table S9. Aortic plaque Aurum codes ... 21

Table S10. Diabetes Aurum codes ... 22

Table S11. Anti-Diabetic Aurum product codes ... 35

Table S12. Hypertension Aurum codes ... 51

Table S13. Anti-Hypertension Aurum product codes ... 54

Table S14. Heart failure selection Aurum codes ... 77

Table S15. Loop Diuretics Aurum product codes ... 79

Table S16. Gastrointestinal and intracranial Bleeding Aurum codes ... 82

Table S17. Gastrointestinal tract ulcer Aurum codes ... 87

Table S18. Brain injury Aurum codes ... 94

Table S19. Spinal Injury Aurum codes ... 114

Table S20. Eye Injury Aurum codes ... 115

Table S21. Estimated glomerular filtration rate Aurum codes ... 116

Table S22. Azole Aurum codes ... 117

Table S23. Dementia Aurum codes ... 118

**Chapter 2. Code lists for trial outcomes** ... **124**

Table S24. Cardivascular mortality HES (ICD10) codes ... 124

Table S25. Ischaemic stroke Aurum codes ... 125

Table S26. Ischaemic stroke HES (ICD10) codes ... 128

Table S27. Transient ischaemic attack outcome Aurum codes ... 129

Table S28. Transient ischaemic attack HES (ICD10) codes ... 131

Table S29. Venous thromoemoblism and pulmonary embolism Aurum codes ... 132

Table S30. Arterial thromboembolic Aurum codes ... 145

Table S31. Thromboembolic HES (ICD10) codes ... 153

Table S32. Myocardial infarction outcome Aurum codes ... 155

Table S33. Myocardial infarction HES (ICD10) codes ... 159

Table S34. Vascular dementia Aurum codes ... 160

Table S35. Vascular dementia HES (ICD10) codes ... 162

Table S36. Gastrointestinal bleeding Aurum codes ... 163

Table S37. Gastrointestinal bleeding HES (ICD10) codes 168
Table S38. Bleeding at other anatomy sites Aurum codes 169
Table S39. Bleeding at other anatomy sites HES (ICD10) codes 188
Table S40. Hospitalisation Aurum codes 190
Table S41. Intracranial haemorrhage Aurum codes 199
Table S42. Intracranial haemorrhage HES (ICD10) codes 205
Table S43. Heart failure outcome Aurum codes 206
Table S44. Heart failure HES (ICD10) codes 209
**Chapter 3. Code lists for baseline characteristics 210**
Table S45. Ethinicity Aurum codes 210
Table S46. Smoking Aurum codes 219
Table S47. Measurements Aurum codes 222
Table S48. Blood test Aurum codes 224
Table S49. Atrial ablation Aurum codes 228
Table S50. Hyperthyoidism Aurum codes 230
Table S51. Hypothyroidism Aurum codes 232
Table S52. Chronic obstructive pulmonary disease Aurum codes 235
Table S53. Diabetic retinopathy Aurum codes 241
Table S54. Retinal vascular occlusion Aurum codes 243
Table S55. Glaucoma Aurum codes 244
Table S56. Age-related macular degeneration Aurum codes 248
Table S57. Retinal Laser Aurum codes 249
Table S58. Vitrectomy Aurum codes 250
Table S59. Scleral buckle Aurum codes 251
Table S60. Intravitreal injection Aurum codes 252
Table S61. Antiplatelet Aurum product codes 253
Table S62. Thiazide diuretics Aurum product codes 256
Table S63. Thiazide like diuretics Aurum product codes 261
Table S64. Potassium sparing diuretics Aurum product codes 262
Table S65. Dihydropyridines Aurum product codes 264
Table S66. Non Dihydropyridines Aurum product codes 269
Table S67. Angiotensin converting enzyme inhibitors Aurum product codes 273
Table S68. Angiotensin II receptor blocker Aurum product codes 275
Table S69. Beta blockers Aurum product codes 279
Table S70. Alpha blockers Aurum product codes 283
Table S71. Aldosterone Aurum product codes 285
Table S72. Other antihypertensive Aurum product codes 286
Table S73. Antiarrhythmic drugs class 1 Aurum product codes 289
Table S74. Antiarrhythmic drugs class 3 Aurum product codes 292
Table S75. Digoxin Aurum product codes 293
Table S76. Sodium-glucose co-transporter-2 inhibitors Aurum product codes 297
Table S77. Nonsteroidal anti-inflammatory drug Aurum product codes 299

### Chapter 1. Code lists for selection criteria

#### Table S1. Atrial fibrillation Aurum codes

| Term | MedCode ID | SNOMED CT Concept ID | SNOMED CT DescriptionID |
|---|---|---|---|
| AF - Atrial fibrillation | 3299911000006116 | 49436004 | 1230726010 |
| AF - Paroxysmal atrial fibrillation | 5669591000006113 | 282825002 | 421232012 |
| Atrial fibrillation | 82343012 | 49436004 | 82343012 |
| Atrial fibrillation and flutter | 300130013 | 195080001 | 300130013 |
| Atrial fibrillation and flutter NOS | 300132017 | 195080001 | 300130013 |
| Atrial fibrillation annual review | 406861000000119 | 248411000000105 | 406861000000119 |
| Atrial fibrillation confirmed | 1823951000006111 | 1823951000006107 | 1823951000006111 |
| Atrial fibrillation detected | 8439861000006115 | 1066831000000104 | 2679661000000115 |
| Atrial fibrillation monitoring first letter | 408081000000116 | 717221000000101 | 1569761000000112 |
| Atrial fibrillation monitoring invitation | 8214691000006119 | 711411000000101 | 1555841000000110 |
| Atrial fibrillation monitoring second letter | 408101000000110 | 716981000000106 | 1569531000000111 |
| Atrial fibrillation monitoring telephone invitation | 8218281000006110 | 716721000000107 | 1569271000000111 |
| Atrial fibrillation monitoring telephone invite | 408241000000115 | 716721000000107 | 1570381000000119 |
| Atrial fibrillation monitoring third letter | 408121000000118 | 716181000000109 | 1568741000000115 |
| Atrial fibrillation monitoring verbal invitation | 8218591000006111 | 717011000000100 | 1569561000000118 |
| Atrial fibrillation monitoring verbal invite | 408141000000113 | 717011000000100 | 1570401000000119 |
| Atrial fibrillation resolved | 294601000000110 | 196371000000102 | 294601000000110 |
| Atrial fibrillation with rapid ventricular response | 8037691000006113 | 120041000119109 | 2981207015 |
| Chronic atrial fibrillation | 2675253013 | 426749004 | 2675253013 |
| Controlled atrial fibrillation | 5887651000006116 | 300996004 | 442207015 |
| ECG: atrial fibrillation | 256478018 | 164889003 | 256478018 |
| Electrocardiographic atrial fibrillation | 4586611000006112 | 164889003 | 3300048013 |
| Evaluation of AF (atrial fibrillation) burden | 12734511000006115 | 143601000000104 | 1628691000000119 |
| Evaluation of atrial fibrillation rate control | 12736851000006112 | 1127481000000104 | 1855311000000119 |
| H/O: atrial fibrillation | 456154015 | 312442005 | 456154015 |
| History of atrial fibrillation | 6016401000006115 | 312442005 | 2986292016 |
| History of maze procedure for atrial fibrillation | 7100011000006116 | 429218009 | 2695416012 |
| Insertion of pacemaker for control of atrial fibrillation | 7375931000006117 | 449863006 | 2912517016 |
| Intermittent atrial fibrillation | 5669611000006119 | 282825002 | 421234013 |
| Lone atrial fibrillation | 5057261000006110 | 233910005 | 350464013 |
| Longstanding persistent atrial fibrillation | 7610331000006110 | 706923002 | 3027950014 |
| Maze procedure for atrial fibrillation | 7099921000006111 | 429211003 | 2694655012 |
| Non-rheumatic atrial fibrillation | 350465014 | 233911009 | 350465014 |
| NRAF - Non-rheumatic atrial fibrillation | 5057281000006117 | 233911009 | 350466010 |
| PAF - Paroxysmal atrial fibrillation | 5669601000006117 | 282825002 | 421233019 |
| Paroxysmal atrial fibrillation | 421235014 | 282825002 | 421235014 |

| | | | |
|---|---|---|---|
| Permanent atrial fibrillation | 636721000000112 | 440028005 | 2793259018 |
| Persistent atrial fibrillation | 636701000000115 | 440059007 | 2793372019 |
| QOF (Quality and Outcomes Framework) atrial fibrillation quality indicator-related care invitation | 12626271000006110 | 1110851000000100 | 2779681000000117 |
| QOF (Quality and Outcomes Framework) atrial fibrillation quality indicator-related care invitation using preferred method of communication | 12734281000006112 | 133111000000106 | 1628151000000119 |
| Rapid atrial fibrillation | 6043781000006115 | 314208002 | 458527016 |
| Transient cerebral ischaemia due to atrial fibrillation | 7064691000006118 | 426814001 | 2675245017 |
| Transient cerebral ischemia due to atrial fibrillation | 7064701000006118 | 426814001 | 2675244018 |

#### Table S2. Anticoagulant Aurum product codes

| Term from EMIS | Prod code ID | dmd ID | Drug substance name |
|---|---|---|---|
| warfarin wbp tablets | 1531941000033113 | | |
| warfarin wbp tablets 1 mg | 1532041000033119 | | |
| warfarin wbp tablets 3 mg | 1532141000033115 | | |
| warfarin wbp tablets 5 mg | 1532241000033110 | | |
| warfarin (evans) tablets 3 mg | 1532341000033117 | | |
| sinthrome tablets 4 mg | 1337441000033110 | | |
| warfarin 1mg tablets | 1531641000033118 | 319733000 | Warfarin sodium |
| marevan 1mg tablets (advanz pharma) | 868141000033114 | 933311000001105 | Warfarin sodium |
| marevan 3mg tablets (advanz pharma) | 868241000033119 | 284011000001104 | Warfarin sodium |
| warfarin 3mg tablets | 1531741000033110 | 319734006 | Warfarin sodium |
| warfarin 5mg tablets | 1531841000033117 | 319735007 | Warfarin sodium |
| marevan 5mg tablets (advanz pharma) | 868341000033112 | 461911000001100 | Warfarin sodium |
| marevan 500microgram tablets (advanz pharma) | 1819741000033117 | 204811000001109 | Warfarin sodium |
| warfarin 500microgram tablets | 1819641000033114 | 319736008 | Warfarin sodium |
| acenocoumarol 1mg tablets | 3097541000033112 | 319740004 | Acenocoumarol |
| sinthrome 1mg tablets (norgine pharmaceuticals ltd) | 1337341000033116 | 738511000001103 | Acenocoumarol |
| phenindione 10mg tablets | 1081541000033110 | 319745009 | Phenindione |
| phenindione 25mg tablets | 1082341000033112 | 319746005 | Phenindione |
| phenindione 50mg tablets | 1082441000033118 | 319747001 | Phenindione |
| coumadin 4mg tablets (imported (canada)) | 6112441000033111 | 18509011000001102 | Warfarin sodium |
| apixaban 2.5mg tablets | 6444441000033114 | 703907006 | Apixaban |
| eliquis 2.5mg tablets (bristol-myers squibb pharmaceuticals ltd) | 6444541000033110 | 19506911000001105 | Apixaban |
| eliquis 5mg tablets (bristol-myers squibb pharmaceuticals ltd) | 8233041000033112 | 21677511000001105 | Apixaban |
| apixaban 5mg tablets | 8232941000033119 | 703908001 | Apixaban |
| warfarin 10mg/5ml oral suspension | 2639041000033119 | 8798011000001109 | Warfarin sodium |
| warfarin 1mg/5ml oral solution | 6000541000033119 | 8798111000001105 | Warfarin sodium |
| warfarin 1mg/5ml oral suspension | 6000841000033117 | 8798211000001104 | Warfarin sodium |
| warfarin 3mg/5ml oral solution | 6000641000033118 | 8798511000001101 | Warfarin sodium |
| warfarin 3mg/5ml oral suspension | 6000941000033113 | 8798611000001102 | Warfarin sodium |
| warfarin 5mg/5ml oral solution | 6000741000033110 | 8798711000001106 | Warfarin sodium |
| warfarin 5mg/5ml oral suspension | 2620141000033114 | 8798811000001103 | Warfarin sodium |
| dabigatran etexilate 110mg capsules | 4500341000033115 | 13532811000001109 | Dabigatran etexilate mesilate |
| pradaxa 110mg capsules (boehringer ingelheim ltd) | 4500541000033110 | 13505411000001109 | Dabigatran etexilate mesilate |
| dabigatran etexilate 75mg capsules | 4500241000033113 | 13532911000001104 | Dabigatran etexilate mesilate |
| pradaxa 75mg capsules (boehringer ingelheim ltd) | 4500441000033114 | 13504711000001102 | Dabigatran etexilate mesilate |
| rivaroxaban 10mg tablets | 4656341000033115 | 14254711000001104 | Rivaroxaban |
| xarelto 10mg tablets (bayer plc) | 4656441000033114 | 14237311000001106 | Rivaroxaban |
| warfarin 1mg/ml oral suspension sugar free | 6066441000033118 | 18290011000001102 | Warfarin sodium |

| | | | |
|---|---|---|---|
| dabigatran etexilate 150mg capsules | 6436141000033111 | 19469811000001101 | Dabigatran etexilate mesilate |
| pradaxa 150mg capsules (boehringer ingelheim ltd) | 6436241000033116 | 19465811000001104 | Dabigatran etexilate mesilate |
| rivaroxaban 15mg tablets | 6511341000033117 | 19842111000001101 | Rivaroxaban |
| xarelto 15mg tablets (bayer plc) | 6511541000033112 | 19840811000001107 | Rivaroxaban |
| xarelto 20mg tablets (bayer plc) | 6511641000033113 | 19841411000001101 | Rivaroxaban |
| rivaroxaban 20mg tablets | 6511441000033111 | 19842211000001107 | Rivaroxaban |
| xarelto 2.5mg tablets (bayer plc) | 9704641000033115 | 27160311000001106 | Rivaroxaban |
| rivaroxaban 2.5mg tablets | 9704541000033116 | 27810711000001104 | Rivaroxaban |
| lixiana 15mg tablets (daiichi sankyo uk ltd) | 10493841000033112 | 29902111000001100 | Edoxaban tosilate |
| edoxaban 15mg tablets | 10493541000033110 | 29903211000001100 | Edoxaban tosilate |
| edoxaban 30mg tablets | 10493641000033111 | 29903311000001108 | Edoxaban tosilate |
| lixiana 30mg tablets (daiichi sankyo uk ltd) | 10493941000033116 | 29902411000001105 | Edoxaban tosilate |
| lixiana 60mg tablets (daiichi sankyo uk ltd) | 10494041000033119 | 29902711000001104 | Edoxaban tosilate |
| edoxaban 60mg tablets | 10493741000033119 | 29903411000001101 | Edoxaban tosilate |
| xarelto 15mg / 20mg treatment initiation pack (bayer plc) | 12407141000033118 | 34793111000001102 | |
| rivaroxaban 15mg tablets and rivaroxaban 20mg tablets | 12407041000033117 | 34819111000001102 | |

#### Table S3. Low molecular weight heparin Aurum product codes

| Term from EMIS | Prod code ID | dmd ID | Drug substance name |
|---|---|---|---|
| Clexane 300mg/3ml solution for injection multidose vials (Sanofi) | 4153241000033118 | 11507011000001102 | Enoxaparin sodium |
| Enoxaparin sodium 300mg/3ml solution for injection vials | 4153141000033113 | 11509311000001109 | Enoxaparin sodium |
| Inhixa 300mg/3ml solution for injection multidose vials (Techdow Pharma England Ltd) | 13711341000033114 | 38232211000001100 | Enoxaparin sodium |
| Clexane Forte 120mg/0.8ml solution for injection pre-filled syringes (Sanofi) | 2732541000033114 | 4198711000001100 | Enoxaparin sodium |
| Enoxaparin sodium 120mg/0.8ml solution for injection pre-filled syringes | 2041741000033113 | 36563911000001108 | Enoxaparin sodium |
| Inhixa 120mg/0.8ml solution for injection pre-filled syringes (Techdow Pharma England Ltd) | 12988541000033114 | 36784211000001100 | Enoxaparin sodium |
| Arovi 120mg/0.8ml solution for injection pre-filled syringes (ROVI Biotech Ltd) | 12584741000033119 | 35559211000001107 | Enoxaparin sodium |
| Clexane 100mg/1ml solution for injection pre-filled syringes (Sanofi) | 274941000033116 | 4199111000001108 | Enoxaparin sodium |
| Arovi 100mg/1ml solution for injection pre-filled syringes (ROVI Biotech Ltd) | 12584641000033111 | 35566911000001104 | Enoxaparin sodium |
| Inhixa 100mg/1ml solution for injection pre-filled syringes (Techdow Pharma England Ltd) | 12373141000033110 | 34701011000001102 | Enoxaparin sodium |
| Enoxaparin Becat 100mg/1ml solution for injection pre-filled syringes (ROVI Biotech Ltd) | 12476641000033118 | 35084011000001109 | Enoxaparin sodium |
| Enoxaparin sodium 100mg/1ml solution for injection pre-filled syringes | 516641000033117 | 36564011000001106 | Enoxaparin sodium |
| Clexane 40mg/0.4ml solution for injection pre-filled syringes (Sanofi) | 276541000033115 | 3825411000001104 | Enoxaparin sodium |
| Enoxaparin sodium 40mg/0.4ml solution for injection pre-filled syringes | 517141000033112 | 36565711000001107 | Enoxaparin sodium |
| Enoxaparin Becat 40mg/0.4ml solution for injection pre-filled syringes (ROVI Biotech Ltd) | 12476241000033116 | 35083411000001103 | Enoxaparin sodium |
| Inhixa 40mg/0.4ml solution for injection pre-filled syringes (Techdow Pharma England Ltd) | 12372841000033114 | 34700211000001101 | Enoxaparin sodium |
| Arovi 40mg/0.4ml solution for injection pre-filled syringes (ROVI Biotech Ltd) | 12585041000033116 | 35569011000001109 | Enoxaparin sodium |
| Clexane 60mg/0.6ml solution for injection pre-filled syringes (Sanofi) | 275041000033116 | 4199811000001101 | Enoxaparin sodium |
| Enoxaparin sodium 60mg/0.6ml solution for injection pre-filled syringes | 516741000033114 | 38895011000001100 | Enoxaparin sodium |
| Enoxaparin Becat 60mg/0.6ml solution for injection pre-filled syringes (ROVI Biotech Ltd) | 12476341000033114 | 35083611000001100 | Enoxaparin sodium |
| Inhixa 60mg/0.6ml solution for injection pre-filled syringes (Techdow Pharma England Ltd) | 12372941000033118 | 34700511000001103 | Enoxaparin sodium |
| Arovi 60mg/0.6ml solution for injection pre-filled syringes (ROVI Biotech Ltd) | 12585141000033117 | 35568611000001106 | Enoxaparin sodium |
| Clexane 80mg/0.8ml solution for injection pre-filled syringes (Sanofi) | 275141000033117 | 4199511000001104 | Enoxaparin sodium |
| Arovi 80mg/0.8ml solution for injection pre-filled syringes (ROVI Biotech Ltd) | 12585241000033112 | 35567911000001101 | Enoxaparin sodium |
| Inhixa 80mg/0.8ml solution for injection pre-filled syringes (Techdow Pharma England Ltd) | 12373041000033111 | 34700811000001100 | Enoxaparin sodium |
| Enoxaparin Becat 80mg/0.8ml solution for injection pre-filled syringes (ROVI Biotech Ltd) | 12476441000033115 | 35083811000001101 | Enoxaparin sodium |
| Enoxaparin sodium 80mg/0.8ml solution for injection pre-filled syringes | 516841000033116 | 38895111000001104 | Enoxaparin sodium |
| Clexane Forte 150mg/1ml solution for injection pre-filled syringes (Sanofi) | 2732641000033110 | 4198211000001107 | Enoxaparin sodium |
| Enoxaparin sodium 150mg/1ml solution for injection pre-filled syringes | 2041941000033111 | 38895211000001105 | Enoxaparin sodium |

| | | | |
|---|---|---|---|
| Arovi 150mg/1ml solution for injection pre-filled syringes (ROVI Biotech Ltd) | 12584841000033112 | 35559411000001106 | Enoxaparin sodium |
| Inhixa 150mg/1ml solution for injection pre-filled syringes (Techdow Pharma England Ltd) | 12988641000033110 | 36784411000001101 | Enoxaparin sodium |
| Clexane 20mg/0.2ml solution for injection pre-filled syringes (Sanofi) | 276441000033116 | 3826311000001101 | Enoxaparin sodium |
| Enoxaparin sodium 20mg/0.2ml solution for injection pre-filled syringes | 517041000033113 | 38896011000001109 | Enoxaparin sodium |
| Arovi 20mg/0.2ml solution for injection pre-filled syringes (ROVI Biotech Ltd) | 12584941000033116 | 35569511000001101 | Enoxaparin sodium |
| Enoxaparin Becat 20mg/0.2ml solution for injection pre-filled syringes (ROVI Biotech Ltd) | 12476141000033111 | 35083211000001102 | Enoxaparin sodium |
| Inhixa 20mg/0.2ml solution for injection pre-filled syringes (Techdow Pharma England Ltd) | 12372741000033116 | 34699911000001109 | Enoxaparin sodium |
| Tinzaparin sodium 12,000units/0.6ml solution for injection pre-filled syringes | 10046341000033113 | 28566111000001107 | Tinzaparin sodium |
| Innohep 12,000units/0.6ml solution for injection pre-filled syringes (LEO Pharma) | 10046641000033117 | 28499911000001109 | Tinzaparin sodium |
| Innohep 16,000units/0.8ml solution for injection pre-filled syringes (LEO Pharma) | 10046741000033114 | 28498711000001103 | Tinzaparin sodium |
| Tinzaparin sodium 16,000units/0.8ml solution for injection pre-filled syringes | 10046441000033119 | 28566311000001109 | Tinzaparin sodium |
| Tinzaparin sodium 8,000units/0.4ml solution for injection pre-filled syringes | 10046241000033115 | 28566511000001103 | Tinzaparin sodium |
| Innohep 8,000units/0.4ml solution for injection pre-filled syringes (LEO Pharma) | 10046541000033118 | 28501011000001100 | Tinzaparin sodium |
| Innohep 10,000units/0.5ml solution for injection pre-filled syringes (LEO Pharma) | 763041000033118 | 3918011000001100 | Tinzaparin sodium |
| Tinzaparin sodium 10,000units/0.5ml solution for injection pre-filled syringes | 1440541000033116 | 35918811000001101 | Tinzaparin sodium |
| Innohep 14,000units/0.7ml solution for injection pre-filled syringes (LEO Pharma) | 763141000033119 | 3919011000001105 | Tinzaparin sodium |
| Tinzaparin sodium 14,000units/0.7ml solution for injection pre-filled syringes | 1440641000033115 | 35918911000001106 | Tinzaparin sodium |
| Innohep 18,000units/0.9ml solution for injection pre-filled syringes (LEO Pharma) | 762941000033111 | 3920011000001106 | Tinzaparin sodium |
| Tinzaparin sodium 18,000units/0.9ml solution for injection pre-filled syringes | 1440741000033112 | 35919011000001102 | Tinzaparin sodium |
| Innohep 2,500units/0.25ml solution for injection pre-filled syringes (LEO Pharma) | 2149141000033117 | 4120211000001102 | Tinzaparin sodium |
| Tinzaparin sodium 2,500units/0.25ml solution for injection pre-filled syringes | 2149041000033116 | 35919111000001101 | Tinzaparin sodium |
| Innohep 20,000units/2ml solution for injection vials (LEO Pharma) | 1781941000033119 | 4121311000001100 | Tinzaparin sodium |
| Tinzaparin sodium 20,000units/2ml solution for injection vials | 1781841000033110 | 35919211000001107 | Tinzaparin sodium |
| Innohep 3,500units/0.35ml solution for injection pre-filled syringes (LEO Pharma) | 1620241000033118 | 3916311000001101 | Tinzaparin sodium |
| Tinzaparin sodium 3,500units/0.35ml solution for injection pre-filled syringes | 1700641000033119 | 35919311000001104 | Tinzaparin sodium |
| Innohep 4,500units/0.45ml solution for injection pre-filled syringes (LEO Pharma) | 1620341000033111 | 4120611000001100 | Tinzaparin sodium |
| Tinzaparin sodium 4,500units/0.45ml solution for injection pre-filled syringes | 1700741000033111 | 35919411000001106 | Tinzaparin sodium |
| Innohep 40,000units/2ml solution for injection vials (LEO Pharma) | 762041000033110 | 4121711000001101 | Tinzaparin sodium |
| Tinzaparin sodium 40,000units/2ml solution for injection vials | 1439841000033110 | 35919511000001105 | Tinzaparin sodium |
| Dalteparin sodium 7,500units/0.3ml solution for injection pre-filled syringes | 2640441000033116 | 4865111000001103 | Dalteparin sodium |
| Fragmin 7,500units/0.3ml solution for injection pre-filled syringes (Pfizer Ltd) | 2640341000033110 | 4864211000001108 | Dalteparin sodium |

| | | | |
|---|---|---|---|
| Fragmin 10,000units/1ml solution for injection ampoules (Pfizer Ltd) | 610941000033116 | 3968911000001100 | Dalteparin sodium |
| Dalteparin sodium 10,000units/1ml solution for injection ampoules | 404041000033110 | 36092911000001101 | Dalteparin sodium |
| Fragmin 10,000units/1ml solution for injection pre-filled syringes (Pfizer Ltd) | 1781641000033114 | 3970311000001105 | Dalteparin sodium |
| Dalteparin sodium 10,000units/1ml solution for injection pre-filled syringes | 1781541000033113 | 36093011000001109 | Dalteparin sodium |
| Fragmin 10,000units/4ml solution for injection ampoules (Pfizer Ltd) | 611141000033113 | 3969411000001100 | Dalteparin sodium |
| Dalteparin sodium 10,000units/4ml solution for injection ampoules | 404241000033119 | 36093111000001105 | Dalteparin sodium |
| Fragmin 100,000units/4ml solution for injection vials (Pfizer Ltd) | 1822041000033115 | 3970611000001100 | Dalteparin sodium |
| Dalteparin sodium 100,000units/4ml solution for injection vials | 1821941000033114 | 36093311000001107 | Dalteparin sodium |
| Fragmin 12,500units/0.5ml solution for injection pre-filled syringes (Pfizer Ltd) | 1731141000033112 | 3838711000001100 | Dalteparin sodium |
| Dalteparin sodium 12,500units/0.5ml solution for injection pre-filled syringes | 1726841000033111 | 36563811000001103 | Dalteparin sodium |
| Fragmin 15,000units/0.6ml solution for injection pre-filled syringes (Pfizer Ltd) | 1731241000033117 | 3968311000001101 | Dalteparin sodium |
| Dalteparin sodium 15,000units/0.6ml solution for injection pre-filled syringes | 1726941000033115 | 36564211000001101 | Dalteparin sodium |
| Fragmin 5,000units/0.2ml solution for injection pre-filled syringes (Pfizer Ltd) | 611241000033118 | 3723611000001103 | Dalteparin sodium |
| Dalteparin sodium 5,000units/0.2ml solution for injection pre-filled syringes | 404341000033112 | 36566011000001101 | Dalteparin sodium |
| Fragmin 10,000units/0.4ml solution for injection pre-filled syringes (Pfizer Ltd) | 1731041000033113 | 3967611000001107 | Dalteparin sodium |
| Dalteparin sodium 10,000units/0.4ml solution for injection pre-filled syringes | 1726741000033118 | 37084311000001106 | Dalteparin sodium |
| Fragmin 18,000units/0.72ml solution for injection pre-filled syringes (Pfizer Ltd) | 1731341000033110 | 3968611000001106 | Dalteparin sodium |
| Dalteparin sodium 18,000units/0.72ml solution for injection pre-filled syringes | 1727041000033119 | 37084411000001104 | Dalteparin sodium |
| Fragmin 2,500units/0.2ml solution for injection pre-filled syringes (Pfizer Ltd) | 611041000033114 | 3840411000001109 | Dalteparin sodium |
| Dalteparin sodium 2,500units/0.2ml solution for injection pre-filled syringes | 404141000033114 | 37084511000001100 | Dalteparin sodium |

#### Table S4. Stroke Aurum codes

| Term | MedCode ID | SNOMED CT Concept ID | SNOMED CT Description ID |
|---|---|---|---|
| Cerebral haemorrhage | 884421000006119 | 274100004 | 884421000006119 |
| External capsule haemorrhage | 300276019 | 195167002 | 300276019 |
| Basilar artery occlusion | 300303013 | 195180004 | 300303013 |
| Cerebral infarction due to thrombosis of cerebral | 300321011 | 195189003 | 300321011 |
| Anterior cerebral artery syndrome | 300363010 | 195210002 | 300363010 |
| Occlusion and stenosis of anterior cerebral artery | 300396012 | 195233001 | 300396012 |
| Occlusion and stenosis of posterior cerebral arter | 300398013 | 195234007 | 300398013 |
| Occlusion and stenosis of cerebellar arteries | 300399017 | 195235008 | 300399017 |
| Right sided cerebral infarction | 451134017 | 307767006 | 451134017 |
| H/O: Stroke in last year | 451371010 | 308067002 | 451371010 |
| Basal ganglion stroke | 6837051000006119 | 413102000 | 2966612019 |
| Subarachnoid haemorrhage from middle cerebral arte | 123511000006114 | 21454007 | 36011016 |
| Subarachnoid haemorrhage from posterior communicat | 123521000006118 | 21454007 | 36011016 |
| Left sided intracerebral haemorrhage, unspecified | 748941000006115 | 274100004 | 409859018 |
| Cerebrl infarctn due/unspcf occlusn or sten/cerebr | 543141000006110 | 20059004 | 33759015 |
| Internal capsule haemorrhage | 496232015 | 52201006 | 496232015 |
| Stroke due to cerebral arterial occlusion | 122361000006113 | 230691006 | 345638019 |
| Subarachnoid haemorrhage from basilar artery | 123491000006115 | 276284000 | 412361011 |
| [V]Personal history of transient ischaemic attack | 1667741000000110 | 751371000000107 | 1667741000000110 |
| Cerebral haemorrhage | 989201000006117 | 274100004 | 989201000006117 |
| Lobar cerebral haemorrhage | 345675012 | 230710000 | 345675012 |
| Cerebral A. occlusion NOS | 884501000006113 | 682621000000105 | 884501000006113 |
| Subdural haemorrhage | 884441000006114 | 195176009 | 884441000006114 |
| Lateral medullary syndrome | 130375018 | 78569004 | 130375018 |
| [RFC] Stroke | 907591000006116 | 907591000006100 | 907591000006116 |
| [RFC] Stroke | 908801000006114 | 908801000006105 | 908801000006114 |
| Right sided intracerebral haemorrhage, unspecified | 163261000006119 | 195168007 | 300277011 |
| Subarachnoid haemorrh from intracranial artery, un | 123441000006112 | 21454007 | 481028017 |
| [X]Other intracerebral haemorrhage | 11919571000006110 | 274100004 | 11919571000006110 |
| Infarction of basal ganglia | 2474651019 | 413102000 | 2474651019 |
| [V]Personal history of stroke | 1227591017 | 275526006 | 2476091017 |
| Basilar artery occluded | 884461000006113 | 195180004 | 884461000006113 |
| H/O: CVA/stroke | 809421000006116 | 275526006 | 2986886017 |
| Intracerebral haemorrhage NOS | 300287010 | 274100004 | 2819959010 |
| Cerebral thrombosis | 118689010 | 71444005 | 118689010 |
| H/O: CVA | 411518010 | 275526006 | 411518010 |
| Left sided cerebral infarction | 451133011 | 307766002 | 451133011 |
| Brain stem stroke | 4057041000006110 | 95457000 | 2966556014 |
| Lacunar stroke | 5011161000006115 | 230698000 | 345652017 |

| | | | |
|---|---|---|---|
| Cereb infarct due unsp occlus/stenos precerebr art | 542261000006114 | 125081000119106 | 3042974014 |
| [X]Other transnt cerebral ischaemic attacks+relate | 416991000006112 | 266257000 | 395788015 |
| Stroke due to intracerebral haemorrhage | 11903571000006110 | 274100004 | 11903571000006110 |
| Subdural haematoma - nontraumatic | 2475119012 | 195176009 | 2469010019 |
| Stroke due to intracerebral haemorrhage | 122371000006118 | 274100004 | 409860011 |
| [X]Other subarachnoid haemorrhage | 300936018 | 21454007 | 481028017 |
| Carotid territory transient ischaemic attack | 345684012 | 230716006 | 345684012 |
| Cerebral amyloid angiopathy | 345701012 | 230724001 | 345701012 |
| Subarachnoid haemorrhage from basilar artery aneur | 5583101000006116 | 276284000 | 412361011 |
| Posterior cerebral artery syndrome | 300364016 | 195211003 | 300364016 |
| Sequelae of intracerebral haemorrhage | 300407010 | 195241001 | 300407010 |
| Brainstem infarction | 524541000006117 | 95457000 | 158113017 |
| Cereb infarct due cerebral venous thrombosis, nonp | 542251000006112 | 195230003 | 300393016 |
| Cerebral haemorrhage NOS | 989211000006119 | 274100004 | 989211000006119 |
| Stroke and cerebrovascular accident unspecified | 12727691000006117 | 685631000000102 | 1501361000000113 |
| Intracerebral haemorrhage | 744901000006114 | 274100004 | 2819959010 |
| Cortical haemorrhage | 495394013 | 49422009 | 495394013 |
| Cerebellar haemorrhage | 502878012 | 75038005 | 502878012 |
| Bulbar haemorrhage | 483988011 | 732923001 | 3467313018 |
| CVA - cerebral artery occlusion | 605461000006117 | 230691006 | 345638019 |
| CVA - Cerebrovascular accident unspecified | 605491000006113 | 230690007 | 345635016 |
| Extradural haemorrhage - nontraumatic | 660111000006117 | 397809001 | 1773153015 |
| Subarachnoid haemorrhage from anterior communicati | 123481000006118 | 21454007 | 36011016 |
| Stroke | 884531000006117 | 685631000000102 | 884531000006117 |
| Ruptured berry aneurysm | 300242011 | 195154000 | 300242011 |
| Subarachnoid haemorrhage from carotid siphon and b | 300244012 | 195155004 | 300244012 |
| Subarachnoid haemorrhage from vertebral artery | 300253017 | 195160000 | 300253017 |
| Subdural haemorrhage - nontraumatic | 300294013 | 195176009 | 300294013 |
| Cerebral infarction due to embolism of precerebral | 300313017 | 195186005 | 300313017 |
| Sequelae of subarachnoid haemorrhage | 300406018 | 195240000 | 300406018 |
| Sequelae of cerebral infarction | 300411016 | 195243003 | 300411016 |
| [X]Cereb infarct due unsp occlus/stenos precerebr | 370661000006114 | 125081000119106 | 3042974014 |
| Cerebral infarction due to embolism of cerebral ar | 300322016 | 195190007 | 300322016 |
| Cerebral embolus | 542831000006116 | 75543006 | 125470015 |
| [X]Other cerebral infarction | 300941014 | 432504007 | 2770034014 |
| Intracerebral haemorrhage in hemisphere, unspecifi | 744921000006116 | 274100004 | 409860011 |
| Intracerebral haemorrhage, multiple localized | 746571000006116 | 195169004 | 300280012 |
| [X]Occlusion and stenosis of other cerebral arteri | 300943012 | 20059004 | 33759015 |
| [X]Other intracerebral haemorrhage | 300939013 | 274100004 | 409860011 |
| Cerebral haemorrhage NOS | 884451000006111 | 700251000000105 | 884451000006111 |
| CVA unspecified | 605501000006117 | 230690007 | 345637012 |

| | | | |
|---|---|---|---|
| [X]Subarachnoid haemorrh from intracranial artery, | 428181000006115 | 21454007 | 481028017 |
| Sequelae of other nontraumatic intracranial haemor | 149551000006111 | 363302008 | 482447013 |
| Sequelae of stroke,not specfd as h'morrhage or inf | 149571000006118 | 195239002 | 300403014 |
| Infarction - cerebral | 218511000000117 | 432504007 | 2770034014 |
| Cerebral infarction NOS | 12727431000006119 | 682621000000105 | 1495311000000117 |
| Basal nucleus haemorrhage | 503791000006114 | 195165005 | 300272017 |
| Stroke/CVA - undefined | 884521000006115 | 685631000000102 | 884521000006115 |
| [X]Cerebrl infarctn due/unspcf occlusn or sten/cer | 370701000006118 | 20059004 | 33759015 |
| History of stroke in last year | 5974521000006116 | 308067002 | 2986393012 |
| [RFC] Stroke/CVA | 907581000006119 | 907581000006103 | 907581000006119 |
| Brainstem infarction NOS | 345650013 | 95457000 | 158113017 |
| Intracerebral haemorrhage, intraventricular | 300277011 | 195168007 | 300277011 |
| Cerebral infarct due to thrombosis of precerebral | 300312010 | 195185009 | 300312010 |
| Middle cerebral artery syndrome | 300362017 | 195209007 | 300362017 |
| Right sided CVA | 300371014 | 195217004 | 300371014 |
| Intracranial haemorrhage NOS | 300298011 | 1386000 | 475553012 |
| Subarachnoid haemorrhage NOS | 300257016 | 21454007 | 481028017 |
| [V]Personal history of cerebrovascular accident (C | 1227592012 | 266995000 | 397829016 |
| Right sided intracerebral haemorrhage, unspecified | 12722481000006116 | 308128006 | 451441015 |
| Subarachnoid haemorrhage | 481028017 | 21454007 | 481028017 |
| [X]Intracerebral haemorrhage in hemisphere, unspec | 300956017 | 274100004 | 409860011 |
| Pontine haemorrhage | 503469016 | 7713009 | 503469016 |
| H/O: stroke | 2476091017 | 275526006 | 2476091017 |
| Cerebellar infarction | 158118014 | 95460007 | 158118014 |
| Intracranial subarachnoid haemorrhage from vertebr | 4777761000006112 | 195160000 | 2916525016 |
| Stroke and cerebrovascular accident unspecified | 405339016 | 230690007 | 345637012 |
| Wallenberg syndrome | 57341000006119 | 78569004 | 130374019 |
| Other and unspecified intracranial haemorrhage | 300290016 | 62914000 | 104563015 |
| CVA - cerebrovascular accid due to intracerebral h | 605471000006112 | 274100004 | 409859018 |
| Stroke | 5010981000006119 | 230690007 | 345636015 |
| CT - Cerebral thrombosis | 3662331000006119 | 71444005 | 1233388010 |
| Occlusive stroke | 1212072018 | 373606000 | 1212072018 |
| Cerebral arterial occlusion | 1222398015 | 20059004 | 1222398015 |
| H/O: TIA | 251692018 | 161511000 | 251692018 |
| Cerebral infarction NOS | 395780010 | 432504007 | 2770034014 |
| Stroke unspecified | 122401000006115 | 230690007 | 345637012 |
| Cerebral embolism | 125470015 | 75543006 | 125470015 |
| Left sided CVA | 300370010 | 195216008 | 300370010 |
| Occlusion and stenosis of middle cerebral artery | 300395011 | 195232006 | 300395011 |
| [X]Subarachnoid haemorrhage from other intracrania | 300935019 | 21454007 | 36011016 |

#### Table S5. Transient ischaemic attack selection Aurum codes

| Term | Medcode ID | SNOMED CT Concept ID | SNOMED CT Description ID |
|---|---|---|---|
| Anterior circulation transient ischaemic attack | 5011491000006115 | 230716006 | 345683018 |
| Transient cerebral ischemia | 11920121000006117 | 266257000 | 395785017 |
| Personal history of transient ischaemic attack | 8231151000006110 | 751371000000107 | 1653121000000119 |
| [V]Personal history of transient ischaemic attack | 1667741000000110 | 751371000000107 | 1667741000000110 |
| Transient ischaemic attack | 395783012 | 266257000 | 395783012 |
| Transient cerebral ischaemia | 395788015 | 266257000 | 395788015 |
| Carotid territory transient ischaemic attack | 345684012 | 230716006 | 345684012 |
| Transient Ischaemic Attacks | 884511000006111 | 584181000000100 | 884511000006111 |
| H/O: TIA | 251692018 | 161511000 | 251692018 |
| Transient Ischaemic Attacks | 988951000006117 | 266257000 | 988951000006117 |
| TIA - Transient ischaemic attack | 5492201000006111 | 266257000 | 395784018 |

#### Table S6. Arterial thromboembolism Aurum codes

| Term | MedCode ID | SNOMED CT Concept ID | SNOMED CT Description ID |
|---|---|---|---|
| Embolism and/or thrombosis of the internal iliac artery | 300554012 | 734298005 | 3503835018 |
| Cerebral infarction due to thrombosis of cerebral arteries | 300321011 | 195189003 | 300321011 |
| Embolism and thrombosis of the brachial artery | 300535012 | 195319003 | 300535012 |
| Embolism and thrombosis of the posterior tibial artery | 300543019 | 195327007 | 300543019 |
| Recurrent pulmonary embolism | 632111000000118 | 438773007 | 2793990014 |
| Embolism and thrombosis of the celiac artery | 4778981000006116 | 195342005 | 300560012 |
| Superior mesenteric artery embolus | 4785631000006110 | 196999001 | 302978019 |
| Cerebral arterial thrombosis | 3662321000006117 | 71444005 | 118691019 |
| Saddle embolus of abdominal aorta | 5058281000006118 | 233972005 | 350565011 |
| Peripheral arterial embolism | 884661000006110 | 583731000000103 | 884661000006110 |
| Embolus/thrombus artery NOS | 884671000006115 | 583761000000108 | 884671000006115 |
| Arterial embolism and thrombosis NOS | 12733161000006111 | 583761000000108 | 1295451000000114 |
| Thrombosis - coronary | 1786198013 | 398274000 | 1786198013 |
| Thrombosis of cerebral arteries | 3662311000006113 | 71444005 | 118690018 |
| Cerebral arterial embolism | 3728451000006116 | 75543006 | 125471016 |
| Renal artery thrombosis | 158317012 | 95579008 | 158317012 |
| Embolism and thrombosis of other specified artery | 300552011 | 266262004 | 395794011 |
| Arterial embolism and thrombosis NOS | 300564015 | 266262004 | 395794011 |
| Aortic bifurcation embolus | 5058291000006115 | 233972005 | 350566012 |
| Embolism and thrombosis of a leg artery NOS | 300547018 | 195318006 | 300534011 |
| Peripheral arterial embolism and thrombosis NOS | 300550015 | 195318006 | 300534011 |
| Renal artery embolism | 158318019 | 95580006 | 158318019 |
| Thromboembolus of internal iliac artery | 3503835018 | 734298005 | 3503835018 |
| SMAE - Superior mesenteric artery embolus | 4785641000006117 | 196999001 | 302979010 |
| Renal artery stent thrombosis | 1848121000006110 | 840961000000108 | 2182101000000113 |
| Thrombosis, carotid artery | 100771000006112 | 86003009 | 142588012 |
| Embolism and thrombosis of an arm artery NOS | 300538014 | 195318006 | 300534011 |
| Saddle embolus | 350563016 | 233972005 | 350563016 |
| Embolism and/or thrombosis of the external iliac artery | 300555013 | 734299002 | 3503839012 |
| Embolism and thrombosis of an arm or leg artery | 300534011 | 195318006 | 300534011 |
| Embolism and/or thrombosis of the common iliac artery | 300553018 | 195335005 | 300553018 |
| Embolism and thrombosis of the splenic artery | 300558010 | 195340002 | 300558010 |
| Embolus/thrombosis abd. aorta | 884641000006111 | 266263009 | 884641000006111 |
| Embolus/thrombosis aorta NOS | 884651000006113 | 195317001 | 884651000006113 |
| Embolism and thrombosis of other and unspec parts aorta | 638871000006114 | 274101000 | 409861010 |
| Embolus of the superior mesenteric artery | 639241000006110 | 196999001 | 302978019 |
| Arterial embolus and thrombosis | 395795012 | 266262004 | 395795012 |

| | | | |
|---|---|---|---|
| Embolism and thrombosis of the iliac artery unspecified | 300556014 | 266262004 | 395794011 |
| [X]Embolism and thrombosis of other arteries | 300964011 | 266262004 | 395794011 |
| Cerebral infarction due to embolism of precerebral arteries | 300313017 | 195186005 | 300313017 |
| Embolism and thrombosis of the anterior tibial artery | 300541017 | 195325004 | 300541017 |
| Embolism and thrombosis of the axillary artery | 300559019 | 195341003 | 300559019 |
| Incomplete spontaneous abortion with embolism | 305331019 | 198650006 | 305331019 |
| Embolism and thrombosis of other arteries NOS | 300563014 | 266262004 | 395794011 |
| Cerebral infarction due to embolism of cerebral arteries | 300322016 | 195190007 | 300322016 |
| Embolism and thrombosis of the radial artery | 300536013 | 195320009 | 300536013 |
| Embolism and thrombosis of the coeliac artery | 300561011 | 195342005 | 300561011 |
| Arterial embolism/thrombosis | 884631000006118 | 266262004 | 884631000006118 |
| Peripheral arterial embolism and thrombosis NOS | 12733151000006114 | 583731000000103 | 1295391000000110 |
| Thrombosis of renal artery | 4059051000006119 | 95579008 | 158316015 |
| Aortic thromboembolism | 5560171000006116 | 274101000 | 409861010 |
| Embolism and thrombosis of the abdominal aorta | 395796013 | 266263009 | 395796013 |
| Other embolism and thrombosis | 300712012 | 429098002 | 2692478010 |
| Thrombosis of artery of transplanted kidney | 2234541000000115 | 864271000000105 | 2234541000000115 |
| Chronic thromboembolic pulmonary hypertension | 5057801000006113 | 233947005 | 2968345015 |
| Thromboembolic pulmonary hypertension | 350517010 | 233947005 | 350517010 |
| [V] Personal history of pulmonary embolism | 451479013 | 161512007 | 251693011 |
| Cerebral infarct due to thrombosis of precerebral arteries | 300312010 | 195185009 | 300312010 |
| Embolism and thrombosis of the thoracic aorta | 300533017 | 195317001 | 300533017 |
| Embolism and thrombosis of the ulnar artery | 300537016 | 195321008 | 300537016 |
| Embolism and thrombosis of the popliteal artery | 300540016 | 195324000 | 300540016 |
| Pulmonary embolism | 98484016 | 59282003 | 98484016 |
| Embolism and thrombosis of the hepatic artery | 300562016 | 195343000 | 3491726018 |
| Embolism and thrombosis NOS | 300713019 | 429098002 | 2692478010 |
| Anterior spinal artery thrombosis | 345723011 | 230740003 | 345723011 |
| Embolism and thrombosis of hepatic artery | 4778991000006118 | 195343000 | 3491726018 |
| Arteriovenous fistula thrombosis | 350892013 | 234205007 | 350892013 |
| Renal artery stent thrombosis | 1809151000006116 | 1809151000006100 | 1809151000006116 |
| Thrombus of the superior mesenteric artery | 100811000006112 | 197000003 | 302980013 |
| Thromboembolus of external iliac artery | 3503839012 | 734299002 | 3503839012 |
| Embolism and thrombosis NOS | 638821000006113 | 429098002 | 2692478010 |
| Thrombosis - arterial | 218541000000116 | 65198009 | 108347015 |
| Arterial embolism and thrombosis | 491241000006118 | 266262004 | 395795012 |
| PE - Pulmonary embolism | 3462921000006119 | 59282003 | 1231937012 |
| Renal artery embolus | 1235925019 | 95580006 | 1235925019 |
| [RFC] Arterial embolism of limbs | 905541000006119 | 905541000006103 | 905541000006119 |
| Arterial embolic and thrombotic occlusion | 395794011 | 266262004 | 395794011 |

| | | | |
|---|---|---|---|
| Embolism and thrombosis of the femoral artery | 300539018 | 195323006 | 300539018 |
| Embolism and thrombosis of the dorsalis pedis artery | 300542012 | 195326003 | 300542012 |
| Embolism and thrombosis of the subclavian artery | 300557017 | 195339004 | 300557017 |
| Thromboembolic pulmonary hypertension | 1749291000006113 | 1749291000006109 | 1749291000006113 |

#### Table S7. Myocadial infarction Aurum codes

| Term | Medcode ID | SNOMED CT Concept ID | SNOMED CT Description ID |
|---|---|---|---|
| Acute anteroapical infarction | 299708014 | 52035003 | 3038718019 |
| Subsequent myocardial infarction of inferior wall | 299812011 | 194858006 | 299812011 |
| Acute Q-wave infarct | 447324018 | 304914007 | 447324018 |
| Acute transmural myocardial infarction of unspecif site | 460681000006116 | 57054005 | 94884017 |
| Postmyocardial infarction syndrome | 3576371000006117 | 66189004 | 109915012 |
| Acute non-Q wave infarction | 450322013 | 307140009 | 450322013 |
| Myocardial infarct | 2855351000006111 | 22298006 | 1784873012 |
| Acute myocardial infarction of inferolateral wall | 3565871000006113 | 65547006 | 108912018 |
| Acute inferior myocardial infarction | 3699921000006110 | 73795002 | 1233665015 |
| Subsequent myocardial infarction of unspecified site | 118831000006118 | 194856005 | 299808017 |
| Postoperative transmural myocardial infarction other sites | 212081000006112 | 129574000 | 208365015 |
| Postoperative transmural myocardial infarction unspec site | 212091000006110 | 129574000 | 208365015 |
| Acute myocardial infarction of atrium | 4775891000006119 | 194809007 | 2470032018 |
| Acute myocardial infarction of septum | 3784911000006111 | 79009004 | 131106018 |
| Other acute myocardial infarction NOS | 299720018 | 57054005 | 94884017 |
| History of myocardial infarction in last year | 5974481000006116 | 308065005 | 2986726013 |
| Acute papillary muscle infarction | 1218860015 | 10273003 | 1218860015 |
| Acute myocardial infarction of anterolateral wall | 3641641000006116 | 70211005 | 116613011 |
| Personal history of myocardial infarction | 230021000006115 | 399211009 | 2986492018 |
| Acute Q wave myocardial infarction | 5935321000006111 | 304914007 | 447323012 |
| Silent myocardial infarction | 350376014 | 233843008 | 350376014 |
| Acute ST segment elevation myocardial infarction | 1780491019 | 401303003 | 1780491019 |
| MI - Myocardial infarction | 2855341000006114 | 22298006 | 1784872019 |
| History of myocardial infarction | 1738171000006114 | 399211009 | 2986492018 |
| Pericarditis following myocardial infarction | 5056951000006117 | 233885007 | 350432018 |
| Postoperative myocardial infarction | 208365015 | 129574000 | 208365015 |
| MI - Silent myocardial infarction | 5056461000006113 | 233843008 | 350377017 |
| Acute myocardial infarction of septum alone | 3784921000006115 | 79009004 | 131107010 |
| Acute posterolateral myocardial infarction | 967931000006114 | 15990001 | 27071012 |
| Previous myocardial infarction | 6619201000006117 | 399211009 | 545891000000115 |
| Acute myocardial infarction of lateral wall | 3452181000006112 | 58612006 | 97399018 |
| Myocardial infarction with complication | 6348651000006112 | 371068009 | 6348651000006112 |
| Lateral myocardial infarction NOS | 299711010 | 58612006 | 1231860015 |
| Hemopericardium due to and following acute myocardial infarction | 12220751000006113 | 194862000 | 3673281015 |
| Ventric septal defect/curr comp fol acut myocardal infarctn | 67081000006119 | 233846000 | 350381017 |
| Postmyocardial infarction syndrome | 109915012 | 66189004 | 500341014 |
| AMI - Acute myocardial infarction | 3427201000006111 | 57054005 | 1231678011 |

| | | | |
|---|---|---|---|
| Postoperative transmural myocardial infarction inferior wall | 212071000006114 | 311793000 | 455419012 |
| Other specified anterior myocardial infarction | 299707016 | 54329005 | 1231324017 |
| Posterior myocardial infarction NOS | 299710011 | 233838001 | 350371016 |
| Other acute myocardial infarction | 299718016 | 57054005 | 94884017 |
| Acute myocardial infarction of anterior wall | 3381601000006117 | 54329005 | 90302019 |
| Anterior myocardial infarction NOS | 299709018 | 54329005 | 1231324017 |
| Past history of myocardial infarction | 6619191000006115 | 399211009 | 1786753012 |
| True posterior myocardial infarction | 299712015 | 194802003 | 299712015 |
| Subsequent myocardial infarction of anterior wall | 299811016 | 194857001 | 299811016 |
| Acute inferolateral infarction | 457531000006110 | 65547006 | 1232697013 |
| Acute myocardial infarction of inferoposterior wall | 3745741000006117 | 76593002 | 127193015 |
| [RFC] Myocardial infarction (MI) | 905351000006113 | 905351000006109 | 905351000006113 |
| Cardiac rupture following myocardial infarction (MI) | 537751000006115 | 233847009 | 350383019 |
| Old myocardial infarction | 4031011 | 1755008 | 4031011 |
| Subsequent myocardial infarction | 299808017 | 194856005 | 299808017 |
| Acute anteroseptal infarction | 455651000006114 | 62695002 | 104192010 |
| [X]Acute transmural myocardial infarction of unspecif site | 362461000006119 | 57054005 | 94884017 |
| First myocardial infarction | 932081000006118 | 932081000006102 | 932081000006118 |
| STEMI - ST elevation myocardial infarction | 6651221000006117 | 401303003 | 2840840013 |
| Postoperative subendocardial myocardial infarction | 455422014 | 311796008 | 455422014 |
| Acute anterolateral infarction | 455641000006112 | 70211005 | 1233238016 |
| [X]Subsequent myocardial infarction of unspecified site | 300882013 | 194856005 | 299808017 |
| Postoperative myocardial infarction, unspecified | 455423016 | 129574000 | 208365015 |
| Acute myocardial infarction of diaphragmatic wall | 3699911000006119 | 73795002 | 122559017 |
| Acute subendocardial infarction | 116992017 | 70422006 | 116992017 |
| Inferior myocardial infarction NOS | 299714019 | 73795002 | 122557015 |
| Acute myocardial infarction NOS | 299721019 | 57054005 | 94884017 |
| Subsequent myocardial infarction of other sites | 299813018 | 194856005 | 299808017 |
| Cardiac rupture due to and following acute myocardial infarction | 12221301000006119 | 233847009 | 3673284011 |
| Hemopericardium as current complication following acute myocardial infarction | 4776221000006110 | 194862000 | 299817017 |
| Acute septal infarction | 1234306015 | 79009004 | 1234306015 |
| Acute non-ST segment elevation myocardial infarction | 1780501013 | 401314000 | 1780501013 |
| History of myocardial infarct at age greater than sixty | 4540481000006111 | 161503005 | 2986493011 |
| Postoperative transmural myocardial infarction anterior wall | 212061000006119 | 311792005 | 455418016 |
| Acute anteroapical myocardial infarction | 3343471000006116 | 52035003 | 86618010 |
| [X]Subsequent myocardial infarction of other sites | 300881018 | 194856005 | 299808017 |
| Postmyocardial infarction pericarditis | 3576391000006116 | 66189004 | 109917016 |
| History of myocardial infarct at age less than sixty | 4540461000006118 | 161502000 | 2986996013 |
| Acute inferoposterior infarction | 1234005010 | 76593002 | 1234005010 |

| | | | |
|---|---|---|---|
| Myocardial Infarction | 884151000006119 | 57054005 | 884151000006119 |
| MI - acute myocardial infarction | 219531000000117 | 57054005 | 94884017 |
| Acute myocardial infarction | 94884017 | 57054005 | 94884017 |
| Myocardial infarction | 2855301000006112 | 22298006 | 37436014 |

#### Table S8. Periperal arterial disease Aurum codes

| Term | Medcode ID | SNOMED CT Concept ID | SNOMED CT Description ID |
|---|---|---|---|
| Ischaemic foot | 443199013 | 301755001 | 443199013 |
| Peripheral angiopathic disease EC NOS | 300511019 | 400047006 | 1779317016 |
| Other specified peripheral vascular disease NOS | 300514010 | 400047006 | 1779317016 |
| Peripheral vascular disease NOS | 300515011 | 400047006 | 1779317016 |
| Peripheral arterial disease confirmed | 1823971000006118 | 1823971000006102 | 1823971000006118 |
| Peripheral ischaemic vascular disease | 350535018 | 233958001 | 350535018 |
| Ischaemic foot | 1699111000006110 | 1699111000006106 | 1699111000006110 |
| Other specified peripheral vascular disease | 300510018 | 400047006 | 1779317016 |
| Peripheral vascular disease NOS | 235911000006116 | 400047006 | 1779317016 |
| Ischaemia of legs | 742481000006118 | 233961000 | 350546015 |
| Other peripheral vascular disease | 395791015 | 400047006 | 1779317016 |
| [X]Other specified peripheral vascular diseases | 300963017 | 400047006 | 1779317016 |
| Peripheral ischaemia | 350533013 | 233958001 | 350533013 |
| Peripheral vascular disease NOS | 12729241000006119 | 646351000000103 | 1422221000000115 |
| Peripheral arterial disease | 1847121000006114 | 399957001 | 1787050010 |
| Aortic atherosclerosis | 218531000000113 | 81817003 | 135728018 |

#### Table S9. Aortic plaque Aurum codes

| Term | MedCode ID | SNOMED CT Concept ID | SNOMED CT Description ID |
|---|---|---|---|
| Atherosclerosis aorta | 3830431000006112 | 81817003 | 2923328010 |
| Aortic thromboembolism | 5560171000006116 | 274101000 | 409861010 |
| Aortoiliac atherosclerosis | 5057931000006110 | 233956002 | 350530011 |
| Aortic atherosclerosis | 218531000000113 | 81817003 | 135728018 |

#### Table S10. Diabetes Aurum codes

| Term | Medcode ID | SNOMED CT Concept ID | SNOMED CT Description ID |
|---|---|---|---|
| Pre-existing type 2 diabetes mellitus in pregnancy | 11931861000006112 | 237627000 | 1988741000006117 |
| Unstable type I diabetes mellitus | 429970018 | 290002008 | 429970018 |
| Diabetes mellitus with ketoacidosis | 2622193012 | 420422005 | 2622193012 |
| Lipoatrophic diabetes mellitus | 5687751000006119 | 284449005 | 1495355018 |
| Other specified diabetes mellitus with multiple comps | 13751000006117 | 385041000000108 | 760111000000115 |
| Other specified diabetes mellitus with periph circ comps | 13791000006111 | 421895002 | 2618203015 |
| Unstable type 1 diabetes mellitus | 72711000006117 | 290002008 | 429971019 |
| Type 1 diabetes mellitus - poor control | 84291000006117 | 444073006 | 2872487013 |
| Type 1 diabetes mellitus with nephropathy | 84371000006110 | 421893009 | 2623054013 |
| Type 1 diabetes mellitus with polyneuropathy | 84421000006115 | 713705003 | 3297342019 |
| Type 1 diabetes mellitus with ulcer | 84451000006112 | 190368000 | 292540012 |
| Type 2 diabetes mellitus - poor control | 84481000006116 | 443694000 | 2921019012 |
| Type 2 diabetes mellitus with arthropathy | 84491000006118 | 314903002 | 459310018 |
| Type 2 diabetes mellitus with neuropathic arthropathy | 84571000006115 | 314904008 | 459313016 |
| Type 2 diabetes mellitus with peripheral angiopathy | 84591000006119 | 314902007 | 459308015 |
| Type 2 diabetes mellitus with polyneuropathy | 84601000006110 | 713706002 | 3297353013 |
| Type I diabetes mellitus with arthropathy | 84681000006118 | 314893005 | 459294012 |
| Type I diabetes mellitus with hypoglycaemic coma | 84711000006117 | 314771006 | 459163017 |
| Type I diabetes mellitus with ulcer | 84821000006118 | 190368000 | 292538019 |
| Type II diabetes mellitus with polyneuropathy | 84971000006111 | 713706002 | 3297353013 |
| Type II diabetes mellitus with renal complications | 84981000006114 | 420279001 | 3013392012 |
| Insulin dependent diabetes mellitus with arthropathy | 6051171000006117 | 314893005 | 459293018 |
| Insulin-dependent diabetes mellitus secretory diarrhea syndrome | 5109851000006119 | 237618001 | 356116016 |
| Type I diabetes mellitus without complication | 457325013 | 313435000 | 457325013 |
| Non-insulin-dependent diabetes mellitus without complication | 457328010 | 313436004 | 457328010 |
| MODY - Maturity onset diabetes glucokinase-related | 5109621000006112 | 237604008 | 356087019 |
| DKA - diabetic ketoacidosis | 6953901000006115 | 420422005 | 3303994013 |
| Maturity onset diabetes of the young type 5 | 1968641000006114 | 609572000 | 2967853015 |
| Diabetes mellitus, juvenile type, + unspecified complication | 616611000006111 | 420868002 | 3013528019 |
| Pre-existing diabetes mellitus in mother complicating childbirth | 8033291000006116 | 106281000119103 | 3005790010 |
| ADM - atypical diabetes mellitus | 11633221000006116 | 530558861000132104 | 3334355014 |
| Type I diabetes mellitus with hypoglycemic coma | 6050061000006116 | 314771006 | 459164011 |
| Type 1 diabetes mellitus with multiple complications | 84361000006115 | 422228004 | 2618232017 |
| Type I diabetes mellitus with neurological complications | 84751000006116 | 421468001 | 3695401013 |
| Non-insulin dependent diabetes mellitus with mononeuropathy | 280521000006117 | 420436000 | 3697667019 |

| | | | |
|---|---|---|---|
| Non-insulin-dependent diabetes mellitus with ophthalm comps | 587111000006111 | 422099009 | 3698440016 |
| Diabetes mellitus, adult onset, with renal manifestation | 616551000006111 | 420279001 | 2615535015 |
| Diabetes mellitus, adult with gangrene | 616561000006113 | 421631007 | 2618206011 |
| Diabetes mellitus, juvenile +peripheral circulatory disorder | 616591000006117 | 421365002 | 2618205010 |
| Type 1 diabetes mellitus with multiple complications | 913541000006118 | 422228004 | 2618232017 |
| Type I diabetes mellitus with retinopathy | 913671000006113 | 420789003 | 3699407019 |
| Type I diabetes mellitus with mononeuropathy | 913791000006119 | 420918009 | 3697664014 |
| Type II diabetes mellitus with neurological complications | 914081000006117 | 421326000 | 3695406015 |
| Type II diabetes mellitus with retinopathy | 914161000006114 | 422034002 | 3699410014 |
| Type II diabetes mellitus with mononeuropathy | 914231000006111 | 420436000 | 3697667019 |
| Type II diabetes mellitus with diabetic cataract | 914311000006114 | 420756003 | 3688522015 |
| Type II diabetes mellitus with gastroparesis | 1667921000000117 | 713703005 | 3698412014 |
| Type I diabetes mellitus with gastroparesis | 1667941000000112 | 713702000 | 3698406016 |
| Insulin-dependent diabetes mellitus with neurological comps | 771481000006112 | 422088007 | 3688477014 |
| Insulin-dependent diabetes mellitus with ophthalmic comps | 771491000006110 | 25093002 | 3688467012 |
| Other specified diabetes mellitus with ketoacidosis | 292478012 | 420422005 | 2616611010 |
| Diabetes mellitus NOS with ketoacidotic coma | 292489017 | 420422005 | 2616611010 |
| Other specified diabetes mellitus with renal complications | 292495016 | 127013003 | 301016 |
| Type I diabetes mellitus - poor control | 292550013 | 444073006 | 2872487013 |
| Type II diabetes mellitus - poor control | 292589019 | 443694000 | 2921019012 |
| [X]Other specified diabetes mellitus | 293756010 | 73211009 | 121589010 |
| Insulin-dependent diabetes mellitus with ulcer | 4758061000006110 | 190368000 | 292539010 |
| Diabetes type 2 with cataract | 6959811000006110 | 420756003 | 3035281015 |
| Diabetes type 2 with retinopathy | 6982521000006119 | 422034002 | 3035295017 |
| Diabetes mellitus, Addison's disease and myxoedema | 3862341000006115 | 83728000 | 505747013 |
| Type 1 diabetes mellitus with arthropathy | 459292011 | 314893005 | 459292011 |
| Type I diabetes mellitus with arthropathy | 459294012 | 314893005 | 459294012 |
| Type II diabetes mellitus with peripheral angiopathy | 459306016 | 314902007 | 459306016 |
| Type 2 diabetes mellitus with peripheral angiopathy | 459308015 | 314902007 | 459308015 |
| Non-insulin dependent diabetes mellitus with arthropathy | 459309011 | 314903002 | 459309011 |
| NIDDM - Non-insulin dependent diabetes mellitus | 493773010 | 44054006 | 493773010 |
| Secondary diabetes mellitus without complication | 189721000000113 | 8801005 | 15518018 |
| Pre-existing diabetes mellitus, unspecified | 213141000006111 | 385051000000106 | 760131000000111 |
| Type 1 diabetes mellitus with hyperosmolar coma | 4757991000006116 | 190330002 | 2967854014 |
| Type 1 diabetes mellitus with hypoglycemic coma | 6050081000006114 | 314771006 | 459166013 |
| Diabetes type 1 with ketoacidosis | 6951091000006110 | 420270002 | 3035606019 |
| NIDDM with peripheral circulatory disorder | 12485441000006119 | 422166005 | 674961000006118 |
| DM - Diabetes mellitus | 3690681000006114 | 73211009 | 502372015 |

| | | | |
|---|---|---|---|
| Diabetes mellitus type 2 with ketoacidotic coma | 6979251000006110 | 421847006 | 3035601012 |
| Diet controlled diabetes mellitus | 4636411000006110 | 170745003 | 264680017 |
| Type I diabetes mellitus with ophthalmic complications | 913491000006111 | 739681000 | 3698438014 |
| Diabetes mellitus NOS with neurological manifestation | 292512019 | 422088007 | 2618195014 |
| Other specified diabetes mellitus with neurological comps | 13761000006115 | 422088007 | 2618195014 |
| Type 1 diabetes mellitus with diabetic cataract | 84321000006114 | 421920002 | 2618234016 |
| Type 2 diabetes mellitus with gangrene | 84511000006112 | 421631007 | 2618206011 |
| Type 2 diabetes mellitus with mononeuropathy | 84531000006118 | 420436000 | 2618201018 |
| Diabetes mellitus, adult onset, + ophthalmic manifestation | 616491000006116 | 422099009 | 2618233010 |
| Type 1 diabetes mellitus with mononeuropathy | 913781000006117 | 420918009 | 2618199015 |
| Type 1 diabetes mellitus with ophthalmic complications | 84401000006113 | 739681000 | 3537386015 |
| Diabetes mellitus, juvenile type, + ophthalmic manifestation | 616601000006113 | 739681000 | 3537386015 |
| Diabetes mellitus with polyneuropathy | 616451000006110 | 49455004 | 82373015 |
| Diabetes mellitus, juvenile, + other specified manifestation | 616681000006116 | 73211009 | 121589010 |
| Diabetes mellitus confirmed | 1823921000006119 | 1823921000006103 | 1823921000006119 |
| Diabetic on insulin | 264682013 | 170747006 | 264682013 |
| Diabetic - cooperative patient | 264717015 | 170769004 | 264717015 |
| Insulin treated non-insulin dependent diabetes mellitus | 1223148019 | 237599002 | 1223148019 |
| Maturity onset diabetes | 223291000000111 | 44054006 | 493774016 |
| Newly diagnosed diabetes | 1694761000006113 | 405749004 | 2157525015 |
| Diabetic acidosis | 6953891000006119 | 420422005 | 2622194018 |
| MODY - Maturity onset diabetes in youth type 2 | 5109611000006116 | 237604008 | 356086011 |
| Diabetes mellitus with neuropathy | 345487013 | 230572002 | 345487013 |
| Diabetes type 2 with neurological disorder | 6969931000006116 | 421326000 | 3035455016 |
| Type 2 diabetes mellitus with hyperosmolar coma | 4758011000006112 | 190331003 | 2967818010 |
| Diabetes with coma | 932641000006116 | 630521000000101 | 932641000006116 |
| DM + persist microalbuminuria | 933211000006112 | 658061000000102 | 933211000006112 |
| Diabetes + nephropathy | 881481000006117 | 127013003 | 881481000006117 |
| Diabetes + eye manifestation | 881491000006119 | 25093002 | 881491000006119 |
| Diabetes + periph.circulat.dis | 881511000006113 | 422275004 | 881511000006113 |
| Lipodystrophic diabetes with partial lipoatrophy | 3730121000006115 | 75659004 | 125663012 |
| Secondary endocrine diabetes mellitus | 356078011 | 237601000 | 356078011 |
| Diabetes mellitus associated with genetic syndrome | 10928019 | 5969009 | 10928019 |
| Type 1 diabetes mellitus with persistent microalbuminuria | 1780311019 | 401110002 | 1780311019 |
| Diabetes mellitus | 121589010 | 73211009 | 121589010 |
| Maturity onset diabetes in youth type II | 5109581000006112 | 237604008 | 356083015 |
| Insulin treated Type II diabetes mellitus | 841351000006110 | 237599002 | 1223147012 |
| Type I diabetes mellitus with renal complications | 913461000006115 | 421893009 | 2623054013 |
| Type I diabetes mellitus with nephropathy | 913851000006119 | 421893009 | 2623054013 |

| | | | |
|---|---|---|---|
| Type II diabetes mellitus with polyneuropathy | 914251000006116 | 713706002 | 3297353013 |
| Insulin treated Type 2 diabetes mellitus | 914391000006116 | 237599002 | 2967820013 |
| Type I diabetes mellitus with persistent microalbuminuria | 928491000006110 | 401110002 | 1780311019 |
| Type I diabetes mellitus with ketoacidosis | 928511000006116 | 420270002 | 2967817017 |
| Type I diabetes mellitus with ketoacidotic coma | 928531000006110 | 421075007 | 2967758019 |
| Type II diabetes mellitus with ketoacidosis | 928591000006114 | 421750000 | 2967754017 |
| Type I diabetes mellitus with exudative maculopathy | 938311000006112 | 420486006 | 2618236019 |
| Insulin dependent diabetes mellitus with hypoglycaemic coma | 459162010 | 237632004 | 356141011 |
| Pre-existing type 1 diabetes mellitus in pregnancy | 2967831017 | 199223000 | 306106018 |
| Type 1 diabetes mellitus with peripheral angiopathy | 84411000006111 | 31211000119101 | 3315042015 |
| Insulin dependent diabetes mellitus with multiple complicatn | 771381000006115 | 385041000000108 | 760111000000115 |
| Insulin dependent diabetes mellitus with polyneuropathy | 771401000006115 | 49455004 | 82373015 |
| Insulin-dependent diabetes mellitus with renal complications | 771501000006119 | 127013003 | 301016 |
| Insulin dependent diab mell with peripheral angiopathy | 772141000006118 | 421895002 | 2618203015 |
| IDDM with peripheral circulatory disorder | 787101000006114 | 421895002 | 2618203015 |
| Insulin-dependent diabetes mellitus with renal complications | 913441000006119 | 127013003 | 301016 |
| Insulin dependent diabetes mellitus with retinopathy | 913651000006115 | 4855003 | 9093013 |
| Insulin dependent diabetes mellitus with nephropathy | 913831000006114 | 127013003 | 301016 |
| Insulin dependent diab mell with peripheral angiopathy | 913921000006119 | 421895002 | 2618203015 |
| Type 1 diabetes mellitus with peripheral angiopathy | 913931000006116 | 31211000119101 | 3315042015 |
| Insulin dependent diab mell with neuropathic arthropathy | 913981000006115 | 201724008 | 309740011 |
| [RFC] Diabetes | 905621000006113 | 905621000006109 | 905621000006113 |
| [RFC] Diabetes mellitus | 908831000006118 | 908831000006102 | 908831000006118 |
| Diabetes mellitus co-occurrent and due to cystic fibrosis | 7063141000006118 | 426705001 | 3440582013 |
| [X]Diabetes mellitus | 377001000006117 | 73211009 | 121589010 |
| Type I diabetes mellitus maturity onset | 292551012 | 190372001 | 292551012 |
| Type 1 diabetes mellitus maturity onset | 292553010 | 190372001 | 292553010 |
| Diabetes type 1 with gangrene | 6961101000006114 | 420825003 | 3035408014 |
| Maturity onset diabetes in youth | 483886014 | 609561005 | 2967884018 |
| Diabetes mellitus, Addison's disease and myxedema | 3862421000006116 | 83728000 | 505755018 |
| Diabetes mellitus type 1 | 3253241000006117 | 46635009 | 197985011 |
| Diabetes with coma | 881471000006115 | 26298008 | 881471000006115 |
| Diabetes mellitus autosomal dominant type 2 | 840971000006112 | 237604008 | 356082013 |
| Type 1 diabetes mellitus with nephropathy | 913841000006116 | 421893009 | 2623054013 |
| Type 2 diabetes mellitus with renal complications | 914031000006118 | 420279001 | 2618208012 |
| Type 1 diabetes mellitus with ketoacidosis | 928501000006119 | 420270002 | 2967817017 |

| | | | |
|---|---|---|---|
| Type 2 diabetes mellitus with persistent proteinuria | 928541000006117 | 421986006 | 2967751013 |
| Type 2 diabetes mellitus | 197761014 | 44054006 | 197761014 |
| Latent diabetes | 2649991000006110 | 9414007 | 16500010 |
| Type 1 diabetes mellitus without complication | 457326014 | 313435000 | 457326014 |
| Type 2 diabetes mellitus with arthropathy | 459310018 | 314903002 | 459310018 |
| Diabetic on insulin and glucagon-like peptide 1 | 1966321000006112 | 976361000000100 | 2483761000000117 |
| Diabetes mellitus NOS with hyperosmolar coma | 292484010 | 422126006 | 2618221017 |
| [X]Unspecified diabetes mellitus with renal complications | 293759015 | 127013003 | 301016 |
| DM caused by insulin receptor ab | 3296773014 | 75682002 | 3296773014 |
| DM due to insulin receptor ab | 2164134013 | 75682002 | 2164134013 |
| Non-insulin dependent diabetes mellitus | 73466011 | 44054006 | 493773010 |
| Diabetes with peripheral circulatory disorder | 4392211000006114 | 127014009 | 3313383012 |
| Ketoacidosis in type II diabetes mellitus | 6977421000006110 | 421750000 | 2618220016 |
| Diabetes mellitus, adult onset, + unspecified complication | 616501000006112 | 422014003 | 3013072014 |
| Diabetes mellitus, juvenile type, no mention of complication | 616621000006115 | 313435000 | 457326014 |
| Insulin dependent diabetes mellitus - poor control | 772181000006112 | 268519009 | 401531012 |
| Insulin dependent diabetes mellitus - poor control | 913681000006111 | 268519009 | 401531012 |
| Type I diabetes mellitus with peripheral angiopathy | 913941000006114 | 31211000119101 | 3315042015 |
| Type II diabetes mellitus with hypoglycaemic coma | 459169018 | 719216001 | 3316336015 |
| Brittle diabetes mellitus | 2683561000006119 | 11530004 | 2658852011 |
| Non-insulin dependent diabetes mellitus with gangrene | 281211000006117 | 421631007 | 2967779019 |
| Type II diabetes mellitus with exudative maculopathy | 12489851000006117 | 421779007 | 938331000006118 |
| Diabetes type 2 on insulin | 5109461000006115 | 237599002 | 3037103012 |
| Diabetes mellitus NOS with ketoacidosis | 292479016 | 420422005 | 2616611010 |
| Ketoacidosis in diabetes mellitus | 6953871000006115 | 420422005 | 2622192019 |
| Jamaica type diabetes | 1550571000000114 | 75524006 | 1550571000000114 |
| Non-insulin dependent d m with neuropathic arthropathy | 281181000006116 | 314904008 | 459314010 |
| Type II diabetes mellitus with renal complications | 914041000006111 | 420279001 | 3013392012 |
| Type II diabetes mellitus with persistent microalbuminuria | 928571000006113 | 420715001 | 2967769016 |
| Type I diabetes mellitus with mononeuropathy | 84721000006113 | 420918009 | 3697664014 |
| Type II diabetes mellitus with diabetic cataract | 84871000006117 | 420756003 | 3688522015 |
| Type II diabetes mellitus with mononeuropathy | 84901000006117 | 420436000 | 3697667019 |
| Type II diabetes mellitus with neurological complications | 84931000006113 | 421326000 | 3695406015 |
| Diabetes mellitis with nephropathy NOS | 292496015 | 127013003 | 301016 |
| Diabetes mellitus NOS with unspecified complication | 292626017 | 74627003 | 123939015 |
| Type 1 diabetes mellitus uncontrolled | 7287611000006113 | 444073006 | 2967763015 |
| Insulin-dependent diabetes without complication | 6030001000006116 | 313435000 | 457327017 |
| Type I diabetes mellitus with neurological complications | 913521000006113 | 421468001 | 3695401013 |

| | | | |
|---|---|---|---|
| Type 1 diabetes mellitus with retinopathy | 913661000006118 | 420789003 | 3699407019 |
| Type 2 diabetes mellitus with ophthalmic complications | 914051000006113 | 422099009 | 3698440016 |
| Type I diabetes mellitus with ophthalmic complications | 84771000006114 | 739681000 | 3698438014 |
| MODY - Maturity onset diabetes in youth type II | 5109591000006110 | 237604008 | 356084014 |
| Type 1 diabetes mellitus with polyneuropathy | 913811000006115 | 713705003 | 3297342019 |
| Type II diabetes mellitus with nephropathy | 914271000006114 | 420279001 | 3013392012 |
| Type I diabetes mellitus with persistent proteinuria | 928471000006114 | 420514000 | 2967914011 |
| Diabetes mellitus with hyperosmolar coma | 292480018 | 422126006 | 2618221017 |
| Diabetes mellitus with other specified manifestation | 292617015 | 74627003 | 123939015 |
| Type I diabetes mellitus with peripheral angiopathy | 84781000006112 | 31211000119101 | 3315042015 |
| Insulin-dependent diabetes without complication | 746791000006111 | 111552007 | 178796018 |
| Insulin dependent diabetes mellitus with arthropathy | 771331000006116 | 39710007 | 1229552016 |
| Insulin dependent diabetes mellitus with mononeuropathy | 771371000006118 | 230577008 | 345492010 |
| Insulin dependent diabetes mellitus with retinopathy | 771411000006117 | 4855003 | 9093013 |
| Insulin dependent diab mell with neuropathic arthropathy | 772131000006111 | 201724008 | 309740011 |
| Insulin dependent diabetes maturity onset | 772151000006116 | 73211009 | 121589010 |
| IDDM-Insulin dependent diabetes mellitus | 787111000006112 | 73211009 | 121589010 |
| Diabetes mellitus type II | 3209431000006116 | 44054006 | 73465010 |
| Unstable diabetes | 411891014 | 11530004 | 19931010 |
| Diabetes mellitus autosomal dominant | 483882011 | 609561005 | 2967884018 |
| Type I diabetes mellitus - poor control | 84661000006111 | 444073006 | 2872487013 |
| Type I diabetes mellitus with multiple complications | 84731000006111 | 422228004 | 2967771016 |
| Diabetes mellitus, adult onset, with no mention of complication | 12718861000006118 | 532411000000102 | 1188841000000119 |
| Type 1 diabetes mellitus with ulcer | 292540012 | 190368000 | 292540012 |
| Type II diabetes mellitus with multiple complications | 292576013 | 190388001 | 292576013 |
| Lipoatrophic diabetes | 4392141000006116 | 127012008 | 300015 |
| Diabetes mellitus induced by steroids | 616241000006112 | 190447002 | 292667010 |
| Insulin dependent diabetes mellitus with ulcer | 771421000006113 | 422183001 | 2623058011 |
| Pre-existing diabetes mellitus in childbirth | 8033301000006115 | 106281000119103 | 3005777015 |
| Unstable diabetes mellitus | 2683591000006110 | 11530004 | 2871747019 |
| [X]Unspecified diabetes mellitus with renal complications | 12703991000006112 | 127013003 | 430881000006111 |
| Type 2 diabetes mellitus with retinopathy | 84621000006117 | 422034002 | 3699410014 |
| Non-insulin dependent diabetes mellitus with ulcer | 280551000006114 | 190389009 | 292580015 |
| Non-insulin-dependent d m with peripheral angiopath | 280561000006111 | 314902007 | 459307013 |
| Non-insulin-dependent diabetes mellitus with multiple comps | 280581000006118 | 190388001 | 292578014 |
| Diabetes mellitus - juvenile | 881441000006111 | 73211009 | 881441000006111 |
| Diabetes+ketoacidosis -no coma | 881461000006110 | 420422005 | 881461000006110 |

| | | | |
|---|---|---|---|
| Diabetes + neuropathy | 881501000006110 | 230572002 | 881501000006110 |
| Insulin-dependent diabetes maturity onset | 4758121000006115 | 190372001 | 292552017 |
| DM + persistent proteinuria | 933221000006116 | 658061000000102 | 933221000006116 |
| Diabetes mellitus NOS with other specified manifestation | 12730981000006113 | 658061000000102 | 1445831000000112 |
| Cystic fibrosis related diabetes mellitus | 494831000000119 | 426705001 | 2674608014 |
| Diabetes mellitus with ketoacidotic coma | 616381000006111 | 26298008 | 44045012 |
| Non-insulin-dependent diabetes mellitus with retinopathy | 641581000006115 | 422034002 | 3035295017 |
| Type 1 diabetes mellitus with persistent proteinuria | 928461000006119 | 420514000 | 2967914011 |
| Diabetes mellitus type 1 with diabetic Charcot's arthropathy | 8022471000006114 | 71771000119100 | 3028508015 |
| Diabetes type 2 with ketoacidosis | 6977431000006113 | 421750000 | 3035594017 |
| Diabetes mellitus, adult onset, + neurological manifestation | 616481000006119 | 421326000 | 2618196010 |
| Diabetes mellitus, adult onset, with ketoacidosis | 616531000006116 | 421750000 | 2618220016 |
| Diabetes mellitus, adult onset, with ketoacidotic coma | 616541000006114 | 421847006 | 2618218019 |
| Diabetes mellitus, juvenile type, with ketoacidosis | 616641000006110 | 420270002 | 2618216015 |
| Diabetes mellitus, juvenile type, with ketoacidotic coma | 616651000006112 | 421075007 | 2618217012 |
| Type 1 diabetes mellitus with gangrene | 292541011 | 420825003 | 2618207019 |
| Diabetes mellitus caused by non-steroid drugs without complication | 6837991000006112 | 413183008 | 3290307015 |
| Ketoacidosis in type I diabetes mellitus | 6951081000006112 | 420270002 | 2618216015 |
| Type II diabetes mellitus with exudative maculopathy | 938331000006118 | 421779007 | 3698529010 |
| Type 1 diabetes mellitus with hypoglycaemic coma | 84341000006119 | 314771006 | 459161015 |
| Type 2 diabetes mellitus | 84471000006119 | 44054006 | 197761014 |
| Type 2 diabetes mellitus with hypoglycaemic coma | 84521000006116 | 719216001 | 3316336015 |
| Type 2 diabetes mellitus with nephropathy | 84551000006113 | 420279001 | 3035432019 |
| Type 2 diabetes mellitus with ulcer | 84631000006119 | 190389009 | 292579018 |
| Type I diabetes mellitus with renal complications | 84801000006111 | 421893009 | 2623054013 |
| Type II diabetes mellitus with ulcer | 85001000006117 | 190389009 | 292581016 |
| Diabetes mellitus type 2 | 3209441000006114 | 44054006 | 197763012 |
| Diabetes mellitus type 1 with ketoacidotic coma | 6965571000006111 | 421075007 | 3035596015 |
| Maturity onset diabetes in youth type 2 | 356085010 | 237604008 | 356085010 |
| NIDDM with peripheral circulatory disorder | 674961000006118 | 422166005 | 2623039019 |
| Non-insulin-dependent diabetes mellitus with renal comps | 587521000006111 | 420279001 | 2615535015 |
| Diabetes mellitus, adult, + peripheral circulatory disorder | 616581000006115 | 422166005 | 2618204014 |
| Type I diabetes mellitus with diabetic cataract | 913911000006110 | 421920002 | 3688521010 |
| Type 2 diabetes mellitus with diabetic cataract | 914301000006111 | 420756003 | 3688522015 |
| Insulin-dependent diabetes mellitus with ophthalmic comps | 913471000006110 | 25093002 | 3688467012 |
| Type I diabetes mellitus with gangrene | 292543014 | 420825003 | 2618207019 |
| Diabetes mellitus, juvenile, + neurological manifestation | 616671000006119 | 421468001 | 2618197018 |

| | | | |
|---|---|---|---|
| Pre-existing diabetes mellitus, non-insulin-dependent | 306113018 | 199230006 | 306113018 |
| Type I diabetes mellitus with polyneuropathy | 84791000006110 | 713705003 | 3297390014 |
| Diabetes mellitus NOS with ophthalmic manifestation | 292503016 | 25093002 | 42062018 |
| Type 1 diabetes mellitus with diabetic cataract | 913901000006112 | 421920002 | 2618234016 |
| Type 2 diabetes mellitus with mononeuropathy | 914221000006113 | 420436000 | 2618201018 |
| Labile type I diabetes mellitus | 5754621000006110 | 290002008 | 2872768015 |
| Diabetes mellitus NOS with no mention of complication | 292475010 | 111552007 | 178796018 |
| Insulin dependent diabetes mellitus with diabetic cataract | 771341000006114 | 43959009 | 73294018 |
| Insulin dependent diabetes mellitus | 772161000006119 | 73211009 | 121589010 |
| Insulin dependent diabetes mellitus with multiple complicat | 913531000006111 | 385041000000108 | 760111000000115 |
| Type II diabetes mellitus with hypoglycaemic coma | 84891000006116 | 719216001 | 3316336015 |
| Atypical diabetes mellitus | 11633211000006112 | 530558861000132104 | 3334354013 |
| Acute complication co-occurrent and due to diabetes mellitus | 3636752017 | 762489000 | 3636752017 |
| Type II diabetes mellitus with ophthalmic complications | 12702421000006111 | 422099009 | 12702421000006111 |
| Type 2 diabetes mellitus with diabetic cataract | 84501000006114 | 420756003 | 3688522015 |
| Type I diabetes mellitus with retinopathy | 84811000006114 | 420789003 | 3699407019 |
| Diabetes mellitus, juvenile type, with renal manifestation | 616661000006114 | 421893009 | 2618211013 |
| Type 1 diabetes mellitus with renal complications | 913451000006117 | 421893009 | 2618211013 |
| Type II diabetes mellitus with ophthalmic complications | 914061000006110 | 422099009 | 3698440016 |
| Insulin-dependent diabetes mellitus with neurological comps | 913501000006115 | 422088007 | 3688477014 |
| Type 2 diabetes mellitus with gangrene | 292583018 | 421631007 | 2618206011 |
| Other specified diabetes mellitus with ophthalmic complicatn | 13771000006110 | 25093002 | 42062018 |
| Type 1 diabetes mellitus with mononeuropathy | 84351000006117 | 420918009 | 2618199015 |
| Type 1 diabetes mellitus with neurological complications | 84381000006113 | 421468001 | 2618197018 |
| Type I diabetes mellitus with gangrene | 84701000006115 | 420825003 | 2618207019 |
| Type 1 diabetes mellitus with neurological complications | 913511000006117 | 421468001 | 2618197018 |
| Diabetes mellitus, juvenile type, with hyperosmolar coma | 292482014 | 190330002 | 292482014 |
| Diabetes mellitus, adult onset, with hyperosmolar coma | 292483016 | 190331003 | 292483016 |
| Type 2 diabetes mellitus with multiple complications | 292577016 | 190388001 | 292577016 |
| Diabetes mellitus caused by insulin receptor antibodies | 3291658019 | 75682002 | 3291658019 |
| History of diabetes mellitus | 4539561000006114 | 161445009 | 2986596017 |
| Type 1 diabetes mellitus with gastroparesis | 299601000000114 | 713702000 | 3698406016 |
| Diabetes mellitus with ophthalmic manifestation | 616421000006118 | 25093002 | 3688467012 |
| Diabetes mellitus with renal manifestation | 616461000006112 | 127013003 | 301016 |
| Insulin treated Type 2 diabetes mellitus | 840951000006119 | 237599002 | 2967820013 |
| Lipoatrophic diabetes mellitus | 967701000006116 | 127012008 | 300015 |

| | | | |
|---|---|---|---|
| Pre-existing diabetes mellitus, insulin-dependent | 306112011 | 385051000000106 | 760131000000111 |
| Diabetes mellitus, adult onset, no mention of complication | 616511000006110 | 313436004 | 457329019 |
| Insulin dependent diabetes mellitus | 772171000006114 | 73211009 | 121589010 |
| Insulin dependent diabetes maturity onset | 913711000006112 | 73211009 | 121589010 |
| Insulin dependent diabetes mellitus with diabetic cataract | 913891000006113 | 43959009 | 73294018 |
| Type 2 diabetes mellitus with nephropathy | 914261000006119 | 420279001 | 3035432019 |
| DM induced by non-steroid drugs without complication | 967631000006115 | 413183008 | 2474729016 |
| Other specified diabetes mellitus with coma | 12725191000006116 | 630521000000101 | 1390421000000119 |
| Diabetes mellitus with other specified manifestation | 12730971000006110 | 658011000000104 | 1445721000000115 |
| Ketoacidotic coma in type 2 diabetes mellitus | 12704991000006114 | 421847006 | 2967777017 |
| Type II diabetes mellitus with neurological complications | 12702411000006115 | 421326000 | 84931000006113 |
| Insulin dependent diabetes mellitus with ulcer | 913591000006110 | 422183001 | 2623058011 |
| H/O: diabetes mellitus | 251591016 | 161445009 | 251591016 |
| Type 1 diabetes mellitus with retinopathy | 84441000006110 | 420789003 | 3699407019 |
| Type II diabetes mellitus with retinopathy | 84991000006112 | 422034002 | 3699410014 |
| Type 1 diabetes mellitus with gangrene | 84331000006112 | 420825003 | 2618207019 |
| Type II diabetes mellitus with gangrene | 84881000006119 | 421631007 | 2618206011 |
| Type 1 diabetes mellitus with ophthalmic complications | 913481000006113 | 739681000 | 3537386015 |
| Diabetic on oral treatment | 264681018 | 170746002 | 264681018 |
| Insulin dependent diabetes mellitus with ulcer | 11927551000006115 | 19429009 | 913591000006110 |
| Latent autoimmune diabetes mellitus in adult (LADA) | 7065531000006115 | 426875007 | 2674066012 |
| Diabetic ketoacidosis | 6953861000006110 | 420422005 | 2616611010 |
| MODY5 - maturity-onset diabetes of the young type 5 | 7500341000006113 | 609572000 | 3435008016 |
| Diabetes type 1 with neurological disorder | 6972451000006115 | 421468001 | 3035338016 |
| Type II diabetes mellitus with ophthalmic complications | 84951000006118 | 422099009 | 3698440016 |
| Diabetes mellitus with peripheral circulatory disorder | 616441000006113 | 421895002 | 3688501014 |
| Type 2 diabetes mellitus with neurological complications | 914071000006115 | 421326000 | 3695406015 |
| Type 2 diabetes mellitus with retinopathy | 914151000006112 | 422034002 | 3699410014 |
| Type 2 diabetes mellitus with ketoacidotic coma | 928601000006118 | 421847006 | 2618218019 |
| Type 1 diabetes mellitus | 84281000006115 | 46635009 | 197984010 |
| Type 1 diabetes mellitus maturity onset | 84301000006116 | 190372001 | 292553010 |
| Type 1 diabetes mellitus with arthropathy | 84311000006118 | 314893005 | 459292011 |
| Type 2 diabetes mellitus without complication | 84641000006112 | 313436004 | 457329019 |
| Type I diabetes mellitus without complication | 84831000006115 | 313435000 | 457325013 |
| Type II diabetes mellitus with arthropathy | 84861000006112 | 314903002 | 459311019 |
| Non-insulin dependent diabetes mellitus with nephropathy | 280531000006119 | 420279001 | 3035432019 |
| Type 1 diabetes mellitus without complication | 84461000006114 | 313435000 | 457326014 |
| Type I diabetes mellitus with nephropathy | 84741000006118 | 421893009 | 2623054013 |

| | | | |
|---|---|---|---|
| Type II diabetes mellitus with multiple complications | 84911000006119 | 190388001 | 292576013 |
| Type II diabetes mellitus without complication | 85011000006119 | 313436004 | 457330012 |
| Non-insulin dependent diabetes mellitus with polyneuropathy | 280541000006112 | 713706002 | 3297353013 |
| Non-insulin depend diabetes mellitus with diabetic cataract | 281161000006114 | 420756003 | 3035281015 |
| Diabetes mellitus NOS with other specified manifestation | 292621010 | 73211009 | 121589010 |
| Diabetes mellitus with unspecified complication | 292622015 | 74627003 | 123939015 |
| Type I diabetes mellitus uncontrolled | 7287621000006117 | 444073006 | 2842075017 |
| Type 1 diabetes mellitus with hypoglycaemic coma | 459161015 | 314771006 | 459161015 |
| Type 1 diabetes mellitus with neuropathic arthropathy | 459296014 | 71771000119100 | 3010513018 |
| Type II diabetes mellitus with arthropathy | 459311019 | 314903002 | 459311019 |
| Diabetes mellitus -adult onset | 881451000006113 | 532411000000102 | 881451000006113 |
| Diabetes + other complications | 881521000006117 | 658011000000104 | 881521000006117 |
| Diabetic on diet only | 264679015 | 170745003 | 264679015 |
| Type 2 diabetes mellitus without complication | 457329019 | 313436004 | 457329019 |
| Type II diabetes mellitus without complication | 457330012 | 313436004 | 457330012 |
| Diabetic on insulin and oral treatment | 458512016 | 314194001 | 458512016 |
| Type II diabetes mellitus with neuropathic arthropathy | 459312014 | 314904008 | 459312014 |
| Type 2 diabetes mellitus with neuropathic arthropathy | 459313016 | 314904008 | 459313016 |
| Maturity onset diabetes of the young, type 2 | 5109561000006119 | 237604008 | 2967875018 |
| Diabetic ketoacidosis without coma | 178798017 | 111556005 | 178798017 |
| Type 2 diabetes mellitus with hypoglycaemic coma | 459167016 | 719216001 | 3316336015 |
| Maturity onset diabetes in youth type 1 | 2476117016 | 609562003 | 2967867012 |
| Other specified diabetes mellitus with unspecified comps | 13811000006110 | 74627003 | 123939015 |
| Diabetes mellitus type I | 3253231000006110 | 46635009 | 77728011 |
| Type I diabetes mellitus with ulcer | 292538019 | 190368000 | 292538019 |
| Type 2 diabetes mellitus with ulcer | 292579018 | 190389009 | 292579018 |
| Type II diabetes mellitus with ulcer | 292581016 | 190389009 | 292581016 |
| Pre-existing malnutrition-related diabetes mellitus | 306114012 | 199231005 | 306114012 |
| Type 1 diabetes mellitus - poor control | 292548017 | 444073006 | 2872487013 |
| Lipoatrophic diabetes mellitus without complication | 189711000000119 | 112991000000101 | 189711000000119 |
| Secondary diabetes mellitus | 15518018 | 8801005 | 15518018 |
| Type 2 diabetes mellitus - poor control | 292590011 | 443694000 | 2921019012 |
| Brittle diabetes | 19931010 | 11530004 | 19931010 |
| [X]Pre-existing diabetes mellitus, unspecified | 308110013 | 385051000000106 | 760131000000111 |
| Insulin-dependent diabetes without complication | 457327017 | 111552007 | 178796018 |
| Insulin dependent diabetes mellitus with arthropathy | 459293018 | 39710007 | 1229552016 |
| Insulin dependent diabetes mellitus with hypoglycaemic coma | 771361000006113 | 237632004 | 356141011 |

| | | | |
|---|---|---|---|
| Insulin dependent diabetes mellitus with mononeuropathy | 913771000006115 | 230577008 | 345492010 |
| Insulin dependent diabetes mellitus with polyneuropathy | 913801000006118 | 49455004 | 82373015 |
| Ketoacidotic coma in type I diabetes mellitus | 6965561000006116 | 421075007 | 2618217012 |
| Type 1 diabetes mellitus | 197984010 | 46635009 | 197984010 |
| Type II diabetes mellitus | 493774016 | 44054006 | 493774016 |
| Diabetes mellitus due to insulin receptor antibodies | 125705011 | 75682002 | 125705011 |
| Insulin-dependent diabetes mellitus secretory diarrhoea syndrome | 356111014 | 237618001 | 356111014 |
| Lipodystrophic diabetes | 4392151000006119 | 127012008 | 205214013 |
| Maturity onset diabetes of the young, type 1 | 7500191000006118 | 609562003 | 2967867012 |
| Latent autoimmune diabetes mellitus in adult | 2674067015 | 426875007 | 2674067015 |
| Other specified diabetes mellitus with other spec comps | 13781000006113 | 73211009 | 121589010 |
| Unstable type I diabetes mellitus | 72721000006113 | 290002008 | 429970018 |
| Type 2 diabetes mellitus with multiple complications | 84541000006111 | 190388001 | 292577016 |
| Type I diabetes mellitus | 84651000006114 | 46635009 | 494564012 |
| Type I diabetes mellitus maturity onset | 84671000006116 | 190372001 | 292551012 |
| Type I diabetes mellitus with neuropathic arthropathy | 84761000006119 | 71771000119100 | 3010513018 |
| Type II diabetes mellitus - poor control | 84851000006110 | 443694000 | 2921019012 |
| Type II diabetes mellitus with neuropathic arthropathy | 84941000006115 | 314904008 | 459312014 |
| Type I diabetes mellitus with polyneuropathy | 913821000006111 | 713705003 | 3297390014 |
| Insulin dependent diabetes mellitus | 77727018 | 73211009 | 121589010 |
| Insulin dependent diabetes mellitus with gangrene | 771351000006111 | 422275004 | 2616612015 |
| Insulin dependent diabetes mellitus with nephropathy | 771391000006117 | 127013003 | 301016 |
| Insulin dependent diabetes mellitus with gangrene | 913621000006112 | 422275004 | 2616612015 |
| Pre-existing diabetes mellitus in pregnancy | 7500211000006117 | 609563008 | 2967860014 |
| Secondary pancreatic diabetes mellitus | 1230929011 | 51002006 | 1230929011 |
| Diabetes mellitus induced by non-steroid drugs | 2160090014 | 408540003 | 2160090014 |
| Acute complication with diabetes mellitus | 3636751012 | 762489000 | 3636751012 |
| Insulin dependent diabetes mellitus with hypoglycaemic coma | 6050041000006115 | 314771006 | 459162010 |
| Secondary pancreatic diabetes mellitus without complication | 198461000000116 | 51002006 | 84990019 |
| Brittle type 1 diabetes mellitus | 5754581000006110 | 290002008 | 2872766016 |
| Diabetes mellitus with gangrene | 616351000006115 | 422275004 | 2616612015 |
| Diabetes mellitus, adult, + other specified manifestation | 616571000006118 | 73211009 | 121589010 |
| Diabetes with gangrene | 616831000006118 | 422275004 | 2616612015 |
| Type I diabetes mellitus with multiple complications | 913551000006116 | 422228004 | 2967771016 |
| Type II diabetes mellitus with persistent proteinuria | 928551000006115 | 421986006 | 2967751013 |
| Diabetic acidosis with coma | 2920971000006115 | 26298008 | 44046013 |
| Insulin dependent diabetes mellitus with hypoglycemic coma | 6050071000006111 | 314771006 | 459165012 |

| | | | |
|---|---|---|---|
| Diabetes mellitus caused by non-steroid drugs | 6763751000006119 | 408540003 | 3290374011 |
| Type 2 diabetes mellitus with persistent microalbuminuria | 928561000006118 | 420715001 | 2967769016 |
| Type I diabetes mellitus | 494564012 | 46635009 | 494564012 |
| Type II diabetes mellitus with nephropathy | 84921000006110 | 420279001 | 3013392012 |
| Non-insulin dependent diabetes mellitus | 280571000006116 | 44054006 | 493773010 |
| Type 2 diabetes mellitus with polyneuropathy | 914241000006118 | 713706002 | 3297353013 |
| Type 1 diabetes mellitus with ketoacidotic coma | 928521000006112 | 421075007 | 2967758019 |
| Type II diabetes mellitus with ketoacidotic coma | 928611000006115 | 421847006 | 2618218019 |
| Diabetic on oral treatment and glucagon-like peptide 1 | 1966311000006116 | 976341000000101 | 2483721000000113 |
| Pre-existing type 2 diabetes mellitus in pregnancy | 1988741000006117 | 237627000 | 356133011 |
| Other specified diabetes mellitus with coma | 292488013 | 420662003 | 2618230013 |
| Unstable insulin dependent diabetes mellitus | 429972014 | 11530004 | 19931010 |
| Diabetic on subcutaneous treatment | 1177721000000112 | 527691000000107 | 1177721000000112 |
| Type 2 diabetes mellitus uncontrolled | 7281511000006118 | 443694000 | 3321322011 |
| Diabetes type 1 with cataract | 6980441000006114 | 421920002 | 3035484013 |
| Diabetes mellitus NOS with peripheral circulatory disorder | 292523015 | 421895002 | 2618203015 |
| Pre-existing type 2 diabetes mellitus | 4796321000006113 | 199230006 | 2967812011 |
| Non-insulin dependent diabetes mellitus - poor control | 281171000006119 | 443694000 | 2842387018 |
| Diabetes mellitus with no mention of complication | 292466013 | 111552007 | 178796018 |
| Unstable insulin dependent diabetes mellitus | 72651000006114 | 11530004 | 19931010 |
| Unstable type 1 diabetes mellitus | 429971019 | 290002008 | 429971019 |
| Insulin treated Type II diabetes mellitus | 1223147012 | 237599002 | 1223147012 |
| Pre-existing diabetes mellitus | 4539551000006112 | 161445009 | 2820057017 |
| Type 2 diabetes mellitus with ketoacidosis | 928581000006111 | 421750000 | 2967754017 |
| Unspecified diabetes mellitus with multiple complications | 292565014 | 385041000000108 | 760111000000115 |
| Diabetic on non-insulin injectable | 1780981000006115 | 1780981000006104 | 1780981000006115 |
| Type I diabetes mellitus with hypoglycaemic coma | 459163017 | 314771006 | 459163017 |
| Type 2 diabetes mellitus with gastroparesis | 299621000000117 | 713703005 | 3698412014 |
| Type 1 diabetes mellitus with exudative maculopathy | 938301000006114 | 420486006 | 3698521013 |
| Type I diabetes mellitus with neuropathic arthropathy | 459295013 | 71771000119100 | 3010513018 |
| Type 1 diabetes mellitus with neuropathic arthropathy | 84391000006111 | 71771000119100 | 3010513018 |
| Type 2 diabetes mellitus with renal complications | 84611000006113 | 420279001 | 2618208012 |
| Type II diabetes mellitus | 84841000006113 | 44054006 | 493774016 |
| Type II diabetes mellitus with peripheral angiopathy | 84961000006116 | 314902007 | 459306016 |
| Type I diabetes mellitus with diabetic cataract | 84691000006115 | 421920002 | 3688521010 |
| Diabetes mellitus with neurological manifestation | 616391000006114 | 422088007 | 3688477014 |
| Type 2 diabetes mellitus with exudative maculopathy | 938321000006116 | 421779007 | 2618237011 |
| Non-insulin-dependent diabetes mellitus with neuro comps | 280591000006115 | 421326000 | 3695406015 |

| | | | |
|---|---|---|---|
| Non-insulin dependent diabetes mellitus with hypoglyca coma | 280511000006113 | 719216001 | 3316336015 |
| Diabetes mellitus uncontrolled | 5505651000006113 | 268519009 | 2920470017 |
| Diabetic on non-insulin injectable medication | 2460251000000115 | 719566006 | 3316952014 |
| Brittle type I diabetes mellitus | 5754611000006119 | 290002008 | 2872767013 |
| Diabetes type 1 with retinopathy | 6960441000006112 | 420789003 | 3035448018 |
| Type 1 diabetes mellitus with renal complications | 84431000006117 | 421893009 | 2618211013 |
| Type 2 diabetes mellitus with neurological complications | 84561000006110 | 421326000 | 3695406015 |
| Type 2 diabetes mellitus with ophthalmic complications | 84581000006117 | 422099009 | 3698440016 |
| Type II diabetes mellitus with gangrene | 292582011 | 421631007 | 2618206011 |

#### Table S11. Anti-Diabetic Aurum product codes

| Term from EMIS | Prod code ID | dmd ID | Drug substance name |
|---|---|---|---|
| Insulin aspart 100units/ml solution for injection 3ml cartridges | 1862041000033113 | 3468611000001108 | Insulin aspart |
| NovoRapid Penfill 100units/ml solution for injection 3ml cartridges (Novo Nordisk Ltd) | 1862341000033110 | 3279211000001105 | Insulin aspart |
| Fiasp Penfill 100units/ml solution for injection 3ml cartridges (Novo Nordisk Ltd) | 12186641000033113 | 34043211000001101 | Insulin aspart |
| NovoRapid Novolet 100units/ml solution for injection (Novo Nordisk Ltd) | 1862441000033116 | 3281611000001102 | Insulin aspart |
| NovoRapid FlexPen 100units/ml solution for injection 3ml pre-filled pens (Novo Nordisk Ltd) | 2796641000033115 | 3282211000001106 | Insulin aspart |
| Insulin aspart 100units/ml solution for injection 3ml pre-filled disposable devices | 3277741000033118 | 3468711000001104 | Insulin aspart |
| NovoRapid FlexTouch 100units/ml solution for injection 3ml pre-filled pens (Novo Nordisk Ltd) | 6456441000033119 | 19570211000001106 | Insulin aspart |
| Fiasp FlexTouch 100units/ml solution for injection 3ml pre-filled pens (Novo Nordisk Ltd) | 12186541000033112 | 34043011000001106 | Insulin aspart |
| NovoMix 30 Penfill 100units/ml suspension for injection 3ml cartridges (Novo Nordisk Ltd) | 2724741000033111 | 3277811000001103 | Insulin aspart/ Insulin aspart protamine |
| NovoMix 30 FlexPen 100units/ml suspension for injection 3ml pre-filled pens (Novo Nordisk Ltd) | 2724841000033118 | 3277211000001104 | Insulin aspart/ Insulin aspart protamine |
| Human Mixtard 10 Penfill 100units/ml suspension for injection 1.5ml cartridges (Novo Nordisk Ltd) | 1818141000033110 | 3268711000001103 | Insulin isophane human/ Insulin soluble human |
| Mixtard 10 Penfill 100units/ml suspension for injection 3ml cartridges (Novo Nordisk Ltd) | 2953541000033112 | 3264711000001106 | Insulin isophane human/ Insulin soluble human |
| Mixtard 10 NovoLet 100units/ml suspension for injection (Novo Nordisk Ltd) | 2953441000033111 | 3269711000001107 | Insulin isophane human/ Insulin soluble human |
| Insuman Comb 15 100units/ml suspension for injection 3ml cartridges (Sanofi) | 2158541000033117 | 3273911000001104 | Insulin isophane human/ Insulin soluble human |
| Insuman Comb 15 100units/ml suspension for injection 3ml pre-filled OptiSet pens (Sanofi) | 2267041000033118 | 3270211000001103 | Insulin isophane human/ Insulin soluble human |
| Insuman Comb 15 100units/ml suspension for injection 5ml vials (Aventis Pharma) | 2158441000033118 | 3271711000001102 | Insulin isophane human/ Insulin soluble human |
| Human Mixtard 20 Penfill 100units/ml suspension for injection 1.5ml cartridges (Novo Nordisk Ltd) | 1818241000033115 | 3267811000001107 | Insulin isophane human/ Insulin soluble human |
| Mixtard 20 Penfill 100units/ml suspension for injection 3ml cartridges (Novo Nordisk Ltd) | 2953741000033116 | 3264411000001100 | Insulin isophane human/ Insulin soluble human |
| Humulin M2 100units/ml suspension for injection 3ml cartridges (Eli Lilly and Company Ltd) | 1616241000033111 | 3263611000001100 | Insulin isophane human/ Insulin soluble human |
| Mixtard 20 NovoLet 100units/ml suspension for injection (Novo Nordisk Ltd) | 2953641000033113 | 3266111000001103 | Insulin isophane human/ Insulin soluble human |
| Insuman Comb 25 100units/ml suspension for injection 3ml cartridges (Sanofi) | 2035641000033115 | 3264111000001105 | Insulin isophane human/ Insulin soluble human |
| Insuman Comb 25 100units/ml suspension for injection 3ml pre-filled OptiSet pens (Sanofi) | 2267141000033119 | 3259811000001109 | Insulin isophane human/ Insulin soluble human |

| | | | |
|---|---|---|---|
| Insuman Comb 25 100units/ml suspension for injection 3ml pre-filled SoloStar pens (Sanofi) | 6011841000033112 | 18150311000001109 | Insulin isophane human/ Insulin soluble human |
| Insuman Comb 25 100units/ml suspension for injection 5ml vials (Sanofi) | 2035541000033116 | 3263011000001107 | Insulin isophane human/ Insulin soluble human |
| Human Mixtard 30 Penfill 100units/ml suspension for injection 1.5ml cartridges (Novo Nordisk Ltd) | 1818341000033113 | 3275311000001104 | Insulin isophane human/ Insulin soluble human |
| Humulin M3 100units/ml suspension for injection 10ml vials (Eli Lilly and Company Ltd) | 725341000033110 | 3274811000001107 | Insulin isophane human/ Insulin soluble human |
| Mixtard 30 100units/ml suspension for injection 10ml vials (Novo Nordisk Ltd) | 2953941000033118 | 3275011000001102 | Insulin isophane human/ Insulin soluble human |
| Mixtard 30 Penfill 100units/ml suspension for injection 3ml cartridges (Novo Nordisk Ltd) | 2954041000033116 | 3273111000001102 | Insulin isophane human/ Insulin soluble human |
| Humulin M3 100units/ml suspension for injection 3ml cartridges (Eli Lilly and Company Ltd) | 1616341000033118 | 3273611000001105 | Insulin isophane human/ Insulin soluble human |
| HumaJect M3 Pen 100units/ml suspension for injection (Eli Lilly and Company Ltd) | 727341000033117 | 3272211000001102 | Insulin isophane human/ Insulin soluble human |
| Mixtard 30 InnoLet 100units/ml suspension for injection 3ml pre-filled pens (Novo Nordisk Ltd) | 2644741000033116 | 3271611000001106 | Insulin isophane human/ Insulin soluble human |
| Mixtard 30 NovoLet 100units/ml suspension for injection (Novo Nordisk Ltd) | 2953841000033114 | 3270511000001100 | Insulin isophane human/ Insulin soluble human |
| Humulin M3 Pen 100units/ml suspension for injection 3ml pre-filled pens (Eli Lilly and Company Ltd) | 3333841000033113 | 9437511000001101 | Insulin isophane human/ Insulin soluble human |
| Humulin M3 KwikPen 100units/ml suspension for injection 3ml pre-filled pens (Eli Lilly and Company Ltd) | 5910741000033117 | 17609511000001109 | Insulin isophane human/ Insulin soluble human |
| Human Mixtard 40 Penfill 100units/ml suspension for injection 1.5ml cartridges (Novo Nordisk Ltd) | 1818441000033119 | 3285511000001103 | Insulin isophane human/ Insulin soluble human |
| Mixtard 40 Penfill 100units/ml suspension for injection 3ml cartridges (Novo Nordisk Ltd) | 2954241000033112 | 3281211000001104 | Insulin isophane human/ Insulin soluble human |
| Mixtard 40 NovoLet 100units/ml suspension for injection (Novo Nordisk Ltd) | 2954141000033117 | 3272011000001107 | Insulin isophane human/ Insulin soluble human |
| Human Mixtard 50 Penfill 100units/ml suspension for injection 1.5ml cartridges (Novo Nordisk Ltd) | 1818541000033118 | 3273411000001107 | Insulin isophane human/ Insulin soluble human |
| Humulin M5 100units/ml suspension for injection 10ml vials (Eli Lilly and Company Ltd) | 722841000033112 | 3271011000001104 | Insulin isophane human/ Insulin soluble human |
| Insuman Comb 50 100units/ml suspension for injection 3ml cartridges (Sanofi) | 2158741000033113 | 3272411000001103 | Insulin isophane human/ Insulin soluble human |
| Mixtard 50 Penfill 100units/ml suspension for injection 3ml cartridges (Novo Nordisk Ltd) | 2954541000033114 | 3272811000001101 | Insulin isophane human/ Insulin soluble human |
| Mixtard 50 NovoLet 100units/ml suspension for injection (Novo Nordisk Ltd) | 2954341000033119 | 3278311000001108 | Insulin isophane human/ Insulin soluble human |
| Insuman Comb 50 100units/ml suspension for injection 3ml pre-filled OptiSet pens (Sanofi) | 2267241000033114 | 3278611000001103 | Insulin isophane human/ Insulin soluble human |

| | | | |
|---|---|---|---|
| Insuman Comb 50 100units/ml suspension for injection 5ml vials (Aventis Pharma) | 2158641000033116 | 3274511000001109 | Insulin isophane human/ Insulin soluble human |
| Hypurin Porcine 30/70 Mix 100units/ml suspension for injection 1.5ml cartridges (C P Pharmaceuticals Ltd) | 2018941000033112 | 3277711000001106 | Insulin isophane porcine/ Insulin soluble porcine |
| Hypurin Porcine 30/70 Mix 100units/ml suspension for injection 10ml vials (Wockhardt UK Ltd) | 2018841000033116 | 3266811000001105 | Insulin isophane porcine/ Insulin soluble porcine |
| Pork Mixtard 30 100units/ml suspension for injection 10ml vials (Novo Nordisk Ltd) | 915641000033117 | 3267911000001102 | Insulin isophane porcine/ Insulin soluble porcine |
| Hypurin Bovine Isophane 100units/ml suspension for injection 10ml vials (Wockhardt UK Ltd) | 735841000033116 | 3280011000001100 | Insulin isophane bovine |
| Insulin isophane bovine 100units/ml suspension for injection 10ml vials | 1821641000033119 | 3472311000001106 | Insulin isophane bovine |
| Human Insulatard Penfill 100units/ml suspension for injection 1.5ml cartridges (Novo Nordisk Ltd) | 1714041000033116 | 3259411000001107 | Insulin isophane human |
| Humulin I 100units/ml suspension for injection 10ml vials (Eli Lilly and Company Ltd) | 724041000033112 | 3255911000001101 | Insulin isophane human |
| Insulatard 100units/ml suspension for injection 10ml vials (Novo Nordisk Ltd) | 2953241000033110 | 3256111000001105 | Insulin isophane human |
| Insulin isophane human 100units/ml suspension for injection 10ml vials | 779141000033111 | 3472511000001100 | Insulin isophane human |
| Insuman Basal 100units/ml suspension for injection 5ml vials (Sanofi) | 2035241000033118 | 3258411000001104 | Insulin isophane human |
| Insulin isophane porcine 100units/ml suspension for injection 10ml vials | 1821441000033116 | 3473011000001104 | Insulin isophane porcine |
| Hypurin Porcine Isophane 100units/ml suspension for injection 10ml vials (Wockhardt UK Ltd) | 734941000033111 | 3284011000001107 | Insulin isophane porcine |
| Pork Insulatard 100units/ml suspension for injection 10ml vials (Novo Nordisk Ltd) | 763341000033116 | 3333111000001107 | Insulin isophane porcine |
| Humalog Mix25 Pen 100units/ml suspension for injection 3ml pre-filled pens (Waymade Healthcare Plc) | 1737041000033117 | 5268611000001103 | Insulin lispro/ Insulin lispro protamine |
| Humalog Mix25 KwikPen 100units/ml suspension for injection 3ml pre-filled pens (Eli Lilly and Company Ltd) | 4608641000033119 | 13884711000001104 | Insulin lispro/ Insulin lispro protamine |
| Humalog Mix50 KwikPen 100units/ml suspension for injection 3ml pre-filled pens (Eli Lilly and Company Ltd) | 4608741000033111 | 13884911000001102 | Insulin lispro/ Insulin lispro protamine |
| Humalog Mix50 Pen 100units/ml suspension for injection 3ml pre-filled pens (Sigma Pharmaceuticals Plc) | 2182141000033117 | 14248811000001102 | Insulin lispro/ Insulin lispro protamine |
| Humalog Mix25 100units/ml suspension for injection 3ml cartridges (Eli Lilly and Company Ltd) | 1736941000033118 | 3275711000001100 | Insulin lispro/ Insulin lispro protamine |
| Hypurin Bovine Lente 100units/ml suspension for injection 10ml vials (Wockhardt UK Ltd) | 735941000033112 | 3280611000001107 | Insulin zinc suspension mixed bovine |
| Hypurin Porcine 30/70 Mix 100units/ml suspension for injection 3ml cartridges (Wockhardt UK Ltd) | 2917341000033116 | 4029411000001100 | Insulin isophane porcine/ Insulin soluble porcine |
| Hypurin Bovine Isophane 100units/ml suspension for injection 3ml cartridges (Wockhardt UK Ltd) | 2917041000033118 | 4028311000001101 | Insulin isophane bovine |
| Insulin isophane bovine 100units/ml suspension for injection 3ml cartridges | 1821741000033111 | 4033211000001103 | Insulin isophane bovine |
| Hypurin Bovine Neutral 100units/ml solution for injection 3ml cartridges (Wockhardt UK Ltd) | 2916941000033119 | 4028811000001105 | Insulin soluble bovine |
| Insulin isophane porcine 100units/ml suspension for injection 3ml cartridges | 1821541000033115 | 4053611000001100 | Insulin isophane porcine |
| Hypurin Porcine Isophane 100units/ml suspension for injection 3ml cartridges (Wockhardt UK Ltd) | 2917241000033114 | 4034311000001108 | Insulin isophane porcine |

| | | | |
|---|---|---|---|
| Hypurin Porcine Neutral 100units/ml solution for injection 3ml cartridges (Wockhardt UK Ltd) | 2917141000033119 | 4034911000001109 | Insulin soluble porcine |
| Insulin detemir 100units/ml solution for injection 3ml pre-filled disposable devices | 3136941000033116 | 7594211000001102 | Insulin detemir |
| Levemir FlexPen 100units/ml solution for injection 3ml pre-filled pens (Novo Nordisk Ltd) | 3137241000033111 | 7589411000001100 | Insulin detemir |
| Levemir InnoLet 100units/ml solution for injection 3ml pre-filled pens (Novo Nordisk Ltd) | 4126541000033110 | 11148111000001101 | Insulin detemir |
| Insulin detemir 100units/ml solution for injection 3ml cartridges | 3137041000033115 | 7597611000001106 | Insulin detemir |
| Levemir Penfill 100units/ml solution for injection 3ml cartridges (Novo Nordisk Ltd) | 3137141000033116 | 7589911000001108 | Insulin detemir |
| Insulin glulisine 100units/ml solution for injection 3ml cartridges | 3345741000033117 | 9532111000001100 | Insulin glulisine |
| Apidra 100units/ml solution for injection 3ml cartridges (Sanofi) | 3345941000033119 | 9528311000001103 | Insulin glulisine |
| Apidra 100units/ml solution for injection 3ml OptiClik cartridges (Sanofi) | 3963941000033117 | 10898411000001103 | Insulin glulisine |
| Insulin glulisine 100units/ml solution for injection 3ml pre-filled disposable devices | 3914241000033112 | 10097211000001102 | Insulin glulisine |
| Apidra 100units/ml solution for injection 3ml pre-filled SoloStar pens (Sanofi) | 4258641000033112 | 12144611000001100 | Insulin glulisine |
| Humalog Mix50 100units/ml suspension for injection 3ml cartridges (Eli Lilly and Company Ltd) | 3922641000033111 | 10344911000001108 | Insulin lispro/ Insulin lispro protamine |
| Exubera 1mg inhalation powder blisters (Pfizer Ltd) | 3945141000033114 | 10690511000001105 | Insulin human |
| Exubera 3mg inhalation powder blisters (Pfizer Ltd) | 3945241000033119 | 10690811000001108 | Insulin human |
| Insulin human 100units/ml solution for injection 10ml vials | 10673941000033110 | 13859411000001101 | Insulin human |
| Insuman Infusat 100units/ml solution for injection 10ml vials (Sanofi) | 10674141000033111 | 18030311000001100 | Insulin human |
| Humulin R 500units/ml solution for injection 20ml vials (Imported (United States)) | 4423041000033114 | 15603111000001104 | Insulin human |
| Humalog Mix25 100units/ml suspension for injection 10ml vials (Eli Lilly and Company Ltd) | 5403541000033118 | 16530311000001107 | Insulin lispro/ Insulin lispro protamine |
| Insulin human 100units/ml solution for injection 3.15ml cartridges | 10674041000033112 | 18083911000001104 | Insulin human |
| Insuman Infusat 100units/ml solution for injection 3.15ml cartridges (Sanofi) | 10674341000033114 | 18046311000001103 | Insulin human |
| Insulin degludec 100units/ml solution for injection 3ml cartridges | 8264341000033118 | 21939511000001100 | Insulin degludec |
| Tresiba Penfill 100units/ml solution for injection 3ml cartridges (Novo Nordisk Ltd) | 8264441000033112 | 21928511000001109 | Insulin degludec |
| Tresiba FlexTouch 100units/ml solution for injection 3ml pre-filled pens (Novo Nordisk Ltd) | 8264141000033116 | 21930011000001100 | Insulin degludec |
| Insulin degludec 100units/ml solution for injection 3ml pre-filled disposable devices | 8263941000033117 | 21939611000001101 | Insulin degludec |
| Insulin degludec 200units/ml solution for injection 3ml pre-filled disposable devices | 8264041000033115 | 21939711000001105 | Insulin degludec |
| Tresiba FlexTouch 200units/ml solution for injection 3ml pre-filled pens (Novo Nordisk Ltd) | 8264241000033111 | 21931911000001108 | Insulin degludec |
| Insulin aspart 100units/ml solution for injection 1.6ml cartridges | 9677041000033116 | 26655811000001102 | Insulin aspart |
| NovoRapid PumpCart 100units/ml solution for injection 1.6ml cartridges (Novo Nordisk Ltd) | 9677141000033117 | 26209611000001109 | Insulin aspart |
| Insulin degludec 100units/ml / Liraglutide 3.6mg/ml solution for injection 3ml pre-filled disposable devices | 10044541000033113 | 28279611000001109 | Insulin degludec/ Liraglutide |
| Xultophy 100units/ml / 3.6mg/ml solution for injection 3ml pre-filled pens (Novo Nordisk Ltd) | 10044641000033114 | 28054311000001106 | Insulin degludec/ Liraglutide |

| | | | |
|---|---|---|---|
| Lyumjev KwikPen 200units/ml solution for injection 3ml pre-filled pens (Eli Lilly and Company Ltd) | 13604941000033116 | 38501011000001104 | Insulin lispro |
| Insulin lispro 200units/ml solution for injection 3ml pre-filled disposable devices | 10252641000033111 | 28989711000001106 | Insulin lispro |
| Humalog KwikPen 200units/ml solution for injection 3ml pre-filled pens (Eli Lilly and Company Ltd) | 10252741000033119 | 28926811000001100 | Insulin lispro |
| Insulin glargine 300units/ml solution for injection 1.5ml pre-filled disposable devices | 10494241000033110 | 29903611000001103 | Insulin glargine |
| Toujeo 300units/ml solution for injection 1.5ml pre-filled SoloStar pens (Sanofi) | 10494441000033111 | 29866811000001104 | Insulin glargine |
| Humulin R KwikPen 500units/ml solution for injection 3ml pre-filled pens (Imported (United States)) | 12481641000033115 | 35214311000001104 | Insulin human |
| NovoRapid 100units/ml solution for injection 10ml vials (Novo Nordisk Ltd) | 1862241000033117 | 3280711000001103 | Insulin aspart |
| Insulin aspart 100units/ml solution for injection 10ml vials | 1861941000033119 | 36047011000001105 | Insulin aspart |
| Fiasp 100units/ml solution for injection 10ml vials (Novo Nordisk Ltd) | 12186441000033111 | 34043411000001102 | Insulin aspart |
| Lantus 100units/ml solution for injection 10ml vials (Sanofi) | 2780741000033111 | 3287911000001100 | Insulin glargine |
| Insulin glargine 100units/ml solution for injection 10ml vials | 2798841000033115 | 36047111000001106 | Insulin glargine |
| Apidra 100units/ml solution for injection 10ml vials (Sanofi) | 3345841000033110 | 9528811000001107 | Insulin glulisine |
| Insulin glulisine 100units/ml solution for injection 10ml vials | 3345641000033114 | 36047211000001100 | Insulin glulisine |
| Insuman Basal 100units/ml suspension for injection 3ml cartridges (Sanofi) | 2035441000033117 | 3265011000001108 | Insulin isophane human |
| Humulin I 100units/ml suspension for injection 3ml cartridges (Eli Lilly and Company Ltd) | 1615941000033113 | 3264211000001104 | Insulin isophane human |
| Insulatard Penfill 100units/ml suspension for injection 3ml cartridges (Novo Nordisk Ltd) | 2953341000033117 | 3263711000001109 | Insulin isophane human |
| Insulatard InnoLet 100units/ml suspension for injection 3ml pre-filled pens (Novo Nordisk Ltd) | 2644641000033113 | 3261411000001102 | Insulin isophane human |
| Insulatard FlexPen 100units/ml suspension for injection (Novo Nordisk Ltd) | 2796741000033112 | 3262011000001103 | Insulin isophane human |
| Insulatard NovoLet 100units/ml suspension for injection (Novo Nordisk Ltd) | 2953141000033115 | 3262511000001106 | Insulin isophane human |
| Insuman Basal 100units/ml suspension for injection 3ml pre-filled OptiSet pens (Sanofi) | 2266841000033110 | 3260611000001109 | Insulin isophane human |
| Humulin I Pen 100units/ml suspension for injection 3ml pre-filled pens (Eli Lilly and Company Ltd) | 2182441000033113 | 3260811000001108 | Insulin isophane human |
| Insuman Basal 100units/ml suspension for injection 3ml pre-filled SoloStar pens (Sanofi) | 6389641000033111 | 19354411000001104 | Insulin isophane human |
| Humulin I KwikPen 100units/ml suspension for injection 3ml pre-filled pens (Eli Lilly and Company Ltd) | 5911541000033119 | 17608511000001103 | Insulin isophane human |
| Humalog 100units/ml solution for injection 1.5ml cartridges (Eli Lilly and Company Ltd) | 720541000033111 | 3282711000001104 | Insulin lispro |
| Insulin lispro 100units/ml solution for injection 1.5ml cartridges | 757641000033112 | 36047511000001102 | Insulin lispro |
| Humalog 100units/ml solution for injection 10ml vials (Eli Lilly and Company Ltd) | 723241000033118 | 3280111000001104 | Insulin lispro |
| Insulin lispro 100units/ml solution for injection 10ml vials | 762441000033118 | 36047611000001103 | Insulin lispro |
| Insulin lispro Sanofi 100units/ml solution for injection 10ml vials (Sanofi) | 12600741000033116 | 35776411000001102 | Insulin lispro |
| Lyumjev 100units/ml solution for injection 10ml vials (Eli Lilly and Company Ltd) | 13605141000033117 | 38456911000001105 | Insulin lispro |
| Humalog 100units/ml solution for injection 3ml cartridges (Eli Lilly and Company Ltd) | 1736841000033114 | 3284311000001105 | Insulin lispro |

| | | | |
|---|---|---|---|
| Lyumjev 100units/ml solution for injection 3ml cartridges (Eli Lilly and Company Ltd) | 13605041000033116 | 38456111000001107 | Insulin lispro |
| Insulin lispro Sanofi 100units/ml solution for injection 3ml cartridges (Sanofi) | 12600841000033114 | 35776511000001103 | Insulin lispro |
| Insulin lispro 100units/ml solution for injection 3ml cartridges | 1739941000033110 | 36047711000001107 | Insulin lispro |
| Humalog KwikPen 100units/ml solution for injection 3ml pre-filled pens (Eli Lilly and Company Ltd) | 4608541000033115 | 13877811000001105 | Insulin lispro |
| Humalog Pen 100units/ml solution for injection 3ml pre-filled pens (Sigma Pharmaceuticals Plc) | 2182341000033119 | 14249011000001103 | Insulin lispro |
| Insulin lispro 100units/ml solution for injection 3ml pre-filled disposable devices | 2182241000033112 | 36047811000001104 | Insulin lispro |
| Insulin lispro Sanofi 100units/ml solution for injection 3ml pre-filled pens (Sanofi) | 12600941000033118 | 35776811000001100 | Insulin lispro |
| Humalog Junior KwikPen 100units/ml solution for injection 3ml pre-filled pens (Eli Lilly and Company Ltd) | 12486241000033111 | 35216811000001101 | Insulin lispro |
| Lyumjev KwikPen 100units/ml solution for injection 3ml pre-filled pens (Eli Lilly and Company Ltd) | 13604841000033112 | 38465811000001104 | Insulin lispro |
| Lyumjev Junior KwikPen 100units/ml solution for injection 3ml pre-filled pens (Eli Lilly and Company Ltd) | 13604741000033119 | 38479611000001109 | Insulin lispro |
| Hypurin Bovine Protamine Zinc 100units/ml suspension for injection 10ml vials (Wockhardt UK Ltd) | 736141000033115 | 3269911000001109 | Insulin protamine zinc bovine |
| Insulin protamine zinc bovine 100units/ml suspension for injection 10ml vials | 1119741000033114 | 36047911000001109 | Insulin protamine zinc bovine |
| Hypurin Bovine Neutral 100units/ml solution for injection 10ml vials (Wockhardt UK Ltd) | 736041000033119 | 3271311000001101 | Insulin soluble bovine |
| Human Actrapid Penfill 100units/ml solution for injection 1.5ml cartridges (Novo Nordisk Ltd) | 721041000033110 | 3309511000001105 | Insulin soluble human |
| Actrapid 100units/ml solution for injection 10ml vials (Novo Nordisk Ltd) | 2952841000033116 | 3312111000001108 | Insulin soluble human |
| Velosulin 100units/ml solution for injection 10ml vials (Novo Nordisk Ltd) | 2954641000033110 | 3312411000001103 | Insulin soluble human |
| Humulin S 100units/ml solution for injection 10ml vials (Eli Lilly and Company Ltd) | 724341000033114 | 3312611000001100 | Insulin soluble human |
| Actrapid Penfill 100units/ml solution for injection 3ml cartridges (Novo Nordisk Ltd) | 2953041000033119 | 3310711000001105 | Insulin soluble human |
| Humulin S 100units/ml solution for injection 3ml cartridges (Eli Lilly and Company Ltd) | 1616041000033115 | 3311311000001101 | Insulin soluble human |
| Insuman Rapid 100units/ml solution for injection 3ml cartridges (Sanofi) | 2034641000033119 | 3311611000001106 | Insulin soluble human |
| Actrapid NovoLet 100units/ml solution for injection (Novo Nordisk Ltd) | 2952941000033112 | 3288511000001106 | Insulin soluble human |
| HumaJect S Pen 100units/ml solution for injection (Eli Lilly and Company Ltd) | 727041000033119 | 3290411000001108 | Insulin soluble human |
| Insuman Rapid 100units/ml solution for injection 3ml pre-filled OptiSet pens (Sanofi) | 2266941000033119 | 3291711000001105 | Insulin soluble human |
| Insuman Rapid 100units/ml solution for injection 5ml vials (Aventis Pharma) | 2034441000033116 | 3294911000001105 | Insulin soluble human |
| Hypurin Porcine Neutral 100units/ml solution for injection 10ml vials (Wockhardt UK Ltd) | 735441000033119 | 3285011000001106 | Insulin soluble porcine |
| Pork Actrapid 100units/ml solution for injection 10ml vials (Novo Nordisk Ltd) | 1749441000033116 | 3285611000001104 | Insulin soluble porcine |
| Insulin soluble porcine 100units/ml solution for injection 10ml vials | 12624241000033111 | 36048711000001108 | Insulin soluble porcine |
| Ultratard 100units/ml suspension for injection 10ml vials (Novo Nordisk Ltd) | 2954841000033111 | 3282311000001103 | Insulin zinc suspension crystalline human |

| | | | |
|---|---|---|---|
| Humulin Zn 100units/ml suspension for injection 10ml vials (Eli Lilly and Company Ltd) | 724541000033119 | 3281811000001103 | Insulin zinc suspension crystalline human |
| Humulin Lente 100units/ml suspension for injection 10ml vials (Eli Lilly and Company Ltd) | 725841000033118 | 3284911000001106 | Insulin zinc suspension mixed human |
| Monotard 100units/ml suspension for injection 10ml vials (Novo Nordisk Ltd) | 2954741000033118 | 3284111000001108 | Insulin zinc suspension mixed human |
| Suliqua 100units/ml / 33micrograms/ml solution for injection 3ml pre-filled SoloStar pens (Sanofi) | 12902541000033110 | 36620611000001107 | Insulin glargine/ Lixisenatide |
| Insulin glargine 100units/ml / Lixisenatide 33micrograms/ml solution for injection 3ml pre-filled disposable devices | 12902741000033119 | 36630511000001105 | Insulin glargine/ Lixisenatide |
| Insulin glargine 100units/ml / Lixisenatide 50micrograms/ml solution for injection 3ml pre-filled disposable devices | 12902841000033112 | 36630611000001109 | Insulin glargine/ Lixisenatide |
| Suliqua 100units/ml / 50micrograms/ml solution for injection 3ml pre-filled SoloStar pens (Sanofi) | 12902641000033111 | 36618311000001109 | Insulin glargine/ Lixisenatide |
| Toujeo 300units/ml solution for injection 3ml pre-filled DoubleStar pens (Sanofi) | 12998841000033112 | 36911311000001101 | Insulin glargine |
| Insulin glargine 300units/ml solution for injection 3ml pre-filled disposable devices | 12998741000033119 | 36931811000001104 | Insulin glargine |
| Lantus 100units/ml solution for injection 3ml cartridges (Sanofi) | 2780841000033118 | 3284211000001102 | Insulin glargine |
| Lantus 100units/ml solution for injection 3ml OptiClik cartridges (Sanofi) | 3869341000033114 | 10250211000001101 | Insulin glargine |
| Abasaglar 100units/ml solution for injection 3ml cartridges (Eli Lilly and Company Ltd) | 10600641000033118 | 30172211000001102 | Insulin glargine |
| Insulin glargine 100units/ml solution for injection 3ml cartridges | 2798941000033111 | 38896911000001108 | Insulin glargine |
| Lantus 100units/ml solution for injection 3ml pre-filled OptiSet pens (Sanofi) | 2780941000033114 | 3283211000001100 | Insulin glargine |
| Lantus 100units/ml solution for injection 3ml pre-filled SoloStar pens (Sanofi) | 4258541000033111 | 11933011000001106 | Insulin glargine |
| Insulin glargine 100units/ml solution for injection 3ml pre-filled disposable devices | 2799041000033119 | 38897011000001107 | Insulin glargine |
| Abasaglar KwikPen 100units/ml solution for injection 3ml pre-filled pens (Eli Lilly and Company Ltd) | 10600741000033110 | 30171811000001105 | Insulin glargine |
| Semglee 100units/ml solution for injection 3ml pre-filled pens (Mylan) | 12686641000033112 | 36082811000001107 | Insulin glargine |
| Acarbose 50mg tablets | 11541000033110 | 326037007 | Acarbose |
| Glucobay 50mg tablets (Bayer Plc) | 645341000033115 | 781411000001104 | Acarbose |
| Glucobay 100mg tablets (Bayer Plc) | 645241000033113 | 444311000001106 | Acarbose |
| Acarbose 100mg tablets | 11441000033114 | 326038002 | Acarbose |
| Eperzan 30mg powder and solvent for solution for injection pre-filled pens (GlaxoSmithKline UK Ltd) | 10952341000033119 | 31014311000001106 | Albiglutide |
| Albiglutide 30mg powder and solvent for solution for injection pre-filled disposable devices | 10951041000033114 | 31015711000001109 | Albiglutide |
| Albiglutide 50mg powder and solvent for solution for injection pre-filled disposable devices | 10951141000033113 | 31015811000001101 | Albiglutide |
| Eperzan 50mg powder and solvent for solution for injection pre-filled pens (GlaxoSmithKline UK Ltd) | 10953041000033113 | 31014611000001101 | Albiglutide |
| Vipdomet 12.5mg/1000mg tablets (Takeda UK Ltd) | 8959941000033110 | 23632611000001103 | Alogliptin benzoate/ Metformin hydrochloride |
| Alogliptin 12.5mg / Metformin 1g tablets | 8959841000033119 | 23637211000001102 | Alogliptin benzoate/ |

| | | | |
|---|---|---|---|
| | | | Metformin hydrochloride |
| Alogliptin 12.5mg tablets | 8959341000033111 | 23637311000001105 | Alogliptin benzoate |
| Vipidia 12.5mg tablets (Takeda UK Ltd) | 8959641000033115 | 23636011000001109 | Alogliptin benzoate |
| Vipidia 25mg tablets (Takeda UK Ltd) | 8959741000033112 | 23636311000001107 | Alogliptin benzoate |
| Alogliptin 25mg tablets | 8959441000033117 | 23637411000001103 | Alogliptin benzoate |
| Alogliptin 6.25mg tablets | 8959241000033118 | 23637511000001104 | Alogliptin benzoate |
| Vipidia 6.25mg tablets (Takeda UK Ltd) | 8959541000033116 | 23634111000001103 | Alogliptin benzoate |
| Canagliflozin 100mg tablets | 9110341000033112 | 703682001 | Canagliflozin hemihydrate |
| Invokana 100mg tablets (Napp Pharmaceuticals Ltd) | 9110541000033117 | 24088611000001101 | Canagliflozin hemihydrate |
| Canagliflozin 300mg tablets | 9110441000033118 | 24104511000001103 | Canagliflozin hemihydrate |
| Invokana 300mg tablets (Napp Pharmaceuticals Ltd) | 9110641000033116 | 24088311000001106 | Canagliflozin hemihydrate |
| Canagliflozin 50mg / Metformin 1g tablets | 9851541000033114 | 28049211000001101 | Canagliflozin hemihydrate/ Metformin hydrochloride |
| Vokanamet 50mg/1000mg tablets (Napp Pharmaceuticals Ltd) | 9851841000033111 | 28024411000001103 | Canagliflozin hemihydrate/ Metformin hydrochloride |
| Vokanamet 50mg/850mg tablets (Napp Pharmaceuticals Ltd) | 9851741000033118 | 28022511000001109 | Canagliflozin hemihydrate/ Metformin hydrochloride |
| Canagliflozin 50mg / Metformin 850mg tablets | 9851641000033110 | 28049311000001109 | Canagliflozin hemihydrate/ Metformin hydrochloride |
| Chlorpropamide 100mg tablets | 249441000033114 | 325213005 | Chlorpropamide |
| Chlorpropamide 250mg tablets | 249541000033110 | 325214004 | Chlorpropamide |
| Forxiga 5mg tablets (AstraZeneca UK Ltd) | 8199541000033113 | 21609511000001105 | Dapagliflozin propanediol monohydrate |
| Dapagliflozin 5mg tablets | 8199341000033118 | 703679006 | Dapagliflozin propanediol monohydrate |
| Dapagliflozin 10mg tablets | 8199441000033112 | 703680009 | Dapagliflozin propanediol monohydrate |
| Forxiga 10mg tablets (AstraZeneca UK Ltd) | 8199641000033114 | 21609811000001108 | Dapagliflozin propanediol monohydrate |
| Xigduo 5mg/1000mg tablets (AstraZeneca UK Ltd) | 9106241000033119 | 24018511000001102 | Dapagliflozin propanediol monohydrate/ Metformin hydrochloride |
| Dapagliflozin 5mg / Metformin 1g tablets | 9106041000033110 | 24054611000001100 | Dapagliflozin propanediol monohydrate/ Metformin hydrochloride |

| | | | |
|---|---|---|---|
| Dapagliflozin 5mg / Metformin 850mg tablets | 9106141000033114 | 24054711000001109 | Dapagliflozin propanediol monohydrate/ Metformin hydrochloride |
| Xigduo 5mg/850mg tablets (AstraZeneca UK Ltd) | 9106341000033112 | 24018111000001106 | Dapagliflozin propanediol monohydrate/ Metformin hydrochloride |
| Saxagliptin 5mg / Dapagliflozin 10mg tablets | 11898041000033111 | 33745311000001109 | Dapagliflozin propanediol monohydrate/ Saxagliptin hydrochloride |
| Qtern 5mg/10mg tablets (AstraZeneca UK Ltd) | 11898141000033110 | 33682311000001103 | Dapagliflozin propanediol monohydrate/ Saxagliptin hydrochloride |
| Dulaglutide 0.75mg/0.5ml solution for injection pre-filled disposable devices | 10207841000033118 | 28789611000001108 | Dulaglutide |
| Trulicity 0.75mg/0.5ml solution for injection pre-filled pens (Eli Lilly and Company Ltd) | 10208041000033112 | 28461011000001109 | Dulaglutide |
| Trulicity 1.5mg/0.5ml solution for injection pre-filled pens (Eli Lilly and Company Ltd) | 10208141000033111 | 28462311000001103 | Dulaglutide |
| Dulaglutide 1.5mg/0.5ml solution for injection pre-filled disposable devices | 10207941000033114 | 28789711000001104 | Dulaglutide |
| Jardiance 10mg tablets (Boehringer Ingelheim Ltd) | 9337141000033114 | 25238811000001107 | Empagliflozin |
| Empagliflozin 10mg tablets | 9336641000033119 | 25290511000001101 | Empagliflozin |
| Empagliflozin 25mg tablets | 9336841000033118 | 25290611000001102 | Empagliflozin |
| Jardiance 25mg tablets (Boehringer Ingelheim Ltd) | 9337241000033119 | 25239711000001108 | Empagliflozin |
| Empagliflozin 12.5mg / Metformin 1g tablets | 10614441000033112 | 30318111000001109 | Empagliflozin/ Metformin hydrochloride |
| Synjardy 12.5mg/1000mg tablets (Boehringer Ingelheim Ltd) | 10614841000033110 | 30175711000001100 | Empagliflozin/ Metformin hydrochloride |
| Synjardy 12.5mg/850mg tablets (Boehringer Ingelheim Ltd) | 10614741000033117 | 30175011000001102 | Empagliflozin/ Metformin hydrochloride |
| Empagliflozin 12.5mg / Metformin 850mg tablets | 10614341000033118 | 30318211000001103 | Empagliflozin/ Metformin hydrochloride |
| Empagliflozin 5mg / Metformin 1g tablets | 10614241000033111 | 30318311000001106 | Empagliflozin/ Metformin hydrochloride |
| Synjardy 5mg/1000mg tablets (Boehringer Ingelheim Ltd) | 10614641000033114 | 30174111000001100 | Empagliflozin/ Metformin hydrochloride |
| Synjardy 5mg/850mg tablets (Boehringer Ingelheim Ltd) | 10614541000033113 | 30173411000001107 | Empagliflozin/ Metformin hydrochloride |
| Empagliflozin 5mg / Metformin 850mg tablets | 10614141000033116 | 30318411000001104 | Empagliflozin/ Metformin hydrochloride |
| Ertugliflozin 15mg tablets | 12881041000033118 | 36529411000001105 | Ertugliflozin L-pyroglutamic acid |
| Steglatro 15mg tablets (Merck Sharp & Dohme Ltd) | 12881241000033114 | 36515111000001107 | Ertugliflozin L-pyroglutamic acid |

| | | | |
|---|---|---|---|
| Ertugliflozin 5mg tablets | 12880941000033111 | 36529511000001109 | Ertugliflozin L-pyroglutamic acid |
| Steglatro 5mg tablets (Merck Sharp & Dohme Ltd) | 12881141000033119 | 36514811000001101 | Ertugliflozin L-pyroglutamic acid |
| Exenatide 10micrograms/0.04ml solution for injection 2.4ml pre-filled disposable devices | 4149441000033117 | 11494111000001102 | Exenatide |
| Byetta 10micrograms/0.04ml solution for injection 2.4ml pre-filled pens (AstraZeneca UK Ltd) | 4149641000033115 | 11494611000001105 | Exenatide |
| Byetta 5micrograms/0.02ml solution for injection 1.2ml pre-filled pens (AstraZeneca UK Ltd) | 4149541000033116 | 11494811000001109 | Exenatide |
| Exenatide 5micrograms/0.02ml solution for injection 1.2ml pre-filled disposable devices | 4149341000033111 | 11494211000001108 | Exenatide |
| Exenatide 2mg powder and solvent for prolonged-release suspension for injection vials | 6388241000033117 | 19275411000001107 | Exenatide |
| Bydureon 2mg powder and solvent for prolonged-release suspension for injection vials (AstraZeneca UK Ltd) | 6388341000033110 | 19274811000001108 | Exenatide |
| Bydureon 2mg powder and solvent for prolonged-release suspension for injection pre-filled pens (AstraZeneca UK Ltd) | 13427441000033112 | 28426011000001108 | Exenatide |
| Exenatide 2mg powder and solvent for prolonged-release suspension for injection pre-filled disposable devices | 13427341000033118 | 28440211000001102 | Exenatide |
| Exenatide 2mg/0.85ml prolonged-release suspension for injection pre-filled disposable devices | 13427541000033113 | 38082811000001104 | Exenatide |
| Bydureon BCise 2mg/0.85ml prolonged-release suspension for injection pre-filled pens (AstraZeneca UK Ltd) | 13427641000033114 | 38060511000001101 | Exenatide |
| Glibenclamide 2.5mg tablets | 644541000033111 | 325218001 | Glibenclamide |
| Euglucon 2.5mg tablets (Aventis Pharma) | 557241000033113 | 233411000001102 | Glibenclamide |
| Semi-Daonil 2.5mg tablets (Sanofi) | 1278041000033114 | 140711000001103 | Glibenclamide |
| Diabetamide 2.5mg tablets (Ashbourne Pharmaceuticals Ltd) | 462041000033119 | 13511000001103 | Glibenclamide |
| Daonil 5mg tablets (Sanofi) | 406841000033114 | 234011000001108 | Glibenclamide |
| Diabetamide 5mg tablets (Ashbourne Pharmaceuticals Ltd) | 462141000033115 | 287811000001101 | Glibenclamide |
| Euglucon 5mg tablets (Sanofi) | 557341000033115 | 322511000001103 | Glibenclamide |
| Glibenclamide 5mg tablets | 644641000033112 | 325219009 | Glibenclamide |
| Glibenclamide 5mg/5ml oral solution | 5968741000033115 | 8523711000001107 | Glibenclamide |
| Glibenclamide 5mg/5ml oral suspension | 5968841000033113 | 8523811000001104 | Glibenclamide |
| Amglidia 0.6mg/ml oral suspension with 1ml oral syringe (Amring Pharmaceuticals Ltd) | 13122741000033116 | 37337011000001106 | Glibenclamide |
| Amglidia 0.6mg/ml oral suspension with 5ml oral syringe (Amring Pharmaceuticals Ltd) | 13122841000033114 | 37337211000001101 | Glibenclamide |
| Glibenclamide 600micrograms/ml oral suspension sugar free | 13122541000033112 | 37405911000001104 | Glibenclamide |
| Glibenclamide 6mg/ml oral suspension sugar free | 13122641000033113 | 37406011000001107 | Glibenclamide |
| Amglidia 6mg/ml oral suspension with 1ml oral syringe (Amring Pharmaceuticals Ltd) | 13122941000033118 | 37337511000001103 | Glibenclamide |
| Amglidia 6mg/ml oral suspension with 5ml oral syringe (Amring Pharmaceuticals Ltd) | 13123041000033111 | 37337711000001108 | Glibenclamide |
| Gliclazide 80mg tablets | 646441000033117 | 325242002 | Gliclazide |
| Diaglyk 80mg tablets (Ashbourne Pharmaceuticals Ltd) | 1602941000033117 | 49111000001103 | Gliclazide |
| Diamicron 80mg tablets (Servier Laboratories Ltd) | 463541000033118 | 745911000001103 | Gliclazide |
| Gliclazide 40mg/5ml oral suspension | 5869241000033114 | 8524211000001102 | Gliclazide |
| Gliclazide 80mg/5ml oral suspension | 3953041000033113 | 8524311000001105 | Gliclazide |

| | | | |
|---|---|---|---|
| Gliclazide 40mg tablets | 5709741000033116 | 16702011000001102 | Gliclazide |
| Zicron 40mg tablets (Bristol Laboratories Ltd) | 5709841000033114 | 16677511000001108 | Gliclazide |
| Laaglyda MR 60mg tablets (Consilient Health Ltd) | 8298241000033118 | 22225011000001109 | Gliclazide |
| Gliclazide 60mg modified-release tablets | 8298141000033113 | 22226111000001104 | Gliclazide |
| Vamju 60mg modified-release tablets (Advanz Pharma) | 10043441000033110 | 28420911000001109 | Gliclazide |
| Lamzarin 60mg modified-release tablets (Key Pharmaceuticals Ltd) | 13345341000033117 | 38018511000001102 | Gliclazide |
| Bilxona 60mg modified-release tablets (Accord Healthcare Ltd) | 10701741000033113 | 37618711000001109 | Gliclazide |
| Glydex 160mg tablets (Medreich Plc) | 12999041000033113 | 36910511000001100 | Gliclazide |
| Gliclazide 160mg tablets | 12998941000033116 | 36914911000001102 | Gliclazide |
| Diamicron 30mg MR tablets (Servier Laboratories Ltd) | 2289941000033113 | 3661311000001107 | Gliclazide |
| Nazdol MR 30mg tablets (Consilient Health Ltd) | 4941241000033119 | 13456711000001106 | Gliclazide |
| Edicil MR 30mg tablets (Teva UK Ltd) | 5815341000033115 | 15334511000001109 | Gliclazide |
| Vitile XL 30mg tablets (Actavis UK Ltd) | 6135341000033117 | 18678711000001108 | Gliclazide |
| Dacadis MR 30mg tablets (Mylan) | 5378141000033117 | 16536211000001102 | Gliclazide |
| Gliclazide 30mg modified-release tablets | 2289441000033115 | 38896211000001104 | Gliclazide |
| Lamzarin 30mg modified-release tablets (Key Pharmaceuticals Ltd) | 13345241000033110 | 38018211000001100 | Gliclazide |
| Zicron PR 30mg tablets (Bristol Laboratories Ltd) | 11918941000033116 | 33766211000001100 | Gliclazide |
| Bilxona 30mg modified-release tablets (Accord Healthcare Ltd) | 10701641000033116 | 30982411000001107 | Gliclazide |
| Vamju 30mg modified-release tablets (Advanz Pharma) | 10043341000033116 | 28420711000001107 | Gliclazide |
| Glimepiride 2mg tablets | 645841000033112 | 325259007 | Glimepiride |
| Amaryl 2mg tablets (Zentiva) | 58841000033113 | 938211000001105 | Glimepiride |
| Niddaryl 2mg tablets (Dee Pharmaceuticals Ltd) | 4522141000033112 | 13626511000001104 | Glimepiride |
| Niddaryl 1mg tablets (Dee Pharmaceuticals Ltd) | 4522041000033113 | 13626011000001107 | Glimepiride |
| Amaryl 1mg tablets (Zentiva) | 59541000033116 | 228111000001101 | Glimepiride |
| Glimepiride 1mg tablets | 645941000033116 | 325261003 | Glimepiride |
| Glimepiride 3mg tablets | 646041000033114 | 325262005 | Glimepiride |
| Amaryl 3mg tablets (Zentiva) | 59641000033115 | 164511000001108 | Glimepiride |
| Niddaryl 3mg tablets (Dee Pharmaceuticals Ltd) | 4522241000033117 | 13626911000001106 | Glimepiride |
| Niddaryl 4mg tablets (Dee Pharmaceuticals Ltd) | 4522341000033110 | 13627211000001100 | Glimepiride |
| Amaryl 4mg tablets (Zentiva) | 59741000033112 | 259611000001101 | Glimepiride |
| Glimepiride 4mg tablets | 646141000033113 | 325263000 | Glimepiride |
| Glimepiride 6mg/5ml oral suspension | 8125241000033119 | 21366911000001103 | Glimepiride |
| Minodiab 5mg tablets (Pfizer Ltd) | 920241000033117 | 652411000001107 | Glipizide |
| Glibenese 5mg tablets (Pfizer Ltd) | 644741000033115 | 515111000001108 | Glipizide |
| Glipizide 5mg tablets | 644841000033113 | 325243007 | Glipizide |
| Glipizide 2.5mg tablets | 646541000033116 | 325248003 | Glipizide |
| Minodiab 2.5mg tablets (Pfizer Ltd) | 920141000033112 | 808711000001104 | Glipizide |
| Guar gum 5g granules sachets sugar free | 655141000033116 | 325287003 | Guar gum |
| Guarem 5g granules sachets (Shire Pharmaceuticals Ltd) | 654941000033115 | 4667411000001106 | Guar gum |

| | | | |
|---|---|---|---|
| Linagliptin 5mg tablets | 6444041000033117 | 19525211000001103 | Linagliptin |
| Trajenta 5mg tablets (Boehringer Ingelheim Ltd) | 6444141000033118 | 19492811000001108 | Linagliptin |
| Jentadueto 2.5mg/1000mg tablets (Boehringer Ingelheim Ltd) | 8115941000033116 | 21208511000001102 | Linagliptin/ Metformin hydrochloride |
| Linagliptin 2.5mg / Metformin 1g tablets | 8115741000033119 | 21245011000001106 | Linagliptin/ Metformin hydrochloride |
| Linagliptin 2.5mg / Metformin 850mg tablets | 8115841000033112 | 21245111000001107 | Linagliptin/ Metformin hydrochloride |
| Jentadueto 2.5mg/850mg tablets (Boehringer Ingelheim Ltd) | 8116041000033114 | 21208211000001100 | Linagliptin/ Metformin hydrochloride |
| Empagliflozin 10mg / Linagliptin 5mg tablets | 13116741000033112 | 37280311000001109 | Empagliflozin/ Linagliptin |
| Glyxambi 10mg/5mg tablets (Boehringer Ingelheim Ltd) | 13116941000033110 | 37225211000001100 | Empagliflozin/ Linagliptin |
| Glyxambi 25mg/5mg tablets (Boehringer Ingelheim Ltd) | 13117041000033111 | 37225511000001102 | Empagliflozin/ Linagliptin |
| Empagliflozin 25mg / Linagliptin 5mg tablets | 13116841000033119 | 37280511000001103 | Empagliflozin/ Linagliptin |
| Victoza 6mg/ml solution for injection 3ml pre-filled pens (Novo Nordisk Ltd) | 5131341000033117 | 15858611000001101 | Liraglutide |
| Liraglutide 6mg/ml solution for injection 3ml pre-filled disposable devices | 5131241000033110 | 15859111000001102 | Liraglutide |
| Saxenda 6mg/ml solution for injection 3ml pre-filled pens (Novo Nordisk Ltd) | 11919041000033113 | 33747711000001102 | Liraglutide |
| Lixisenatide 10micrograms/0.2ml solution for injection 3ml pre-filled disposable devices | 8267341000033110 | 21994611000001106 | Lixisenatide |
| Lyxumia 10micrograms/0.2ml solution for injection 3ml pre-filled pens (Sanofi) | 8267541000033115 | 21941511000001102 | Lixisenatide |
| Lixisenatide 20micrograms/0.2ml solution for injection 3ml pre-filled disposable devices | 8267441000033116 | 21994811000001105 | Lixisenatide |
| Lyxumia 20micrograms/0.2ml solution for injection 3ml pre-filled pens (Sanofi) | 8267641000033119 | 21941011000001105 | Lixisenatide |
| Metformin 500mg tablets | 896941000033112 | 325278007 | Metformin hydrochloride |
| Glucophage 500mg tablets (Merck Serono Ltd) | 644941000033117 | 363211000001102 | Metformin hydrochloride |
| Glucophage 850mg tablets (Merck Serono Ltd) | 645041000033117 | 365111000001109 | Metformin hydrochloride |
| Metformin 850mg tablets | 897041000033113 | 325279004 | Metformin hydrochloride |
| Rosiglitazone 1mg / Metformin 500mg tablets | 3191341000033115 | 409121008 | Metformin hydrochloride/ Rosiglitazone maleate |
| Avandamet 1mg/500mg tablets (GlaxoSmithKline UK Ltd) | 2995141000033110 | 5302911000001102 | Metformin hydrochloride/ Rosiglitazone maleate |
| Avandamet 2mg/500mg tablets (GlaxoSmithKline UK Ltd) | 2995241000033115 | 5303611000001103 | Metformin hydrochloride/ Rosiglitazone maleate |
| Rosiglitazone 2mg / Metformin 500mg tablets | 3191441000033114 | 409122001 | Metformin hydrochloride/ Rosiglitazone maleate |
| Rosiglitazone 2mg / Metformin 1g tablets | 3200541000033116 | 409124000 | Metformin hydrochloride/ |

| | | | |
|---|---|---|---|
| | | | Rosiglitazone maleate |
| Avandamet 2mg/1000mg tablets (GlaxoSmithKline UK Ltd) | 3200741000033112 | 8176311000001105 | Metformin hydrochloride/ Rosiglitazone maleate |
| Avandamet 4mg/1000mg tablets (GlaxoSmithKline UK Ltd) | 3200841000033119 | 8174611000001109 | Metformin hydrochloride/ Rosiglitazone maleate |
| Rosiglitazone 4mg / Metformin 1g tablets | 3200641000033115 | 409125004 | Metformin hydrochloride/ Rosiglitazone maleate |
| Metformin 500mg/5ml oral solution | 5997041000033115 | 409197000 | Metformin hydrochloride |
| Pioglitazone 15mg / Metformin 850mg tablets | 3983941000033115 | 419873003 | Metformin hydrochloride/ Pioglitazone hydrochloride |
| Competact 15mg/850mg tablets (Takeda UK Ltd) | 3984241000033114 | 10922311000001108 | Metformin hydrochloride/ Pioglitazone hydrochloride |
| Metformin 500mg/5ml oral suspension | 2620241000033119 | 8664411000001102 | Metformin hydrochloride |
| Metformin 500mg/5ml oral solution sugar free | 3982341000033110 | 10750111000001100 | Metformin hydrochloride |
| Metsol 500mg/5ml oral solution (Kappin Ltd) | 3982441000033116 | 10741611000001100 | Metformin hydrochloride |
| Vildagliptin 50mg / Metformin 1g tablets | 4452441000033113 | 13413011000001108 | Metformin hydrochloride/ Vildagliptin |
| Eucreas 50mg/1000mg tablets (Novartis Pharmaceuticals UK Ltd) | 4452541000033114 | 13412311000001101 | Metformin hydrochloride/ Vildagliptin |
| Eucreas 50mg/850mg tablets (Novartis Pharmaceuticals UK Ltd) | 4452641000033110 | 13412611000001106 | Metformin hydrochloride/ Vildagliptin |
| Vildagliptin 50mg / Metformin 850mg tablets | 4452341000033119 | 13413111000001109 | Metformin hydrochloride/ Vildagliptin |
| Metformin 1g oral powder sachets sugar free | 5007741000033111 | 15411211000001106 | Metformin hydrochloride |
| Glucophage 1000mg oral powder sachets (Merck Serono Ltd) | 5007941000033114 | 15374311000001101 | Metformin hydrochloride |
| Metformin 500mg oral powder sachets sugar free | 5007641000033119 | 15411311000001103 | Metformin hydrochloride |
| Glucophage 500mg oral powder sachets (Merck Serono Ltd) | 5007841000033118 | 15373711000001104 | Metformin hydrochloride |
| Janumet 50mg/1000mg tablets (Merck Sharp & Dohme Ltd) | 5132241000033116 | 17060511000001108 | Metformin hydrochloride/ Sitagliptin phosphate |
| Metformin 1g / Sitagliptin 50mg tablets | 5576041000033115 | 17071811000001108 | Metformin hydrochloride/ Sitagliptin phosphate |
| Saxagliptin 2.5mg / Metformin 1g tablets | 8242641000033113 | 21711411000001100 | Metformin hydrochloride/ Saxagliptin hydrochloride |

| | | | |
|---|---|---|---|
| Komboglyze 2.5mg/1000mg tablets (AstraZeneca UK Ltd) | 8242841000033114 | 21705611000001102 | Metformin hydrochloride/ Saxagliptin hydrochloride |
| Komboglyze 2.5mg/850mg tablets (AstraZeneca UK Ltd) | 8242741000033116 | 21705311000001107 | Metformin hydrochloride/ Saxagliptin hydrochloride |
| Saxagliptin 2.5mg / Metformin 850mg tablets | 8242541000033112 | 21711511000001101 | Metformin hydrochloride/ Saxagliptin hydrochloride |
| Metformin 1g/5ml oral solution sugar free | 11781341000033117 | 33550811000001105 | Metformin hydrochloride |
| Metformin 850mg/5ml oral solution sugar free | 11781441000033111 | 33550911000001100 | Metformin hydrochloride |
| Bolamyn SR 1000mg tablets (Teva UK Ltd) | 8348941000033113 | 22308811000001109 | Metformin hydrochloride |
| Metabet SR 1000mg tablets (Morningside Healthcare Ltd) | 6279741000033117 | 18885611000001108 | Metformin hydrochloride |
| Glucophage SR 1000mg tablets (Merck Serono Ltd) | 4945341000033110 | 15367811000001105 | Metformin hydrochloride |
| Metformin 1g modified-release tablets | 4945241000033117 | 38893711000001104 | Metformin hydrochloride |
| Yaltormin SR 1000mg tablets (Wockhardt UK Ltd) | 12593341000033117 | 35548311000001108 | Metformin hydrochloride |
| Meijumet 1000mg modified-release tablets (Medreich Plc) | 12326741000033119 | 34553311000001105 | Metformin hydrochloride |
| Sukkarto SR 1000mg tablets (Morningside Healthcare Ltd) | 9230841000033114 | 24568211000001104 | Metformin hydrochloride |
| Glucient SR 1000mg tablets (Consilient Health Ltd) | 10598941000033117 | 30012311000001107 | Metformin hydrochloride |
| Glucophage SR 750mg tablets (Merck Serono Ltd) | 4549241000033113 | 13748611000001107 | Metformin hydrochloride |
| Glucient SR 750mg tablets (Consilient Health Ltd) | 10598841000033113 | 30012111000001105 | Metformin hydrochloride |
| Meijumet 750mg modified-release tablets (Medreich Plc) | 12326641000033111 | 34552811000001109 | Metformin hydrochloride |
| Yaltormin SR 750mg tablets (Wockhardt UK Ltd) | 12593241000033110 | 35548011000001105 | Metformin hydrochloride |
| Metformin 750mg modified-release tablets | 4549141000033118 | 38893811000001107 | Metformin hydrochloride |
| Sukkarto SR 750mg tablets (Morningside Healthcare Ltd) | 13429041000033112 | 38238111000001109 | Metformin hydrochloride |
| Glucophage SR 500mg tablets (Merck Serono Ltd) | 3228241000033112 | 8990711000001100 | Metformin hydrochloride |
| Bolamyn SR 500mg tablets (Teva UK Ltd) | 4957741000033118 | 14183911000001108 | Metformin hydrochloride |
| Metabet SR 500mg tablets (Morningside Healthcare Ltd) | 6029841000033115 | 18141911000001101 | Metformin hydrochloride |
| Glucient SR 500mg tablets (Consilient Health Ltd) | 6391041000033117 | 19308111000001109 | Metformin hydrochloride |
| Diagemet XL 500mg tablets (Genus Pharmaceuticals Ltd) | 7874941000033110 | 20552511000001105 | Metformin hydrochloride |
| Metformin 500mg modified-release tablets | 3228141000033117 | 39113511000001101 | Metformin hydrochloride |
| Glucorex SR 500mg tablets (GlucoRx Ltd) | 13606641000033119 | 38749111000001108 | Metformin hydrochloride |
| Sukkarto SR 500mg tablets (Morningside Healthcare Ltd) | 9230641000033113 | 24568411000001100 | Metformin hydrochloride |
| Yaltormin SR 500mg tablets (Wockhardt UK Ltd) | 12593141000033115 | 35547511000001101 | Metformin hydrochloride |

| | | | |
|---|---|---|---|
| Meijumet 500mg modified-release tablets (Medreich Plc) | 12326541000033110 | 34552411000001107 | Metformin hydrochloride |
| Metuxtan SR 500mg tablets (Accord Healthcare Ltd) | 12664441000033115 | 35849011000001104 | Metformin hydrochloride |
| Starlix 180mg tablets (Novartis Pharmaceuticals UK Ltd) | 2288741000033118 | 3883511000001107 | Nateglinide |
| Nateglinide 180mg tablets | 2288441000033113 | 134608004 | Nateglinide |
| Nateglinide 120mg tablets | 2288341000033119 | 134609007 | Nateglinide |
| Starlix 120mg tablets (Novartis Pharmaceuticals UK Ltd) | 2288641000033110 | 3651211000001105 | Nateglinide |
| Starlix 60mg tablets (Novartis Pharmaceuticals UK Ltd) | 2288541000033114 | 3650611000001102 | Nateglinide |
| Nateglinide 60mg tablets | 2288241000033112 | 134610002 | Nateglinide |
| Actos 30mg tablets (Takeda UK Ltd) | 2191041000033110 | 446711000001101 | Pioglitazone hydrochloride |
| Pioglitazone 30mg tablets | 2190841000033113 | 326061000 | Pioglitazone hydrochloride |
| Glizofar 30mg tablets (Arrow Generics Ltd) | 6526041000033117 | 20023111000001108 | Pioglitazone hydrochloride |
| Diabiom 30mg tablets (Tillomed Laboratories Ltd) | 10337041000033113 | 29743011000001108 | Pioglitazone hydrochloride |
| Glidipion 30mg tablets (Actavis UK Ltd) | 8839341000033116 | 23369611000001106 | Pioglitazone hydrochloride |
| Glizofar 15mg tablets (Arrow Generics Ltd) | 6525941000033110 | 20022911000001104 | Pioglitazone hydrochloride |
| Pioglitazone 15mg tablets | 2190741000033115 | 326062007 | Pioglitazone hydrochloride |
| Actos 15mg tablets (Takeda UK Ltd) | 2190941000033117 | 58011000001106 | Pioglitazone hydrochloride |
| Diabiom 15mg tablets (Tillomed Laboratories Ltd) | 10336941000033112 | 29742811000001105 | Pioglitazone hydrochloride |
| Glidipion 15mg tablets (Actavis UK Ltd) | 10251341000033112 | 28940811000001104 | Pioglitazone hydrochloride |
| Actos 45mg tablets (Takeda UK Ltd) | 2996041000033119 | 5199411000001105 | Pioglitazone hydrochloride |
| Pioglitazone 45mg tablets | 2995941000033112 | 374897009 | Pioglitazone hydrochloride |
| Glizofar 45mg tablets (Arrow Generics Ltd) | 6526141000033118 | 20023411000001103 | Pioglitazone hydrochloride |
| Glidipion 45mg tablets (Actavis UK Ltd) | 8839441000033110 | 23372211000001107 | Pioglitazone hydrochloride |
| Diabiom 45mg tablets (Tillomed Laboratories Ltd) | 10337141000033112 | 29743211000001103 | Pioglitazone hydrochloride |
| Repaglinide 500microgram tablets | 1672941000033110 | 326047005 | Repaglinide |
| NovoNorm 500microgram tablets (Novo Nordisk Ltd) | 1659941000033116 | 152311000001101 | Repaglinide |
| Prandin 0.5mg tablets (Novo Nordisk Ltd) | 3993941000033119 | 10952411000001100 | Repaglinide |
| Enyglid 0.5mg tablets (Consilient Health Ltd) | 6391141000033118 | 19306711000001106 | Repaglinide |
| Enyglid 1mg tablets (Consilient Health Ltd) | 6391241000033113 | 19306911000001108 | Repaglinide |
| Prandin 1mg tablets (Novo Nordisk Ltd) | 3994041000033117 | 10956911000001107 | Repaglinide |
| NovoNorm 1mg tablets (Novo Nordisk Ltd) | 1660041000033118 | 494111000001103 | Repaglinide |
| Repaglinide 1mg tablets | 1672741000033112 | 326048000 | Repaglinide |
| Repaglinide 2mg tablets | 1672841000033119 | 326049008 | Repaglinide |
| NovoNorm 2mg tablets (Novo Nordisk Ltd) | 1660141000033119 | 840811000001100 | Repaglinide |
| Prandin 2mg tablets (Novo Nordisk Ltd) | 3994141000033118 | 10957811000001100 | Repaglinide |
| Enyglid 2mg tablets (Consilient Health Ltd) | 6391341000033115 | 19307311000001105 | Repaglinide |

| | | | |
|---|---|---|---|
| Saxagliptin 2.5mg tablets | 6123241000033112 | 443067000 | Saxagliptin hydrochloride |
| Onglyza 2.5mg tablets (AstraZeneca UK Ltd) | 6123341000033119 | 18596311000001108 | Saxagliptin hydrochloride |
| Saxagliptin 5mg tablets | 5316641000033118 | 443713000 | Saxagliptin hydrochloride |
| Onglyza 5mg tablets (AstraZeneca UK Ltd) | 5316741000033110 | 15993011000001101 | Saxagliptin hydrochloride |
| Ozempic 0.25mg/0.19ml solution for injection 1.5ml pre-filled pens (Novo Nordisk Ltd) | 12876541000033113 | 36470811000001107 | Semaglutide |
| Semaglutide 0.25mg/0.19ml solution for injection 1.5ml pre-filled disposable device | 12876241000033111 | 36490911000001109 | Semaglutide |
| Semaglutide 0.5mg/0.37ml solution for injection 1.5ml pre-filled disposable device | 12876341000033118 | 36491011000001101 | Semaglutide |
| Ozempic 0.5mg/0.37ml solution for injection 1.5ml pre-filled pens (Novo Nordisk Ltd) | 12876641000033114 | 36471111000001106 | Semaglutide |
| Ozempic 1mg/0.74ml solution for injection 3ml pre-filled pens (Novo Nordisk Ltd) | 12876441000033112 | 36471411000001101 | Semaglutide |
| Semaglutide 1mg/0.74ml solution for injection 3ml pre-filled disposable device | 12876141000033116 | 36491111000001100 | Semaglutide |
| Rybelsus 14mg tablets (Novo Nordisk Ltd) | 13712741000033116 | 38732411000001108 | Semaglutide |
| Semaglutide 14mg tablets | 13712441000033111 | 38840111000001100 | Semaglutide |
| Semaglutide 3mg tablets | 13712241000033110 | 38840211000001106 | Semaglutide |
| Rybelsus 3mg tablets (Novo Nordisk Ltd) | 13712541000033112 | 38731811000001101 | Semaglutide |
| Rybelsus 7mg tablets (Novo Nordisk Ltd) | 13712641000033113 | 38732111000001103 | Semaglutide |
| Semaglutide 7mg tablets | 13712341000033117 | 38840311000001103 | Semaglutide |
| Sitagliptin 25mg tablets | 7687441000033111 | 423962004 | Sitagliptin phosphate |
| Januvia 25mg tablets (Merck Sharp & Dohme Ltd) | 7687541000033112 | 20115111000001106 | Sitagliptin phosphate |
| Sitagliptin 100mg tablets | 4133241000033117 | 424345005 | Sitagliptin phosphate |
| Januvia 100mg tablets (Merck Sharp & Dohme Ltd) | 4133341000033110 | 11473711000001108 | Sitagliptin phosphate |
| Januvia 50mg tablets (Merck Sharp & Dohme Ltd) | 7687341000033117 | 20114811000001100 | Sitagliptin phosphate |
| Sitagliptin 50mg tablets | 7687241000033110 | 424513004 | Sitagliptin phosphate |
| Tolbutamide 500mg tablets | 1450941000033113 | 325267004 | Tolbutamide |

#### Table S12. Hypertension Aurum codes

| Term | Medcode ID | SNOMED CT Concept ID | SNOMED CT Description ID |
|---|---|---|---|
| HTN - Hypertension | 3117511000006117 | 38341003 | 2164904016 |
| Accelerated essential hypertension | 3784371000006115 | 78975002 | 503982017 |
| High blood pressure | 3117411000006118 | 38341003 | 64172013 |
| Malignant hypertensive heart disease NOS | 299650019 | 54225002 | 90135019 |
| History of hypertension | 4540441000006117 | 161501007 | 2987063016 |
| Benign hypertensive heart disease NOS | 299654011 | 36221001 | 60444016 |
| Secondary malignant hypertension NOS | 299677011 | 89242004 | 147988014 |
| Other specified hypertensive disease | 299686018 | 38341003 | 1215744012 |
| [X]Hypertension secondary to other renal disorders | 300871017 | 31992008 | 53452019 |
| Benign hypertensive renal disease | 1409014 | 193003 | 1409014 |
| HHD - Hypertensive heart disease | 3552531000006110 | 64715009 | 499930018 |
| Hypertensive disease | 64168014 | 38341003 | 64168014 |
| Hypertensive renal disease | 64282015 | 38481006 | 64282015 |
| Malignant hypertensive heart disease | 90135019 | 54225002 | 90135019 |
| Systolic hypertension | 93494011 | 56218007 | 93494011 |
| Primary hypertension | 99047018 | 59621000 | 99047018 |
| Hypertensive heart disease NOS | 741661000006118 | 64715009 | 107545013 |
| Hypertensive heart disease NOS with CCF | 741681000006111 | 64715009 | 107545013 |
| Hypertensive heart disease NOS without CCF | 741691000006114 | 64715009 | 107545013 |
| Malignant hypertension | 884121000006111 | 78975002 | 884121000006111 |
| Renal hypertension | 47076011 | 38481006 | 64282015 |
| Hypertensive crisis | 84112010 | 50490005 | 84111015 |
| Hypertensive heart disease | 107545013 | 64715009 | 107545013 |
| HT - Hypertension | 3117481000006113 | 38341003 | 490281011 |
| Stage 2 hypertension | 8286581000006114 | 846371000000103 | 2211211000000110 |
| Benign hypertensive heart disease without CCF | 504911000006115 | 77970009 | 129404019 |
| Hypertensive heart disease NOS | 299655012 | 64715009 | 107545013 |
| Hypertensive heart and renal disease NOS | 299675015 | 86234004 | 143003017 |
| Hypertensive renal failure | 3296351000006114 | 49220004 | 81984019 |
| Cardiomegaly - hypertensive | 411508017 | 275516004 | 411508017 |
| Severe hypertension (Nat Inst for Health Clinical Ex 2011) | 1846961000006115 | 843841000000109 | 2189451000000110 |
| Stage 2 hypertension (NICE - Nat Ins for Hth Clin Excl 2011) | 1846991000006111 | 846371000000103 | 2194941000000119 |
| Stage 1 hyperten (NICE 2011) with evidnce end organ damge | 1908721000006111 | 908651000000101 | 2335801000000114 |
| Systemic arterial hypertension | 3117461000006115 | 38341003 | 490278018 |
| High blood pressure | 64172013 | 24184005 | 196353013 |
| Secondary malignant hypertension | 151161000006115 | 89242004 | 147988014 |
| Benign hypertensive heart disease without congestive heart failure | 3768511000006114 | 77970009 | 129404019 |
| Malignant hypertension | 3642801000006112 | 70272006 | 116721012 |

| | | | |
|---|---|---|---|
| Hypertensive cardiopathy | 3552501000006119 | 64715009 | 107547017 |
| Hypertensive cardiomegaly | 3552511000006116 | 64715009 | 107548010 |
| Hypertensive heart&renal dis wth (congestive) heart failure | 741701000006114 | 194779001 | 299672017 |
| Hyperten heart&renal dis+both(congestv)heart and renal fail | 789941000006117 | 194781004 | 299674016 |
| Hypertensive heart and renal disease with renal failure | 299673010 | 194780003 | 299673010 |
| [X]Hypertensive diseases | 300869017 | 38341003 | 1215744012 |
| Hypertensive renal disease with renal failure | 299665018 | 49220004 | 81984019 |
| Systemic primary arterial hypertension | 3468501000006117 | 59621000 | 99046010 |
| Benign hypertensive heart disease with CCF | 504901000006118 | 194767001 | 299653017 |
| Stage 2 hypertension | 1806081000006115 | 1806081000006104 | 1806081000006115 |
| HBP - High blood pressure | 3117471000006110 | 38341003 | 490280012 |
| Accelerated secondary hypertension | 3950631000006119 | 89242004 | 508416019 |
| Hypertensive cardiovascular disease | 3552521000006112 | 64715009 | 107549019 |
| Benign essential hypertension | 3135013 | 1201005 | 3135013 |
| [RFC] Hypertension | 909441000006118 | 909441000006102 | 909441000006118 |
| Essential hypertension NOS | 395751018 | 59621000 | 99042012 |
| Secondary malignant renovascular hypertension | 299676019 | 194783001 | 299676019 |
| Essential hypertension | 99042012 | 59621000 | 99042012 |
| Severe hypertension | 2193021000000110 | 843841000000109 | 2193021000000110 |
| Hypertension resistant to drug therapy | 2193971000000110 | 845891000000103 | 2193971000000110 |
| Malignant hypertensive renal disease | 108730018 | 65443008 | 108730018 |
| Malignant hypertensive heart AND renal disease | 110659019 | 66610008 | 110659019 |
| H/O: hypertension | 251674014 | 161501007 | 251674014 |
| [X]Other secondary hypertension | 300870016 | 31992008 | 53452019 |
| Hypertension confirmed | 1823901000006112 | 1823901000006108 | 1823901000006112 |
| BP+ - Hypertension | 3117501000006115 | 38341003 | 490283014 |
| Hypertensive vascular disease | 3117421000006114 | 38341003 | 64173015 |
| Benign hypertensive heart AND renal disease | 109700019 | 66052004 | 109700019 |
| Malignant essential hypertension | 131046010 | 78975002 | 131046010 |
| Hypertensive heart AND renal disease | 143003017 | 86234004 | 143003017 |
| Hypertensive disorder, systemic arterial | 3117521000006113 | 38341003 | 2671386015 |
| BP - High blood pressure | 3117451000006117 | 38341003 | 490277011 |
| Resistant hypertension | 8286321000006117 | 845891000000103 | 2194001000000116 |
| Severe hypertension | 1806141000006113 | 1806141000006109 | 1806141000006113 |
| Hypertensive disease NOS | 299687010 | 38341003 | 1215744012 |
| Hypertension NOS | 790121000006116 | 38341003 | 64176011 |
| BP - hypertensive disease | 523801000006119 | 38341003 | 64168014 |
| Hypertensive nephropathy | 3119661000006114 | 38481006 | 64285018 |
| Hypertensive renal disease NOS | 395753015 | 38481006 | 64282015 |
| Hypertension due to renovascular disease | 4356081000006115 | 123799005 | 1220673019 |
| Malignant hypertensive heart disease with CCF | 728671000006119 | 83105008 | 1236017010 |

| | | | |
|---|---|---|---|
| Malignant hypertensive heart disease without CCF | 728681000006116 | 36315003 | 60617018 |
| Hypertensive renal + heart dis | 884131000006114 | 86234004 | 884131000006114 |
| Hypertensive retinopathy | 12496011 | 6962006 | 12496011 |
| Malignant hypertensive heart disease with congestive heart failure | 3852501000006115 | 83105008 | 137848017 |
| Benign hypertensive heart disease | 60444016 | 36221001 | 60444016 |
| Idiopathic hypertension | 3468491000006113 | 59621000 | 99044013 |

#### Table S13. Anti-Hypertension Aurum product codes

| Term from EMIS | Prod code ID | dmd ID | Drug substance name |
|---|---|---|---|
| Rasilez 300mg tablets (Noden Pharma DAC) | 4213941000033117 | 11961711000001103 | Aliskiren hemifumarate |
| Aliskiren 300mg tablets | 4213741000033115 | 425669009 | Aliskiren hemifumarate |
| Rasilez 150mg tablets (Noden Pharma DAC) | 4213841000033113 | 11960911000001108 | Aliskiren hemifumarate |
| Aliskiren 150mg tablets | 4213641000033112 | 425960005 | Aliskiren hemifumarate |
| Amoride 5mg tablets (Dr Reddy's Laboratories (UK) Ltd) | 2956041000033112 | 220711000001102 | Amiloride hydrochloride |
| Amiloride 5mg tablets | 57141000033118 | 318052005 | Amiloride hydrochloride |
| Amiloride 2.5mg / Cyclopenthiazide 250microgram tablets | 1903541000033116 | 318096005 | Amiloride hydrochloride/ Cyclopenthiazide |
| Navispare 2.5mg/250microgram tablets (Advanz Pharma) | 956741000033114 | 535711000001100 | Amiloride hydrochloride/ Cyclopenthiazide |
| Burinex A 5mg/1mg tablets (LEO Pharma) | 172341000033112 | 33911000001104 | Amiloride hydrochloride/ Bumetanide |
| Amiloride 5mg / Bumetanide 1mg tablets | 1845641000033119 | 318097001 | Amiloride hydrochloride/ Bumetanide |
| Co-amilozide 2.5mg/25mg tablets | 376941000033117 | 318121006 | Amiloride hydrochloride/ Hydrochlorothiazide |
| Moduret 25 tablets (Merck Sharp & Dohme Ltd) | 935441000033112 | 314211000001100 | Amiloride hydrochloride/ Hydrochlorothiazide |
| Synuretic 25 tablets (Dr Reddy's Laboratories (UK) Ltd) | 2955841000033110 | 465111000001101 | Amiloride hydrochloride/ Hydrochlorothiazide |
| Frumil LS 20mg/2.5mg tablets (Sanofi) | 614341000033113 | 550711000001106 | Amiloride hydrochloride/ Furosemide |
| Co-amilofruse 2.5mg/20mg tablets | 376841000033113 | 318135008 | Amiloride hydrochloride/ Furosemide |
| Co-amilofruse 5mg/40mg tablets | 376641000033112 | 318136009 | Amiloride hydrochloride/ Furosemide |
| Frumil 40mg/5mg tablets (Sanofi) | 614641000033117 | 427411000001106 | Amiloride hydrochloride/ Furosemide |
| Froop Co 5mg/40mg tablets (Ashbourne Pharmaceuticals Ltd) | 1611441000033117 | 331311000001103 | Amiloride hydrochloride/ Furosemide |
| Lasoride 5mg/40mg tablets (Sanofi) | 818941000033112 | 678511000001106 | Amiloride hydrochloride/ Furosemide |
| Fru-Co 5mg/40mg tablets (Teva UK Ltd) | 615241000033116 | 818511000001106 | Amiloride hydrochloride/ Furosemide |
| Frumil Forte 10mg/80mg tablets (Sanofi) | 615641000033118 | 82611000001106 | Amiloride hydrochloride/ Furosemide |

| | | | |
|---|---|---|---|
| Co-amilofruse 10mg/80mg tablets | 376741000033115 | 318137000 | Amiloride hydrochloride/ Furosemide |
| Moducren tablets (Merck Sharp & Dohme Ltd) | 935341000033118 | 74311000001101 | Amiloride hydrochloride/ Hydrochlorothiazide/ Timolol maleate |
| Kalten capsules (M & A Pharmachem Ltd) | 793741000033117 | 237011000001100 | Amiloride hydrochloride/ Atenolol/ Hydrochlorothiazide |
| Co-amilozide 5mg/50mg tablets | 369141000033118 | 377566005 | Amiloride hydrochloride/ Hydrochlorothiazide |
| Amilmaxco 5mg/50mg tablets (Ashbourne Pharmaceuticals Ltd) | 57341000033115 | 712211000001108 | Amiloride hydrochloride/ Hydrochlorothiazide |
| Moduretic 5mg/50mg tablets (Merck Sharp & Dohme Ltd) | 935541000033113 | 453811000001103 | Amiloride hydrochloride/ Hydrochlorothiazide |
| Amil-Co 5mg/50mg tablets (IVAX Pharmaceuticals UK Ltd) | 60341000033110 | 636611000001107 | Amiloride hydrochloride/ Hydrochlorothiazide |
| Synuretic 50 tablets (Dr Reddy's Laboratories (UK) Ltd) | 2955741000033117 | 601011000001103 | Amiloride hydrochloride/ Hydrochlorothiazide |
| Co-amilofruse 5mg/40mg/5ml oral suspension | 5710441000033117 | 8427011000001108 | Amiloride hydrochloride/ Furosemide |
| Co-amilozide 5mg/50mg/5ml oral solution | 351941000033111 | 8792311000001109 | Amiloride hydrochloride/ Hydrochlorothiazide |
| Timolol 10mg / Amiloride 2.5mg / Hydrochlorothiazide 25mg tablets (Essential Generics Ltd) | 5376441000033114 | 16458411000001107 | Amiloride hydrochloride/ Hydrochlorothiazide/ Timolol maleate |
| Amilamont 5mg/5ml oral solution sugar free (Rosemont Pharmaceuticals Ltd) | 2101641000033119 | 799711000001108 | Amiloride hydrochloride |
| Amiloride 5mg/5ml oral solution sugar free | 53841000033117 | 35900111000001108 | Amiloride hydrochloride |
| Amlodipine 5mg tablets | 3038041000033111 | 319283006 | Amlodipine |
| Istin 5mg tablets (Upjohn UK Ltd) | 787941000033112 | 172711000001100 | Amlodipine |
| Amlostin 5mg tablets (Discovery Pharmaceuticals) | 3188741000033111 | 8046211000001107 | Amlodipine |
| Amlostin 10mg tablets (Discovery Pharmaceuticals) | 3188841000033118 | 8046411000001106 | Amlodipine |
| Istin 10mg tablets (Upjohn UK Ltd) | 788041000033110 | 408111000001107 | Amlodipine |
| Amlodipine 10mg tablets | 3038141000033110 | 319284000 | Amlodipine |
| Sevikar 20mg/5mg tablets (Daiichi Sankyo UK Ltd) | 5006141000033110 | 15773211000001105 | Amlodipine besilate/ Olmesartan medoxomil |
| Olmesartan medoxomil 20mg / Amlodipine 5mg tablets | 5005841000033114 | 429502004 | Amlodipine besilate/ Olmesartan medoxomil |

| | | | |
|---|---|---|---|
| Olmesartan medoxomil 40mg / Amlodipine 5mg tablets | 5005941000033118 | 429503009 | Amlodipine besilate/ Olmesartan medoxomil |
| Sevikar 40mg/5mg tablets (Daiichi Sankyo UK Ltd) | 5006241000033115 | 15772911000001108 | Amlodipine besilate/ Olmesartan medoxomil |
| Sevikar 40mg/10mg tablets (Daiichi Sankyo UK Ltd) | 5006341000033113 | 15772611000001102 | Amlodipine besilate/ Olmesartan medoxomil |
| Olmesartan medoxomil 40mg / Amlodipine 10mg tablets | 5006041000033111 | 429678006 | Amlodipine besilate/ Olmesartan medoxomil |
| Amlodipine 2.5mg tablets | 13422241000033114 | 429828006 | Amlodipine |
| Amlodipine 10mg/5ml oral suspension | 3963741000033115 | 8278111000001105 | Amlodipine |
| Amlodipine 5mg/5ml oral suspension | 5888741000033110 | 8278311000001107 | Amlodipine |
| Amlodipine 10mg / Valsartan 160mg tablets | 4021641000033111 | 11160111000001107 | Amlodipine besilate/ Valsartan |
| Exforge 10mg/160mg tablets (Novartis Pharmaceuticals UK Ltd) | 4021941000033116 | 11160711000001108 | Amlodipine besilate/ Valsartan |
| Exforge 5mg/160mg tablets (Novartis Pharmaceuticals UK Ltd) | 4021841000033112 | 11161511000001105 | Amlodipine besilate/ Valsartan |
| Amlodipine 5mg / Valsartan 160mg tablets | 4021541000033110 | 11160211000001101 | Amlodipine besilate/ Valsartan |
| Amlodipine 5mg / Valsartan 80mg tablets | 4021441000033114 | 11160311000001109 | Amlodipine besilate/ Valsartan |
| Exforge 5mg/80mg tablets (Novartis Pharmaceuticals UK Ltd) | 4021741000033119 | 11161811000001108 | Amlodipine besilate/ Valsartan |
| Amlodipine 5mg/5ml oral solution | 5490241000033113 | 13892511000001100 | Amlodipine |
| Amlodipine 1.5mg/5ml oral suspension | 4897741000033118 | 15773511000001108 | Amlodipine |
| Sevikar HCT 20mg/5mg/12.5mg tablets (Daiichi Sankyo UK Ltd) | 6180641000033112 | 18986411000001108 | Amlodipine besilate/ Hydrochlorothiazide/ Olmesartan medoxomil |
| Sevikar HCT 40mg/10mg/12.5mg tablets (Daiichi Sankyo UK Ltd) | 6180841000033113 | 18987011000001101 | Amlodipine besilate/ Hydrochlorothiazide/ Olmesartan medoxomil |
| Sevikar HCT 40mg/10mg/25mg tablets (Daiichi Sankyo UK Ltd) | 6181041000033110 | 18987611000001108 | Amlodipine besilate/ Hydrochlorothiazide/ Olmesartan medoxomil |
| Sevikar HCT 40mg/5mg/12.5mg tablets (Daiichi Sankyo UK Ltd) | 6180741000033115 | 18986711000001102 | Amlodipine besilate/ Hydrochlorothiazide/ Olmesartan medoxomil |
| Sevikar HCT 40mg/5mg/25mg tablets (Daiichi Sankyo UK Ltd) | 6180941000033117 | 18987311000001103 | Amlodipine besilate/ Hydrochlorothiazide/ Olmesartan medoxomil |
| Amlodipine 10mg/5ml oral solution | 7740641000033112 | 20478011000001105 | Amlodipine |
| Perindopril erbumine 4mg / Amlodipine 10mg tablets | 9105341000033116 | 23984911000001108 | Amlodipine besilate/ |

| | | | |
|---|---|---|---|
| | | | Perindopril erbumine |
| Perindopril erbumine 4mg / Amlodipine 5mg tablets | 9105241000033114 | 23985011000001108 | Amlodipine besilate/ Perindopril erbumine |
| Perindopril erbumine 8mg / Amlodipine 10mg tablets | 9105541000033111 | 23985111000001109 | Amlodipine besilate/ Perindopril erbumine |
| Perindopril erbumine 8mg / Amlodipine 5mg tablets | 9105441000033110 | 23985211000001103 | Amlodipine besilate/ Perindopril erbumine |
| Amlodipine 10mg/5ml oral solution sugar free | 10387441000033111 | 29826211000001109 | Amlodipine |
| Amlodipine 5mg/5ml oral solution sugar free | 4508041000033119 | 29826311000001101 | Amlodipine |
| Amlodipine 5mg/5ml oral suspension sugar free | 12682241000033110 | 36237311000001101 | Amlodipine |
| Edarbi 40mg tablets (Takeda UK Ltd) | 7755241000033119 | 20351211000001103 | Azilsartan medoxomil |
| Azilsartan medoxomil 40mg tablets | 7754941000033110 | 449109006 | Azilsartan medoxomil |
| Azilsartan medoxomil 80mg tablets | 7755041000033110 | 449333009 | Azilsartan medoxomil |
| Edarbi 80mg tablets (Takeda UK Ltd) | 7755341000033112 | 20351811000001102 | Azilsartan medoxomil |
| Azilsartan medoxomil 20mg tablets | 7754841000033119 | 20418711000001104 | Azilsartan medoxomil |
| Edarbi 20mg tablets (Takeda UK Ltd) | 7755141000033114 | 20350911000001100 | Azilsartan medoxomil |
| Bendroflumethiazide 2.5mg tablets | 3083141000033112 | 317919004 | Bendroflumethiazide |
| Urizide 2.5mg tablets (Dr Reddy's Laboratories (UK) Ltd) | 2955941000033119 | 485311000001101 | Bendroflumethiazide |
| Aprinox 2.5mg tablets (Advanz Pharma) | 74541000033119 | 672111000001100 | Bendroflumethiazide |
| Neo-Naclex 2.5mg tablets (Advanz Pharma) | 6045641000033115 | 18149011000001101 | Bendroflumethiazide |
| Urizide 5mg tablets (Dr Reddy's Laboratories (UK) Ltd) | 1495441000033116 | 898211000001109 | Bendroflumethiazide |
| Neo-Naclex 5mg tablets (Mercury Pharma Group Ltd) | 965341000033114 | 817911000001100 | Bendroflumethiazide |
| Aprinox 5mg tablets (Amdipharm Plc) | 74641000033118 | 120911000001103 | Bendroflumethiazide |
| Bendroflumethiazide 5mg tablets | 3083241000033117 | 317920005 | Bendroflumethiazide |
| Corgaretic 40mg tablets (Sanofi-Synthelabo Ltd) | 375441000033118 | 3886211000001102 | Bendroflumethiazide/ Nadolol |
| Corgaretic 80mg tablets (Sanofi-Synthelabo Ltd) | 375541000033117 | 4057911000001102 | Bendroflumethiazide/ Nadolol |
| Prestim tablets (Meda Pharmaceuticals Ltd) | 1132741000033111 | 98411000001109 | Bendroflumethiazide/ Timolol maleate |
| Timolol 10mg / Bendroflumethiazide 2.5mg tablets | 3346641000033118 | 318556002 | Bendroflumethiazide/ Timolol maleate |
| Tenben 25mg/1.25mg capsules (Galen Ltd) | 1414241000033117 | 721411000001106 | Atenolol/ Bendroflumethiazide |
| Inderetic 80mg/2.5mg capsules (AstraZeneca UK Ltd) | 756641000033114 | 333111000001104 | Bendroflumethiazide/ Propranolol hydrochloride |

| | | | |
|---|---|---|---|
| Bendroflumethiazide 5mg/5ml oral suspension | 5401741000033110 | 8306711000001109 | Bendroflumethiazide |
| Bendroflumethiazide 2.5mg/5ml oral suspension | 3151841000033117 | 8306811000001101 | Bendroflumethiazide |
| Bendroflumethiazide 1.25mg/5ml oral suspension | 5453941000033119 | 8307011000001105 | Bendroflumethiazide |
| Centyl K modified-release tablets (LEO Pharma) | 234841000033110 | 3932711000001105 | Bendroflumethiazide/ Potassium chloride |
| Bendroflumethiazide 2.5mg / Potassium chloride 573mg (potassium 7.7mmol) modified-release tablets | 3161041000033115 | 35910011000001105 | Bendroflumethiazide/ Potassium chloride |
| Neo-Naclex-K modified-release tablets (Mercury Pharma Group Ltd) | 967041000033118 | 3638211000001100 | Bendroflumethiazide/ Potassium chloride |
| Bendroflumethiazide 2.5mg / Potassium chloride 630mg (potassium 8.4mmol) modified-release tablets | 3162841000033118 | 35910111000001106 | Bendroflumethiazide/ Potassium chloride |
| Inderex 160mg/5mg modified-release capsules (AstraZeneca UK Ltd) | 756741000033117 | 350811000001100 | Bendroflumethiazide/ Propranolol hydrochloride |
| Amias 2mg tablets (Takeda UK Ltd) | 59241000033118 | 97311000001103 | Candesartan cilexetil |
| Candesartan 2mg tablets | 211641000033118 | 318977009 | Candesartan cilexetil |
| Candesartan 4mg tablets | 211741000033110 | 318978004 | Candesartan cilexetil |
| Amias 4mg tablets (Takeda UK Ltd) | 59341000033111 | 857411000001100 | Candesartan cilexetil |
| Amias 8mg tablets (Takeda UK Ltd) | 59441000033117 | 36011000001106 | Candesartan cilexetil |
| Candesartan 8mg tablets | 211841000033117 | 318979007 | Candesartan cilexetil |
| Candesartan 16mg tablets | 211541000033119 | 318980005 | Candesartan cilexetil |
| Amias 16mg tablets (Takeda UK Ltd) | 59141000033113 | 908511000001100 | Candesartan cilexetil |
| Candesartan 32mg tablets | 3227241000033111 | 376998003 | Candesartan cilexetil |
| Amias 32mg tablets (Takeda UK Ltd) | 3227341000033118 | 8983911000001107 | Candesartan cilexetil |
| Capozide LS 12.5mg/25mg tablets (Bristol-Myers Squibb Pharmaceuticals Ltd) | 209241000033119 | 546711000001107 | Captopril/ Hydrochlorothiazide |
| Co-zidocapt 12.5mg/25mg tablets | 1893141000033119 | 318806002 | Captopril/ Hydrochlorothiazide |
| Co-zidocapt 25mg/50mg tablets | 1893241000033114 | 318807006 | Captopril/ Hydrochlorothiazide |
| Acezide 25mg/50mg tablets (Bristol-Myers Squibb Pharmaceuticals Ltd) | 13141000033119 | 263311000001101 | Captopril/ Hydrochlorothiazide |
| Capozide 25mg/50mg tablets (Bristol-Myers Squibb Pharmaceuticals Ltd) | 216441000033110 | 17311000001102 | Captopril/ Hydrochlorothiazide |
| Capoten 12.5mg tablets (Bristol-Myers Squibb Pharmaceuticals Ltd) | 214141000033113 | 134511000001103 | Captopril |
| Ecopace 12.5mg tablets (Advanz Pharma) | 2945841000033112 | 221411000001104 | Captopril |
| Acepril 12.5mg tablets (Bristol-Myers Squibb Pharmaceuticals Ltd) | 10841000033119 | 660111000001107 | Captopril |
| Tensopril 12.5mg tablets (Teva UK Ltd) | 2928341000033114 | 673711000001107 | Captopril |

| | | | |
|---|---|---|---|
| Kaplon 12.5mg tablets (Teva UK Ltd) | 1622041000033110 | 777711000001105 | Captopril |
| Captopril 12.5mg tablets | 213441000033119 | 318820009 | Captopril |
| Captopril 25mg tablets | 214441000033117 | 318821008 | Captopril |
| Ecopace 25mg tablets (Advanz Pharma) | 2945941000033116 | 817411000001108 | Captopril |
| Acepril 25mg tablets (Bristol-Myers Squibb Pharmaceuticals Ltd) | 10941000033110 | 517711000001103 | Captopril |
| Kaplon 25mg tablets (Teva UK Ltd) | 1622141000033114 | 477111000001100 | Captopril |
| Tensopril 25mg tablets (Teva UK Ltd) | 2928441000033115 | 597511000001109 | Captopril |
| Capoten 25mg tablets (Bristol-Myers Squibb Pharmaceuticals Ltd) | 214241000033118 | 455611000001103 | Captopril |
| Capoten 50mg tablets (Bristol-Myers Squibb Pharmaceuticals Ltd) | 214341000033111 | 386611000001109 | Captopril |
| Kaplon 50mg tablets (Teva UK Ltd) | 1622241000033119 | 572511000001102 | Captopril |
| Acepril 50mg tablets (Bristol-Myers Squibb Pharmaceuticals Ltd) | 11041000033117 | 814111000001108 | Captopril |
| Ecopace 50mg tablets (Advanz Pharma) | 2946041000033114 | 767911000001101 | Captopril |
| Tensopril 50mg tablets (Teva UK Ltd) | 2928541000033119 | 653111000001108 | Captopril |
| Captopril 50mg tablets | 214541000033116 | 318824000 | Captopril |
| Captopril 10mg/5ml oral suspension | 5008141000033111 | 8346811000001109 | Captopril |
| Captopril 12.5mg/5ml oral solution | 5898741000033114 | 8347111000001104 | Captopril |
| Captopril 12.5mg/5ml oral suspension | 5898841000033116 | 8347211000001105 | Captopril |
| Captopril 15mg/5ml oral suspension | 5401641000033118 | 8347411000001109 | Captopril |
| Captopril 20mg/5ml oral suspension | 2719841000033113 | 8348311000001101 | Captopril |
| Captopril 25mg/5ml oral solution | 3012741000033115 | 8348511000001107 | Captopril |
| Captopril 25mg/5ml oral suspension | 5132041000033112 | 8348611000001106 | Captopril |
| Captopril 3mg/5ml oral solution | 3851341000033117 | 8350511000001107 | Captopril |
| Captopril 5mg/5ml oral solution | 6001041000033115 | 8351611000001101 | Captopril |
| Captopril 5mg/5ml oral suspension | 2656341000033110 | 8351811000001102 | Captopril |
| Captopril 6.25mg/5ml oral solution | 5898941000033112 | 8351911000001107 | Captopril |
| Captopril 6.25mg/5ml oral suspension | 5899041000033115 | 8352011000001100 | Captopril |
| Captopril 2mg capsules | 3940841000033110 | 8791811000001109 | Captopril |
| Captopril 4mg capsules | 5454041000033117 | 8791911000001104 | Captopril |
| Captopril 8mg/5ml oral suspension | 3333941000033117 | 19820711000001101 | Captopril |
| Noyada 25mg/5ml oral solution (Martindale Pharmaceuticals Ltd) | 8961341000033119 | 23681711000001107 | Captopril |
| Captopril 25mg/5ml oral solution sugar free | 9108041000033111 | 23707311000001108 | Captopril |
| Noyada 5mg/5ml oral solution (Martindale Pharmaceuticals Ltd) | 8961241000033112 | 23682011000001102 | Captopril |
| Captopril 5mg/5ml oral solution sugar free | 9108141000033110 | 23707511000001102 | Captopril |
| Irbesartan 300mg / Hydrochlorothiazide 12.5mg tablets | 3161241000033111 | 134460003 | Hydrochlorothiazide/ Irbesartan |
| CoAprovel 300mg/12.5mg tablets (Sanofi) | 2216241000033110 | 682711000001109 | Hydrochlorothiazide/ Irbesartan |
| CoAprovel 150mg/12.5mg tablets (Sanofi) | 2216141000033115 | 792411000001108 | Hydrochlorothiazide/ Irbesartan |
| Irbesartan 150mg / Hydrochlorothiazide 12.5mg tablets | 3161141000033116 | 134461004 | Hydrochlorothiazide/ Irbesartan |
| Co-Betaloc tablets (Pfizer Ltd) | 372741000033116 | 2977611000001106 | Hydrochlorothiazide/ Metoprolol tartrate |

| | | | |
|---|---|---|---|
| Metoprolol 100mg / Hydrochlorothiazide 12.5mg tablets | 3161341000033118 | 318546001 | Hydrochlorothiazide/ Metoprolol tartrate |
| Secadrex 200mg/12.5mg tablets (Sanofi) | 1274641000033111 | 878911000001106 | Acebutolol hydrochloride/ Hydrochlorothiazide |
| Acebutolol 200mg / Hydrochlorothiazide 12.5mg tablets | 3162941000033114 | 318586009 | Acebutolol hydrochloride/ Hydrochlorothiazide |
| Monozide 10 tablets (Wyeth Pharmaceuticals) | 937341000033111 | 4542911000001104 | Bisoprolol fumarate/ Hydrochlorothiazide |
| Enalapril 20mg / Hydrochlorothiazide 12.5mg tablets | 3162241000033117 | 318849001 | Enalapril maleate/ Hydrochlorothiazide |
| Innozide 20mg/12.5mg tablets (Merck Sharp & Dohme Ltd) | 770141000033115 | 146811000001108 | Enalapril maleate/ Hydrochlorothiazide |
| Zestoretic 20 tablets (AstraZeneca UK Ltd) | 1552341000033118 | 3143111000001100 | Hydrochlorothiazide/ Lisinopril |
| Carace 20 Plus tablets (Merck Sharp & Dohme Ltd) | 217041000033116 | 3143511000001109 | Hydrochlorothiazide/ Lisinopril |
| Caralpha 20mg/12.5mg tablets (Actavis UK Ltd) | 3136141000033118 | 7385711000001106 | Hydrochlorothiazide/ Lisinopril |
| Lisicostad 20mg/12.5mg tablets (Genus Pharmaceuticals Ltd) | 3279641000033119 | 8145911000001100 | Hydrochlorothiazide/ Lisinopril |
| Lisinopril 20mg / Hydrochlorothiazide 12.5mg tablets | 3160941000033113 | 318880006 | Hydrochlorothiazide/ Lisinopril |
| Lisoretic 20mg/12.5mg tablets (Bristol Laboratories Ltd) | 13443541000033119 | 38240011000001105 | Hydrochlorothiazide/ Lisinopril |
| Lisinopril 10mg / Hydrochlorothiazide 12.5mg tablets | 3160841000033117 | 318884002 | Hydrochlorothiazide/ Lisinopril |
| Caralpha 10mg/12.5mg tablets (Actavis UK Ltd) | 3136041000033117 | 7385911000001108 | Hydrochlorothiazide/ Lisinopril |
| Lisicostad 10mg/12.5mg tablets (Genus Pharmaceuticals Ltd) | 3279541000033115 | 8145711000001102 | Hydrochlorothiazide/ Lisinopril |
| Zestoretic 10 tablets (AstraZeneca UK Ltd) | 1551841000033118 | 3144311000001101 | Hydrochlorothiazide/ Lisinopril |
| Carace 10 Plus tablets (Merck Sharp & Dohme Ltd) | 209841000033115 | 3144511000001107 | Hydrochlorothiazide/ Lisinopril |
| Lisoretic 10mg/12.5mg tablets (Bristol Laboratories Ltd) | 13443441000033115 | 38240211000001100 | Hydrochlorothiazide/ Lisinopril |
| Quinapril 10mg / Hydrochlorothiazide 12.5mg tablets | 3160641000033118 | 318892006 | Hydrochlorothiazide/ Quinapril hydrochloride |
| Accuretic 10mg/12.5mg tablets (Pfizer Ltd) | 11241000033113 | 260211000001104 | Hydrochlorothiazide/ Quinapril hydrochloride |
| Losartan 50mg / Hydrochlorothiazide 12.5mg tablets | 3161741000033117 | 318959004 | Hydrochlorothiazide/ Losartan potassium |
| Cozaar-Comp 50mg/12.5mg tablets (Merck Sharp & Dohme Ltd) | 370841000033110 | 255911000001105 | Hydrochlorothiazide/ Losartan potassium |
| Hydrochlorothiazide 25mg tablets | 744941000033110 | 376209006 | Hydrochlorothiazide |
| Hydrosaluric 25mg tablets (Merck Sharp & Dohme Ltd) | 742541000033112 | 4544011000001109 | Hydrochlorothiazide |
| Hydrochlorothiazide 50mg tablets | 745441000033118 | 376508004 | Hydrochlorothiazide |

| | | | |
|---|---|---|---|
| Hydrosaluric 50mg tablets (Merck Sharp & Dohme Ltd) | 742641000033113 | 4546211000001109 | Hydrochlorothiazide |
| Co-Diovan 80mg/12.5mg tablets (Novartis Pharmaceuticals UK Ltd) | 3190841000033119 | 8150111000001108 | Hydrochlorothiazide/ Valsartan |
| Valsartan 80mg / Hydrochlorothiazide 12.5mg tablets | 3190741000033112 | 377488008 | Hydrochlorothiazide/ Valsartan |
| Cozaar-Comp 100mg/25mg tablets (Merck Sharp & Dohme Ltd) | 3346941000033113 | 9566911000001105 | Hydrochlorothiazide/ Losartan potassium |
| Losartan 100mg / Hydrochlorothiazide 25mg tablets | 3346841000033117 | 395497004 | Hydrochlorothiazide/ Losartan potassium |
| Chlorothiazide 250mg tablets | 5093641000033110 | 395516007 | Chlorothiazide |
| Valsartan 160mg / Hydrochlorothiazide 12.5mg tablets | 3161441000033112 | 395521005 | Hydrochlorothiazide/ Valsartan |
| Co-Diovan 160mg/12.5mg tablets (Novartis Pharmaceuticals UK Ltd) | 3152541000033112 | 7668611000001104 | Hydrochlorothiazide/ Valsartan |
| Telmisartan 40mg / Hydrochlorothiazide 12.5mg tablets | 3162641000033119 | 407855002 | Hydrochlorothiazide/ Telmisartan |
| MicardisPlus 40mg/12.5mg tablets (Boehringer Ingelheim Ltd) | 2864241000033110 | 3806311000001109 | Hydrochlorothiazide/ Telmisartan |
| Tolucombi 40mg/12.5mg tablets (Consilient Health Ltd) | 9292641000033116 | 24573211000001101 | Hydrochlorothiazide/ Telmisartan |
| Actelsar HCT 40mg/12.5mg tablets (Actavis UK Ltd) | 9159341000033112 | 24411411000001100 | Hydrochlorothiazide/ Telmisartan |
| MicardisPlus 80mg/12.5mg tablets (Boehringer Ingelheim Ltd) | 2864341000033117 | 3806911000001105 | Hydrochlorothiazide/ Telmisartan |
| Telmisartan 80mg / Hydrochlorothiazide 12.5mg tablets | 3162741000033111 | 407856001 | Hydrochlorothiazide/ Telmisartan |
| Actelsar HCT 80mg/12.5mg tablets (Actavis UK Ltd) | 9159441000033118 | 24412511000001109 | Hydrochlorothiazide/ Telmisartan |
| Tolucombi 80mg/12.5mg tablets (Consilient Health Ltd) | 9292741000033113 | 24573411000001102 | Hydrochlorothiazide/ Telmisartan |
| Chlorothiazide 250mg/5ml oral suspension | 246241000033116 | 408039005 | Chlorothiazide |
| Olmetec Plus 20mg/12.5mg tablets (Daiichi Sankyo UK Ltd) | 3909141000033116 | 10261511000001103 | Hydrochlorothiazide/ Olmesartan medoxomil |
| Olmesartan medoxomil 20mg / Hydrochlorothiazide 12.5mg tablets | 3908941000033112 | 409184002 | Hydrochlorothiazide/ Olmesartan medoxomil |
| Olmesartan medoxomil 40mg / Hydrochlorothiazide 12.5mg tablets | 5566341000033111 | 409185001 | Hydrochlorothiazide/ Olmesartan medoxomil |
| Olmetec Plus 40mg/12.5mg tablets (Daiichi Sankyo UK Ltd) | 5566441000033117 | 17220911000001102 | Hydrochlorothiazide/ Olmesartan medoxomil |
| Valsartan 160mg / Hydrochlorothiazide 25mg tablets | 3161541000033113 | 409298002 | Hydrochlorothiazide/ Valsartan |
| Co-Diovan 160mg/25mg tablets (Novartis Pharmaceuticals UK Ltd) | 3152641000033113 | 7668911000001105 | Hydrochlorothiazide/ Valsartan |
| Co-triamterzide 50mg/25mg tablets | 370241000033111 | 410896007 | Hydrochlorothiazide/ Triamterene |
| Triamaxco 50mg/25mg tablets (Ashbourne Pharmaceuticals Ltd) | 1468141000033118 | 721311000001104 | Hydrochlorothiazide/ Triamterene |
| Dyazide 50mg/25mg tablets (Advanz Pharma) | 492341000033115 | 132811000001106 | Hydrochlorothiazide/ Triamterene |
| Triam-Co 50mg/25mg tablets (IVAX Pharmaceuticals UK Ltd) | 1470341000033112 | 191211000001106 | Hydrochlorothiazide/ Triamterene |
| Chlorothiazide 150mg/5ml oral suspension | 5899341000033118 | 8358511000001105 | Chlorothiazide |
| Chlorothiazide 25mg/5ml oral suspension | 5899441000033112 | 8358711000001100 | Chlorothiazide |

| | | | |
|---|---|---|---|
| Hydrochlorothiazide 50mg/5ml oral solution | 2746441000033111 | 8529611000001101 | Hydrochlorothiazide |
| Olmesartan medoxomil 20mg / Hydrochlorothiazide 25mg tablets | 3909041000033115 | 10270711000001105 | Hydrochlorothiazide/ Olmesartan medoxomil |
| Olmetec Plus 20mg/25mg tablets (Daiichi Sankyo UK Ltd) | 3909241000033111 | 10261811000001100 | Hydrochlorothiazide/ Olmesartan medoxomil |
| Irbesartan 300mg / Hydrochlorothiazide 25mg tablets | 3995241000033112 | 10970311000001105 | Hydrochlorothiazide/ Irbesartan |
| CoAprovel 300mg/25mg tablets (Sanofi) | 3995341000033119 | 10968611000001106 | Hydrochlorothiazide/ Irbesartan |
| Chlorothiazide 200mg/5ml oral suspension | 3924141000033111 | 12502311000001102 | Chlorothiazide |
| Chlorothiazide 250mg/5ml oral solution | 5991141000033110 | 12503011000001109 | Chlorothiazide |
| Hydrochlorothiazide 5mg/5ml oral solution | 5969841000033119 | 12538711000001107 | Hydrochlorothiazide |
| Hydrochlorothiazide 5mg/5ml oral suspension | 5969941000033110 | 12538811000001104 | Hydrochlorothiazide |
| Losartan 100mg / Hydrochlorothiazide 12.5mg tablets | 4424741000033110 | 13112711000001103 | Hydrochlorothiazide/ Losartan potassium |
| Cozaar-Comp 100mg/12.5mg tablets (Merck Sharp & Dohme Ltd) | 4424841000033117 | 13094111000001102 | Hydrochlorothiazide/ Losartan potassium |
| MicardisPlus 80mg/25mg tablets (Boehringer Ingelheim Ltd) | 4548841000033118 | 13719711000001103 | Hydrochlorothiazide/ Telmisartan |
| Telmisartan 80mg / Hydrochlorothiazide 25mg tablets | 4548741000033111 | 13731911000001109 | Hydrochlorothiazide/ Telmisartan |
| Actelsar HCT 80mg/25mg tablets (Accord Healthcare Ltd) | 9159541000033117 | 24413611000001103 | Hydrochlorothiazide/ Telmisartan |
| Tolucombi 80mg/25mg tablets (Consilient Health Ltd) | 9292841000033115 | 24573611000001104 | Hydrochlorothiazide/ Telmisartan |
| Spironolactone 3mg / Chlorothiazide 30mg capsules | 9278541000033112 | 16072811000001103 | Chlorothiazide/ Spironolactone |
| Spironolactone 4mg / Chlorothiazide 40mg capsules | 7681541000033117 | 18520511000001107 | Chlorothiazide/ Spironolactone |
| Co-Betaloc SA tablets (Pfizer Ltd) | 372841000033114 | 3853411000001104 | Hydrochlorothiazide/ Metoprolol tartrate |
| Chlortalidone 50mg tablets | 3086241000033111 | 317935006 | Chlortalidone |
| Hygroton 50mg tablets (Alliance Pharmaceuticals Ltd) | 742841000033114 | 285911000001101 | Chlortalidone |
| Cilazapril 500microgram tablets | 258441000033119 | 318915008 | Cilazapril monohydrate |
| Vascace 500microgram tablets (Roche Products Ltd) | 1504941000033117 | 3740211000001108 | Cilazapril monohydrate |
| Vascace 1mg tablets (Roche Products Ltd) | 1504641000033112 | 3669711000001105 | Cilazapril monohydrate |
| Cilazapril 1mg tablets | 258141000033110 | 318916009 | Cilazapril monohydrate |
| Cilazapril 2.5mg tablets | 258241000033115 | 318917000 | Cilazapril monohydrate |
| Vascace 2.5mg tablets (Roche Products Ltd) | 1504741000033115 | 3672411000001101 | Cilazapril monohydrate |
| Vascace 5mg tablets (Roche Products Ltd) | 1503441000033113 | 3671311000001104 | Cilazapril monohydrate |
| Cilazapril 5mg tablets | 258641000033117 | 318923005 | Cilazapril monohydrate |
| Catapres 100microgram tablets (Boehringer Ingelheim Ltd) | 215341000033112 | 215111000001101 | Clonidine hydrochloride |
| Clonidine 100microgram tablets | 285841000033118 | 318667005 | Clonidine hydrochloride |

| | | | |
|---|---|---|---|
| Clonidine 300microgram tablets | 287241000033110 | 318668000 | Clonidine hydrochloride |
| Catapres 300microgram tablets (Boehringer Ingelheim Ltd) | 215441000033118 | 368711000001103 | Clonidine hydrochloride |
| Dixarit 25microgram tablets (Boehringer Ingelheim Ltd) | 464341000033111 | 344511000001105 | Clonidine hydrochloride |
| Clonidine 25microgram tablets | 285941000033114 | 322840006 | Clonidine hydrochloride |
| Clonidine 50micrograms/5ml oral solution | 5900141000033118 | 8398511000001100 | Clonidine hydrochloride |
| Clonidine 50micrograms/5ml oral suspension | 5900241000033113 | 8398611000001101 | Clonidine hydrochloride |
| Clonidine 100micrograms/5ml oral suspension | 12569041000033115 | 11813011000001109 | Clonidine hydrochloride |
| Catapres 150micrograms/1ml solution for injection ampoules (Boehringer Ingelheim Ltd) | 196241000033111 | 364911000001108 | Clonidine hydrochloride |
| Clonidine 150micrograms/1ml solution for injection ampoules | 275641000033110 | 36089211000001103 | Clonidine hydrochloride |
| Clonidine 250microgram modified-release capsules | 261841000033118 | 36089511000001100 | Clonidine hydrochloride |
| Clonidine 50micrograms/5ml oral solution sugar free | 12684041000033114 | 36392711000001102 | Clonidine hydrochloride |
| Aldactide 25 tablets (Pfizer Ltd) | 40241000033110 | 762511000001105 | Hydroflumethiazide/ Spironolactone |
| Co-flumactone 25mg/25mg tablets | 377041000033116 | 318127005 | Hydroflumethiazide/ Spironolactone |
| Co-flumactone 50mg/50mg tablets | 377141000033117 | 318128000 | Hydroflumethiazide/ Spironolactone |
| Aldactide 50 tablets (Pfizer Ltd) | 40341000033117 | 4669111000001107 | Hydroflumethiazide/ Spironolactone |
| Dytac 50mg capsules (Advanz Pharma) | 491641000033116 | 3907911000001100 | Triamterene |
| Triamterene 50mg capsules | 1455341000033115 | 318082004 | Triamterene |
| Triamterene 50mg / Benzthiazide 25mg capsules | 3161941000033119 | 318098006 | Benzthiazide/ Triamterene |
| Dytide capsules (Mercury Pharma Group Ltd) | 491741000033113 | 714911000001108 | Benzthiazide/ Triamterene |
| Kalspare tablets (DHP Healthcare Ltd) | 796541000033115 | 3252011000001105 | Chlortalidone/ Triamterene |
| Triamterene 50mg / Chlortalidone 50mg tablets | 3162341000033110 | 318100006 | Chlortalidone/ Triamterene |
| Triamterene 50mg / Furosemide 40mg tablets | 3092741000033119 | 318101005 | Furosemide/ Triamterene |
| Frusene 50mg/40mg tablets (Orion Pharma (UK) Ltd) | 615041000033112 | 25411000001108 | Furosemide/ Triamterene |
| Navidrex 500microgram tablets (Advanz Pharma) | 955441000033111 | 487811000001100 | Cyclopenthiazide |
| Cyclopenthiazide 500microgram tablets | 395041000033117 | 317940003 | Cyclopenthiazide |
| Trasidrex modified-release tablets (Mercury Pharma Group Ltd) | 1469941000033114 | 3444411000001107 | Cyclopenthiazide/ Oxprenolol hydrochloride |
| Co-prenozide 160mg/0.25mg modified-release tablets | 377341000033119 | 36091611000001101 | Cyclopenthiazide/ Oxprenolol hydrochloride |
| Pralenal 2.5 tablets (Opus Pharmaceuticals Ltd) | 1918641000033112 | 448611000001106 | Enalapril maleate |
| Innovace 2.5mg tablets (Merck Sharp & Dohme Ltd) | 769141000033117 | 749611000001107 | Enalapril maleate |
| Enalapril 2.5mg tablets | 523541000033115 | 318850001 | Enalapril maleate |
| Enalapril 5mg tablets | 522841000033111 | 318851002 | Enalapril maleate |
| Innovace 5mg tablets (Merck Sharp & Dohme Ltd) | 769341000033119 | 730211000001104 | Enalapril maleate |
| Pralenal 5 tablets (Opus Pharmaceuticals Ltd) | 1918741000033115 | 578511000001100 | Enalapril maleate |

| | | | |
|---|---|---|---|
| Innovace 10mg tablets (Merck Sharp & Dohme Ltd) | 769041000033116 | 316111000001104 | Enalapril maleate |
| Pralenal 10 tablets (Opus Pharmaceuticals Ltd) | 1918841000033113 | 761411000001101 | Enalapril maleate |
| Enalapril 10mg tablets | 522641000033110 | 318853004 | Enalapril maleate |
| Enalapril 20mg tablets | 522741000033118 | 318855006 | Enalapril maleate |
| Pralenal 20 tablets (Opus Pharmaceuticals Ltd) | 1918941000033117 | 508511000001105 | Enalapril maleate |
| Innovace 20mg tablets (Merck Sharp & Dohme Ltd) | 769241000033112 | 302311000001104 | Enalapril maleate |
| Enalapril 1.25mg/5ml oral solution | 6012441000033118 | 8485511000001103 | Enalapril maleate |
| Enalapril 1.25mg/5ml oral suspension | 6012541000033117 | 8485611000001104 | Enalapril maleate |
| Enalapril 10mg/5ml oral solution | 5967041000033117 | 8485711000001108 | Enalapril maleate |
| Enalapril 10mg/5ml oral suspension | 5967141000033118 | 8485811000001100 | Enalapril maleate |
| Enalapril 5mg/5ml oral solution | 5992541000033111 | 8486911000001103 | Enalapril maleate |
| Enalapril 5mg/5ml oral suspension | 5992641000033112 | 8487011000001104 | Enalapril maleate |
| Enalapril 5mg/5ml oral suspension sugar free | 3279241000033117 | 20092911000001108 | Enalapril maleate |
| Inspra 25mg tablets (Upjohn UK Ltd) | 3199041000033116 | 8477211000001104 | Eplerenone |
| Eplerenone 25mg tablets | 3198841000033117 | 407011007 | Eplerenone |
| Eplerenone 50mg tablets | 3198941000033113 | 407012000 | Eplerenone |
| Inspra 50mg tablets (Upjohn UK Ltd) | 3199141000033117 | 8479811000001106 | Eplerenone |
| Teveten 300mg tablets (Mylan) | 2040841000033113 | 401211000001105 | Eprosartan mesilate |
| Eprosartan 300mg tablets | 2036341000033115 | 318994006 | Eprosartan mesilate |
| Eprosartan 400mg tablets | 2036441000033114 | 318995007 | Eprosartan mesilate |
| Teveten 400mg tablets (Abbott Healthcare Products Ltd) | 2040941000033117 | 151411000001103 | Eprosartan mesilate |
| Teveten 600mg tablets (Mylan) | 2041041000033110 | 872011000001109 | Eprosartan mesilate |
| Eprosartan 600mg tablets | 2036541000033110 | 318996008 | Eprosartan mesilate |
| Felodipine 2.5mg modified-release / Ramipril 2.5mg tablets | 3163341000033119 | 318176004 | Felodipine/ Ramipril |
| Triapin 2.5mg/2.5mg modified-release tablets (Sanofi) | 8264941000033119 | 4093211000001109 | Felodipine/ Ramipril |
| Triapin 5mg/5mg modified-release tablets (Sanofi) | 1834341000033110 | 3887911000001109 | Felodipine/ Ramipril |
| Felodipine 5mg modified-release / Ramipril 5mg tablets | 3163241000033112 | 318177008 | Felodipine/ Ramipril |
| Plendil 2.5mg modified-release tablets (AstraZeneca UK Ltd) | 1093941000033116 | 562711000001104 | Felodipine |
| Cabren 2.5mg modified-release tablets (Teva UK Ltd) | 2928841000033117 | 3800311000001106 | Felodipine |
| Neofel XL 2.5mg tablets (Actavis UK Ltd) | 4521541000033113 | 18167311000001103 | Felodipine |
| Felotens XL 2.5mg tablets (Thornton & Ross Ltd) | 4429241000033115 | 13127311000001107 | Felodipine |
| Folpik XL 2.5mg tablets (Teva UK Ltd) | 5576441000033112 | 13565311000001101 | Felodipine |
| Cardioplen XL 2.5mg tablets (Chiesi Ltd) | 4152841000033112 | 11506711000001103 | Felodipine |
| Felodipine 2.5mg modified-release tablets | 568041000033111 | 39020311000001104 | Felodipine |
| Parmid XL 2.5mg tablets (Sandoz Ltd) | 9121541000033117 | 24221811000001105 | Felodipine |
| Plendil 10mg modified-release tablets (AstraZeneca UK Ltd) | 1094041000033119 | 48511000001101 | Felodipine |
| Cabren 10mg modified-release tablets (Teva UK Ltd) | 2929041000033116 | 3800711000001105 | Felodipine |
| Felogen XL 10mg tablets (Mylan) | 3177541000033111 | 4973011000001100 | Felodipine |
| Folpik XL 10mg tablets (Teva UK Ltd) | 5576341000033118 | 5008911000001100 | Felodipine |

| | | | |
|---|---|---|---|
| Felotens XL 10mg tablets (Thornton & Ross Ltd) | 2980041000033115 | 4785511000001107 | Felodipine |
| Vascalpha 10mg modified-release tablets (Accord Healthcare Ltd) | 3034441000033111 | 5638811000001106 | Felodipine |
| Parmid XL 10mg tablets (Sandoz Ltd) | 9121741000033113 | 7388311000001103 | Felodipine |
| Neofel XL 10mg tablets (Kent Pharmaceuticals Ltd) | 4521741000033117 | 8089811000001107 | Felodipine |
| Cardioplen XL 10mg tablets (Chiesi Ltd) | 3154941000033110 | 7887511000001107 | Felodipine |
| Felodipine 10mg modified-release tablets | 569041000033115 | 39020511000001105 | Felodipine |
| Cardioplen XL 5mg tablets (Chiesi Ltd) | 3154841000033119 | 7887011000001104 | Felodipine |
| Neofel XL 5mg tablets (Kent Pharmaceuticals Ltd) | 4521641000033114 | 8090111000001106 | Felodipine |
| Parmid XL 5mg tablets (Sandoz Ltd) | 9121641000033116 | 7387911000001103 | Felodipine |
| Vascalpha 5mg modified-release tablets (Accord Healthcare Ltd) | 3034341000033117 | 5638311000001102 | Felodipine |
| Felotens XL 5mg tablets (Thornton & Ross Ltd) | 2979941000033119 | 4785111000001103 | Felodipine |
| Folpik XL 5mg tablets (Teva UK Ltd) | 5576541000033113 | 5008511000001107 | Felodipine |
| Felogen XL 5mg tablets (Mylan) | 3177441000033110 | 4972811000001103 | Felodipine |
| Cabren 5mg modified-release tablets (Teva UK Ltd) | 2928941000033113 | 3800511000001100 | Felodipine |
| Plendil 5mg modified-release tablets (AstraZeneca UK Ltd) | 1094141000033115 | 490211000001101 | Felodipine |
| Felodipine 5mg modified-release tablets | 569141000033116 | 39020611000001109 | Felodipine |
| Fosinopril 10mg tablets | 609141000033111 | 318909008 | Fosinopril sodium |
| Staril 10mg tablets (Bristol-Myers Squibb Pharmaceuticals Ltd) | 1387341000033114 | 462511000001104 | Fosinopril sodium |
| Staril 20mg tablets (Bristol-Myers Squibb Pharmaceuticals Ltd) | 1387441000033115 | 348111000001107 | Fosinopril sodium |
| Fosinopril 20mg tablets | 609241000033116 | 318910003 | Fosinopril sodium |
| Hydralazine 25mg tablets | 741941000033113 | 318649003 | Hydralazine hydrochloride |
| Apresoline 25mg tablets (Advanz Pharma) | 74341000033114 | 657011000001101 | Hydralazine hydrochloride |
| Hydralazine 50mg tablets | 742041000033119 | 318650003 | Hydralazine hydrochloride |
| Hydralazine 10mg tablets | 12877141000033119 | 414426001 | Hydralazine hydrochloride |
| Apo-Hydralazine 10mg tablets (Imported (United States)) | 12877241000033114 | 10637411000001100 | Hydralazine hydrochloride |
| Hydralazine 10mg/5ml oral suspension | 4456241000033111 | 8528611000001105 | Hydralazine hydrochloride |
| Hydralazine 25mg/5ml oral solution | 11072741000033116 | 8580911000001106 | Hydralazine hydrochloride |
| Apresoline 20mg powder for solution for injection ampoules (Advanz Pharma) | 73441000033117 | 3925011000001100 | Hydralazine hydrochloride |
| Hydralazine 20mg powder for solution for injection ampoules | 734141000033114 | 34193811000001100 | Hydralazine hydrochloride |
| Tanatril 20mg tablets (Mitsubishi Tanabe Pharma Europe Ltd) | 2145741000033116 | 533511000001106 | Imidapril hydrochloride |
| Imidapril 20mg tablets | 2145841000033114 | 318942009 | Imidapril hydrochloride |
| Imidapril 5mg tablets | 1739841000033119 | 318943004 | Imidapril hydrochloride |
| Tanatril 5mg tablets (Mitsubishi Tanabe Pharma Europe Ltd) | 1753941000033114 | 797911000001103 | Imidapril hydrochloride |
| Tanatril 10mg tablets (Mitsubishi Tanabe Pharma Europe Ltd) | 1753841000033118 | 680811000001102 | Imidapril hydrochloride |
| Imidapril 10mg tablets | 1739741000033112 | 318944005 | Imidapril hydrochloride |

| | | | |
|---|---|---|---|
| Indapamide 2.5mg tablets | 768641000033118 | 317956008 | Indapamide hemihydrate |
| Nindaxa 2.5 tablets (Ashbourne Pharmaceuticals Ltd) | 769741000033118 | 424311000001108 | Indapamide hemihydrate |
| Natrilix 2.5mg tablets (Servier Laboratories Ltd) | 955341000033117 | 321811000001109 | Indapamide hemihydrate |
| Coversyl Plus tablets (Servier Laboratories Ltd) | 2739541000033117 | 562511000001109 | Indapamide/ Perindopril erbumine |
| Perindopril erbumine 4mg / Indapamide 1.25mg tablets | 3161641000033114 | 3437611000001100 | Indapamide/ Perindopril erbumine |
| Perindopril arginine 5mg / Indapamide 1.25mg tablets | 4454541000033118 | 13454311000001101 | Indapamide/ Perindopril arginine |
| Coversyl Arginine Plus 5mg/1.25mg tablets (Servier Laboratories Ltd) | 4454641000033117 | 13444311000001104 | Indapamide/ Perindopril arginine |
| Indapamide 2.5mg/5ml oral suspension | 8197041000033112 | 21578911000001105 | Indapamide |
| Perindopril tosilate 5mg / Indapamide 1.25mg tablets | 8263641000033112 | 21940011000001101 | Indapamide/ Perindopril tosilate |
| Natrilix SR 1.5mg tablets (Servier Laboratories Ltd) | 953141000033119 | 456611000001108 | Indapamide |
| Rawel XL 1.5mg tablets (Consilient Health Ltd) | 5566041000033114 | 16737911000001105 | Indapamide |
| Ethibide XL 1.5mg tablets (Genus Pharmaceuticals Ltd) | 4571241000033110 | 13824811000001102 | Indapamide |
| Tensaid XL 1.5mg tablets (Mylan) | 4656741000033119 | 14242211000001100 | Indapamide |
| Varbim XL 1.5mg tablets (Teva UK Ltd) | 5816741000033112 | 14693611000001102 | Indapamide |
| Indipam XL 1.5mg tablets (Accord Healthcare Ltd) | 5053141000033113 | 15436111000001104 | Indapamide |
| Mapemid XL 1.5mg tablets (Teva UK Ltd) | 5128741000033110 | 15600711000001107 | Indapamide |
| Indapamide 1.5mg modified-release tablets | 765741000033116 | 39020711000001100 | Indapamide |
| Cardide SR 1.5mg tablets (Teva UK Ltd) | 9121841000033115 | 24331611000001104 | Indapamide |
| Alkapamid XL 1.5mg tablets (HBS Healthcare Ltd) | 12312641000033118 | 34494911000001106 | Indapamide |
| Lorvacs XL 1.5mg tablets (Torrent Pharma (UK) Ltd) | 12988441000033113 | 36812711000001103 | Indapamide |
| Indoramin 20mg tablets | 768541000033119 | 318739007 | Indoramin hydrochloride |
| Doralese Tiltab 20mg tablets (Chemidex Pharma Ltd) | 480541000033110 | 3354611000001100 | Indoramin hydrochloride |
| Baratol 25mg tablets (Amdipharm Plc) | 116741000033118 | 3689111000001107 | Indoramin hydrochloride |
| Indoramin 25mg tablets | 769841000033111 | 318740009 | Indoramin hydrochloride |
| Irbesartan 75mg tablets | 775641000033115 | 318968002 | Irbesartan |
| Aprovel 75mg tablets (Sanofi) | 76141000033117 | 434511000001104 | Irbesartan |
| Sabervel 75mg tablets (Aspire Pharma Ltd) | 8193941000033116 | 21522211000001102 | Irbesartan |
| Ifirmasta 75mg tablets (Consilient Health Ltd) | 8554141000033116 | 22720311000001108 | Irbesartan |
| Ifirmasta 150mg tablets (Consilient Health Ltd) | 8554241000033111 | 22720611000001103 | Irbesartan |
| Sabervel 150mg tablets (Aspire Pharma Ltd) | 8194041000033119 | 21522411000001103 | Irbesartan |
| Aprovel 150mg tablets (Sanofi) | 75941000033114 | 859711000001103 | Irbesartan |
| Irbesartan 150mg tablets | 775441000033117 | 318969005 | Irbesartan |
| Irbesartan 300mg tablets | 775541000033116 | 318970006 | Irbesartan |
| Aprovel 300mg tablets (Sanofi) | 76041000033116 | 323211000001107 | Irbesartan |
| Sabervel 300mg tablets (Aspire Pharma Ltd) | 8194141000033115 | 21522611000001100 | Irbesartan |

| | | | |
|---|---|---|---|
| Ifirmasta 300mg tablets (Consilient Health Ltd) | 8554341000033118 | 22720811000001104 | Irbesartan |
| Irbesartan 150mg/5ml oral suspension | 5970641000033117 | 8580811000001101 | Irbesartan |
| Irbesartan 300mg/5ml oral suspension | 4954341000033111 | 12639511000001103 | Irbesartan |
| Irbesartan 20mg oral powder sachets | 6167841000033112 | 18852211000001107 | Irbesartan |
| Irbesartan 30mg oral powder sachets | 6423941000033116 | 19281511000001102 | Irbesartan |
| Irbesartan 37.5mg oral powder sachets | 6435741000033117 | 19481911000001105 | Irbesartan |
| Prescal 2.5mg tablets (Novartis Pharmaceuticals UK Ltd) | 1140241000033117 | 3689711000001108 | Isradipine |
| Isradipine 2.5mg tablets | 784641000033112 | 319280009 | Isradipine |
| Lacidipine 2mg tablets | 817541000033119 | 319300008 | Lacidipine |
| Motens 2mg tablets (GlaxoSmithKline UK Ltd) | 937041000033114 | 910911000001108 | Lacidipine |
| Motens 4mg tablets (GlaxoSmithKline UK Ltd) | 937141000033113 | 333011000001100 | Lacidipine |
| Lacidipine 4mg tablets | 817641000033118 | 319301007 | Lacidipine |
| Molap 4mg tablets (Rivopharm (UK) Ltd) | 10217641000033116 | 28882211000001100 | Lacidipine |
| Lacidipine 6mg tablets | 13344441000033112 | 38019311000001102 | Lacidipine |
| Lercanidipine 10mg tablets | 825941000033112 | 319316005 | Lercanidipine hydrochloride |
| Zanidip 10mg tablets (Recordati Pharmaceuticals Ltd) | 1550841000033111 | 20011000001105 | Lercanidipine hydrochloride |
| Lercanidipine 20mg tablets | 3908741000033114 | 10225911000001102 | Lercanidipine hydrochloride |
| Zanidip 20mg tablets (Recordati Pharmaceuticals Ltd) | 3908841000033116 | 10198711000001102 | Lercanidipine hydrochloride |
| Zestril 2.5mg tablets (AstraZeneca UK Ltd) | 1552041000033115 | 593111000001108 | Lisinopril |
| Carace 2.5mg tablets (Bristol-Myers Squibb Pharmaceuticals Ltd) | 216041000033118 | 321011000001103 | Lisinopril |
| Lisinopril 2.5mg tablets | 837941000033119 | 318857003 | Lisinopril |
| Lisinopril 5mg tablets | 838141000033117 | 318858008 | Lisinopril |
| Carace 5mg tablets (Bristol-Myers Squibb Pharmaceuticals Ltd) | 216241000033114 | 778011000001109 | Lisinopril |
| Zestril 5mg tablets (AstraZeneca UK Ltd) | 1552241000033111 | 823211000001109 | Lisinopril |
| Zestril 10mg tablets (AstraZeneca UK Ltd) | 1551941000033114 | 825311000001100 | Lisinopril |
| Carace 10mg tablets (Bristol-Myers Squibb Pharmaceuticals Ltd) | 215941000033111 | 315211000001104 | Lisinopril |
| Lisinopril 10mg tablets | 837841000033110 | 318859000 | Lisinopril |
| Lisinopril 20mg tablets | 838041000033116 | 318860005 | Lisinopril |
| Carace 20mg tablets (Bristol-Myers Squibb Pharmaceuticals Ltd) | 216141000033119 | 56711000001109 | Lisinopril |
| Zestril 20mg tablets (AstraZeneca UK Ltd) | 1552141000033116 | 891711000001107 | Lisinopril |
| Lisinopril 2.5mg/5ml oral solution | 5971341000033117 | 8622111000001109 | Lisinopril |
| Lisinopril 2.5mg/5ml oral suspension | 5971441000033111 | 8622211000001103 | Lisinopril |
| Lisinopril 20mg/5ml oral solution | 6044641000033119 | 8622311000001106 | Lisinopril |
| Lisinopril 20mg/5ml oral suspension | 6044541000033115 | 8622411000001104 | Lisinopril |
| Lisinopril 5mg/5ml oral solution | 5890241000033117 | 8622511000001100 | Lisinopril |
| Lisinopril 5mg/5ml oral suspension | 3152141000033115 | 8622611000001101 | Lisinopril |
| Lisinopril 7.5mg/5ml oral solution | 5971541000033112 | 8622811000001102 | Lisinopril |
| Lisinopril 7.5mg/5ml oral suspension | 5971641000033113 | 8622911000001107 | Lisinopril |
| Lisinopril 40mg/5ml oral suspension | 5568741000033113 | 20556011000001103 | Lisinopril |

| | | | |
|---|---|---|---|
| Lisinopril 5mg/5ml oral solution sugar free | 10645041000033114 | 30251811000001106 | Lisinopril |
| Losartan 25mg tablets | 851841000033117 | 318955005 | Losartan potassium |
| Cozaar 25mg tablets (Merck Sharp & Dohme Ltd) | 370441000033112 | 266511000001104 | Losartan potassium |
| Cozaar 50mg tablets (Merck Sharp & Dohme Ltd) | 370541000033113 | 53611000001106 | Losartan potassium |
| Losartan 50mg tablets | 851941000033113 | 318956006 | Losartan potassium |
| Losartan 100mg tablets | 2720141000033115 | 407784004 | Losartan potassium |
| Cozaar 100mg tablets (Merck Sharp & Dohme Ltd) | 2720241000033110 | 245811000001102 | Losartan potassium |
| Losartan 50mg/5ml oral solution | 5971941000033118 | 14159411000001106 | Losartan potassium |
| Losartan 50mg/5ml oral suspension | 5972041000033112 | 14159511000001105 | Losartan potassium |
| Losartan 12.5mg tablets | 4957541000033114 | 15148111000001100 | Losartan potassium |
| Cozaar 12.5mg tablets (Merck Sharp & Dohme Ltd) | 4957641000033110 | 15138911000001101 | Losartan potassium |
| Losartan 100mg/5ml oral solution | 5971741000033116 | 15451111000001107 | Losartan potassium |
| Losartan 100mg/5ml oral suspension | 5971841000033114 | 15451211000001101 | Losartan potassium |
| Cozaar 2.5mg/ml oral suspension (Merck Sharp & Dohme Ltd) | 5150041000033110 | 15506811000001105 | Losartan potassium |
| Losartan 2.5mg/ml oral suspension sugar free | 5149941000033118 | 15507411000001105 | Losartan potassium |
| Methyldopa 125mg tablets | 897341000033110 | 318671008 | Methyldopa anhydrous |
| Methyldopa 250mg tablets | 897441000033116 | 318672001 | Methyldopa anhydrous |
| Aldomet 250mg tablets (Aspen Pharma Trading Ltd) | 40941000033118 | 253711000001107 | Methyldopa anhydrous |
| Aldomet 500mg tablets (Aspen Pharma Trading Ltd) | 41041000033111 | 73611000001108 | Methyldopa anhydrous |
| Methyldopa 500mg tablets | 897541000033115 | 318673006 | Methyldopa anhydrous |
| Methyldopa 250mg/5ml oral suspension | 892341000033112 | 8667311000001100 | Methyldopa anhydrous |
| Metenix 5mg tablets (Sanofi) | 896841000033116 | 375611000001109 | Metolazone |
| Metolazone 5mg tablets | 903541000033116 | 317965001 | Metolazone |
| Zaroxolyn 5mg tablets (Imported (Canada)) | 8196641000033116 | 21574611000001109 | Metolazone |
| Metolazone 2.5mg tablets | 8196441000033118 | 374173002 | Metolazone |
| Zaroxolyn 2.5mg tablets (Imported (Canada)) | 8196541000033117 | 21574911000001103 | Metolazone |
| Loniten 10mg tablets (Pfizer Ltd) | 850441000033112 | 3666711000001100 | Minoxidil |
| Minoxidil 10mg tablets | 920641000033119 | 318657000 | Minoxidil |
| Minoxidil 5mg tablets | 920941000033114 | 318656009 | Minoxidil |
| Loniten 5mg tablets (Pfizer Ltd) | 850641000033114 | 3666411000001106 | Minoxidil |
| Loniten 2.5mg tablets (Pfizer Ltd) | 850541000033113 | 3667011000001104 | Minoxidil |
| Minoxidil 2.5mg tablets | 920841000033118 | 318655008 | Minoxidil |
| Perdix 7.5mg tablets (UCB Pharma Ltd) | 1066041000033113 | 4041111000001109 | Moexipril hydrochloride |
| Moexipril 7.5mg tablets | 938141000033112 | 318934008 | Moexipril hydrochloride |
| Moexipril 15mg tablets | 938041000033113 | 318935009 | Moexipril hydrochloride |
| Perdix 15mg tablets (UCB Pharma Ltd) | 1065941000033115 | 4040311000001101 | Moexipril hydrochloride |
| Moxonidine 200microgram tablets | 938541000033115 | 318707000 | Moxonidine |
| Physiotens 200microgram tablets (Mylan) | 1080841000033119 | 41111000001102 | Moxonidine |

| | | | |
|---|---|---|---|
| Physiotens 400microgram tablets (Mylan) | 1080941000033110 | 142811000001107 | Moxonidine |
| Moxonidine 400microgram tablets | 938641000033119 | 318708005 | Moxonidine |
| Moxonidine 300microgram tablets | 2635641000033111 | 408604009 | Moxonidine |
| Physiotens 300microgram tablets (Mylan) | 2635741000033119 | 522011000001109 | Moxonidine |
| Cardene 20mg capsules (Astellas Pharma Ltd) | 176741000033114 | 344811000001108 | Nicardipine hydrochloride |
| Nicardipine 20mg capsules | 967741000033115 | 319217004 | Nicardipine hydrochloride |
| Nicardipine 30mg capsules | 967141000033119 | 319218009 | Nicardipine hydrochloride |
| Cardene 30mg capsules (Astellas Pharma Ltd) | 176841000033116 | 291111000001102 | Nicardipine hydrochloride |
| Nicardipine 10mg/10ml solution for infusion ampoules | 9697741000033114 | 27126711000001103 | Nicardipine hydrochloride |
| Cardene SR 30mg capsules (Astellas Pharma Ltd) | 198941000033118 | 540311000001105 | Nicardipine hydrochloride |
| Nicardipine 30mg modified-release capsules | 971441000033112 | 39021711000001108 | Nicardipine hydrochloride |
| Cardene SR 45mg capsules (Astellas Pharma Ltd) | 199041000033110 | 118811000001102 | Nicardipine hydrochloride |
| Nicardipine 45mg modified-release capsules | 971541000033113 | 39021811000001100 | Nicardipine hydrochloride |
| Adalat 5mg capsules (Bayer Plc) | 14141000033116 | 27111000001107 | Nifedipine |
| Nifedipine 5mg capsules | 967341000033116 | 319222004 | Nifedipine |
| Nifedipine 10mg capsules | 967241000033114 | 319223009 | Nifedipine |
| Adalat 10mg capsules (Bayer Plc) | 14041000033115 | 782511000001108 | Nifedipine |
| Angiopine 10 capsules (Ashbourne Pharmaceuticals Ltd) | 61841000033111 | 811311000001107 | Nifedipine |
| Nifedipine 10mg/5ml oral suspension | 5973741000033117 | 8670111000001106 | Nifedipine |
| Nifedipine 5mg/5ml oral suspension | 5056241000033112 | 8670311000001108 | Nifedipine |
| Nifedipine 20mg/ml oral drops | 2639341000033117 | 9096811000001100 | Nifedipine |
| Nifedipine 2.5mg/5ml oral suspension | 5402541000033112 | 12303311000001109 | Nifedipine |
| Coracten XL 60mg capsules (UCB Pharma Ltd) | 1725041000033113 | 3381511000001105 | Nifedipine |
| Nifedipine 60mg modified-release capsules | 1778841000033111 | 38896511000001101 | Nifedipine |
| Coracten SR 20mg capsules (UCB Pharma Ltd) | 364241000033112 | 389611000001101 | Nifedipine |
| Nifedipine 20mg modified-release capsules | 974141000033119 | 39022511000001106 | Nifedipine |
| Slofedipine XL 60 tablets (Zentiva) | 1753041000033113 | 630411000001107 | Nifedipine |
| Adalat LA 60mg tablets (Bayer Plc) | 17241000033117 | 235511000001104 | Nifedipine |
| Adipine XL 60mg tablets (Chiesi Ltd) | 3225541000033116 | 9049911000001105 | Nifedipine |
| Nimodrel XL 60mg tablets (Zurich Pharmaceuticals) | 3952741000033118 | 10189311000001108 | Nifedipine |
| Neozipine XL 60mg tablets (Kent Pharmaceuticals Ltd) | 5817541000033118 | 10751411000001101 | Nifedipine |
| Valni XL 60mg tablets (Zentiva) | 4452041000033116 | 13402111000001101 | Nifedipine |
| Adanif XL 60mg tablets (Advanz Pharma) | 5892041000033114 | 17666211000001101 | Nifedipine |
| Nifedipine 60mg modified-release tablets | 971341000033118 | 39022611000001105 | Nifedipine |
| Nidef 60mg modified-release tablets (Morningside Healthcare Ltd) | 12372441000033111 | 34685811000001103 | Nifedipine |
| Cardilate MR 10mg tablets (Teva UK Ltd) | 1580841000033116 | 25911000001100 | Nifedipine |
| Calchan MR 10 tablets (Ranbaxy (UK) Ltd) | 2295641000033116 | 627111000001104 | Nifedipine |
| Angiopine MR 10mg tablets (Ashbourne Pharmaceuticals Ltd) | 66141000033114 | 568911000001104 | Nifedipine |

| | | | |
|---|---|---|---|
| Adalat retard 10mg tablets (Bayer Plc) | 19441000033113 | 569011000001108 | Nifedipine |
| Tensipine MR 10 tablets (Genus Pharmaceuticals Ltd) | 1421241000033110 | 413111000001101 | Nifedipine |
| Adipine MR 10 tablets (Chiesi Ltd) | 18341000033114 | 833611000001109 | Nifedipine |
| Nifedipress MR 10 tablets (Dexcel-Pharma Ltd) | 1745041000033110 | 904011000001104 | Nifedipine |
| Nifedipine 10mg modified-release tablets | 971841000033110 | 39022711000001101 | Nifedipine |
| Cardilate MR 20mg tablets (IVAX Pharmaceuticals UK Ltd) | 199141000033114 | 905711000001103 | Nifedipine |
| Slofedipine 20mg tablets (Sterwin Medicines) | 1697641000033111 | 843411000001105 | Nifedipine |
| Adalat LA 20mg tablets (Bayer Plc) | 1766341000033113 | 881811000001100 | Nifedipine |
| Angiopine MR 20mg tablets (Ashbourne Pharmaceuticals Ltd) | 65241000033114 | 865011000001105 | Nifedipine |
| Hypolar Retard 20 tablets (Sandoz Ltd) | 738141000033119 | 677411000001108 | Nifedipine |
| Valni 20 Retard tablets (Tillomed Laboratories Ltd) | 2955041000033115 | 693311000001101 | Nifedipine |
| Tensipine MR 20 tablets (Genus Pharmaceuticals Ltd) | 1421341000033117 | 385611000001104 | Nifedipine |
| Nifedipress MR 20 tablets (Dexcel-Pharma Ltd) | 1745141000033114 | 619111000001101 | Nifedipine |
| Calchan MR 20 tablets (Ranbaxy (UK) Ltd) | 2295741000033113 | 17011000001100 | Nifedipine |
| Adalat retard 20mg tablets (Bayer Plc) | 19341000033119 | 5011000001109 | Nifedipine |
| Adipine MR 20 tablets (Chiesi Ltd) | 18241000033116 | 74111000001103 | Nifedipine |
| Nifopress Retard 20mg tablets (Advanz Pharma) | 2189541000033117 | 280811000001100 | Nifedipine |
| Coroday MR 20mg tablets (Mylan) | 2051241000033116 | 309611000001102 | Nifedipine |
| Nifedipine 20mg modified-release tablets | 970941000033113 | 39022811000001109 | Nifedipine |
| Coracten SR 10mg capsules (UCB Pharma Ltd) | 336641000033114 | 126411000001108 | Nifedipine |
| Nifedipine 10mg modified-release capsules | 971141000033116 | 39022911000001104 | Nifedipine |
| Coracten XL 30mg capsules (UCB Pharma Ltd) | 1724941000033113 | 162811000001100 | Nifedipine |
| Nifedipine 30mg modified-release capsules | 1778741000033118 | 39107511000001109 | Nifedipine |
| Fortipine LA 40 tablets (Advanz Pharma) | 605941000033110 | 188711000001108 | Nifedipine |
| Nifedipine 40mg modified-release tablets | 972441000033116 | 39107611000001108 | Nifedipine |
| Adalat LA 30mg tablets (Bayer Plc) | 17141000033112 | 2881311000001105 | Nifedipine |
| Hypolar XL 30 tablets (Sandoz Ltd) | 2776541000033111 | 2881811000001101 | Nifedipine |
| Slofedipine XL 30mg tablets (Zentiva) | 1752941000033115 | 2882011000001104 | Nifedipine |
| Nimodrel XL 30mg tablets (Zurich Pharmaceuticals) | 3952641000033110 | 10189111000001106 | Nifedipine |
| Adipine XL 30mg tablets (Chiesi Ltd) | 3225441000033117 | 9049711000001108 | Nifedipine |
| Neozipine XL 30mg tablets (Kent Pharmaceuticals Ltd) | 5817441000033119 | 10751211000001100 | Nifedipine |
| Valni XL 30mg tablets (Zentiva) | 4451941000033110 | 13401911000001109 | Nifedipine |
| Adanif XL 30mg tablets (Advanz Pharma) | 5891941000033115 | 17666011000001106 | Nifedipine |
| Nifedipine 30mg modified-release tablets | 971241000033111 | 39111711000001105 | Nifedipine |
| Nidef 30mg modified-release tablets (Morningside Healthcare Ltd) | 12372341000033117 | 34685211000001104 | Nifedipine |
| Nimotop 30mg tablets (Bayer Plc) | 977741000033114 | 3879211000001104 | Nimodipine |
| Nimodipine 30mg tablets | 977441000033119 | 323273000 | Nimodipine |
| Nimotop 0.02% solution for infusion 50ml vials (Bayer Plc) | 970841000033117 | 4387211000001100 | Nimodipine |
| Nimodipine 10mg/50ml solution for infusion vials | 970741000033110 | 36031711000001104 | Nimodipine |
| Syscor MR 10 tablets (Forest Laboratories UK Ltd) | 1401341000033110 | 3877011000001100 | Nisoldipine |

| | | | |
|---|---|---|---|
| Nisoldipine 10mg modified-release tablets | 972141000033112 | 36031811000001107 | Nisoldipine |
| Syscor MR 20 tablets (Forest Laboratories UK Ltd) | 1401441000033116 | 4070011000001100 | Nisoldipine |
| Nisoldipine 20mg modified-release tablets | 972241000033117 | 36031911000001102 | Nisoldipine |
| Syscor MR 30 tablets (Forest Laboratories UK Ltd) | 1401541000033115 | 4069211000001108 | Nisoldipine |
| Nisoldipine 30mg modified-release tablets | 972341000033110 | 36032011000001109 | Nisoldipine |
| Olmesartan medoxomil 20mg tablets | 2944641000033112 | 385542009 | Olmesartan medoxomil |
| Olmetec 20mg tablets (Daiichi Sankyo UK Ltd) | 2944941000033117 | 4624311000001103 | Olmesartan medoxomil |
| Olmetec 40mg tablets (Daiichi Sankyo UK Ltd) | 2945041000033117 | 4624611000001108 | Olmesartan medoxomil |
| Olmesartan medoxomil 40mg tablets | 2944741000033115 | 385543004 | Olmesartan medoxomil |
| Olmesartan medoxomil 10mg tablets | 2944541000033111 | 408055003 | Olmesartan medoxomil |
| Olmetec 10mg tablets (Daiichi Sankyo UK Ltd) | 2944841000033113 | 4624011000001101 | Olmesartan medoxomil |
| Olmesartan medoxomil 10mg/5ml oral suspension | 4273241000033118 | 14680711000001103 | Olmesartan medoxomil |
| Perindopril erbumine 2mg tablets | 1067841000033115 | 318896009 | Perindopril erbumine |
| Coversyl 2mg tablets (Servier Laboratories Ltd) | 377541000033114 | 48211000001104 | Perindopril erbumine |
| Coversyl 4mg tablets (Servier Laboratories Ltd) | 377641000033110 | 902211000001100 | Perindopril erbumine |
| Perindopril erbumine 4mg tablets | 1067941000033111 | 318897000 | Perindopril erbumine |
| Perindopril erbumine 8mg tablets | 2846741000033119 | 374667004 | Perindopril erbumine |
| Coversyl 8mg tablets (Servier Laboratories Ltd) | 2846641000033111 | 3803711000001101 | Perindopril erbumine |
| Perindopril erbumine 4mg/5ml oral suspension | 8276041000033117 | 8671311000001103 | Perindopril erbumine |
| Coversyl Arginine 10mg tablets (Servier Laboratories Ltd) | 4454441000033119 | 13444611000001109 | Perindopril arginine |
| Perindopril arginine 10mg tablets | 4454141000033110 | 13454111000001103 | Perindopril arginine |
| Perindopril arginine 2.5mg tablets | 4453941000033114 | 13454211000001109 | Perindopril arginine |
| Coversyl Arginine 2.5mg tablets (Servier Laboratories Ltd) | 4454241000033115 | 13445211000001108 | Perindopril arginine |
| Perindopril arginine 5mg tablets | 4454041000033111 | 13454411000001108 | Perindopril arginine |
| Coversyl Arginine 5mg tablets (Servier Laboratories Ltd) | 4454341000033113 | 13444911000001103 | Perindopril arginine |
| Perindopril erbumine 8mg/5ml oral solution | 8275941000033110 | 14057311000001102 | Perindopril erbumine |
| Perindopril erbumine 8mg/5ml oral suspension | 8276141000033118 | 14057411000001109 | Perindopril erbumine |
| Perindopril tosilate 10mg tablets | 8263541000033111 | 21939811000001102 | Perindopril tosilate |
| Perindopril tosilate 2.5mg tablets | 8263341000033116 | 21939911000001107 | Perindopril tosilate |
| Perindopril tosilate 5mg tablets | 8263441000033110 | 21940111000001100 | Perindopril tosilate |
| Perindopril erbumine 4mg/5ml oral solution | 8869441000033119 | 23471511000001106 | Perindopril erbumine |
| Polythiazide 1mg tablets | 1110141000033119 | 317967009 | Polythiazide |
| Nephril 1mg tablets (Pfizer Ltd) | 965441000033115 | 4549211000001100 | Polythiazide |
| Hypovase 500microgram tablets (Pfizer Ltd) | 743541000033118 | 321311000001100 | Prazosin hydrochloride |

| | | | |
|---|---|---|---|
| Prazosin 500microgram tablets | 1131341000033118 | 318767003 | Prazosin hydrochloride |
| Prazosin 1mg tablets | 1131441000033112 | 318768008 | Prazosin hydrochloride |
| Hypovase 1mg tablets (Pfizer Ltd) | 743641000033117 | 347411000001101 | Prazosin hydrochloride |
| Alphavase 1 tablets (Ashbourne Pharmaceuticals Ltd) | 37741000033118 | 934411000001100 | Prazosin hydrochloride |
| Alphavase 2 tablets (Ashbourne Pharmaceuticals Ltd) | 37841000033111 | 570011000001106 | Prazosin hydrochloride |
| Hypovase 2mg tablets (Pfizer Ltd) | 743741000033114 | 150911000001104 | Prazosin hydrochloride |
| Prazosin 2mg tablets | 1131541000033113 | 318769000 | Prazosin hydrochloride |
| Prazosin 5mg tablets | 1131641000033114 | 318770004 | Prazosin hydrochloride |
| Alphavase 5 tablets (Ashbourne Pharmaceuticals Ltd) | 37941000033115 | 74711000001102 | Prazosin hydrochloride |
| Prazosin 500micrograms/5ml oral solution | 11567441000033119 | 13078011000001104 | Prazosin hydrochloride |
| Accupro 5mg tablets (Pfizer Ltd) | 13441000033110 | 582611000001106 | Quinapril hydrochloride |
| Quinil 5mg tablets (Tillomed Laboratories Ltd) | 3283141000033118 | 9207711000001100 | Quinapril hydrochloride |
| Quinapril 5mg tablets | 1149241000033111 | 318885001 | Quinapril hydrochloride |
| Quinapril 10mg tablets | 1149341000033118 | 318886000 | Quinapril hydrochloride |
| Quinil 10mg tablets (Tillomed Laboratories Ltd) | 3283241000033113 | 9208111000001100 | Quinapril hydrochloride |
| Accupro 10mg tablets (Pfizer Ltd) | 13241000033114 | 231111000001106 | Quinapril hydrochloride |
| Accupro 20mg tablets (Pfizer Ltd) | 13341000033116 | 829111000001100 | Quinapril hydrochloride |
| Quinil 20mg tablets (Tillomed Laboratories Ltd) | 3283341000033115 | 9208411000001105 | Quinapril hydrochloride |
| Quinapril 20mg tablets | 1149441000033112 | 318887009 | Quinapril hydrochloride |
| Accupro 40mg tablets (Pfizer Ltd) | 11641000033111 | 86411000001109 | Quinapril hydrochloride |
| Quinil 40mg tablets (Tillomed Laboratories Ltd) | 3283441000033114 | 9208711000001104 | Quinapril hydrochloride |
| Quinapril 40mg tablets | 1149541000033113 | 318894007 | Quinapril hydrochloride |
| Ramipril 1.25mg capsules | 1151341000033117 | 318900007 | Ramipril |
| Tritace 1.25mg capsules (Aventis Pharma) | 1455541000033110 | 111611000001109 | Ramipril |
| Tritace 2.5mg capsules (Sanofi) | 1455641000033111 | 835411000001105 | Ramipril |
| Lopace 2.5mg capsules (Discovery Pharmaceuticals) | 3159741000033111 | 7948711000001102 | Ramipril |
| Ramipril 2.5mg capsules | 1151441000033111 | 318901006 | Ramipril |
| Ramipril 5mg capsules | 1151541000033112 | 318902004 | Ramipril |
| Lopace 5mg capsules (Discovery Pharmaceuticals) | 3159841000033118 | 7948911000001100 | Ramipril |
| Tritace 5mg capsules (Sanofi) | 1455741000033119 | 802311000001101 | Ramipril |
| Tritace 10mg capsules (Sanofi) | 1769941000033115 | 43711000001100 | Ramipril |
| Lopace 10mg capsules (Discovery Pharmaceuticals) | 3159941000033114 | 7949111000001105 | Ramipril |
| Ramipril 10mg capsules | 1769841000033111 | 318906001 | Ramipril |
| Ramipril 1.25mg tablets | 2989541000033115 | 408040007 | Ramipril |
| Tritace 1.25mg tablets (Sanofi) | 2989941000033114 | 5010511000001106 | Ramipril |

| | | | |
|---|---|---|---|
| Tritace 2.5mg tablets (Sanofi) | 2990041000033116 | 5010811000001109 | Ramipril |
| Ramipril 2.5mg tablets | 2989641000033119 | 408050008 | Ramipril |
| Ramipril 5mg tablets | 2989741000033111 | 408051007 | Ramipril |
| Tritace 5mg tablets (Sanofi) | 2990141000033117 | 5011111000001108 | Ramipril |
| Tritace 10mg tablets (Sanofi) | 2990241000033112 | 5011411000001103 | Ramipril |
| Ramipril 10mg tablets | 2989841000033118 | 408052000 | Ramipril |
| Ramipril 1.25mg/5ml oral solution | 5998241000033112 | 8720211000001109 | Ramipril |
| Ramipril 1.25mg/5ml oral suspension | 4152241000033113 | 8720311000001101 | Ramipril |
| Ramipril 10mg/5ml oral solution | 5887841000033118 | 8720411000001108 | Ramipril |
| Ramipril 10mg/5ml oral suspension | 5887941000033114 | 8720511000001107 | Ramipril |
| Ramipril 2.5mg/5ml oral solution | 5890541000033115 | 8720611000001106 | Ramipril |
| Ramipril 2.5mg/5ml oral suspension | 4805941000033114 | 8720711000001102 | Ramipril |
| Ramipril 5mg/5ml oral solution | 5890341000033110 | 8720811000001105 | Ramipril |
| Ramipril 5mg/5ml oral suspension | 2883341000033118 | 8720911000001100 | Ramipril |
| Ramipril 2.5mg/5ml oral solution sugar free | 6517041000033110 | 19877111000001100 | Ramipril |
| Sacubitril 24mg / Valsartan 26mg tablets | 10943641000033117 | 31142011000001103 | Sacubitril/ Valsartan |
| Entresto 24mg/26mg tablets (Novartis Pharmaceuticals UK Ltd) | 10943941000033112 | 31136811000001109 | Sacubitril/ Valsartan |
| Sacubitril 49mg / Valsartan 51mg tablets | 10943741000033114 | 31142111000001102 | Sacubitril/ Valsartan |
| Entresto 49mg/51mg tablets (Novartis Pharmaceuticals UK Ltd) | 10944041000033114 | 31136411000001107 | Sacubitril/ Valsartan |
| Sacubitril 97mg / Valsartan 103mg tablets | 10943841000033116 | 31142211000001108 | Sacubitril/ Valsartan |
| Entresto 97mg/103mg tablets (Novartis Pharmaceuticals UK Ltd) | 10944141000033113 | 31138011000001104 | Sacubitril/ Valsartan |
| Spironolactone 25mg tablets | 1374241000033118 | 318056008 | Spironolactone |
| Aldactone 25mg tablets (Pfizer Ltd) | 40541000033112 | 930511000001105 | Spironolactone |
| Aldactone 50mg tablets (Pfizer Ltd) | 40641000033113 | 921811000001103 | Spironolactone |
| Spironolactone 50mg tablets | 1374341000033111 | 318057004 | Spironolactone |
| Spironolactone 100mg tablets | 1374141000033113 | 318058009 | Spironolactone |
| Spirospare 100 tablets (Ashbourne Pharmaceuticals Ltd) | 1374541000033116 | 3411000001104 | Spironolactone |
| Aldactone 100mg tablets (Pfizer Ltd) | 40441000033111 | 421611000001100 | Spironolactone |
| Lasilactone 20mg/50mg capsules (Sanofi) | 808841000033119 | 3645811000001107 | Furosemide/ Spironolactone |
| Spironolactone 50mg / Furosemide 20mg capsules | 3162541000033115 | 318102003 | Furosemide/ Spironolactone |
| Spironolactone 5mg/5ml oral suspension | 1371141000033111 | 8726311000001103 | Spironolactone |
| Spironolactone 50mg/5ml oral suspension | 1371341000033114 | 8726411000001105 | Spironolactone |
| Spironolactone 25mg/5ml oral suspension | 1371041000033112 | 8726511000001109 | Spironolactone |
| Spironolactone 15mg/5ml oral suspension | 5975441000033113 | 8727011000001103 | Spironolactone |
| Spironolactone 10mg/5ml oral suspension | 1371241000033116 | 8727311000001100 | Spironolactone |
| Spironolactone 100mg/5ml oral suspension | 2011541000033114 | 8727611000001105 | Spironolactone |
| Spironolactone 250mg/5ml oral solution | 5975541000033114 | 13353511000001101 | Spironolactone |
| Spironolactone 250mg/5ml oral suspension | 5975641000033110 | 13353611000001102 | Spironolactone |
| Spironolactone 100mg/5ml oral solution | 5999141000033111 | 13894611000001109 | Spironolactone |

| | | | |
|---|---|---|---|
| Spironolactone 10mg/5ml oral solution | 5999241000033116 | 13894711000001100 | Spironolactone |
| Spironolactone 25mg/5ml oral solution | 5890141000033112 | 13894811000001108 | Spironolactone |
| Spironolactone 50mg/5ml oral solution | 5890441000033116 | 13894911000001103 | Spironolactone |
| Spironolactone 5mg/5ml oral solution | 5999341000033114 | 13895011000001103 | Spironolactone |
| Telmisartan 20mg tablets | 2272641000033119 | 134463001 | Telmisartan |
| Micardis 20mg tablets (Boehringer Ingelheim Ltd) | 2272741000033111 | 648711000001100 | Telmisartan |
| Tolura 20mg tablets (Consilient Health Ltd) | 9209341000033116 | 24555211000001104 | Telmisartan |
| Micardis 40mg tablets (Boehringer Ingelheim Ltd) | 1849741000033114 | 924911000001106 | Telmisartan |
| Telmisartan 40mg tablets | 1849541000033118 | 318986004 | Telmisartan |
| Tolura 40mg tablets (Consilient Health Ltd) | 9209441000033110 | 24555411000001100 | Telmisartan |
| Telmisartan 80mg tablets | 1849641000033117 | 318987008 | Telmisartan |
| Micardis 80mg tablets (Boehringer Ingelheim Ltd) | 1849841000033116 | 527411000001102 | Telmisartan |
| Tolura 80mg tablets (Consilient Health Ltd) | 9209541000033111 | 24555611000001102 | Telmisartan |
| Hytrin 5mg tablets (Advanz Pharma) | 745841000033115 | 3154311000001103 | Terazosin hydrochloride |
| Terazosin 5mg tablets | 1429141000033119 | 318776005 | Terazosin hydrochloride |
| Benph 5mg tablets (Mylan) | 11807841000033119 | 33628611000001102 | Terazosin hydrochloride |
| Terazosin 10mg tablets | 1429041000033118 | 318777001 | Terazosin hydrochloride |
| Hytrin 10mg tablets (Advanz Pharma) | 745641000033116 | 3150911000001107 | Terazosin hydrochloride |
| Hytrin 2mg tablets (Advanz Pharma) | 745741000033113 | 3147611000001100 | Terazosin hydrochloride |
| Terazosin 2mg tablets | 1425341000033119 | 318779003 | Terazosin hydrochloride |
| Benph 2mg tablets (Mylan) | 11799841000033112 | 33630711000001107 | Terazosin hydrochloride |
| Trandolapril 500microgram capsules | 1454141000033119 | 318924004 | Trandolapril |
| Gopten 500microgram capsules (Abbott Laboratories Ltd) | 648541000033117 | 253511000001102 | Trandolapril |
| Odrik 500microgram capsules (Aventis Pharma) | 998741000033119 | 227511000001106 | Trandolapril |
| Gopten 1mg capsules (Abbott Laboratories Ltd) | 648341000033112 | 346811000001101 | Trandolapril |
| Odrik 1mg capsules (Aventis Pharma) | 998541000033110 | 432911000001102 | Trandolapril |
| Trandolapril 1mg capsules | 1453941000033115 | 318925003 | Trandolapril |
| Trandolapril 2mg capsules | 1454041000033118 | 318926002 | Trandolapril |
| Gopten 2mg capsules (Abbott Laboratories Ltd) | 648441000033118 | 273111000001109 | Trandolapril |
| Odrik 2mg capsules (Aventis Pharma) | 998641000033111 | 140511000001108 | Trandolapril |
| Gopten 4mg capsules (Abbott Laboratories Ltd) | 3014841000033118 | 5651611000001103 | Trandolapril |
| Trandolapril 4mg capsules | 3014741000033111 | 410958005 | Trandolapril |
| Tarka modified-release capsules (Abbott Laboratories Ltd) | 1406341000033117 | 3691211000001101 | Trandolapril/ Verapamil hydrochloride |
| Verapamil 180mg modified-release / Trandolapril 2mg capsules | 3163041000033116 | 36149211000001102 | Trandolapril/ Verapamil hydrochloride |
| Trimetazidine 20mg tablets | 7747541000033112 | 15243911000001109 | Trimetazidine dihydrochloride |
| Trimetazidine 35mg modified-release tablets | 10495841000033113 | 20456111000001105 | Trimetazidine dihydrochloride |

| | | | |
|---|---|---|---|
| Vastarel MR 35mg tablets (Imported (France)) | 5492841000033111 | 20434311000001100 | Trimetazidine dihydrochloride |
| Diovan 40mg capsules (Novartis Pharmaceuticals UK Ltd) | 437741000033110 | 777611000001101 | Valsartan |
| Valsartan 40mg capsules | 1498341000033118 | 318961008 | Valsartan |
| Valsartan 80mg capsules | 1498441000033112 | 318962001 | Valsartan |
| Diovan 80mg capsules (Novartis Pharmaceuticals UK Ltd) | 437841000033117 | 554511000001105 | Valsartan |
| Diovan 160mg capsules (Novartis Pharmaceuticals UK Ltd) | 437641000033118 | 117011000001107 | Valsartan |
| Valsartan 160mg capsules | 1498241000033111 | 318963006 | Valsartan |
| Valsartan 80mg tablets | 6515441000033113 | 375034009 | Valsartan |
| Valsartan 160mg tablets | 6515541000033114 | 375035005 | Valsartan |
| Valsartan 320mg tablets | 4424941000033113 | 376487009 | Valsartan |
| Diovan 320mg tablets (Novartis Pharmaceuticals UK Ltd) | 4425041000033113 | 13143311000001102 | Valsartan |
| Valsartan 40mg tablets | 3201341000033115 | 416515008 | Valsartan |
| Diovan 40mg tablets (Novartis Pharmaceuticals UK Ltd) | 3201441000033114 | 8263211000001101 | Valsartan |
| Valsartan 3mg/ml oral solution | 6528441000033113 | 20007411000001100 | Valsartan |
| Diovan 3mg/1ml oral solution (Novartis Pharmaceuticals UK Ltd) | 6528541000033114 | 20001711000001102 | Valsartan |
| Diurexan 20mg tablets (Mylan) | 470741000033113 | 348911000001105 | Xipamide |
| Xipamide 20mg tablets | 1543941000033111 | 317970008 | Xipamide |
| Doxazosin 1mg tablets | 480641000033111 | 318781001 | Doxazosin mesilate |
| Cardura 1mg tablets (Upjohn UK Ltd) | 216741000033115 | 907711000001109 | Doxazosin mesilate |
| Doxadura 1mg tablets (Dexcel-Pharma Ltd) | 2957941000033110 | 4857511000001108 | Doxazosin mesilate |
| Doxadura 2mg tablets (Dexcel-Pharma Ltd) | 2958041000033113 | 4857711000001103 | Doxazosin mesilate |
| Cascor 2mg tablets (Ranbaxy (UK) Ltd) | 2760041000033110 | 904511000001107 | Doxazosin mesilate |
| Cardura 2mg tablets (Upjohn UK Ltd) | 216841000033113 | 41811000001109 | Doxazosin mesilate |
| Doxazosin 2mg tablets | 480741000033119 | 318782008 | Doxazosin mesilate |
| Doxazosin 4mg tablets | 480841000033112 | 318783003 | Doxazosin mesilate |
| Cascor 4mg tablets (Ranbaxy (UK) Ltd) | 2760141000033114 | 179311000001107 | Doxazosin mesilate |
| Doxadura 4mg tablets (Dexcel-Pharma Ltd) | 2958141000033112 | 4858111000001103 | Doxazosin mesilate |
| Doxazosin 8mg tablets | 12682141000033115 | 421069003 | Doxazosin mesilate |
| Doxazosin 1mg/5ml oral suspension | 4432941000033119 | 8483411000001101 | Doxazosin mesilate |
| Doxazosin 4mg/5ml oral suspension | 5138341000033111 | 8483511000001102 | Doxazosin mesilate |
| Doxazosin 4mg/5ml oral solution | 8870641000033119 | 23466611000001105 | Doxazosin mesilate |
| Doxazosin 1mg/5ml oral solution | 9173941000033111 | 24509711000001103 | Doxazosin mesilate |
| Oxandosin XL 4mg tablets (Ratiopharm UK Ltd) | 4288341000033111 | 11812211000001105 | Doxazosin mesilate |
| Colixil XL 4mg tablets (Sandoz Ltd) | 5403641000033117 | 11757711000001107 | Doxazosin mesilate |
| Doxadura XL 4mg tablets (Dexcel-Pharma Ltd) | 4063741000033112 | 11269911000001101 | Doxazosin mesilate |
| Slocinx XL 4mg tablets (Zentiva) | 4021141000033118 | 11098311000001104 | Doxazosin mesilate |
| Doxzogen XL 4mg tablets (Mylan) | 4575941000033115 | 13811611000001108 | Doxazosin mesilate |
| Cardozin XL 4mg tablets (Almus Pharmaceuticals Ltd) | 4936841000033112 | 18197411000001107 | Doxazosin mesilate |
| Raporsin XL 4mg tablets (Actavis UK Ltd) | 6036141000033119 | 18164911000001108 | Doxazosin mesilate |

| | | | |
|---|---|---|---|
| Larbex XL 4mg tablets (Teva UK Ltd) | 5809641000033111 | 17338211000001104 | Doxazosin mesilate |
| Cardura XL 4mg tablets (Upjohn UK Ltd) | 2274041000033118 | 123911000001106 | Doxazosin mesilate |
| Doxazosin 4mg modified-release tablets | 2273841000033111 | 39020411000001106 | Doxazosin mesilate |
| Cardura XL 8mg tablets (Upjohn UK Ltd) | 2274141000033119 | 873411000001109 | Doxazosin mesilate |
| Doxazosin 8mg modified-release tablets | 2273941000033115 | 39021111000001107 | Doxazosin mesilate |

#### Table S14. Heart failure selection Aurum codes

| Term | Medcode ID | SNOMED CT Concept ID | SNOMED CT Description ID |
|---|---|---|---|
| Acute left ventricular failure | 300190010 | 195114002 | 300190010 |
| Impaired left ventricular function | 411506018 | 275514001 | 411506018 |
| H/O: heart failure | 251680018 | 161505003 | 251680018 |
| Congestive heart failure due to valvular disease | 2675255018 | 426611007 | 2675255018 |
| Left ventricular cardiac dysfunction | 2694523019 | 429589006 | 2694523019 |
| CCF - Congestive cardiac failure | 3182551000006115 | 42343007 | 493288018 |
| Chronic pulmonary oedema | 494669012 | 46847001 | 494669012 |
| Acute congestive heart failure | 18472010 | 10633002 | 18472010 |
| COCM - Congestive cardiomyopathy | 6615881000006112 | 399020009 | 1786620011 |
| Hypertensive heart disease NOS with CCF | 741681000006111 | 64715009 | 107545013 |
| Congestive heart failure | 70653017 | 42343007 | 70653017 |
| Congestive cardiomyopathy | 1778488011 | 399020009 | 1778488011 |
| Cardiac failure | 139482012 | 84114007 | 139482012 |
| Heart failure with preserved ejection fraction | 7321121000006119 | 446221000 | 3496968011 |
| Congestive cardiac failure | 3182541000006117 | 42343007 | 493287011 |
| CCM - Congestive cardiomyopathy | 6615871000006114 | 399020009 | 1786619017 |
| Heart failure with preserved ejection fraction | 2227501000000110 | 446221000 | 2227501000000110 |
| Left ventricular failure | 141306010 | 85232009 | 141306010 |
| Chronic congestive heart failure | 147247018 | 88805009 | 147247018 |
| Heart failure NOS | 395772015 | 84114007 | 139475013 |
| HF - Heart failure | 3868341000006118 | 84114007 | 1234906013 |
| History of heart failure in last year | 5990971000006113 | 309634009 | 2986867013 |
| Pulmonary oedema NOS | 301694014 | 19242006 | 479262018 |
| Congestive obstructive cardiomyopathy | 350413012 | 233871002 | 350413012 |
| Biventricular failure | 4005301000006110 | 92506005 | 510016018 |
| Hypertensive heart&renal dis wth (congestive) heart failure | 741701000006114 | 194779001 | 299672017 |
| Left-sided heart failure | 3886061000006119 | 85232009 | 201199018 |
| Heart failure confirmed | 1488804017 | 395105005 | 1488804017 |
| Hyperten heart&renal dis+both(congestv)heart and renal fail | 789941000006117 | 194781004 | 299674016 |
| Decompensated cardiac failure | 300179017 | 195111005 | 300179017 |
| Compensated cardiac failure | 300180019 | 195112003 | 300180019 |
| Benign hypertensive heart disease with CCF | 504901000006118 | 194767001 | 299653017 |
| HFNEF - heart failure with normal ejection fraction | 1661371000000112 | 446221000 | 1713091000000115 |
| Severe left ventricular systolic dysfunction | 1991651000006115 | 1991651000006104 | 1991651000006115 |
| LVF - Left ventricular failure | 3886071000006114 | 85232009 | 1235017018 |
| Left heart failure | 3886041000006118 | 85232009 | 141303019 |
| Cardiac failure NOS | 223981000000118 | 84114007 | 139482012 |
| [RFC] Cardiac failure | 905391000006119 | 905391000006103 | 905391000006119 |
| Congestive cardiac failure | 493287011 | 42343007 | 70653017 |

| | | | |
|---|---|---|---|
| Acute heart failure | 94251011 | 56675007 | 94251011 |
| Post cardiac operation heart failure NOS | 300217019 | 195130005 | 300214014 |
| CHF - Congestive heart failure | 3182561000006118 | 42343007 | 493289014 |
| Heart failure | 139475013 | 84114007 | 139475013 |
| Heart failure with normal ejection fraction | 1647701000000118 | 446221000 | 2883808011 |
| Echocardiogram shows left ventricular systolic dysfunction | 2159197017 | 407596008 | 2159197017 |
| Echocardiogram shows left ventricular diastolic dysfunction | 2159198010 | 407597004 | 2159198010 |
| Pulmonary oedema - acute | 1490256017 | 40541001 | 492666016 |
| Malignant hypertensive heart disease with CCF | 728671000006119 | 83105008 | 1236017010 |
| H/O: Heart failure in last year | 453099015 | 309634009 | 453099015 |
| Biventricular failure | 510016018 | 92506005 | 153058012 |
| Impaired left ventricular function | 784191000006110 | 275514001 | 411506018 |
| History of heart failure | 4540521000006111 | 161505003 | 2986453013 |
| Malignant hypertensive heart disease with congestive heart failure | 3852501000006115 | 83105008 | 137848017 |
| Left ventricular systolic dysfunction | 216207010 | 134401001 | 216207010 |

#### Table S15. Loop Diuretics Aurum product codes

| Term from EMIS | Prod code ID | dmd ID | Drug substance name |
|---|---|---|---|
| Burinex 1mg tablets (LEO Pharma) | 172741000033113 | 846411000001100 | Bumetanide |
| Bumetanide 1mg tablets | 172441000033118 | 318021009 | Bumetanide |
| Bumetanide 5mg tablets | 172541000033117 | 318022002 | Bumetanide |
| Burinex 5mg tablets (LEO Pharma) | 172841000033115 | 521811000001107 | Bumetanide |
| Bumetanide 1mg/5ml oral solution sugar free | 169041000033112 | 318023007 | Bumetanide |
| Burinex A 5mg/1mg tablets (LEO Pharma) | 172341000033112 | 33911000001104 | Amiloride hydrochloride/ Bumetanide |
| Amiloride 5mg / Bumetanide 1mg tablets | 1845641000033119 | 318097001 | Amiloride hydrochloride/ Bumetanide |
| Bumetanide 5mg/5ml oral suspension | 10713941000033113 | 30798811000001102 | Bumetanide |
| Bumetanide 2mg/4ml solution for injection ampoules | 3858741000033112 | 35912911000001108 | Bumetanide |
| Burinex K modified-release tablets (LEO Pharma) | 172941000033111 | 3290211000001109 | Bumetanide/ Potassium chloride |
| Bumetanide 500microgram / Potassium chloride 573mg (potassium 7.7mmol) modified-release tablets | 1845741000033111 | 35913011000001100 | Bumetanide/ Potassium chloride |
| Furosemide 20mg tablets | 3092241000033113 | 317971007 | Furosemide |
| Lasix 20mg tablets (Borg Medicare) | 816341000033112 | 701611000001102 | Furosemide |
| Froop 40mg tablets (Ashbourne Pharmaceuticals Ltd) | 615541000033119 | 714311000001107 | Furosemide |
| Lasix 40mg tablets (Sanofi) | 816441000033118 | 79411000001107 | Furosemide |
| Frusid 40mg tablets (Dr Reddy's Laboratories (UK) Ltd) | 615141000033111 | 99311000001108 | Furosemide |
| Furosemide 40mg tablets | 3092341000033115 | 317972000 | Furosemide |
| Furosemide 500mg tablets | 3092441000033114 | 317973005 | Furosemide |
| Lasix 500mg tablets (Sanofi) | 816541000033117 | 829911000001102 | Furosemide |
| Diuresal 500mg tablets (Ennogen Pharma Ltd) | 9808241000033110 | 27992311000001106 | Furosemide |
| Lasix 5mg/5ml oral solution (Borg Medicare) | 812041000033114 | 701111000001105 | Furosemide |
| Triamterene 50mg / Furosemide 40mg tablets | 3092741000033119 | 318101005 | Furosemide/ Triamterene |
| Frusene 50mg/40mg tablets (Orion Pharma (UK) Ltd) | 615041000033112 | 25411000001108 | Furosemide/ Triamterene |
| Lasilactone 20mg/50mg capsules (Sanofi) | 808841000033119 | 3645811000001107 | Furosemide/ Spironolactone |
| Spironolactone 50mg / Furosemide 20mg capsules | 3162541000033115 | 318102003 | Furosemide/ Spironolactone |
| Frumil LS 20mg/2.5mg tablets (Sanofi) | 614341000033113 | 550711000001106 | Amiloride hydrochloride/ Furosemide |
| Co-amilofruse 2.5mg/20mg tablets | 376841000033113 | 318135008 | Amiloride hydrochloride/ Furosemide |
| Co-amilofruse 5mg/40mg tablets | 376641000033112 | 318136009 | Amiloride hydrochloride/ Furosemide |
| Frumil 40mg/5mg tablets (Sanofi) | 614641000033117 | 427411000001106 | Amiloride hydrochloride/ Furosemide |

| | | | |
|---|---|---|---|
| Froop Co 5mg/40mg tablets (Ashbourne Pharmaceuticals Ltd) | 1611441000033117 | 331311000001103 | Amiloride hydrochloride/ Furosemide |
| Lasoride 5mg/40mg tablets (Sanofi) | 818941000033112 | 678511000001106 | Amiloride hydrochloride/ Furosemide |
| Fru-Co 5mg/40mg tablets (Teva UK Ltd) | 615241000033116 | 818511000001106 | Amiloride hydrochloride/ Furosemide |
| Frumil Forte 10mg/80mg tablets (Sanofi) | 615641000033118 | 82611000001106 | Amiloride hydrochloride/ Furosemide |
| Co-amilofruse 10mg/80mg tablets | 376741000033115 | 318137000 | Amiloride hydrochloride/ Furosemide |
| Diumide-K Continus tablets (Teofarma) | 470541000033117 | 4540011000001102 | Furosemide/ Potassium chloride |
| Furosemide 40mg / Potassium chloride 600mg (potassium 8mmol) modified-release tablets | 3161841000033110 | 4557711000001102 | Furosemide/ Potassium chloride |
| Co-amilofruse 5mg/40mg/5ml oral suspension | 5710441000033117 | 8427011000001108 | Amiloride hydrochloride/ Furosemide |
| Furosemide 5mg/5ml oral solution | 3924941000033113 | 13893411000001108 | Furosemide |
| Furosemide 5mg/5ml oral suspension | 5993341000033112 | 13893511000001107 | Furosemide |
| Furosemide 250mg/25ml solution for injection ampoules | 3954441000033119 | 34193711000001108 | Furosemide |
| Lasikal modified-release tablets (Borg Medicare) | 816141000033114 | 3704811000001105 | Furosemide/ Potassium chloride |
| Furosemide 20mg / Potassium chloride 750mg (potassium 10mmol) modified-release tablets | 3162441000033116 | 36061311000001109 | Furosemide/ Potassium chloride |
| Lasix 20mg/2ml solution for injection ampoules (Sanofi) | 811841000033111 | 9611000001104 | Furosemide |
| Furosemide 20mg/2ml solution for injection ampoules | 3091841000033115 | 36061411000001102 | Furosemide |
| Furosemide 50mg/5ml solution for injection ampoules | 3091741000033113 | 36061511000001103 | Furosemide |
| Furosemide 80mg/8ml solution for injection Minijet pre-filled syringes (UCB Pharma Ltd) | 3092641000033111 | 2898811000001106 | Furosemide |
| Furosemide 80mg/8ml solution for injection pre-filled syringes | 3092541000033110 | 36061611000001104 | Furosemide |
| Frusol 50mg/5ml oral solution (Rosemont Pharmaceuticals Ltd) | 1731741000033111 | 855611000001109 | Furosemide |
| Furosemide 50mg/5ml oral solution sugar free | 3092141000033118 | 36564411000001102 | Furosemide |
| Frusol 40mg/5ml oral solution (Rosemont Pharmaceuticals Ltd) | 1731641000033119 | 494811000001105 | Furosemide |
| Furosemide 40mg/5ml oral solution sugar free | 3092041000033117 | 39021211000001101 | Furosemide |
| Frusol 20mg/5ml oral solution (Rosemont Pharmaceuticals Ltd) | 1731541000033115 | 602811000001100 | Furosemide |
| Furosemide 20mg/5ml oral solution sugar free | 3091941000033111 | 39108611000001107 | Furosemide |
| Torasemide 2.5mg tablets | 1450241000033116 | 318040003 | Torasemide |
| Torem 2.5mg tablets (Mylan) | 1449941000033115 | 3699711000001107 | Torasemide |
| Torem 5mg tablets (Mylan) | 1450041000033112 | 3700311000001109 | Torasemide |
| Torasemide 5mg tablets | 1450341000033114 | 318041004 | Torasemide |
| Torasemide 10mg tablets | 1450141000033111 | 318042006 | Torasemide |

| | | | |
|---|---|---|---|
| Torem 10mg tablets (Mylan) | 1449841000033111 | 3700811000001100 | Torasemide |

#### Table S16. Gastrointestinal and intracranial Bleeding Aurum codes

| Term | MedCode ID | SNOMED CT Concept ID | SNOMED CT Description ID |
|---|---|---|---|
| Cerebral haemorrhage | 884421000006119 | 274100004 | 884421000006119 |
| External capsule haemorrhage | 300276019 | 195167002 | 300276019 |
| Oesophageal varices with bleeding in diseases EC | 300776018 | 195475003 | 300776018 |
| Oesophageal varices without bleeding in diseases EC | 300779013 | 195476002 | 300779013 |
| H/O: GI Bleed | 411546012 | 275551007 | 411546012 |
| Cerebral infarction with haemorrhagic transformation | 1573101000006112 | 1573101000006108 | 1573101000006112 |
| H/O: upper GIT bleed | 456218012 | 312489007 | 456218012 |
| History of subarachnoid haemorrhage | 4540691000006112 | 161515009 | 2986875019 |
| H/O: GIT bleed | 5574481000006112 | 275551007 | 3036845016 |
| Duodenal ulcer with haemorrhage | 302534015 | 27281001 | 483490017 |
| Subarachnoid haemorrhage from middle cerebral artery | 123511000006114 | 21454007 | 36011016 |
| Subarachnoid haemorrhage from posterior communicating artery | 123521000006118 | 21454007 | 36011016 |
| Left sided intracerebral haemorrhage, unspecified | 748941000006115 | 274100004 | 409859018 |
| Traumatic subdural haematoma without open intracranial wound | 459511018 | 315046005 | 459511018 |
| Acute haemorrhagic pancreatitis | 493743018 | 4399003 | 493743018 |
| Chronic gastrojejunal ulcer with haemorrhage AND perforation | 494304013 | 45640006 | 494304013 |
| Internal capsule haemorrhage | 496232015 | 52201006 | 496232015 |
| Subarachnoid haemorrhage from basilar artery | 123491000006115 | 276284000 | 412361011 |
| Traumatic intracerebral haemorrhage | 7378421000006118 | 450418003 | 2915664016 |
| Unspecified duodenal ulcer with haemorrhage and perforation | 302537010 | 51868009 | 86362013 |
| Unspecified gastrojejunal ulcer with haemorrhage | 302606019 | 16121001 | 27283019 |
| Gastro-oesophageal laceration-haemorrhage syndrome | 3068361000006115 | 35265002 | 1216440014 |
| Intrapontine haemorrhage | 2622631000006114 | 7713009 | 503468012 |
| Bleeding internal haemorrhoids | 3733931000006110 | 75884004 | 200553019 |
| Evacuation of extradural haematoma | 649541000006113 | 171713001 | 265968017 |
| Evacuation of haematoma from temporal lobe of brain | 265689012 | 171473007 | 265689012 |
| Chronic peptic ulcer with haemorrhage AND perforation | 498977018 | 61300005 | 498977018 |
| Haematemesis | 507679012 | 8765009 | 507679012 |
| Acute gastric ulcer with haemorrhage | 508664015 | 89748001 | 508664015 |
| Oesophageal varices with bleeding | 1216060010 | 17709002 | 1216060010 |
| Subdural haematoma evacuation | 3469891000006117 | 59712006 | 498519016 |
| Cerebral haemorrhage | 989201000006117 | 274100004 | 989201000006117 |
| Evacuation of subdural haematoma | 3469851000006111 | 59712006 | 498516011 |
| Lobar cerebral haemorrhage | 345675012 | 230710000 | 345675012 |
| SAH - Subarachnoid haemorrhage | 2841141000006111 | 21454007 | 1216125018 |
| Gastrointestinal bleeding | 3710521000006115 | 74474003 | 123695010 |

| | | | |
|---|---|---|---|
| GI bleeding | 3710501000006113 | 74474003 | 123689014 |
| Oversew of blood vessel of duodenal ulcer | 362951000000115 | 226891000000104 | 362951000000115 |
| Subperiosteal haematoma | 933081000006112 | 669341000000103 | 933081000006112 |
| Subdural haemorrhage | 884441000006114 | 195176009 | 884441000006114 |
| DU - acute + haemorrhage | 885851000006113 | 12847006 | 885851000006113 |
| PU - acute + haemorrhage | 885941000006118 | 12274003 | 885941000006118 |
| PU - acute + h'ge + perf. | 885961000006119 | 111353003 | 885961000006119 |
| PU - chronic + haemorrhage | 885981000006112 | 49232000 | 885981000006112 |
| Upper GI - gastrointestinal haemorrhage | 3102201000006117 | 37372002 | 1216567017 |
| Right sided intracerebral haemorrhage, unspecified | 163261000006119 | 195168007 | 300277011 |
| Subarachnoid haemorrh from intracranial artery, unspecif | 123441000006112 | 21454007 | 481028017 |
| Intracerebral haemorrhage | 12223101000006118 | 274100004 | 3673215016 |
| Traumatic subdural haematoma | 5459261000006114 | 262952002 | 2913152010 |
| Extradural haemorrhage | 884431000006116 | 397809001 | 884431000006116 |
| Gastrointestinal bleed | 802131000006119 | 74474003 | 1233753013 |
| Intracerebral haemorrhage NOS | 300287010 | 274100004 | 2819959010 |
| Unspecified peptic ulcer with haemorrhage and perforation | 302571018 | 13200003 | 22592012 |
| Intracerebral haemorrhage with intraventricular haemorrhage | 4777861000006119 | 195168007 | 2915438018 |
| Intracranial haemorrhage following injury | 7378271000006116 | 450410005 | 2915426010 |
| Endoscopic injection haemostasis of duodenal ulcer | 459010010 | 314627002 | 459010010 |
| Oesophageal varices with bleeding, associated with another disorder | 4779721000006113 | 195475003 | 2575715017 |
| Non-traumatic subdural haemorrhage | 2475119012 | 195176009 | 2469010019 |
| Upper GI haemorrhage | 3102191000006115 | 37372002 | 486703016 |
| Evacuation of intracerebral haematoma | 265692011 | 10458001 | 279429014 |
| Gastric haemorrhage | 303660017 | 61401005 | 499003013 |
| Cerebral haemorrhage | 122371000006118 | 274100004 | 409860011 |
| [X]Other subarachnoid haemorrhage | 300936018 | 21454007 | 481028017 |
| Subarachnoid haemorrhage following injury with open intracranial wound | 320735017 | 5251007 | 496337012 |
| Traumatic subdural hematoma | 391035012 | 262952002 | 2912898019 |
| Subarachnoid haemorrhage from basilar artery aneurysm | 5583101000006116 | 276284000 | 412361011 |
| Bleeding gastric ulcer | 2752321000006114 | 15902003 | 1221244018 |
| Sequelae of intracerebral haemorrhage | 300407010 | 195241001 | 300407010 |
| Traumatic cranial subdural haematoma | 5459221000006115 | 262952002 | 391036013 |
| Oesophageal haemorrhage | 2741641000006118 | 15238002 | 477249012 |
| Intestinal haemorrhage NOS | 303663015 | 74474003 | 123688018 |
| Intracerebral haemorrhage, multiple localised | 4777881000006112 | 195169004 | 300280012 |
| Cerebral haemorrhage NOS | 989211000006119 | 274100004 | 989211000006119 |
| GU - acute + h'ge + perf. | 885791000006113 | 48974009 | 885791000006113 |
| GU - chronic + haemorrhage | 885811000006112 | 57246001 | 885811000006112 |
| GU - chronic + h'ge + perf. | 885831000006118 | 76181002 | 885831000006118 |

| | | | |
|---|---|---|---|
| PU - chronic + h'ge + perf. | 886001000006111 | 61300005 | 886001000006111 |
| Intracerebral haemorrhage (ICH) | 744901000006114 | 274100004 | 2819959010 |
| Cortical haemorrhage | 495394013 | 49422009 | 495394013 |
| Cerebellar haemorrhage | 502878012 | 75038005 | 502878012 |
| Bleeding chronic gastric ulcer | 1231704018 | 57246001 | 1231704018 |
| ICH - intracerebral haemorrhage | 12223121000006111 | 274100004 | 3673217012 |
| Bulbar haemorrhage | 483988011 | 732923001 | 3467313018 |
| Acute peptic ulcer with haemorrhage and perforation | 361430016 | 111353003 | 361430016 |
| Choroidal haemorrhage | 297943011 | 122003 | 1256014 |
| Chronic gastric ulcer with haemorrhage AND with perforation | 554311000006114 | 76181002 | 503219010 |
| Nontraumatic extradural haemorrhage | 660111000006117 | 397809001 | 1773153015 |
| Evacuation of haematoma from cerebellum | 265690015 | 171474001 | 265690015 |
| Gastrotomy and ligation of bleeding point of stomach | 269184010 | 173778009 | 269184010 |
| Melaena | 5913016 | 2901004 | 5913016 |
| Unspec peptic ulcer; unspec haemorrhage and/or perforation | 81501000006111 | 13200003 | 22592012 |
| Bleeding chronic duodenal ulcer | 1235462014 | 89469000 | 1235462014 |
| Gastrointestinal tract haemorrhage NOS | 303667019 | 74474003 | 123688018 |
| Subarachnoid haemorrhage from anterior communicating artery | 123481000006118 | 21454007 | 36011016 |
| Ruptured berry aneurysm | 300242011 | 195154000 | 300242011 |
| Subarachnoid haemorrhage from carotid siphon and bifurcation | 300244012 | 195155004 | 300244012 |
| Subarachnoid haemorrhage from vertebral artery | 300253017 | 195160000 | 300253017 |
| Subdural haemorrhage - nontraumatic | 300294013 | 195176009 | 300294013 |
| Sequelae of subarachnoid haemorrhage | 300406018 | 195240000 | 300406018 |
| Duodenal ulcer with perforation | 302535019 | 88968005 | 147516013 |
| Subdural haemorrhage | 2534198011 | 35486000 | 486181011 |
| Gastric ulcer with haemorrhage AND perforation | 81311000006115 | 62366003 | 499256017 |
| Corpus cavernosum haematoma | 304493010 | 198031007 | 304493010 |
| History of gastrointestinal bleed | 397836015 | 275551007 | 411546012 |
| GI - Gastrointestinal haemorrhage | 3710541000006110 | 74474003 | 1216914011 |
| Intracerebral haemorrhage in hemisphere, unspecified | 744921000006116 | 274100004 | 409860011 |
| Intracerebral haemorrhage, multiple localized | 746571000006116 | 195169004 | 300280012 |
| Cerebral haemorrhage | 300939013 | 274100004 | 409860011 |
| H/O: haematemesis | 251739019 | 161538007 | 251739019 |
| Cerebral haemorrhage NOS | 884451000006111 | 700251000000105 | 884451000006111 |
| Bleeding oesophageal varices | 2781011000006119 | 17709002 | 478405010 |
| Gastrointestinal hemorrhage | 396403016 | 74474003 | 502715010 |
| Unspec duodenal ulcer; unspec haemorrhage and/or perforation | 81301000006118 | 51868009 | 86362013 |
| Sequelae of other nontraumatic intracranial haemorrhage | 149551000006111 | 363302008 | 482447013 |
| DU - chronic + haemorrhage | 885891000006119 | 89469000 | 885891000006119 |

| | | | |
|---|---|---|---|
| Acute duodenal ulcer with haemorrhage | 474430017 | 12847006 | 474430017 |
| Basal ganglia haemorrhage | 503791000006114 | 195165005 | 300272017 |
| H/O: upper GIT haemorrhage | 456217019 | 312489007 | 456217019 |
| Acute peptic ulcer with haemorrhage | 470365013 | 12274003 | 470365013 |
| Gastric bleeding | 3497621000006112 | 61401005 | 102011018 |
| Intracerebral haemorrhage, intraventricular | 300277011 | 195168007 | 300277011 |
| Intracranial haemorrhage | 300298011 | 1386000 | 475553012 |
| Haemorrhage | 300840014 | 131148009 | 3035879011 |
| Subarachnoid haemorrhage NOS | 300257016 | 21454007 | 481028017 |
| Unspecified gastric ulcer with haemorrhage and perforation | 302490012 | 62366003 | 499256017 |
| Unspecified peptic ulcer with haemorrhage | 302568014 | 13200003 | 22592012 |
| Subarachnoid haemorrhage | 481028017 | 21454007 | 481028017 |
| [X]Intracerebral haemorrhage in hemisphere, unspecified | 300956017 | 274100004 | 409860011 |
| Pontine haemorrhage | 503469016 | 7713009 | 503469016 |
| Acute gastrojejunal ulcer with haemorrhage AND perforation | 504674011 | 81387001 | 504674011 |
| Cerebral haemorrhage following injury | 505322013 | 450418003 | 2916363013 |
| Traumatic cerebral haemorrhage | 505324014 | 450418003 | 2916058017 |
| Lower gastrointestinal haemorrhage | 507736019 | 87763006 | 507736019 |
| Evacuation of intracranial extradural haematoma | 4645641000006117 | 171713001 | 265968017 |
| Bleeding haemorrhoids | 300752014 | 51551000 | 198339016 |
| Gastric ulcer with haemorrhage | 302487018 | 15902003 | 477573015 |
| GI haemorrhage | 3710531000006117 | 74474003 | 502714014 |
| H/O subarachnoid haemorrhage | 808111000006110 | 161515009 | 251701010 |
| Traumatic subdural haematoma with open intracranial wound | 459514014 | 315047001 | 459514014 |
| Upper gastrointestinal haemorrhage | 486704010 | 37372002 | 486704010 |
| Acute gastric ulcer with haemorrhage AND perforation | 495273013 | 48974009 | 495273013 |
| Chronic gastric ulcer with haemorrhage | 497833010 | 57246001 | 497833010 |
| Evacuation of intracranial subdural haematoma | 498516011 | 59712006 | 498520010 |
| Oesophageal bleeding | 2741601000006115 | 15238002 | 477250012 |
| Other and unspecified intracranial haemorrhage | 300290016 | 62914000 | 104563015 |
| Epidural haemorrhage | 505381016 | 82999001 | 505381016 |
| Cerebral hemorrhage | 605471000006112 | 274100004 | 409859018 |
| Haemorrhage of oesophagus | 477248016 | 15238002 | 477248016 |
| Acute haemorrhagic gastritis | 478553011 | 2367005 | 481682019 |
| Chronic duodenal ulcer with haemorrhage AND perforation | 486596015 | 36975000 | 486596015 |
| Chronic peptic ulcer with haemorrhage | 495344012 | 49232000 | 495344012 |
| Acute gastrojejunal ulcer with haemorrhage | 499719010 | 63954007 | 499719010 |
| Gastrointestinal haemorrhage | 502715010 | 74474003 | 502715010 |
| Acute duodenal ulcer with haemorrhage AND perforation | 507303017 | 86895006 | 507303017 |
| Chronic gastrojejunal ulcer with haemorrhage | 499386010 | 62838000 | 499386010 |

| | | | |
|---|---|---|---|
| Chronic duodenal ulcer with haemorrhage | 508524014 | 89469000 | 508524014 |
| Bleeding acute gastric ulcer | 1235493010 | 89748001 | 1235493010 |
| Extradural haemorrhage following injury without open intracranial wound | 402930018 | 43216008 | 493552012 |
| Upper GI bleeding | 3102181000006118 | 37372002 | 62348012 |
| GU - acute + haemorrhage | 885771000006112 | 89748001 | 885771000006112 |
| History of upper gastrointestinal tract haemorrhage | 6017201000006118 | 312489007 | 2986370018 |

#### Table S17. Gastrointestinal tract ulcer Aurum codes

| Term | Medcode ID | SNOMED CT Concept ID | SNOMED CT Description ID |
|---|---|---|---|
| Acute duodenal ulcer with hemorrhage AND obstruction | 3926261000006112 | 87756006 | 145494013 |
| Acute gastrojejunal ulcer with hemorrhage AND perforation | 3823411000006113 | 81387001 | 135007015 |
| Acute gastrojejunal ulcer with perforation but without obstruction | 3677391000006113 | 72395008 | 120279015 |
| Chronic duodenal ulcer with hemorrhage | 3954241000006119 | 89469000 | 148344012 |
| Unspec gastrojejunal ulcer; unspec haemorrhage/perforation | 81331000006114 | 16121001 | 27283019 |
| Acute gastric ulcer with haemorrhage, with perforation AND with obstruction | 3365071000006116 | 53337006 | 496625019 |
| Duodenal ulcer with haemorrhage | 2937121000006115 | 27281001 | 2937121000006115 |
| Duodenal ulcer with haemorrhage | 302534015 | 27281001 | 483490017 |
| Gastrojejunal ulcer with hemorrhage but without obstruction | 3320581000006112 | 50663005 | 84434015 |
| Chronic gastrojejunal ulcer with haemorrhage, with perforation AND with obstruction | 2882871000006112 | 24001002 | 481784011 |
| Peptic ulcer with hemorrhage, with perforation AND with obstruction | 2919721000006119 | 26221006 | 43926010 |
| Chronic duodenal ulcer with perforation but without obstruction | 3057061000006111 | 34602004 | 57742016 |
| Perforated chronic gastric ulcer | 484836012 | 31301004 | 484836012 |
| Chronic gastrojejunal ulcer with haemorrhage AND perforation | 494304013 | 45640006 | 494304013 |
| Unspecified duodenal ulcer with haemorrhage and perforation | 302537010 | 51868009 | 86362013 |
| Unspecified peptic ulcer with perforation | 302569018 | 13200003 | 22592012 |
| Unspecified gastrojejunal ulcer with haemorrhage | 302606019 | 16121001 | 27283019 |
| Duodenal ulcer with hemorrhage | 11902881000006118 | 27281001 | 45608011 |
| Chronic peptic ulcer with haemorrhage AND perforation | 498977018 | 61300005 | 498977018 |
| Acute gastric ulcer with haemorrhage | 508664015 | 89748001 | 508664015 |
| Duodenal ulcer with hemorrhage but without obstruction | 3072881000006114 | 35560008 | 59330018 |
| Acute peptic ulcer with hemorrhage AND with perforation but without obstruction | 3260961000006111 | 47064007 | 78447013 |
| Acute peptic ulcer NOS | 12721881000006113 | 591741000000105 | 1312001000000116 |
| Chronic gastric ulcer with haemorrhage AND with obstruction | 3895591000006110 | 85859006 | 506779011 |
| DU - acute NOS | 885881000006117 | 618291000000105 | 885881000006117 |
| DU - acute + haemorrhage | 885851000006113 | 12847006 | 885851000006113 |
| DU - acute + perforation | 885861000006110 | 61347001 | 885861000006110 |
| DU - chronic + h'ge + perf. | 885911000006117 | 36975000 | 885911000006117 |
| PU - acute + haemorrhage | 885941000006118 | 12274003 | 885941000006118 |
| PU - acute + perforation | 885951000006116 | 79118000 | 885951000006116 |
| PU - acute + h'ge + perf. | 885961000006119 | 111353003 | 885961000006119 |
| PU - chronic + haemorrhage | 885981000006112 | 49232000 | 885981000006112 |
| Acute peptic ulcer with hemorrhage and perforation | 4194481000006117 | 111353003 | 90139013 |
| Gastrojejunal ulcer with haemorrhage but without obstruction | 3320571000006114 | 50663005 | 495743015 |

| | | | |
|---|---|---|---|
| Acute gastric ulcer with hemorrhage, with perforation AND with obstruction | 3365081000006118 | 53337006 | 88690011 |
| Gastrojejunal ulcer with haemorrhage AND with perforation but without obstruction | 3926841000006112 | 87796008 | 507754015 |
| Esophageal bleeding due to ulcerative esophagitis | 7222991000006116 | 439442003 | 2794960012 |
| Chronic duodenal ulcer with haemorrhage but without obstruction | 3513251000006119 | 62341002 | 499253013 |
| Acute duodenal ulcer with hemorrhage AND with perforation but without obstruction | 3340401000006119 | 51847008 | 86331014 |
| Peptic ulcer with hemorrhage | 3542821000006112 | 64121000 | 106588013 |
| Gastric ulcer with haemorrhage AND perforation but without obstruction | 2531561000006115 | 2066005 | 480818019 |
| Acute gastrojejunal ulcer with hemorrhage | 3539941000006116 | 63954007 | 106302011 |
| Chronic gastrojejunal ulcer with hemorrhage AND perforation | 3236981000006112 | 45640006 | 76108010 |
| GU - acute + perforation | 885781000006110 | 19850005 | 885781000006110 |
| GU - chronic + perforation | 885821000006116 | 31301004 | 885821000006116 |
| Acute duodenal ulcer with haemorrhage AND obstruction | 3926251000006110 | 87756006 | 507729015 |
| Acute gastric ulcer with perforation AND obstruction | 3203661000006112 | 43694004 | 72854019 |
| Unspecified peptic ulcer with haemorrhage and perforation | 302571018 | 13200003 | 22592012 |
| Unspecified gastrojejunal ulcer with perforation | 302608018 | 16121001 | 27283019 |
| Acute peptic ulcer with perforation | 131281018 | 79118000 | 131281018 |
| Chronic peptic ulcer with hemorrhage AND obstruction | 3417381000006115 | 56461008 | 93897011 |
| Acute gastric ulcer with hemorrhage AND perforation | 3292411000006119 | 48974009 | 81590012 |
| Chronic peptic ulcer with hemorrhage | 3296621000006114 | 49232000 | 82009014 |
| Acute duodenal ulcer with perforation but without obstruction | 2858711000006118 | 22511002 | 37788015 |
| Chronic gastrojejunal ulcer with hemorrhage but without perforation | 3463971000006114 | 59356009 | 98589012 |
| Acute peptic ulcer without hemorrhage, without perforation AND without obstruction | 2547461000006113 | 3023008 | 6131012 |
| Gastric ulcer with haemorrhage, with perforation AND with obstruction | 2779151000006110 | 17593008 | 478382011 |
| Chronic duodenal ulcer with haemorrhage AND with perforation but without obstruction | 3819871000006111 | 81142005 | 504625010 |
| Chronic gastric ulcer without haemorrhage, without perforation AND without obstruction | 2523571000006111 | 1567007 | 477462013 |
| Chronic gastric ulcer with haemorrhage AND with perforation but without obstruction | 3708391000006114 | 74341002 | 502673018 |
| Acute gastrojejunal ulcer with haemorrhage but without obstruction | 3466621000006113 | 59515005 | 498459015 |
| Duodenal ulcer with haemorrhage AND perforation | 2879911000006114 | 23812009 | 481717012 |
| Acute gastrojejunal ulcer with haemorrhage AND obstruction | 3677531000006115 | 72408002 | 502115017 |
| Acute gastrojejunal ulcer with haemorrhage AND with perforation but without obstruction | 3584201000006119 | 66673003 | 500495015 |
| Chronic peptic ulcer with hemorrhage but without obstruction | 3825521000006119 | 81518000 | 135230017 |
| Chronic peptic ulcer with haemorrhage AND with perforation but without obstruction | 3405521000006119 | 55746001 | 497387014 |
| Bleeding ulcer of oesophagus | 3438281000006118 | 57748001 | 497976013 |
| Gastric ulcer with hemorrhage, with perforation AND with obstruction | 2779161000006112 | 17593008 | 29732013 |

| | | | |
|---|---|---|---|
| Gastrojejunal ulcer with hemorrhage AND with perforation but without obstruction | 3926851000006114 | 87796008 | 145560014 |
| Acute peptic ulcer with perforation AND obstruction | 3074551000006112 | 35681000 | 59518011 |
| Chronic gastric ulcer with hemorrhage AND with perforation | 3739191000006114 | 76181002 | 126521010 |
| Chronic duodenal ulcer with hemorrhage, with perforation AND with obstruction | 3902501000006117 | 86258000 | 143049014 |
| Chronic duodenal ulcer with hemorrhage but without obstruction | 3513261000006117 | 62341002 | 103623018 |
| Acute gastric ulcer with haemorrhage AND obstruction | 3254501000006115 | 46708007 | 494593012 |
| Acute bleeding ulcer | 3239141000006110 | 45771005 | 76330018 |
| Acute gastric ulcer with haemorrhage but without obstruction | 3645271000006113 | 70418001 | 501576019 |
| Bleeding peptic ulcer | 3542831000006110 | 64121000 | 1232530018 |
| Gastrojejunal ulcer without hemorrhage AND without perforation | 2543301000006112 | 2783007 | 5708013 |
| Duodenal ulcer with hemorrhage AND obstruction | 2791911000006117 | 18367003 | 31008016 |
| Bleeding stress ulcer of stomach | 5982901000006117 | 308882008 | 452259016 |
| Gastric ulcer with hemorrhage | 2752311000006118 | 15902003 | 26937011 |
| Chronic gastric ulcer with hemorrhage, with perforation AND with obstruction | 3894481000006112 | 85787009 | 142209015 |
| Chronic duodenal ulcer with hemorrhage AND with perforation but without obstruction | 3819881000006114 | 81142005 | 134627014 |
| Peptic ulcer with haemorrhage AND with perforation but without obstruction | 3970661000006113 | 90489006 | 509021016 |
| Gastrojejunal ulcer without haemorrhage AND without perforation | 2543291000006111 | 2783007 | 483682014 |
| Chronic gastrojejunal ulcer with hemorrhage AND obstruction | 3967191000006116 | 90257004 | 149607014 |
| Chronic gastric ulcer with haemorrhage but without obstruction | 3737501000006117 | 76078009 | 503190016 |
| Acute duodenal ulcer with hemorrhage | 2704011000006118 | 12847006 | 22034017 |
| Acute peptic ulcer with haemorrhage but without obstruction | 2852931000006111 | 22157005 | 481240010 |
| Acute duodenal ulcer with hemorrhage but without obstruction | 3585601000006117 | 66767006 | 110913018 |
| Gastric ulcer with hemorrhage but without obstruction | 2764921000006114 | 16694003 | 28271019 |
| Acute duodenal ulcer with haemorrhage but without obstruction | 3585591000006113 | 66767006 | 500520016 |
| Acute peptic ulcer with perforation but without obstruction | 3062531000006115 | 34921009 | 58276010 |
| Peptic ulcer with hemorrhage AND perforation | 3403141000006119 | 55617001 | 92455015 |
| Peptic ulcer with haemorrhage AND obstruction | 3547581000006111 | 64398008 | 499847018 |
| Haemorrhagic ulcer of oesophagus | 3438311000006116 | 57748001 | 497977016 |
| Chronic gastric ulcer with hemorrhage AND with obstruction | 3895601000006119 | 85859006 | 142332019 |
| Giant duodenal ulcer | 3619491000006117 | 68834009 | 114337018 |
| Chronic peptic ulcer with perforation but without obstruction | 3816621000006116 | 80953005 | 134291011 |
| Bleeding duodenal ulcer | 2937141000006110 | 27281001 | 1225269012 |
| Bleeding gastric ulcer | 2752321000006114 | 15902003 | 1221244018 |
| Gastrojejunal ulcer with haemorrhage AND obstruction | 3188241000006116 | 42698006 | 493421013 |

| | | | |
|---|---|---|---|
| Chronic peptic ulcer with hemorrhage, with perforation AND with obstruction | 3763481000006114 | 77661009 | 128895010 |
| Acute gastrojejunal ulcer with hemorrhage AND obstruction | 3677541000006113 | 72408002 | 120295015 |
| Chronic duodenal ulcer with haemorrhage AND obstruction | 3047841000006117 | 34021006 | 485720014 |
| Acute gastric ulcer with hemorrhage but without obstruction | 3645281000006111 | 70418001 | 116984014 |
| Gastric ulcer with haemorrhage AND obstruction | 3373651000006112 | 53877005 | 496788013 |
| Chronic gastric ulcer with perforation but without obstruction | 3084181000006117 | 36246001 | 60497010 |
| Acute duodenal ulcer with perforation AND obstruction | 3523071000006113 | 62936002 | 104600011 |
| Gastric ulcer with hemorrhage AND obstruction | 3373661000006114 | 53877005 | 89552015 |
| Peptic ulcer with haemorrhage but without obstruction | 3287291000006112 | 48658001 | 495168013 |
| Chronic peptic ulcer with hemorrhage AND with perforation but without obstruction | 3405531000006116 | 55746001 | 92690019 |
| Peptic ulcer with haemorrhage | 3542811000006116 | 64121000 | 499762011 |
| Gastrojejunal ulcer with hemorrhage AND perforation | 3542351000006113 | 64094003 | 106539013 |
| Duodenal ulcer with haemorrhage, with perforation AND with obstruction | 2696661000006114 | 12355008 | 472362019 |
| Acute gastric ulcer NOS | 12724281000006114 | 606391000000100 | 1341841000000112 |
| Chronic duodenal ulcer with haemorrhage, with perforation AND with obstruction | 3902491000006113 | 86258000 | 507005018 |
| Duodenal ulcer with haemorrhage AND obstruction | 2791901000006115 | 18367003 | 478566011 |
| Duodenal ulcer with haemorrhage but without obstruction | 3072871000006111 | 35560008 | 486208012 |
| GU - acute + h'ge + perf. | 885791000006113 | 48974009 | 885791000006113 |
| GU - chronic + haemorrhage | 885811000006112 | 57246001 | 885811000006112 |
| GU - chronic + h'ge + perf. | 885831000006118 | 76181002 | 885831000006118 |
| DU - chronic + perforation | 885901000006115 | 49916007 | 885901000006115 |
| PU - chronic + h'ge + perf. | 886001000006111 | 61300005 | 886001000006111 |
| Acute gastric ulcer with haemorrhage AND with perforation but without obstruction | 2770951000006116 | 17067009 | 478052011 |
| Acute duodenal ulcer with haemorrhage, with perforation AND with obstruction | 3176721000006111 | 41986000 | 493172010 |
| Acute duodenal ulcer NOS | 12730031000006113 | 618291000000105 | 1365771000000117 |
| Acute peptic ulcer with hemorrhage but without obstruction | 2852941000006118 | 22157005 | 37201012 |
| Perforated ulcer | 3981721000006111 | 91182001 | 151086014 |
| Bleeding chronic gastric ulcer | 1231704018 | 57246001 | 1231704018 |
| Oesophageal bleeding due to ulcerative oesophagitis | 7222981000006119 | 439442003 | 2790933013 |
| Acute peptic ulcer with haemorrhage AND with perforation but without obstruction | 3260951000006114 | 47064007 | 494746019 |
| Bleeding oesophageal ulcer | 3438321000006112 | 57748001 | 3027281014 |
| Acute peptic ulcer with haemorrhage and perforation | 361430016 | 111353003 | 361430016 |
| Chronic duodenal ulcer with hemorrhage AND perforation | 3095701000006110 | 36975000 | 61676011 |
| Gastric ulcer with hemorrhage AND perforation | 3513611000006117 | 62366003 | 103655018 |
| Acute gastric ulcer with bleeding | 3958891000006118 | 89748001 | 148790018 |

| | | | |
|---|---|---|---|
| Chronic gastric ulcer with haemorrhage AND with perforation | 554311000006114 | 76181002 | 503219010 |
| Chronic gastric ulcer with perforation AND with obstruction | 3400861000006115 | 55483002 | 92236018 |
| Chronic gastrojejunal ulcer with perforation | 5742016 | 2807004 | 5742016 |
| Unspec peptic ulcer; unspec haemorrhage and/or perforation | 81501000006111 | 13200003 | 22592012 |
| Bleeding chronic duodenal ulcer | 1235462014 | 89469000 | 1235462014 |
| Peptic ulcer with hemorrhage but without obstruction | 3287301000006113 | 48658001 | 81073011 |
| Bleeding esophageal ulcer | 3438331000006110 | 57748001 | 3027309010 |
| Chronic gastrojejunal ulcer with hemorrhage | 3521501000006114 | 62838000 | 104433010 |
| Duodenal ulcer with perforation | 302535019 | 88968005 | 147516013 |
| Gastric ulcer with haemorrhage AND perforation | 81311000006115 | 62366003 | 499256017 |
| PU - acute NOS | 885971000006114 | 591741000000105 | 885971000006114 |
| Peptic ulcer with hemorrhage AND obstruction | 3547591000006114 | 64398008 | 107045019 |
| Bleeding ulcer of esophagus | 3438291000006115 | 57748001 | 96029011 |
| Acute gastrojejunal ulcer with hemorrhage, with perforation AND with obstruction | 3453731000006116 | 58711008 | 97562017 |
| Acute gastrojejunal ulcer with hemorrhage but without obstruction | 3466631000006111 | 59515005 | 98850013 |
| Chronic gastric ulcer with hemorrhage but without obstruction | 3737511000006119 | 76078009 | 126363010 |
| Acute gastric ulcer with hemorrhage AND obstruction | 3254511000006117 | 46708007 | 77847018 |
| Acute gastric ulcer with hemorrhage AND with perforation but without obstruction | 2770961000006119 | 17067009 | 28881013 |
| DU - acute NOS | 990721000006111 | 196652006 | 990721000006111 |
| Unspec duodenal ulcer; unspec haemorrhage and/or perforation | 81301000006118 | 51868009 | 86362013 |
| DU - chronic + haemorrhage | 885891000006119 | 89469000 | 885891000006119 |
| PU - chronic + perforation | 885991000006110 | 3483000 | 885991000006110 |
| Acute duodenal ulcer with haemorrhage | 474430017 | 12847006 | 474430017 |
| Chronic gastrojejunal ulcer with hemorrhage, with perforation AND with obstruction | 2882881000006110 | 24001002 | 40293019 |
| Chronic duodenal ulcer with perforation | 83140013 | 49916007 | 83140013 |
| Acute peptic ulcer with haemorrhage | 470365013 | 12274003 | 470365013 |
| Duodenal ulcer with perforation AND obstruction | 3759241000006113 | 77410006 | 128499012 |
| Chronic peptic ulcer with perforation AND obstruction | 3440281000006115 | 57871005 | 96224010 |
| Acute peptic ulcer with hemorrhage AND obstruction | 3199311000006117 | 43406003 | 72407011 |
| Duodenal ulcer with haemorrhage AND with perforation but without obstruction | 2739541000006116 | 15115006 | 477132014 |
| Barrett's ulcer of oesophagus | 302387019 | 196609006 | 302387019 |
| Acute gastrojejunal ulcer with hemorrhage AND with perforation but without obstruction | 3584211000006116 | 66673003 | 110759011 |
| Gastric ulcer with perforation | 302489015 | 9829001 | 17164015 |
| Chronic gastric ulcer with hemorrhage AND with perforation but without obstruction | 3708401000006111 | 74341002 | 123454019 |
| Gastrojejunal ulcer with haemorrhage, with perforation AND with obstruction | 3389331000006116 | 54798007 | 497049011 |
| Unspecified gastric ulcer with haemorrhage and perforation | 302490012 | 62366003 | 499256017 |

| | | | |
|---|---|---|---|
| Unspecified peptic ulcer with haemorrhage | 302568014 | 13200003 | 22592012 |
| Chronic gastric ulcer with haemorrhage, with perforation AND with obstruction | 3894471000006114 | 85787009 | 506732011 |
| Peptic ulcer with perforation AND obstruction | 3281961000006115 | 48336009 | 80529013 |
| Gastrojejunal ulcer with haemorrhage AND perforation | 3542341000006111 | 64094003 | 499758017 |
| Acute gastric ulcer | 158227015 | 95529005 | 158227015 |
| Gastrojejunal ulcer with haemorrhage | 3868431000006116 | 84124004 | 505930017 |
| Acute peptic ulcer with hemorrhage, with perforation AND with obstruction | 2965441000006115 | 28945005 | 48463010 |
| Gastrojejunal ulcer with hemorrhage AND obstruction | 3188251000006119 | 42698006 | 71244011 |
| Peptic ulcer with haemorrhage, with perforation AND with obstruction | 2919711000006110 | 26221006 | 483152019 |
| Gastric ulcer with perforation AND obstruction | 3678661000006114 | 72486001 | 120408012 |
| Duodenal ulcer with hemorrhage, with perforation AND with obstruction | 2696671000006119 | 12355008 | 21266019 |
| Acute gastrojejunal ulcer with haemorrhage AND perforation | 504674011 | 81387001 | 504674011 |
| Gastric ulcer with hemorrhage AND perforation but without obstruction | 2531571000006110 | 2066005 | 4556011 |
| Acute peptic ulcer without haemorrhage, without perforation AND without obstruction | 2547451000006111 | 3023008 | 484524010 |
| Acute gastric ulcer with hemorrhage | 3958881000006116 | 89748001 | 148789010 |
| Chronic gastrojejunal ulcer with hemorrhage AND with perforation but without obstruction | 3251351000006110 | 46523000 | 77544017 |
| Chronic gastrojejunal ulcer with perforation but without obstruction | 3515751000006113 | 62477005 | 103838019 |
| Acute duodenal ulcer with hemorrhage AND perforation | 3912771000006119 | 86895006 | 144111018 |
| Gastric ulcer with haemorrhage | 302487018 | 15902003 | 477573015 |
| Acute peptic ulcer with haemorrhage, with perforation AND with obstruction | 2965431000006113 | 28945005 | 484017015 |
| Chronic gastrojejunal ulcer with haemorrhage AND obstruction | 3967181000006119 | 90257004 | 508918010 |
| Peptic ulcer with haemorrhage AND perforation | 3403131000006112 | 55617001 | 497320014 |
| Chronic duodenal ulcer with hemorrhage AND obstruction | 3047851000006115 | 34021006 | 56818016 |
| Chronic peptic ulcer with haemorrhage but without obstruction | 3825511000006110 | 81518000 | 504712012 |
| Hemorrhagic ulcer of esophagus | 3438301000006119 | 57748001 | 96030018 |
| Chronic peptic ulcer with hemorrhage AND perforation | 3496021000006116 | 61300005 | 101859019 |
| Acute gastrojejunal ulcer with perforation AND obstruction | 3674881000006110 | 72219001 | 120001014 |
| Gastrojejunal ulcer with perforation AND obstruction | 2846221000006119 | 21759003 | 36512017 |
| Acute gastrojejunal ulcer with haemorrhage, with perforation AND with obstruction | 3453721000006119 | 58711008 | 498229010 |
| Acute gastric ulcer with haemorrhage AND perforation | 495273013 | 48974009 | 495273013 |
| Chronic gastric ulcer with haemorrhage | 497833010 | 57246001 | 497833010 |
| Acute duodenal ulcer with hemorrhage, with perforation AND with obstruction | 3176731000006114 | 41986000 | 70069011 |
| Chronic gastric ulcer with hemorrhage | 3430281000006116 | 57246001 | 95214011 |
| Bleeding ulcer | 3393951000006114 | 55075001 | 91564012 |

| | | | |
|---|---|---|---|
| Chronic gastric ulcer without hemorrhage, without perforation AND without obstruction | 2523581000006114 | 1567007 | 3721015 |
| Chronic peptic ulcer with haemorrhage, with perforation AND with obstruction | 3763471000006111 | 77661009 | 503637019 |
| Duodenal ulcer with hemorrhage AND perforation | 2879921000006118 | 23812009 | 39977013 |
| Chronic duodenal ulcer with perforation AND obstruction | 3483491000006119 | 60551006 | 100598014 |
| Duodenal ulcer with perforation but without obstruction | 3914141000006116 | 86983005 | 144261016 |
| Peptic ulcer with hemorrhage AND with perforation but without obstruction | 3970671000006118 | 90489006 | 149975019 |
| Chronic duodenal ulcer with haemorrhage AND perforation | 486596015 | 36975000 | 486596015 |
| Chronic peptic ulcer with haemorrhage | 495344012 | 49232000 | 495344012 |
| Acute gastrojejunal ulcer with haemorrhage | 499719010 | 63954007 | 499719010 |
| Acute duodenal ulcer with haemorrhage AND perforation | 507303017 | 86895006 | 507303017 |
| Gastric ulcer with haemorrhage but without obstruction | 2764911000006118 | 16694003 | 477900010 |
| Acute duodenal ulcer with haemorrhage AND with perforation but without obstruction | 3340391000006116 | 51847008 | 496129014 |
| Chronic gastrojejunal ulcer with haemorrhage | 499386010 | 62838000 | 499386010 |
| Chronic duodenal ulcer with haemorrhage | 508524014 | 89469000 | 508524014 |
| Bleeding acute gastric ulcer | 1235493010 | 89748001 | 1235493010 |
| Acute peptic ulcer with haemorrhage AND obstruction | 3199301000006115 | 43406003 | 493597015 |
| Chronic gastrojejunal ulcer with haemorrhage but without perforation | 3463961000006119 | 59356009 | 498412010 |
| Gastrojejunal ulcer with hemorrhage | 3868441000006114 | 84124004 | 139505014 |
| Unspec gastrojejunal ulcer with haemorrhage and perforation | 81321000006111 | 16121001 | 27283019 |
| Chronic peptic ulcer with haemorrhage AND obstruction | 3417371000006118 | 56461008 | 497616018 |
| GU - acute + haemorrhage | 885771000006112 | 89748001 | 885771000006112 |
| DU - acute + h'ge + perf. | 885871000006115 | 86895006 | 885871000006115 |
| Acute gastric ulcer with perforation | 33419017 | 19850005 | 33419017 |
| Chronic gastrojejunal ulcer with haemorrhage AND with perforation but without obstruction | 3251341000006113 | 46523000 | 494529015 |
| Acute duodenal ulcer with perforation | 101926016 | 61347001 | 101926016 |
| Acute gastrojejunal ulcer with perforation | 110704012 | 66636001 | 110704012 |
| Gastrojejunal ulcer with hemorrhage, with perforation AND with obstruction | 3389341000006114 | 54798007 | 91074015 |

#### Table S18. Brain injury Aurum codes

| Term | MedCode ID | SNOMED CT Concept ID | SNOMED CT Description ID |
|---|---|---|---|
| Traumatic subdural hemorrhage confined to region of tentorium | 7834491000006110 | 722628000 | 3325389012 |
| Open skull fracture with intracranial injury | 4198071000006115 | 111617009 | 178860014 |
| Subdural hemorrhage following injury without open intracranial wound AND with moderate loss of consciousness (1-24 hours) | 3529801000006110 | 63323000 | 105259018 |
| Brain injury without open intracranial wound AND with moderate loss of consciousness (1-24 hours) | 3267211000006116 | 47450003 | 79110011 |
| Subarachnoid haemorrhage following injury without open intracranial wound | 2950231000006111 | 28048009 | 2950231000006111 |
| Oth cerebral laceration/contusion no open intracranial wound | 39851000006115 | 269144002 | 402921014 |
| Cerebellar contusion with open intracranial wound AND loss of consciousness | 2805231000006116 | 19233004 | 32422011 |
| Intracranial injury with prolonged coma without open wound | 455459010 | 311827003 | 455459010 |
| Brain stem contusion with open intracranial wound AND moderate loss of consciousness (1-24 hours) | 2867121000006117 | 23026001 | 38683019 |
| Subarachnoid hemorrhage following injury with open intracranial wound | 2583451000006111 | 5251007 | 9804011 |
| Intracranial hematoma following injury | 3111241000006113 | 37955001 | 63313013 |
| Traumatic extradural hematoma with open intracranial wound | 6053351000006112 | 315049003 | 459517019 |
| Non-traumatic intracranial subdural haemorrhage | 4777951000006113 | 195176009 | 2913186018 |
| Traumatic cranial subdural hematoma | 5459251000006112 | 262952002 | 391039018 |
| Open traumatic subdural hemorrhage | 4854201000006110 | 209956005 | 320770015 |
| Other cerebral h'ge after injury + open intracranial wound | 35791000006117 | 450418003 | 2915288012 |
| Other cerebral h'ge after injury no open intracranial wound | 35801000006116 | 450418003 | 2915288012 |
| Oth cereb h'ge inj + open intracran wnd+LOC unspec duration | 39721000006117 | 450418003 | 2915288012 |
| Oth cereb h'ge inj no open intracran wnd+concussion unspec | 39751000006114 | 450418003 | 2915288012 |
| Subarach h'ge inj no open intracran wnd + concussion unspec | 123311000006115 | 28048009 | 46957015 |
| Subarachnoid h'ge inj + open intracran wnd+concussion unspec | 123351000006119 | 5251007 | 9804011 |
| Subarachnoid h'ge inj no open intracran wnd+<1hr loss consc | 123391000006113 | 28048009 | 46957015 |
| Subarachnoid h'ge inj no open intracran wound + 1-24hr LOC | 123421000006117 | 28048009 | 46957015 |
| Subdural h'ge inj + open intracranial wound + unspec consc | 124091000006112 | 209956005 | 320770015 |
| Extradural h'ge inj + open intracran wnd+LOC unspec duration | 659991000006112 | 65189006 | 108332012 |
| Extradural h'ge inj + open intracranial wnd + unspec consc | 660001000006119 | 65189006 | 108332012 |
| Extradural h'ge inj no open intracra wnd+LOC unspec duration | 660031000006110 | 262949005 | 391026017 |
| Extradural h'ge inj no open intracran wnd+1-24hr loss consc | 660061000006118 | 43216008 | 72101019 |
| Extradural h'ge inj no open intracranial wnd + no loss consc | 660081000006111 | 43216008 | 72101019 |
| Extradural h'ge inj no open intracranial wnd + unspec consc | 660091000006114 | 262949005 | 391026017 |

| | | | |
|---|---|---|---|
| Extradural h'ge inj no open intracranial wnd+<1hr loss consc | 660101000006115 | 43216008 | 72101019 |
| [V]Personal history of malignant neoplasm of brain | 1227581011 | 266987004 | 397818015 |
| Nontraumatic subarachnoid intracranial hemorrhage | 5518101000006114 | 270907008 | 3449601019 |
| Intracranial hemorrhage following injury without open intracranial wound | 4198351000006110 | 111668007 | 178881014 |
| Intracranial haemorrhage following injury without open intracranial wound AND with brief loss of consciousness (less than one hour) | 2780311000006112 | 17667005 | 478394019 |
| Biopsy of lesion of brain stem NEC | 512541000006116 | 230842002 | 345857013 |
| Brain cont + open intracranial wound + concussion unspec | 524321000006119 | 269144002 | 402921014 |
| Brain cont + open intracranial wound + unspec state consc | 524351000006111 | 269144002 | 402921014 |
| Closed #skull bse + intracranial injury + concussion unspec | 559971000006115 | 111603000 | 178853015 |
| Closed #skull bse + intracranial injury, LOC unspec duration | 560011000006110 | 111603000 | 178853015 |
| Closed #skull bse no intracranial inj,>24hr LOC not restored | 560041000006114 | 27644009 | 46274013 |
| Closed #skull bse no intracranial injury + concussion unspec | 560051000006111 | 27644009 | 46274013 |
| Closed #skull NOS + intracranial inj, <1hr loss of consc | 560121000006118 | 371162008 | 1209864014 |
| Closed #skull NOS + intracranial inj, >24hr LOC not restored | 560131000006115 | 371162008 | 1209864014 |
| Closed #skull NOS + intracranial inj, >24hrs LOC + recovery | 560141000006113 | 371162008 | 1209864014 |
| Closed #skull NOS no intracranial inj, >24hrs LOC + recovery | 560211000006115 | 371162008 | 1209864014 |
| Closed #skull NOS no intracranial inj, LOC unspec duration | 560231000006114 | 371162008 | 1209864014 |
| Closed #skull vlt + intracranial injury, LOC unspec duration | 560311000006112 | 207687004 | 318117015 |
| Closed #skull vlt + intracranial injury, unspec state consc | 560331000006118 | 207687004 | 318117015 |
| Closed #skull/face, mult + intracranial inj, no loss consc | 560451000006118 | 5468008 | 10127011 |
| Closed #skull/face,mult + intracran inj, concussion unspec | 560541000006119 | 5468008 | 10127011 |
| Cortex lacn + open intracranial wound + concussion unspec | 597901000006114 | 59748008 | 99251014 |
| Cortex lacn + open intracranial wound + unspec state consc | 597921000006116 | 59748008 | 99251014 |
| Crushing injury of head, part unspecified | 602701000006116 | 283851005 | 422396012 |
| Non-traumatic intracranial subdural hemorrhage | 4777941000006111 | 195176009 | 2913068019 |
| Brain damage - traumatic | 12483101000006111 | 127295002 | 524451000006116 |
| Subdural h'ge after injury | 991811000006115 | 209987007 | 991811000006115 |
| Closed traumatic subdural haemorrhage | 320752018 | 209947002 | 320752018 |
| Open traumatic subdural haemorrhage | 320771016 | 209956005 | 320771016 |
| Traumatic subdural haemorrhage | 320835014 | 209987007 | 320835014 |
| Subdural haemorrhage following injury | 320836010 | 209987007 | 320836010 |
| Traumatic subdural haematoma without open intracranial wound | 459511018 | 315046005 | 459511018 |
| Traumatic extradural haematoma with open intracranial wound | 459518012 | 315049003 | 459518012 |
| Subdural h'ge inj no open intracran wnd+>24hr LOC -restored | 124121000006111 | 209947002 | 320752018 |

| | | | |
|---|---|---|---|
| Subdural h'ge inj no open intracran wnd+1-24hr loss consc | 124131000006114 | 209947002 | 320752018 |
| Subdural h'ge inj no open intracranial wound+no loss consc | 124191000006113 | 209947002 | 320752018 |
| Subdural or cerebral haemorrhage due to birth trauma OS | 124261000006112 | 240312009 | 359984010 |
| Open traumatic subdural intracranial haemorrhage | 4854211000006113 | 209956005 | 2912603017 |
| Traumatic intracerebral haemorrhage | 7378421000006118 | 450418003 | 2915664016 |
| Contusion of cerebral cortex with open intracranial wound | 3928361000006118 | 87888006 | 145701014 |
| Extradural haemorrhage following injury with open intracranial wound AND prolonged loss of consciousness (more than 24 hours) AND return to pre-existing conscious level | 3825541000006114 | 81520002 | 3825541000006114 |
| Traumatic brain injury with no loss of consciousness | 4396321000006112 | 127302008 | 590014 |
| Epidural haematoma following injury | 647051000006114 | 262949005 | 391028016 |
| Evacuation of extradural haematoma | 649541000006113 | 171713001 | 265968017 |
| Clipping of aneurysm of cerebral artery | 506344012 | 85028008 | 506344012 |
| Open #skull/face, mult + intracranial inj, unspec consc | 263941000006114 | 5468008 | 10127011 |
| Open #skull bse + intracranial injury, unspec state of consc | 264171000006119 | 111607004 | 178855010 |
| Open #skull NOS + intracranial inj, <1hr loss of consc | 264271000006114 | 371161001 | 1209863015 |
| Open #skull NOS + intracranial inj, >24hrs LOC not restored | 264281000006112 | 371161001 | 1209863015 |
| Traumatic cerebral edema with open intracranial wound | 6009911000006116 | 311826007 | 455457012 |
| Traumatic extradural hematoma without open intracranial wound | 6053331000006117 | 315048006 | 459516011 |
| Brain stem laceration without open intracranial wound AND with concussion | 2782671000006110 | 17819003 | 30102010 |
| Traumatic brain injury with moderate loss of consciousness | 4396301000006119 | 127300000 | 588013 |
| Subdural haemorrhage following injury without open intracranial wound | 4854181000006114 | 209947002 | 2915859018 |
| Traumatic intracranial extradural hematoma | 5459191000006110 | 262949005 | 2915790014 |
| Cerebral haemorrhage following injury NOS | 320852013 | 450418003 | 2916363013 |
| Other cerebral haemorrhage following injury | 320853015 | 450418003 | 2916363013 |
| Brain stem laceration without open intracranial wound AND with prolonged loss of consciousness (more than 24 hours) AND return to pre-existing conscious level | 3649561000006119 | 70686002 | 117408014 |
| Middle meningeal haemmorhage following injury | 702641000006119 | 30400005 | 484576014 |
| Intracran inj NOS no open intracran wnd+>24hr LOC -restored | 746611000006114 | 127296001 | 584010 |
| Intracranial inj NOS + open intracran wnd+>24hr LOC+recovery | 769311000006117 | 127296001 | 584010 |
| Intracranial inj NOS + open intracran wnd+concussion unspec | 769321000006113 | 127296001 | 584010 |
| Intracranial inj NOS + open intracranial wnd+<1hr loss consc | 769351000006116 | 127296001 | 584010 |
| Intracranial inj NOS + open intracranial wound+no loss consc | 769361000006119 | 127296001 | 584010 |
| Extradural hemorrhage following injury with open intracranial wound, with no loss of consciousness | 4854301000006119 | 209978003 | 320818010 |
| Intracerebral injury NOS | 990131000006113 | 566951000000103 | 990131000006113 |

| | | | |
|---|---|---|---|
| Traumatic intracranial haemorrhage | 12757921000006111 | 82894007 | 505323015 |
| Cerebral haemorrhage following injury | 12757911000006115 | 82894007 | 505322013 |
| Subarachnoid hemorrhage following injury without open intracranial wound | 11923791000006114 | 28048009 | 46957015 |
| Craniectomy with treatment of penetrating wound of brain | 2546441000006111 | 2968008 | 6037013 |
| Subarachnoid hemorrhage following injury without open intracranial wound AND with concussion | 2567831000006110 | 4332009 | 8298019 |
| Traumatic extradural haematoma | 391028016 | 262949005 | 391028016 |
| Traumatic cranial subarachnoid hemorrhage | 5459281000006116 | 262955000 | 391046010 |
| Closed skull fracture with intracranial injury | 4198041000006111 | 111613008 | 4198041000006111 |
| Subarachnoid hemorrhage following injury with open intracranial wound AND concussion | 3284991000006114 | 48518008 | 80833014 |
| Traumatic intracranial subdural haematoma with brief loss of consciousness | 6348441000006113 | 371050006 | 1207227019 |
| Extradural hemorrhage following injury with open intracranial wound AND prolonged loss of consciousness (more than 24 hours) AND return to pre-existing conscious level | 11924651000006110 | 81520002 | 135232013 |
| Ligation of aneurysm of cerebral artery NEC | 271559016 | 230899005 | 345929019 |
| Subarachnoid haemorrhage following injury without open intracranial wound AND with no loss of consciousness | 3074431000006116 | 35672006 | 486236019 |
| Intracranial hemorrhage following injury | 12757871000006118 | 82894007 | 137497019 |
| Lacerating brain injury | 3783541000006116 | 78914008 | 503949015 |
| Traumatic subdural haematoma | 5459261000006114 | 262952002 | 2913152010 |
| Cerebellar contusion without open intracranial wound | 3438141000006115 | 57739006 | 96013015 |
| Intracranial hemorrhage following injury without intracranial wound AND with moderate loss of consciousness (1-24 hours) | 3589031000006116 | 66976009 | 111248010 |
| Intracranial haemorrhage following injury with loss of consciousness | 4396431000006115 | 127308007 | 474109017 |
| Brain stem contusion without open intracranial wound AND with concussion | 3788431000006116 | 79220008 | 131450012 |
| Subdural haemorrhage following injury without open intracranial wound AND with moderate loss of consciousness (1-24 hours) | 3529791000006114 | 63323000 | 499552019 |
| Non-traumatic subdural hemorrhage | 4777931000006118 | 195176009 | 2469434017 |
| Nontraumatic intracranial haemorrhage | 3533578018 | 738779002 | 3533578018 |
| Hind brain contusion with open intracranial wound, with more than 24 hours loss of consciousness without return to pre-existing conscious level | 823341000006110 | 209887008 | 320658014 |
| Hind brain contusion with open intracranial wound, with 1-24 hours loss of consciousness | 823351000006112 | 209885000 | 320656013 |
| Hind brain contusion with open intracranial wound, with more than 24 hours loss of consciousness and return to pre-existing conscious level | 823381000006116 | 209886004 | 320657016 |
| Chronic non-traumatic intracranial subdural haemorrhage | 7497191000006114 | 609382000 | 2959633012 |
| Subarachnoid hemorrhage following injury with open intracranial wound AND moderate loss of consciousness (1-24 hours) | 11903611000006117 | 87253004 | 144684019 |
| Extradural hemorrhage following injury without open intracranial wound AND with concussion | 3760981000006114 | 77498000 | 128652013 |
| Brain stem laceration without open intracranial wound AND with brief loss of consciousness (less than one hour) | 3467421000006112 | 59561005 | 98926012 |

| | | | |
|---|---|---|---|
| Traumatic brain injury with prolonged loss of consciousness | 4396311000006116 | 127301001 | 589017 |
| Nontraumatic hemorrhage of subarachnoid space from intracranial artery | 3511284017 | 291371000119100 | 3511284017 |
| Brain injury with open intracranial wound AND brief loss of consciousness (less than one hour) | 2951821000006119 | 28156009 | 47135017 |
| Intracranial haemorrhage following injury | 7378271000006116 | 450410005 | 2915426010 |
| Nontraumatic extradural intracranial hemorrhage | 6593141000006116 | 397809001 | 2915638010 |
| Extradural hemorrhage following injury with open intracranial wound AND moderate loss of consciousness (1-24 hours) | 3468981000006117 | 59648004 | 99099016 |
| Intracranial hemorrhage following injury with intracranial wound AND moderate loss of consciousness (1-24 hours) | 3055551000006113 | 34501004 | 57585011 |
| Non-traumatic extradural intracranial hematoma | 5656951000006110 | 281865000 | 419997012 |
| Cerebellar laceration with open intracranial wound AND prolonged loss of consciousness (more than 24 hours) AND return to pre-existing conscious level | 3366571000006117 | 53429002 | 88842018 |
| Traumatic haemorrhage of subdural space of infratentorial region | 7834471000006114 | 722628000 | 3325385018 |
| Traumatic cerebral oedema without open intracranial wound | 455455016 | 311825006 | 455455016 |
| Excision of aneurysm of cerebral artery | 271556011 | 175385008 | 271556011 |
| Intracranial haemorrhage following injury with open intracranial wound AND concussion | 3782881000006115 | 78879009 | 503942012 |
| Craniotomy and evacuation of traumatic haematoma | 7330521000006119 | 446816008 | 2883668012 |
| Intracranial hemorrhage following injury without open intracranial wound AND with prolonged loss of consciousness (more than 24 hours) without return to pre-existing level | 2521231000006111 | 1430001 | 3487015 |
| Brain stem laceration with open intracranial wound AND moderate loss of consciousness (1-24 hours) | 3160801000006118 | 41025001 | 68439012 |
| Brain stem laceration without open intracranial wound AND with no loss of consciousness | 3777371000006118 | 78525006 | 130297019 |
| Subarachnoid haemorrhage following injury without open intracranial wound AND with loss of consciousness | 3629571000006112 | 69458009 | 501279013 |
| Multiple open fractures of skull AND/OR face with cerebral laceration AND/OR contusion | 3930061000006116 | 88008005 | 145885012 |
| Cerebellar contusion with open intracranial wound AND brief loss of consciousness (less than one hour) | 3683771000006112 | 72797002 | 120920014 |
| Intracranial hemorrhage following injury with open intracranial wound and prolonged loss of consciousness (more than 24 hours) without return to pre-existing level | 4198371000006117 | 111671004 | 135096017 |
| Cerebellar laceration with open intracranial wound | 2997431000006117 | 30858001 | 51649013 |
| Intracranial hemorrhage co-occurrent and due to complex wound of head | 7840901000006116 | 723141009 | 3335258015 |
| Brain stem contusion without open intracranial wound AND with loss of consciousness | 2979771000006112 | 29807001 | 49879010 |
| Traumatic injury of blood vessel of head | 7317871000006113 | 446029009 | 2883183018 |
| Extradural haemorrhage following injury without open intracranial wound AND with concussion | 3760971000006111 | 77498000 | 503596015 |
| Intracranial haemorrhage following injury without open intracranial wound | 4198341000006113 | 111668007 | 189842012 |
| Secondary traumatic haemorrhage of brainstem | 7840851000006114 | 723139008 | 3334700013 |

| | | | |
|---|---|---|---|
| Extradural haemorrhage following injury with open intracranial wound | 320814012 | 65189006 | 500043017 |
| Subarachnoid haemorrhage following injury with open intracranial wound AND prolonged loss of consciousness (more than 24 hours) without return to pre-existing conscious level | 123291000006119 | 69178005 | 501220014 |
| Subarachnoid haemorrhage following injury with open intracranial wound AND moderate loss of consciousness (1-24 hours) | 123341000006116 | 87253004 | 507490011 |
| Subdural haemorrhage following injury with open intracranial wound AND moderate loss of consciousness (1-24 hours) | 124071000006111 | 90165008 | 508869011 |
| Subdural haemorrhage following injury without open intracranial wound AND with loss of consciousness | 3049841000006112 | 34135005 | 485760010 |
| Subdural haemorrhage following injury with open intracranial wound AND brief loss of consciousness (less than one hour) | 124101000006118 | 29635000 | 484345019 |
| Subdural haemorrhage following injury with open intracranial wound, with no loss of consciousness | 124111000006115 | 209958006 | 320775013 |
| Extradural haemorrhage following injury without open intracranial wound AND with no loss of consciousness | 4198421000006112 | 111679002 | 364069017 |
| Brain stem laceration without open intracranial wound AND with moderate loss of consciousness (1-24 hours) | 2794111000006117 | 18531006 | 31261015 |
| Nontraumatic subarachnoid haemorrhage with brain compression | 8044541000006113 | 141091000119105 | 3043206010 |
| Extradural haemorrhage following injury without open intracranial wound AND with prolonged loss of consciousness (more than 24 hours) AND return to pre-existing conscious level | 3879081000006114 | 84792001 | 506232012 |
| Intracranial hemorrhage following injury with brief loss of consciousness | 4396461000006112 | 127309004 | 597012 |
| Subdural hemorrhage following injury without open intracranial wound AND with prolonged loss of consciousness (more than 24 hours) AND return to pre-existing conscious level | 3920061000006118 | 87345009 | 144822016 |
| Subacute non-traumatic intracranial subdural hemorrhage | 8089881000006112 | 291591000119107 | 2959779014 |
| Intracranial hemorrhage following injury without open intracranial wound AND with prolonged loss of consciousness (more than 24 hours) AND return to pre-existing conscious level | 3776601000006116 | 78477003 | 130220011 |
| Brain stem laceration with open intracranial wound AND prolonged loss of consciousness (more than 24 hours) without return to pre-existing conscious level | 3547841000006115 | 64413001 | 107072010 |
| Open fracture of base of skull with cerebral laceration AND contusion | 2989141000006113 | 30371007 | 50833015 |
| Subarachnoid hemorrhage following injury without open intracranial wound AND with no loss of consciousness | 3074441000006114 | 35672006 | 59500013 |
| Subarachnoid hemorrhage following injury without open intracranial wound AND with prolonged loss of consciousness (more than 24 hours) AND return to pre-existing conscious level | 3942121000006114 | 88747000 | 147146018 |
| Brain injury with open intracranial wound AND no loss of consciousness | 2705081000006112 | 12912004 | 22137010 |
| Subarachnoid haemorrhage following injury without open intracranial wound AND with moderate loss of consciousness (1-24 hours) | 4198381000006119 | 111673001 | 364055017 |

| | | | |
|---|---|---|---|
| Cerebellar laceration without open intracranial wound | 2983451000006116 | 30031007 | 50251013 |
| Subdural haemorrhage following injury without open intracranial wound AND with no loss of consciousness | 3145681000006112 | 40135004 | 492453011 |
| Extradural hemorrhage following injury with open intracranial wound AND loss of consciousness | 4198451000006115 | 111681000 | 189006017 |
| Traumatic intracranial subdural hematoma with brief loss of consciousness | 6348451000006110 | 371050006 | 1208039019 |
| Cranial puncture | 5559151000006110 | 273989008 | 409716019 |
| Compression of brain co-occurrent and due to nontraumatic subarachnoid haemorrhage | 8044551000006110 | 141091000119105 | 3042983016 |
| Subdural hemorrhage following injury without open intracranial wound AND with brief loss of consciousness (less than one hour) | 2919471000006113 | 26205001 | 43899010 |
| Subarachnoid haemorrhage following injury without open intracranial wound AND with brief loss of consciousness (less than one hour) | 3914851000006114 | 87020000 | 507372011 |
| Focal haemorrhagic contusion of cerebrum | 7834261000006115 | 722615007 | 3332903015 |
| Subarachnoid haemorrhage following injury with open intracranial wound AND moderate loss of consciousness (1-24 hours) | 3918591000006111 | 87253004 | 3918591000006111 |
| Cerebellar laceration with open intracranial wound AND prolonged loss of consciousness (more than 24 hours) without return to pre-existing conscious level | 3546541000006115 | 64337000 | 106943019 |
| Subarachnoid haemorrhage following injury without open intracranial wound AND with prolonged loss of consciousness (more than 24 hours) AND return to pre-existing conscious level | 3942111000006118 | 88747000 | 508175017 |
| Injury of head with otorrhagia | 4063221000006112 | 95848000 | 158756016 |
| Open fracture of base of skull with intracranial injury, with less than 1 hour loss of consciousness | 264121000006115 | 207746007 | 318190017 |
| Intracranial hemorrhage following injury with open intracranial wound AND brief loss of consciousness (less than one hour) | 2795161000006118 | 18605003 | 31370015 |
| Traumatic brain injury with prolonged loss of consciousness (more than 24 hours) and return to pre-existing conscious level | 7380341000006114 | 450551009 | 2916174013 |
| Subdural haemorrhage following injury without open intracranial wound AND with prolonged loss of consciousness (more than 24 hours) AND return to pre-existing conscious level | 3920051000006115 | 87345009 | 507531010 |
| Compression of brain co-occurrent and due to nontraumatic subarachnoid hemorrhage | 8044561000006112 | 141091000119105 | 3043035013 |
| Traumatic brain injury with loss of consciousness one hour or more | 7380591000006114 | 450569000 | 2915718018 |
| Intracranial hemorrhage following injury without open intracranial wound AND with brief loss of consciousness (less than one hour) | 2780321000006116 | 17667005 | 29855017 |
| Intracranial hemorrhage following injury with loss of consciousness | 4396441000006113 | 127308007 | 596015 |
| Traumatic subdural hematoma without open intracranial wound | 6053291000006112 | 315046005 | 459512013 |
| Intracranial hemorrhage following injury with open intracranial wound | 3765301000006113 | 77768006 | 129069015 |
| Subarachnoid hemorrhage following injury without open intracranial wound AND with brief loss of consciousness (less than one hour) | 3914861000006111 | 87020000 | 144313014 |
| Intracranial hemorrhage following injury without open intracranial wound AND with concussion | 3781111000006112 | 78757009 | 130692016 |

| | | | |
|---|---|---|---|
| Brain injury with open intracranial wound | 2952391000006117 | 28188001 | 47189015 |
| Cerebellar contusion with open intracranial wound AND concussion | 3900201000006118 | 86125000 | 142812014 |
| Traumatic brain injury with prolonged loss of consciousness (more than 24 hours) without return to pre-existing conscious level | 7380351000006111 | 450552002 | 2915596018 |
| Non-traumatic spinal subdural haematoma | 6028571000006117 | 313304007 | 457177012 |
| Extradural hemorrhage following injury without open intracranial wound AND with brief loss of consciousness (less than one hour) | 3152821000006115 | 40549004 | 67610019 |
| Brain damage - traumatic | 11923371000006114 | 275272006 | 411234012 |
| Subdural hemorrhage following injury without open intracranial wound AND with concussion | 2606541000006112 | 6666006 | 12042015 |
| Head injury with hemorrhage from nose | 4395861000006117 | 127276009 | 205229010 |
| Intracranial haemorrhage following injury with prolonged loss of consciousness without return to pre-existing conscious level | 4396511000006110 | 127312001 | 474113012 |
| Non-traumatic intracranial subdural hematoma | 5656931000006115 | 281864001 | 419995016 |
| Focal traumatic haematoma of brainstem | 7834421000006113 | 722624003 | 3334493017 |
| Nontraumatic subdural haematoma with brain compression | 8044591000006116 | 141151000119101 | 2967571019 |
| Subacute non-traumatic intracranial subdural haemorrhage | 8089861000006119 | 291591000119107 | 2959517016 |
| Subarachnoid haemorrhage following injury with open intracranial wound | 320735017 | 5251007 | 496337012 |
| Traumatic subdural hematoma | 391035012 | 262952002 | 2912898019 |
| Lobotomy of brain | 5559171000006117 | 273991000 | 409719014 |
| Intracranial haemorrhage following injury with open intracranial wound AND brief loss of consciousness (less than one hour) | 2795151000006115 | 18605003 | 478724012 |
| Brain stem laceration with open intracranial wound | 2700331000006119 | 12589008 | 21640014 |
| Traumatic subdural intracranial haemorrhage | 4854361000006118 | 209987007 | 2912530015 |
| Subarachnoid hemorrhage following injury with open intracranial wound AND brief loss of consciousness (less than one hour) | 3468691000006111 | 59633005 | 99064015 |
| Closed traumatic subdural hemorrhage | 4854141000006115 | 209947002 | 320753011 |
| Subdural haemorrhage following injury without open intracranial wound AND with concussion | 2606531000006119 | 6666006 | 500492017 |
| Extradural haemorrhage following injury without open intracranial wound AND with prolonged loss of consciousness (more than 24 hours) without return to pre-existing conscious level | 2768441000006115 | 16907002 | 477986019 |
| Acute nontraumatic subdural haemorrhage | 3515795019 | 291581000119109 | 3515795019 |
| Intracran inj NOS + open intracran wnd+LOC unspec duration | 746601000006111 | 127296001 | 584010 |
| Intracranial inj NOS no open intracran wnd+<1hr loss consc | 768831000006114 | 127296001 | 584010 |
| Intracranial inj NOS no open intracran wnd+1-24hr loss consc | 768851000006119 | 127296001 | 584010 |
| Subdural hemorrhage following injury without open intracranial wound AND with loss of consciousness | 3049851000006114 | 34135005 | 57003014 |
| Closed fracture of skull NOS with intracranial injury | 12484051000006114 | 371162008 | 563781000006112 |
| Subdural hemorrhage following injury with open intracranial wound AND prolonged loss of consciousness (more than 24 hours) AND return to pre-existing conscious level | 3357881000006110 | 52902005 | 88030010 |

| | | | |
|---|---|---|---|
| Focal traumatic hematoma of brainstem | 7834431000006111 | 722624003 | 3332924013 |
| Nontraumatic subdural hematoma with brain compression | 8044601000006112 | 141151000119101 | 2967694013 |
| Open fracture of vault of skull with intracranial injury, with 1-24 hours loss of consciousness | 52941000006118 | 207709008 | 318139018 |
| Cerebellar contusion without open intracranial wound AND with loss of consciousness | 2772451000006114 | 17169009 | 29035017 |
| Cerebral hemorrhage following injury | 12757891000006117 | 82894007 | 505319011 |
| Traumatic intracranial hemorrhage | 12757901000006118 | 82894007 | 505320017 |
| Subdural hemorrhage following injury without open intracranial wound AND with no loss of consciousness | 3145691000006110 | 40135004 | 63714017 |
| Subarachnoid haemorrhage following injury with open intracranial wound AND loss of consciousness | 3727871000006112 | 75507000 | 503040018 |
| Subdural hemorrhage following injury with open intracranial wound AND concussion | 3197131000006115 | 43262000 | 72179018 |
| Extradural hemorrhage following injury without open intracranial wound AND with prolonged loss of consciousness (more than 24 hours) AND return to pre-existing conscious level | 3879091000006112 | 84792001 | 140578018 |
| Brain injury with open intracranial wound AND moderate loss of consciousness (1-24 hours) | 3898191000006111 | 86010003 | 142597011 |
| Non-traumatic extradural intracranial haematoma | 5656941000006113 | 281865000 | 419996015 |
| Traumatic haemorrhage of brainstem | 7834611000006115 | 722632006 | 3332940011 |
| Cerebellar contusion with open intracranial wound AND prolonged loss of consciousness (more than 24 hours) without return to pre-existing conscious level | 2863721000006113 | 22825007 | 38315013 |
| Traumatic spinal subdural hematoma | 5456261000006117 | 262721007 | 390720010 |
| Cortex contusion with open intracranial wound | 3928351000006115 | 87888006 | 145700010 |
| Traumatic cranial subdural haematoma | 5459221000006115 | 262952002 | 391036013 |
| Subarachnoid haemorrhage following injury with open intracranial wound AND brief loss of consciousness (less than one hour) | 123371000006112 | 59633005 | 498500014 |
| Subarachnoid haemorrhage following injury with open intracranial wound, with no loss of consciousness | 123381000006110 | 209940000 | 320739011 |
| Subdural haemorrhage following injury with open intracranial wound AND prolonged loss of consciousness (more than 24 hours) AND return to pre-existing conscious level | 124061000006116 | 52902005 | 496480019 |
| Brain stem contusion with open intracranial wound AND brief loss of consciousness (less than one hour) | 3426401000006114 | 57012007 | 94810019 |
| Subarachnoid hemorrhage following injury without open intracranial wound AND with prolonged loss of consciousness (more than 24 hours) AND without return to pre-existing conscious level | 2822141000006116 | 20276007 | 34108017 |
| Subdural hemorrhage following injury with open intracranial wound, with no loss of consciousness | 4854261000006111 | 209958006 | 320774012 |
| Cerebellar laceration with open intracranial wound AND loss of consciousness | 3880781000006111 | 84900008 | 140766015 |
| Closed fracture of vault of skull with intracranial injury, with no loss of consciousness | 560321000006116 | 207689001 | 318119017 |
| Brain stem contusion with open intracranial wound AND no loss of consciousness | 3358011000006113 | 52913008 | 88047011 |

| | | | |
|---|---|---|---|
| Extradural haemorrhage following injury without open intracranial wound AND with loss of consciousness | 3965881000006116 | 90178008 | 508877010 |
| Oth cereb h'ge inj no open intracran wnd+LOC unspec duration | 39761000006111 | 450418003 | 2915288012 |
| Subarachnoid h'ge inj no open intracran wnd+no loss consc | 123411000006113 | 28048009 | 46957015 |
| Traumatic hemorrhage of brainstem | 7834621000006111 | 722632006 | 3332939014 |
| Subarachnoid haemorrhage following injury without open intracranial wound AND with prolonged loss of consciousness (more than 24 hours) AND without return to pre-existing conscious level | 2822131000006114 | 20276007 | 480562018 |
| Intracranial hemorrhage following injury without open intracranial wound AND with loss of consciousness | 2793491000006117 | 18485009 | 31186017 |
| Cerebellar laceration with open intracranial wound AND no loss of consciousness | 2728701000006119 | 14434006 | 24535010 |
| Multiple traumatic hemorrhages of brain tissue | 7834661000006117 | 722633001 | 3332944019 |
| Intracranial hemorrhage following injury with open intracranial wound AND loss of consciousness | 2710751000006112 | 13289004 | 22738011 |
| Open fracture of base of skull with intracranial injury | 259601000006113 | 111607004 | 178855010 |
| Open fracture of vault of skull with intracranial injury, with less than 1 hour loss of consciousness | 264431000006110 | 207708000 | 318138014 |
| Open fracture of base of skull with intracranial injury, with more than 24 hours loss of consciousness without return to pre-existing conscious level | 265451000006116 | 207749000 | 318193015 |
| Head injury with haemorrhage from ear | 4063241000006117 | 95848000 | 512328010 |
| Intracranial hemorrhage following injury with prolonged loss of consciousness AND return to pre-existing conscious level | 4396501000006112 | 127311008 | 599010 |
| Brain stem laceration without open intracranial wound | 2832491000006115 | 20899000 | 35146017 |
| Extradural haemorrhage following injury without open intracranial wound AND with brief loss of consciousness (less than one hour) | 3152811000006111 | 40549004 | 492668015 |
| Brain injury with open intracranial wound AND prolonged loss of consciousness (more than 24 hours) AND return to pre-existing conscious level | 2948171000006119 | 27923006 | 46743012 |
| Closed fracture of base of skull with cerebral laceration AND/OR contusion | 3988021000006116 | 91589002 | 151735014 |
| Nontraumatic haemorrhage of subarachnoid space from intracranial artery | 3511285016 | 291371000119100 | 3511285016 |
| Intracranial haemorrhage following injury with open intracranial wound AND no loss of consciousness | 2624481000006115 | 7819003 | 503779018 |
| Cerebellar laceration without open intracranial wound AND with prolonged loss of consciousness (more than 24 hours) AND return to pre-existing conscious level | 3981471000006117 | 91168001 | 151064013 |
| Subdural hemorrhage following injury with open intracranial wound AND loss of consciousness | 3237321000006110 | 45659008 | 76138016 |
| Traumatic cerebral hemorrhage | 7378441000006113 | 450418003 | 2916150010 |
| Subarachnoid hemorrhage following injury with open intracranial wound AND prolonged loss of consciousness (more than 24 hours) without return to pre-existing conscious level | 11903601000006115 | 69178005 | 114903015 |
| Traumatic intracerebral haemorrhage | 12757941000006116 | 82894007 | 2475153011 |

| | | | |
|---|---|---|---|
| Subarachnoid hemorrhage following injury with open intracranial wound, with no loss of consciousness | 4854121000006110 | 209940000 | 320738015 |
| Intracranial nontraumatic haemorrhage of foetus and newborn | 394431000006118 | 206417009 | 2791314016 |
| Brain contusion with open intracranial wound, with more than 1 hour loss of consciousness | 524291000006111 | 209921006 | 320693019 |
| Brain contusion with open intracranial wound, with 1-24 hours loss of consciousness | 524311000006110 | 209922004 | 320694013 |
| Brain contusion with open intracranial wound, with no loss of consciousness | 524341000006114 | 209920007 | 320692012 |
| Biopsy of lesion of cranial nerve | 512591000006113 | 171687004 | 265940016 |
| Brain cont + open intracranial wound + LOC unspec duration | 524331000006116 | 269144002 | 402921014 |
| Closed #skull bse no intracranial injury, <1hr loss of consc | 560061000006113 | 27644009 | 46274013 |
| Closed #skull vlt with intracranial injury+concussion unspec | 560421000006110 | 207687004 | 318117015 |
| Cortex cont + open intracranial wnd + LOC unspec duration | 597681000006113 | 209843004 | 320611018 |
| Cortex cont + open intracranial wound + unspec state consc | 597731000006115 | 87888006 | 145700010 |
| Cortex lacn + open intracranial wnd + LOC unspec duration | 597861000006119 | 59748008 | 99251014 |
| Cortex lacn no open intracranial wnd + LOC unspec duration | 597951000006113 | 78914008 | 130942015 |
| Late effect of injury to blood vessel of head, neck and extremities | 750631000006119 | 210950000 | 322021015 |
| Injuries of brain and cranial nerves with injuries of nerves and spinal cord at neck level | 777751000006119 | 213383004 | 325280017 |
| Intracranial haemorrhage following injury with brief loss of consciousness | 4396451000006110 | 127309004 | 474110010 |
| Intracranial hemorrhage following injury with open intracranial wound AND concussion | 3782891000006117 | 78879009 | 130868014 |
| Traction injury of cranial nerve | 5459341000006115 | 262960001 | 391054012 |
| Extradural hemorrhage following injury without open intracranial wound AND with prolonged loss of consciousness (more than 24 hours) without return to pre-existing conscious level | 2768451000006118 | 16907002 | 28621013 |
| Nontraumatic subarachnoid hemorrhage with brain compression | 8044571000006117 | 141091000119105 | 3043074018 |
| Closed skull fracture with cerebral laceration AND/OR contusion | 3940751000006119 | 88651008 | 146986016 |
| Cerebellar laceration without open intracranial wound AND with no loss of consciousness | 3887091000006113 | 85314006 | 141420019 |
| Craniotomy and removal of hematoma from extradural space | 7312961000006111 | 445746008 | 2884424011 |
| Closed fracture of vault of skull with intracranial injury, with less than 1 hour loss of consciousness | 560281000006110 | 207690005 | 318120011 |
| Cortex contusion with open intracranial wound, with more than 24 hours loss of consciousness and return to pre-existing conscious level | 597661000006115 | 209849000 | 320617019 |
| Cortex laceration with open intracranial wound, with more than 24 hours loss of consciousness and return to pre-existing conscious level | 597881000006112 | 209867007 | 320635011 |
| Cortex laceration with open intracranial wound, with 1-24 hours loss of consciousness | 597891000006110 | 209866003 | 320634010 |
| Cortex laceration with open intracranial wound, with no loss of consciousness | 597911000006112 | 209864000 | 320632014 |
| Nontraumatic extradural haemorrhage | 660111000006117 | 397809001 | 1773153015 |
| Intracranial haemorrhage following injury without open intracranial wound AND with prolonged loss | 2521221000006113 | 1430001 | 476216010 |

| | | | |
|---|---|---|---|
| of consciousness (more than 24 hours) without return to pre-existing level | | | |
| Traumatic spinal extradural haematoma | 5456211000006115 | 262720008 | 390718012 |
| Open #skull vlt with intracranial injury + concussion unspec | 263881000006110 | 207705002 | 318135012 |
| Intracranial hemorrhage following injury with moderate loss of consciousness | 4396481000006119 | 127310009 | 598019 |
| Traumatic hemorrhage of subdural space of infratentorial region | 7834481000006112 | 722628000 | 3325383013 |
| Intracranial haemorrhage co-occurrent and due to complex wound of head | 7840891000006115 | 723141009 | 3335259011 |
| Traumatic spinal subdural haematoma | 5456251000006119 | 262721007 | 390721014 |
| Brain stem laceration with open intracranial wound AND concussion | 3164101000006115 | 41222005 | 68763015 |
| Cerebellar laceration without open intracranial wound AND with brief loss of consciousness (less than one hour) | 3059661000006113 | 34751006 | 58002019 |
| Extradural hemorrhage following injury without open intracranial wound AND with moderate loss of consciousness (1-24 hours) | 3523681000006119 | 62973004 | 104678015 |
| Subdural haemorrhage following injury without open intracranial wound AND with prolonged loss of consciousness (more than 24 hours) without return to pre-existing conscious level | 3652481000006119 | 70861009 | 501707010 |
| Primary traumatic haemorrhage of brainstem | 7840831000006119 | 723138000 | 3334697014 |
| Oth cerebral h'ge inj + open intracranial wnd+no loss consc | 39801000006119 | 450418003 | 2915288012 |
| Subarach h'ge inj no open intracran wnd+>24hrs LOC-restored | 123321000006111 | 28048009 | 46957015 |
| Subarachnoid h'ge inj + open intracran wound + unspec consc | 123361000006117 | 5251007 | 9804011 |
| Extradural h'ge inj no open intracran wnd+>24hr LOC+recovery | 660041000006117 | 43216008 | 72101019 |
| Subdural haemorrhage - nontraumatic | 300294013 | 195176009 | 300294013 |
| Traumatic intracerebral hemorrhage | 12757951000006119 | 82894007 | 2475363012 |
| Traumatic intracranial subdural haematoma | 5459231000006117 | 262952002 | 391037016 |
| Extradural hemorrhage following injury without open intracranial wound AND with no loss of consciousness | 4198431000006110 | 111679002 | 199709019 |
| Cerebellar laceration without open intracranial wound AND with loss of consciousness | 3969771000006113 | 90429009 | 149873010 |
| Subarachnoid h'ge inj no open intracran wound + unspec consc | 123431000006119 | 28048009 | 46957015 |
| Open fracture of base of skull with intracranial injury, with more than 24 hours loss of consciousness and return to pre-existing conscious level | 264131000006117 | 207748008 | 318192013 |
| Subdural haemorrhage following injury without open intracranial wound AND with brief loss of consciousness (less than one hour) | 2919461000006118 | 26205001 | 483148019 |
| Open #skull/face, mult + intracran inj + concussion, unspec | 263891000006113 | 5468008 | 10127011 |
| Open #skull NOS + intracranial inj, LOC unspec duration | 264301000006111 | 371161001 | 1209863015 |
| Traumatic injury of third cranial nerve | 3370341000006114 | 53675004 | 198460011 |
| Closed #skull bse no intracranial injury, 1-24hr loss consc | 560081000006115 | 27644009 | 46274013 |
| Closed #skull NOS + intracranial inj, 1-24hrs loss of consc | 560151000006110 | 371162008 | 1209864014 |

| | | | |
|---|---|---|---|
| Closed #skull NOS no intracranial inj, <1hr loss of consc | 560201000006118 | 371162008 | 1209864014 |
| Closed #skull NOS no intracranial inj, unspec state of consc | 560251000006119 | 371162008 | 1209864014 |
| Closed #skull/face,mult + intracran inj, LOC unspec duration | 560551000006117 | 5468008 | 10127011 |
| Cortex lacn no open intracranial wnd + 1-24hr loss of consc | 597941000006111 | 78914008 | 130942015 |
| Mult #skull/face+other bones, closed + intracranial injury | 696141000006119 | 5468008 | 10127011 |
| Hind brain cont + open intracranial wnd +LOC unspec duration | 823391000006118 | 209881009 | 320652010 |
| Subdural haemorrhage following injury with open intracranial wound AND concussion | 3197121000006118 | 43262000 | 493561012 |
| Closed fracture of base of skull with intracranial injury, with less than 1 hour loss of consciousness | 559981000006117 | 207728001 | 318172018 |
| Closed fracture of vault of skull with intracranial injury, with more than 24 hours loss of consciousness without return to pre-existing conscious level | 560271000006112 | 207693007 | 318123013 |
| Cortex laceration with open intracranial wound, with less than 1 hour loss of consciousness | 597871000006114 | 209865004 | 320633016 |
| Intracranial haemorrhage following injury | 12716261000006116 | 82894007 | 505321018 |
| Nontraumatic extradural intracranial haemorrhage | 6593151000006119 | 397809001 | 2916444013 |
| Intracran inj NOS + open intracran wnd+>24hr LOC -restored | 746591000006115 | 127296001 | 584010 |
| Intracranial inj NOS no open intracran wnd+>24hr LOC+recover | 768841000006116 | 127296001 | 584010 |
| Intracranial injury, excluding those with skull fracture NOS | 768961000006113 | 54355006 | 90339018 |
| Intracranial inj NOS + open intracranial wnd + 1-24hr LOC | 769331000006111 | 127296001 | 584010 |
| Intracranial inj NOS + open intracranial wnd + unspec consc | 769341000006118 | 127296001 | 584010 |
| Injury of blood vessels of head, NEC | 776021000006111 | 85564003 | 141827011 |
| Focal traumatic haemorrhage of brainstem | 7834401000006115 | 722623009 | 3332922012 |
| Extradural haemorrhage following injury with open intracranial wound AND prolonged loss of consciousness (more than 24 hours) without return to pre-existing conscious level | 659951000006118 | 86182004 | 506960017 |
| Extradural haemorrhage following injury with open intracranial wound AND brief loss of consciousness (less than one hour) | 660011000006116 | 73308006 | 502406019 |
| Secondary hemorrhage of brainstem due to traumatic injury | 7840871000006116 | 723139008 | 3334701012 |
| Cerebellar contusion with open intracranial wound AND moderate loss of consciousness (1-24 hours) | 3427941000006119 | 57099002 | 94960012 |
| Subarach h'ge inj + open intracran wnd+LOC unspec duration | 123301000006118 | 5251007 | 9804011 |
| Subarachnoid h'ge inj no open intracran wnd+>24 LOC+recovery | 123401000006110 | 28048009 | 46957015 |
| Subdural h'ge inj + open intracran wnd+LOC unspec duration | 124051000006118 | 209956005 | 320770015 |
| Extradural h'ge inj + open intracran wnd+concussion unspec | 659981000006114 | 65189006 | 108332012 |
| Extradural h'ge inj no open intracran wnd+>24hr LOC-restored | 660051000006115 | 43216008 | 72101019 |
| Sequela of traumatic intracranial hemorrhage | 8031561000006113 | 100581000119102 | 2967672017 |
| Closed #skull bse no intracranial injury, >24hr LOC+recovery | 560071000006118 | 27644009 | 46274013 |

| | | | |
|---|---|---|---|
| Closed #skull NOS + intracranial inj, no loss of consc | 560171000006117 | 371162008 | 1209864014 |
| Closed #skull NOS no intracranial inj, no loss of consc | 560241000006116 | 371162008 | 1209864014 |
| Closed #skull NOS no intracranial inj,>24hr LOC not restored | 560261000006117 | 371162008 | 1209864014 |
| Extradural haemorrhage after injury | 5459181000006112 | 262949005 | 391029012 |
| Extradural hemorrhage following injury with open intracranial wound AND concussion | 2951811000006110 | 28155008 | 47133012 |
| Head injury with hemorrhage from ear | 4063231000006110 | 95848000 | 158757013 |
| Subarachnoid hemorrhage following injury with open intracranial wound AND prolonged loss of consciousness (more than 24 hours) AND return to pre-existing conscious level | 11903591000006111 | 73439007 | 121954015 |
| Traumatic subdural hematoma with open intracranial wound | 6053311000006111 | 315047001 | 459513015 |
| Traumatic subdural intracranial hemorrhage | 4854351000006115 | 209987007 | 2912507014 |
| Traumatic subdural hemorrhage | 4854331000006110 | 209987007 | 320837018 |
| Fracture of frontonasoethmoidal complex with increased intercanthal distance | 5461381000006115 | 263154009 | 391284014 |
| Traumatic brain injury with loss of consciousness | 4396281000006118 | 127298000 | 586012 |
| Subdural h'ge inj no open intracran wnd+LOC unspec duration | 124141000006116 | 209947002 | 320752018 |
| Sequelae of other nontraumatic intracranial haemorrhage | 149551000006111 | 363302008 | 482447013 |
| Open #skull vlt + intracranial injury, unspec state of consc | 263791000006119 | 207705002 | 318135012 |
| Open #skull/face, mult + intracranial inj, no loss consc | 263931000006116 | 5468008 | 10127011 |
| Open #skull NOS + intracranial inj, 1-24hrs loss of consc | 264291000006110 | 371161001 | 1209863015 |
| Open #skull NOS + intracranial inj, no loss of consc | 264311000006114 | 371161001 | 1209863015 |
| Open #skull NOS + intracranial inj, unspec state of consc | 264321000006118 | 371161001 | 1209863015 |
| Focal traumatic hemorrhage of brainstem | 7834411000006117 | 722623009 | 3332921017 |
| Open #skull NOS + intracranial inj + concussion unspec | 264261000006119 | 371161001 | 1209863015 |
| Intracranial haemorrhage following injury with open intracranial wound AND loss of consciousness | 2710741000006110 | 13289004 | 475195013 |
| Subarachnoid haemorrhage following injury with open intracranial wound AND prolonged loss of consciousness (more than 24 hours) without return to pre-existing conscious level | 3625151000006114 | 69178005 | 3625151000006114 |
| Intracranial hemorrhage following injury | 7378281000006118 | 450410005 | 2915467017 |
| Intracranial injury with prolonged coma with open wound | 455461018 | 311829000 | 455461018 |
| Head injury with haemorrhage from nose | 4395871000006112 | 127276009 | 474087019 |
| Temporal resolution test | 5598691000006114 | 277391005 | 413892012 |
| Closed fracture of vault of skull with intracranial injury, with more than 24 hours loss of consciousness and return to pre-existing conscious level | 560291000006113 | 207692002 | 318122015 |
| Cortex contusion with open intracranial wound, with no loss of consciousness | 597721000006118 | 209845006 | 320613015 |
| Extradural haemorrhage following injury with open intracranial wound AND moderate loss of consciousness (1-24 hours) | 659971000006111 | 59648004 | 498503011 |

| | | | |
|---|---|---|---|
| Traumatic intracranial haemorrhage | 660121000006113 | 450410005 | 2915482010 |
| Intracranial haemorrhage following injury with open intracranial wound and prolonged loss of consciousness (more than 24 hours) without return to pre-existing level | 4198361000006112 | 111671004 | 364052019 |
| Injury of multiple blood vessels of head AND/OR neck | 2601561000006119 | 6378004 | 11573012 |
| Multiple open fractures of skull AND/OR face without intracranial injury | 2829351000006110 | 20714001 | 34857016 |
| Closed traumatic subdural intracranial hemorrhage | 4854151000006118 | 209947002 | 2912529013 |
| Traumatic intracranial extradural haematoma | 5459201000006113 | 262949005 | 2916480013 |
| Traumatic subdural haemorrhage confined to region of tentorium | 7834501000006119 | 722628000 | 3327785019 |
| Extradural haemorrhage following injury with open intracranial wound AND loss of consciousness | 4198441000006117 | 111681000 | 364077018 |
| Other cerebral haemorrhage following injury NOS | 320897016 | 450418003 | 2916363013 |
| Non-traumatic hematoma of subdural space of neuraxis | 7375011000006110 | 449797005 | 2913008017 |
| Craniotomy and removal of haematoma from extradural space | 7312951000006114 | 445746008 | 2883205017 |
| Extradural haemorrhage following injury without open intracranial wound AND with moderate loss of consciousness (1-24 hours) | 3523671000006117 | 62973004 | 499442016 |
| Steroid injection of greater occipital nerve | 552331000000115 | 307951000000101 | 552331000000115 |
| Subdural haemorrhage following injury with open intracranial wound AND moderate loss of consciousness (1-24 hours) | 3965681000006115 | 90165008 | 3965681000006115 |
| Extradural hemorrhage following injury without open intracranial wound | 3196371000006118 | 43216008 | 72101019 |
| Traumatic brain injury with brief loss of consciousness | 4396291000006115 | 127299008 | 587015 |
| Traumatic intracranial subarachnoid haemorrhage | 5459271000006119 | 262955000 | 391048011 |
| Subdural hemorrhage following injury without open intracranial wound | 4854171000006111 | 209947002 | 2915477015 |
| Traumatic intracranial subarachnoid hemorrhage | 5459291000006118 | 262955000 | 391047018 |
| Non-traumatic spinal subdural hematoma | 6028581000006119 | 313304007 | 457178019 |
| Laceration of brain with open intracranial wound | 2863601000006110 | 22819008 | 38306018 |
| Injury of extracranial vessel of head | 7863871000006119 | 725002006 | 3436884013 |
| Cerebral hemorrhage following injury | 7378411000006114 | 450418003 | 2915447014 |
| Fracture of frontonasoethmoidal complex | 5461371000006118 | 263153003 | 391283015 |
| Chronic non-traumatic intracranial subdural hemorrhage | 7497201000006112 | 609382000 | 2959747017 |
| Brain stem contusion with open intracranial wound | 3784231000006114 | 78968003 | 131037016 |
| Traumatic hematoma of subdural space of neuraxis | 7374991000006117 | 449796001 | 2913015013 |
| Traumatic cranial subarachnoid haemorrhage | 5459301000006117 | 262955000 | 391049015 |
| Subdural hemorrhage following injury with open intracranial wound AND moderate loss of consciousness (1-24 hours) | 11903641000006118 | 90165008 | 149462011 |
| Traumatic intracranial hemorrhage | 7378301000006119 | 450410005 | 2915928019 |
| Lobotomy - brain | 5559181000006119 | 273991000 | 409720015 |
| Subarachnoid haemorrhage following injury with open intracranial wound AND concussion | 3284981000006111 | 48518008 | 495117010 |
| Extradural hemorrhage following injury without open intracranial wound AND with loss of consciousness | 3965891000006118 | 90178008 | 149485012 |

| | | | |
|---|---|---|---|
| Traumatic spinal extradural hematoma | 5456221000006111 | 262720008 | 390719016 |
| Cerebral haemorrhage following injury | 505322013 | 450418003 | 2916363013 |
| Traumatic cerebral haemorrhage | 505324014 | 450418003 | 2916058017 |
| Traumatic haematoma of subdural space of neuraxis | 7374981000006115 | 449796001 | 2912448014 |
| Extradural haemorrhage following injury with open intracranial wound | 3559941000006115 | 65189006 | 3559941000006115 |
| Subdural haemorrhage following injury with open intracranial wound and prolonged loss of consciousness (more than 24 hours) and without return to pre-existing conscious level | 124041000006115 | 111677000 | 364064010 |
| Open fracture of vault of skull with intracranial injury, with no loss of consciousness | 263781000006117 | 207707005 | 318137016 |
| Open fracture of base of skull with intracranial injury, with no loss of consciousness | 264161000006114 | 207745006 | 318189014 |
| Open traumatic subdural intracranial hemorrhage | 4854221000006117 | 209956005 | 2912899010 |
| Extradural haemorrhage following injury with open intracranial wound AND prolonged loss of consciousness (more than 24 hours) without return to pre-existing conscious level | 3901101000006118 | 86182004 | 3901101000006118 |
| Vestibular trauma due to head injury | 4062681000006118 | 95817008 | 512313010 |
| Brain injury without open intracranial wound AND with prolonged loss of consciousness (more than 24 hours) without return to pre-existing conscious level | 3267451000006119 | 47462004 | 79134018 |
| Intracranial haemorrhage following injury with open intracranial wound AND prolonged loss of consciousness (more than 24 hours) AND return to pre-existing conscious level | 3982281000006110 | 91216000 | 509357015 |
| Haemorrhage of brainstem due to traumatic injury | 7834641000006116 | 722632006 | 3332942015 |
| Intracranial haemorrhage following injury without open intracranial wound AND with loss of consciousness | 2793481000006115 | 18485009 | 478644014 |
| Extradural hemorrhage following injury with open intracranial wound AND prolonged loss of consciousness (more than 24 hours) without return to pre-existing conscious level | 11924641000006113 | 86182004 | 142911018 |
| Intracerebral injury | 4396221000006117 | 127294003 | 474102014 |
| Closed #skull bse + intracranial inj, unspec state of consc | 559961000006110 | 111603000 | 178853015 |
| Closed #skull NOS + intracranial inj, LOC unspec duration | 560161000006112 | 371162008 | 1209864014 |
| Closed #skull NOS + intracranial inj, unspec state of consc | 560181000006119 | 371162008 | 1209864014 |
| Closed #skull NOS no intracranial inj, 1-24hr loss of consc | 560221000006111 | 371162008 | 1209864014 |
| Closed #skull/face, mult + intracranial inj, 1-24hrs LOC | 560441000006115 | 5468008 | 10127011 |
| Closed #skull/face, mult + intracranial inj, unspec consc | 560461000006116 | 5468008 | 10127011 |
| Mult #skull/face + other bones, open + intracranial injury | 696121000006114 | 5468008 | 10127011 |
| Extradural haemorrhage following injury with open intracranial wound AND brief loss of consciousness (less than one hour) | 3692151000006113 | 73308006 | 3692151000006113 |
| Subdural hemorrhage following injury with open intracranial wound and prolonged loss of consciousness (more than 24 hours) and without return to pre-existing conscious level | 4198411000006116 | 111677000 | 61538012 |

| | | | |
|---|---|---|---|
| Evacuation of intracerebral hematoma | 2666411000006111 | 10458001 | 18195013 |
| Nontraumatic extradural hemorrhage | 6593111000006115 | 397809001 | 1774439018 |
| Closed fracture of base of skull with intracranial injury, with more than 24 hours loss of consciousness and return to pre-existing conscious level | 559991000006119 | 207730004 | 318174017 |
| Closed fracture of base of skull with intracranial injury, with 1-24 hours loss of consciousness | 560001000006112 | 207729009 | 318173011 |
| Closed fracture of base of skull with intracranial injury, with no loss of consciousness | 560021000006119 | 207727006 | 318171013 |
| Closed fracture of vault of skull with intracranial injury, with 1-24 hours loss of consciousness | 560301000006114 | 207691009 | 318121010 |
| Cortex contusion with open intracranial wound, with 1-24 hours loss of consciousness | 597701000006111 | 209848008 | 320616011 |
| Intracranial nontraumatic haemorrhage of foetus and newborn | 744871000006114 | 206417009 | 2791314016 |
| Hind brain contusion with open intracranial wound, with less than 1 hour loss of consciousness | 823401000006116 | 209884001 | 320655012 |
| Hind brain laceration with open intracranial wound, with less than 1 hour loss of consciousness | 823531000006111 | 209903008 | 320675015 |
| Subarachnoid hemorrhage following injury without open intracranial wound AND with loss of consciousness | 3629581000006110 | 69458009 | 115392016 |
| Traumatic subdural haematoma with open intracranial wound | 459514014 | 315047001 | 459514014 |
| Intracranial haematoma following injury | 486896012 | 37955001 | 486896012 |
| Open fracture of vault of skull with intracranial injury, with more than 24 hours loss of consciousness and return to pre-existing conscious level | 52931000006111 | 207710003 | 318140016 |
| Brain stem contusion with open intracranial wound AND loss of consciousness | 3579631000006117 | 66393002 | 110266011 |
| Intracranial haemorrhage following injury without open intracranial wound AND with prolonged loss of consciousness (more than 24 hours) AND return to pre-existing conscious level | 3776591000006112 | 78477003 | 503848012 |
| Intracranial haemorrhage following injury without open intracranial wound AND with no loss of consciousness | 3098131000006113 | 37134004 | 486631018 |
| Open #skull/face, mult + intracran inj, LOC unspec duration | 263911000006110 | 5468008 | 10127011 |
| Open #skull/face, mult + intracranial inj, <1hr LOC | 263921000006119 | 5468008 | 10127011 |
| Open #skull bse + intracranial injury, LOC unspec duration | 264151000006112 | 111607004 | 178855010 |
| Hind brain laceration with open intracranial wound, with no loss of consciousness | 823561000006119 | 209902003 | 320674016 |
| Subarachnoid haemorrhage following injury with open intracranial wound AND prolonged loss of consciousness (more than 24 hours) AND return to pre-existing conscious level | 123281000006117 | 73439007 | 502436011 |
| Extradural haemorrhage following injury with open intracranial wound AND prolonged loss of consciousness (more than 24 hours) AND return to pre-existing conscious level | 659961000006116 | 81520002 | 504713019 |
| Intracranial haemorrhage following injury with open intracranial wound | 3765291000006112 | 77768006 | 503672016 |
| Extradural hemorrhage following injury with open intracranial wound AND brief loss of consciousness (less than one hour) | 11924661000006112 | 73308006 | 121738013 |
| Intracranial haemorrhage following injury without open intracranial wound AND with concussion | 3781101000006114 | 78757009 | 503910018 |

| | | | |
|---|---|---|---|
| Intracranial haemorrhage following injury with moderate loss of consciousness | 4396471000006117 | 127310009 | 474111014 |
| Birth brain damage NOS | 513921000006111 | 240312009 | 359984010 |
| Closed #skull bse no intracranial inj, LOC unspec duration | 560031000006116 | 27644009 | 46274013 |
| Closed #skull bse no intracranial injury, no loss of consc | 560091000006117 | 27644009 | 46274013 |
| Closed #skull bse no intracranial injury, unspec state consc | 560101000006111 | 24063002 | 40395011 |
| Closed #skull NOS + intracranial inj + concussion unspec | 560111000006114 | 371162008 | 1209864014 |
| Closed #skull NOS no intracranial inj + concussion unspec | 560191000006116 | 371162008 | 1209864014 |
| Closed #skull/face, mult + intracranial inj, <1hr LOC | 560431000006113 | 5468008 | 10127011 |
| Cortex cont + open intracranial wound + concussion unspec | 597711000006114 | 209843004 | 320611018 |
| Cortex lacn no open intracranial wnd + >24hr LOC + recovery | 597931000006118 | 78914008 | 130942015 |
| Cortex lacn no open intracranial wnd +>24hr LOC not restored | 597961000006110 | 78914008 | 130942015 |
| Non-traumatic intracranial subdural haematoma | 5656921000006118 | 281864001 | 419994017 |
| Multiple traumatic haemorrhages of brain tissue | 7834651000006119 | 722633001 | 3332945018 |
| Epidural haemorrhage | 505381016 | 82999001 | 505381016 |
| Subarachnoid hemorrhage following injury with open intracranial wound AND loss of consciousness | 3727881000006110 | 75507000 | 125412015 |
| Subdural haemorrhage following injury with open intracranial wound AND loss of consciousness | 3237311000006119 | 45659008 | 494311012 |
| Open fracture of base of skull with intracranial injury, with 1-24 hours loss of consciousness | 264141000006110 | 207747003 | 318191018 |
| Brain contusion with open intracranial wound, with more than 24 hours loss of consciousness and return to pre-existing conscious level | 524301000006112 | 209923009 | 320695014 |
| Closed fracture of base of skull with intracranial injury, with more than 24 hours loss of consciousness without return to pre-existing conscious level | 559951000006113 | 207731000 | 318175016 |
| Cortex contusion with open intracranial wound, with less than 1 hour loss of consciousness | 597691000006111 | 209847003 | 320615010 |
| Extradural haemorrhage following injury with open intracranial wound, with no loss of consciousness | 660021000006112 | 209978003 | 320817017 |
| Subdural hemorrhage following injury | 4854341000006117 | 209987007 | 320838011 |
| Closed traumatic subdural intracranial haemorrhage | 4854161000006116 | 209947002 | 2912797018 |
| Non-traumatic haematoma of subdural space of neuraxis | 7375001000006112 | 449797005 | 2912549016 |
| Cerebellar laceration with open intracranial wound AND moderate loss of consciousness (1-24 hours) | 3879281000006119 | 84803006 | 140595014 |
| Intracranial haemorrhage following injury with prolonged loss of consciousness AND return to pre-existing conscious level | 4396491000006116 | 127311008 | 474112019 |
| Subarachnoid haemorrhage following injury without open intracranial wound | 402929011 | 28048009 | 483737017 |
| Subdural hemorrhage - nontraumatic | 4777911000006112 | 195176009 | 300293019 |
| Extradural hemorrhage after injury | 5459171000006114 | 262949005 | 391027014 |
| Subdural haemorrhage following injury with open intracranial wound | 4854241000006112 | 209956005 | 2916362015 |

| | | | |
|---|---|---|---|
| Intracranial haemorrhage following injury without intracranial wound AND with moderate loss of consciousness (1-24 hours) | 3589021000006119 | 66976009 | 500582010 |
| Craniotomy with treatment of penetrating wound of brain | 3786211000006110 | 79093007 | 131239012 |
| Subarachnoid hemorrhage following injury without open intracranial wound AND with moderate loss of consciousness (1-24 hours) | 4198391000006116 | 111673001 | 195613013 |
| Brain stem laceration without open intracranial wound AND with prolonged loss of consciousness (more than 24 hours) without return to pre-existing conscious level | 3596181000006113 | 67378005 | 111973015 |
| Brain stem contusion with open intracranial wound AND prolonged loss of consciousness (more than 24 hours) AND return to pre-existing conscious level | 2767251000006115 | 16837005 | 28508011 |
| Subdural hemorrhage following injury with open intracranial wound | 4854231000006119 | 209956005 | 2915474010 |
| Concussion of periodontal ligament | 5462911000006115 | 263275009 | 391448016 |
| Extradural haemorrhage following injury with open intracranial wound AND concussion | 2951801000006112 | 28155008 | 483768018 |
| Hind brain contusion with open intracranial wound, with no loss of consciousness | 823411000006118 | 209883007 | 320654011 |
| Hind brain laceration with open intracranial wound, with 1-24 hours loss of consciousness | 823541000006118 | 209904002 | 320676019 |
| Subdural hemorrhage following injury without open intracranial wound AND with prolonged loss of consciousness (more than 24 hours) without return to pre-existing conscious level | 3652491000006116 | 70861009 | 117705011 |
| Traumatic extradural hematoma | 5459161000006119 | 262949005 | 391026017 |
| Subarachnoid haemorrhage following injury without open intracranial wound AND with concussion | 2567821000006112 | 4332009 | 493572019 |
| Intracranial hemorrhage following injury with open intracranial wound AND no loss of consciousness | 2624491000006117 | 7819003 | 13934013 |
| Intracranial haemorrhage following injury with intracranial wound AND moderate loss of consciousness (1-24 hours) | 3055541000006111 | 34501004 | 485859011 |
| Auditory cranial nerve injury | 511287019 | 9441002 | 511287019 |
| Traumatic cerebral oedema with open intracranial wound | 455458019 | 311826007 | 455458019 |
| Injury of intracranial vessel of head | 7815641000006111 | 721328009 | 3324974017 |
| Oth cerebral laceration/contusion + open intracranial wound | 39841000006117 | 269144002 | 402921014 |
| Subdural h'ge inj no open intracranial wound+<1hr loss consc | 124181000006110 | 209947002 | 320752018 |
| Extradural haemorrhage following injury without open intracranial wound | 402930018 | 43216008 | 493552012 |
| Open fracture of vault of skull with intracranial injury, with more than 24 hours loss of consciousness without return to pre-existing conscious level | 264421000006112 | 207711004 | 318141017 |
| Primary traumatic hemorrhage of brainstem | 7840841000006112 | 723138000 | 3334696017 |
| Traumatic brain injury of unknown intent | 7638391000006118 | 708728007 | 3083294016 |
| Intracranial hemorrhage following injury with open intracranial wound AND prolonged loss of consciousness (more than 24 hours) AND return to pre-existing conscious level | 3982291000006113 | 91216000 | 151139013 |
| Sequela of traumatic intracranial haemorrhage | 8031551000006111 | 100581000119102 | 2967472012 |

| | | | |
|---|---|---|---|
| Intracranial hemorrhage following injury without open intracranial wound AND with no loss of consciousness | 3098141000006115 | 37134004 | 61953011 |
| Brain stem laceration with open intracranial wound AND no loss of consciousness | 2804791000006111 | 19210000 | 32368010 |
| Intracranial inj NOS no open intracran wnd + unspec consc | 768821000006111 | 127296001 | 584010 |
| Intracranial injury NOS + open intracranial wound | 768911000006110 | 127296001 | 584010 |
| Intracranial injury NOS no open intracranial wound | 768921000006119 | 127296001 | 584010 |
| Injury to blood vessels of head and neck | 774541000006119 | 85564003 | 141827011 |
| Hind brain lacn + open intracranial wnd + concussion unspec | 823551000006116 | 209900006 | 320672017 |
| Hind brain lacn + open intracranial wnd + unspec state consc | 823571000006114 | 209900006 | 320672017 |
| Subdural h'ge inj no open intracran wound+concussion unspec | 124151000006119 | 209947002 | 320752018 |
| Subdural h'ge inj no open intracranial wnd + unspec consc | 124161000006117 | 209947002 | 320752018 |
| Subdural h'ge inj no open intracranial wnd+>24 LOC +recovery | 124171000006112 | 209947002 | 320752018 |
| Traumatic cerebral haemorrhage | 12757931000006114 | 82894007 | 505324014 |
| Traumatic cerebral hemorrhage | 12757881000006115 | 82894007 | 137503010 |
| Complex wound of head with avulsive loss of part of skull and cranial contents | 7840931000006112 | 723143007 | 3334711017 |
| Nontraumatic subarachnoid intracranial haemorrhage | 5518111000006112 | 270907008 | 3449602014 |
| Hemorrhage of brainstem due to traumatic injury | 7834631000006114 | 722632006 | 3332941010 |
| Subdural hemorrhage following injury with open intracranial wound AND brief loss of consciousness (less than one hour) | 2976661000006116 | 29635000 | 49572012 |
| Nontraumatic intracranial hemorrhage | 3533577011 | 738779002 | 3533577011 |
| Traumatic intracranial subdural hematoma | 5459241000006110 | 262952002 | 391038014 |
| Secondary traumatic hemorrhage of brainstem | 7840861000006111 | 723139008 | 3334699012 |
| Oth cereb h'ge inj + open intracran wnd+concussion unspec | 39711000006113 | 450418003 | 2915288012 |
| Oth cerebral h'ge inj no open intracranial wnd+no loss consc | 39831000006110 | 450418003 | 2915288012 |
| Subarach h'ge inj no open intracran wnd+LOC unspec duration | 123331000006114 | 28048009 | 46957015 |
| Subdural h'ge inj + open intracranial wnd+concussion unspec | 124081000006114 | 209956005 | 320770015 |
| Extradural h'ge inj no open intracran wnd+concussion unspec | 660071000006113 | 262949005 | 391026017 |
| Cortex contusion without open intracranial wound AND with loss of consciousness | 2777751000006115 | 17498002 | 29575016 |
| Cerebellar laceration with open intracranial wound AND brief loss of consciousness (less than one hour) | 2651541000006113 | 9501009 | 16628012 |
| Focal hemorrhagic contusion of cerebrum | 7834271000006110 | 722615007 | 3332902013 |

##### Table S19. Spinal Injury Aurum codes

| Term | Medcode ID | SNOMED CT Concept ID | SNOMED CT Description ID |
|---|---|---|---|
| Injury of spinal cord vasculature | 5456141000006113 | 262715008 | 390711018 |

#### Table S20. Eye Injury Aurum codes

| Term | Medcode ID | SNOMED CT Concept ID | SNOMED CT Description ID |
|---|---|---|---|
| Haemophthalmos, except current injury | 4771081000006111 | 193287005 | 2550868017 |
| Hemophthalmos, except current injury | 4771091000006114 | 193287005 | 2550869013 |
| Perforating injury of both eyeballs | 3515113016 | 735658005 | 3515113016 |
| Haemophthalmos without current injury | 4771041000006117 | 193287005 | 2547718010 |
| Bilateral penetrating injury of eyeball | 3515111019 | 735657000 | 3515111019 |
| Repair of penetrating eye injury | 8255451000006117 | 800491000000104 | 1789891000000113 |
| Haemophthalmos (excluding current injury) | 297686013 | 193287005 | 297686013 |
| Penetrating injury of both eyeballs | 3515110018 | 735657000 | 3515110018 |
| Repair of penetrating eye injury | 11999811000006115 | 763097008 | 3637861018 |
| Hemophthalmos without current injury | 4771071000006113 | 193287005 | 2547719019 |
| Haemorrhage into cornea | 5029221000006110 | 231916005 | 347514014 |

#### Table S21. Estimated glomerular filtration rate Aurum codes

| Term | Medcode ID | SNOMED CT Concept ID | SNOMED CT Description ID |
|---|---|---|---|
| EgfrUsingCreatinine(ckd-epi)Per1.73SquareMetres | 1942831000006114 | 1011481000000105 | 2579541000000113 |
| EstimatedCreatinineClearance | 2470380013 | 395680003 | 2470380013 |
| EstimatedCreatinineClearance(cockcroft-gaultFormula) | 2465711000000116 | 968191000000100 | 2465711000000116 |
| GlomerularFiltrationRate | 133205018 | 80274001 | 133205018 |
| SerumCreatinine | 380389013 | 1000731000000107 | 2577271000000111 |
| GfrCalculatedAbbreviatedMdrd | 976481000006110 | 1020291000000106 | 2569781000000116 |

#### Table S22. Azole Aurum codes

| Term | Medcode ID | SNOMED CT Concept ID | SNOMED CT Description ID |
|---|---|---|---|
| Atherosclerosis aorta | 3830431000006112 | 81817003 | 2923328010 |
| Aortic thromboembolism | 5560171000006116 | 274101000 | 409861010 |
| Aortoiliac atherosclerosis | 5057931000006110 | 233956002 | 350530011 |
| Aortic atherosclerosis | 218531000000113 | 81817003 | 135728018 |

#### Table S23. Dementia Aurum codes

| Term | Medcode ID | SNOMED CT Concept ID | SNOMED CT Description ID |
|---|---|---|---|
| H/O: dementia | 251625013 | 161465002 | 251625013 |
| [X]Predominantly cortical dementia | 423221000006117 | 56267009 | 93568017 |
| [X]Presenile dementia,Alzheimer's type | 423351000006115 | 416780008 | 2957138011 |
| [X]Primary degen dementia, Alzheimer's type, presenile onset | 423391000006114 | 416780008 | 2957119019 |
| Lewy body variant of Alzheimer's disease | 3802621000006119 | 80098002 | 132894011 |
| SDLT - Senile dementia of the Lewy body type | 3802631000006116 | 80098002 | 1234435012 |
| [X]Senile dementia,Alzheimer's type | 425901000006116 | 416975007 | 2957124016 |
| Dementia associated with alcoholism | 2502971000006115 | 281004 | 1557018 |
| Senile dementia with depressive or paranoid features NOS | 294647019 | 191457008 | 294644014 |
| [X]Vascular dementia, unspecified | 295681016 | 429998004 | 2770951017 |
| [X]Dementia in other specified diseases classif elsewhere | 295690011 | 191519005 | 2547722017 |
| Dementia infantilis | 3670601000006111 | 71961003 | 119578017 |
| Dementia co-occurrent and due to Pick's disease | 8009541000006119 | 21921000119103 | 3333186018 |
| Dementia due to Picks disease | 8009521000006114 | 21921000119103 | 2987132018 |
| ADC - Acquired immune deficiency syndrome dementia complex | 6973451000006113 | 421529006 | 2622766016 |
| Alzheimer's disease with early onset | 499946014 | 416780008 | 2957141019 |
| Alzheimer's disease with late onset | 500317011 | 416975007 | 2957124016 |
| Dementia in Alzheimer's disease with early onset, other symptoms, predominantly delusional | 1971541000006114 | 1971541000006105 | 1971541000006114 |
| Dementia in Alzheimer's disease with early onset, other symptoms, predominantly hallucinatory | 1971771000006112 | 1971771000006108 | 1971771000006112 |
| Unspecified dementia, other symptoms, predominantly delusional | 1972041000006114 | 1972041000006105 | 1972041000006114 |
| Unspecified dementia, other symptoms, predominantly hallucinatory | 1972061000006113 | 1972061000006109 | 1972061000006113 |
| Dementia in Alzheimer's disease with late onset, other symptoms, predominantly delusional | 1972181000006119 | 1972181000006103 | 1972181000006119 |
| Dementia in Alzheimer's disease with late onset, other symptoms, predominantly depressive | 1972201000006118 | 1972201000006102 | 1972201000006118 |
| Dementia in Alzheimer's disease with late onset, other mixed symptoms | 1972211000006115 | 1972211000006104 | 1972211000006115 |
| Dementia in Alzheimer's dis, atypical or mixed type, other symptoms, predominantly hallucinatory | 1972291000006113 | 1972291000006109 | 1972291000006113 |
| Vascular dementia of acute onset, other symptoms, predominantly hallucinatory | 1972521000006114 | 1972521000006105 | 1972521000006114 |
| Subcortical vascular dementia, other symptoms, predominantly depressive | 1972771000006116 | 1972771000006100 | 1972771000006116 |
| Mixed cortical and subcortical vascular dementia, other symptoms, predominantly hallucinatory | 1972871000006113 | 1972871000006109 | 1972871000006113 |
| Mixed cortical and subcortical vascular dementia, other symptoms, predominantly depressive | 1972911000006111 | 1972911000006107 | 1972911000006111 |
| Mental and behav dis due to hallucinogens: resid & late-onset psychot dis, dementia | 1973711000006116 | 1973711000006100 | 1973711000006116 |
| Presenile dementia | 21256010 | 12348006 | 21256010 |
| Senile dementia | 26545010 | 15662003 | 26545010 |

| | | | |
|---|---|---|---|
| Alzheimer's disease | 45046017 | 26929004 | 45046017 |
| VAD - Vascular dementia | 3414251000006112 | 56267009 | 497560014 |
| Dementia due to Creutzfeldt-Jakob disease | 7103601000006116 | 429458009 | 2693170019 |
| Primary degenerative dementia of the Alzheimer type, late onset | 6900191000006112 | 416975007 | 2554010014 |
| Transmissible virus dementia | 2511001000006119 | 792004 | 2164150017 |
| Binswanger's dementia | 3964611000006111 | 90099008 | 149345019 |
| Lewy body disease | 745381000006119 | 80098002 | 132893017 |
| Mental and behav dis due to other stimulants inc caffeine: resid/late-onset psycht dis, dementia | 1976091000006118 | 1976091000006102 | 1976091000006118 |
| Primary degenerative dementia of the Alzheimer type, early onset | 6897221000006113 | 416780008 | 2553939016 |
| Acquired immune deficiency syndrome dementia complex | 6973461000006110 | 421529006 | 2622767013 |
| Dementia associated with AIDS | 6973421000006116 | 421529006 | 2617355019 |
| MID - Multi-infarct dementia | 3414231000006117 | 56267009 | 497558012 |
| Arteriosclerotic dementia NOS | 294656010 | 56267009 | 497559016 |
| [X]Alzheimer's dementia unspec | 363021000006113 | 26929004 | 45046017 |
| [D] Dementia with Lewy bodies | 914931000006119 | 914931000006103 | 914931000006119 |
| [D] Dementia | 914941000006112 | 914941000006108 | 914941000006112 |
| [X] Senile dementia NOS | 359141000006111 | 15662003 | 26545010 |
| [X]Alzheimer's disease type 2 | 363041000006118 | 416780008 | 2957119019 |
| [X]Dementia in human immunodef virus [HIV] disease | 376571000006116 | 421529006 | 2622767013 |
| [X]Dementia infantalis | 376631000006115 | 71961003 | 119576018 |
| Uncomplicated senile dementia | 294635013 | 191449005 | 294635013 |
| Presenile dementia with delirium | 294638010 | 191452002 | 294638010 |
| Uncomplicated arteriosclerotic dementia | 294652012 | 191463004 | 294652012 |
| Arteriosclerotic dementia with delirium | 294653019 | 191464005 | 294653019 |
| Arteriosclerotic dementia with paranoia | 294654013 | 191465006 | 294654013 |
| Arteriosclerotic dementia with depression | 294655014 | 191466007 | 294655014 |
| Dementia | 882171000006115 | 268612007 | 882171000006115 |
| Dementia in Alzheimer's disease - type 2 | 6897271000006114 | 416780008 | 2957156011 |
| LBD - Lewy body disease | 3802641000006114 | 80098002 | 1234436013 |
| DLBD - Diffuse Lewy body disease | 3802661000006113 | 80098002 | 1234438014 |
| Primary degenerative dementia of the Alzheimer type, senile onset | 6900181000006114 | 416975007 | 2549518019 |
| Dementia of the Alzheimers type with early onset | 6897241000006118 | 416780008 | 2957133019 |
| Presenile dementia, Alzheimer's type | 6897251000006116 | 416780008 | 2957138011 |
| [X]Dementia in other diseases classified elsewhere | 295684012 | 191519005 | 2547722017 |
| [X]Dementia in Pick's disease | 295685013 | 21921000119103 | 2968126015 |
| [X]Dementia in Creutzfeldt-Jakob disease | 295686014 | 429458009 | 2695885013 |
| [X]Dementia in Parkinson's disease | 295688010 | 425390006 | 2921040013 |
| AD - Alzheimer's disease | 2931231000006118 | 26929004 | 1225144019 |
| Dementia of the Alzheimers type, late onset | 6900201000006110 | 416975007 | 2957123010 |
| [X]Dementia in Alzheimer's disease | 295668011 | 26929004 | 45046017 |

| | | | |
|---|---|---|---|
| [X]Dementia in Alzheimer's disease, unspecified | 295672010 | 26929004 | 45046017 |
| [X] Senile dementia, depressed or paranoid type | 359151000006113 | 191457008 | 294644014 |
| Nuchal dystonia-dementia syndrome | 2966081000006114 | 28978003 | 48514014 |
| Dementia in Alzheimer's disease with early onset, without additional symptoms | 1971401000006111 | 1971401000006107 | 1971401000006111 |
| Mental & behav dis due to seds/hypntcs: resid & late-onset psychot dis, dementia | 1971661000006114 | 1971661000006105 | 1971661000006114 |
| Dementia in Alzheimer's dis, atypical or mixed type, other symptoms, predominantly depressive | 1972311000006112 | 1972311000006108 | 1972311000006112 |
| Dementia in Alzheimer's dis, atypical or mixed type, other mixed symptoms | 1972341000006111 | 1972341000006107 | 1972341000006111 |
| Dementia in Alzheimer's disease, unspecified, without additional symptoms | 1972371000006115 | 1972371000006104 | 1972371000006115 |
| Mental & behav dis due to cannabinoids: resid & late-onset psychot dis, dementia | 1972431000006113 | 1972431000006109 | 1972431000006113 |
| Vascular dementia of acute onset, other symptoms, predominantly delusional | 1972501000006116 | 1972501000006100 | 1972501000006116 |
| Other vascular dementia, other symptoms, predominantly depressive | 1973381000006112 | 1973381000006108 | 1973381000006112 |
| [RFC] Dementia | 939491000006118 | 939491000006102 | 939491000006118 |
| Drug-induced dementia | 294688019 | 191493005 | 294688019 |
| JCD - Jakob-Creutzfeldt disease | 2510991000006115 | 792004 | 1234326016 |
| [X] Unspecified dementia | 359241000006119 | 52448006 | 87274019 |
| Senile dementia-acute confused | 882211000006118 | 191461002 | 882211000006118 |
| [X]Dementia in Alzheimer's disease with early onset | 376531000006119 | 416780008 | 2957119019 |
| [X]Dementia in Alzheimer's disease with late onset | 376541000006112 | 416975007 | 2957137018 |
| [X]Vascular dementia | 431681000006117 | 429998004 | 2770951017 |
| [X]Vascular dementia of acute onset | 431691000006119 | 230285003 | 345110014 |
| Multi infarct dementia | 3414261000006114 | 56267009 | 2921000019 |
| Alzheimer dementia | 2931251000006113 | 26929004 | 3424952012 |
| [D] Vascular dementia | 914921000006117 | 914921000006101 | 914921000006117 |
| [D] Dementia in Alzheimer's disease | 914951000006114 | 914951000006105 | 914951000006114 |
| Dementia associated with Parkinson Disease | 7043661000006117 | 425390006 | 2842012010 |
| Senile dementia with delirium | 294648012 | 191461002 | 294648012 |
| [X]Dementia in Huntington's disease | 295687017 | 442344002 | 2820374011 |
| Alcoholic dementia NOS | 346929012 | 281004 | 1225776018 |
| Uncomplicated presenile dementia | 294637017 | 191451009 | 294637017 |
| Senile dementia with depression | 294646011 | 191459006 | 294646011 |
| [X] Primary degenerative dementia NOS | 359101000006114 | 279982005 | 417465012 |
| [X]Alzheimer's disease type 1 | 363031000006111 | 416975007 | 2957137018 |
| Dementia due to Huntingtons disease | 7263021000006116 | 442344002 | 2951827019 |
| [X]Primary degen dementia of Alzheimer's type, senile onset | 423381000006111 | 416975007 | 2957137018 |
| Presenile dementia NOS | 294643015 | 12348006 | 21256010 |
| Mental & behav dis due to use opioids: resid & late-onset psychot dis, dementia | 1971701000006118 | 1971701000006102 | 1971701000006118 |
| Unspecified dementia, without additional symptoms | 1972021000006119 | 1972021000006103 | 1972021000006119 |
| Unspecified dementia, other mixed symptoms | 1972081000006115 | 1972081000006104 | 1972081000006115 |

| | | | |
|---|---|---|---|
| Dementia in Alzheimer's disease with early onset, other symptoms, predominantly depressive | 1972131000006115 | 1972131000006104 | 1972131000006115 |
| Dementia in Alzheimer's disease with early onset, other mixed symptoms | 1972141000006113 | 1972141000006109 | 1972141000006113 |
| Dementia in Alzheimer's dis, atypical or mixed type, without additional symptoms | 1972231000006114 | 1972231000006105 | 1972231000006114 |
| Dementia in Alzheimer's dis, atypical or mixed type, other symptoms, predominantly delusional | 1972251000006119 | 1972251000006103 | 1972251000006119 |
| Dementia in Alzheimer's disease, unspecified, other mixed symptoms | 1972471000006111 | 1972471000006107 | 1972471000006111 |
| Vascular dementia of acute onset, without additional symptoms | 1972481000006114 | 1972481000006105 | 1972481000006114 |
| Multi-infarct dementia, other symptoms, predominantly delusional | 1972621000006113 | 1972621000006109 | 1972621000006113 |
| Multi-infarct dementia, other symptoms, predominantly depressive | 1972661000006119 | 1972661000006103 | 1972661000006119 |
| Subcortical vascular dementia, without additional symptoms | 1972711000006113 | 1972711000006109 | 1972711000006113 |
| Subcortical vascular dementia, other symptoms, predominantly delusional | 1972731000006119 | 1972731000006103 | 1972731000006119 |
| Other vascular dementia, without additional symptoms | 1973221000006118 | 1973221000006102 | 1973221000006118 |
| Other vascular dementia, other symptoms, predominantly delusional | 1973271000006117 | 1973271000006101 | 1973271000006117 |
| Other vascular dementia, other mixed symptoms | 1973401000006112 | 1973401000006108 | 1973401000006112 |
| Mental & behav dis due to use cocaine: resid & late-onset psychot dis, dementia | 1973941000006119 | 1973941000006103 | 1973941000006119 |
| Multi infarct dementia | 696161000006115 | 56267009 | 93568017 |
| Subcortical atherosclerotic dementia | 3964661000006114 | 90099008 | 1235536019 |
| Dementia associated with acquired immunodeficiency syndrome | 6973471000006115 | 421529006 | 2970885015 |
| CLBD - Cortical Lewy body disease | 3802681000006115 | 80098002 | 1234440016 |
| [X]Lewy body dementia | 299641000000112 | 80098002 | 132893017 |
| Mental & behav dis due to tobacco: resid & late-onset psychot dis, dementia | 1974931000006114 | 1974931000006105 | 1974931000006114 |
| Mental & behav dis due to use alcohol: resid & late-onset psychot dis, dementia | 1975591000006119 | 1975591000006103 | 1975591000006119 |
| Dementia associated with Parkinson's Disease | 7043651000006119 | 425390006 | 2645368017 |
| Organic dementia | 3350441000006115 | 52448006 | 2970981011 |
| Senile/presenile dementia | 148381000006115 | 52448006 | 87274019 |
| Senile and presenile dementias | 882191000006119 | 268612007 | 882191000006119 |
| Jakob-Creutzfeldt disease | 2386018 | 792004 | 2386018 |
| Pick's disease | 22408016 | 13092008 | 22408016 |
| [X]Delirium superimposed on dementia | 295714013 | 2776000 | 5694012 |
| Dementia due to Pick disease | 8009531000006112 | 21921000119103 | 2987352013 |
| Primary degenerative dementia of the Alzheimer type, presenile onset | 6897211000006117 | 416780008 | 2549310013 |
| [RFC] Alzheimer's disease | 905791000006115 | 905791000006104 | 905791000006115 |
| Presenile dementia with depression | 294642013 | 191455000 | 294642013 |
| Senile dementia with depressive or paranoid features | 294644014 | 191457008 | 294644014 |
| Senile dementia with paranoia | 294645010 | 191458003 | 294645010 |
| Dementia in Alzheimer's disease with late onset, other symptoms, predominantly hallucinatory | 1972191000006116 | 1972191000006100 | 1972191000006116 |

| | | | |
|---|---|---|---|
| Vascular dementia of acute onset, other symptoms, predominantly depressive | 1972541000006119 | 1972541000006103 | 1972541000006119 |
| Multi-infarct dementia, without additional symptoms | 1972601000006115 | 1972601000006104 | 1972601000006115 |
| Subcortical vascular dementia, other mixed symptoms | 1972791000006115 | 1972791000006104 | 1972791000006115 |
| Mixed cortical and subcortical vascular dementia, without additional symptoms | 1972821000006112 | 1972821000006108 | 1972821000006112 |
| Mental and behav dis due to vol solvents: resid & late-onset psychotic dis, dementia | 1973171000006112 | 1973171000006108 | 1973171000006112 |
| Other vascular dementia, other symptoms, predominantly hallucinatory | 1973341000006118 | 1973341000006102 | 1973341000006118 |
| Vascular dementia, unspecified, without additional symptoms | 1973461000006113 | 1973461000006109 | 1973461000006113 |
| Vascular dementia, unspecified, other symptoms, predominantly hallucinatory | 1973531000006117 | 1973531000006101 | 1973531000006117 |
| SD - Senile dementia | 2748441000006111 | 15662003 | 1221222011 |
| [X]Other vascular dementia | 295680015 | 429998004 | 2770951017 |
| [X]Other Alzheimer's disease | 299325013 | 26929004 | 45046017 |
| [X] Presenile dementia NOS | 359081000006118 | 12348006 | 21256010 |
| [X]Arteriosclerotic dementia | 363791000006112 | 56267009 | 497559016 |
| [X]Dementia in Alzheimer's dis, atypical or mixed type | 295671015 | 26929004 | 45046017 |
| Dementia in Alzheimer's disease, unspecified, other symptoms, predominantly delusional | 1972401000006117 | 1972401000006101 | 1972401000006117 |
| Dementia in Alzheimer's disease, unspecified, other symptoms, predominantly hallucinatory | 1972421000006110 | 1972421000006106 | 1972421000006110 |
| Multi-infarct dementia, other symptoms, predominantly hallucinatory | 1972641000006118 | 1972641000006102 | 1972641000006118 |
| Multi-infarct dementia, other mixed symptoms | 1972681000006112 | 1972681000006108 | 1972681000006112 |
| Vascular dementia, unspecified, other symptoms, predominantly depressive | 1973551000006112 | 1973551000006108 | 1973551000006112 |
| Vascular dementia, unspecified, other mixed symptoms | 1976831000006111 | 1976831000006107 | 1976831000006111 |
| Arteriosclerotic dementia | 497559016 | 56267009 | 497559016 |
| Dementia due to Huntington disease | 7263011000006112 | 442344002 | 2820375012 |
| Dementia in Alzheimer's disease - type 1 | 6900241000006112 | 416975007 | 2957155010 |
| Unspecified dementia, other symptoms, predominantly depressive | 1972071000006118 | 1972071000006102 | 1972071000006118 |
| Dementia in Alzheimer's disease with late onset, without additional symptoms | 1972171000006117 | 1972171000006101 | 1972171000006117 |
| Dementia in Alzheimer's disease, unspecified, other symptoms, predominantly depressive | 1972451000006118 | 1972451000006102 | 1972451000006118 |
| Vascular dementia of acute onset, other mixed symptoms | 1972571000006110 | 1972571000006106 | 1972571000006110 |
| Subcortical vascular dementia, other symptoms, predominantly hallucinatory | 1972751000006114 | 1972751000006105 | 1972751000006114 |
| Mixed cortical and subcortical vascular dementia, other symptoms, predominantly delusional | 1972831000006110 | 1972831000006106 | 1972831000006110 |
| Mixed cortical and subcortical vascular dementia, other mixed symptoms | 1972931000006117 | 1972931000006101 | 1972931000006117 |
| Vascular dementia, unspecified, other symptoms, predominantly delusional | 1973501000006113 | 1973501000006109 | 1973501000006113 |
| Mental and behav dis mlti drg use/oth psych subs: resid/late psychot dis, dementia | 1974271000006119 | 1974271000006103 | 1974271000006119 |
| Dementia confirmed | 1823871000006112 | 1823871000006108 | 1823871000006112 |
| AIDS - Acquired immune deficiency syndrome dementia complex | 6973441000006111 | 421529006 | 2622765017 |

| | | | |
|---|---|---|---|
| Alcohol-induced persisting dementia | 2502981000006117 | 281004 | 1558011 |
| Acquired immune deficiency syndrome-related dementia | 6973431000006118 | 421529006 | 2622764018 |
| Dementia paralytica | 3341641000006113 | 51928006 | 86446018 |
| SDAT - Senile dementia, Alzheimer's type | 6900221000006117 | 416975007 | 2957134013 |
| [X]Mixed cortical and subcortical vascular dementia | 398571000006112 | 230287006 | 345112018 |
| [X]Alcoholic dementia NOS | 362941000006113 | 281004 | 1225776018 |
| Dementia of the Lewy body type | 3802651000006111 | 80098002 | 1234437016 |
| [X]Subcortical vascular dementia | 428201000006119 | 230286002 | 345111013 |
| Cortical Lewy body disease | 3802671000006118 | 80098002 | 1234439018 |
| Paralytic dementia | 3341701000006116 | 51928006 | 1231057014 |
| [X]Multi-infarct dementia | 399031000006111 | 56267009 | 93568017 |
| Other alcoholic dementia | 401760017 | 281004 | 1225776018 |
| Senile dementia - simple type | 882201000006116 | 191449005 | 882201000006116 |
| Presenile dementia with paranoia | 294641018 | 191454001 | 294641018 |
| Dementia in conditions EC | 294718018 | 191519005 | 294718018 |

### Chapter 2. Code lists for trial outcomes

#### Table S24. Cardivascular mortality HES (ICD10) codes

| ICD10 codes | Term |
|---|---|
| I00-I02 | Acute rheumatic fever |
| I05-I09 | Chronic rheumatic heart diseases |
| I10-I15 | Hypertensive diseases |
| I20-I25 | Ischaemic heart diseases |
| I26-I28 | Pulmonary heart disease and diseases of pulmonary circulation |
| I30-I52 | Other forms of heart disease |
| I60-I69 | Cerebrovascular diseases |
| I70-I78 (exclude I78.0, I78.1) | Diseases of arteries, arterioles and capillaries |
| G46 | Vascular syndromes of brain in cerebrovascular diseases |
| G45 | Transient cerebral ischaemic attacks and related syndromes |
| R57.0 | Cardiogenic shock |
| K76.1 | Chronic passive congestion of liver |
| J81 | Pulmonary oedema |

#### Table S25. Ischaemic stroke Aurum codes

| Term | Medcode ID | SNOMED CT Concept ID | SNOMED CT Description ID |
|---|---|---|---|
| [X]Other cerebral infarction | 300941014 | 432504007 | 2770034014 |
| Multiple lacunar infarcts | 5967391000006111 | 307363008 | 450609012 |
| Stroke due to basilar artery thrombus | 3508956015 | 329641000119104 | 3508956015 |
| Cerebrovascular accident due to thrombus of right middle cerebral artery | 3508961018 | 16002031000119102 | 3508961018 |
| Total anterior cerebral circulation stroke | 5011071000006114 | 230694003 | 345642016 |
| [RFC] Stroke/CVA | 907581000006119 | 907581000006103 | 907581000006119 |
| H/O: stroke | 2476091017 | 275526006 | 2476091017 |
| Right sided CVA | 300371014 | 195217004 | 300371014 |
| Brain stem infarct | 4057031000006117 | 95457000 | 158114011 |
| History of stroke in last year | 5974521000006116 | 308067002 | 2986393012 |
| Left sided cerebral hemisphere cerebrovascular accident | 4778181000006119 | 195216008 | 300369014 |
| Stroke due to thrombus of right middle cerebral artery | 3508962013 | 16002031000119102 | 3508962013 |
| Cerebral infarction due to embolism of precerebral arteries | 300313017 | 195186005 | 300313017 |
| PACI - Partial anterior cerebral circulation infarction | 5011101000006116 | 230695002 | 345646018 |
| Cerebellar stroke | 9912271000006112 | 16371781000119100 | 3320493016 |
| Cerebellar infarction | 158118014 | 95460007 | 158118014 |
| TACS - Total anterior cerebral circulation stroke | 5011081000006112 | 230694003 | 345643014 |
| Infarction of brain stem | 4057051000006112 | 95457000 | 2966602014 |
| Total anterior cerebral circulation infarction | 5011051000006116 | 230694003 | 345644015 |
| Cerebrovascular accident | 405339016 | 230690007 | 345637012 |
| History of cerebrovascular accident | 809421000006116 | 275526006 | 2986886017 |
| CVA unspecified | 605501000006117 | 230690007 | 345637012 |
| Posterior cerebral circulation stroke | 5011141000006119 | 230696001 | 2966565019 |
| Thalamic stroke | 7071911000006114 | 427296003 | 2966596011 |
| Partial anterior cerebral circulation stroke | 5011111000006118 | 230695002 | 345647010 |
| Right sided cerebral infarction | 451134017 | 307767006 | 451134017 |
| Left sided cerebral infarction | 451133011 | 307766002 | 451133011 |
| Multi-infarct state | 5011261000006110 | 230704000 | 345662012 |
| Thalamic infarction | 7071901000006111 | 427296003 | 2674127019 |
| Choroidal infarct | 6041261000006115 | 314003000 | 458273015 |
| LI - Lacunar infarction | 5011181000006113 | 230698000 | 345654016 |
| [RFC] Stroke | 907591000006116 | 907591000006100 | 907591000006116 |
| [V]Personal history of stroke | 1227591017 | 275526006 | 2476091017 |
| [X]Cerebrl infarctn due/unspcf occlusn or sten/cerebrl artrs | 370701000006118 | 20059004 | 33759015 |
| Cerebral infarct due to thrombosis of precerebral arteries | 300312010 | 195185009 | 300312010 |
| Cerebral infarction NOS | 12727431000006119 | 682621000000105 | 1495311000000117 |
| Lacunar stroke | 5011161000006115 | 230698000 | 345652017 |

| | | | |
|---|---|---|---|
| Cerebral infarction due to occlusion of precerebral artery | 542261000006114 | 125081000119106 | 3042974014 |
| Cerebral infarction due to thrombosis of cerebral arteries | 300321011 | 195189003 | 300321011 |
| Cerebral ischaemic stroke due to small artery occlusion | 7855631000006111 | 724424009 | 3438036013 |
| Haemorrhagic cerebral infarction | 5011291000006119 | 230706003 | 345667018 |
| Cerebral infarction due to stenosis of cerebral artery | 2729571000000119 | 1089421000000105 | 2729571000000119 |
| Left sided CVA | 300370010 | 195216008 | 300370010 |
| Stroke due to thrombus of left middle cerebral artery | 3508965010 | 16002111000119106 | 3508965010 |
| Cerebral infarction due to embolism of cerebral arteries | 300322016 | 195190007 | 300322016 |
| Posterior cerebral circulation infarction | 5011131000006112 | 230696001 | 345649013 |
| Stroke | 884531000006117 | 685631000000102 | 884531000006117 |
| [RFC] CVA | 909171000006115 | 909171000006104 | 909171000006115 |
| Brainstem infarction NOS | 345650013 | 95457000 | 158113017 |
| Lacunar infarction | 299342019 | 230698000 | 345651012 |
| Sequelae of stroke,not specfd as h'morrhage or infarction | 149571000006118 | 195239002 | 300403014 |
| Infarction of basal ganglia | 2474651019 | 413102000 | 2474651019 |
| Brain stem infarction | 524541000006117 | 95457000 | 158113017 |
| [V]Personal history of cerebrovascular accident (CVA) | 1227592012 | 266995000 | 397829016 |
| Occipital cerebral infarction | 5582311000006110 | 276219001 | 412270012 |
| Anterior circulation stroke of uncertain pathology | 5011461000006111 | 230714009 | 345681016 |
| Occlusive stroke | 1212072018 | 373606000 | 1212072018 |
| Stroke of uncertain pathology | 5011451000006114 | 230713003 | 345680015 |
| Cerebral infarction | 395780010 | 432504007 | 2770034014 |
| PACS - Partial anterior cerebral circulation stroke | 5011121000006114 | 230695002 | 345648017 |
| Cerebrovascular accident due to cerebral artery occlusion | 5011021000006113 | 230691006 | 2914970017 |
| Cerebral infarction with haemorrhagic transformation | 1573101000006112 | 1573101000006108 | 1573101000006112 |
| [X]Sequelae of stroke,not specfd as h'morrhage or infarction | 426321000006116 | 195239002 | 300403014 |
| CVA - Cerebrovascular accident | 605491000006113 | 230690007 | 345635016 |
| Stroke/CVA - undefined | 884521000006115 | 685631000000102 | 884521000006115 |
| Anterior cerebral circulation haemorrhagic infarction | 5011331000006114 | 230707007 | 345668011 |
| H/O: CVA | 411518010 | 275526006 | 411518010 |
| Cerebral infarction due to thrombosis of middle cerebral artery | 7599001000006116 | 705130002 | 3023374015 |
| Ischemic stroke | 6990991000006112 | 422504002 | 2644233012 |
| Stroke unspecified | 122401000006115 | 230690007 | 345637012 |
| Partial anterior cerebral circulation infarction | 5011091000006110 | 230695002 | 345645019 |
| Basal ganglion stroke | 6837051000006119 | 413102000 | 2966612019 |
| LACI - Lacunar infarction | 5011171000006110 | 230698000 | 345653010 |
| Brainstem stroke syndrome | 524511000006116 | 195212005 | 300365015 |
| Right sided cerebral hemisphere cerebrovascular accident | 4778201000006118 | 195217004 | 300372019 |

| | | | |
|---|---|---|---|
| Infarction - cerebral | 218511000000117 | 432504007 | 2770034014 |
| Stroke and TIA | 8221321000006118 | 720191000000104 | 1574361000000119 |
| Sequela of ischaemic cerebral infarction | 8011371000006117 | 23671000119107 | 2967516015 |
| Cerebral infarction due to occlusion of cerebral artery | 2729551000000111 | 1089411000000104 | 2729551000000111 |
| Cerebral infarction due to embolism of middle cerebral artery | 7598981000006119 | 705128004 | 3023367013 |
| Infarction - precerebral | 345639010 | 230692004 | 345639010 |
| Stroke and cerebrovascular accident unspecified | 12727691000006117 | 685631000000102 | 1501361000000113 |
| H/O: Stroke in last year | 451371010 | 308067002 | 451371010 |
| Cerebrovascular accident due to occlusion of cerebral artery | 11878511000006113 | 230691006 | 3636108019 |
| CVA - cerebrovascular accident due to cerebral artery occlusion | 605461000006117 | 230691006 | 345638019 |
| [X]Cereb infarct due unsp occlus/stenos precerebr arteries | 370661000006114 | 125081000119106 | 3042974014 |
| Stroke due to cerebral arterial occlusion | 122361000006113 | 230691006 | 345638019 |
| Stroke | 5010981000006119 | 230690007 | 345636015 |
| Left hemispheric cerebellar artery embolism with stroke | 3509817019 | 329461000119102 | 3509817019 |
| Posterior circulation stroke of uncertain pathology | 5011471000006116 | 230715005 | 345682011 |
| Ischaemic stroke | 6990981000006114 | 422504002 | 2644234018 |
| Anterior cerebral circulation infarction | 5011041000006118 | 230693009 | 345640012 |
| TACI - Total anterior cerebral circulation infarction | 5011061000006119 | 230694003 | 345641011 |
| Sequelae of cerebral infarction | 300411016 | 195243003 | 300411016 |
| Embolic stroke | 6348301000006119 | 371041009 | 1209751018 |
| Brain stem stroke | 4057041000006110 | 95457000 | 2966556014 |
| Cause of Death- Cerebral Infarct | 1576261000006112 | 1576261000006108 | 1576261000006112 |
| [RFC] Stroke | 908801000006114 | 908801000006105 | 908801000006114 |
| Cerebrl infarctn due/unspcf occlusn or sten/cerebrl artrs | 543141000006110 | 20059004 | 33759015 |
| Sequela of ischemic cerebral infarction | 8011381000006119 | 23671000119107 | 2967553014 |

#### Table S26. Ischaemic stroke HES (ICD10) codes

| ICD10 codes | Term |
|---|---|
| I63.0 | Cerebral infarction due to thrombosis of precerebral arteries |
| I63.1 | Cerebral infarction due to embolism of precerebral arteries |
| I63.2 | Cerebral infarction due to unspecified occlusion or stenosis of precerebral arteries |
| I63.3 | Cerebral infarction due to thrombosis of cerebral arteries |
| I63.4 | Cerebral infarction due to embolism of cerebral arteries |
| I63.5 | Cerebral infarction due to unspecified occlusion or stenosis of cerebral arteries |
| I63.6 | Cerebral infarction due to cerebral venous thrombosis, nonpyogenic |
| I63.8 | Other cerebral infarction |
| I63.9 | Cerebral infarction, unspecified |
| I64 | Stroke, not specified as haemorrhage or infarction |

#### Table S27. Transient ischaemic attack outcome Aurum codes

| Term | Medcode ID | SNOMED CT Concept ID | SNOMED CT Description ID |
|---|---|---|---|
| Intermittent cerebral ischaemia | 300353019 | 195206000 | 300353019 |
| Anterior cerebral artery syndrome | 300363010 | 195210002 | 300363010 |
| Anterior circulation transient ischaemic attack | 5011491000006115 | 230716006 | 345683018 |
| Transient cerebral ischemia | 11920121000006117 | 266257000 | 395785017 |
| Personal history of transient ischaemic attack | 8231151000006110 | 751371000000107 | 1653121000000119 |
| Generalised ischaemic cerebrovascular disease NOS | 300379011 | 302909007 | 444905019 |
| Insufficiency - basilar artery | 499739014 | 64009001 | 499739014 |
| [V]Personal history of transient ischaemic attack | 1667741000000110 | 751371000000107 | 1667741000000110 |
| Vertebral artery syndrome | 58046010 | 34781003 | 58046010 |
| Chronic cerebral ischaemia | 360778011 | 111298007 | 360778011 |
| Basilar artery syndrome | 106392017 | 64009001 | 106392017 |
| Vertebrobasilar insufficiency | 106394016 | 64009001 | 106394016 |
| Transient ischaemic attack | 395783012 | 266257000 | 395783012 |
| Transient cerebral ischaemia | 395788015 | 266257000 | 395788015 |
| Other transient cerebral ischaemia | 300348012 | 266257000 | 395788015 |
| [X]Other transnt cerebral ischaemic attacks+related syndroms | 416991000006112 | 266257000 | 395788015 |
| Carotid territory transient ischaemic attack | 345684012 | 230716006 | 345684012 |
| Vertebrobasilar territory transient ischaemic attack | 5011521000006118 | 230717002 | 345689019 |
| Posterior cerebral artery syndrome | 300364016 | 195211003 | 300364016 |
| Occlusion and stenosis of cerebral arteries, not resulting in cerebral infarction | 267321000006114 | 195231004 | 300394010 |
| History of transient ischemic attack | 4540591000006113 | 161511000 | 2986735018 |
| Impending cerebral ischaemia | 300352012 | 195205001 | 300352012 |
| AF - Amaurosis fugax | 3930471000006118 | 88032003 | 507856014 |
| Transient cerebral ischaemia NOS | 12733221000006110 | 584181000000100 | 1296321000000119 |
| Transient Ischaemic Attacks | 884511000006111 | 584181000000100 | 884511000006111 |
| Transient cerebral ischaemia NOS | 95931000006111 | 266257000 | 395788015 |
| AFx - Amaurosis fugax | 3930481000006115 | 88032003 | 507857017 |
| Transient cerebral ischaemia NOS | 300349016 | 266257000 | 395788015 |
| Middle cerebral artery syndrome | 300362017 | 195209007 | 300362017 |
| Amaurosis fugax | 145925010 | 88032003 | 145925010 |
| Posterior circulation transient ischemic attack | 5011541000006113 | 230717002 | 345688010 |
| Amaurosis fugax of right eye | 9478331000006110 | 12237951000119105 | 3326553010 |
| Vertebro-basilar insufficiency | 67511000006117 | 64009001 | 106394016 |
| Carotid territory transient ischemic attack | 5011511000006114 | 230716006 | 345686014 |
| Anterior circulation transient ischemic attack | 5011501000006111 | 230716006 | 345685013 |
| Vertebrobasilar arterial insufficiency | 3540781000006117 | 64009001 | 106393010 |
| Suspected transient ischaemic attack | 1704681000000110 | 473129008 | 2955917017 |
| Amaurosis fugax (one sided temporary vision loss) | 3930501000006113 | 88032003 | 2966609017 |
| Suspected transient ischemic attack | 7483201000006116 | 473129008 | 2955838012 |

| | | | |
|---|---|---|---|
| Amaurosis | 3645721000006118 | 70449006 | 117033019 |
| H/O: TIA | 251692018 | 161511000 | 251692018 |
| Vertebro-basilar artery syndrome | 67501000006115 | 195199008 | 300343015 |
| Transient Ischaemic Attacks | 988951000006117 | 266257000 | 988951000006117 |
| TIA - Transient ischaemic attack | 5492201000006111 | 266257000 | 395784018 |

#### Table S28. Transient ischaemic attack HES (ICD10) codes

| ICD10 codes | Term |
|---|---|
| G45.2 | Multiple and bilateral precerebral artery syndromes |
| G45.3 | Amaurosis fugax |
| G45.8 | Other transient cerebral ischaemic attacks and related syndromes |
| G45.9 | Transient cerebral ischaemic attack, unspecified |
| G46.0 | Middle cerebral artery syndrome |
| G46.1 | Anterior cerebral artery syndrome |
| G46.2 | Posterior cerebral artery syndrome |
| G46.3 | Brain stem stroke syndrome |
| G46.4 | Cerebellar stroke syndrome |
| G46.5 | Pure motor lacunar syndrome |
| G46.6 | Pure sensory lacunar syndrome |
| G46.7 | Other lacunar syndromes |
| G46.8 | Other vascular syndromes of brain in cerebrovascular diseases |
| I65.0 | Occlusion and stenosis of vertebral artery |
| I65.1 | Occlusion and stenosis of basilar artery |
| I65.2 | Occlusion and stenosis of carotid artery |
| I65.3 | Occlusion and stenosis of multiple and bilateral precerebral arteries |
| I65.8 | Occlusion and stenosis of other precerebral artery |
| I65.9 | Occlusion and stenosis of unspecified precerebral artery |
| I66.0 | Occlusion and stenosis of middle cerebral artery |
| I66.1 | Occlusion and stenosis of anterior cerebral artery |
| I66.2 | Occlusion and stenosis of posterior cerebral artery |
| I66.3 | Occlusion and stenosis of cerebellar arteries |
| I66.4 | Occlusion and stenosis of multiple and bilateral cerebral arteries |
| I66.8 | Occlusion and stenosis of other cerebral artery |
| I66.9 | Occlusion and stenosis of unspecified cerebral artery |
| I67.2 | Cerebral atherosclerosis |
| I67.9 | Cerebrovascular disease, unspecified |

#### Table S29. Venous thromoemoblism and pulmonary embolism Aurum codes

| Term | Medcode ID | SNOMED CT Concept ID | SNOMED CT Description ID |
|---|---|---|---|
| Embolism and/or thrombosis of the internal iliac artery | 300554012 | 734298005 | 3503835018 |
| CRVT - Central retinal vein thrombosis | 3614311000006116 | 68478007 | 501026011 |
| H/O: Deep vein thrombosis | 4540531000006114 | 161508001 | 4540531000006114 |
| Thrombosis transverse sinus | 296939018 | 192761004 | 296939018 |
| Coronary thrombosis not resulting in myocardial infarction | 299742017 | 194821006 | 299742017 |
| Embolism and thrombosis of the brachial artery | 300535012 | 195319003 | 300535012 |
| Embolism and thrombosis of the posterior tibial artery | 300543019 | 195327007 | 300543019 |
| Mesenteric embolus | 412653014 | 276500007 | 412653014 |
| Ischaemic foot | 443199013 | 301755001 | 443199013 |
| Open removal of cardiac thrombus | 2675367019 | 426067000 | 2675367019 |
| Percutaneous transluminal thrombolysis of artery | 2675878019 | 426485003 | 2675878019 |
| Open thrombectomy of renal vein | 2675885015 | 426937008 | 2675885015 |
| Ischaemic disease of gut | 3836741000006113 | 82196007 | 504940010 |
| Acute intestinal ischaemic syndrome | 3986491000006113 | 91489000 | 509476019 |
| Recurrent pulmonary embolism | 632111000000118 | 438773007 | 2793990014 |
| Intestinal ischaemia | 3836721000006118 | 82196007 | 504938017 |
| Embolism and thrombosis of the celiac artery | 4778981000006116 | 195342005 | 300560012 |
| Ischaemia reperfusion injury of liver | 7821701000006113 | 721715000 | 3325835015 |
| AMI - Acute mesenteric ischaemia | 3986481000006110 | 91489000 | 509475015 |
| Deep venous thrombosis of lower extremity | 6694741000006111 | 404223003 | 2156120013 |
| Inner retinal ischaemic spots | 3140891000006117 | 39832008 | 2620723015 |
| Thrombosis of renal artery bypass graft | 449993015 | 306857000 | 449993015 |
| Ischaemic toe | 450665015 | 307408003 | 450665015 |
| Mesenteric thrombus and/or embolus | 450682018 | 307420006 | 450682018 |
| Thrombosed haemorrhoid incised | 5727361000006116 | 287799004 | 2871742013 |
| Infarct of liver | 2783881000006119 | 17890003 | 30227010 |
| TL - Transluminal pulmonary embolectomy | 4666351000006113 | 175268008 | 271403011 |
| Ischaemic bowel disease | 3836731000006115 | 82196007 | 504939013 |
| Thrombosis of central nervous system venous sinus NOS | 296940016 | 192759008 | 296937016 |
| Sagittal sinus thrombosis | 3648391000006110 | 70607008 | 501619010 |
| History of recurrent deep vein thrombosis | 8044851000006112 | 141911000119100 | 3043613018 |
| Thrombosed external haemorrhoid | 2922231000006111 | 26373009 | 483208017 |
| Venous thromboembolic disease | 7098321000006117 | 429098002 | 2695294011 |
| Deep venous thrombosis - leg | 989221000006110 | 404223003 | 989221000006110 |
| Cerebral venous thrombosis of lateral sinus | 2838001000006118 | 21258007 | 1222620011 |
| Ischaemic optic neuropathy | 476238012 | 14357004 | 476238012 |
| Pulmonary embolus | 193601000006115 | 59282003 | 98484016 |
| Mesenteric thrombosis | 215511000000110 | 95446005 | 158099015 |

| | | | |
|---|---|---|---|
| Post radiological embolism of upper limb artery | 216711000006118 | 312377009 | 456079019 |
| Thrombosis of cavernous venous sinus | 3962581000006110 | 89980009 | 149150017 |
| Unprovoked deep vein thrombosis | 2488341000000112 | 978421000000101 | 2488341000000112 |
| Ischaemic arm | 5058031000006112 | 233959009 | 350538016 |
| Thrombosis of transverse sinus | 2837991000006111 | 21258007 | 35719015 |
| Pulmonary arterial thrombosis | 3439711000006111 | 57834008 | 96166019 |
| Male genital thrombosis NOS | 304536019 | 198057005 | 304524013 |
| Ischaemic necrosis | 3901731000006110 | 86217007 | 142974016 |
| Vascular graft embolus | 5925451000006112 | 304090004 | 446358019 |
| Deep venous thrombosis of leg | 6694761000006110 | 404223003 | 2162420015 |
| Superior mesenteric artery embolus | 4785631000006110 | 196999001 | 302978019 |
| Embolus of central nervous system venous sinus | 639231000006117 | 14246007 | 1221072019 |
| Acute ischaemic colitis | 503091015 | 75700000 | 503091015 |
| Thyroid infarction | 1220407012 | 11859008 | 1220407012 |
| Embolism of central nervous system venous sinus | 1221072019 | 14246007 | 1221072019 |
| Capillary thrombosis | 30091013 | 17810004 | 30091013 |
| Acute infarction of papillary muscle | 2663441000006117 | 10273003 | 17900011 |
| Removal of vessel clot | 866201000006119 | 392031002 | 866201000006119 |
| Acute deep venous thrombosis of internal jugular vein | 8042511000006119 | 135001000119100 | 2971631010 |
| PVT - Portal vein thrombosis | 2784351000006119 | 17920008 | 1221613018 |
| Arteritic ischaemic optic neuropathy | 5008941000006114 | 230508004 | 345412011 |
| Peripheral ischaemic vascular disease | 350535018 | 233958001 | 350535018 |
| Upper limb ischaemia | 350540014 | 233959009 | 350540014 |
| Trendelenburg pulmonary embolectomy | 271392011 | 175262009 | 271392011 |
| Open embolectomy of bifurcation of aorta | 271509013 | 175348000 | 271509013 |
| Open embolectomy of carotid artery | 271540018 | 175374006 | 271540018 |
| Open embolectomy of axillary artery | 271639012 | 175446000 | 271639012 |
| Percutaneous transluminal embolectomy of subclavian artery | 271654018 | 175456001 | 271654018 |
| Percutaneous transluminal embolectomy of brachial artery | 271656016 | 175457005 | 271656016 |
| Percutaneous transluminal embolectomy of axillary artery | 271671017 | 175465008 | 271671017 |
| Percutaneous transluminal embolectomy of iliac artery | 271901019 | 175613000 | 271901019 |
| Open thrombectomy of femoral artery | 272032018 | 175704008 | 272032018 |
| Percutaneous transluminal embolectomy of femoral artery | 272055015 | 175717006 | 272055015 |
| Percutaneous transluminal embolectomy of popliteal artery | 272056019 | 175718001 | 272056019 |
| Prosthetic graft thrombectomy | 272305017 | 175891000 | 272305017 |
| Corpus cavernosum embolism | 1231107016 | 52303006 | 1231107016 |
| Vein graft thrombectomy | 1233341011 | 71007004 | 1233341011 |
| Cerebral vein thrombosis | 1235913017 | 95455008 | 1235913017 |
| Infarction of fallopian tube | 94629010 | 56900004 | 94629010 |
| Infarct of lung | 3551761000006115 | 64662007 | 107460013 |

| | | | |
|---|---|---|---|
| Cerebral arterial thrombosis | 3662321000006117 | 71444005 | 118691019 |
| Ischaemic foot | 1699111000006110 | 1699111000006106 | 1699111000006110 |
| Percutaneous mechanical thromboembolectomy | 359731000000112 | 225291000000108 | 359731000000112 |
| Unprovoked DVT (deep vein thrombosis) | 8361011000006114 | 978421000000101 | 2518391000000114 |
| Carotid thromboendarterectomy | 3588601000006110 | 66951008 | 111209010 |
| Saddle embolus of abdominal aorta | 5058281000006118 | 233972005 | 350565011 |
| Peripheral arterial embolism | 884661000006110 | 583731000000103 | 884661000006110 |
| Embolus/thrombus artery NOS | 884671000006115 | 583761000000108 | 884671000006115 |
| Venous embolism NOS | 884741000006117 | 587851000000108 | 884741000006117 |
| Embolism | 887751000006110 | 198850008 | 887751000006110 |
| Evacuation of thrombosed haemorrhoids | 3984761000006115 | 91392002 | 509429011 |
| Embolus of vein NOS | 12719591000006119 | 587851000000108 | 1303841000000119 |
| Arterial embolism and thrombosis NOS | 12733161000006111 | 583761000000108 | 1295451000000114 |
| Cholesterol embolus syndrome | 354495010 | 236489002 | 354495010 |
| Fogarty embolectomy of artery | 1776773010 | 397047005 | 1776773010 |
| Thrombosis - coronary | 1786198013 | 398274000 | 1786198013 |
| Central retinal vein occlusion | 113737013 | 68478007 | 113737013 |
| Thrombosis of cerebral arteries | 3662311000006113 | 71444005 | 118690018 |
| BRVT - Branch retinal vein thrombosis | 2892681000006110 | 24596005 | 481963013 |
| Thrombosed external pile | 2922241000006118 | 26373009 | 483209013 |
| Thromboendarterectomy of aorta | 3295821000006118 | 49187005 | 81937019 |
| PC TL embolectomy popliteal artery | 4668191000006117 | 175718001 | 272057011 |
| Percutaneous transluminal thrombolysis of vein with reconstruction | 7098901000006114 | 429141007 | 2692268010 |
| Right ventricular thrombus | 5989831000006113 | 309518001 | 452962013 |
| Thrombosis of testis | 3605531000006118 | 67962003 | 112890010 |
| Cerebral arterial embolism | 3728451000006116 | 75543006 | 125471016 |
| IRIS - Inner retinal ischaemic spots | 3140901000006118 | 39832008 | 2620724014 |
| Deep vein thrombosis of peroneal vein | 1119161000000115 | 443210003 | 1664901000000117 |
| Renal infarction | 1848131000006113 | 45456005 | 75778012 |
| [RFC] Venous thrombosis | 905491000006112 | 905491000006108 | 905491000006112 |
| Ischaemic nephropathy | 1160091000000115 | 710565001 | 3082872011 |
| Release of ischaemic contracture of forearm | 4682421000006112 | 178253006 | 275890019 |
| Open embolectomy of inferior mesenteric artery NEC | 271760012 | 397045002 | 1776771012 |
| Open embolectomy of suprarenal artery NEC | 271761011 | 397045002 | 1776771012 |
| Open embolectomy of visceral branch of abdominal aorta NEC | 271780013 | 397045002 | 1776771012 |
| Open embolectomy of artery NEC | 272097014 | 397045002 | 1776771012 |
| Open removal of thrombus from vein NOS | 272248014 | 175843001 | 272243017 |
| Open embolectomy of iliac artery | 394332017 | 265521000 | 394332017 |
| Renal artery thrombosis | 158317012 | 95579008 | 158317012 |
| Deep vein thrombosis of leg related to air travel | 216205019 | 134399007 | 216205019 |
| Thrombosis of lateral venous sinus | 2837981000006113 | 21258007 | 35718011 |

| | | | |
|---|---|---|---|
| Percutaneous thrombolysis of vein | 5593341000006115 | 276985008 | 413389019 |
| Brachial embolectomy | 5048991000006113 | 233313002 | 349588014 |
| Percutaneous transluminal embolectomy of artery NEC | 394347014 | 397045002 | 1776771012 |
| Cerebral venous thrombosis | 1729331000006116 | 1729331000006100 | 1729331000006116 |
| Embolism of corpus cavernosum | 3347911000006112 | 52303006 | 87035015 |
| Chronic intestinal ischaemia | 4194521000006117 | 111354009 | 361442014 |
| Deep vein thrombosis of portal vein | 2784341000006116 | 17920008 | 1221612011 |
| Embolism and thrombosis of other specified artery | 300552011 | 266262004 | 395794011 |
| Arterial embolism and thrombosis NOS | 300564015 | 266262004 | 395794011 |
| Infarction of thyroid | 2688811000006110 | 11859008 | 20465014 |
| Thrombosis of intracranial venous sinus | 4768491000006118 | 192759008 | 1784812010 |
| Thrombosis of penile vein | 3745871000006116 | 76598006 | 127203016 |
| Thrombosed hemorrhoid incised | 5727411000006112 | 287799004 | 2871743015 |
| Occlusive thromboarteriopathy | 6206441000006110 | 359789008 | 475141015 |
| Percutaneous transluminal thrombolysis of blood vessel of liver | 1545591000006112 | 426059008 | 2675452017 |
| Percutaneous transluminal thrombolysis and reconstruction of artery | 1548401000006112 | 428068004 | 2696045014 |
| Aortic bifurcation embolus | 5058291000006115 | 233972005 | 350566012 |
| External thrombosed hemorrhoids | 2922201000006115 | 26373009 | 483203014 |
| Acute colonic ischaemia | 3730871000006110 | 75700000 | 2310141000000117 |
| Thrombosis of cerebral veins | 4056971000006118 | 95455008 | 158110019 |
| Embolectomy | 3668011000006113 | 71815002 | 119306015 |
| Embolism and thrombosis of a leg artery NOS | 300547018 | 195318006 | 300534011 |
| Peripheral arterial embolism and thrombosis NOS | 300550015 | 195318006 | 300534011 |
| Renal artery embolism | 158318019 | 95580006 | 158318019 |
| Embolectomy of popliteal artery | 5049001000006113 | 233314008 | 349592019 |
| Multiple infarcts | 2947401000006115 | 27876002 | 46655014 |
| DVT - Deep vein thrombosis of lower limb | 6694751000006113 | 404223003 | 2162419014 |
| Infarction of kidney | 3233381000006117 | 45456005 | 75780018 |
| Acute deep vein thrombosis of lower limb | 7966351000006118 | 651000119108 | 2967724010 |
| Superficial venous thrombosis of leg | 7103191000006117 | 429434005 | 2692641015 |
| Ocular ischaemic syndrome | 6045981000006117 | 314436005 | 458797013 |
| Mesenteric embolism | 412652016 | 276500007 | 412652016 |
| Thrombosis of inferior vena cava | 419640012 | 281595001 | 419640012 |
| Thrombosis of vein of leg | 453210011 | 309735004 | 453210011 |
| Vertebrobasilar ischaemic vertigo | 5034291000006111 | 232288005 | 348027016 |
| Infarction | 3403531000006115 | 55641003 | 92491010 |
| Embolism in vascular graft | 446359010 | 304090004 | 446359010 |
| Thromboembolus of internal iliac artery | 3503835018 | 734298005 | 3503835018 |
| SMAE - Superior mesenteric artery embolus | 4785641000006117 | 196999001 | 302979010 |
| Thrombosis of stent of renal artery | 1848121000006110 | 840961000000108 | 2182101000000113 |
| Deep venous thrombosis of the popliteal vein | 7078601000006114 | 427776007 | 2691844011 |

| | | | |
|---|---|---|---|
| Thrombosis of vein of lower leg | 5059271000006110 | 234043001 | 2820167012 |
| Saddle embolus of pulmonary artery | 8106001000006111 | 328511000119109 | 3322848011 |
| Popliteal artery thrombosis | 3918361000006110 | 87239004 | 144656012 |
| Saddle embolus | 3108321000006117 | 37778000 | 63025019 |
| Arterial embolism | 3387371000006112 | 54687002 | 90875013 |
| Brachiocephalic vein thrombosis | 5653441000006111 | 281597009 | 419642016 |
| Stroke due to basilar artery thrombus | 3508956015 | 329641000119104 | 3508956015 |
| Femoral embolectomy | 2634331000006113 | 8409001 | 1234899012 |
| Hemopericardium due to and following acute myocardial infarction | 12220751000006113 | 194862000 | 3673281015 |
| Penile venous thrombosis | 3745891000006115 | 76598006 | 3036947013 |
| Thrombosed external haemorrhoids | 2922181000006116 | 26373009 | 196520016 |
| Cerebral infarction due to occlusion of precerebral artery | 542261000006114 | 125081000119106 | 3042974014 |
| Deep vein thrombosis | 2162148012 | 128053003 | 2162149016 |
| Carotid artery thrombosis | 100771000006112 | 86003009 | 142588012 |
| Occlusive mesenteric ischaemia | 5086201000006115 | 235842000 | 353530010 |
| Multiple lacunar infarcts | 5967391000006111 | 307363008 | 450609012 |
| Venous thrombosis, phlebitis and thrombophlebitis | 6018501000006115 | 312585004 | 456332011 |
| Thyroid hemorrhage and infarction | 4757961000006112 | 190305000 | 292442019 |
| Non-occlusive thrombus | 2927841000006117 | 26713004 | 44719012 |
| Open embolectomy of coeliac artery NEC | 271757017 | 397045002 | 1776771012 |
| Multifocal infarct | 3474711000006110 | 60007006 | 99691019 |
| Infarction of spleen | 38632019 | 22996003 | 1223010014 |
| Pulmonary thromboembolism | 5057591000006116 | 233935004 | 350497014 |
| Thrombectomy | 3205401000006110 | 43810009 | 73061013 |
| Thrombosis of superior sagittal sinus | 3648381000006112 | 70607008 | 117287016 |
| Ischaemic hepatitis | 5086761000006113 | 235877000 | 353591011 |
| Ischaemic gangrene | 6670921000006117 | 402861007 | 1773672019 |
| Suspected pulmonary embolism | 2549664019 | 417113001 | 2549664019 |
| Pulmonary thromboendarterectomy | 2675439010 | 426900002 | 2675439010 |
| Intracranial venous thrombosis | 5836951000006117 | 297157005 | 437623015 |
| Thrombosis of atrium, auricular appendage, and ventricle due to and following acute myocardial infarction | 12220791000006119 | 194868001 | 3673277015 |
| Recurrent DVT (deep vein thrombosis) | 7665071000006116 | 710167004 | 2496081000000115 |
| Lacunar infarction | 299342019 | 230698000 | 345651012 |
| Venous embolism | 300975010 | 234049002 | 350676017 |
| Embolectomy of iliac artery | 5153821000006115 | 240928009 | 360865017 |
| Embolectomy of femoral artery | 2634321000006110 | 8409001 | 14892019 |
| Ischaemic ulcer of toe | 7108391000006114 | 429768000 | 2693575017 |
| Acute deep venous thrombosis of femoral vein | 8041581000006116 | 132291000119106 | 2971883017 |
| Removal of thrombus of arteriovenous fistula | 7081801000006114 | 427992007 | 2691808018 |
| Ischemic infarction of muscle | 5143791000006114 | 240126009 | 359749018 |

| | | | |
|---|---|---|---|
| Thyroid haemorrhage or infarction NOS | 292445017 | 190305000 | 292443012 |
| Thromboangiitis obliterans NOS | 300503012 | 52403007 | 87208017 |
| Embolism and thrombosis of an arm artery NOS | 300538014 | 195318006 | 300534011 |
| Thrombosis | 7218941000006118 | 439127006 | 2794470015 |
| Clotted venous saccule | 5086421000006112 | 235854000 | 353554017 |
| Venous thrombosis | 395803017 | 111293003 | 178522012 |
| Thrombosis | 5477661000006119 | 264579008 | 393059012 |
| Saddle embolus | 350563016 | 233972005 | 350563016 |
| Infarction of liver | 2783891000006116 | 17890003 | 1221609013 |
| Provoked DVT (deep vein thrombosis) | 8361031000006115 | 978441000000108 | 2518461000000111 |
| Femoral artery embolus | 5058301000006119 | 233973000 | 350567015 |
| Critical lower limb ischaemia | 5058111000006119 | 233962007 | 350547012 |
| Subclavian artery thrombosis | 5836921000006114 | 297154003 | 437620017 |
| Lacunar infarct | 3818181000006114 | 81037000 | 134436018 |
| Ischaemia of feet | 5886711000006111 | 300917007 | 442114010 |
| Embolism and/or thrombosis of the external iliac artery | 300555013 | 734299002 | 3503839012 |
| Renal infarct | 3233391000006119 | 45456005 | 75781019 |
| Embolectomy of carotid artery | 5902211000006114 | 302053004 | 443591015 |
| Embolism of vein NOS | 12759341000006113 | 587851000000108 | 1303861000000118 |
| Lower limb arterial embolus | 6015571000006117 | 312378004 | 456080016 |
| Embolectomy of lower limb artery | 3688071000006119 | 73038007 | 121315013 |
| Iliofemoral deep vein thrombosis | 5059301000006112 | 234044007 | 3464381018 |
| Thrombolysis of vein | 5050621000006118 | 233428003 | 349770015 |
| Thrombolysis of artery | 5049421000006111 | 233335005 | 349633016 |
| Thyroid haemorrhage and infarction | 292443012 | 190305000 | 292443012 |
| Embolism cavernous sinus | 296932010 | 192754003 | 296932010 |
| Embolism lateral sinus | 296934011 | 192756001 | 296934011 |
| Embolism and thrombosis of an arm or leg artery | 300534011 | 195318006 | 300534011 |
| Embolism and/or thrombosis of the common iliac artery | 300553018 | 195335005 | 300553018 |
| Embolism and thrombosis of the splenic artery | 300558010 | 195340002 | 300558010 |
| Thrombophlebitis of the femoral vein | 300672010 | 195410000 | 300672010 |
| Thrombophlebitis of the anterior tibial vein | 300674011 | 195412008 | 300674011 |
| History of thromboembolism | 5497011000006116 | 266996004 | 2986452015 |
| Acute intestinal ischaemia | 3986461000006117 | 91489000 | 509473010 |
| History of deep vein thrombosis | 4540551000006119 | 161508001 | 2986938011 |
| Incision of thrombosed hemorrhoid | 3984721000006114 | 91392002 | 151416013 |
| Post-radiological embolism of upper limb artery | 6015561000006112 | 312377009 | 456079019 |
| Colonic ischaemia | 2992791000006113 | 30588004 | 1216341014 |
| Cerebral venous thrombosis of cavernous sinus | 100691000006118 | 89980009 | 1235519010 |
| Cerebral venous sinus thrombosis | 100721000006111 | 192759008 | 296937016 |
| Superior mesenteric vein thrombosis | 100821000006116 | 197001004 | 302981012 |

| | | | |
|---|---|---|---|
| Postoperative deep vein thrombosis | 216571000006116 | 213220000 | 325066018 |
| Postoperative pulmonary embolus | 216591000006115 | 194883006 | 299847015 |
| Ischaemic priapism | 7252011000006114 | 441575009 | 2817057019 |
| Central retinal vein thrombosis | 3614331000006110 | 68478007 | 501028012 |
| Digital arterial thrombosis | 5886741000006110 | 300919005 | 442117015 |
| Cerebral infarction due to cerebral venous thrombosis, non-pyogenic | 542251000006112 | 195230003 | 300393016 |
| Percutaneous transluminal venous thrombolysis NEC | 358041000000114 | 276985008 | 413389019 |
| Percutaneous removal of thrombus from vein NOS | 359571000000118 | 276977004 | 413381016 |
| Other specified percutaneous removal of thrombus from vein | 359771000000114 | 276977004 | 413381016 |
| Evacuation of thrombosed hemorrhoid | 3984741000006119 | 91392002 | 509427013 |
| Lower limb ischaemia | 5058071000006110 | 233961000 | 350544017 |
| Arterial embolus | 3387391000006113 | 54687002 | 497013010 |
| Embolism | 3402611000006115 | 55584005 | 92392016 |
| Chronic deep vein thrombosis of left iliac vein | 12107671000006115 | 293461000119100 | 3646306012 |
| Splenic vein thrombosis | 2730141000006117 | 14534009 | 24693011 |
| Percutaneous transluminal embolectomy of bifurcation of aorta | 232881000006113 | 175356002 | 271519019 |
| Branch retinal vein thrombosis | 2892651000006119 | 24596005 | 481960011 |
| Embolus/thrombosis abd. aorta | 884641000006111 | 266263009 | 884641000006111 |
| Embolus/thrombosis aorta NOS | 884651000006113 | 195317001 | 884651000006113 |
| Removal of thrombus from intracranial artery | 2845481000006110 | 21710002 | 36433012 |
| History of thromboembolism of vein | 7095381000006112 | 428904003 | 2695935017 |
| Percutaneous embolectomy of brachial artery | 4667301000006111 | 175457005 | 271655017 |
| Transluminal pulmonary embolectomy | 4666341000006111 | 175268008 | 271402018 |
| Acute mesenteric ischaemia | 457631000006114 | 91489000 | 509474016 |
| Mesenteric infarction | 523601000006115 | 3558002 | 7001012 |
| Deep venous thrombosis of deep femoral vein | 7078581000006116 | 427775006 | 2695171017 |
| Embolism and thrombosis of other and unspec parts aorta | 638871000006114 | 274101000 | 409861010 |
| Embolus of the superior mesenteric artery | 639241000006110 | 196999001 | 302978019 |
| Thrombosed external hemorrhoid | 2922211000006117 | 26373009 | 483205019 |
| Pulmonary infarction | 728841000006111 | 64662007 | 107459015 |
| Thrombosed internal haemorrhoids | 770161000006115 | 52931009 | 198419016 |
| Percutaneous transluminal thrombectomy of blood vessel of liver | 1545581000006114 | 428953003 | 2692834012 |
| Percutaneous transluminal venous thrombolysis with reconstruction | 1549761000006110 | 429141007 | 1659501000000119 |
| Retinal ischaemia | 483237018 | 26468004 | 483237018 |
| Ischaemic | 5360051000006112 | 255426005 | 380631011 |
| Asymptomatic ischaemia | 5056261000006112 | 233823002 | 350356015 |
| History of pulmonary embolus | 4540611000006119 | 161512007 | 2986421016 |
| Central retinal vein occlusion - ischaemic | 6024101000006114 | 312997008 | 456819013 |
| Embolism of iliac artery | 3807321000006113 | 80383008 | 133386017 |
| Fogarty embolectomy of vein | 1776763013 | 397035004 | 1776763013 |

| | | | |
|---|---|---|---|
| Arterial embolus and thrombosis | 395795012 | 266262004 | 395795012 |
| Occlusive thrombus | 3771791000006113 | 78195007 | 129763011 |
| Pulmonary embolism with pulmonary infarction | 7967281000006119 | 1001000119102 | 2967075019 |
| Non-pyogenic venous sinus thrombosis | 586241000006116 | 195229008 | 300392014 |
| Arterial ischaemia | 5057951000006115 | 233957006 | 350532015 |
| Intracranial thrombectomy | 2845491000006113 | 21710002 | 36434018 |
| Deep vein thrombosis, leg | 218551000000118 | 266267005 | 395801015 |
| Deep venous thrombosis of the superior mesenteric vein | 4785671000006113 | 197001004 | 302982017 |
| Acute ischaemia of small intestine | 8321371000006112 | 906191000000102 | 2329961000000119 |
| Rupture of chordae tendinae due to and following acute myocardial infarction | 12220781000006117 | 194866002 | 3673278013 |
| Central retinal vein occlusion - non-ischaemic | 6024121000006116 | 312998003 | 456822010 |
| Ischaemic finger | 5967971000006111 | 307409006 | 450668018 |
| Ischaemic ulcer | 2721231000006112 | 13954005 | 475615013 |
| Mural thrombosis | 1235655012 | 91335003 | 1235655012 |
| Ischaemic leg ulcer | 357895013 | 238793001 | 357895013 |
| Ischaemic foot ulcer | 5126451000006116 | 238794007 | 357897017 |
| Left ventricular thrombus | 748061000006117 | 309519009 | 452964014 |
| Ischaemia | 3353921000006110 | 52674009 | 496400017 |
| Acute deep venous thrombosis | 8041571000006119 | 132281000119108 | 2968256017 |
| Embolism and thrombosis of the iliac artery unspecified | 300556014 | 266262004 | 395794011 |
| [X]Embolism and thrombosis of other arteries | 300964011 | 266262004 | 395794011 |
| Vessel incision - embolectomy | 866261000006118 | 597641000000105 | 866261000006118 |
| Venous thrombosis NOS | 884751000006115 | 690131000000103 | 884751000006115 |
| Pulmonary embolus care | 6452821000006114 | 386280004 | 1480446016 |
| Thrombosis of pelvic vein | 4056781000006117 | 95448006 | 158101010 |
| Thrombosis of superior longitudinal sinus | 296938014 | 192760003 | 296938014 |
| Embolism and thrombosis of the anterior tibial artery | 300541017 | 195325004 | 300541017 |
| Embolism and thrombosis of the axillary artery | 300559019 | 195341003 | 300559019 |
| Deep venous thrombosis | 2162149016 | 128053003 | 194647015 |
| Embolectomy of subclavian artery | 5888791000006119 | 301080008 | 442323018 |
| [X] Embolism from prosthetic heart valve | 358931000006111 | 35688006 | 59527012 |
| Acute massive pulmonary embolism | 5057621000006119 | 233936003 | 350502011 |
| Omental infarction | 3722741000006113 | 75204008 | 124899011 |
| Embolism and thrombosis of other arteries NOS | 300563014 | 266262004 | 395794011 |
| Acute atrial infarction | 299719012 | 194809007 | 299719012 |
| Atrial thrombosis | 300234010 | 195147006 | 300234010 |
| Embolism and thrombosis of the radial artery | 300536013 | 195320009 | 300536013 |
| Embolism and thrombosis of the coeliac artery | 300561011 | 195342005 | 300561011 |
| Embolism and thrombosis of the vena cava | 300710016 | 195437003 | 300710016 |
| History of embolism | 5574351000006111 | 275545002 | 2986809014 |
| Provoked deep vein thrombosis | 2488381000000116 | 978441000000108 | 2488381000000116 |

| | | | |
|---|---|---|---|
| Acute deep vein thrombosis of left iliac vein | 12107691000006119 | 293491000119107 | 3644949016 |
| Cerebral venous thrombosis | 4056981000006115 | 95455008 | 158111015 |
| Arterial embolism/thrombosis | 884631000006118 | 266262004 | 884631000006118 |
| Thromboembolism | 6348271000006116 | 371039008 | 1228915016 |
| Percutaneous aspiration thromboembolectomy | 359871000000116 | 225361000000106 | 359871000000116 |
| Thrombectomy of arteriovenous fistula | 360031000000113 | 427992007 | 2694416018 |
| Ischaemic leg | 742481000006118 | 233961000 | 350546015 |
| Percutaneous venous thrombectomy | 1549601000006119 | 276977004 | 413381016 |
| Arterial embolectomy | 6580991000006118 | 397045002 | 1776771012 |
| Peripheral arterial embolism and thrombosis NOS | 12733151000006114 | 583731000000103 | 1295391000000110 |
| Thrombosis of vein NOS | 12728641000006110 | 690131000000103 | 1510411000000110 |
| Embolus of vein | 5059381000006115 | 234049002 | 350677014 |
| Incision of thrombosed haemorrhoid | 3984711000006118 | 91392002 | 201577017 |
| Thrombectomy of venous graft | 3654851000006117 | 71007004 | 117941017 |
| Transjugular intrahepatic thrombectomy of portal vein | 2691870012 | 429788001 | 2691870012 |
| Thrombectomy of vein | 6580651000006116 | 397028007 | 1776761010 |
| Thrombus | 6568121000006118 | 396339007 | 1776291018 |
| H/O: pulmonary embolus | 251693011 | 161512007 | 251693011 |
| Recurrent deep vein thrombosis | 2144741000000111 | 710167004 | 3043650017 |
| Open embolectomy of subclavian artery | 271633013 | 175440006 | 271633013 |
| Multi-infarct state | 5011261000006110 | 230704000 | 345662012 |
| Thrombosis of renal artery | 4059051000006119 | 95579008 | 158316015 |
| Deep venous thrombosis of lower limb | 6694771000006115 | 404223003 | 2162421016 |
| Thrombosis of vein of lower limb | 5992311000006119 | 309735004 | 453209018 |
| Aortic thromboembolism | 5560171000006116 | 274101000 | 409861010 |
| Incision and evacuation of thrombosed hemorrhoid | 5727391000006112 | 287799004 | 2547808014 |
| Femoral artery thrombosis | 3345371000006113 | 52156004 | 86808015 |
| Internal thrombosed piles | 884851000006114 | 52931009 | 884851000006114 |
| Open embolectomy of artery NEC | 12723621000006114 | 597641000000105 | 1324221000000116 |
| Ischaemic colitis | 12705101000006115 | 30588004 | 484643015 |
| Embolism and thrombosis of the abdominal aorta | 395796013 | 266263009 | 395796013 |
| Open embolectomy of common iliac artery | 411214011 | 275254000 | 411214011 |
| Right ventricular thrombosis | 452963015 | 309518001 | 452963015 |
| H/O: embolism | 411539015 | 275545002 | 411539015 |
| External thrombosed haemorrhoids | 483206018 | 26373009 | 483206018 |
| Infarction of ovary | 6297018 | 3129009 | 6297018 |
| Renal infarction | 75778012 | 45456005 | 75778012 |
| Other embolism and thrombosis | 300712012 | 429098002 | 2692478010 |
| Embolus of vein NOS | 300714013 | 234049002 | 350676017 |
| Mesenteric embolus NOS | 302983010 | 276500007 | 412653014 |
| Thrombosis of external jugular vein | 2234411000000115 | 864211000000100 | 2234411000000115 |
| Thrombosis of artery of transplanted kidney | 2234541000000115 | 864271000000105 | 2234541000000115 |

| | | | |
|---|---|---|---|
| Excision of external thrombotic haemorrhoid | 2591991000006114 | 5796001 | 194958014 |
| Percutaneous embolectomy of popliteal artery | 4668171000006118 | 175718001 | 272058018 |
| Chronic thromboembolic pulmonary hypertension | 5057801000006113 | 233947005 | 2968345015 |
| Incision and evacuation of thrombosed haemorrhoid | 5727401000006114 | 287799004 | 2547809018 |
| Critical ischaemia of foot | 6021731000006115 | 312822006 | 456609017 |
| Splenic infarction | 2866521000006111 | 22996003 | 38632019 |
| [RFC] Deep vein thrombosis | 909471000006114 | 909471000006105 | 909471000006114 |
| Thrombosed haemorrhoid - incised | 914791000006117 | 287799004 | 427359014 |
| Thromboembolic pulmonary hypertension | 350517010 | 233947005 | 350517010 |
| Percutaneous transluminal embolectomy of renal artery | 271705017 | 175489001 | 271705017 |
| Open embolectomy of femoral artery | 272031013 | 175704008 | 272031013 |
| Open femoral embolectomy | 272033011 | 175704008 | 272033011 |
| Open vein thrombectomy | 272244011 | 175843001 | 272244011 |
| Embolism of vein NOS | 1222321017 | 234049002 | 350676017 |
| [V] Personal history of pulmonary embolism | 451479013 | 161512007 | 251693011 |
| Percutaneous thrombolysis of intracranial artery | 5014421000006115 | 230935003 | 345980018 |
| Vertebral artery thrombosis | 3553621000006119 | 64775002 | 107661016 |
| Embolism of intracranial venous sinus | 2725621000006114 | 14246007 | 24228018 |
| Embolism superior longitudinal sinus | 296933017 | 192755002 | 296933017 |
| Embolism transverse sinus | 296935012 | 192757005 | 296935012 |
| Myelopathy due to arterial thrombosis of spinal cord | 297138017 | 192898007 | 297138017 |
| Embolism and thrombosis of the thoracic aorta | 300533017 | 195317001 | 300533017 |
| Embolism and thrombosis of the ulnar artery | 300537016 | 195321008 | 300537016 |
| Embolism and thrombosis of the popliteal artery | 300540016 | 195324000 | 300540016 |
| Other specified open removal of thrombus from vein | 272247016 | 175843001 | 272243017 |
| Thrombosed aneurysm | 4374441000006113 | 125271003 | 193761013 |
| Thrombosis of vein of transplanted kidney | 2234621000000111 | 864311000000105 | 2234621000000111 |
| Thrombosed external hemorrhoids | 2922191000006118 | 26373009 | 44177012 |
| LACI - Lacunar infarction | 5011171000006110 | 230698000 | 345653010 |
| Portal vein thrombosis | 30273012 | 17920008 | 30273012 |
| Pulmonary embolism | 98484016 | 59282003 | 98484016 |
| Infarction of breast | 128289017 | 77296004 | 128289017 |
| Thrombotic infarction | 3756351000006111 | 77255002 | 128233014 |
| Microvascular embolism of arteriole | 7855891000006114 | 724439005 | 3438135014 |
| Percutaneous thrombolysis of artery | 5593331000006113 | 276984007 | 413388010 |
| Ischaemic foot pain when walking | 2377911000000114 | 713674006 | 3295993015 |
| Thrombosed haemorrhoids | 300748014 | 75955007 | 200560013 |
| Thrombosis of mesenteric vein | 302984016 | 95446005 | 158099015 |
| Thromboembolism of vein | 300704016 | 429098002 | 2692478010 |
| Embolism central nervous system venous sinus NOS | 296936013 | 14246007 | 24228018 |
| Embolism and thrombosis of the hepatic artery | 300562016 | 195343000 | 3491726018 |

| | | | |
|---|---|---|---|
| Thrombotic microangiopathy NOS | 300587011 | 126729006 | 135665013 |
| Embolism and thrombosis NOS | 300713019 | 429098002 | 2692478010 |
| Thrombosis of internal jugular vein | 2234371000000119 | 864191000000104 | 2234371000000119 |
| Ischaemic foot pain at rest | 2377871000000112 | 713412006 | 3289284017 |
| Hepatic infarction | 30226018 | 17890003 | 30226018 |
| Thromboangiitis obliterans | 87208017 | 52403007 | 87208017 |
| Cardiac rupture due to and following acute myocardial infarction | 12221301000006119 | 233847009 | 3673284011 |
| DVT - Deep vein thrombosis | 4403261000006113 | 128053003 | 2162148012 |
| Thrombosis of corpus cavernosum | 3215871000006116 | 44437005 | 74102013 |
| Anterior spinal artery thrombosis | 345723011 | 230740003 | 345723011 |
| Peripheral ischaemia | 350533013 | 233958001 | 350533013 |
| Thrombosis of retinal vein | 3244261000006119 | 46085004 | 76829016 |
| Nephropathy due to ischaemia | 7674011000006115 | 710565001 | 3044790019 |
| Irreversible ischaemic colitis | 2936461000006116 | 27241008 | 483469013 |
| Axillary vein thrombosis | 851241000006115 | 851241000006104 | 851241000006115 |
| Embolus due to internal prosthetic device | 324950011 | 213131003 | 324950011 |
| Pulmonary infarct | 499917019 | 64662007 | 499917019 |
| Thromboembolic disease | 6348281000006118 | 371039008 | 1228916015 |
| Perianal venous thrombosis | 2608401000000118 | 449815008 | 2608401000000118 |
| Hepatic vein thrombosis | 64466019 | 82385007 | 136654014 |
| Infarct | 3403521000006118 | 55641003 | 92488010 |
| Deep vein thrombosis of lower limb | 2162422011 | 404223003 | 2162422011 |
| Retinal embolus | 6702781000006116 | 404667009 | 2156561018 |
| Embolism and thrombosis of hepatic artery | 4778991000006118 | 195343000 | 3491726018 |
| Arteriovenous fistula thrombosis | 350892013 | 234205007 | 350892013 |
| Renal artery stent thrombosis | 1809151000006116 | 1809151000006100 | 1809151000006116 |
| Superior mesenteric artery thrombosis | 100811000006112 | 197000003 | 302980013 |
| H/O: thromboembolism | 217601000000119 | 266996004 | 397830014 |
| Choroidal infarct | 6041261000006115 | 314003000 | 458273015 |
| Thromboembolus of external iliac artery | 3503839012 | 734299002 | 3503839012 |
| Acute lacunar infarction | 7054171000006115 | 426107000 | 2674093010 |
| Acute infarct | 3400671000006111 | 55470003 | 92217011 |
| Acute pulmonary embolism | 7609581000006110 | 706870000 | 3027244010 |
| Superficial femoral embolectomy | 6019821000006117 | 312689003 | 456443016 |
| Iliac vein thrombosis | 3636257010 | 762256003 | 3636257010 |
| Reason for referral: Deep Vein Thrombosis | 1776621000006117 | 1776621000006101 | 1776621000006117 |
| Percutaneous embolectomy of pulmonary artery | 4666331000006118 | 175268008 | 271401013 |
| Percutaneous embolectomy of aortoiliac segment | 4666711000006113 | 175356002 | 271520013 |
| Embolism and thrombosis NOS | 638821000006113 | 429098002 | 2692478010 |
| Retinal vein occlusion | 3244281000006112 | 46085004 | 494432015 |
| Thrombosis of vessels of cord | 2962491000006118 | 28773001 | 48182014 |
| Embolectomy operation | 5559451000006118 | 274021001 | 409761015 |

| | | | |
|---|---|---|---|
| Thrombosis of subclavian vein | 2795114019 | 438647008 | 2795114019 |
| History of thrombosis | 5574371000006118 | 275546001 | 2986432016 |
| Ischaemic nephropathy | 1757951000006119 | 1757951000006103 | 1757951000006119 |
| Arterial thrombosis | 218541000000116 | 65198009 | 108347015 |
| Thromboendarterectomy | 866221000006112 | 392031002 | 866221000006112 |
| Open thrombectomy of popliteal artery | 272034017 | 175705009 | 272034017 |
| Open embolectomy popliteal artery | 272035016 | 175705009 | 272035016 |
| Open thrombectomy of vein of lower limb | 272246013 | 175845008 | 272246013 |
| Declotting of thigh vein loop | 272283013 | 175873005 | 272283013 |
| High probability of deep vein thrombosis | 1141351000000111 | 513291000000109 | 1141351000000111 |
| Arterial embolism and thrombosis | 491241000006118 | 266262004 | 395795012 |
| Corpus cavernosum embolus | 1231106013 | 52303006 | 1231106013 |
| Blood clot | 3731671000006117 | 75753009 | 125815012 |
| Retinal microembolism | 169951000006111 | 76975009 | 127782012 |
| Cerebral venous thrombosis of sigmoid sinus | 5011651000006111 | 230721009 | 345698016 |
| Evacuation of thrombosed hemorrhoids | 3984731000006112 | 91392002 | 151417016 |
| Thrombosed hemorrhoids | 3735111000006119 | 75955007 | 126159013 |
| PE - Pulmonary embolism | 3462921000006119 | 59282003 | 1231937012 |
| Ischaemic lower limb pain at rest | 8278561000006114 | 836711000000108 | 2172511000000111 |
| H/O: thrombosis | 411540018 | 275546001 | 411540018 |
| Axillary vein thrombosis | 437622013 | 297156001 | 437622013 |
| Retinal vein thrombosis | 494431010 | 46085004 | 494431010 |
| Corpus cavernosum thrombosis | 1230092014 | 44437005 | 1230092014 |
| Renal artery embolus | 1235925019 | 95580006 | 1235925019 |
| Deep venous thrombosis of peroneal vein | 7273741000006119 | 443210003 | 2839752017 |
| Thrombosed hemorrhoid - incised | 5727371000006111 | 287799004 | 427358018 |
| Blood clot in eye | 5233761000006119 | 246682000 | 368495017 |
| Recurrent deep vein thrombosis | 1786921000006116 | 1786921000006100 | 1786921000006116 |
| Thrombosis of middle cerebral artery | 7598991000006116 | 705129007 | 3023377010 |
| Haemopericardium due to and following acute myocardial infarction | 12220761000006110 | 194862000 | 3673282010 |
| Penis vein thrombosis | 503317010 | 76598006 | 503317010 |
| Testicular thrombosis | 1232993010 | 67962003 | 1232993010 |
| H/O: Deep vein thrombosis | 451478017 | 161508001 | 2547692019 |
| [V] Personal history DVT (deep vein thrombosis) | 1227369013 | 161508001 | 2547692019 |
| Thrombosis lateral sinus | 100701000006118 | 21258007 | 1222620011 |
| Percutaneous transluminal thrombolysis of femoral graft using streptokinase | 238621000006112 | 309449009 | 452880015 |
| Open pulmonary embolectomy | 261021000006119 | 175262009 | 271393018 |
| Thrombosis of arteries of lower extremity | 3978101000006118 | 90958004 | 150730010 |
| Deep venous thrombosis - leg | 884731000006110 | 266267005 | 884731000006110 |
| External thrombosed piles | 884881000006118 | 26373009 | 884881000006118 |
| [RFC] Pulmonary embolism/pulmonary hypertension | 905451000006118 | 905451000006102 | 905451000006118 |

| | | | |
|---|---|---|---|
| [RFC] Arterial embolism of limbs | 905541000006119 | 905541000006103 | 905541000006119 |
| Evacuation of thrombosed haemorrhoid | 649671000006118 | 91392002 | 509428015 |
| Infarction of prostate | 36968017 | 22035000 | 36968017 |
| Spinal cord infarct | 7156221000006114 | 432249006 | 2770833019 |
| Post radiological embolism of lower limb artery | 216701000006116 | 312380005 | 456083019 |
| Percutaneous transluminal embolectomy of pulmonary artery | 232951000006114 | 175268008 | 271401013 |
| Renal vein thrombosis | 2751261000006119 | 15842009 | 26843012 |
| Ischaemic infarction of muscle | 359748014 | 240126009 | 359748014 |
| Arterial embolic and thrombotic occlusion | 395794011 | 266262004 | 395794011 |
| CRVO - Central retinal vein occlusion | 3614321000006112 | 68478007 | 501027019 |
| Limb ischaemia | 8009281000006110 | 21631000119105 | 2923092011 |
| Open embolectomy of superior mesenteric artery NEC | 271759019 | 397045002 | 1776771012 |
| Embolism and thrombosis of the femoral artery | 300539018 | 195323006 | 300539018 |
| Embolism and thrombosis of the dorsalis pedis artery | 300542012 | 195326003 | 300542012 |
| Embolism and thrombosis of the subclavian artery | 300557017 | 195339004 | 300557017 |
| Embolism and thrombosis of the renal vein | 300711017 | 195438008 | 300711017 |
| Scrotal thrombosis | 304527018 | 198059008 | 304527018 |
| H/O: Deep Vein Thrombosis | 251689017 | 161508001 | 251689017 |
| Percutaneous embolectomy of femoral artery | 4668151000006111 | 175717006 | 272054016 |
| Thromboembolic pulmonary hypertension | 1749291000006113 | 1749291000006109 | 1749291000006113 |
| Post-radiological embolism of lower limb artery | 6015601000006112 | 312380005 | 456083019 |
| Thrombosed internal hemorrhoids | 3358351000006111 | 52931009 | 88072011 |
| Infarct of prostate | 2850821000006117 | 22035000 | 1222704012 |
| Superficial vein thrombosis | 5574011000006113 | 275517008 | 411509013 |
| Open embolectomy of brachial artery | 271635018 | 175442003 | 271635018 |
| Open embolectomy of vertebral artery | 271636017 | 175443008 | 271636017 |
| Percutaneous transluminal embolectomy of vertebral artery | 271657013 | 175458000 | 271657013 |
| Open embolectomy of renal artery | 271695015 | 175482005 | 271695015 |
| Open removal of thrombus from vein | 272243017 | 175843001 | 272243017 |
| Open thrombectomy of vein of upper limb | 272245012 | 175844007 | 272245012 |

#### Table S30. Arterial thromboembolic Aurum codes

| Term | Medcode ID | SNOMED CT Concept ID | SNOMED CT Description ID |
|---|---|---|---|
| Embolism and/or thrombosis of the internal iliac artery | 300554012 | 734298005 | 3503835018 |
| Coronary thrombosis not resulting in myocardial infarction | 299742017 | 194821006 | 299742017 |
| Embolism and thrombosis of the brachial artery | 300535012 | 195319003 | 300535012 |
| Embolism and thrombosis of the posterior tibial artery | 300543019 | 195327007 | 300543019 |
| Mesenteric embolus | 412653014 | 276500007 | 412653014 |
| Ischaemic foot | 443199013 | 301755001 | 443199013 |
| Open removal of cardiac thrombus | 2675367019 | 426067000 | 2675367019 |
| Percutaneous transluminal thrombolysis of artery | 2675878019 | 426485003 | 2675878019 |
| Ischaemic disease of gut | 3836741000006113 | 82196007 | 504940010 |
| Acute intestinal ischaemic syndrome | 3986491000006113 | 91489000 | 509476019 |
| Intestinal ischaemia | 3836721000006118 | 82196007 | 504938017 |
| Embolism and thrombosis of the celiac artery | 4778981000006116 | 195342005 | 300560012 |
| Ischaemia reperfusion injury of liver | 7821701000006113 | 721715000 | 3325835015 |
| AMI - Acute mesenteric ischaemia | 3986481000006110 | 91489000 | 509475015 |
| Inner retinal ischaemic spots | 3140891000006117 | 39832008 | 2620723015 |
| Thrombosis of renal artery bypass graft | 449993015 | 306857000 | 449993015 |
| Ischaemic toe | 450665015 | 307408003 | 450665015 |
| Mesenteric thrombus and/or embolus | 450682018 | 307420006 | 450682018 |
| Infarct of liver | 2783881000006119 | 17890003 | 30227010 |
| Ischaemic bowel disease | 3836731000006115 | 82196007 | 504939013 |
| Ischaemic optic neuropathy | 476238012 | 14357004 | 476238012 |
| Mesenteric thrombosis | 215511000000110 | 95446005 | 158099015 |
| Post radiological embolism of upper limb artery | 216711000006118 | 312377009 | 456079019 |
| Ischaemic arm | 5058031000006112 | 233959009 | 350538016 |
| Pulmonary arterial thrombosis | 3439711000006111 | 57834008 | 96166019 |
| Male genital thrombosis NOS | 304536019 | 198057005 | 304524013 |
| Ischaemic necrosis | 3901731000006110 | 86217007 | 142974016 |
| Vascular graft embolus | 5925451000006112 | 304090004 | 446358019 |
| Superior mesenteric artery embolus | 4785631000006110 | 196999001 | 302978019 |
| Acute ischaemic colitis | 503091015 | 75700000 | 503091015 |
| Thyroid infarction | 1220407012 | 11859008 | 1220407012 |
| Acute infarction of papillary muscle | 2663441000006117 | 10273003 | 17900011 |
| Removal of vessel clot | 866201000006119 | 392031002 | 866201000006119 |
| Arteritic ischaemic optic neuropathy | 5008941000006114 | 230508004 | 345412011 |
| Peripheral ischaemic vascular disease | 350535018 | 233958001 | 350535018 |
| Upper limb ischaemia | 350540014 | 233959009 | 350540014 |
| Open embolectomy of bifurcation of aorta | 271509013 | 175348000 | 271509013 |
| Open embolectomy of carotid artery | 271540018 | 175374006 | 271540018 |
| Open embolectomy of axillary artery | 271639012 | 175446000 | 271639012 |

| | | | |
|---|---|---|---|
| Percutaneous transluminal embolectomy of subclavian artery | 271654018 | 175456001 | 271654018 |
| Percutaneous transluminal embolectomy of brachial artery | 271656016 | 175457005 | 271656016 |
| Percutaneous transluminal embolectomy of axillary artery | 271671017 | 175465008 | 271671017 |
| Percutaneous transluminal embolectomy of iliac artery | 271901019 | 175613000 | 271901019 |
| Open thrombectomy of femoral artery | 272032018 | 175704008 | 272032018 |
| Percutaneous transluminal embolectomy of femoral artery | 272055015 | 175717006 | 272055015 |
| Percutaneous transluminal embolectomy of popliteal artery | 272056019 | 175718001 | 272056019 |
| Prosthetic graft thrombectomy | 272305017 | 175891000 | 272305017 |
| Infarction of fallopian tube | 94629010 | 56900004 | 94629010 |
| Infarct of lung | 3551761000006115 | 64662007 | 107460013 |
| Cerebral arterial thrombosis | 3662321000006117 | 71444005 | 118691019 |
| Ischaemic foot | 1699111000006110 | 1699111000006106 | 1699111000006110 |
| Percutaneous mechanical thromboembolectomy | 359731000000112 | 225291000000108 | 359731000000112 |
| Carotid thromboendarterectomy | 3588601000006110 | 66951008 | 111209010 |
| Saddle embolus of abdominal aorta | 5058281000006118 | 233972005 | 350565011 |
| Peripheral arterial embolism | 884661000006110 | 583731000000103 | 884661000006110 |
| Embolus/thrombus artery NOS | 884671000006115 | 583761000000108 | 884671000006115 |
| Embolism | 887751000006110 | 198850008 | 887751000006110 |
| Arterial embolism and thrombosis NOS | 12733161000006111 | 583761000000108 | 1295451000000114 |
| Cholesterol embolus syndrome | 354495010 | 236489002 | 354495010 |
| Fogarty embolectomy of artery | 1776773010 | 397047005 | 1776773010 |
| Thrombosis - coronary | 1786198013 | 398274000 | 1786198013 |
| Thrombosis of cerebral arteries | 3662311000006113 | 71444005 | 118690018 |
| Thromboendarterectomy of aorta | 3295821000006118 | 49187005 | 81937019 |
| PC TL embolectomy popliteal artery | 4668191000006117 | 175718001 | 272057011 |
| Thrombosis of testis | 3605531000006118 | 67962003 | 112890010 |
| Cerebral arterial embolism | 3728451000006116 | 75543006 | 125471016 |
| IRIS - Inner retinal ischaemic spots | 3140901000006118 | 39832008 | 2620724014 |
| Renal infarction | 1848131000006113 | 45456005 | 75778012 |
| Ischaemic nephropathy | 1160091000000115 | 710565001 | 3082872011 |
| Release of ischaemic contracture of forearm | 4682421000006112 | 178253006 | 275890019 |
| Open embolectomy of inferior mesenteric artery NEC | 271760012 | 397045002 | 1776771012 |
| Open embolectomy of suprarenal artery NEC | 271761011 | 397045002 | 1776771012 |
| Open embolectomy of visceral branch of abdominal aorta NEC | 271780013 | 397045002 | 1776771012 |
| Open embolectomy of artery NEC | 272097014 | 397045002 | 1776771012 |
| Open embolectomy of iliac artery | 394332017 | 265521000 | 394332017 |
| Renal artery thrombosis | 158317012 | 95579008 | 158317012 |
| Percutaneous thrombolysis of vein | 5593341000006115 | 276985008 | 413389019 |
| Brachial embolectomy | 5048991000006113 | 233313002 | 349588014 |

| | | | |
|---|---|---|---|
| Percutaneous transluminal embolectomy of artery NEC | 394347014 | 397045002 | 1776771012 |
| Chronic intestinal ischaemia | 4194521000006117 | 111354009 | 361442014 |
| Embolism and thrombosis of other specified artery | 300552011 | 266262004 | 395794011 |
| Arterial embolism and thrombosis NOS | 300564015 | 266262004 | 395794011 |
| Infarction of thyroid | 2688811000006110 | 11859008 | 20465014 |
| Occlusive thromboarteriopathy | 6206441000006110 | 359789008 | 475141015 |
| Percutaneous transluminal thrombolysis of blood vessel of liver | 1545591000006112 | 426059008 | 2675452017 |
| Percutaneous transluminal thrombolysis and reconstruction of artery | 1548401000006112 | 428068004 | 2696045014 |
| Aortic bifurcation embolus | 5058291000006115 | 233972005 | 350566012 |
| Acute colonic ischaemia | 3730871000006110 | 75700000 | 2310141000000117 |
| Embolectomy | 3668011000006113 | 71815002 | 119306015 |
| Embolism and thrombosis of a leg artery NOS | 300547018 | 195318006 | 300534011 |
| Peripheral arterial embolism and thrombosis NOS | 300550015 | 195318006 | 300534011 |
| Renal artery embolism | 158318019 | 95580006 | 158318019 |
| Embolectomy of popliteal artery | 5049001000006113 | 233314008 | 349592019 |
| Multiple infarcts | 2947401000006115 | 27876002 | 46655014 |
| Infarction of kidney | 3233381000006117 | 45456005 | 75780018 |
| Ocular ischaemic syndrome | 6045981000006117 | 314436005 | 458797013 |
| Mesenteric embolism | 412652016 | 276500007 | 412652016 |
| Vertebrobasilar ischaemic vertigo | 5034291000006111 | 232288005 | 348027016 |
| Infarction | 3403531000006115 | 55641003 | 92491010 |
| Embolism in vascular graft | 446359010 | 304090004 | 446359010 |
| Thromboembolus of internal iliac artery | 3503835018 | 734298005 | 3503835018 |
| SMAE - Superior mesenteric artery embolus | 4785641000006117 | 196999001 | 302979010 |
| Thrombosis of stent of renal artery | 1848121000006110 | 840961000000108 | 2182101000000113 |
| Popliteal artery thrombosis | 3918361000006110 | 87239004 | 144656012 |
| Arterial embolism | 3387371000006112 | 54687002 | 90875013 |
| Stroke due to basilar artery thrombus | 3508956015 | 329641000119104 | 3508956015 |
| Femoral embolectomy | 2634331000006113 | 8409001 | 1234899012 |
| Hemopericardium due to and following acute myocardial infarction | 12220751000006113 | 194862000 | 3673281015 |
| Cerebral infarction due to occlusion of precerebral artery | 542261000006114 | 125081000119106 | 3042974014 |
| Carotid artery thrombosis | 100771000006112 | 86003009 | 142588012 |
| Occlusive mesenteric ischaemia | 5086201000006115 | 235842000 | 353530010 |
| Multiple lacunar infarcts | 5967391000006111 | 307363008 | 450609012 |
| Thyroid hemorrhage and infarction | 4757961000006112 | 190305000 | 292442019 |
| Non-occlusive thrombus | 2927841000006117 | 26713004 | 44719012 |
| Open embolectomy of coeliac artery NEC | 271757017 | 397045002 | 1776771012 |
| Multifocal infarct | 3474711000006110 | 60007006 | 99691019 |
| Infarction of spleen | 38632019 | 22996003 | 1223010014 |
| Thrombectomy | 3205401000006110 | 43810009 | 73061013 |

| | | | |
|---|---|---|---|
| Ischaemic hepatitis | 5086761000006113 | 235877000 | 353591011 |
| Ischaemic gangrene | 6670921000006117 | 402861007 | 1773672019 |
| Thrombosis of atrium, auricular appendage, and ventricle due to and following acute myocardial infarction | 12220791000006119 | 194868001 | 3673277015 |
| Lacunar infarction | 299342019 | 230698000 | 345651012 |
| Embolectomy of iliac artery | 5153821000006115 | 240928009 | 360865017 |
| Embolectomy of femoral artery | 2634321000006110 | 8409001 | 14892019 |
| Ischaemic ulcer of toe | 7108391000006114 | 429768000 | 2693575017 |
| Removal of thrombus of arteriovenous fistula | 7081801000006114 | 427992007 | 2691808018 |
| Ischemic infarction of muscle | 5143791000006114 | 240126009 | 359749018 |
| Thyroid haemorrhage or infarction NOS | 292445017 | 190305000 | 292443012 |
| Thromboangiitis obliterans NOS | 300503012 | 52403007 | 87208017 |
| Embolism and thrombosis of an arm artery NOS | 300538014 | 195318006 | 300534011 |
| Thrombosis | 7218941000006118 | 439127006 | 2794470015 |
| Thrombosis | 5477661000006119 | 264579008 | 393059012 |
| Saddle embolus | 350563016 | 233972005 | 350563016 |
| Infarction of liver | 2783891000006116 | 17890003 | 1221609013 |
| Femoral artery embolus | 5058301000006119 | 233973000 | 350567015 |
| Critical lower limb ischaemia | 5058111000006119 | 233962007 | 350547012 |
| Subclavian artery thrombosis | 5836921000006114 | 297154003 | 437620017 |
| Lacunar infarct | 3818181000006114 | 81037000 | 134436018 |
| Ischaemia of feet | 5886711000006111 | 300917007 | 442114010 |
| Embolism and/or thrombosis of the external iliac artery | 300555013 | 734299002 | 3503839012 |
| Renal infarct | 3233391000006119 | 45456005 | 75781019 |
| Embolectomy of carotid artery | 5902211000006114 | 302053004 | 443591015 |
| Lower limb arterial embolus | 6015571000006117 | 312378004 | 456080016 |
| Embolectomy of lower limb artery | 3688071000006119 | 73038007 | 121315013 |
| Thrombolysis of artery | 5049421000006111 | 233335005 | 349633016 |
| Thyroid haemorrhage and infarction | 292443012 | 190305000 | 292443012 |
| Embolism and thrombosis of an arm or leg artery | 300534011 | 195318006 | 300534011 |
| Embolism and/or thrombosis of the common iliac artery | 300553018 | 195335005 | 300553018 |
| Embolism and thrombosis of the splenic artery | 300558010 | 195340002 | 300558010 |
| Acute intestinal ischaemia | 3986461000006117 | 91489000 | 509473010 |
| Post-radiological embolism of upper limb artery | 6015561000006112 | 312377009 | 456079019 |
| Colonic ischaemia | 2992791000006113 | 30588004 | 1216341014 |
| Ischaemic priapism | 7252011000006114 | 441575009 | 2817057019 |
| Digital arterial thrombosis | 5886741000006110 | 300919005 | 442117015 |
| Percutaneous transluminal venous thrombolysis NEC | 358041000000114 | 276985008 | 413389019 |
| Percutaneous removal of thrombus from vein NOS | 359571000000118 | 276977004 | 413381016 |
| Lower limb ischaemia | 5058071000006110 | 233961000 | 350544017 |
| Arterial embolus | 3387391000006113 | 54687002 | 497013010 |

| | | | |
|---|---|---|---|
| Embolism | 3402611000006115 | 55584005 | 92392016 |
| Percutaneous transluminal embolectomy of bifurcation of aorta | 232881000006113 | 175356002 | 271519019 |
| Embolus/thrombosis abd. aorta | 884641000006111 | 266263009 | 884641000006111 |
| Embolus/thrombosis aorta NOS | 884651000006113 | 195317001 | 884651000006113 |
| Removal of thrombus from intracranial artery | 2845481000006110 | 21710002 | 36433012 |
| Percutaneous embolectomy of brachial artery | 4667301000006111 | 175457005 | 271655017 |
| Acute mesenteric ischaemia | 457631000006114 | 91489000 | 509474016 |
| Mesenteric infarction | 523601000006115 | 3558002 | 7001012 |
| Embolism and thrombosis of other and unspec parts aorta | 638871000006114 | 274101000 | 409861010 |
| Embolus of the superior mesenteric artery | 639241000006110 | 196999001 | 302978019 |
| Percutaneous transluminal thrombectomy of blood vessel of liver | 1545581000006114 | 428953003 | 2692834012 |
| Percutaneous transluminal venous thrombolysis with reconstruction | 1549761000006110 | 429141007 | 1659501000000119 |
| Retinal ischaemia | 483237018 | 26468004 | 483237018 |
| Ischaemic | 5360051000006112 | 255426005 | 380631011 |
| Asymptomatic ischaemia | 5056261000006112 | 233823002 | 350356015 |
| Embolism of iliac artery | 3807321000006113 | 80383008 | 133386017 |
| Arterial embolus and thrombosis | 395795012 | 266262004 | 395795012 |
| Occlusive thrombus | 3771791000006113 | 78195007 | 129763011 |
| Arterial ischaemia | 5057951000006115 | 233957006 | 350532015 |
| Intracranial thrombectomy | 2845491000006113 | 21710002 | 36434018 |
| Acute ischaemia of small intestine | 8321371000006112 | 906191000000102 | 2329961000000119 |
| Rupture of chordae tendinae due to and following acute myocardial infarction | 12220781000006117 | 194866002 | 3673278013 |
| Ischaemic finger | 5967971000006111 | 307409006 | 450668018 |
| Ischaemic ulcer | 2721231000006112 | 13954005 | 475615013 |
| Mural thrombosis | 1235655012 | 91335003 | 1235655012 |
| Ischaemic leg ulcer | 357895013 | 238793001 | 357895013 |
| Ischaemic foot ulcer | 5126451000006116 | 238794007 | 357897017 |
| Left ventricular thrombus | 748061000006117 | 309519009 | 452964014 |
| Ischaemia | 3353921000006110 | 52674009 | 496400017 |
| Embolism and thrombosis of the iliac artery unspecified | 300556014 | 266262004 | 395794011 |
| [X]Embolism and thrombosis of other arteries | 300964011 | 266262004 | 395794011 |
| Vessel incision - embolectomy | 866261000006118 | 597641000000105 | 866261000006118 |
| Embolism and thrombosis of the anterior tibial artery | 300541017 | 195325004 | 300541017 |
| Embolism and thrombosis of the axillary artery | 300559019 | 195341003 | 300559019 |
| Embolectomy of subclavian artery | 5888791000006119 | 301080008 | 442323018 |
| [X] Embolism from prosthetic heart valve | 358931000006111 | 35688006 | 59527012 |
| Omental infarction | 3722741000006113 | 75204008 | 124899011 |
| Embolism and thrombosis of other arteries NOS | 300563014 | 266262004 | 395794011 |
| Acute atrial infarction | 299719012 | 194809007 | 299719012 |
| Atrial thrombosis | 300234010 | 195147006 | 300234010 |

| | | | |
|---|---|---|---|
| Embolism and thrombosis of the radial artery | 300536013 | 195320009 | 300536013 |
| Embolism and thrombosis of the coeliac artery | 300561011 | 195342005 | 300561011 |
| Arterial embolism/thrombosis | 884631000006118 | 266262004 | 884631000006118 |
| Thromboembolism | 6348271000006116 | 371039008 | 1228915016 |
| Percutaneous aspiration thromboembolectomy | 359871000000116 | 225361000000106 | 359871000000116 |
| Thrombectomy of arteriovenous fistula | 360031000000113 | 427992007 | 2694416018 |
| Ischaemic leg | 742481000006118 | 233961000 | 350546015 |
| Arterial embolectomy | 6580991000006118 | 397045002 | 1776771012 |
| Peripheral arterial embolism and thrombosis NOS | 12733151000006114 | 583731000000103 | 1295391000000110 |
| Thrombus | 6568121000006118 | 396339007 | 1776291018 |
| Open embolectomy of subclavian artery | 271633013 | 175440006 | 271633013 |
| Multi-infarct state | 5011261000006110 | 230704000 | 345662012 |
| Thrombosis of renal artery | 4059051000006119 | 95579008 | 158316015 |
| Aortic thromboembolism | 5560171000006116 | 274101000 | 409861010 |
| Femoral artery thrombosis | 3345371000006113 | 52156004 | 86808015 |
| Open embolectomy of artery NEC | 12723621000006114 | 597641000000105 | 1324221000000116 |
| Ischaemic colitis | 12705101000006115 | 30588004 | 484643015 |
| Embolism and thrombosis of the abdominal aorta | 395796013 | 266263009 | 395796013 |
| Open embolectomy of common iliac artery | 411214011 | 275254000 | 411214011 |
| H/O: embolism | 411539015 | 275545002 | 411539015 |
| Infarction of ovary | 6297018 | 3129009 | 6297018 |
| Renal infarction | 75778012 | 45456005 | 75778012 |
| Other embolism and thrombosis | 300712012 | 429098002 | 2692478010 |
| Mesenteric embolus NOS | 302983010 | 276500007 | 412653014 |
| Thrombosis of artery of transplanted kidney | 2234541000000115 | 864271000000105 | 2234541000000115 |
| Percutaneous embolectomy of popliteal artery | 4668171000006118 | 175718001 | 272058018 |
| Critical ischaemia of foot | 6021731000006115 | 312822006 | 456609017 |
| Splenic infarction | 2866521000006111 | 22996003 | 38632019 |
| Percutaneous transluminal embolectomy of renal artery | 271705017 | 175489001 | 271705017 |
| Open embolectomy of femoral artery | 272031013 | 175704008 | 272031013 |
| Open femoral embolectomy | 272033011 | 175704008 | 272033011 |
| Percutaneous thrombolysis of intracranial artery | 5014421000006115 | 230935003 | 345980018 |
| Vertebral artery thrombosis | 3553621000006119 | 64775002 | 107661016 |
| Myelopathy due to arterial thrombosis of spinal cord | 297138017 | 192898007 | 297138017 |
| Embolism and thrombosis of the thoracic aorta | 300533017 | 195317001 | 300533017 |
| Embolism and thrombosis of the ulnar artery | 300537016 | 195321008 | 300537016 |
| Embolism and thrombosis of the popliteal artery | 300540016 | 195324000 | 300540016 |
| Thrombosed aneurysm | 4374441000006113 | 125271003 | 193761013 |
| LACI - Lacunar infarction | 5011171000006110 | 230698000 | 345653010 |
| Infarction of breast | 128289017 | 77296004 | 128289017 |
| Thrombotic infarction | 3756351000006111 | 77255002 | 128233014 |

| | | | |
|---|---|---|---|
| Microvascular embolism of arteriole | 7855891000006114 | 724439005 | 3438135014 |
| Percutaneous thrombolysis of artery | 5593331000006113 | 276984007 | 413388010 |
| Ischaemic foot pain when walking | 2377911000000114 | 713674006 | 3295993015 |
| Embolism and thrombosis of the hepatic artery | 300562016 | 195343000 | 3491726018 |
| Thrombotic microangiopathy NOS | 300587011 | 126729006 | 135665013 |
| Embolism and thrombosis NOS | 300713019 | 429098002 | 2692478010 |
| Ischaemic foot pain at rest | 2377871000000112 | 713412006 | 3289284017 |
| Hepatic infarction | 30226018 | 17890003 | 30226018 |
| Thromboangiitis obliterans | 87208017 | 52403007 | 87208017 |
| Cardiac rupture due to and following acute myocardial infarction | 12221301000006119 | 233847009 | 3673284011 |
| Thrombosis of corpus cavernosum | 3215871000006116 | 44437005 | 74102013 |
| Anterior spinal artery thrombosis | 345723011 | 230740003 | 345723011 |
| Peripheral ischaemia | 350533013 | 233958001 | 350533013 |
| Nephropathy due to ischaemia | 7674011000006115 | 710565001 | 3044790019 |
| Irreversible ischaemic colitis | 2936461000006116 | 27241008 | 483469013 |
| Embolus due to internal prosthetic device | 324950011 | 213131003 | 324950011 |
| Thromboembolic disease | 6348281000006118 | 371039008 | 1228916015 |
| Infarct | 3403521000006118 | 55641003 | 92488010 |
| Retinal embolus | 6702781000006116 | 404667009 | 2156561018 |
| Embolism and thrombosis of hepatic artery | 4778991000006118 | 195343000 | 3491726018 |
| Arteriovenous fistula thrombosis | 350892013 | 234205007 | 350892013 |
| Renal artery stent thrombosis | 1809151000006116 | 1809151000006100 | 1809151000006116 |
| Superior mesenteric artery thrombosis | 100811000006112 | 197000003 | 302980013 |
| Choroidal infarct | 6041261000006115 | 314003000 | 458273015 |
| Thromboembolus of external iliac artery | 3503839012 | 734299002 | 3503839012 |
| Acute lacunar infarction | 7054171000006115 | 426107000 | 2674093010 |
| Acute infarct | 3400671000006111 | 55470003 | 92217011 |
| Superficial femoral embolectomy | 6019821000006117 | 312689003 | 456443016 |
| Percutaneous embolectomy of aortoiliac segment | 4666711000006113 | 175356002 | 271520013 |
| Embolism and thrombosis NOS | 638821000006113 | 429098002 | 2692478010 |
| Thrombosis of vessels of cord | 2962491000006118 | 28773001 | 48182014 |
| Embolectomy operation | 5559451000006118 | 274021001 | 409761015 |
| History of thrombosis | 5574371000006118 | 275546001 | 2986432016 |
| Ischaemic nephropathy | 1757951000006119 | 1757951000006103 | 1757951000006119 |
| Arterial thrombosis | 218541000000116 | 65198009 | 108347015 |
| Thromboendarterectomy | 866221000006112 | 392031002 | 866221000006112 |
| Open thrombectomy of popliteal artery | 272034017 | 175705009 | 272034017 |
| Open embolectomy popliteal artery | 272035016 | 175705009 | 272035016 |
| Arterial embolism and thrombosis | 491241000006118 | 266262004 | 395795012 |
| Retinal microembolism | 169951000006111 | 76975009 | 127782012 |
| Ischaemic lower limb pain at rest | 8278561000006114 | 836711000000108 | 2172511000000111 |
| Corpus cavernosum thrombosis | 1230092014 | 44437005 | 1230092014 |

| | | | |
|---|---|---|---|
| Renal artery embolus | 1235925019 | 95580006 | 1235925019 |
| Thrombosis of middle cerebral artery | 7598991000006116 | 705129007 | 3023377010 |
| Haemopericardium due to and following acute myocardial infarction | 12220761000006110 | 194862000 | 3673282010 |
| Testicular thrombosis | 1232993010 | 67962003 | 1232993010 |
| Percutaneous transluminal thrombolysis of femoral graft using streptokinase | 238621000006112 | 309449009 | 452880015 |
| Thrombosis of arteries of lower extremity | 3978101000006118 | 90958004 | 150730010 |
| [RFC] Arterial embolism of limbs | 905541000006119 | 905541000006103 | 905541000006119 |
| Infarction of prostate | 36968017 | 22035000 | 36968017 |
| Spinal cord infarct | 7156221000006114 | 432249006 | 2770833019 |
| Post radiological embolism of lower limb artery | 216701000006116 | 312380005 | 456083019 |
| Ischaemic infarction of muscle | 359748014 | 240126009 | 359748014 |
| Arterial embolic and thrombotic occlusion | 395794011 | 266262004 | 395794011 |
| Limb ischaemia | 8009281000006110 | 21631000119105 | 2923092011 |
| Open embolectomy of superior mesenteric artery NEC | 271759019 | 397045002 | 1776771012 |
| Embolism and thrombosis of the femoral artery | 300539018 | 195323006 | 300539018 |
| Embolism and thrombosis of the dorsalis pedis artery | 300542012 | 195326003 | 300542012 |
| Embolism and thrombosis of the subclavian artery | 300557017 | 195339004 | 300557017 |
| Scrotal thrombosis | 304527018 | 198059008 | 304527018 |
| Percutaneous embolectomy of femoral artery | 4668151000006111 | 175717006 | 272054016 |
| Post-radiological embolism of lower limb artery | 6015601000006112 | 312380005 | 456083019 |
| Infarct of prostate | 2850821000006117 | 22035000 | 1222704012 |
| Open embolectomy of brachial artery | 271635018 | 175442003 | 271635018 |
| Open embolectomy of vertebral artery | 271636017 | 175443008 | 271636017 |
| Percutaneous transluminal embolectomy of vertebral artery | 271657013 | 175458000 | 271657013 |
| Open embolectomy of renal artery | 271695015 | 175482005 | 271695015 |

#### Table S31. Thromboembolic HES (ICD10) codes

| ICD10 codes | Term |
|---|---|
| D73.5 | Infarction of spleen |
| H34.0 | Transient retinal artery occlusion |
| H34.1 | Central retinal artery occlusion |
| H34.2 | Other retinal artery occlusions |
| H34.8 | Other retinal vascular occlusions |
| H34.9 | Retinal vascular occlusion, unspecified |
| I23.6 | Thrombosis of atrium, auricular appendage, and ventricle as current complications following acute myocardial infarction |
| I26.0 | Pulmonary embolism with mention of acute cor pulmonale |
| I26.9 | Pulmonary embolism without mention of acute cor pulmonale |
| I51.3 | Intracardiac thrombosis, not elsewhere classified |
| I65.0 | Occlusion and stenosis of vertebral artery |
| I65.1 | Occlusion and stenosis of basilar artery |
| I65.2 | Occlusion and stenosis of carotid artery |
| I65.3 | Occlusion and stenosis of multiple and bilateral precerebral arteries |
| I65.8 | Occlusion and stenosis of other precerebral artery |
| I65.9 | Occlusion and stenosis of unspecified precerebral artery |
| I66.0 | Occlusion and stenosis of middle cerebral artery |
| I66.1 | Occlusion and stenosis of anterior cerebral artery |
| I66.2 | Occlusion and stenosis of posterior cerebral artery |
| I66.3 | Occlusion and stenosis of cerebellar arteries |
| I66.4 | Occlusion and stenosis of multiple and bilateral cerebral arteries |
| I66.8 | Occlusion and stenosis of other cerebral artery |
| I66.9 | Occlusion and stenosis of unspecified cerebral artery |
| I67.6 | Nonpyogenic thrombosis of intracranial venous system |
| I73.1 | Thromboangiitis obliterans [Buerger] |
| I74.0 | Embolism and thrombosis of abdominal aorta |
| I74.1 | Embolism and thrombosis of other and unspecified parts of aorta |
| I74.2 | Embolism and thrombosis of arteries of upper extremities |
| I74.3 | Embolism and thrombosis of arteries of lower extremities |
| I74.4 | Embolism and thrombosis of arteries of extremities, unspecified |
| I74.5 | Embolism and thrombosis of iliac artery |
| I74.8 | Embolism and thrombosis of other arteries |
| I74.9 | Embolism and thrombosis of unspecified artery |
| I80.0 | Phlebitis and thrombophlebitis of superficial vessels of lower extremities |
| I80.1 | Phlebitis and thrombophlebitis of femoral vein |
| I80.2 | Phlebitis and thrombophlebitis of other deep vessels of lower extremities |
| I80.3 | Phlebitis and thrombophlebitis of lower extremities, unspecified |
| I80.8 | Phlebitis and thrombophlebitis of other sites |
| I80.9 | Phlebitis and thrombophlebitis of unspecified site |

| | |
|---|---|
| I81 | Portal vein thrombosis |
| I82.1 | Thrombophlebitis migrans |
| I82.2 | Embolism and thrombosis of vena cava |
| I82.3 | Embolism and thrombosis of renal vein |
| I82.8 | Embolism and thrombosis of other specified veins |
| I82.9 | Embolism and thrombosis of unspecified vein |
| I84.0 | Internal thrombosed haemorrhoids |
| I84.3 | External thrombosed haemorrhoids |
| I84.7 | Unspecified thrombosed haemorrhoids |
| K55.0 | Acute vascular disorders of intestine |
| K55.9 | Vascular disorder of intestine, unspecified |
| K64.5 | Perianal venous thrombosis |
| K76.3 | Infarction of liver |
| M31.1 | Thrombotic microangiopathy |
| M62.2 | Ischaemic infarction of muscle |
| M62.20 | Ischaemic infarction of muscle, unspecified site |
| M62.21 | Ischaemic infarction of muscle, shoulder |
| M62.22 | Ischaemic infarction of muscle, upper arm |
| M62.23 | Ischaemic infarction of muscle, forearm |
| M62.24 | Ischaemic infarction of muscle, hand |
| M62.25 | Ischaemic infarction of muscle, thigh |
| M62.26 | Ischaemic infarction of muscle, lower leg |
| M62.27 | Ischaemic infarction of muscle, ankle and foot |
| M62.28 | Ischaemic infarction of muscle, other site |
| N28.0 | Ischaemia and infarction of kidney |

#### Table S32. Myocardial infarction outcome Aurum codes

| Term | Medcode ID | SNOMED CT Concept ID | SNOMED CT Description ID |
|---|---|---|---|
| Electrocardiogram: lateral infarction | 4586241000006113 | 164871004 | 2619220010 |
| Acute anteroapical infarction | 299708014 | 52035003 | 3038718019 |
| Subsequent myocardial infarction of inferior wall | 299812011 | 194858006 | 299812011 |
| Acute Q-wave infarct | 447324018 | 304914007 | 447324018 |
| Electrocardiographic subendocardial infarct | 4586211000006114 | 164870003 | 3300442017 |
| Acute transmural myocardial infarction of unspecif site | 460681000006116 | 57054005 | 94884017 |
| Postmyocardial infarction syndrome | 3576371000006117 | 66189004 | 109915012 |
| Acute non-Q wave infarction | 450322013 | 307140009 | 450322013 |
| H/O: Myocardial infarction in last year | 451369010 | 308065005 | 451369010 |
| Post infarct angina | 458410010 | 314116003 | 458410010 |
| ECG: antero-septal infarct. | 256455012 | 164868007 | 256455012 |
| Myocardial infarct | 2855351000006111 | 22298006 | 1784873012 |
| Acute myocardial infarction of inferolateral wall | 3565871000006113 | 65547006 | 108912018 |
| Acute inferior myocardial infarction | 3699921000006110 | 73795002 | 1233665015 |
| Other acute and subacute ischaemic heart disease | 39111000006114 | 414545008 | 2534663012 |
| Lateral infarction on electrocardiogram | 4586221000006118 | 164871004 | 2692104017 |
| Electrocardiographic myocardial infarction | 4586041000006115 | 164865005 | 3300376010 |
| Post-infarction pericarditis | 5056941000006119 | 233885007 | 350431013 |
| CT - Coronary thrombosis | 6601131000006115 | 398274000 | 1786196012 |
| Subsequent myocardial infarction of unspecified site | 118831000006118 | 194856005 | 299808017 |
| Postoperative transmural myocardial infarction other sites | 212081000006112 | 129574000 | 208365015 |
| Postoperative transmural myocardial infarction unspec site | 212091000006110 | 129574000 | 208365015 |
| Post infarction pericarditis | 216351000006118 | 233885007 | 350431013 |
| Acute myocardial infarction of atrium | 4775891000006119 | 194809007 | 2470032018 |
| Defect of ventricular septum following myocardial infarction | 5056521000006115 | 233846000 | 3333538017 |
| Acute myocardial infarction of septum | 3784911000006111 | 79009004 | 131106018 |
| EKG:posterior/inferior infarct | 4586171000006112 | 164869004 | 3300251011 |
| Aborted myocardial infarction | 4775931000006111 | 194821006 | 2619483012 |
| EKG: myocardial infarction | 4586051000006118 | 164865005 | 3300377018 |
| Other acute myocardial infarction NOS | 299720018 | 57054005 | 94884017 |
| History of myocardial infarction in last year | 5974481000006116 | 308065005 | 2986726013 |
| Acute papillary muscle infarction | 1218860015 | 10273003 | 1218860015 |
| Acute myocardial infarction of anterolateral wall | 3641641000006116 | 70211005 | 116613011 |
| Acute infarction of papillary muscle | 2663441000006117 | 10273003 | 17900011 |
| Acute Q wave myocardial infarction | 5935321000006111 | 304914007 | 447323012 |
| Silent myocardial infarction | 350376014 | 233843008 | 350376014 |
| Microinfarction of heart | 1229885017 | 42531007 | 1229885017 |
| Coronary artery thrombosis | 6601121000006118 | 398274000 | 1777817018 |

| | | | |
|---|---|---|---|
| EKG: subendocardial infarct | 4586201000006111 | 164870003 | 3300441012 |
| Coronary thrombosis | 884141000006116 | 57054005 | 884141000006116 |
| ECG: myocardial infarct NOS | 256460011 | 164865005 | 256452010 |
| Acute ST segment elevation myocardial infarction | 1780491019 | 401303003 | 1780491019 |
| Thrombosis - coronary | 1786198013 | 398274000 | 1786198013 |
| Electrocardiogram finding of infarction | 4586031000006113 | 164865005 | 2693159014 |
| MI - Myocardial infarction | 2855341000006114 | 22298006 | 1784872019 |
| True posterior wall infarction | 4775881000006117 | 194802003 | 2535851011 |
| Pericarditis following myocardial infarction | 5056951000006117 | 233885007 | 350432018 |
| Postoperative myocardial infarction | 208365015 | 129574000 | 208365015 |
| MI - Silent myocardial infarction | 5056461000006113 | 233843008 | 350377017 |
| Acute myocardial infarction of septum alone | 3784921000006115 | 79009004 | 131107010 |
| Acute myocardial infarction of posterolateral wall | 967931000006114 | 15990001 | 27071012 |
| Previous myocardial infarction | 6619201000006117 | 399211009 | 545891000000115 |
| Acute myocardial infarction of lateral wall | 3452181000006112 | 58612006 | 97399018 |
| Subsequent NSTEMI (non-ST segment elevation myocardial infarction) | 7574491000006114 | 703360004 | 3008532019 |
| Myocardial infarction with complication | 6348651000006112 | 371068009 | 6348651000006112 |
| Acute lateral myocardial infarction | 299711010 | 58612006 | 1231860015 |
| Hemopericardium due to and following acute myocardial infarction | 12220751000006113 | 194862000 | 3673281015 |
| Infarction of heart | 2855311000006110 | 22298006 | 37441018 |
| Ventricular septal defect as current complication following acute myocardial infarction | 67081000006119 | 233846000 | 350381017 |
| Acute inferior ST segment elevation myocardial infarction | 7572341000006117 | 703213009 | 3007793013 |
| Post-myocardial infarction syndrome | 109915012 | 66189004 | 500341014 |
| Coronary artery embolism | 2981141000006112 | 29899005 | 50041016 |
| AMI - Acute myocardial infarction | 3427201000006111 | 57054005 | 1231678011 |
| Cause of Death- Myocardial Infarction | 1576271000006117 | 1576271000006101 | 1576271000006117 |
| Rupture of chordae tendinae as current complication following acute myocardial infarction | 158611000006118 | 194866002 | 299822017 |
| Postoperative transmural myocardial infarction of inferior wall | 212071000006114 | 311793000 | 455419012 |
| Thrombosis of atrium, auricular appendage, and ventricle due to and following acute myocardial infarction | 12220791000006119 | 194868001 | 3673277015 |
| Acute anterior myocardial infarction | 299707016 | 54329005 | 1231324017 |
| Acute posterior myocardial infarction | 299710011 | 233838001 | 350371016 |
| Rupture of cardiac wall without hemopericardium as current complication following acute myocardial infarction | 4776251000006118 | 194865003 | 299820013 |
| Other acute myocardial infarction | 299718016 | 57054005 | 94884017 |
| Acute myocardial infarction of anterior wall | 3381601000006117 | 54329005 | 90302019 |
| Acute ST segment elevation myocardial infarction of inferior wall | 7572321000006112 | 703213009 | 3007915011 |
| Anterior myocardial infarction NOS | 299709018 | 54329005 | 1231324017 |
| True posterior myocardial infarction | 299712015 | 194802003 | 299712015 |
| Subsequent myocardial infarction of anterior wall | 299811016 | 194857001 | 299811016 |

| | | | |
|---|---|---|---|
| Cardiac infarction | 2855321000006119 | 22298006 | 37442013 |
| Rupture of papillary muscle as current complication following acute myocardial infarction | 159001000006119 | 194867006 | 299823010 |
| Acute inferolateral myocardial infarction | 457531000006110 | 65547006 | 1232697013 |
| Acute STEMI (ST elevation myocardial infarction) of inferior wall | 7572331000006110 | 703213009 | 3007397011 |
| ECG: subendocardial infarct | 256458014 | 164870003 | 256458014 |
| Electrocardiogram: anteroseptal infarction | 4586141000006116 | 164868007 | 2619217019 |
| Acute myocardial infarction of inferoposterior wall | 3745741000006117 | 76593002 | 127193015 |
| Acute STEMI (ST elevation myocardial infarction) of anterior wall | 7571601000006115 | 703164000 | 3007542019 |
| Electrocardiographic posterior/inferior infarct | 4586181000006110 | 164869004 | 3300252016 |
| [RFC] Myocardial infarction (MI) | 905351000006113 | 905351000006109 | 905351000006113 |
| Septal infarction by electrocardiogram | 2515471000006117 | 1077002 | 2972454013 |
| Cardiac rupture after acute myocardial infarction | 537751000006115 | 233847009 | 350383019 |
| Diabetes mellitus insulin-glucose infusion in acute myocardial infarction | 616081000006113 | 315287002 | 459791015 |
| Rupture of chordae tendinae due to and following acute myocardial infarction | 12220781000006117 | 194866002 | 3673278013 |
| Infarct myocardiectomy | 865031000006114 | 585461000000102 | 865031000006114 |
| Acute atrial infarction | 299719012 | 194809007 | 299719012 |
| Subsequent myocardial infarction | 299808017 | 194856005 | 299808017 |
| Acute anteroseptal myocardial infarction | 455651000006114 | 62695002 | 104192010 |
| Heart attack | 37443015 | 22298006 | 37443015 |
| Myocardial infarction with complication | 543291000006110 | 371068009 | 1209776017 |
| Acute anterior ST segment elevation myocardial infarction | 7571581000006113 | 703164000 | 3007817016 |
| ECG: anteroseptal infarction | 4586131000006114 | 164868007 | 256456013 |
| [X]Acute transmural myocardial infarction of unspecif site | 362461000006119 | 57054005 | 94884017 |
| First myocardial infarction | 932081000006118 | 932081000006102 | 932081000006118 |
| ECG:posterior/inferior infarct | 256457016 | 164869004 | 256457016 |
| STEMI - ST elevation myocardial infarction | 6651221000006117 | 401303003 | 2840840013 |
| Acute nontransmural infarction | 3645351000006114 | 70422006 | 116993010 |
| Postoperative subendocardial myocardial infarction | 455422014 | 311796008 | 455422014 |
| Acute anterolateral myocardial infarction | 455641000006112 | 70211005 | 1233238016 |
| Atrial septal defect as current complication following acute myocardial infarction | 498031000006112 | 194863005 | 299818010 |
| [X]Subsequent myocardial infarction of unspecified site | 300882013 | 194856005 | 299808017 |
| Reinfarction of myocardium | 4776171000006116 | 194856005 | 299809013 |
| Postoperative myocardial infarction, unspecified | 455423016 | 129574000 | 208365015 |
| Acute myocardial infarction of diaphragmatic wall | 3699911000006119 | 73795002 | 122559017 |
| Dressler's syndrome | 109916013 | 66189004 | 109916013 |
| Post-infarction ventricular septal defect | 5056501000006113 | 233846000 | 350382012 |
| Anteroseptal infarction on electrocardiogram | 4586111000006115 | 164868007 | 2695135018 |
| Acute subendocardial infarction | 116992017 | 70422006 | 116992017 |
| ECG: lateral infarction | 256459018 | 164871004 | 256459018 |

| | | | |
|---|---|---|---|
| Acute myocardial infarction of inferior wall | 299714019 | 73795002 | 122557015 |
| Acute myocardial infarction NOS | 299721019 | 57054005 | 94884017 |
| Subsequent myocardial infarction of other sites | 299813018 | 194856005 | 299808017 |
| Attack - heart | 219521000000119 | 22298006 | 37443015 |
| Cardiac rupture due to and following acute myocardial infarction | 12221301000006119 | 233847009 | 3673284011 |
| NSTEMI - Non-ST segment elevation MI | 6651391000006114 | 401314000 | 1787486017 |
| Hemopericardium as current complication following acute myocardial infarction | 4776221000006110 | 194862000 | 299817017 |
| ECG: antero-septal infarct | 4586151000006119 | 164868007 | 2820892012 |
| Acute septal infarction | 1234306015 | 79009004 | 1234306015 |
| Acute non-ST segment elevation myocardial infarction | 1780501013 | 401314000 | 1780501013 |
| Rupture of cardiac wall without haemopericardium as current complication following acute myocardial infarction | 158601000006116 | 194865003 | 299821012 |
| Postoperative transmural myocardial infarction of anterior wall | 212061000006119 | 311792005 | 455418016 |
| Atrial septal defect due to and following acute myocardial infarction | 12220771000006115 | 194863005 | 3673273016 |
| First myocardial infarction | 6546751000006118 | 394710008 | 1488431012 |
| Anterolateral infarction by EKG | 3202711000006110 | 43630006 | 72749019 |
| Acute anteroapical myocardial infarction | 3343471000006116 | 52035003 | 86618010 |
| Non-Q wave myocardial infarction | 6043771000006118 | 314207007 | 458526013 |
| MI - Myocardial infarction aborted | 2619484018 | 194821006 | 2619484018 |
| Haemopericardium as current complication following acute myocardial infarction | 813961000006116 | 194862000 | 299816014 |
| [X]Subsequent myocardial infarction of other sites | 300881018 | 194856005 | 299808017 |
| Acute transmural myocardial infarction | 2729671000000118 | 1089471000000109 | 2729671000000118 |
| Postmyocardial infarction pericarditis | 3576391000006116 | 66189004 | 109917016 |
| Cause of Death- Acute Myocardial Infarction | 1576301000006115 | 1576301000006104 | 1576301000006115 |
| Haemopericardium due to and following acute myocardial infarction | 12220761000006110 | 194862000 | 3673282010 |
| Acute inferoposterior infarction | 1234005010 | 76593002 | 1234005010 |
| Coronary thrombosis | 1786197015 | 398274000 | 1786197015 |
| ECG: myocardial infarction | 256452010 | 164865005 | 256452010 |
| Thrombosis of atrium, auricular appendage, and ventricle as current complications following acute myocardial infarction | 100681000006116 | 194868001 | 299824016 |
| Myocardial Infarction | 884151000006119 | 57054005 | 884151000006119 |
| Myocardial infarction aborted | 682481000006118 | 194821006 | 2619484018 |
| MI - acute myocardial infarction | 219531000000117 | 57054005 | 94884017 |
| Acute myocardial infarction | 94884017 | 57054005 | 94884017 |
| Myocardial infarction | 2855301000006112 | 22298006 | 37436014 |
| Microinfarct of heart | 3185771000006112 | 42531007 | 70952012 |
| [X]Other current complicatns following acute myocard infarct | 408571000006116 | 302049001 | 443587013 |

#### Table S33. Myocardial infarction HES (ICD10) codes

| ICD10 codes | Term |
|---|---|
| I21.0 | Acute transmural myocardial infarction of anterior wall |
| I21.1 | Acute transmural myocardial infarction of inferior wall |
| I21.2 | Acute transmural myocardial infarction of other sites |
| I21.3 | Acute transmural myocardial infarction of unspecified site |
| I21.4 | Acute subendocardial myocardial infarction |
| I21.9 | Acute myocardial infarction, unspecified |
| I22.0 | Subsequent myocardial infarction of anterior wall |
| I22.1 | Subsequent myocardial infarction of inferior wall |
| I22.8 | Subsequent myocardial infarction of other sites |
| I22.9 | Subsequent myocardial infarction of unspecified site |
| I23.0 | Haemopericardium as current complication following acute myocardial infarction |
| I23.1 | Atrial septal defect as current complication following acute myocardial infarction |
| I23.2 | Ventricular septal defect as current complication following acute myocardial infarction |
| I23.3 | Rupture of cardiac wall without haemopericardium as current complication following acute myocardial infarction |
| I23.4 | Rupture of chordae tendineae as current complication following acute myocardial infarction |
| I23.5 | Rupture of papillary muscle as current complication following acute myocardial infarction |
| I23.6 | Thrombosis of atrium, auricular appendage, and ventricle as current complications following acute myocardial infarction |
| I23.8 | Other current complications following acute myocardial infarction |
| I24.1 | Dressler syndrome |

#### Table S34. Vascular dementia Aurum codes

| Term | Medcode ID | SNOMED CT Concept ID | SNOMED CT Description ID |
|---|---|---|---|
| [X]Vascular dementia, unspecified | 295681016 | 429998004 | 2770951017 |
| Vascular dementia of acute onset, other symptoms, predominantly hallucinatory | 1972521000006114 | 1972521000006105 | 1972521000006114 |
| Subcortical vascular dementia, other symptoms, predominantly depressive | 1972771000006116 | 1972771000006100 | 1972771000006116 |
| Mixed cortical and subcortical vascular dementia, other symptoms, predominantly hallucinatory | 1972871000006113 | 1972871000006109 | 1972871000006113 |
| Mixed cortical and subcortical vascular dementia, other symptoms, predominantly depressive | 1972911000006111 | 1972911000006107 | 1972911000006111 |
| VAD - Vascular dementia | 3414251000006112 | 56267009 | 497560014 |
| Subcortical leukoencephalopathy | 3964601000006113 | 90099008 | 149344015 |
| Binswanger's dementia | 3964611000006111 | 90099008 | 149345019 |
| Binswanger's encephalopathy | 1235534016 | 90099008 | 1235534016 |
| Binswanger's disease | 149347010 | 90099008 | 149347010 |
| MID - Multi-infarct dementia | 3414231000006117 | 56267009 | 497558012 |
| Arteriosclerotic dementia NOS | 294656010 | 56267009 | 497559016 |
| Subcortical leucoencephalopathy | 3964591000006117 | 90099008 | 508841015 |
| Uncomplicated arteriosclerotic dementia | 294652012 | 191463004 | 294652012 |
| Arteriosclerotic dementia with delirium | 294653019 | 191464005 | 294653019 |
| Arteriosclerotic dementia with paranoia | 294654013 | 191465006 | 294654013 |
| Arteriosclerotic dementia with depression | 294655014 | 191466007 | 294655014 |
| Ischaemic vascular dementia | 7840631000006118 | 723123001 | 3334662012 |
| Vascular dementia of acute onset, other symptoms, predominantly delusional | 1972501000006116 | 1972501000006100 | 1972501000006116 |
| Other vascular dementia, other symptoms, predominantly depressive | 1973381000006112 | 1973381000006108 | 1973381000006112 |
| Vascular dementia, with delirium | 2664611000006117 | 10349009 | 18021011 |
| [X]Vascular dementia | 431681000006117 | 429998004 | 2770951017 |
| [X]Vascular dementia of acute onset | 431691000006119 | 230285003 | 345110014 |
| Multi infarct dementia | 3414261000006114 | 56267009 | 2921000019 |
| [D] Vascular dementia | 914921000006117 | 914921000006101 | 914921000006117 |
| Vascular dementia of acute onset, without additional symptoms | 1972481000006114 | 1972481000006105 | 1972481000006114 |
| Multi-infarct dementia, other symptoms, predominantly delusional | 1972621000006113 | 1972621000006109 | 1972621000006113 |
| Multi-infarct dementia, other symptoms, predominantly depressive | 1972661000006119 | 1972661000006103 | 1972661000006119 |
| Subcortical vascular dementia, without additional symptoms | 1972711000006113 | 1972711000006109 | 1972711000006113 |
| Subcortical vascular dementia, other symptoms, predominantly delusional | 1972731000006119 | 1972731000006103 | 1972731000006119 |
| Other vascular dementia, without additional symptoms | 1973221000006118 | 1973221000006102 | 1973221000006118 |
| Other vascular dementia, other symptoms, predominantly delusional | 1973271000006117 | 1973271000006101 | 1973271000006117 |
| Other vascular dementia, other mixed symptoms | 1973401000006112 | 1973401000006108 | 1973401000006112 |
| Multi infarct dementia | 696161000006115 | 56267009 | 93568017 |
| Vascular dementia of acute onset, other symptoms, predominantly depressive | 1972541000006119 | 1972541000006103 | 1972541000006119 |

| | | | |
|---|---|---|---|
| Multi-infarct dementia, without additional symptoms | 1972601000006115 | 1972601000006104 | 1972601000006115 |
| Subcortical vascular dementia, other mixed symptoms | 1972791000006115 | 1972791000006104 | 1972791000006115 |
| Mixed cortical and subcortical vascular dementia, without additional symptoms | 1972821000006112 | 1972821000006108 | 1972821000006112 |
| Other vascular dementia, other symptoms, predominantly hallucinatory | 1973341000006118 | 1973341000006102 | 1973341000006118 |
| Vascular dementia, unspecified, without additional symptoms | 1973461000006113 | 1973461000006109 | 1973461000006113 |
| Vascular dementia, unspecified, other symptoms, predominantly hallucinatory | 1973531000006117 | 1973531000006101 | 1973531000006117 |
| [X]Other vascular dementia | 295680015 | 429998004 | 2770951017 |
| [X]Arteriosclerotic dementia | 363791000006112 | 56267009 | 497559016 |
| Multi-infarct dementia, other symptoms, predominantly hallucinatory | 1972641000006118 | 1972641000006102 | 1972641000006118 |
| Multi-infarct dementia, other mixed symptoms | 1972681000006112 | 1972681000006108 | 1972681000006112 |
| Vascular dementia, unspecified, other symptoms, predominantly depressive | 1973551000006112 | 1973551000006108 | 1973551000006112 |
| Vascular dementia, unspecified, other mixed symptoms | 1976831000006111 | 1976831000006107 | 1976831000006111 |
| Arteriosclerotic dementia | 497559016 | 56267009 | 497559016 |
| Vascular dementia of acute onset, other mixed symptoms | 1972571000006110 | 1972571000006106 | 1972571000006110 |
| Subcortical vascular dementia, other symptoms, predominantly hallucinatory | 1972751000006114 | 1972751000006105 | 1972751000006114 |
| Mixed cortical and subcortical vascular dementia, other symptoms, predominantly delusional | 1972831000006110 | 1972831000006106 | 1972831000006110 |
| Mixed cortical and subcortical vascular dementia, other mixed symptoms | 1972931000006117 | 1972931000006101 | 1972931000006117 |
| Vascular dementia, unspecified, other symptoms, predominantly delusional | 1973501000006113 | 1973501000006109 | 1973501000006113 |
| [X]Mixed cortical and subcortical vascular dementia | 398571000006112 | 230287006 | 345112018 |
| [X]Subcortical vascular dementia | 428201000006119 | 230286002 | 345111013 |
| [X]Multi-infarct dementia | 399031000006111 | 56267009 | 93568017 |
| Subcortical arteriosclerotic encephalopathy | 3964651000006112 | 90099008 | 1235535015 |
| Vascular dementia with behavioral disturbance | 8089241000006117 | 288631000119104 | 3290002019 |

#### Table S35. Vascular dementia HES (ICD10) codes

| ICD10 codes | Term |
|---|---|
| F01.0 | Vascular dementia of acute onset |
| F01.1 | Multi-infarct dementia |
| F01.2 | Subcortical vascular dementia |
| F01.3 | Mixed cortical and subcortical vascular dementia |
| F01.8 | Other vascular dementia |
| F01.9 | Vascular dementia, unspecified |

#### Table S36. Gastrointestinal bleeding Aurum codes

| Term | Medcode ID | SNOMED CT Concept ID | SNOMED CT Description ID |
|---|---|---|---|
| Gastric ulcer with hemorrhage AND perforation | 3513611000006117 | 62366003 | 103655018 |
| Haemorrhage of large intestine co-occurrent and due to diverticular disease of large intestine | 11823161000006110 | 197092000 | 3512394014 |
| Bleeding duodenal ulcer | 2937141000006110 | 27281001 | 1225269012 |
| DU - acute + h'ge + perf. | 885871000006115 | 86895006 | 885871000006115 |
| GU - acute + haemorrhage | 885771000006112 | 89748001 | 885771000006112 |
| Bleeding diverticulosis of large intestine | 11823171000006115 | 197092000 | 3512393015 |
| Bleeding chronic duodenal ulcer | 1235462014 | 89469000 | 1235462014 |
| DU - acute + haemorrhage | 885851000006113 | 12847006 | 885851000006113 |
| Haemorrhagic duodenitis | 4058111000006114 | 95531001 | 512171013 |
| Bleeding external hemorrhoids | 2923111000006117 | 26421009 | 44253010 |
| Bleeding internal hemorrhoids | 3733941000006117 | 75884004 | 126038015 |
| Vomiting blood - fresh | 397912015 | 267051003 | 397912015 |
| Chronic peptic ulcer with hemorrhage | 3296621000006114 | 49232000 | 82009014 |
| RB - Rectal bleeding | 2692031000006113 | 12063002 | 464498014 |
| GI bleeding | 3710501000006113 | 74474003 | 123689014 |
| Rectal haemorrhage | 464500010 | 12063002 | 464500010 |
| Gastric haemorrhage | 303660017 | 61401005 | 499003013 |
| Acute peptic ulcer with haemorrhage and perforation | 361430016 | 111353003 | 361430016 |
| Lower GI bleeding | 3926401000006112 | 87763006 | 145512011 |
| Vomiting blood - coffee ground | 492804019 | 40835002 | 492804019 |
| Haemorrhage of large intestine with diverticular disease of large intestine | 11823191000006119 | 197092000 | 3512395010 |
| PU - chronic + h'ge + perf. | 886001000006111 | 61300005 | 886001000006111 |
| Chronic peptic ulcer with hemorrhage AND perforation | 3496021000006116 | 61300005 | 101859019 |
| GI - Gastrointestinal hemorrhage | 3710571000006119 | 74474003 | 1218401013 |
| Chronic gastrojejunal ulcer with hemorrhage AND perforation | 3236981000006112 | 45640006 | 76108010 |
| Upper GI - gastrointestinal haemorrhage | 3102201000006117 | 37372002 | 1216567017 |
| Hemorrhage of large intestine co-occurrent and due to diverticular disease of large intestine | 11823151000006113 | 197092000 | 3512397019 |
| DU - chronic + h'ge + perf. | 885911000006117 | 36975000 | 885911000006117 |
| Bleeding gastric ulcer | 2752321000006114 | 15902003 | 1221244018 |
| Rectorrhagia | 2691961000006110 | 12063002 | 20789018 |
| Chronic gastric ulcer with hemorrhage AND with perforation | 3739191000006114 | 76181002 | 126521010 |
| Chronic gastric ulcer with hemorrhage | 3430281000006116 | 57246001 | 95214011 |
| Haematemesis | 507679012 | 8765009 | 507679012 |
| Duodenal ulcer with haemorrhage | 302534015 | 27281001 | 483490017 |
| Oesophageal varices with bleeding in diseases EC | 300776018 | 195475003 | 300776018 |
| Acute upper GI bleeding | 3126711000006119 | 38938002 | 65309016 |
| Unspecified peptic ulcer with haemorrhage and perforation | 302571018 | 13200003 | 22592012 |

| | | | |
|---|---|---|---|
| Unspecified peptic ulcer with haemorrhage | 302568014 | 13200003 | 22592012 |
| Hematoma of perianal region | 7375281000006110 | 449815008 | 2913034013 |
| Rectal bleeding | 20792019 | 12063002 | 20792019 |
| Chronic peptic ulcer with haemorrhage | 495344012 | 49232000 | 495344012 |
| Melaena - O/E of faeces | 411734011 | 275782008 | 411734011 |
| Upper GI hemorrhage | 3102171000006116 | 37372002 | 62347019 |
| Bleeding acute gastric ulcer | 1235493010 | 89748001 | 1235493010 |
| Haemorrhage of rectum and anus | 396382012 | 266464001 | 396382012 |
| Melena | 2545151000006112 | 2901004 | 5911019 |
| GU - chronic + h'ge + perf. | 885831000006118 | 76181002 | 885831000006118 |
| GU - chronic + haemorrhage | 885811000006112 | 57246001 | 885811000006112 |
| History of lower GIT haemorrhage | 1819591000006117 | 1819591000006101 | 1819591000006117 |
| Gastrooesophageal laceration-hemorrhage syndrome | 3068381000006113 | 35265002 | 2646918012 |
| PR - Bleeding per rectum | 2692001000006117 | 12063002 | 464495012 |
| Upper GI bleeding | 3102181000006118 | 37372002 | 62348012 |
| Acute gastric ulcer with hemorrhage | 3958881000006116 | 89748001 | 148789010 |
| Duodenal ulcer with hemorrhage | 11902881000006118 | 27281001 | 45608011 |
| Painless rectal bleeding | 2534250012 | 414992000 | 2534250012 |
| Acute peptic ulcer with haemorrhage | 470365013 | 12274003 | 470365013 |
| Rectal hemorrhage | 2692051000006118 | 12063002 | 464501014 |
| Hematemesis | 2639821000006115 | 8765009 | 15458019 |
| Acute peptic ulcer with hemorrhage and perforation | 4194481000006117 | 111353003 | 90139013 |
| Rectal packing for hemorrhage | 4716091000006113 | 182606003 | 282271018 |
| Chronic gastric ulcer with haemorrhage AND with perforation | 554311000006114 | 76181002 | 503219010 |
| GI haemorrhage | 3710531000006117 | 74474003 | 502714014 |
| Haemorrhage of rectum and anus NOS | 303317017 | 266464001 | 396382012 |
| GI hemorrhage | 3710511000006111 | 74474003 | 123690017 |
| Evacuation of perianal hematoma | 5789981000006110 | 293018003 | 433185011 |
| Bleeding hemorrhoid | 3335301000006118 | 51551000 | 1218192016 |
| Melaena on examination of faeces | 5576661000006114 | 275782008 | 2771057011 |
| Chronic gastrojejunal ulcer with haemorrhage AND perforation | 494304013 | 45640006 | 494304013 |
| Chronic gastric ulcer with haemorrhage | 497833010 | 57246001 | 497833010 |
| Chronic peptic ulcer with haemorrhage AND perforation | 498977018 | 61300005 | 498977018 |
| Chronic gastrojejunal ulcer with haemorrhage | 499386010 | 62838000 | 499386010 |
| Oesophageal varices with haemorrhage | 2781041000006115 | 17709002 | 478406011 |
| Gastric hemorrhage | 3497601000006119 | 61401005 | 102007012 |
| Oesophageal haemorrhage | 2741641000006118 | 15238002 | 477249012 |
| Acute duodenal ulcer with hemorrhage AND perforation | 3912771000006119 | 86895006 | 144111018 |
| Acute duodenal ulcer with hemorrhage | 2704011000006118 | 12847006 | 22034017 |
| Haematoma of perianal region | 7375271000006112 | 449815008 | 2912591019 |

| | | | |
|---|---|---|---|
| Duodenal ulcer with haemorrhage | 2937121000006115 | 27281001 | 2937121000006115 |
| Perianal haematoma | 233701000006116 | 449815008 | 2213971000000113 |
| Hemorrhage of rectum and anus | 5493021000006119 | 266464001 | 396383019 |
| Unspec gastrojejunal ulcer with haemorrhage and perforation | 81321000006111 | 16121001 | 27283019 |
| Oesophageal bleeding | 2741601000006115 | 15238002 | 477250012 |
| Bleeding from anus | 481141000006114 | 6072007 | 498827015 |
| Internal bleeding haemorrhoids | 1216930015 | 75884004 | 1216930015 |
| Unspecified gastrojejunal ulcer with haemorrhage | 302606019 | 16121001 | 27283019 |
| Unspecified gastric ulcer with haemorrhage and perforation | 302490012 | 62366003 | 499256017 |
| Acute gastric ulcer with bleeding | 3958891000006118 | 89748001 | 148790018 |
| Melena on examination of feces | 5576651000006112 | 275782008 | 2770255015 |
| Melaena | 5913016 | 2901004 | 5913016 |
| Acute gastrojejunal ulcer with haemorrhage | 499719010 | 63954007 | 499719010 |
| Melena - O/E of feces | 5576641000006110 | 275782008 | 411735012 |
| Chronic gastrojejunal ulcer with hemorrhage | 3521501000006114 | 62838000 | 104433010 |
| Gastric bleeding | 3497621000006112 | 61401005 | 102011018 |
| Anal/rectal haemorrhage | 886471000006110 | 266464001 | 886471000006110 |
| PU - acute + haemorrhage | 885941000006118 | 12274003 | 885941000006118 |
| Gastric ulcer with hemorrhage | 2752311000006118 | 15902003 | 26937011 |
| Internal bleeding hemorrhoids | 3733961000006118 | 75884004 | 1218417019 |
| Upper GI - gastrointestinal hemorrhage | 3102211000006119 | 37372002 | 1218058016 |
| Bleeding pile | 3335311000006115 | 51551000 | 1230996010 |
| Unspecified duodenal ulcer with haemorrhage and perforation | 302537010 | 51868009 | 86362013 |
| GI - Gastrointestinal haemorrhage | 3710541000006110 | 74474003 | 1216914011 |
| Bleeding hemorrhoids | 3335281000006117 | 51551000 | 85856015 |
| Bleeding per rectum | 515491000006115 | 12063002 | 464494011 |
| Anal margin haematoma | 5585681000006116 | 276465001 | 412609016 |
| Peptic ulcer with haemorrhage | 3542811000006116 | 64121000 | 499762011 |
| Esophageal varices with hemorrhage | 2781031000006113 | 17709002 | 29938017 |
| Chronic duodenal ulcer with hemorrhage AND perforation | 3095701000006110 | 36975000 | 61676011 |
| Acute duodenal ulcer with haemorrhage | 474430017 | 12847006 | 474430017 |
| Acute haemorrhagic gastritis | 478553011 | 2367005 | 481682019 |
| Chronic duodenal ulcer with haemorrhage | 508524014 | 89469000 | 508524014 |
| Esophageal bleeding | 2741651000006116 | 15238002 | 477251011 |
| PU - acute + h'ge + perf. | 885961000006119 | 111353003 | 885961000006119 |
| Upper GI haemorrhage | 3102191000006115 | 37372002 | 486703016 |
| H/O: upper GIT hemorrhage | 6017191000006116 | 312489007 | 456219016 |
| PU - acute + perforation | 885951000006116 | 79118000 | 885951000006116 |
| Proctorrhagia | 2691971000006115 | 12063002 | 20790010 |
| Gastroesophageal laceration-hemorrhage syndrome | 3068351000006117 | 35265002 | 58844012 |

| | | | |
|---|---|---|---|
| Unspec peptic ulcer; unspec haemorrhage and/or perforation | 81501000006111 | 13200003 | 22592012 |
| Unspec gastrojejunal ulcer; unspec haemorrhage/perforation | 81331000006114 | 16121001 | 27283019 |
| Lower GI hemorrhage | 3926391000006110 | 87763006 | 145511016 |
| Lower GI haemorrhage | 3926411000006110 | 87763006 | 507737011 |
| Perianal haematoma | 483207010 | 237331002 | 355700019 |
| Bleeding internal haemorrhoids | 3733931000006110 | 75884004 | 200553019 |
| Acute duodenal ulcer with haemorrhage AND perforation | 507303017 | 86895006 | 507303017 |
| Gastric hemorrhage due to erosive gastritis | 7987031000006110 | 7071000119102 | 3015365015 |
| Chronic duodenal ulcer with hemorrhage | 3954241000006119 | 89469000 | 148344012 |
| Haemorrhage of oesophagus | 477248016 | 15238002 | 477248016 |
| Gastrotomy and ligation of bleeding point of stomach | 269184010 | 173778009 | 269184010 |
| Chronic duodenal ulcer with haemorrhage AND perforation | 486596015 | 36975000 | 486596015 |
| Acute peptic ulcer with hemorrhage | 2695521000006113 | 12274003 | 21136016 |
| GU - acute + perforation | 885781000006110 | 19850005 | 885781000006110 |
| Rectal packing for haemorrhage | 4716081000006110 | 182606003 | 282269018 |
| BOV - Bleeding oesophageal varices | 2781061000006116 | 17709002 | 1216061014 |
| Perianal haematoma | 1757561000000116 | 449815008 | 2213971000000113 |
| Bleeding external haemorrhoids | 2923101000006115 | 26421009 | 196528011 |
| Esophageal hemorrhage | 2741621000006113 | 15238002 | 25860014 |
| Bleeding esophageal varices | 2781021000006110 | 17709002 | 29937010 |
| Unspec duodenal ulcer; unspec haemorrhage and/or perforation | 81301000006118 | 51868009 | 86362013 |
| Acute gastrojejunal ulcer with haemorrhage AND perforation | 504674011 | 81387001 | 504674011 |
| H/O: upper GIT haemorrhage | 456217019 | 312489007 | 456217019 |
| Acute gastric ulcer with hemorrhage AND perforation | 3292411000006119 | 48974009 | 81590012 |
| PRB - Rectal bleeding | 464499018 | 12063002 | 464499018 |
| Gastric ulcer with haemorrhage AND perforation | 81311000006115 | 62366003 | 499256017 |
| Bleeding oesophageal varices | 2781011000006119 | 17709002 | 478405010 |
| Acute hemorrhagic gastritis | 2536601000006112 | 2367005 | 5056011 |
| Esophageal varices with bleeding, associated with another disorder | 4779741000006118 | 195475003 | 2575714018 |
| Gastric ulcer with haemorrhage | 302487018 | 15902003 | 477573015 |
| Bloody diarrhoea | 1566381000006119 | 95545007 | 201773015 |
| Perinatal gastrointestinal haemorrhage | 495197011 | 48729005 | 495197011 |
| Acute gastrojejunal ulcer with hemorrhage | 3539941000006116 | 63954007 | 106302011 |
| Bleeding diverticulosis | 303134017 | 197092000 | 303134017 |
| Hemorrhage of large intestine with diverticular disease of large intestine | 11823181000006117 | 197092000 | 3512398012 |
| Acute gastric ulcer with haemorrhage | 508664015 | 89748001 | 508664015 |
| Bleeding ulcer | 3393951000006114 | 55075001 | 91564012 |
| Bleeding chronic gastric ulcer | 1231704018 | 57246001 | 1231704018 |
| Bleeding oesophageal ulcer | 3438321000006112 | 57748001 | 3027281014 |

| | | | |
|---|---|---|---|
| Vomiting of blood | 2639831000006117 | 8765009 | 15459010 |
| External bleeding hemorrhoids | 2923131000006111 | 26421009 | 1217729010 |
| Bleeding haemorrhoid | 3335291000006119 | 51551000 | 1216702010 |
| GU - acute + h'ge + perf. | 885791000006113 | 48974009 | 885791000006113 |
| DU - acute + perforation | 885861000006110 | 61347001 | 885861000006110 |
| PU - chronic + haemorrhage | 885981000006112 | 49232000 | 885981000006112 |
| DU - chronic + haemorrhage | 885891000006119 | 89469000 | 885891000006119 |
| PR - Blood per rectum | 2692021000006110 | 12063002 | 464497016 |
| Vomiting blood | 15459010 | 8765009 | 507678016 |
| Blood per rectum | 2692011000006119 | 12063002 | 464496013 |
| Painful rectal bleeding | 2534219010 | 414991007 | 2534219010 |
| BOV - Bleeding esophageal varices | 2781081000006114 | 17709002 | 1217568017 |
| Manchester triage - GI bleeding | 1984161000006112 | 1984161000006108 | 1984161000006112 |
| Oesophageal varices with bleeding, associated with another disorder | 4779721000006113 | 195475003 | 2575715017 |
| Esophageal varices with bleeding | 2781071000006111 | 17709002 | 1217567010 |
| Acute gastrojejunal ulcer with hemorrhage AND perforation | 3823411000006113 | 81387001 | 135007015 |
| Perianal hematoma | 1988561000006114 | 449815008 | 2213971000000113 |
| Oesophageal varices with bleeding | 1216060010 | 17709002 | 1216060010 |
| Acute gastric ulcer with haemorrhage AND perforation | 495273013 | 48974009 | 495273013 |
| Intestinal haemorrhage NOS | 303663015 | 74474003 | 123688018 |
| Hemorrhage of esophagus | 2741611000006117 | 15238002 | 25859016 |
| Gastro-oesophageal laceration-haemorrhage syndrome | 3068361000006115 | 35265002 | 1216440014 |
| Bleeding haemorrhoids | 300752014 | 51551000 | 198339016 |
| Acute lower GI bleeding | 3890061000006117 | 85521005 | 141749014 |

#### Table S37. Gastrointestinal bleeding HES (ICD10) codes

| ICD10 codes | Term |
|---|---|
| I85.0 | Oesophageal varices with bleeding |
| I98.3 | Oesophageal varices with bleeding in diseases classified elsewhere |
| K22.6 | Gastro-oesophageal laceration-haemorrhage syndrome |
| K25.0 | Gastric ulcer. Acute with haemorrhage |
| K25.2 | Gastric ulcer. Acute with both haemorrhage and perforation |
| K25.4 | Gastric ulcer. Chronic or unspecified with haemorrhage |
| K25.6 | Gastric ulcer. Chronic or unspecified with both haemorrhage and perforation |
| K26.0 | Duodenal ulcer. Acute with haemorrhage |
| K26.2 | Duodenal ulcer. Acute with both haemorrhage and perforation |
| K26.4 | Duodenal ulcer. Chronic or unspecified with haemorrhage |
| K26.6 | Duodenal ulcer. Chronic or unspecified with both haemorrhage and perforation |
| K27.0 | Peptic ulcer, site unspecified. Acute with haemorrhage |
| K27.2 | Peptic ulcer, site unspecified. Acute with both haemorrhage and perforation |
| K27.4 | Peptic ulcer, site unspecified. Chronic or unspecified with haemorrhage |
| K27.6 | Peptic ulcer, site unspecified. Chronic or unspecified with both haemorrhage and perforation |
| K28.0 | Gastrojejunal ulcer. Acute with haemorrhage |
| K28.2 | Gastrojejunal ulcer. Acute with both haemorrhage and perforation |
| K28.4 | Gastrojejunal ulcer. Chronic or unspecified with haemorrhage |
| K28.6 | Gastrojejunal ulcer. Chronic or unspecified with both haemorrhage and perforation |
| K29.0 | Acute haemorrhagic gastritis |
| K62.5 | Haemorrhage of anus and rectum |
| K66.1 | Haemoperitoneum |
| K92.0 | Haematemesis |
| K92.1 | Melaena |
| K92.2 | Gastrointestinal haemorrhage, unspecified |

#### Table S38. Bleeding at other anatomy sites Aurum codes

| Term | Medcode ID | SNOMED CT Concept ID | SNOMED CT Description ID |
|---|---|---|---|
| Bleeding from nose | 5270631000006113 | 249366005 | 5270631000006113 |
| O/E - retinal microaneurysms | 255335015 | 163986001 | 255335015 |
| Nontraumatic scrotal hematoma | 4790941000006116 | 198058000 | 3036672010 |
| Bleeding into joint | 3830271000006111 | 81808003 | 1234640010 |
| Drainage of haematoma of pinna and insertion of bolster sutures | 4650061000006119 | 172619007 | 267229016 |
| Control of hemorrhage | 3330351000006114 | 51241000 | 85365019 |
| Haemarthrosis of PIP joint of finger | 310710010 | 202409003 | 310710010 |
| Haemarthrosis of wrist | 310705011 | 202407001 | 310705011 |
| Haemarthrosis of the ankle | 310724014 | 202415003 | 310724014 |
| Primary post tonsillectomy hemorrhage | 6046521000006116 | 314483007 | 458849015 |
| Injury of heart with haemopericardium | 320982018 | 210078001 | 320982018 |
| Subungual haemorrhage | 5125561000006112 | 238722001 | 357791012 |
| Postcoital bleeding | 81454015 | 48880000 | 81454015 |
| Pelvic haematoma | 5477481000006114 | 264558008 | 393037010 |
| Corneal haemorrhage | 5029211000006119 | 231916005 | 347513015 |
| Bleeding stoma | 353962019 | 236129005 | 353962019 |
| Acute posthaemorrhagic anaemia | 399228011 | 267530009 | 399228011 |
| Vitreous hemorrhage | 3005171000006115 | 31341008 | 52406015 |
| Hematoma of pinna | 3930851000006114 | 88050005 | 145963019 |
| Haematoma of lower leg | 7373861000006119 | 449704006 | 2912608014 |
| Bleeding in mouth | 2858361000006116 | 22490002 | 481357012 |
| Haematoma of tongue | 5455391000006110 | 262651006 | 390627015 |
| Extradural haematoma | 7085881000006119 | 428268007 | 2899390010 |
| Retinal blot haemorrhage | 5239831000006118 | 247133006 | 2619788016 |
| Haematoma of penis | 5681941000006113 | 283948004 | 422520011 |
| Perioperative haemorrhage | 3636041019 | 762957002 | 3636041019 |
| Persistent haematuria | 5656871000006111 | 281859000 | 419988013 |
| Corpus cavernosum haematoma | 304493010 | 198031007 | 304493010 |
| Thyroid hemorrhage and infarction | 4757961000006112 | 190305000 | 292442019 |
| Subretinal haemorrhage | 475607010 | 13937002 | 475607010 |
| Corpus cavernosum haemorrhage | 1216873013 | 69452005 | 1216873013 |
| Scrotal haemorrhage | 1216798017 | 62182006 | 1216798017 |
| Bleeding tooth socket | 5700021000006118 | 285335006 | 424307013 |
| Retinal hemorrhages | 2966421000006119 | 28998008 | 1217788016 |
| Massive haemoptysis | 2229721000000116 | 862231000000103 | 2229721000000116 |
| Head injury with haemorrhage from nose | 4395871000006112 | 127276009 | 474087019 |
| Spontaneous intraperitoneal hemorrhage | 3236641000006110 | 45626005 | 76080019 |
| Deformity/haematoma of pinna | 883651000006111 | 286943000 | 883651000006111 |
| Nontraumatic haemoperitoneum | 3236611000006111 | 45626005 | 2816649012 |

| | | | |
|---|---|---|---|
| Reopening of chest and re-exploration of intra-abdominal operation site and surgical arrest of postoperative bleeding | 4679481000006110 | 177768005 | 275176015 |
| Nipple bleeding | 5755901000006111 | 290103001 | 430097013 |
| Contact bleeding | 5103751000006111 | 237133008 | 355447012 |
| Haematoma of thigh | 5967731000006111 | 307391000 | 450646011 |
| Moderate vaginal bleeding | 5748541000006110 | 289539007 | 429439016 |
| Extradural hemorrhage | 3850511000006116 | 82999001 | 137670017 |
| On examination - epistaxis | 4576361000006110 | 164187003 | 4576361000006110 |
| Intraperitoneal haematoma | 5088411000006117 | 236000006 | 353764011 |
| External capsule haemorrhage | 300276019 | 195167002 | 300276019 |
| Haemophthalmos, except current injury | 4771081000006111 | 193287005 | 2550868017 |
| Haemorrhage from pharynx | 6119831000006110 | 324618004 | 465452016 |
| Liver hematoma | 5457191000006118 | 262796008 | 390818018 |
| Subungual haematoma, hand | 5561121000006110 | 274202000 | 409975019 |
| Expulsive haemorrhage | 2684991000006115 | 11623000 | 2474957017 |
| Hemopneumothorax | 2763811000006111 | 16632002 | 28149019 |
| History of abnormal uterine bleeding | 5497401000006114 | 267016006 | 2986860010 |
| History of epistaxis | 1137551000000110 | 511511000000101 | 1137551000000110 |
| Hematoma of spleen | 5457481000006111 | 262818004 | 390852015 |
| Non-traumatic testicular haematoma | 3074001000006117 | 35644004 | 1216444017 |
| Deep intra-retinal hemorrhages | 3288931000006115 | 48742005 | 1490493013 |
| Wrist haemarthrosis | 310706012 | 202407001 | 310706012 |
| Elbow haemarthrosis | 310700018 | 202405009 | 310700018 |
| Spontaneous intraperitoneal haemorrhage | 3236701000006118 | 45626005 | 494285013 |
| Traumatic haematuria | 512189014 | 95567008 | 512189014 |
| Optic nerve sheath hemorrhage | 2729121000006117 | 14460007 | 1217537016 |
| Adrenocortical haemorrhage | 356328018 | 237766002 | 356328018 |
| Haemarthrosis of DIP joint of finger | 310711014 | 202410008 | 310711014 |
| Liposuction removal of haematoma | 275087014 | 177714000 | 275087014 |
| Haemorrhage | 3325451000006112 | 50960005 | 495845019 |
| Bleeding skin | 5846621000006115 | 297968009 | 438563011 |
| Bleeding gums | 143080016 | 86276007 | 143080016 |
| Infected haematoma | 7510021000006118 | 698573001 | 2974732011 |
| Hemarthrosis of wrist | 4813321000006115 | 202407001 | 310703016 |
| Haemorrhage of cervix | 504705015 | 81488005 | 504705015 |
| External bleeding haemorrhoids | 1216226011 | 26421009 | 1216226011 |
| H/O: haematuria | 251768018 | 161550001 | 251768018 |
| Haematoma of kidney | 5458501000006117 | 262891006 | 390956018 |
| O/E - vitreous hemorrhages | 4573231000006119 | 163990004 | 255340011 |
| Evacuation of haematoma from vagina | 649591000006116 | 392256003 | 1490236018 |
| Bleeding | 4454741000006110 | 131148009 | 210860014 |
| Haemarthrosis of multiple sites | 812741000006113 | 76427005 | 503277015 |

| | | | |
|---|---|---|---|
| Haematoma - postoperative | 813261000006113 | 213262007 | 325137018 |
| On examination - subconjunctival hemorrhage | 4552921000006116 | 162815002 | 2667822015 |
| Haematoma of temporal region | 5965691000006113 | 307214007 | 450417012 |
| Bleeding varices | 3427151000006119 | 57052009 | 94882018 |
| Vulval haematoma evacuated | 869431000006119 | 65687007 | 869431000006119 |
| PU - chronic + haemorrhage | 885981000006112 | 49232000 | 885981000006112 |
| Bleeding of ear canal | 5875341000006114 | 300129004 | 441138011 |
| Haemarthrosis - hand joint | 890591000006112 | 27507004 | 890591000006112 |
| Optic disc haemorrhage | 5241101000006119 | 247233000 | 369184014 |
| Vulval hematoma | 3628381000006110 | 69385001 | 1218357015 |
| Blood in anterior chamber | 3723211000006110 | 75229002 | 1233846014 |
| Normocytic anemia following acute bleed | 5914361000006111 | 303060002 | 445120015 |
| Breast hematoma | 5913301000006112 | 302924003 | 444948013 |
| Corpus cavernosum hematoma | 4790761000006115 | 198031007 | 304492017 |
| Fresh bleeding from vagina | 5748511000006111 | 289536000 | 429436011 |
| Hemorrhage of cervix | 3825061000006115 | 81488005 | 135183016 |
| Scrotal hemorrhage | 3510231000006116 | 62182006 | 1218286015 |
| Bleeding haemorrhoid | 3335291000006119 | 51551000 | 1216702010 |
| Hemarthrosis of glenohumeral joint | 7529911000006111 | 700018009 | 2987396013 |
| Perineal hematoma | 5105871000006114 | 237331002 | 355701015 |
| Haemophthalmos without current injury | 4771041000006117 | 193287005 | 2547718010 |
| Hemarthrosis of acromioclavicular joint | 4813241000006115 | 202404008 | 310696014 |
| Haemorrhage of testis | 3764241000006119 | 77708008 | 503651019 |
| O/E - subconjunctival hemorrhage | 4552911000006112 | 162815002 | 253782010 |
| Hemorrhage of scrotum | 3510211000006110 | 62182006 | 103341016 |
| Evacuation of intracranial subdural hematoma | 3469871000006118 | 59712006 | 498517019 |
| Hemarthrosis of MCP joint | 4813361000006114 | 202408006 | 310708013 |
| Hemorrhage in optic nerve sheaths | 2729081000006119 | 14460007 | 24578017 |
| Acute hemorrhagic pancreatitis | 2568871000006112 | 4399003 | 7699015 |
| Retrohyaloid haemorrhage | 2683851000006119 | 11547003 | 411994019 |
| Nontraumatic scrotal haematoma | 4790951000006119 | 198058000 | 3036831010 |
| Evacuation of intracerebral hematoma | 2666411000006111 | 10458001 | 18195013 |
| Haemorrhage in optic nerve sheaths | 2729071000006117 | 14460007 | 476340016 |
| Paroxysmal hematoma of the finger | 5126821000006110 | 238824006 | 357937018 |
| Nontraumatic hematoma of testis | 3073991000006114 | 35644004 | 59462016 |
| Hematoma | 3073011000006117 | 35566002 | 59338013 |
| Haemarthrosis of sternoclavicular joint | 310694012 | 202403002 | 310694012 |
| Haemarthrosis of hip | 310715017 | 202411007 | 310715017 |
| Haemarthrosis of metacarpophalangeal joint | 4813371000006119 | 202408006 | 2692465010 |
| Surgical arrest of bleeding from internal nose | 267550012 | 172811007 | 267550012 |
| Perineal haematoma | 355700019 | 237331002 | 355700019 |
| Haemarthrosis of acromioclavicular joint | 310695013 | 202404008 | 310695013 |

| | | | |
|---|---|---|---|
| Peri-operative haemorrhage or haematoma | 403111013 | 269302003 | 403111013 |
| Thyroid haemorrhage and infarction | 292443012 | 190305000 | 292443012 |
| Haemorrhagic disorder due to circulating anticoagulants | 294307015 | 191287000 | 294307015 |
| Exophthalmos due to orbital haemorrhage | 298692016 | 194016009 | 298692016 |
| Tonsillar haemorrhage | 5036181000006114 | 232425007 | 348218013 |
| Haematemesis - cause unknown | 5983251000006115 | 308904008 | 452291015 |
| Vaginal bleeding problem | 5748581000006116 | 289543006 | 429444011 |
| Haematoma of groin | 5967711000006117 | 307390004 | 450644014 |
| Anticoagulant-induced bleeding | 5612371000006113 | 278365007 | 415242016 |
| Traumatic haematoma | 409951019 | 274179004 | 409951019 |
| Adrenal haemorrhage | 495316011 | 49111001 | 495316011 |
| Thyroid haemorrhage | 1216311012 | 3002002 | 1216311012 |
| Haemorrhage of eyelid | 105913010 | 63720000 | 105913010 |
| Intracranial hematoma following injury | 3111241000006113 | 37955001 | 63313013 |
| Haematoma of lower limb | 7373841000006118 | 449703000 | 2912531016 |
| Intraocular hemorrhage | 4021591000006119 | 93478000 | 154753016 |
| Hemorrhagic detachment of retinal pigment epithelium | 3312501000006111 | 50165004 | 83560014 |
| Haematoma of scrotum | 3962411000006116 | 89966002 | 508761019 |
| Dissection of carotid artery | 7806271000006110 | 720626009 | 3441846017 |
| Orbital haemorrhage | 3000741000006117 | 31056006 | 484766013 |
| Postoperative wound hemorrhage | 7510191000006115 | 698584008 | 2974777013 |
| Haematoma with intact skin | 5518251000006117 | 270911002 | 405394012 |
| Flame-shaped haemorrhage | 12222161000006112 | 247132001 | 3644832012 |
| Respiratory tract haemorrhage | 317993018 | 95431003 | 512121010 |
| Upper urinary tract haematuria | 5097861000006116 | 236717007 | 354811014 |
| Haemarthrosis of distal interphalangeal joint of finger | 4813431000006112 | 202410008 | 2695357016 |
| Nasal septal haematoma | 5035491000006111 | 232377002 | 348145019 |
| Respiratory tract hemorrhage | 4056521000006113 | 95431003 | 158078018 |
| Nontraumatic extradural intracranial haemorrhage | 6593151000006119 | 397809001 | 2916444013 |
| Vaginal hematoma | 3362091000006111 | 53162000 | 88426019 |
| Wrist hemarthrosis | 4813331000006117 | 202407001 | 310704010 |
| Hemorrhage of thyroid | 2547001000006117 | 3002002 | 6091011 |
| Postoperative hemorrhage | 4177071000006111 | 110265006 | 175034010 |
| Hyphema | 3723181000006111 | 75229002 | 124940012 |
| Suture of nose for epistaxis | 5724391000006117 | 287400009 | 2988047014 |
| O/E - epistaxis | 11905171000006116 | 164187003 | 255582011 |
| Nontraumatic epidural haemorrhage | 6593121000006111 | 397809001 | 1783616016 |
| On examination epistaxis | 11873291000006112 | 164187003 | 3634839017 |
| Retinal blot hemorrhages | 3288921000006118 | 48742005 | 1490492015 |
| Evacuation of haematoma of vulva | 3568141000006115 | 65687007 | 500187011 |
| Retrobulbar haematoma | 4774401000006112 | 194179009 | 298915016 |

| | | | |
|---|---|---|---|
| Hemarthrosis of hip | 4813471000006110 | 202411007 | 310713012 |
| Extradural haemorrhage after injury | 5459181000006112 | 262949005 | 391029012 |
| Vulval haematoma | 3628371000006112 | 69385001 | 1216870011 |
| Haematuria associated with urinary catheter | 1938771000006116 | 1938771000006100 | 1938771000006116 |
| Macroscopic haematuria | 4790361000006116 | 197941005 | 304354019 |
| O/E - throat haemorrhage | 255708010 | 164279008 | 255708010 |
| Epistaxis control -cryosurgery | 860731000006118 | 287393001 | 860731000006118 |
| O/E - retinal hemorrhages | 4573161000006116 | 163987005 | 255336019 |
| Bleeding | 3325491000006118 | 50960005 | 84923010 |
| Bleeding cervix | 5751971000006119 | 289799006 | 429729011 |
| Subungual haematoma | 403014019 | 269219004 | 403014019 |
| Haemorrhage of parathyroid | 292686019 | 190460008 | 292686019 |
| Haematoma | 3073001000006115 | 35566002 | 486214017 |
| Intraabdominal hemorrhage | 7283681000006111 | 443826006 | 2841291010 |
| Haematoma of abdominal wall | 7334721000006118 | 447096000 | 2883375018 |
| Wound haemorrhage | 5131561000006112 | 239161005 | 358453011 |
| Surgical arrest of post-extraction haemorrhage | 1006061000006112 | 1006061000006108 | 1006061000006112 |
| Retinal pigment epithelium haemorrhagic detachment | 1216693019 | 50165004 | 1216693019 |
| Intraoperative haemorrhage | 744731000006115 | 213261000 | 325136010 |
| Retinal haemorrhages | 814181000006113 | 28998008 | 1216289019 |
| Haematoma of broad ligament | 813321000006114 | 83294006 | 505536016 |
| Hemorrhage postprocedure | 4177101000006118 | 110265006 | 348212014 |
| Bleeding from vagina | 1786762014 | 289530006 | 2670003011 |
| Chronic hemorrhagic anemia | 6842811000006111 | 413533008 | 2536841015 |
| Haemarthrosis - ankle/foot | 890621000006114 | 267948008 | 890621000006114 |
| Haemarthrosis - hip joint | 890601000006116 | 202411007 | 890601000006116 |
| Vaginal vault haematoma | 5103381000006119 | 237102006 | 355396015 |
| Hemarthrosis of subtalar joint | 4813591000006110 | 202416002 | 310726011 |
| Surgical arrest of post-extraction hemorrhage | 4654761000006111 | 173330006 | 268350014 |
| Posttonsillectomy bleeding | 8015351000006113 | 35101000119106 | 3306550017 |
| Blood in urine - haematuria | 518011000006114 | 53298000 | 1216720018 |
| Pulmonary haemorrhage with glomerulonephritis | 3319101000006111 | 50581000 | 495714011 |
| Male haematocele | 3577551000006116 | 66259004 | 500367016 |
| Hemorrhage from ureter | 5096521000006110 | 236606001 | 354659012 |
| Non-traumatic testicular hematoma | 3074011000006119 | 35644004 | 1217934015 |
| Evacuation of vulval haematoma | 3568161000006116 | 65687007 | 1216830016 |
| Subconjunctival haemorrhage of both eyes | 8110661000006118 | 343341000119108 | 3427632015 |
| Deep retinal hemorrhage | 3288871000006111 | 48742005 | 81229018 |
| Pulmonary hemorrhage | 11989391000006117 | 78144005 | 301491000006118 |
| Dot AND blot hemorrhage | 3288881000006114 | 48742005 | 81231010 |
| Haemorrhage postprocedure | 4177091000006112 | 110265006 | 348211019 |
| Nontraumatic haematoma of breast | 4791291000006110 | 198123000 | 2619486016 |

| | | | |
|---|---|---|---|
| Liver haematoma | 5457181000006116 | 262796008 | 5457181000006116 |
| Pharyngeal hemorrhage | 11905801000006118 | 324618004 | 465449012 |
| Hematoma of vulva | 3628361000006117 | 69385001 | 115258016 |
| Haemorrhage of prostate | 3223401000006119 | 44843000 | 494041015 |
| Haemopericardium due to and following acute myocardial infarction | 12220761000006110 | 194862000 | 3673282010 |
| Hemorrhage at vitreoretinal interface | 5239141000006115 | 247097006 | 369011013 |
| Bleeding reduction: nasal | 3076691000006114 | 35807001 | 1490745018 |
| Expulsive choroidal hemorrhage | 2684981000006118 | 11623000 | 20081013 |
| Extradural haemorrhage | 884431000006116 | 397809001 | 884431000006116 |
| Manchester triage - PV bleeding | 1984281000006110 | 1984281000006106 | 1984281000006110 |
| Pleural haemorrhage | 3013671000006111 | 31892009 | 485062013 |
| Haemorrhage control | 3330361000006111 | 51241000 | 1490265012 |
| Hemarthrosis of first metatarsophalangeal joint | 4813651000006115 | 202419009 | 2692707015 |
| Hemorrhage from the ear | 2921441000006110 | 26322001 | 44087014 |
| Evacuation of hematoma of vagina | 6531331000006119 | 392256003 | 1477262019 |
| Abnormal vaginal bleeding | 456808010 | 301822002 | 443305015 |
| [D]Cough with haemorrhage | 317408012 | 66857006 | 500542015 |
| O/E - bleeding gums | 254280011 | 163164001 | 254280011 |
| Perinephric haematoma | 4789661000006116 | 197824007 | 304163014 |
| Haemarthrosis of elbow | 310699019 | 202405009 | 310699019 |
| Hemarthrosis of PIP joint of finger | 4813401000006116 | 202409003 | 310709017 |
| Haematoma of occipital scalp | 7097721000006119 | 429059007 | 2694431010 |
| Hemarthrosis of knee | 4813531000006111 | 202413005 | 310719011 |
| Urethral bleeding | 371989018 | 249301002 | 371989018 |
| Nontraumatic breast haematoma | 4791311000006114 | 198123000 | 3012832013 |
| Bleeding from tonsillar bed | 5271551000006114 | 249423002 | 372151014 |
| Ecchymosis of eyelid | 7002461000006117 | 423145008 | 2644948013 |
| Hyphaema | 124942016 | 75229002 | 124942016 |
| Incision and drainage of haematoma | 7276661000006117 | 443385002 | 2838330015 |
| Bleeding in mouth and/or pharynx | 5271471000006115 | 249417007 | 1489865014 |
| Post-surgical epistaxis | 5035211000006119 | 232357009 | 348118015 |
| Haemarthrosis of hand | 812861000006119 | 27507004 | 483566016 |
| Postoperative haemorrhage control | 5564161000006119 | 274508002 | 410324010 |
| Liposuction removal of hematoma | 4679251000006115 | 177714000 | 275086017 |
| Haemorrhagic choroidal detachment | 479008012 | 19031009 | 479008012 |
| Superficial retinal haemorrhage | 499542018 | 63297008 | 499542018 |
| Haematoma of pinna | 507866018 | 88050005 | 507866018 |
| Optic nerve sheath haemorrhage | 1216033015 | 14460007 | 1216033015 |
| Extradural h'ge inj no open intracran wnd+>24hr LOC+recovery | 660041000006117 | 43216008 | 72101019 |
| Nasal hemorrhage | 11780441000006118 | 249366005 | 3633923016 |
| Bleeding of subgingival space | 3902881000006112 | 86276007 | 507017016 |

| | | | |
|---|---|---|---|
| Total blood loss | 3279111000006114 | 48149007 | 80230019 |
| Abnormal bleeding/vaginal discharge | 981591000006114 | 981591000006105 | 981591000006114 |
| Subperiosteal haematoma | 989951000006111 | 274179004 | 989951000006111 |
| Recurrent microscopic haematuria | 5656851000006118 | 281858008 | 419987015 |
| [X]Haemorrhage from respiratory passages, unspecified | 317999019 | 95431003 | 512121010 |
| Respiratory tract haemorrhage | 4056511000006117 | 95431003 | 4056511000006117 |
| Haemorrhage at vitreoretinal interface | 5239131000006113 | 247097006 | 369010014 |
| Seen in haematuria clinic | 1682771000006111 | 1682771000006107 | 1682771000006111 |
| Dot AND blot haemorrhage | 3288911000006114 | 48742005 | 495203014 |
| VH - Vitreous hemorrhage | 3005211000006118 | 31341008 | 2579343019 |
| Haemarthrosis of shoulder joint | 7529901000006113 | 700018009 | 2987346012 |
| Pulmonary hemorrhage with glomerulonephritis | 3319071000006118 | 50581000 | 84267019 |
| Tracheostomy hemorrhage | 3848221000006114 | 82872004 | 137462016 |
| Haemorrhagic diarrhoea | 4058311000006112 | 95545007 | 512178019 |
| Hematoma | 6438781000006118 | 385494008 | 1477038019 |
| Adrenal hemorrhage | 3294631000006115 | 49111001 | 81821014 |
| Deformity/haematoma of pinna | 5721171000006118 | 286943000 | 426436012 |
| Intragel vitreous haemorrhage | 3005181000006117 | 31341008 | 1216354019 |
| Hemorrhage control by packing | 4716051000006119 | 182605004 | 282264011 |
| Haemarthrosis of glenohumeral joint | 7529881000006111 | 700018009 | 2987361013 |
| Haemorrhagic pancreatitis | 2568891000006113 | 4399003 | 493744012 |
| Hemarthrosis of hand | 2941131000006113 | 27507004 | 46020011 |
| Evacuation of haematoma of vagina | 6531321000006117 | 392256003 | 1476301016 |
| Pulmonary haemorrhage | 3771001000006116 | 78144005 | 3771001000006116 |
| Retroperitoneal hematoma | 5088461000006119 | 236002003 | 353769018 |
| Minor surgery postoperative complication - haemorrhage | 914911000006113 | 914911000006109 | 914911000006113 |
| Postmenopausal postcoital bleeding | 2534105016 | 415149004 | 2534105016 |
| Hemarthrosis of metacarpophalangeal joint | 4813381000006116 | 202408006 | 2694346016 |
| Acute blood loss anemia | 5500931000006115 | 267530009 | 2536036012 |
| Epistaxis control | 11776911000006118 | 35807001 | 3633933012 |
| Traumatic extradural hematoma without open intracranial wound | 6053331000006117 | 315048006 | 459516011 |
| Haemarthrosis of subtalar joint | 310725010 | 202416002 | 310725010 |
| Hip haemarthrosis | 310716016 | 202411007 | 310716016 |
| Haemarthrosis of talonavicular joint | 310728012 | 202417006 | 310728012 |
| Dysfunctional uterine hemorrhage | 2803941000006119 | 19155002 | 479159011 |
| Intra-abdominal hematoma | 5088381000006115 | 235999009 | 353760019 |
| Posterior epistaxis | 5035191000006115 | 232355001 | 348116016 |
| Evacuation of haematoma from temporal lobe of brain | 265689012 | 171473007 | 265689012 |
| Haemorrhagic fibrinogenolysis | 3596661000006113 | 67406007 | 500723016 |
| Mediastinal haematoma | 2860621000006113 | 22625005 | 481391011 |
| Retinal macroaneurysm | 369052016 | 247124009 | 369052016 |

| | | | |
|---|---|---|---|
| Haematoma of palate | 5455351000006116 | 262649007 | 390623016 |
| Subungual haematoma of foot | 11784831000006115 | 274203005 | 3513825017 |
| Recurrent frank haematuria | 5656811000006119 | 281856007 | 419982014 |
| Evacuation of perianal haematoma | 433186012 | 293018003 | 433186012 |
| Packing for non-obstetric uterine bleeding | 4716101000006119 | 182607007 | 282272013 |
| Testicular haemorrhage | 1216948012 | 77708008 | 1216948012 |
| Extradural haemorrhage following injury without open intracranial wound | 402930018 | 43216008 | 493552012 |
| Haematoma of leg | 2674499019 | 91603007 | 151760014 |
| Prostatic congestion or haemorrhage NOS | 304407017 | 197973004 | 304404012 |
| Retinal flame hemorrhage | 5239731000006114 | 247132001 | 1773842011 |
| Haematoma of testis | 5681911000006114 | 283944002 | 422515011 |
| Epistaxis | 11780411000006117 | 249366005 | 3633921019 |
| Haemarthrosis - wrist joint | 890581000006114 | 202407001 | 890581000006114 |
| Haemarthrosis - knee joint | 890611000006118 | 202413005 | 890611000006118 |
| C/O p.v. bleeding | 12483181000006119 | 275570002 | 411565016 |
| Drainage of hematoma of external ear and control cavity with bolster suture | 4650071000006114 | 172619007 | 267228012 |
| Hemorrhage of testis | 3764251000006117 | 77708008 | 128969012 |
| Acute post-haemorrhagic anaemia | 5500971000006117 | 267530009 | 2536040015 |
| Hemoptysis | 11905461000006115 | 66857006 | 111056015 |
| Persistent microscopic haematuria | 5656831000006113 | 281857003 | 419984010 |
| Anterior epistaxis | 5035151000006114 | 232354002 | 348112019 |
| Recurrent haematuria co-occurrent and due to minor glomerular abnormality | 183201000006117 | 714823009 | 3300550011 |
| Conjunctival hemorrhage | 2835651000006118 | 21117005 | 35492010 |
| Hemophthalmos (excluding current injury) | 4771051000006115 | 193287005 | 297685012 |
| Bleeding varicose vein of leg | 4779581000006117 | 195448005 | 300726015 |
| Nasal haemorrhage | 11780431000006111 | 249366005 | 3633922014 |
| Suprachoroidal haemorrhage | 6939791000006112 | 419596007 | 2578131018 |
| Haemarthrosis of first metatarsophalangeal joint | 4813641000006117 | 202419009 | 2692686017 |
| Gingival haemorrhage | 3902921000006116 | 86276007 | 507021011 |
| Hemorrhagic pancreatitis | 2568881000006110 | 4399003 | 7701015 |
| O/E - speculum bleeding through os | 4563041000006113 | 163415000 | 4563041000006113 |
| Kidney hematoma without rupture of capsule, with open wound into cavity | 4855101000006116 | 210205007 | 321132016 |
| Postoperative control of hemorrhage of adenoids | 3135621000006114 | 39508008 | 66269014 |
| Macular subretinal hemorrhage | 5240211000006116 | 247161000 | 369102011 |
| Acute post-hemorrhagic anemia | 5500921000006118 | 267530009 | 2536035011 |
| Hemorrhage from pharynx | 6119821000006112 | 324618004 | 465450012 |
| Packing for haemorrhage | 4716031000006114 | 182605004 | 282265012 |
| Essential haematuria | 458617017 | 314280007 | 458617017 |
| Control of hemorrhage of nose | 3076641000006117 | 35807001 | 59727010 |
| Drainage of hematoma of pinna | 4650011000006117 | 172617009 | 267222013 |
| Exophthalmos due to orbital hemorrhage | 4773741000006115 | 194016009 | 298691011 |

| | | | |
|---|---|---|---|
| Bleeding from breast | 1805191000006113 | 52297004 | 87019010 |
| Intraretinal hemorrhage | 2966441000006114 | 28998008 | 1783893010 |
| Hematoma of kidney | 5458511000006119 | 262891006 | 390957010 |
| Crevicular bleeding of gum | 3902901000006114 | 86276007 | 507019018 |
| Haemarthrosis of shoulder | 2540431000006115 | 2602008 | 1216220017 |
| O/E - throat hemorrhage | 4577941000006112 | 164279008 | 255707017 |
| Hemorrhage of parathyroid | 4758531000006114 | 190460008 | 292687011 |
| Testicular hemorrhage | 3764271000006110 | 77708008 | 1218435011 |
| Traumatic extradural hematoma | 5459161000006119 | 262949005 | 391026017 |
| Postoperative hematoma formation | 4872571000006110 | 213262007 | 325138011 |
| Hemarthrosis of the ankle and/or foot | 5502491000006111 | 267948008 | 1477399016 |
| Airway hemorrhage | 4056541000006118 | 95431003 | 1784236019 |
| On examination - retinal hemorrhages | 4573171000006111 | 163987005 | 2668326010 |
| Gingival crevice bleeding | 3902891000006110 | 86276007 | 507018014 |
| Haematoma of auricle | 3930871000006116 | 88050005 | 507867010 |
| Bleeding unrelated to menstrual cycle | 3557191000006117 | 64996003 | 108033013 |
| Superficial retinal hemorrhage | 3529381000006112 | 63297008 | 105219016 |
| Adrenocortical hemorrhage | 5112201000006111 | 237766002 | 356329014 |
| Stomal bleeding | 5090211000006119 | 236129005 | 353961014 |
| Other abnormal uterine and vaginal bleeding | 456805013 | 301822002 | 443305015 |
| Pleural hemorrhage | 3013661000006116 | 31892009 | 53279018 |
| Bloody diarrhea | 4058331000006118 | 95545007 | 158255016 |
| On examination - vitreous hemorrhages | 4573251000006114 | 163990004 | 2668347014 |
| History of postmenopausal bleeding | 4544131000006114 | 161788002 | 2986807011 |
| Drainage of hematoma of external ear | 4650031000006111 | 172617009 | 267224014 |
| Normocytic anaemia following acute bleed | 445119014 | 303060002 | 445119014 |
| Surgical arrest of bleeding post dental extraction | 117221000006116 | 173330006 | 268351013 |
| Haematoma of perineal wound | 6357731000006110 | 371610007 | 1207307018 |
| Control of nasal haemorrhage | 3076671000006113 | 35807001 | 1490108018 |
| Haemarthrosis of lesser metatarsophalangeal joint | 812721000006118 | 202420003 | 310733011 |
| Control of epistaxis by suture | 5724371000006118 | 287400009 | 2988411019 |
| On examination - throat hemorrhage | 4577951000006114 | 164279008 | 2667890017 |
| [X]Vitreous haemorrhage in diseases classified elsewhere | 299491012 | 31341008 | 484848010 |
| Secondary and recurrent haemorrhage | 157451000006115 | 212373009 | 323899017 |
| Recurrent haematuria co-occurrent and due to diffuse mesangiocapillary glomerulonephritis | 189181000006115 | 714819001 | 3300538017 |
| Intragel vitreous hemorrhage | 3005191000006119 | 31341008 | 1217853012 |
| Hemarthrosis of shoulder | 2540441000006113 | 2602008 | 1217723011 |
| Hyphaemia | 3723201000006112 | 75229002 | 502959013 |
| Haematoma with intact skin | 405394012 | 105618004 | 169739016 |
| Prostatic haemorrhage | 1216632014 | 44843000 | 1216632014 |
| Prostatic congestion or haemorrhage | 304404012 | 197973004 | 304404012 |

| | | | |
|---|---|---|---|
| Haemarthrosis of other specified site | 12728591000006117 | 687801000000104 | 1505711000000112 |
| Haemarthrosis of the knee | 219591000000116 | 202413005 | 310720017 |
| Perirenal haematoma | 304164015 | 197824007 | 304164015 |
| Epistaxis control - suture | 860721000006116 | 287400009 | 860721000006116 |
| Control of haemorrhage | 878511000006114 | 51241000 | 878511000006114 |
| Recurrent haematuria co-occurrent and due to dense deposit disease | 304001012 | 714813000 | 3300520015 |
| Surgical arrest of postoperative bleeding from tonsillar bed | 268510012 | 173440008 | 268510012 |
| Hemorrhage into bladder wall | 2779531000006112 | 17615006 | 29772016 |
| Re-exploration of organ and surgical arrest of postoperative bleeding NOC | 184131000006117 | 621531000000109 | 1372281000000112 |
| [X]Sequelae of other nontraumatic intracranial haemorrhage | 426231000006114 | 363302008 | 482447013 |
| Uterine hemorrhage | 3116561000006117 | 38280009 | 64072010 |
| Renal artery hemorrhage | 4789521000006111 | 197812000 | 304141017 |
| Haemarthrosis of knee | 310720017 | 202413005 | 310720017 |
| Haemarthrosis of MCP joint | 310707015 | 202408006 | 310707015 |
| Epistaxis control - cautery | 5724261000006115 | 287382003 | 426914018 |
| Pack to control postnatal haemorrhage | 394488013 | 265641004 | 394488013 |
| Hemarthrosis of distal radioulnar joint | 4813301000006113 | 202406005 | 310702014 |
| Choroidal haemorrhage and rupture | 297942018 | 193474002 | 297942018 |
| Painless haematuria | 304345017 | 197938001 | 304345017 |
| Frank haematuria | 304353013 | 197941005 | 304353013 |
| Haemorrhage of seminal vesicle | 2635281000006115 | 8465005 | 506159011 |
| Spontaneous hyphaema | 5914111000006110 | 303019009 | 445086014 |
| Secondary haemorrhage | 5459051000006115 | 262938004 | 391013016 |
| Secondary post tonsillectomy hemorrhage | 6046541000006111 | 314484001 | 458850015 |
| Haemorrhage of blood vessel | 6715161000006118 | 405539004 | 2153971019 |
| Acute haemorrhagic pancreatitis | 493743018 | 4399003 | 493743018 |
| Deep retinal haemorrhage | 495202016 | 48742005 | 495202016 |
| Liver haematoma and contusion with open wound into cavity | 321093013 | 21580006 | 36197018 |
| Epistaxis from anterior nasal septum | 5035171000006116 | 232354002 | 348114018 |
| Periorbital haematoma | 2626321000006115 | 7927006 | 504059010 |
| Traumatic intracranial haemorrhage | 660121000006113 | 450410005 | 2915482010 |
| Scrotal hematoma due to non-traumatic cause | 4790931000006114 | 198058000 | 304526010 |
| Extradural hemorrhage following injury without open intracranial wound | 3196371000006118 | 43216008 | 72101019 |
| Subcutaneous haematoma | 2584481000006117 | 5309003 | 496551013 |
| Conjunctival haemorrhage | 298518014 | 21117005 | 480926011 |
| Haematoma of skin | 5625851000006111 | 279420009 | 416697010 |
| Perioperative haematoma | 3636045011 | 762958007 | 3636045011 |
| Haematuria NOS | 886851000006119 | 34436003 | 886851000006119 |
| Chemical haematuria | 5097881000006114 | 236718002 | 354812019 |
| Haematoma of cervix | 5682141000006118 | 283965008 | 422540016 |

| | | | |
|---|---|---|---|
| Pharyngeal haemorrhage | 317317013 | 324618004 | 465451011 |
| Haematoma of scalp | 7223281000006117 | 439461004 | 2794134010 |
| Perinephric hematoma | 4789671000006111 | 197824007 | 304161011 |
| Evidence of recent epistaxis | 5596491000006110 | 277236000 | 413696012 |
| Epistaxis | 11905371000006119 | 12441001 | 21406015 |
| Macular subretinal haemorrhage | 5240201000006119 | 247161000 | 369103018 |
| Traumatic intracranial extradural hematoma | 5459191000006110 | 262949005 | 2915790014 |
| Intra-alveolar haemorrhage | 3771051000006117 | 78144005 | 503766010 |
| Vaginal introitus bleeding | 5264711000006114 | 248978005 | 371563010 |
| Pelvic hematoma | 5477491000006112 | 264558008 | 393036018 |
| Deformity/hematoma of pinna | 5721181000006115 | 286943000 | 426435011 |
| PCB - Postcoital bleeding | 3290931000006116 | 48880000 | 1230662018 |
| Subungual hematoma | 5510911000006111 | 269219004 | 403015018 |
| Cortical hemorrhage | 3299621000006119 | 49422009 | 82320015 |
| Haematoma of spleen | 5457461000006118 | 262818004 | 390850011 |
| Haematuria of undiagnosed cause | 6044681000006112 | 314280007 | 458614012 |
| Haemorrhage of thyroid | 2546991000006113 | 3002002 | 484460015 |
| Haemarthrosis of proximal interphalangeal joint of finger | 4813391000006118 | 202409003 | 1772566017 |
| Hemorrhage into cornea | 5029241000006115 | 231916005 | 347516011 |
| Bleeding from nipple | 1805151000006119 | 248838002 | 371379012 |
| Hemarthrosis of proximal interphalangeal joint of finger | 4813421000006114 | 202409003 | 1773835011 |
| Complaining of per vaginam bleeding | 5574801000006117 | 275570002 | 5574801000006117 |
| Evacuation of subungual hematoma | 3939701000006111 | 88584000 | 146879013 |
| Haemorrhagic detachment of retinal pigment epithelium | 3312491000006115 | 50165004 | 495600016 |
| Hemarthrosis-1st metatarsophalangeal joint | 4813631000006110 | 202419009 | 310731013 |
| Retinal hemorrhage | 2966401000006112 | 28998008 | 48549019 |
| Haemorrhage from the ear | 2921451000006112 | 26322001 | 483175014 |
| Secondary and recurrent haemorrhage | 323899017 | 212373009 | 323899017 |
| Aspiration of haematoma of organ NOC | 279090012 | 621461000000103 | 1372141000000114 |
| Ureteric haemorrhage | 354662010 | 236606001 | 354662010 |
| Haemorrhage control by packing | 282266013 | 182605004 | 282266013 |
| Profuse vaginal bleeding | 5748551000006112 | 289540009 | 429440019 |
| Secondary post tonsillectomy haemorrhage | 458851016 | 314484001 | 458851016 |
| Breast haematoma | 5913291000006111 | 302924003 | 444949017 |
| Diabetic vitreous haemorrhage | 6022831000006119 | 312910009 | 456713019 |
| Haemophthalmos (excluding current injury) | 297686013 | 193287005 | 297686013 |
| Renal artery haemorrhage | 304142012 | 197812000 | 304142012 |
| Haematuria - cause not known | 6044721000006117 | 314280007 | 458616014 |
| Subhyaloid haemorrhage | 5239151000006118 | 247098001 | 369013011 |
| Haemorrhage - postoperative | 814171000006110 | 110265006 | 202906017 |
| VH - Vitreous haemorrhage | 3005201000006116 | 31341008 | 2579342012 |

| | | | |
|---|---|---|---|
| Hemarthrosis of distal interphalangeal joint of finger | 4813451000006117 | 202410008 | 2694113016 |
| Haemarthrosis - knee joint | 989491000006111 | 202413005 | 989491000006111 |
| Vaginal vault bleeding | 5650051000006112 | 281309004 | 419293019 |
| Abnormal uterine bleeding unrelated to menstrual cycle | 456806014 | 312984006 | 456806014 |
| Vitreous haemorrhage | 484848010 | 31341008 | 484848010 |
| Insertion of Brighton epistaxis balloon | 267563018 | 172821004 | 267563018 |
| Haematuria/abnormal colour | 962031000006113 | 962031000006109 | 962031000006113 |
| Haemarthrosis - elbow joint | 890571000006111 | 202405009 | 890571000006111 |
| Hip hemarthrosis | 4813481000006113 | 202411007 | 310714018 |
| Anaemia due to acute blood loss | 5500951000006110 | 267530009 | 2536038013 |
| Nontraumatic hematoma of breast | 4791281000006112 | 198123000 | 2619485017 |
| Intraocular bleeding | 4021601000006110 | 93478000 | 154754010 |
| Haemoperitoneum | 3236681000006116 | 45626005 | 494283018 |
| Haematoma AND contusion of liver with open wound into abdominal cavity | 2843221000006113 | 21580006 | 481063010 |
| Peritoneal haemorrhage | 3236691000006118 | 45626005 | 494284012 |
| Evacuation of hematoma from temporal lobe of brain | 4644231000006112 | 171473007 | 265688016 |
| Prostatic congestion or hemorrhage | 4790551000006114 | 197973004 | 304405013 |
| Choroidal haemorrhage or rupture NOS | 297946015 | 193474002 | 297942018 |
| Hemarthrosis of talonavicular joint | 4813611000006116 | 202417006 | 310727019 |
| PMB - Postmenopausal bleeding | 3748141000006114 | 76742009 | 503350010 |
| Hemorrhagic fibrinogenolysis | 3596641000006114 | 67406007 | 112019014 |
| Preretinal hemorrhage | 2683841000006116 | 11547003 | 19960018 |
| Evacuation of vaginal haematoma | 6531341000006112 | 392256003 | 1490236018 |
| Haemarthrosis of tibiofibular joint | 4813541000006118 | 202414004 | 310721018 |
| Haematoma | 982751000006117 | 982751000006101 | 982751000006117 |
| Haemarthrosis - shoulder joint | 890561000006116 | 2602008 | 890561000006116 |
| Nontraumatic epidural hemorrhage | 6593131000006114 | 397809001 | 1784005019 |
| EDH - Extradural hematoma | 3850541000006117 | 82999001 | 1218488017 |
| Postoperative bleeding | 4177081000006114 | 110265006 | 175035011 |
| Nontraumatic extradural hemorrhage | 6593111000006115 | 397809001 | 1774439018 |
| Haemorrhage of dialysis arteriovenous shunt | 2240911000000119 | 867031000000105 | 2240911000000119 |
| Drainage of haematoma of pinna | 4650001000006115 | 172617009 | 267225010 |
| Surgical arrest of bleeding from internal nose OS | 117211000006112 | 172811007 | 267550012 |
| On examination - vitreous haemorrhages | 4573241000006112 | 163990004 | 2668346017 |
| Evacuation of hematoma | 4305861000006115 | 118441006 | 176736013 |
| Hemorrhage of eyelid | 3536341000006111 | 63720000 | 105912017 |
| Paroxysmal haematoma of the finger | 1697911000006119 | 238824006 | 357936010 |
| Evacuation of intracranial extradural hematoma | 4645651000006115 | 171713001 | 265969013 |
| [X]Haemorrhage, not elsewhere classified | 318044011 | 131148009 | 210860014 |
| Optic nerve haemorrhage | 2729101000006110 | 14460007 | 476341017 |
| Superficial haematoma | 5518261000006115 | 270911002 | 405395013 |

| | | | |
|---|---|---|---|
| Nasal packing for control of haemorrhage | 5563931000006112 | 274491002 | 2669563011 |
| Surgical arrest of bleeding from internal nose NOS | 267560015 | 172811007 | 267550012 |
| VTE risk assessment - active bleeding | 1992421000006117 | 1992421000006101 | 1992421000006117 |
| Secondary and recurrent hemorrhage | 4868001000006115 | 212373009 | 323900010 |
| Control of epistaxis using nasal packing | 5563951000006117 | 274491002 | 2793090012 |
| Tracheostomy haemorrhage | 505306012 | 82872004 | 505306012 |
| Haematoma of rectus sheath | 451473014 | 308154003 | 451473014 |
| Control of epistaxis by cautery | 5724251000006117 | 287382003 | 2988735013 |
| Hemophthalmos, except current injury | 4771091000006114 | 193287005 | 2550869013 |
| Hematoma of broad ligament | 3855481000006115 | 83294006 | 138135015 |
| Testicular haematoma due to nontraumatic cause | 110341000006115 | 35644004 | 1216444017 |
| Surgical arrest of post-extraction haemorrhage | 117241000006111 | 173330006 | 268351013 |
| Nontraumatic extradural intracranial hemorrhage | 6593141000006116 | 397809001 | 2915638010 |
| Haemarthrosis of sacroiliac joint | 812781000006119 | 202412000 | 310717013 |
| Bleeding from nose | 317316016 | 249366005 | 372073016 |
| Hemarthrosis of lesser metatarsophalangeal joint | 4813671000006113 | 202420003 | 310734017 |
| Splenic haematoma | 5457451000006115 | 262818004 | 390849011 |
| Hemarthrosis of sacroiliac joint | 4813511000006117 | 202412000 | 310718015 |
| Peri-operative haemorrhage or haematoma | 11904821000006110 | 269302003 | 403111013 |
| Hemorrhage of prostate | 3223411000006116 | 44843000 | 74812016 |
| Choroidal haemorrhage | 2500391000006119 | 122003 | 2500391000006119 |
| Pack to control postnatal vaginal bleeding | 394487015 | 265641004 | 394487015 |
| Evacuation of haematoma of vulva | 500187011 | 65687007 | 1216830016 |
| Recurrent haematuria co-occurrent and due to diffuse crescentic glomerulonephritis | 189131000006116 | 714815007 | 3300526014 |
| Postoperative haemorrhage | 217221000006118 | 110265006 | 202906017 |
| Hematoma with intact skin | 5518211000006118 | 270911002 | 405389012 |
| Haemopericardium | 39303019 | 23412002 | 39303019 |
| Gingival crevicular bleeding | 3902871000006114 | 86276007 | 507016013 |
| Intramuscular haematoma | 768621000006112 | 262969000 | 391070014 |
| Intraocular haemorrhage | 769531000006117 | 93478000 | 510363011 |
| Extradural h'ge inj no open intracranial wnd+<1hr loss consc | 660101000006115 | 43216008 | 72101019 |
| Extradural h'ge inj no open intracranial wnd + no loss consc | 660081000006111 | 43216008 | 72101019 |
| Haemorrhage of corpus cavernosum | 3629431000006110 | 69452005 | 501277010 |
| Haemarthrosis of other tarsal joint | 310729016 | 267948008 | 400242010 |
| Preretinal haemorrhage | 207961000006117 | 11547003 | 411993013 |
| Haemarthrosis of the forearm | 812851000006116 | 202407001 | 310703016 |
| Incision and drainage of breast haematoma | 5570921000006110 | 275236004 | 411190012 |
| Evacuation of hematoma of vulva | 3568151000006118 | 65687007 | 109140016 |
| Corpus cavernosum hemorrhage | 3629461000006118 | 69452005 | 1218360010 |
| Packing for hemorrhage | 4716071000006112 | 182605004 | 282267016 |
| Haemarthrosis - multiple joint | 890641000006119 | 76427005 | 890641000006119 |

| | | | |
|---|---|---|---|
| Peritoneal hemorrhage | 3236661000006114 | 45626005 | 76084011 |
| Hemarthrosis of multiple sites | 3743251000006114 | 76427005 | 126927017 |
| Pulmonary hemorrhage | 11905681000006111 | 78144005 | 129684011 |
| Nontraumatic breast hematoma | 4791301000006111 | 198123000 | 3012814013 |
| Acute blood loss anaemia | 5500961000006112 | 267530009 | 2536039017 |
| Retrobulbar hemorrhage | 4774381000006112 | 194179009 | 298913011 |
| Hemorrhagic choroidal detachment | 2802071000006118 | 19031009 | 32080012 |
| Pneumohaemothorax | 2763821000006115 | 16632002 | 28150019 |
| On examination - bleeding gums | 4558931000006115 | 163164001 | 2667369016 |
| Bleeding after menopause | 3748151000006111 | 76742009 | 503351014 |
| Drainage of hematoma of pinna and insertion of bolster sutures | 4650091000006110 | 172619007 | 267231013 |
| Evacuation of vaginal hematoma | 6531351000006114 | 392256003 | 1490465014 |
| Haemorrhage of dialysis arteriovenous fistula | 2240871000000116 | 867011000000102 | 2240871000000116 |
| Haematuria - symptom | 813391000006111 | 34436003 | 57480016 |
| Haemarthrosis of distal radioulnar joint | 812671000006110 | 202406005 | 310701019 |
| Haemarthrosis-1st metatarsophalangeal joint | 812951000006113 | 202419009 | 310732018 |
| H/O: postcoital bleeding | 811291000006116 | 161786003 | 252099013 |
| Scrotal haematoma due to non-traumatic cause | 156461000006116 | 198058000 | 304525014 |
| Recurrent haematuria co-occurrent and due to focal and segmental glomerular lesions | 189151000006111 | 714827005 | 3300562010 |
| Hemarthrosis of shoulder region | 2540421000006118 | 2602008 | 5429017 |
| Retrobulbar hematoma | 4774391000006110 | 194179009 | 298914017 |
| Male genital haematoma NOS | 304528011 | 198057005 | 304524013 |
| Functional uterine haemorrhage NOS | 305075011 | 19155002 | 32278010 |
| Complaining of per vaginam bleeding | 411565016 | 275570002 | 2984033019 |
| Recurrent and persistent haematuria, dense deposit disease | 11904541000006117 | 281860005 | 189231000006110 |
| Subretinal hemorrhage | 2721051000006114 | 13937002 | 23758016 |
| Deep intra-retinal haemorrhages | 3288901000006111 | 48742005 | 495201011 |
| Internal prosthetic device causing hemorrhage | 4871651000006112 | 213133000 | 324953013 |
| Airway haemorrhage | 4056531000006111 | 95431003 | 1783845016 |
| Vaginal haematoma | 496575013 | 53162000 | 496575013 |
| Anti-GBM nephritis with pulmonary hemorrhage | 3319091000006117 | 50581000 | 84270015 |
| Hemarthrosis of the ankle | 4813571000006114 | 202415003 | 310723015 |
| [X]Other specified abnormal uterine and vaginal bleeding | 305260014 | 301822002 | 443305015 |
| Contact bleeding from cervix | 590531000006114 | 248980004 | 371565015 |
| Retinal blot haemorrhages | 3288891000006112 | 48742005 | 495200012 |
| Subconjunctival hemorrhage | 3781231000006112 | 78768009 | 130708019 |
| Microscopic haematuria | 304349011 | 197940006 | 304349011 |
| Liver haematoma | 739711000006114 | 262796008 | 390818018 |
| Liver haematoma | 12704641000006114 | 262796008 | 390819014 |
| Hemarthrosis of shoulder joint | 7529891000006114 | 700018009 | 2987325015 |
| Haemarthrosis - other joint | 890631000006112 | 687801000000104 | 890631000006112 |

| | | | |
|---|---|---|---|
| Choroidal haemorrhage | 297943011 | 122003 | 1256014 |
| Extradural h'ge inj no open intracran wnd+>24hr LOC-restored | 660051000006115 | 43216008 | 72101019 |
| Haemarthrosis of the lower leg | 812881000006112 | 202413005 | 310719011 |
| Epidural haematoma | 7583041000006117 | 703861005 | 3010111010 |
| Bleeding of oral mucosa | 5271491000006119 | 249418002 | 372144017 |
| Control of epistaxis by unlisted technique | 3076661000006118 | 35807001 | 59732011 |
| Control of haemorrhage | 3330341000006112 | 51241000 | 495965010 |
| Hematoma of perineal wound | 6357741000006117 | 371610007 | 1208119015 |
| Closed heart injury with haemopericardium | 455462013 | 311830005 | 455462013 |
| Tanner devascularisation for bleeding varices | 114801000006118 | 173617001 | 268812014 |
| Traumatic retinopathy | 5030871000006117 | 232030000 | 347668010 |
| Haemorrhage of dialysis vascular access | 2237901000000119 | 865801000000100 | 2237901000000119 |
| Extradural haemorrhage | 3850521000006112 | 82999001 | 505380015 |
| Spleen haematoma without mention of open wound into cavity | 135631000006119 | 262818004 | 390849011 |
| Elbow hemarthrosis | 4813271000006111 | 202405009 | 310698010 |
| Haematoma | 813271000006118 | 385494008 | 1476077014 |
| Ureteric hemorrhage | 5096531000006113 | 236606001 | 354661015 |
| Choroidal hemorrhage | 2500401000006117 | 122003 | 1256014 |
| Haemorrhage from ureter | 5096511000006119 | 236606001 | 354660019 |
| [X]Sequelae of stroke,not specfd as h'morrhage or infarction | 426321000006116 | 195239002 | 300403014 |
| Haemopneumothorax | 477873012 | 16632002 | 477873012 |
| Haematoma of vulva | 501268015 | 69385001 | 501268015 |
| Bleeding from urethra | 371988014 | 249301002 | 371988014 |
| Irregular uterine bleeding | 3804361000006111 | 80182007 | 504362017 |
| Kidney haematoma without mention of open wound into cavity | 754831000006113 | 262891006 | 390956018 |
| Kidney haematoma without rupture of capsule, with open wound into cavity | 754821000006110 | 210205007 | 321133014 |
| Hemorrhagic diarrhea | 4058321000006116 | 95545007 | 158254017 |
| Intraretinal haemorrhage | 2966431000006116 | 28998008 | 1783522015 |
| Haemarthrosis of the ankle and/or foot | 5502461000006115 | 267948008 | 1476436016 |
| Choroidal haemorrhage | 12702351000006114 | 122003 | 469795010 |
| Extradural h'ge inj no open intracranial wnd + unspec consc | 660091000006114 | 262949005 | 391026017 |
| Hyphaema of right eye | 3526020015 | 680521000119101 | 3526020015 |
| Haemarthrosis of shoulder | 1216220017 | 700018009 | 2987325015 |
| Epistaxis symptom | 647631000006114 | 249366005 | 372073016 |
| Has nosebleeds - epistaxis | 816991000006115 | 249366005 | 372073016 |
| Subperiosteal haematoma | 933081000006112 | 669341000000103 | 933081000006112 |
| Nontraumatic haematoma of testis | 3073981000006111 | 35644004 | 486230013 |
| Retinal flame haemorrhage | 5239721000006111 | 247132001 | 1772573010 |
| Hemorrhage in uterus | 3116551000006119 | 38280009 | 64071015 |
| Hematoma of auricle | 3930861000006111 | 88050005 | 145964013 |

| | | | |
|---|---|---|---|
| Perirenal hematoma | 4789681000006114 | 197824007 | 304162016 |
| Postoperative hemorrhage control | 5564171000006114 | 274508002 | 410325011 |
| Hemorrhage of corpus cavernosum | 3629441000006117 | 69452005 | 115380016 |
| Secondary and recurrent haemorrhage | 157461000006118 | 212373009 | 323899017 |
| Reopening of abdomen and re-exploration of intra-abdominal operation site and surgical arrest of postoperative bleeding | 177571000006110 | 177956008 | 275461019 |
| Renal haematoma with open wound into cavity | 176891000006112 | 210205007 | 321133014 |
| Postoperative haematoma formation | 217211000006114 | 213262007 | 325137018 |
| Hemarthrosis of interphalangeal joint of toe | 4813691000006114 | 202421004 | 310735016 |
| Gingival hemorrhage | 3902911000006112 | 86276007 | 507020012 |
| Hemarthrosis of the ankle and foot | 5502471000006110 | 267948008 | 400242010 |
| Reopening of abdomen and reexploration of intra-abdominal operation site and surgical arrest of postoperative bleeding | 4680671000006110 | 177956008 | 2164235018 |
| Intramuscular hematoma | 5459481000006117 | 262969000 | 391071013 |
| Chronic haematoma of pinna | 2950691000006112 | 28072004 | 1216250013 |
| Bleeding hemorrhoids | 3335281000006117 | 51551000 | 85856015 |
| Painful haematuria | 304348015 | 197939009 | 304348015 |
| Breast haematoma due to non-traumatic cause | 525281000006114 | 198123000 | 304630018 |
| Extradural h'ge inj no open intracra wnd+LOC unspec duration | 660031000006110 | 262949005 | 391026017 |
| Peri-operative hemorrhage or hematoma | 5511401000006112 | 269302003 | 403112018 |
| Evacuation of extradural haematoma | 649541000006113 | 171713001 | 265968017 |
| [X]Haemorrhage from other sites in respiratory passages | 12469521000006114 | 397271000000104 | 388341000006114 |
| Postoperative control of haemorrhage of adenoids | 12705111000006117 | 39508008 | 492111018 |
| Flame-shaped hemorrhage | 12222151000006110 | 247132001 | 3644831017 |
| Subconjunctival haemorrhage | 503913016 | 78768009 | 503913016 |
| Liver haematoma and contusion without open wound into cavity | 11989661000006117 | 262796008 | 11989661000006117 |
| Thyroid hemorrhage | 2547021000006110 | 3002002 | 1217810012 |
| Haemarthrosis of the upper arm | 812911000006112 | 202405009 | 310697017 |
| Retinal dot haemorrhage | 5239801000006114 | 247133006 | 1772574016 |
| Subchorionic haematoma | 7494711000006110 | 609204004 | 2958468013 |
| Ventricular haemorrhage | 2871241000006118 | 23276006 | 481581018 |
| Vaginal bleeding | 1783697014 | 289530006 | 2983956017 |
| Bleeding from mouth | 2858321000006110 | 22490002 | 37748013 |
| Subchondral haematoma | 5651931000006113 | 281464006 | 419472015 |
| Epistaxis from Little's area | 5035181000006118 | 232354002 | 348115017 |
| Pneumohemothorax | 2763831000006117 | 16632002 | 28152010 |
| Hyphemia | 3723191000006114 | 75229002 | 124941011 |
| Intra-alveolar hemorrhage | 3771021000006114 | 78144005 | 129685012 |
| Hemarthrosis of elbow | 4813261000006116 | 202405009 | 310697017 |
| Abnormal uterine and vaginal bleeding, unspecified | 456809019 | 312984006 | 456806014 |
| Extradural hemorrhage after injury | 5459171000006114 | 262949005 | 391027014 |

| | | | |
|---|---|---|---|
| Thyroid haemorrhage or infarction NOS | 292445017 | 190305000 | 292443012 |
| Corneal hemorrhage | 5029231000006113 | 231916005 | 347515010 |
| Haemarthrosis of interphalangeal joint of toe | 812701000006111 | 202421004 | 310736015 |
| O/E - subconjunctival haemorrhage | 270631000006114 | 162815002 | 253783017 |
| Hemarthrosis of tibiofibular joint | 4813551000006116 | 202414004 | 310722013 |
| Haemorrhage into cornea | 5029221000006110 | 231916005 | 347514014 |
| Superficial hematoma | 5518221000006114 | 270911002 | 405390015 |
| Renal haematoma without mention of open wound into cavity | 176901000006111 | 262891006 | 390956018 |
| Haemarthrosis of shoulder region | 812901000006114 | 2602008 | 483092019 |
| Haemarthrosis of tibio-fibular joint | 812921000006116 | 202414004 | 310721018 |
| Epistaxis control by cryosurgery | 5724341000006114 | 287393001 | 426925010 |
| Hemophthalmos without current injury | 4771071000006113 | 193287005 | 2547719019 |
| O/E - retinal haemorrhages | 255337011 | 163987005 | 255337011 |
| Splenic hematoma | 5457471000006113 | 262818004 | 390851010 |
| Abnormal uterine bleeding | 456807017 | 44991000119100 | 2995739016 |
| Intraoperative hemorrhage | 4872551000006117 | 213261000 | 325135014 |
| Bleeding after intercourse | 3290921000006119 | 48880000 | 1230661013 |
| Male genital haemorrhage NOS | 396748016 | 198057005 | 304524013 |
| Recurrent haematuria co-occurrent and due to diffuse endocapillary proliferative glomerulonephritis | 189161000006113 | 714817004 | 3300532016 |
| Excessive bleeding at onset of menopause | 3936961000006118 | 88424000 | 146605017 |
| Bleeding from ear | 2921411000006111 | 26322001 | 483174013 |
| Hemorrhage | 4454751000006112 | 131148009 | 3035867011 |
| Extradural h'ge inj no open intracran wnd+concussion unspec | 660071000006113 | 262949005 | 391026017 |
| Urethral meatal bleeding during urinary catheterisation | 1938681000006112 | 1938681000006108 | 1938681000006112 |
| Wound haematoma | 5131541000006113 | 239160006 | 358450014 |
| Arterial haemorrhage | 5058751000006119 | 234003006 | 350613018 |
| Delayed/excessive haemorrhage | 887711000006114 | 198827002 | 887711000006114 |
| Drainage of haematoma of external ear and control cavity with bolster suture | 628481000006113 | 172619007 | 267230014 |
| Scrotal hematoma | 3962431000006110 | 89966002 | 149129014 |
| Hemorrhage control | 3330371000006116 | 51241000 | 1490495018 |
| Drainage of subungual haematoma | 508104019 | 88584000 | 508104019 |
| Evacuation of vulval hematoma | 3568171000006111 | 65687007 | 1218318011 |
| Heavy episode of vaginal bleeding | 459713018 | 315224006 | 459713018 |
| Bladder hemorrhage | 4790081000006111 | 197887003 | 304254016 |
| Haemorrhage | 300840014 | 131148009 | 3035879011 |
| Recurrent haematuria | 303986013 | 281860005 | 419991013 |
| Optic nerve hemorrhage | 2729091000006116 | 14460007 | 24580011 |
| Pharyngeal haemorrhage | 6119801000006119 | 324618004 | 6119801000006119 |
| Haematoma - perineal wound | 813251000006111 | 371610007 | 1207307018 |
| Haemarthrosis of ankle | 812651000006117 | 202415003 | 310724014 |

| | | | |
|---|---|---|---|
| Retinal pigment epithelium hemorrhagic detachment | 3312521000006118 | 50165004 | 1218183019 |
| Drainage of haematoma of external ear | 267223015 | 172617009 | 267223015 |
| Haematuria | 57480016 | 34436003 | 57480016 |
| Chronic hematoma of pinna | 2950701000006112 | 28072004 | 1217750018 |
| Epidural intracranial haemorrhage | 3850551000006115 | 82999001 | 2915908018 |
| Haemorrhage into bladder wall | 478383018 | 17615006 | 478383018 |
| Subungual Haematoma | 1576441000006114 | 1576441000006105 | 1576441000006114 |
| Recurrent haematuria co-occurrent and due to diffuse membranous glomerulonephritis | 189141000006114 | 714821006 | 3300544018 |
| Retroperitoneal haematoma | 353768014 | 236002003 | 353768014 |
| On examination - epistaxis | 255582011 | 164187003 | 2667170012 |
| Epistaxis control | 860691000006114 | 35807001 | 860691000006114 |
| Bladder haemorrhage | 304253010 | 197887003 | 304253010 |
| Submembranous retinal haemorrhage | 2683861000006117 | 11547003 | 411995018 |
| Nontraumatic extradural haemorrhage | 660111000006117 | 397809001 | 1773153015 |
| Traumatic extradural haematoma without open intracranial wound | 86021000006119 | 315048006 | 459515010 |
| Prostatic hemorrhage | 3223431000006110 | 44843000 | 1218121019 |
| Gingival bleeding | 3902861000006119 | 86276007 | 143081017 |
| Intrarenal haematoma | 304143019 | 197813005 | 304143019 |
| Clot haematuria | 304355018 | 197942003 | 304355018 |
| Post-op. haemorrhage control | 878521000006118 | 274508002 | 878521000006118 |
| Haemoperitoneum - non-traumatic | 813971000006111 | 45626005 | 494282011 |
| H/O: postmenopausal bleeding | 811301000006115 | 161788002 | 252102013 |
| Haemarthrosis of the pelvic region and thigh | 812891000006110 | 202411007 | 310713012 |
| Postoperative control of haemorrhage of adenoids | 1543141000006110 | 39508008 | 66269014 |
| Bulbar haemorrhage | 483988011 | 732923001 | 3467313018 |
| Haemarthrosis - shoulder joint | 989471000006110 | 2602008 | 989471000006110 |
| Spleen injury with haematoma without rupture of capsule, with open wound into cavity | 135621000006117 | 210190001 | 321114016 |
| Haemorrhage of dialysis arteriovenous graft | 2240831000000118 | 866991000000105 | 2240831000000118 |
| Recurrent haematuria co-occurrent and due to diffuse mesangial proliferative glomerulonephritis | 189171000006118 | 714825002 | 3300556017 |
| Expulsive choroidal haemorrhage | 420970014 | 11623000 | 420970014 |
| Drainage of breast haematoma | 5106761000006119 | 237402002 | 355796018 |
| Acute haematoma of pinna | 5912831000006117 | 302906000 | 444900012 |
| Hemarthrosis of sternoclavicular joint | 4813221000006110 | 202403002 | 310693018 |
| Haemorrhage of scrotum | 3510201000006112 | 62182006 | 499223018 |
| Retinal haemorrhage | 297864014 | 28998008 | 484055013 |
| Bleeding from the ear | 2921431000006117 | 26322001 | 44085018 |
| Hematoma of rectus sheath | 5975711000006116 | 308154003 | 451472016 |
| Bleeding - vaginal NOS | 371562017 | 289530006 | 2983956017 |
| Haemarthrosis - elbow joint | 989481000006113 | 202405009 | 989481000006113 |
| Haemorrhage in uterus | 3116591000006113 | 38280009 | 489888015 |

| | | | |
|---|---|---|---|
| Finding of bleeding of nose | 5270651000006118 | 249366005 | 1224241013 |
| Epidural intracranial hemorrhage | 3850561000006118 | 82999001 | 2916559017 |
| Evacuation of haematoma | 279096018 | 118441006 | 444575012 |
| Choroidal hemorrhage and rupture | 4771941000006116 | 193474002 | 297941013 |
| Intrarenal hematoma | 4789541000006116 | 197813005 | 304144013 |
| Breast hematoma due to non-traumatic cause | 4791271000006114 | 198123000 | 304631019 |
| Haemopericardium as current complication following acute myocardial infarction | 813961000006116 | 194862000 | 299816014 |
| Packing for bleeding | 4716041000006116 | 182605004 | 282263017 |
| Nosebleed/epistaxis symptom | 297191000006112 | 249366005 | 372073016 |
| O/E - vitreous haemorrhages | 255341010 | 163990004 | 255341010 |
| Acute posthemorrhagic anemia | 5500911000006114 | 267530009 | 399227018 |
| Anemia due to acute blood loss | 5500941000006113 | 267530009 | 2536037015 |
| Haemarthrosis of the ankle and foot | 400243017 | 267948008 | 400243017 |
| Epistaxis control - cautery | 860711000006112 | 287382003 | 860711000006112 |
| Retrobulbar haemorrhage | 298916015 | 194179009 | 298916015 |
| Extradural h'ge inj no open intracran wnd+1-24hr loss consc | 660061000006118 | 43216008 | 72101019 |
| Unspecified choroidal haemorrhage | 11989321000006119 | 122003 | 11989321000006119 |
| Postmenopausal bleeding | 127423017 | 76742009 | 127423017 |
| Epistaxis control - suture | 5724381000006115 | 287400009 | 426932018 |
| Hematoma AND contusion of liver with open wound into abdominal cavity | 2843231000006111 | 21580006 | 36197018 |
| Pulmonary haemorrhage | 317409016 | 78144005 | 503767018 |
| Control of epistaxis | 3076651000006115 | 35807001 | 59731016 |
| Drainage of haematoma of nasal septum | 2753321000006117 | 15962000 | 477593011 |
| Epistaxis control NOS | 860741000006111 | 35807001 | 860741000006111 |
| Pericardial haematoma evacuat. | 865621000006117 | 562891000000104 | 865621000006117 |
| Subperiosteal haematoma | 6552151000006115 | 395183000 | 1476344017 |
| Spontaneous haemarthrosis | 5650711000006114 | 281370001 | 419361012 |
| Epistaxis control | 5724171000006115 | 287374003 | 426905011 |
| Persistent frank haematuria | 5656791000006118 | 281855006 | 419981019 |
| Subcapsular haematoma of spleen | 5065651000006112 | 234507003 | 351359017 |

#### Table S39. Bleeding at other anatomy sites HES (ICD10) codes

| ICD10 codes | Term |
|---|---|
| D62 | Acute posthaemorrhagic anaemia |
| D68.3 | Haemorrhagic disorder due to circulating anticoagulants |
| D69.8 | Other specified haemorrhagic conditions |
| D69.9 | Haemorrhagic condition, unspecified |
| H11.3 | Conjunctival haemorrhage |
| H21.0 | Hyphema |
| H31.3 | Choroidal haemorrhage and rupture |
| H35.6 | Retinal haemorrhage |
| H43.1 | Vitreous haemorrhage |
| H45.0 | Vitreous haemorrhage in diseases classified elsewhere |
| H92.2 | Otorrhagia |
| I23.0 | Haemopericardium as current complication following acute myocardial infarction |
| I31.2 | Haemopericardium, not elsewhere classified |
| I71.1 | Thoracic aortic aneurysm, ruptured |
| I71.3 | Abdominal aortic aneurysm, ruptured |
| I71.5 | Thoracoabdominal aortic aneurysm, ruptured |
| J94.2 | Haemothorax |
| K76.2 | Central haemorrhagic necrosis of liver |
| K92.0 | Haematemesis |
| K92.1 | Melaena |
| M25.0 | Haemarthrosis |
| M25.00 | Haemarthrosis, unspecified joint |
| M25.01 | Haemarthrosis, shoulder |
| M25.02 | Haemarthrosis, elbow |
| M25.03 | Haemarthrosis, wrist |
| M25.04 | Haemarthrosis, hand |
| M25.05 | Haemarthrosis, hip |
| M25.06 | Haemarthrosis, knee |
| M25.07 | Haemarthrosis, ankle and foot |
| M25.08 | Haemarthrosis, other specified site |
| N02.0 | Recurrent and persistent haematuria. Minor glomerular abnormality |
| N02.1 | Recurrent and persistent haematuria. Focal and segmental glomerular lesions |
| N02.2 | Recurrent and persistent haematuria. Diffuse membranous glomerulonephritis |
| N02.3 | Recurrent and persistent haematuria. Diffuse mesangial proliferative glomerulonephritis |
| N02.4 | Recurrent and persistent haematuria. Diffuse endocapillary proliferative glomerulonephritis |
| N02.5 | Recurrent and persistent haematuria. Diffuse mesangiocapillary glomerulonephritis |
| N02.6 | Recurrent and persistent haematuria. Dense deposit disease |

| | |
|---|---|
| N02.7 | Recurrent and persistent haematuria. Diffuse crescentic glomerulonephritis |
| N02.8 | Recurrent and persistent haematuria. Other |
| N02.9 | Recurrent and persistent haematuria. Unspecified |
| N42.1 | Congestion and haemorrhage of prostate |
| N83.6 | Hematosalpinx |
| N83.7 | Haematoma of broad ligament |
| N85.7 | Hematometra |
| N89.7 | Hematocolpos |
| N93.0 | Postcoital and contact bleeding |
| N93.8 | Other specified abnormal uterine and vaginal bleeding |
| N93.9 | Abnormal uterine and vaginal bleeding, unspecified |
| N95.0 | Postmenopausal bleeding |
| R04.0 | Epistaxis |
| R04.1 | Haemorrhage from throat |
| R04.2 | Haemoptysis |
| R04.8 | Haemorrhage from other sites in respiratory passages |
| R04.9 | Haemorrhage from respiratory passages, unspecified |
| R31 | Unspecified haematuria |
| R58 | Haemorrhage, not elsewhere classified |
| S26.0 | Injury of heart with haemopericardium |
| T81.0 | Haemorrhage and haematoma complicating a procedure, not elsewhere classified |

#### Table S40. Hospitalisation Aurum codes

| Term | Medcode ID | SNOMED CT Concept ID | SNOMED CT Description ID |
|---|---|---|---|
| Admit geriatric emergency | 12482071000006111 | 183457004 | 283525018 |
| Admission to colorectal surgery department | 5940241000006118 | 305422004 | 447841011 |
| Admission by surgeon | 5938551000006116 | 305293001 | 447669018 |
| Night hospital care | 283555013 | 183483009 | 283555013 |
| Refer to hospital casualty | 283795013 | 183659006 | 283795013 |
| Admission to medical department | 5939401000006112 | 305354007 | 447756010 |
| Admission to ophthalmology department | 5940281000006112 | 305426001 | 447845019 |
| Rheumatology emergency hospital admission | 283536013 | 183465001 | 3082921015 |
| Discharge from Inpatient Care | 8049681000006116 | 163391000000107 | 214171000000113 |
| Emergency hospital admission to general surgical service | 8453071000006119 | 1078041000000108 | 2702931000000114 |
| Patient in hospital | 284397011 | 184091000 | 284397011 |
| Patient died in hospital | 284637019 | 184297005 | 284637019 |
| Refer to hospital | 2476476011 | 310449005 | 2476476011 |
| Discharged from accident and emergency | 2549670013 | 417119002 | 2549670013 |
| Discharge summary awaited | 21521000000115 | 17301000000103 | 21521000000115 |
| Admit to intensive care unit | 447747019 | 305351004 | 447747019 |
| Admission by assistant GP | 967731000000117 | 449201000000100 | 967731000000117 |
| Hospital admission, short-term | 3321091000006111 | 50699000 | 84496015 |
| Place to which Pt admitted (Gen Surg) | 1594641000006111 | 1594641000006107 | 1594641000006111 |
| Hospital admission, transfer from other hospital or health care facility | 2571671000006112 | 4563007 | 7859016 |
| Inpatient stay 6 days | 783711000006116 | 183804009 | 284001011 |
| Admission by locum general practitioner | 1823591000006110 | 476981000000106 | 1690371000000119 |
| Hospital inpatient note | 1898071000006117 | 1898071000006101 | 1898071000006117 |
| Discharge to rented accommodation | 1951041000006116 | 1951041000006100 | 1951041000006116 |
| Hospital Inpatient | 1563381000006113 | 1563381000006109 | 1563381000006113 |
| Accident and Emergency department discharge to neonatal intensive care unit | 8439411000006116 | 1066401000000108 | 2680731000000115 |
| Place to which Pt admitted (Ophth) | 1598011000006110 | 1598011000006106 | 1598011000006110 |
| Hematology emergency hospital admission | 4723611000006111 | 183470008 | 3082879019 |
| Vascular surgery emergency hospital admission | 7517441000006118 | 699122005 | 7517441000006118 |
| Admission by co-operative general practitioner | 8129421000006110 | 388751000000101 | 8129421000006110 |
| Discharged from private hospital | 4725511000006111 | 183670004 | 4725511000006111 |
| Emergency department discharge to ICU (intensive care unit) | 8439381000006119 | 1066391000000105 | 2678241000000110 |
| Post hospital discharge followed up within 3 days | 2391861000000116 | 933601000000101 | 2391861000000116 |
| Admit trauma emergency | 12482281000006119 | 183463008 | 283534011 |
| Post-operative assessment at 4-6 hours after secondary care discharge | 1628141000006112 | 298941000000105 | 528521000000113 |
| Admission by deputising general practitioner | 1845391000006119 | 395581000000108 | 962331000000116 |
| Ophthalmological emergency hospital admission | 4723471000006117 | 183464002 | 4723471000006117 |
| Admit cardiology emergency | 12482021000006110 | 313385005 | 457274011 |

| | | | |
|---|---|---|---|
| Discharged - treatment completed | 968851000006112 | 968851000006108 | 968851000006112 |
| [RFC] Contact after hospital discharge/ A&E visit | 910381000006112 | 910381000006108 | 910381000006112 |
| Refer to hospital eye casualty | 184781000006115 | 183661002 | 283798010 |
| Death in hospital | 608281000006119 | 183676005 | 283820019 |
| Admission to accident and emergency department | 1230914014 | 50849002 | 1230914014 |
| Admission to neurological intensive care unit | 283516012 | 183448005 | 2900917012 |
| Cardiothoracic emergency hospital admission | 283549015 | 183477006 | 3082969017 |
| Inpatient stay 5 days | 783701000006119 | 183803003 | 284000012 |
| Admission by orthopaedic surgeon | 5938821000006111 | 305314005 | 447700011 |
| Admit other unit | 949781000006113 | 949781000006109 | 949781000006113 |
| Emergency hospital admission to general medical service | 8453341000006112 | 1078291000000109 | 2703431000000113 |
| Admission to ward | 5939171000006112 | 305342007 | 447733018 |
| Discharge from hospital | 5976961000006114 | 308283009 | 451609018 |
| Admission to gastroenterology department | 5939531000006117 | 305367005 | 447769014 |
| Facilitated early discharge from hospital | 726311000000112 | 343961000000102 | 726311000000112 |
| Admission by ophthalmologist | 5938791000006119 | 305312009 | 447696015 |
| Hospital inpatient report | 62131000000111 | 24661000000108 | 62131000000111 |
| Discharge from intermediate care | 62671000000115 | 25131000000105 | 62671000000115 |
| Urology emergency hospital admission | 4723551000006111 | 183468004 | 4723551000006111 |
| Admission to gastrointestinal surgery department | 5940211000006117 | 305419001 | 447838019 |
| Admission by casualty doctor | 5937711000006113 | 305226003 | 447581010 |
| Discharge to hospital at home service | 8439271000006114 | 1066351000000102 | 2678131000000114 |
| Admit vascular surgery emergency | 12491011000006112 | 699122005 | 2982822015 |
| Admission to respiratory intensive care unit | 4723181000006114 | 183447000 | 4723181000006114 |
| Date Pt admitted (Musc/Skel) | 1596111000006113 | 1596111000006109 | 1596111000006113 |
| Place to which Pt admitted (Musc/Skel) | 1596121000006117 | 1596121000006101 | 1596121000006117 |
| Renal medicine emergency hospital admission | 4723701000006118 | 183475003 | 4723701000006118 |
| Emergency department discharge to ambulatory emergency care service | 8439251000006116 | 1066341000000100 | 2678111000000118 |
| Emergency room admission | 3323561000006114 | 50849002 | 84735018 |
| Discharge letter faxed to general practitioner | 1756601000006113 | 1756601000006109 | 1756601000006113 |
| [V]Delayed discharge - social services | 1227761013 | 309568009 | 453018012 |
| Discharge to care home | 1951031000006114 | 1951031000006105 | 1951031000006114 |
| Discharge to other establishment | 1951251000006111 | 1951251000006107 | 1951251000006111 |
| Discharge to acute hospital | 1951201000006112 | 1951201000006108 | 1951201000006112 |
| Cardiothoracic emergency hospital admission | 4723741000006116 | 183477006 | 4723741000006116 |
| Acute secondary care intervention on development of symptoms | 2180221000000114 | 840121000000102 | 2180221000000114 |
| Discharge letter given to patient | 1693771000000110 | 763571000000107 | 1693771000000110 |
| Hospital discharge letter received | 2343861000000113 | 911911000000100 | 2343861000000113 |
| Accident and Emergency department discharge to CCU (coronary care unit) | 8439331000006115 | 1066371000000106 | 2680701000000114 |
| Neurology emergency hospital admission | 4723531000006116 | 183467009 | 4723531000006116 |
| Admission by co-op GP | 12491231000006111 | 388751000000101 | 851091000000111 |

| | | | |
|---|---|---|---|
| Admission by accident and emergency doctor | 447580011 | 305226003 | 1775689013 |
| Trauma emergency hospital admission | 4723451000006110 | 183463008 | 4723451000006110 |
| Admission to cardiac intensive care unit | 4723161000006116 | 183446009 | 4723161000006116 |
| Discharge from A & E service | 5957881000006110 | 306563004 | 1785121019 |
| Delayed discharge to nursing home | 283813014 | 183672007 | 283813014 |
| Inpatient care | 1488379018 | 394656005 | 1488379018 |
| Urology emergency hospital admission | 283539018 | 183468004 | 3082876014 |
| Place to which Pt admitted (General Medicine) | 1594091000006112 | 1594091000006108 | 1594091000006112 |
| Long stay hospital inpatient | 736561000006112 | 160741006 | 250569018 |
| Inpatient stay longer than 12 hours | 783601000006114 | 183798007 | 283995016 |
| Place to which Pt admitted (Gastro) | 1593541000006113 | 1593541000006109 | 1593541000006113 |
| Discharge from Accident and Emergency service | 5957861000006117 | 306563004 | 1775696010 |
| Admission to cardiology department | 5939431000006116 | 305357000 | 447759015 |
| Inpatient surgery care | 3524772015 | 736774000 | 3524772015 |
| Discharge from adult intensive care service | 5957951000006111 | 306569000 | 449638012 |
| Emergency hospital admission to gastroenterology service | 8453311000006113 | 1078261000000103 | 2703371000000116 |
| Discharge by Accident and Emergency doctor | 5955681000006119 | 306390007 | 1775695014 |
| Admit haematology emergency | 12482091000006112 | 183470008 | 283541017 |
| Emergency hospital admission to thoracic surgery service | 8453021000006115 | 1077991000000101 | 2702801000000111 |
| Discharge to day ward | 5959671000006119 | 306707002 | 449818014 |
| Discharged from inpatient care | 283805013 | 183667003 | 283805013 |
| Emergency hospital admission to accident and emergency service | 8458471000006117 | 1082421000000101 | 2712531000000116 |
| Neurology emergency hospital admission | 283538014 | 183467009 | 3082917010 |
| Admission by ear, nose and throat surgeon | 5938651000006115 | 305302002 | 447682014 |
| Inpatient Care and Treatment Review | 8433711000006113 | 1060741000000104 | 2666481000000111 |
| Hospital inpatient | 397755014 | 266938001 | 397755014 |
| Admit to respiratory ITU | 12482271000006117 | 183447000 | 283515011 |
| Admission by urologist | 5938901000006112 | 305320006 | 447707014 |
| 24 hours post admission | 5650851000006116 | 281380002 | 419373012 |
| Admission to neurological intensive care unit | 4723201000006110 | 183448005 | 4723201000006110 |
| Refer to hospital casualty NOS | 283801016 | 183659006 | 283795013 |
| Accident and Emergency department discharge to SCBU (special care baby unit) | 8439361000006112 | 1066381000000108 | 2680741000000112 |
| Vascular surgery emergency hospital admission | 1137891000000117 | 699122005 | 3082834018 |
| Admission with abdominal symptoms | 1838001000006117 | 1838001000006101 | 1838001000006117 |
| Date admit other unit | 949961000006117 | 949961000006101 | 949961000006117 |
| DOA (dead on arrival) at hospital | 3528421000006114 | 63238001 | 2121181000000118 |
| Discharged from community hospital | 1562411000000110 | 712671000000101 | 1562411000000110 |
| Admit hospital emergency NOS | 283552011 | 183452005 | 283520011 |
| Admission by general practitioner | 5937781000006118 | 305230000 | 2692897015 |
| Post-operative assessment at 12-24 hours after secondary care discharge | 1627141000006113 | 298971000000104 | 528581000000114 |

| | | | |
|---|---|---|---|
| Discharged from hospital | 283803018 | 183665006 | 283803018 |
| Post-operative assessment at 6 weeks after secondary care discharge | 1768661000006118 | 760301000000104 | 1681501000000110 |
| Post hospital discharge medication reconciliation with medical notes | 1753671000006114 | 730061000000107 | 1601761000000119 |
| Appropriate non-elective hospital admission | 1955941000006117 | 1955941000006101 | 1955941000006117 |
| Accident and Emergency department discharge to ICU (intensive care unit) | 8439391000006116 | 1066391000000105 | 2680721000000117 |
| Admission to hospital | 1227951014 | 32485007 | 1227951014 |
| Discharge letter emailed to general practitioner | 1706631000000118 | 766251000000108 | 1706631000000118 |
| Inpatient stay 3 days | 783681000006117 | 183801001 | 283998019 |
| Discharge letter sent to GP (general practitioner) | 8090731000006111 | 294331000000109 | 1676661000000111 |
| Geriatric emergency hospital admission | 283525018 | 183457004 | 3082885014 |
| Admission to cardiac intensive care unit | 283514010 | 183446009 | 2901401018 |
| Inpatient stay 2 days | 783671000006115 | 183800000 | 283997012 |
| Patient self-discharge | 242581000006111 | 183955003 | 284179014 |
| Adjustment reaction due to hospitalisation | 461701000006114 | 192065003 | 295515011 |
| Discharge to private nursing home | 5959551000006111 | 306695007 | 449802015 |
| Trauma emergency hospital admission | 283534011 | 183463008 | 3082906014 |
| Renal medicine emergency hospital admission | 283547018 | 183475003 | 3082929018 |
| Admit to intensive c.u. NOS | 283519017 | 305351004 | 447746011 |
| Discharge to tertiary referral hospital | 5959621000006115 | 306703003 | 449811015 |
| Admission to neurology department | 5939691000006119 | 305379005 | 447786015 |
| Oral surgical emergency hospital admission | 4723661000006114 | 183473005 | 4723661000006114 |
| Discharged from hospital within 6 hours of delivery | 283814015 | 183673002 | 283814015 |
| Admission to department | 5939271000006117 | 305349003 | 447743015 |
| Cardiology emergency hospital admission | 457274011 | 313385005 | 3082972012 |
| Inpatient stay 4 days | 783691000006119 | 183802008 | 283999010 |
| Admission to stroke unit | 449935012 | 306803007 | 449935012 |
| Admission to general medical department | 5939541000006110 | 305368000 | 447770010 |
| Discharge report | 63261000000118 | 25571000000103 | 63261000000118 |
| Admission by GP registrar | 851111000000118 | 424211000000104 | 851111000000118 |
| Received hospital death discharge letter | 193281000006110 | 184277000 | 284626015 |
| Ophthalmological emergency hospital admission | 462221000006116 | 183464002 | 3082894015 |
| Discharge to ward | 5959661000006114 | 306706006 | 449817016 |
| Emergency department discharge to high dependency unit | 8439281000006112 | 1066361000000104 | 2678161000000116 |
| Admission by A & E doctor | 5937721000006117 | 305226003 | 447582015 |
| Patient transfer, in-hospital, service-to-service | 1846191000006113 | 57976004 | 96383017 |
| Discharged from private hosp'l | 12484901000006111 | 183670004 | 283811011 |
| Admission by own GP | 1823541000006118 | 305231001 | 1823541000006118 |
| Admission by associate GP (general practitioner) | 1823571000006114 | 437171000000107 | 1674951000000110 |
| Admission by assistant GP (general practitioner) | 1823561000006119 | 449201000000100 | 1674411000000118 |
| Patient transfer from hospital to hospital | 1825121000006116 | 239531000000108 | 1825121000006116 |
| Neurosurgical emergency hospital admission | 4723721000006111 | 183476002 | 4723721000006111 |

| | | | |
|---|---|---|---|
| Discharge letter emailed to general practitioner | 1756571000006118 | 1756571000006102 | 1756571000006118 |
| Post hospital discharge medication reconciliation with patient | 1753661000006119 | 730021000000104 | 1601671000000117 |
| Hospital death disch. NOS | 284628019 | 309039003 | 452370013 |
| Hospital death discharge notification | 828231000006117 | 184274007 | 284623011 |
| Admit to ITU | 462351000006119 | 305351004 | 447746011 |
| Admit surgical emergency unsp. | 462321000006111 | 183452005 | 283520011 |
| Admit medical emergency unsp. | 462181000006113 | 183452005 | 283520011 |
| Hospital re-admission | 1782016 | 417005 | 1782016 |
| Admission to respiratory intensive care unit | 283515011 | 183447000 | 2902327017 |
| Haematology emergency hospital admission | 283541017 | 183470008 | 3082962014 |
| Inpatient stay 1 day | 783611000006112 | 183799004 | 283996015 |
| Accident and Emergency department discharge to ambulatory emergency care service | 8439261000006119 | 1066341000000100 | 2680751000000110 |
| Inpatient final discharge letter | 8271641000006116 | 824331000000106 | 2145271000000110 |
| Admission to general gastrointestinal surgery department | 5940221000006113 | 305420007 | 447839010 |
| Taking own discharge | 5957831000006114 | 306560001 | 449625016 |
| Reason for admission | 8310281000006117 | 886861000000108 | 2285241000000112 |
| Admission to adult intensive care unit | 5939341000006118 | 305352006 | 447750016 |
| Admitted other | 950751000006110 | 950751000006106 | 950751000006110 |
| Admission to respiratory medicine department | 5939461000006113 | 305360007 | 447762017 |
| Admit ENT emergency | 12482061000006116 | 183462003 | 283533017 |
| Referral to inpatient unit | 2729321000000110 | 1089301000000106 | 2729321000000110 |
| On admission | 5611821000006112 | 278307001 | 415178019 |
| [V]Delayed discharge - nursing home vacancy awaited | 1227760014 | 183930009 | 284147017 |
| Admission to thoracic medicine department | 5939451000006111 | 305359002 | 447761012 |
| A&E attendance - discharged no follow up treatment required | 1981781000006116 | 1981781000006100 | 1981781000006116 |
| Admission by GP | 447587014 | 305230000 | 447587014 |
| Discharge to community hospital | 449808016 | 306701001 | 449808016 |
| Premature hospital discharge | 1918091000006114 | 1918091000006105 | 1918091000006114 |
| Admit cardiothoracic emergency | 12482031000006113 | 183477006 | 283549015 |
| Date Pt admitted (Cardio) | 1592091000006115 | 1592091000006104 | 1592091000006115 |
| Emergency department discharge to coronary care unit | 8439311000006114 | 1066371000000106 | 2678191000000110 |
| Emergency department discharge to CCU (coronary care unit) | 8439321000006118 | 1066371000000106 | 2678181000000113 |
| Discharged from community hospital | 1744481000006110 | 1744481000006106 | 1744481000006110 |
| Patient transfer from hospital to hospital | 1156751000000119 | 239531000000108 | 1156751000000119 |
| Admission by GP partner | 1823551000006116 | 305232008 | 1823551000006116 |
| Admission by GP (general practitioner) registrar | 1823581000006112 | 424211000000104 | 1675721000000119 |
| Premature hospital discharge | 2444251000000115 | 958801000000109 | 2444251000000115 |
| Discharge letter faxed to general practitioner | 1693731000000113 | 763551000000103 | 1693731000000113 |
| Place to which Pt admitted (Cardio) | 1592101000006114 | 1592101000006105 | 1592101000006114 |
| Date Pt discharged (Gastro) | 1593561000006112 | 1593561000006108 | 1593561000006112 |

| | | | |
|---|---|---|---|
| Date Pt admitted (General Medicine) | 1594081000006114 | 1594081000006105 | 1594081000006114 |
| Admit ophthalmological emergency | 12482131000006114 | 183464002 | 283535012 |
| Admit anticoagulation emergency | 2198591000000112 | 848101000000108 | 2198591000000112 |
| Gynaecological emergency hospital admission | 4723341000006117 | 183459001 | 4723341000006117 |
| Admit neurosurgical emergency | 12482111000006115 | 183476002 | 283548011 |
| Admit to cardiac ITU | 12482241000006113 | 183446009 | 283514010 |
| Admitted | 949761000006115 | 949761000006104 | 949761000006115 |
| Inpatient stay 12 days | 783641000006111 | 183810009 | 284007010 |
| ENT emergency hospital admission | 283533017 | 183462003 | 3082975014 |
| Admission to oral surgery department | 5940291000006110 | 305427005 | 447847010 |
| Orthopaedic emergency hospital admission | 283532010 | 183461005 | 3082948019 |
| Discharge letter | 8270971000006113 | 823701000000103 | 2143921000000116 |
| Emergency hospital admission to stroke service | 8453821000006113 | 1078671000000108 | 2704231000000114 |
| Admission to hepatobiliary surgical department | 5940261000006119 | 305424003 | 447843014 |
| Discharge letter offered to patient | 8306951000006115 | 879761000000102 | 2269661000000111 |
| Emergency department discharge to intensive care unit | 8439371000006117 | 1066391000000105 | 2678251000000113 |
| Discharge summary | 1212385013 | 373942005 | 1212385013 |
| Discharge from intensive care service | 5957941000006114 | 306568008 | 449637019 |
| Admission to gynaecology department | 5939761000006118 | 305385003 | 447793016 |
| Hospital admission, emergency, from emergency room, medical nature | 2816831000006113 | 19951005 | 33588016 |
| Discharge to residential home | 449798016 | 306691003 | 449798016 |
| In-patient admission | 841531000006114 | 841531000006105 | 841531000006114 |
| Admission by GP locum | 851101000000115 | 476981000000106 | 851101000000115 |
| Hospital admission note | 62121000000114 | 24651000000105 | 62121000000114 |
| Admission to emergency department | 3323601000006114 | 50849002 | 2995318014 |
| Intervention - hospital admission | 971111000006119 | 971111000006103 | 971111000006119 |
| Accident and Emergency department discharge to HDU (high dependency unit) | 8439301000006111 | 1066361000000104 | 2680711000000111 |
| Admission to adult ITU | 5939351000006116 | 305352006 | 447751017 |
| Admission by co-operative general practitioner | 851091000000111 | 388751000000101 | 1692241000000117 |
| Emergency department discharge to emergency department short stay ward | 8439231000006111 | 1066331000000109 | 2678091000000111 |
| Date Pt admitted (Oral Medicine and Surgery) | 1597591000006112 | 1597591000006108 | 1597591000006112 |
| Date Pt admitted (Ophth) | 1598001000006112 | 1598001000006108 | 1598001000006112 |
| Inpatient stay 10 days | 783621000006116 | 183808007 | 284005019 |
| Discharge Summary (Full) | 1563121000006112 | 1563121000006108 | 1563121000006112 |
| Admission notification | 2304471000000113 | 895571000000108 | 2304471000000113 |
| Discharged from acute assessment unit | 2172541000000112 | 836721000000102 | 2172541000000112 |
| ENT emergency hospital admission | 4723421000006118 | 183462003 | 4723421000006118 |
| Discharge to nursing home | 449801010 | 306694006 | 449801010 |
| Awaiting hospital death discharge letter | 500831000006114 | 184276009 | 284625016 |
| Ear, nose and throat emergency hospital admission | 4723441000006113 | 183462003 | 3082934019 |
| Reviewed at hospital | 265639016 | 763287008 | 3638423011 |

| | | | |
|---|---|---|---|
| Inpatient discharge summary received | 15051000000111 | 16831000000102 | 15051000000111 |
| Emergency hospital admission to intermediate care service | 8453391000006115 | 1078331000000102 | 2703511000000118 |
| Inpatient stay 7 days | 783721000006112 | 183805005 | 284002016 |
| Inpatient stay | 782791000006115 | 308540004 | 451902013 |
| Transfer of care from hospital | 95461000006112 | 710112000 | 3043107011 |
| Long stay hospital inpatient | 250569018 | 160741006 | 250569018 |
| Admission to high dependency unit | 6598861000006115 | 398162007 | 1777720019 |
| Admission to Inpatient Care | 8101001000006119 | 313071000000104 | 566451000000110 |
| Admission to endocrinology department | 5939521000006115 | 305366001 | 447768018 |
| Admission by doctor | 2476441014 | 305260005 | 2476441014 |
| Discharge to home | 2901225012 | 306689006 | 2901225012 |
| Hospital patient | 397754013 | 266938001 | 397754013 |
| Admission to day ward | 447734012 | 305343002 | 447734012 |
| Hospital admission, emergency, from emergency room, accidental injury | 2665021000006115 | 10378005 | 18064016 |
| Admission by associate GP | 851081000000114 | 437171000000107 | 851081000000114 |
| Admission to private hospital | 5939121000006111 | 305339001 | 447728017 |
| Place to which Pt admitted (Neuro) | 1596671000006113 | 1596671000006109 | 1596671000006113 |
| Admission to cardiac surgery department | 5940121000006115 | 305412005 | 447829016 |
| Admit oral surgical emergency | 12482141000006116 | 183473005 | 283545014 |
| Inpatient stay 11 days | 783631000006118 | 183809004 | 284006018 |
| Discharge letter given to patient | 1756581000006115 | 1756581000006104 | 1756581000006115 |
| Admit neurology emergency | 12482101000006118 | 183467009 | 283538014 |
| Admit to neurological ITU | 12482261000006112 | 183448005 | 283516012 |
| Follow-up appointment offered after hospital discharge | 1882401000006115 | 1882401000006104 | 1882401000006115 |
| ICU - Admission to intensive care unit | 5939331000006111 | 305351004 | 447748012 |
| Date Pt admitted (Urology) | 1599441000006110 | 1599441000006106 | 1599441000006110 |
| Orthopedic emergency hospital admission | 4723411000006114 | 183461005 | 3082890012 |
| Haematology emergency hospital admission | 4723591000006117 | 183470008 | 4723591000006117 |
| Admit urology emergency | 12482291000006116 | 183468004 | 283539018 |
| Emergency department discharge to HDU (high dependency unit) | 8439291000006110 | 1066361000000104 | 2678151000000119 |
| Date Pt admitted (Gastro) | 1593531000006115 | 1593531000006104 | 1593531000006115 |
| Admission by general practitioner partner | 5937821000006112 | 305232008 | 2972101017 |
| Discharge to relative's home | 449797014 | 306690002 | 449797014 |
| Cardiology emergency hospital admission | 6029451000006115 | 313385005 | 6029451000006115 |
| Review at hospital | 265638012 | 763287008 | 3638423011 |
| Discharge letter sent to general practitioner | 517481000000119 | 294331000000109 | 517481000000119 |
| Discharged from private hospital | 283811011 | 183670004 | 3029377018 |
| Seen in hospital ward | 285196018 | 185212007 | 285196018 |
| Accident and Emergency department discharge to Accident and Emergency department short stay ward | 8439241000006118 | 1066331000000109 | 2680761000000113 |
| Death notification from hospital | 608291000006116 | 184275008 | 284624017 |

| | | | |
|---|---|---|---|
| Inpatient stay 9 days | 782781000006118 | 183807002 | 284004015 |
| Emergency hospital admission to endocrinology service | 8453321000006117 | 1078271000000105 | 2703391000000117 |
| Discharge summary awaited | 1676631000006113 | 17301000000103 | 21521000000115 |
| Oral surgical emergency hospital admission | 283545014 | 183473005 | 3082839011 |
| Plastic surgery emergency hospital admission | 283543019 | 183471007 | 3082940014 |
| Neurosurgical emergency hospital admission | 283548011 | 183476002 | 3082967015 |
| Date Pt admitted (Neuro) | 1596661000006118 | 1596661000006102 | 1596661000006118 |
| Hospital admission, emergency, direct | 3779761000006116 | 78680009 | 130558011 |
| Long stay hospital | 3016541000006111 | 32074000 | 1227906014 |
| Admission to orthopaedic department | 5940311000006114 | 305428000 | 447849013 |
| Admission by dental surgeon | 5938601000006119 | 305298005 | 447677019 |
| Admission by own general practitioner | 5937801000006119 | 305231001 | 2972278017 |
| Admission to surgical department | 5940081000006117 | 305408004 | 447825010 |
| Death in hospital | 2769631000006119 | 16983000 | 28745019 |
| Admission to vascular surgery department | 5940401000006119 | 305435008 | 447857011 |
| Emergency hospital admission | 283520011 | 183452005 | 283520011 |
| Died in hospital | 283820019 | 183676005 | 283820019 |
| Seen in hospital casualty | 285194015 | 185210004 | 285194015 |
| Admission for treatment | 5931041000006116 | 304566005 | 446903011 |
| Inpatient | 6897591000006113 | 416800000 | 2549330012 |
| Hospital admission, emergency, from emergency room | 3696791000006119 | 73607007 | 122228018 |
| Admission by own GP | 447588016 | 305231001 | 447588016 |
| Admission by GP partner | 447589012 | 305232008 | 447589012 |
| Discharge to hospital | 449806017 | 306699001 | 449806017 |
| Patient transfer, in-hospital | 3107571000006112 | 37729005 | 62934015 |
| Admission to intensive care unit | 5939301000006115 | 305351004 | 447749016 |
| Discharge summary sent to general practitioner | 8237241000006114 | 763591000000106 | 1693801000000113 |
| Admit orthopaedic emergency | 12482151000006119 | 183461005 | 283532010 |
| Admitted: Hospital | 1583651000006112 | 1583651000006108 | 1583651000006112 |
| Inappropriate non-elective hospital admission | 1955951000006115 | 1955951000006104 | 1955951000006115 |
| Hospital admission, emergency, indirect | 1728541000006114 | 18083007 | 30547015 |
| Admission to acute assessment unit | 2172381000000110 | 836651000000103 | 2172381000000110 |
| Emergency hospital admission from walk-in centre | 2186461000000117 | 842951000000109 | 2186461000000117 |
| Admission to A & E department | 3323591000006118 | 50849002 | 1230916011 |
| Discharge summary report | 63271000000113 | 25581000000101 | 63271000000113 |
| Hospital admission | 283592013 | 32485007 | 54238014 |
| Admission by physician | 5938171000006116 | 305260005 | 447631011 |
| Hospital notified of death | 1780508019 | 401321000 | 1780508019 |
| A&E attendance - admitted to hospital bed/lodged patient | 1981761000006114 | 1981761000006105 | 1981761000006114 |
| Self-referral to hospital | 283832015 | 183687002 | 283832015 |
| Patient transfer, in-hospital, service-to-service | 1824251000006116 | 57976004 | 1824251000006116 |

| | | | |
|---|---|---|---|
| Discharge from Accident and Emergency service with advice for follow up treatment by general practitioner | 8368771000006114 | 989501000000106 | 2514921000000119 |
| Inpatient stay 8 days | 783731000006110 | 183806006 | 284003014 |
| Inpatient stay 14 days | 783661000006110 | 183812001 | 284009013 |
| Inpatient stay 13 days | 783651000006113 | 183811008 | 284008017 |
| Emergency room admission, died in emergency room | 3719361000006111 | 75004002 | 124573013 |
| Listed for Geriatrics admission | 283967016 | 183774000 | 2668801012 |
| Admission to urology department | 5940391000006116 | 305434007 | 447856019 |
| Admission to general surgical department | 5940251000006116 | 305423009 | 447842016 |
| A&E attendance - discharged with follow up treatment by GP | 1981771000006119 | 1981771000006103 | 1981771000006119 |
| Patient died in acute hospital | 1964501000006119 | 1964501000006103 | 1964501000006119 |

#### Table S41. Intracranial haemorrhage Aurum codes

| Term | Medcode ID | SNOMED CT Concept ID | SNOMED CT Description ID |
|---|---|---|---|
| Intrapontine haemorrhage | 2622631000006114 | 7713009 | 503468012 |
| Cerebral hemisphere haemorrhage | 3687811000006118 | 73020009 | 502318010 |
| Traumatic subdural hematoma without open intracranial wound | 6053291000006112 | 315046005 | 459512013 |
| Subarachnoid haemorrhage following injury | 391042012 | 450375008 | 2915309017 |
| Cortical haemorrhage | 495394013 | 49422009 | 495394013 |
| Internal capsule haemorrhage | 496232015 | 52201006 | 496232015 |
| Intracranial haemorrhage | 300298011 | 1386000 | 475553012 |
| Other cerebral haemorrhage following injury NOS | 320897016 | 450418003 | 2916363013 |
| Basal ganglia haemorrhage | 503791000006114 | 195165005 | 300272017 |
| Sequelae of intracerebral haemorrhage | 300407010 | 195241001 | 300407010 |
| Spontaneous subarachnoid haemorrhage | 5518041000006112 | 270907008 | 405380011 |
| Subarachnoid hemorrhage from carotid siphon and bifurcation | 4777721000006118 | 195155004 | 300243018 |
| Cerebellar hemorrhage | 3719851000006116 | 75038005 | 124627016 |
| Cerebral infarction with haemorrhagic transformation | 1573101000006112 | 1573101000006108 | 1573101000006112 |
| Subdural intracranial haemorrhage | 3071681000006119 | 35486000 | 2913218019 |
| Traumatic intracranial subarachnoid haemorrhage | 5459271000006119 | 262955000 | 391048011 |
| Subarachnoid intracranial haemorrhage | 2841161000006110 | 21454007 | 2916299018 |
| Subarachnoid haemorrhage following injury without open intracranial wound | 2950231000006111 | 28048009 | 2950231000006111 |
| Middle meningeal haemmorhage following injury | 702641000006119 | 30400005 | 484576014 |
| Intracerebral haemorrhage with intraventricular haemorrhage | 4777861000006119 | 195168007 | 2915438018 |
| Subdural hemorrhage | 3071661000006112 | 35486000 | 59206018 |
| Epidural hemorrhage | 3850501000006119 | 82999001 | 137667016 |
| Cerebral haemorrhage | 884421000006119 | 274100004 | 884421000006119 |
| Subdural h'ge inj no open intracran wnd+>24hr LOC -restored | 124121000006111 | 209947002 | 320752018 |
| Subdural h'ge inj no open intracranial wound+<1hr loss consc | 124181000006110 | 209947002 | 320752018 |
| Cerebral haemorrhage NOS | 884451000006111 | 700251000000105 | 884451000006111 |
| Traumatic hemorrhage into subarachnoid space of neuraxis | 7377831000006117 | 450375008 | 2915309017 |
| Closed traumatic subdural haemorrhage | 320752018 | 209947002 | 320752018 |
| Subarachnoid haemorrhage from carotid siphon and bifurcation | 300244012 | 195155004 | 300244012 |
| Haematoma of brain | 5898441000006118 | 301764006 | 443223014 |
| SDH - Subdural haematoma | 4056891000006113 | 95453001 | 1217311015 |
| Spontaneous subacute subdural hemorrhage | 8089871000006114 | 291591000119107 | 2959496014 |
| Traumatic subdural intracranial haemorrhage | 4854361000006118 | 209987007 | 2912530015 |
| Subarachnoid hemorrhage | 12761901000006117 | 21454007 | 36011016 |
| Cerebral haemorrhage NOS | 989211000006119 | 274100004 | 989211000006119 |
| Ruptured aneurysm of basilar artery | 5597601000006115 | 277324009 | 413805016 |

| | | | |
|---|---|---|---|
| Subarachnoid haemorrhage from middle cerebral artery aneurysm | 5583021000006113 | 276280009 | 412353018 |
| External capsule haemorrhage | 300276019 | 195167002 | 300276019 |
| Sequelae of intracerebral hemorrhage | 4778351000006113 | 195241001 | 300408017 |
| Subdural hemorrhage following injury | 4854341000006117 | 209987007 | 320838011 |
| Subdural h'ge inj no open intracranial wnd+>24 LOC +recovery | 124171000006112 | 209947002 | 320752018 |
| Intrapontine hemorrhage | 2622641000006116 | 7713009 | 13755017 |
| Subdural haematoma evacuation | 3469891000006117 | 59712006 | 498519016 |
| Other cerebral haemorrhage following injury | 320853015 | 450418003 | 2916363013 |
| Intracranial subarachnoid haemorrhage from vertebral artery | 4777761000006112 | 195160000 | 2916525016 |
| ICH - intracerebral hemorrhage | 12223131000006114 | 274100004 | 3673218019 |
| Intracerebral hemorrhage, multiple localized | 4777891000006110 | 195169004 | 300279014 |
| Intracerebral haemorrhage | 12223101000006118 | 274100004 | 3673215016 |
| Traumatic intracerebral hemorrhage | 7378401000006111 | 450418003 | 2915288012 |
| [X]Other intracerebral haemorrhage | 11919571000006110 | 274100004 | 11919571000006110 |
| Subarachnoid h'ge inj no open intracran wound + 1-24hr LOC | 123421000006117 | 28048009 | 46957015 |
| Subarachnoid hemorrhage from basilar artery aneurysm | 5583111000006118 | 276284000 | 412362016 |
| Ruptured aneurysm of posterior inferior cerebellar artery | 5597611000006117 | 277325005 | 413807012 |
| Thalamic haemorrhage | 5011411000006113 | 230711001 | 345676013 |
| Haemorrhagic cerebral infarction | 5011291000006119 | 230706003 | 345667018 |
| Intracranial haemorrhage following injury without open intracranial wound | 4198341000006113 | 111668007 | 189842012 |
| Pontine hemorrhage | 2622651000006119 | 7713009 | 13756016 |
| Evacuation of subdural haematoma | 3469851000006111 | 59712006 | 498516011 |
| Pontine haemorrhage | 503469016 | 7713009 | 503469016 |
| Non-traumatic subdural hemorrhage | 4777931000006118 | 195176009 | 2469434017 |
| Subdural haemorrhage | 2534198011 | 35486000 | 486181011 |
| Subarachnoid h'ge inj no open intracran wnd+>24 LOC+recovery | 123401000006110 | 28048009 | 46957015 |
| Subarach h'ge inj no open intracran wnd+LOC unspec duration | 123331000006114 | 28048009 | 46957015 |
| Subarach h'ge inj no open intracran wnd + concussion unspec | 123311000006115 | 28048009 | 46957015 |
| Subarachnoid h'ge inj no open intracran wnd+no loss consc | 123411000006113 | 28048009 | 46957015 |
| Subarachnoid haemorrhage following injury without open intracranial wound | 402929011 | 28048009 | 483737017 |
| Cerebral haemorrhage following injury | 12757911000006115 | 82894007 | 505322013 |
| Cerebral hemorrhage following injury | 12757891000006117 | 82894007 | 505319011 |
| Traumatic intracranial hemorrhage | 12757901000006118 | 82894007 | 505320017 |
| Right sided intracerebral haemorrhage, unspecified | 163261000006119 | 195168007 | 300277011 |
| [X]Other subarachnoid haemorrhage | 300936018 | 21454007 | 481028017 |
| Cerebral haemorrhage | 122371000006118 | 274100004 | 409860011 |
| Other and unspecified intracranial haemorrhage | 300290016 | 62914000 | 104563015 |
| Traumatic cerebral haemorrhage | 505324014 | 450418003 | 2916058017 |

| | | | |
|---|---|---|---|
| Cerebral hemorrhage | 605471000006112 | 274100004 | 409859018 |
| Ventricular haemorrhage | 2871241000006118 | 23276006 | 481581018 |
| Subarachnoid haemorrhage due to ruptured aneurysm | 5011581000006119 | 230719004 | 345695018 |
| Cerebral haemorrhage | 300939013 | 274100004 | 409860011 |
| Subdural haemorrhage following injury | 320836010 | 209987007 | 320836010 |
| Hemorrhage of medulla oblongata | 7951281000006118 | 732923001 | 3467313018 |
| Lobar cerebral haemorrhage | 345675012 | 230710000 | 345675012 |
| Ruptured berry aneurysm | 300242011 | 195154000 | 300242011 |
| Sequelae of subarachnoid haemorrhage | 300406018 | 195240000 | 300406018 |
| Subdural h'ge after injury | 991811000006115 | 209987007 | 991811000006115 |
| [X]Intracerebral haemorrhage in hemisphere, unspecified | 300956017 | 274100004 | 409860011 |
| Subdural h'ge inj no open intracranial wnd + unspec consc | 124161000006117 | 209947002 | 320752018 |
| Subdural hemorrhage following injury without open intracranial wound | 4854171000006111 | 209947002 | 2915477015 |
| Pituitary haemorrhage | 5111331000006119 | 237702003 | 356253018 |
| SAH - Subarachnoid hemorrhage | 2841151000006113 | 21454007 | 1217630014 |
| Non-traumatic intracranial subdural haemorrhage | 4777951000006113 | 195176009 | 2913186018 |
| History of subarachnoid haemorrhage | 4540691000006112 | 161515009 | 2986875019 |
| Subdural hemorrhage - nontraumatic | 4777911000006112 | 195176009 | 300293019 |
| Intracerebral hemorrhage with intraventricular hemorrhage | 4777871000006114 | 195168007 | 2916313015 |
| Intracranial hemorrhage following injury | 7378281000006118 | 450410005 | 2915467017 |
| Intracranial haemorrhage following injury | 12716261000006116 | 82894007 | 505321018 |
| Traumatic cranial subdural hematoma | 5459251000006112 | 262952002 | 391039018 |
| Subdural intracranial hemorrhage | 3071671000006117 | 35486000 | 2912630010 |
| Intracerebral haemorrhage NOS | 300287010 | 274100004 | 2819959010 |
| Subdural h'ge inj no open intracranial wound+no loss consc | 124191000006113 | 209947002 | 320752018 |
| Traumatic subdural hematoma | 391035012 | 262952002 | 2912898019 |
| Cerebral hemorrhage | 13031191000006112 | 274100004 | 122371000006118 |
| Subdural hematoma | 4056881000006110 | 95453001 | 158107014 |
| Ruptured cerebral arteriovenous malformation | 5597281000006110 | 277299009 | 413775010 |
| Intracerebral haemorrhage, multiple localised | 4777881000006112 | 195169004 | 300280012 |
| Closed traumatic subdural hemorrhage | 4854141000006115 | 209947002 | 320753011 |
| H/O subarachnoid hemorrhage | 4540671000006111 | 161515009 | 251700011 |
| Subdural hematoma evacuation | 3469881000006115 | 59712006 | 498518012 |
| Traumatic cerebral hemorrhage | 7378441000006113 | 450418003 | 2916150010 |
| Intraventricular haemorrhage | 2871271000006114 | 23276006 | 481580017 |
| Subarach h'ge inj no open intracran wnd+>24hrs LOC-restored | 123321000006111 | 28048009 | 46957015 |
| Traumatic subdural haemorrhage | 320835014 | 209987007 | 320835014 |
| Subdural haemorrhage - nontraumatic | 300294013 | 195176009 | 300294013 |
| Nontraumatic intracerebral haemorrhage | 8089851000006116 | 291571000119106 | 3331108011 |
| Intraventricular hemorrhage | 2871261000006119 | 23276006 | 39068015 |

| | | | |
|---|---|---|---|
| SDH - Subdural hematoma | 4056901000006112 | 95453001 | 1218799016 |
| Brain stem haemorrhage | 4056931000006116 | 95454007 | 512138014 |
| Traumatic cranial subarachnoid hemorrhage | 5459281000006116 | 262955000 | 391046010 |
| Traumatic subdural intracranial hemorrhage | 4854351000006115 | 209987007 | 2912507014 |
| Subarachnoid hemorrhage from any perinatal cause | 2836991000006113 | 21202004 | 35631013 |
| Traumatic cranial subarachnoid haemorrhage | 5459301000006117 | 262955000 | 391049015 |
| Cerebral haemorrhage | 989201000006117 | 274100004 | 989201000006117 |
| Cerebral haemorrhage following injury NOS | 320852013 | 450418003 | 2916363013 |
| Evacuation of subdural hematoma | 3469861000006113 | 59712006 | 99196013 |
| Subarachnoid haemorrhage from basilar artery aneurysm | 5583101000006116 | 276284000 | 412361011 |
| Cerebral haemorrhage following injury | 505322013 | 450418003 | 2916363013 |
| Traumatic intracerebral haemorrhage | 7378421000006118 | 450418003 | 2915664016 |
| External capsule hemorrhage | 4777831000006111 | 195167002 | 300275015 |
| Haemorrhage of medulla oblongata | 7951271000006116 | 732923001 | 3467314012 |
| Cerebral hemorrhage following injury | 7378411000006114 | 450418003 | 2915447014 |
| Intracranial hemorrhage | 2520541000006112 | 1386000 | 3421016 |
| Traumatic intracranial haemorrhage | 12757921000006111 | 82894007 | 505323015 |
| Cerebral h'ge after injury | 896531000006110 | 82894007 | 896531000006110 |
| Subarachnoid haemorrhage NOS | 300257016 | 21454007 | 481028017 |
| Bulbar haemorrhage | 483988011 | 732923001 | 3467313018 |
| Traumatic cerebral haemorrhage | 12757931000006114 | 82894007 | 505324014 |
| Sequelae of other nontraumatic intracranial haemorrhage | 149551000006111 | 363302008 | 482447013 |
| Intracerebral haemorrhage in hemisphere, unspecified | 744921000006116 | 274100004 | 409860011 |
| Thalamic hemorrhage | 5011421000006117 | 230711001 | 345677016 |
| Sequelae of subarachnoid hemorrhage | 4778331000006118 | 195240000 | 300405019 |
| Subdural haemorrhage | 884441000006114 | 195176009 | 884441000006114 |
| Cerebral hemorrhage | 12762021000006116 | 274100004 | 605471000006112 |
| Traumatic intracerebral haemorrhage | 12757941000006116 | 82894007 | 2475153011 |
| Stroke due to intracerebral haemorrhage | 11903571000006110 | 274100004 | 11903571000006110 |
| Subarachnoid hemorrhage | 428181000006115 | 21454007 | 481028017 |
| Subarachnoid haemorrhage from anterior communicating artery aneurysm | 5583061000006119 | 276282001 | 412357017 |
| Traumatic intracranial subarachnoid hemorrhage | 5459291000006118 | 262955000 | 391047018 |
| ICH - intracerebral haemorrhage | 12223121000006111 | 274100004 | 3673217012 |
| Traumatic intracranial subdural hematoma | 5459241000006110 | 262952002 | 391038014 |
| Intracerebral hemorrhage | 12223111000006115 | 274100004 | 3673216015 |
| Traumatic cranial subdural haematoma | 5459221000006115 | 262952002 | 391036013 |
| Lobar cerebral hemorrhage | 5011401000006110 | 230710000 | 345674011 |
| History of subarachnoid hemorrhage | 4540681000006114 | 161515009 | 2986825016 |
| Intracerebral haemorrhage, intraventricular | 300277011 | 195168007 | 300277011 |
| Subdural h'ge inj no open intracran wnd+LOC unspec duration | 124141000006116 | 209947002 | 320752018 |

| | | | |
|---|---|---|---|
| Intracranial subarachnoid hemorrhage from vertebral artery | 4777751000006110 | 195160000 | 2915522014 |
| Intracranial haemorrhage following injury | 7378271000006116 | 450410005 | 2915426010 |
| Subarachnoid hemorrhage from vertebral artery | 4777741000006113 | 195160000 | 300254011 |
| Subarachnoid haemorrhage from posterior communicating artery | 123521000006118 | 21454007 | 36011016 |
| Intracerebral hemorrhage (ICH) | 5560161000006111 | 274100004 | 2819960017 |
| Closed traumatic subdural intracranial hemorrhage | 4854151000006118 | 209947002 | 2912529013 |
| Subarachnoid h'ge inj no open intracran wnd+<1hr loss consc | 123391000006113 | 28048009 | 46957015 |
| Traumatic subdural haematoma without open intracranial wound | 459511018 | 315046005 | 459511018 |
| Non-traumatic intracranial subdural hemorrhage | 4777941000006111 | 195176009 | 2913068019 |
| Intracranial hemorrhage following injury | 12757871000006118 | 82894007 | 137497019 |
| Intracranial haemorrhage NOS | 12729181000006117 | 700251000000105 | 1530681000000111 |
| Evacuation of haematoma from cerebellum | 265690015 | 171474001 | 265690015 |
| Evacuation of hematoma from cerebellum | 4644251000006117 | 171474001 | 265691016 |
| Occipital subdural haematoma | 7090271000006110 | 428561000 | 2694218015 |
| Cerebellar haemorrhage | 502878012 | 75038005 | 502878012 |
| Subarachnoid hemorrhage following injury without open intracranial wound | 11923791000006114 | 28048009 | 46957015 |
| Traumatic subdural haematoma | 5459261000006114 | 262952002 | 2913152010 |
| [X]Subarachnoid haemorrhage from other intracranial arteries | 300935019 | 21454007 | 36011016 |
| Basal ganglia hemorrhage | 4777811000006117 | 195165005 | 300271012 |
| Subdural h'ge inj no open intracran wnd+1-24hr loss consc | 124131000006114 | 209947002 | 320752018 |
| Left sided intracerebral haemorrhage, unspecified | 748941000006115 | 274100004 | 409859018 |
| Subarachnoid haemorrhage from middle cerebral artery | 123511000006114 | 21454007 | 36011016 |
| Intracerebral haemorrhage, multiple localized | 746571000006116 | 195169004 | 300280012 |
| Traumatic cerebral hemorrhage | 12757881000006115 | 82894007 | 137503010 |
| Right sided intracerebral haemorrhage, unspecified | 12722481000006116 | 308128006 | 451441015 |
| Intraparenchymal haematoma of brain | 7134241000006113 | 431266005 | 2768748018 |
| Subarachnoid h'ge inj no open intracran wound + unspec consc | 123431000006119 | 28048009 | 46957015 |
| Subarachnoid haemorrhage from posterior communicating artery aneurysm | 5583081000006112 | 276283006 | 412360012 |
| Inferior cerebellar artery syndrome | 3778011000006112 | 78569004 | 130376017 |
| SAH - Subarachnoid haemorrhage | 2841141000006111 | 21454007 | 1216125018 |
| Ruptured cerebral aneurysm | 5058411000006114 | 233983001 | 350578011 |
| Internal capsule hemorrhage | 3346051000006110 | 52201006 | 86879010 |
| Anterior cerebral circulation haemorrhagic infarction | 5011331000006114 | 230707007 | 345668011 |
| Closed traumatic subdural intracranial haemorrhage | 4854161000006116 | 209947002 | 2912797018 |
| Subcortical hemorrhage | 2832661000006113 | 20908003 | 35158017 |
| Subdural haemorrhage following injury without open intracranial wound | 4854181000006114 | 209947002 | 2915859018 |
| Subdural h'ge inj no open intracran wound+concussion unspec | 124151000006119 | 209947002 | 320752018 |
| Subdural haematoma | 4056871000006112 | 95453001 | 512137016 |

| | | | |
|---|---|---|---|
| Middle meningeal hemorrhage following injury | 2989681000006112 | 30400005 | 50888016 |
| Intracerebral hemorrhage, intraventricular | 4777851000006116 | 195168007 | 300278018 |
| Subarachnoid intracranial hemorrhage | 2841171000006115 | 21454007 | 2916568015 |
| Subarachnoid haemorrhage | 481028017 | 21454007 | 481028017 |
| Non-traumatic subdural haemorrhage | 2475119012 | 195176009 | 2469010019 |
| Traumatic intracranial hemorrhage | 7378301000006119 | 450410005 | 2915928019 |
| H/O subarachnoid haemorrhage | 808111000006110 | 161515009 | 251701010 |
| Subarachnoid haemorrhage from basilar artery | 123491000006115 | 276284000 | 412361011 |
| Traumatic subdural hemorrhage | 4854331000006110 | 209987007 | 320837018 |
| Subarachnoid haemorrhage from anterior communicating artery | 123481000006118 | 21454007 | 36011016 |
| Subarachnoid haemorrh from intracranial artery, unspecif | 123441000006112 | 21454007 | 481028017 |
| Intracerebral haemorrhage (ICH) | 744901000006114 | 274100004 | 2819959010 |
| Traumatic intracerebral hemorrhage | 12757951000006119 | 82894007 | 2475363012 |
| Subarachnoid haemorrhage from vertebral artery | 300253017 | 195160000 | 300253017 |
| Ruptured aneurysm of middle cerebral artery | 5597491000006112 | 277316004 | 413794017 |

#### Table S42. Intracranial haemorrhage HES (ICD10) codes

| ICD10 codes | Term |
|---|---|
| I60.0 | Subarachnoid haemorrhage from carotid siphon and bifurcation |
| I60.1 | Subarachnoid haemorrhage from middle cerebral artery |
| I60.2 | Subarachnoid haemorrhage from anterior communicating artery |
| I60.3 | Subarachnoid haemorrhage from posterior communicating artery |
| I60.4 | Subarachnoid haemorrhage from basilar artery |
| I60.5 | Subarachnoid haemorrhage from vertebral artery |
| I60.6 | Subarachnoid haemorrhage from other intracranial arteries |
| I60.7 | Subarachnoid haemorrhage from intracranial artery, unspecified |
| I60.8 | Other subarachnoid haemorrhage |
| I60.9 | Subarachnoid haemorrhage, unspecified |
| I61.0 | Intracerebral haemorrhage in hemisphere, subcortical |
| I61.1 | Intracerebral haemorrhage in hemisphere, cortical |
| I61.2 | Intracerebral haemorrhage in hemisphere, unspecified |
| I61.3 | Intracerebral haemorrhage in brain stem |
| I61.4 | Intracerebral haemorrhage in cerebellum |
| I61.5 | Intracerebral haemorrhage, intraventricular |
| I61.6 | Intracerebral haemorrhage, multiple localized |
| I61.8 | Other intracerebral haemorrhage |
| I61.9 | Intracerebral haemorrhage, unspecified |
| I62.0 | Subdural haemorrhage (acute)(nontraumatic) |
| I62.1 | Nontraumatic extradural haemorrhage |
| I62.9 | Intracranial haemorrhage (nontraumatic), unspecified |
| S06.4 | Epidural haemorrhage |
| S06.5 | Traumatic subdural haemorrhage |
| S06.6 | Traumatic subarachnoid haemorrhage |
| I69.0 | Sequelae of subarachnoid haemorrhage |
| I69.1 | Sequelae of intracerebral haemorrhage |
| I69.2 | Sequelae of other nontraumatic intracranial haemorrhage |

#### Table S43. Heart failure outcome Aurum codes

| Term | Medcode ID | SNOMED CT Concept ID | SNOMED CT Description ID |
|---|---|---|---|
| Acute left ventricular failure | 300190010 | 195114002 | 300190010 |
| Impaired left ventricular function | 411506018 | 275514001 | 411506018 |
| O/E - pulmonary oedema | 253994013 | 162970000 | 253994013 |
| New York Heart Association Classification - Class II | 2616471011 | 421704003 | 2616471011 |
| Congestive heart failure due to valvular disease | 2675255018 | 426611007 | 2675255018 |
| Left ventricular cardiac dysfunction | 2694523019 | 429589006 | 2694523019 |
| O/E - pulmonary edema | 4555461000006110 | 162970000 | 253993019 |
| CCF - Congestive cardiac failure | 3182551000006115 | 42343007 | 493288018 |
| Chronic pulmonary oedema | 494669012 | 46847001 | 494669012 |
| Pulmonary congestion and hypostasis NOS | 301695010 | 196115007 | 301689014 |
| Admit heart failure emergency | 12490061000006118 | 416683003 | 2549208013 |
| Acute congestive heart failure | 18472010 | 10633002 | 18472010 |
| COCM - Congestive cardiomyopathy | 6615881000006112 | 399020009 | 1786620011 |
| Emergency hospital admission for heart failure | 6895651000006113 | 416683003 | 6895651000006113 |
| Heart failure as a complication of care | 350484012 | 233924009 | 350484012 |
| Hypertensive heart disease NOS with CCF | 741681000006111 | 64715009 | 107545013 |
| Congestive heart failure | 70653017 | 42343007 | 70653017 |
| Other transplantation of heart | 12719481000006116 | 586351000000102 | 1300781000000114 |
| Left ventricular diastolic dysfunction | 1489358014 | 395704004 | 1489358014 |
| Congestive cardiomyopathy | 1778488011 | 399020009 | 1778488011 |
| Cardiac failure | 139482012 | 84114007 | 139482012 |
| Congestive heart disease | 3182531000006110 | 42343007 | 70654011 |
| Right ventricular failure | 1816101000006113 | 367363000 | 490972013 |
| Postoperative pulmonary oedema | 401870011 | 11468004 | 401870011 |
| Right heart failure | 206703015 | 128404006 | 206703015 |
| Heart failure with preserved ejection fraction | 7321121000006119 | 446221000 | 3496968011 |
| Congestive cardiac failure | 3182541000006117 | 42343007 | 493287011 |
| CCM - Congestive cardiomyopathy | 6615871000006114 | 399020009 | 1786619017 |
| Heart failure with preserved ejection fraction | 2227501000000110 | 446221000 | 2227501000000110 |
| Acute pulmonary oedema unspecified | 301741013 | 40541001 | 492666016 |
| Left ventricular failure | 141306010 | 85232009 | 141306010 |
| Chronic congestive heart failure | 147247018 | 88805009 | 147247018 |
| Nonischemic congestive cardiomyopathy | 7964821000006113 | 111000119104 | 2920819015 |
| Acute cardiac pulmonary oedema | 6212521000006115 | 360371003 | 476096017 |
| Heart failure NOS | 395772015 | 84114007 | 139475013 |
| HF - Heart failure | 3868341000006118 | 84114007 | 1234906013 |
| Heart failure with reduced ejection fraction due to heart valve disease | 7573211000006119 | 703276005 | 3008139015 |
| History of heart failure in last year | 5990971000006113 | 309634009 | 2986867013 |
| Cause of Death- Congestive Cardiac Failure | 1576321000006113 | 1576321000006109 | 1576321000006113 |

| | | | |
|---|---|---|---|
| Chronic heart failure | 3283871000006117 | 48447003 | 80720010 |
| Pulmonary edema - acute | 3152731000006113 | 40541001 | 1490485013 |
| New York Heart Association Classification - Class IV | 2616473014 | 422293003 | 2616473014 |
| Pulmonary oedema NOS | 301694014 | 19242006 | 479262018 |
| [D]Cardiorespiratory failure | 317955011 | 410431009 | 2472092014 |
| Chronic left ventricular systolic dysfunction | 7119761000006112 | 430396006 | 2770949016 |
| Impaired left ventricular func | 5573991000006118 | 275514001 | 1495323018 |
| Biventricular failure | 4005301000006110 | 92506005 | 510016018 |
| Hypertensive heart&renal dis wth (congestive) heart failure | 741701000006114 | 194779001 | 299672017 |
| Heart failure due to end stage congenital heart disease | 7475721000006114 | 471880001 | 2950981012 |
| Acute edema of lung | 3152701000006117 | 40541001 | 67601010 |
| Cardiac cirrhosis | 3713561000006119 | 74669004 | 123997016 |
| Left-sided heart failure | 3886061000006119 | 85232009 | 201199018 |
| On examination - pulmonary edema | 4555471000006115 | 162970000 | 2667198010 |
| Heart failure confirmed | 1488804017 | 395105005 | 1488804017 |
| Hyperten heart&renal dis+both(congestv)heart and renal fail | 789941000006117 | 194781004 | 299674016 |
| Acute pulmonary edema | 3152691000006117 | 40541001 | 67598017 |
| Systolic heart failure | 6914191000006115 | 417996009 | 2577903013 |
| Decompensated cardiac failure | 300179017 | 195111005 | 300179017 |
| Compensated cardiac failure | 300180019 | 195112003 | 300180019 |
| HFNEF - heart failure with normal ejection fraction | 1661371000000112 | 446221000 | 1713091000000115 |
| Acute oedema of lung, unspecified | 1216090015 | 40541001 | 67598017 |
| Severe left ventricular systolic dysfunction | 1991651000006115 | 1991651000006104 | 1991651000006115 |
| LVF - Left ventricular failure | 3886071000006114 | 85232009 | 1235017018 |
| Left heart failure | 3886041000006118 | 85232009 | 141303019 |
| Right heart failure due to pulmonary hypertension | 7052811000006113 | 426012001 | 2674125010 |
| Cardiac failure NOS | 223981000000118 | 84114007 | 139482012 |
| Diastolic heart failure | 6919191000006119 | 418304008 | 2577902015 |
| [RFC] Cardiac failure | 905391000006119 | 905391000006103 | 905391000006119 |
| Acute oedema of lung | 3152711000006119 | 40541001 | 492667013 |
| Congestive cardiac failure | 493287011 | 42343007 | 70653017 |
| Acute heart failure | 94251011 | 56675007 | 94251011 |
| Pulmonary congestion | 112265015 | 67599009 | 112265015 |
| Post cardiac operation heart failure NOS | 300217019 | 195130005 | 300214014 |
| CHF - Congestive heart failure | 3182561000006118 | 42343007 | 493289014 |
| Chronic pulmonary edema | 3257341000006116 | 46847001 | 78084015 |
| Heart failure | 139475013 | 84114007 | 139475013 |
| Right ventricular failure | 490972013 | 367363000 | 490972013 |
| Fluid overload pulmonary oedema | 5054791000006119 | 233712009 | 350205014 |
| Fluid overload pulmonary oedema | 5054791000006119 | 233712009 | 350205014 |
| Heart failure with normal ejection fraction | 1647701000000118 | 446221000 | 2883808011 |

| | | | |
|---|---|---|---|
| Postoperative pulmonary edema | 2682661000006114 | 11468004 | 19840013 |
| New York Heart Association Classification - Class III | 2616472016 | 420913000 | 2616472016 |
| Acute pulmonary oedema NOS | 301743011 | 40541001 | 492666016 |
| Echocardiogram shows left ventricular systolic dysfunction | 2159197017 | 407596008 | 2159197017 |
| Echocardiogram shows left ventricular diastolic dysfunction | 2159198010 | 407597004 | 2159198010 |
| Pulmonary oedema - acute | 3152721000006110 | 40541001 | 1490256017 |
| Pulmonary oedema - acute | 1490256017 | 40541001 | 492666016 |
| Suspected heart failure | 1488591011 | 394887005 | 1488591011 |
| Malignant hypertensive heart disease with CCF | 728671000006119 | 83105008 | 1236017010 |
| Exacerbation of congestive heart failure | 8030311000006111 | 96311000119109 | 2921293019 |
| H/O: Heart failure in last year | 453099015 | 309634009 | 453099015 |
| Acute right-sided heart failure | 6204381000006117 | 359617009 | 474047012 |
| Acute right-sided heart failure | 6204381000006117 | 359617009 | 474047012 |
| Biventricular failure | 510016018 | 92506005 | 153058012 |
| Heart failure with reduced ejection fraction | 7573171000006116 | 703272007 | 3008211011 |
| Impaired left ventricular function | 784191000006110 | 275514001 | 411506018 |
| Decompensated chronic heart failure | 7025691000006110 | 424404003 | 2645367010 |
| Left ventricular systolic dysfunction | 216207010 | 134401001 | 216207010 |
| Pulmonary congestion and hypostasis | 301689014 | 196115007 | 301689014 |
| Congestive heart failure with right heart failure | 8011111000006111 | 23341000119109 | 2984250012 |

#### Table S44. Heart failure HES (ICD10) codes

| ICD10 codes | Term |
|---|---|
| I50.0 | Congestive heart failure |
| I50.1 | Left ventricular heart failure |
| I50.9 | Heart failure, unspecified |
| I11.0 | Hypertensive heart disease with (congestive) heart failure |
| I13.0 | Hypertensive heart and renal disease with (congestive) heart failure |
| I13.2 | Hypertensive heart and renal disease with both (congestive) heart failure and renal failure |
| I42.0 | Dilated cardiomyopathy |
| K76.1 | Chronic passive congestion of liver |
| J81 | Pulmonary oedema |
| R57.0 | Cardiogenic shock |

### Chapter 3. Code lists for baseline characteristics

#### Table S45. Ethinicity Aurum codes

| Term | Medcode ID | SNOMED CT Concept ID | SNOMED CT Description ID |
|---|---|---|---|
| Black Arab | 411574019 | 275587000 | 411574019 |
| Black Iranian | 411575018 | 275588005 | 411575018 |
| Black East African Asian | 411576017 | 275589002 | 411576017 |
| Caribbean Island (NMO) | 411578016 | 275591005 | 411578016 |
| Iranian (NMO) | 411582019 | 275595001 | 411582019 |
| Turkish Cypriot (NMO) | 411597016 | 275602002 | 411597016 |
| O/E - Asian origin | 412016016 | 276029002 | 412016016 |
| Australian origin | 250227018 | 160518001 | 250227018 |
| Middle Eastern origin | 250229015 | 160520003 | 250229015 |
| Far Eastern origin | 250230013 | 160521004 | 250230013 |
| West Indian origin | 250231012 | 160522006 | 250231012 |
| O/E - Europeanoid | 253629014 | 162731001 | 253629014 |
| Other ethnic NEC (NMO) | 285988010 | 186005001 | 285958018 |
| New Zealand ethnic group NOS | 286022019 | 186035008 | 286006015 |
| Afghan | 1564151000006117 | 1564151000006101 | 1564151000006117 |
| Australian | 1564241000006111 | 1564241000006107 | 1564241000006111 |
| Bangladeshi | 1564291000006119 | 1564291000006103 | 1564291000006119 |
| Burmese | 1564431000006119 | 1564431000006103 | 1564431000006119 |
| Chilean | 1564511000006114 | 1564511000006105 | 1564511000006114 |
| Cuban | 1564581000006119 | 1564581000006103 | 1564581000006119 |
| Filipino | 1564731000006113 | 1564731000006109 | 1564731000006113 |
| Greek | 1564811000006111 | 1564811000006107 | 1564811000006111 |
| Israeli | 1564971000006113 | 1564971000006109 | 1564971000006113 |
| Italian | 1564981000006111 | 1564981000006107 | 1564981000006111 |
| Korean (North) | 1565071000006113 | 1565071000006109 | 1565071000006113 |
| Malaysian | 1565211000006111 | 1565211000006107 | 1565211000006111 |
| Moroccan | 1565321000006110 | 1565321000006106 | 1565321000006110 |
| New Zealander | 1565391000006112 | 1565391000006108 | 1565391000006112 |
| Saudi Arabian | 1565611000006114 | 1565611000006105 | 1565611000006114 |
| Somali | 1565701000006116 | 1565701000006100 | 1565701000006116 |
| Thai | 1565821000006119 | 1565821000006103 | 1565821000006119 |
| Venezuelan | 1565941000006116 | 1565941000006100 | 1565941000006116 |
| Vietnamese | 1565951000006119 | 1565951000006103 | 1565951000006119 |
| On examination - Asian origin | 5580031000006118 | 276029002 | 2669882010 |
| Ethnic group not recorded | 456652017 | 312861003 | 456652017 |
| Other ethnic, mixed white origin | 4740381000006118 | 186022004 | 2668812012 |
| Mixed multiple ethnic groups: White and Black Caribbean - England and Wales ethnic category 2011 census | 1968091000006110 | 976711000000103 | 2484791000000114 |

| | | | |
|---|---|---|---|
| Asian or Asian British: Bangladeshi - England and Wales ethnic category 2011 census | 1968151000006117 | 976831000000100 | 2485031000000111 |
| Black or African or Caribbean or Black British: other Black or African or Caribbean background - England and Wales ethnic category 2011 census | 1968201000006114 | 976931000000109 | 2485231000000119 |
| Asian or Asian British: Pakistani - Northern Ireland ethnic category 2011 census | 1968311000006115 | 977711000000100 | 2486881000000113 |
| Asian or Asian British: Chinese - Northern Ireland ethnic category 2011 census | 1968331000006114 | 977751000000101 | 2486961000000110 |
| Asian or Asian Scottish or Asian British: Chinese - Scotland ethnic category 2011 census | 1968511000006114 | 978191000000109 | 2487851000000117 |
| Caribbean or Black: any other Black or Caribbean group - Scotland ethnic category 2011 census | 1968571000006117 | 978361000000101 | 2488221000000112 |
| British ethnic minority unspecified (NMO) | 4740261000006118 | 186007009 | 2668698012 |
| On examination - Mongoloid origin | 5507871000006110 | 268914005 | 2669720011 |
| Black Caribbean | 514611000006111 | 185988007 | 285930014 |
| Chinese | 550541000006110 | 33897005 | 56590016 |
| White British | 459726019 | 315236000 | 459726019 |
| White Irish | 459727011 | 315237009 | 459727011 |
| Other Asian ethnic group | 459784018 | 315281001 | 459784018 |
| Irish traveller | 459786016 | 315283003 | 459786016 |
| White: Irish - England and Wales ethnic category 2011 census | 2484671000000118 | 976651000000108 | 2484671000000118 |
| White: Scottish - Scotland ethnic category 2011 census | 2487281000000112 | 977911000000103 | 2487281000000112 |
| White: Polish - Scotland ethnic category 2011 census | 2487481000000113 | 978011000000101 | 2487481000000113 |
| Sri Lankan - ethnic category 2001 census | 136081000000111 | 86461000000107 | 136081000000111 |
| Albanian - ethnic category 2001 census | 138231000000116 | 88971000000106 | 138231000000116 |
| Jewish - ethnic category 2001 census | 138241000000113 | 88991000000105 | 138241000000113 |
| White and Black African - ethnic category 2001 census | 141331000000116 | 92431000000100 | 141331000000116 |
| Other Asian background - ethnic category 2001 census | 141381000000117 | 92481000000101 | 141381000000117 |
| Welsh - ethnic category 2001 census | 141441000000119 | 92551000000106 | 141441000000119 |
| Northern Irish - ethnic category 2001 census | 141451000000116 | 92561000000109 | 141451000000116 |
| Chinese and White - ethnic category 2001 census | 141491000000112 | 92601000000109 | 141491000000112 |
| Kashmiri - ethnic category 2001 census | 141531000000112 | 92651000000105 | 141531000000112 |
| East African Asian - ethnic category 2001 census | 141541000000115 | 92661000000108 | 141541000000115 |
| British Asian - ethnic category 2001 census | 141561000000119 | 92681000000104 | 141561000000119 |
| Caribbean Asian - ethnic category 2001 census | 141571000000114 | 92691000000102 | 141571000000114 |
| Nigerian - ethnic category 2001 census | 141601000000119 | 92731000000108 | 141601000000119 |
| Filipino - ethnic category 2001 census | 141641000000116 | 92771000000105 | 141641000000116 |
| Cypriot (part not stated) - ethnic category 2001 census | 141661000000115 | 92791000000109 | 141661000000115 |
| Greek - ethnic category 2001 census | 142701000000116 | 93931000000104 | 142701000000116 |
| Kosovan - ethnic category 2001 census | 142741000000118 | 93981000000100 | 142741000000118 |
| Croatian - ethnic category 2001 census | 142761000000117 | 94001000000108 | 142761000000117 |
| Mixed Irish and other White - ethnic category 2001 census | 142781000000114 | 94021000000104 | 142781000000114 |
| North African - ethnic category 2001 census | 142811000000112 | 94061000000107 | 142811000000112 |

| | | | |
|---|---|---|---|
| Buddhist - ethnic category 2001 census | 142881000000117 | 94131000000103 | 142881000000117 |
| Caribbean - ethnic category 2001 census | 154401000000118 | 107691000000105 | 154401000000118 |
| English - ethnic category 2001 census | 157281000000117 | 110761000000106 | 157281000000117 |
| Sinhalese - ethnic category 2001 census | 157301000000116 | 110781000000102 | 157301000000116 |
| Serbian - ethnic category 2001 census | 157991000000110 | 88981000000108 | 157991000000110 |
| Asian and Chinese - ethnic category 2001 census | 158361000000116 | 92611000000106 | 158361000000116 |
| Mixed Black - ethnic category 2001 census | 158371000000111 | 92721000000106 | 158371000000111 |
| Italian - ethnic category 2001 census | 158481000000115 | 93961000000109 | 158481000000115 |
| British ethnic minority specified (NMO) | 4740241000006117 | 186006000 | 2668697019 |
| Tongan | 504723011 | 81560001 | 504723011 |
| White Scottish | 1780407014 | 401213008 | 1780407014 |
| Other white British ethnic group | 1780408016 | 401214002 | 1780408016 |
| Caucasian | 6846371000006111 | 413773004 | 2533842018 |
| Romanian | 1158211000000111 | 445343003 | 2872780011 |
| Black Caribbean | 285930014 | 185988007 | 285930014 |
| Black, other, non-mixed origin | 285931013 | 185989004 | 285931013 |
| Black Indian sub-continent | 285948017 | 185995003 | 285948017 |
| Black - other Asian | 285949013 | 185996002 | 285949013 |
| Other Black - Black/White orig | 285952017 | 185999009 | 285952017 |
| Other African countries (NMO) | 285971015 | 186010002 | 285971015 |
| Indian sub-continent (NMO) | 285976013 | 186012005 | 285976013 |
| Other European (NMO) | 285987017 | 186017004 | 285987017 |
| Other ethnic, Black/White orig | 285990011 | 186020007 | 285990011 |
| Other ethnic, Asian/White orig | 285991010 | 186021006 | 285991010 |
| Other ethnic, other mixed orig | 285993013 | 186023009 | 285993013 |
| New Zealand ethnic groups | 286006015 | 186035008 | 286006015 |
| Other European in New Zealand | 286009010 | 186037000 | 286009010 |
| New Zealand Maori | 286012013 | 186039002 | 286012013 |
| Tokelauan | 286015010 | 186042008 | 286015010 |
| Turkish/Turkish Cypriot (NMO) | 405071018 | 270467002 | 405071018 |
| Race: White | 2537217015 | 413773004 | 2537217015 |
| Black Caribbean/West India/Guyana | 5516681000006114 | 270460000 | 2983029017 |
| Commonwealth of (Russian) Independent States - ethnic category 2001 census | 937311000006113 | 88961000000104 | 138221000000118 |
| Other republics which made up the former Yugoslavia - ethnic category 2001 census | 937371000006116 | 94011000000105 | 142771000000112 |
| Middle Eastern (excluding Israeli, Iranian and Arab) - ethnic category 2001 census | 937871000006114 | 94071000000100 | 142821000000118 |
| Multi-ethnic islands: Mauritian or Seychellois or Maldivian or St Helena - ethnic category 2001 census | 937941000006111 | 94121000000100 | 142871000000119 |
| Black N African/Arab/Iranian | 405065012 | 270461001 | 405065012 |
| Indo-Caribbean (NMO) | 411584018 | 275597009 | 411584018 |
| Greek (NMO) | 411594011 | 275599007 | 411594011 |
| Black West Indian | 453109012 | 309643000 | 453109012 |

| | | | |
|---|---|---|---|
| Other black ethnic group | 459782019 | 315279003 | 459782019 |
| Black African and White | 460154012 | 315635008 | 460154012 |
| White: English or Welsh or Scottish or Northern Irish or British - England and Wales ethnic category 2011 census | 1968051000006116 | 976631000000101 | 2484631000000115 |
| Mixed multiple ethnic groups: any other Mixed or multiple ethnic background - England and Wales ethnic category 2011 census | 1968121000006114 | 976771000000108 | 2484911000000115 |
| Asian or Asian British: Indian - England and Wales ethnic category 2011 census | 1968131000006112 | 976791000000107 | 2484951000000116 |
| Asian or Asian British: Pakistani - England and Wales ethnic category 2011 census | 1968141000006119 | 976811000000108 | 2484991000000112 |
| Black or African or Caribbean or Black British: African - England and Wales ethnic category 2011 census | 1968181000006113 | 976891000000104 | 2485151000000115 |
| Black or African or Caribbean or Black British: Caribbean - England and Wales ethnic category 2011 census | 1968191000006111 | 976911000000101 | 2485191000000111 |
| Other ethnic group: Arab - England and Wales ethnic category 2011 census | 1968211000006112 | 976951000000102 | 2485271000000117 |
| Asian or Asian British: Indian - Northern Ireland ethnic category 2011 census | 1968301000006118 | 977591000000103 | 2486641000000116 |
| Asian or Asian British: Bangladeshi - Northern Ireland ethnic category 2011 census | 1968321000006111 | 977731000000108 | 2486921000000119 |
| Asian or Asian British: any other Asian background - Northern Ireland ethnic category 2011 census | 1968341000006116 | 977771000000105 | 2487001000000110 |
| Black or African or Caribbean or Black British: African - Northern Ireland ethnic category 2011 census | 1968351000006119 | 977791000000109 | 2487041000000113 |
| Black or African or Caribbean or Black British: other Black or African or Caribbean background - Northern Ireland ethnic category 2011 census | 1968371000006112 | 977831000000102 | 2487121000000119 |
| Other ethnic group: any other ethnic group - Northern Ireland ethnic category 2011 census | 1968391000006113 | 977871000000100 | 2487201000000119 |
| Asian or Asian Scottish or Asian British: Pakistani, Pakistani Scottish or Pakistani British - Scotland ethnic category 2011 census | 1968481000006118 | 978071000000106 | 2487601000000117 |
| Asian or Asian Scottish or Asian British: any other Asian group - Scotland ethnic category 2011 census | 1968521000006118 | 978211000000108 | 2487891000000113 |
| African: any other African - Scotland ethnic category 2011 census | 1968541000006113 | 978251000000107 | 2487971000000118 |
| Other ethnic group: Arab, Arab Scottish or Arab British - Scotland ethnic category 2011 census | 1968581000006119 | 978381000000105 | 2488261000000116 |
| Other ethnic group: any other ethnic group - Scotland ethnic category 2011 census | 1968591000006116 | 978401000000105 | 2488301000000114 |
| Black - ethnic group | 459730016 | 315240009 | 459730016 |
| Mixed ethnic census group | 459729014 | 315239007 | 459729014 |
| Asian - ethnic group | 286020010 | 315280000 | 459783012 |
| Race | 371005013 | 103579009 | 265558016 |
| White - ethnic group | 459728018 | 185984009 | 285926011 |
| Chinese | 1564521000006118 | 1564521000006102 | 1564521000006118 |
| Colombian | 1564531000006115 | 1564531000006104 | 1564531000006115 |
| Cypriot | 1564591000006116 | 1564591000006100 | 1564591000006116 |
| Peruvian | 1565491000006119 | 1565491000006103 | 1565491000006119 |
| Taiwanese | 1565791000006111 | 1565791000006107 | 1565791000006111 |
| Brazilian | 1573191000006117 | 1573191000006101 | 1573191000006117 |

| | | | |
|---|---|---|---|
| Indian | 781081000006113 | 414481008 | 2533844017 |
| Yemeni | 523591000000116 | 296841000000102 | 523591000000116 |
| Roma ethnic group | 2645811000000115 | 718958002 | 2678971000000113 |
| White Irish - ethnic category 2001 census | 1064041000000111 | 494161000000100 | 1064041000000111 |
| Polish - ethnic category 2001 census | 138201000000110 | 88941000000100 | 138201000000110 |
| Irish - ethnic category 2001 census | 141301000000110 | 92401000000106 | 141301000000110 |
| White and Black Caribbean - ethnic category 2001 census | 141321000000118 | 92421000000102 | 141321000000118 |
| Scottish - ethnic category 2001 census | 141431000000111 | 92541000000108 | 141431000000111 |
| Tamil - ethnic category 2001 census | 141551000000117 | 92671000000101 | 141551000000117 |
| Somali - ethnic category 2001 census | 141591000000113 | 92711000000100 | 141591000000113 |
| Ulster Scots - ethnic category 2001 census | 142691000000116 | 93921000000101 | 142691000000116 |
| Kurdish - ethnic category 2001 census | 142841000000113 | 94091000000101 | 142841000000113 |
| Sikh - ethnic category 2001 census | 142891000000115 | 94141000000107 | 142891000000115 |
| British or mixed British - ethnic category 2001 census | 158341000000117 | 92391000000108 | 158341000000117 |
| Ethnic group | 459785017 | 372148003 | 1210724015 |
| Black British | 285932018 | 185990008 | 285932018 |
| Indian | 285954016 | 414481008 | 2533844017 |
| Pakistani | 285955015 | 186002003 | 285955015 |
| Brit. ethnic minor. spec.(NMO) | 285959014 | 186006000 | 285959014 |
| Other Asian (NMO) | 285977016 | 186013000 | 285977016 |
| Other ethnic, mixed white orig | 285992015 | 186022004 | 285992015 |
| Pakeha | 286008019 | 186036009 | 286008019 |
| South East Asian | 286018012 | 186044009 | 286018012 |
| Arabs | 196621000006115 | 90027003 | 149237014 |
| Caucasian race | 196641000006110 | 413773004 | 2537215011 |
| Mixed racial group | 196681000006116 | 414752008 | 2533847012 |
| Oriental | 196701000006118 | 414978006 | 2533849010 |
| Unknown racial group | 196731000006114 | 415794004 | 2533839012 |
| Black East African Asian/Indo-Caribbean | 514651000006112 | 270462008 | 405066013 |
| Caribbean I./W.I./Guyana (NMO) | 405067016 | 270463003 | 405067016 |
| Black African | 30683015 | 18167009 | 30683015 |
| RACE: Bangladeshi | 196631000006117 | 186003008 | 285956019 |
| White British - ethnic category 2001 census | 1063981000000117 | 494131000000105 | 1063981000000117 |
| Black - other African country | 285943014 | 185993005 | 285943014 |
| Other ethnic, mixed origin | 285989019 | 186019001 | 285989019 |
| Cook Island Maori | 286013015 | 186040000 | 286013015 |
| Niuean | 286014014 | 186041001 | 286014014 |
| Afro-Caribbean | 196601000006113 | 413465009 | 2533840014 |
| Koreans | 196671000006119 | 38361009 | 63432018 |
| Other New Zealand ethnic group | 286021014 | 186035008 | 286006015 |
| Patient ethnicity unknown | 308131000000114 | 202171000000101 | 308131000000114 |

| | | | |
|---|---|---|---|
| Gypsy | 3146431000006116 | 40182006 | 2162400016 |
| Bangladeshi or British Bangladeshi - ethnic category 2001 census | 937541000006115 | 92471000000103 | 141371000000119 |
| Mixed multiple ethnic groups: White and Black African - Northern Ireland ethnic category 2011 census | 1968271000006115 | 977411000000108 | 2486281000000114 |
| Mixed multiple ethnic groups: any other Mixed or multiple ethnic background - Northern Ireland ethnic category 2011 census | 1968291000006119 | 977551000000106 | 2486561000000119 |
| White: any other White ethnic group - Scotland ethnic category 2011 census | 1968461000006111 | 978031000000109 | 2487521000000113 |
| South American origin | 250226010 | 160517006 | 250226010 |
| Japanese - ethnic category 2001 census | 141631000000113 | 92761000000103 | 141631000000113 |
| Bosnian - ethnic category 2001 census | 142751000000115 | 93991000000103 | 142751000000115 |
| O/E - Mongoloid origin | 402434017 | 268914005 | 402434017 |
| N African Arab/Iranian (NMO) | 405068014 | 270464009 | 405068014 |
| Guyana (NMO) | 411580010 | 275593008 | 411580010 |
| Greek Cypriot (NMO) | 411595012 | 275600005 | 411595012 |
| Black Guyana | 453110019 | 309644006 | 453110019 |
| Black Indo-Caribbean | 411577014 | 275590006 | 411577014 |
| West Indian (NMO) | 411579012 | 275592003 | 411579012 |
| Black Caribbean and White | 460153018 | 315634007 | 460153018 |
| Black | 459731017 | 315240009 | 459731017 |
| White | 285925010 | 185984009 | 285925010 |
| Bangladeshi | 285956019 | 186003008 | 285956019 |
| Irish (NMO) | 285978014 | 186014006 | 285978014 |
| Portuguese | 133078012 | 80208004 | 133078012 |
| Pakistani | 1565441000006111 | 1565441000006107 | 1565441000006111 |
| Black Black - other | 285950013 | 185989004 | 285931013 |
| European origin | 250222012 | 160513005 | 250222012 |
| Asian origin | 250224013 | 160515003 | 250224013 |
| White - Northern Ireland ethnic category 2011 census | 2486161000000112 | 977351000000100 | 2486161000000112 |
| White: other British - Scotland ethnic category 2011 census | 2487321000000116 | 977931000000106 | 2487321000000116 |
| White: Irish - Scotland ethnic category 2011 census | 2487361000000112 | 977951000000104 | 2487361000000112 |
| Other Pacific ethnic group | 286017019 | 372148003 | 1210724015 |
| Nepalese | 1745831000006112 | 1745831000006108 | 1745831000006112 |
| Caucasoid race | 6846391000006112 | 413773004 | 2537216012 |
| Japanese | 196661000006114 | 414551003 | 2533843011 |
| Race not stated | 196691000006118 | 415226007 | 2533846015 |
| Other ethnic, Black/White origin | 4740341000006112 | 186020007 | 2763938013 |
| Gypsy/Romany - ethnic category 2001 census | 138191000000113 | 88931000000109 | 138191000000113 |
| Pakistani or British Pakistani - ethnic category 2001 census | 141361000000114 | 92461000000105 | 141361000000114 |
| Black and Asian - ethnic category 2001 census | 141471000000113 | 92581000000100 | 141471000000113 |
| Black and Chinese - ethnic category 2001 census | 141481000000110 | 92591000000103 | 141481000000110 |

| | | | |
|---|---|---|---|
| Mixed Asian - ethnic category 2001 census | 141511000000116 | 92631000000103 | 141511000000116 |
| Vietnamese - ethnic category 2001 census | 141621000000111 | 92751000000101 | 141621000000111 |
| Turkish Cypriot - ethnic category 2001 census | 142721000000113 | 93951000000106 | 142721000000113 |
| Turkish - ethnic category 2001 census | 156921000000110 | 110401000000103 | 156921000000110 |
| Hindu - ethnic category 2001 census | 157351000000115 | 110831000000107 | 157351000000115 |
| Slovak | 1551471000000116 | 36329002 | 3314524013 |
| Other White European or European unspecified or Mixed European - ethnic category 2001 census | 937391000006115 | 94041000000106 | 142801000000110 |
| Other Black or Black unspecified - ethnic category 2001 census | 937731000006115 | 92741000000104 | 141611000000117 |
| Fijian | 501416013 | 69865008 | 501416013 |
| Afro-Caucasian | 196611000006111 | 413466005 | 2533841013 |
| Gypsies | 850671000006119 | 40182006 | 63802019 |
| Baltic States (Estonian or Latvian or Lithuanian) - ethnic category 2001 census | 937301000006110 | 88951000000102 | 138211000000112 |
| Other Mixed or Mixed unspecified - ethnic category 2001 census | 937511000006119 | 92621000000100 | 141501000000118 |
| Other Asian or Asian unspecified - ethnic category 2001 census | 937651000006117 | 92701000000102 | 141581000000111 |
| Asian or Asian British: Chinese - England and Wales ethnic category 2011 census | 1968161000006115 | 976851000000107 | 2485071000000113 |
| Mixed multiple ethnic groups: White and Black Caribbean - Northern Ireland ethnic category 2011 census | 1968261000006110 | 977391000000108 | 2486241000000118 |
| Mixed multiple ethnic groups: White and Asian - Northern Ireland ethnic category 2011 census | 1968281000006117 | 977431000000100 | 2486321000000118 |
| Asian or Asian Scottish or Asian British: Indian, Indian Scottish or Indian British - Scotland ethnic category 2011 census | 1968491000006115 | 978111000000100 | 2487681000000110 |
| Other ethnic non-mixed (NMO) | 285958018 | 186005001 | 285958018 |
| Brit. ethnic minor. unsp (NMO) | 285960016 | 186007009 | 285960016 |
| New Zealand European | 286007012 | 186036009 | 286007012 |
| Czech | 1160331000000119 | 286009 | 3314518019 |
| Bulgarian | 1158301000000115 | 29343004 | 1158301000000115 |
| Irish Traveller - ethnic category 2001 census | 138171000000114 | 88911000000101 | 138171000000114 |
| Traveller - ethnic category 2001 census | 138181000000111 | 88921000000107 | 138181000000111 |
| South and Central American - ethnic category 2001 census | 138271000000119 | 89021000000101 | 138271000000119 |
| Muslim - ethnic category 2001 census | 138281000000117 | 89031000000104 | 138281000000117 |
| Other White background - ethnic category 2001 census | 141311000000112 | 92411000000108 | 141311000000112 |
| Other Mixed background - ethnic category 2001 census | 141351000000111 | 92451000000107 | 141351000000111 |
| African - ethnic category 2001 census | 141391000000115 | 92491000000104 | 141391000000115 |
| Chinese - ethnic category 2001 census | 141401000000117 | 92511000000107 | 141401000000117 |
| Other - ethnic category 2001 census | 141411000000115 | 92521000000101 | 141411000000115 |
| Cornish - ethnic category 2001 census | 141461000000118 | 92571000000102 | 141461000000118 |
| Punjabi - ethnic category 2001 census | 141521000000110 | 92641000000107 | 141521000000110 |
| Malaysian - ethnic category 2001 census | 141651000000118 | 92781000000107 | 141651000000118 |
| Greek Cypriot - ethnic category 2001 census | 142711000000119 | 93941000000108 | 142711000000119 |
| Latin American - ethnic category 2001 census | 142861000000114 | 94111000000106 | 142861000000114 |

| | | | |
|---|---|---|---|
| Black and White - ethnic category 2001 census | 157291000000115 | 110771000000104 | 157291000000115 |
| Other Black background - ethnic category 2001 census | 158351000000119 | 92501000000105 | 158351000000119 |
| White: any other White background - England and Wales ethnic category 2011 census | 1968081000006112 | 976691000000100 | 2484751000000118 |
| Mixed multiple ethnic groups: White and Black African - England and Wales ethnic category 2011 census | 1968101000006116 | 976731000000106 | 2484831000000119 |
| Asian or Asian British: any other Asian background - England and Wales ethnic category 2011 census | 1968171000006110 | 976871000000103 | 2485111000000119 |
| Other ethnic group: Arab - Northern Ireland ethnic category 2011 census | 1968381000006110 | 977851000000109 | 2487161000000110 |
| Mixed or multiple ethnic groups: any Mixed or multiple ethnic group - Scotland ethnic category 2011 census | 1968471000006116 | 978051000000102 | 2487561000000117 |
| Albanian | 1564161000006115 | 1564161000006104 | 1564161000006115 |
| Brazilian | 1564381000006110 | 1564381000006106 | 1564381000006110 |
| Indian | 1564921000006112 | 1564921000006108 | 1564921000006112 |
| Iranian | 1564941000006117 | 1564941000006101 | 1564941000006117 |
| Mexican | 1565281000006116 | 1565281000006100 | 1565281000006116 |
| E Afric Asian/Indo-Carib (NMO) | 405069018 | 270465005 | 405069018 |
| Greek/Greek Cypriot (NMO) | 405070017 | 270466006 | 405070017 |
| North African Arab (NMO) | 411581014 | 275594002 | 411581014 |
| East African Asian (NMO) | 411583012 | 275596000 | 411583012 |
| Turkish (NMO) | 411596013 | 275601009 | 411596013 |
| American | 1564181000006113 | 1564181000006109 | 1564181000006113 |
| Cambodian | 1564451000006114 | 1564451000006105 | 1564451000006114 |
| Central African | 1564491000006115 | 1564491000006104 | 1564491000006115 |
| Japanese | 1565011000006116 | 1565011000006100 | 1565011000006116 |
| Sri Lankan | 1565731000006112 | 1565731000006108 | 1565731000006112 |
| Yugoslavian | 1565971000006112 | 1565971000006108 | 1565971000006112 |
| Nepali | 1572831000000110 | 718131000000106 | 1572831000000110 |
| Other White or White unspecified - ethnic category 2001 census | 937411000006115 | 94051000000109 | 158491000000118 |
| White: Gypsy or Irish Traveller - England and Wales ethnic category 2011 census | 1968071000006114 | 976671000000104 | 2484711000000117 |
| Mixed multiple ethnic groups: White and Asian - England and Wales ethnic category 2011 census | 1968111000006118 | 976751000000104 | 2484871000000117 |
| Other ethnic group: any other ethnic group - England and Wales ethnic category 2011 census | 1968221000006116 | 976971000000106 | 2485311000000117 |
| Irish Traveller - Northern Ireland ethnic category 2011 census | 1968251000006113 | 977371000000109 | 2486201000000116 |
| Black or African or Caribbean or Black British: Caribbean - Northern Ireland ethnic category 2011 census | 1968361000006117 | 977811000000105 | 2487081000000117 |
| White: Gypsy or Irish Traveller - Scotland ethnic category 2011 census | 1968441000006112 | 977971000000108 | 2487401000000115 |
| Asian or Asian Scottish or Asian British: Bangladeshi, Bangladeshi Scottish or Bangladeshi British - Scotland ethnic category 2011 census | 1968501000006111 | 978171000000105 | 2487801000000118 |
| African: African, African Scottish or African British - Scotland ethnic category 2011 census | 1968531000006115 | 978231000000100 | 2487931000000115 |

| | | | |
|---|---|---|---|
| Caribbean or Black: Caribbean, Caribbean Scottish or Caribbean British - Scotland ethnic category 2011 census | 1968551000006110 | 978271000000103 | 2488011000000113 |
| Caribbean or Black: Black, Black Scottish or Black British - Scotland ethnic category 2011 census | 1968561000006112 | 978341000000102 | 2488181000000119 |
| Other ethnic, other mixed origin | 4740401000006118 | 186023009 | 2668813019 |
| Samoan | 507015012 | 86275006 | 507015012 |
| Vietnamese | 456650013 | 312859007 | 456650013 |
| African origin | 250223019 | 160514004 | 250223019 |
| North American origin | 250225014 | 160516002 | 250225014 |
| Indian origin | 250228011 | 160519009 | 250228011 |
| Race: West indian | 250243013 | 160531006 | 250243013 |
| Refusal by patient to provide information about ethnic group | 456651012 | 763726001 | 3643639011 |
| Arab - ethnic category 2001 census | 138251000000111 | 89001000000105 | 138251000000111 |
| Iranian - ethnic category 2001 census | 138261000000114 | 89011000000107 | 138261000000114 |
| White and Asian - ethnic category 2001 census | 141341000000113 | 92441000000109 | 141341000000113 |
| Ethnic category not stated - 2001 census | 141421000000114 | 92531000000104 | 141421000000114 |
| Other mixed White - ethnic category 2001 census | 142791000000111 | 94031000000102 | 142791000000111 |
| Israeli - ethnic category 2001 census | 142831000000116 | 94081000000103 | 142831000000116 |
| Moroccan - ethnic category 2001 census | 142851000000111 | 94101000000109 | 142851000000111 |
| Any other group - ethnic category 2001 census | 142901000000119 | 94151000000105 | 142901000000119 |
| Indian or British Indian - ethnic category 2001 census | 157271000000119 | 110751000000108 | 157271000000119 |
| Black British - ethnic category 2001 census | 157311000000119 | 110791000000100 | 157311000000119 |
| RACE: Chinese | 196651000006112 | 33897005 | 56590016 |
| RACE: Pakistani | 196721000006111 | 186002003 | 285955015 |
| Irish | 1564961000006118 | 1564961000006102 | 1564961000006118 |
| Korean (South) | 1565081000006111 | 1565081000006107 | 1565081000006111 |
| Micronesian | 1565291000006118 | 1565291000006102 | 1565291000006118 |
| Nepalese | 1565371000006111 | 1565371000006107 | 1565371000006111 |
| Black Caribbean/W.I./Guyana | 405064011 | 270460000 | 405064011 |
| Black North African | 411573013 | 275586009 | 411573013 |
| Other ethnic, Asian/White origin | 4740361000006111 | 186021006 | 2764123018 |
| Chinese | 56590016 | 33897005 | 56590016 |
| Patient declined to provide information about ethnic group | 12009371000006110 | 763726001 | 3644134011 |
| Black - other, mixed | 285951012 | 185998001 | 285951012 |
| Other Black - Black/Asian orig | 285953010 | 186000006 | 285953010 |

#### Table S46. Smoking Aurum codes

| Term | Medcode ID | SNOMED CT Concept ID | SNOMED CT Description ID |
|---|---|---|---|
| **Current smoker** | | | |
| user of electronic cigarette | 1879431000006110 | 1879431000006106 | 1879431000006110 |
| tobacco abuse | 3959141000006110 | 89765005 | 148823015 |
| tobacco use, continuous | 4765711000006110 | 191887008 | 2951660019 |
| ready to stop smoking | 1488577017 | 394872000 | 1488577017 |
| tobacco dependence | 102951000006115 | 89765005 | 2951656017 |
| grade c moderate smoker (11-20/day) | 854981000006117 | 854981000006101 | 854981000006117 |
| smoking cessation advice | 338608011 | 225323000 | 338608011 |
| grade d heavy smoker (>20 day) | 855001000006114 | 855001000006105 | 855001000006114 |
| referred for copd structured smoking assessment | 1704551000006110 | 375851000000108 | 739301000000115 |
| current smoker | 503483019 | 77176002 | 503483019 |
| smoking free weeks | 1488873010 | 395177003 | 1488873010 |
| light smoker - 1-9 cigs/day | 743331000006116 | 160603005 | 250358015 |
| smokes drugs in cigarette form | 342444018 | 228378005 | 342444018 |
| smokes drugs through a pipe | 342445017 | 228379002 | 342445017 |
| cigar consumption | 344794017 | 230057008 | 344794017 |
| pipe tobacco consumption | 344795016 | 230058003 | 344795016 |
| [v]tobacco abuse counselling | 461114016 | 711028002 | 3047058010 |
| very heavy smoker - 40+cigs/d | 67621000006112 | 160606002 | 250362014 |
| tobacco consumption | 102921000006112 | 266918002 | 397730015 |
| cigarette consumption | 344793011 | 230056004 | 344793011 |
| tobacco dependence, unspecified | 295256013 | 89765005 | 148820017 |
| referral to smoking cessation advisor | 1489355012 | 395700008 | 1489355012 |
| smokers' cough | 78013015 | 46802002 | 78013015 |
| pipe smoker | 136515019 | 82302008 | 136515019 |
| tobacco dependence, episodic | 295258014 | 191888003 | 295258014 |
| rolls own cigarettes | 250375014 | 160619003 | 250375014 |
| keeps trying to stop smoking | 250368013 | 160612007 | 250368013 |
| smoking started | 2669652019 | 266929003 | 2669652019 |
| provision of smoking cessation leaflet | 2982426011 | 699033005 | 2982426011 |
| smokes/uses tobacco products | 961581000006114 | 961581000006105 | 961581000006114 |
| smoking cessation referral declined | 1591651000006110 | 1591651000006106 | 1591651000006110 |
| referral to stop-smoking clinic | 459722017 | 315232003 | 459722017 |
| smoking restarted | 137791000006118 | 308438006 | 2670126018 |
| smoker (read codes) | 137711000006111 | 137711000006107 | 137711000006111 |
| cigarette smoker | 854021000006115 | 854021000006104 | 854021000006115 |
| cigar smoker | 99639019 | 59978006 | 99639019 |
| not interested in stopping smoking | 1488578010 | 394873005 | 1488578010 |
| reason for restarting smoking | 1780360012 | 401159003 | 1780360012 |

| | | | |
|---|---|---|---|
| current smoker annual review - enhanced services admin | 1714541000006110 | 505651000000103 | 1152111000000118 |
| not interested in stopping smoking | 903981000006117 | 903981000006101 | 903981000006117 |
| previous smoking quit attempts | 904021000006118 | 904021000006102 | 904021000006118 |
| thinking about stopping smoking | 904031000006115 | 904031000006104 | 904031000006115 |
| carbon monoxide validation of smoking status | 904051000006110 | 904051000006106 | 904051000006110 |
| smoking cessation bupropion therapy | 904221000006110 | 904221000006106 | 904221000006110 |
| [rfc] smoking cessation | 909391000006117 | 909391000006101 | 909391000006117 |
| smoking reduced | 216212011 | 134406006 | 216212011 |
| current smoker nos | 604961000006114 | 604961000006105 | 604961000006114 |
| smoker | 128130017 | 77176002 | 128130017 |
| ready to stop smoking | 904001000006111 | 904001000006107 | 904001000006111 |
| waking time to first cigarette | 904041000006113 | 904041000006109 | 904041000006113 |
| carbon monoxide validation confirms smoker | 904121000006117 | 904121000006101 | 904121000006117 |
| cigarette smoker | 108938018 | 65568007 | 108938018 |
| failed attempt to stop smoking | 1592611000000110 | 446172000 | 2882460017 |
| [v]tobacco use | 460828018 | 110483000 | 175325014 |
| [x]mental and behavioural disorder due to use of tobacco | 295951010 | 30310000 | 50736017 |
| varenicline smoking cessation therapy offered | 2462431000000110 | 966991000000104 | 2462431000000110 |
| occasional smoker | 397733018 | 428041000124106 | 2717161000000119 |
| smoking restarted | 2670126018 | 308438006 | 2670126018 |
| minutes from waking to first tobacco consumption | 2474719011 | 413173009 | 2474719011 |
| smoker - amount smoked | 137721000006115 | 266918002 | 397730015 |
| thinking about stopping smoking | 1488576014 | 394871007 | 1488576014 |
| moderate smoker - 10-19 cigs/d | 700121000006118 | 160604004 | 250359011 |
| tobacco dependence, continuous | 295257016 | 191887008 | 295257016 |
| trivial smoker - < 1 cig/day | 88471000006112 | 266920004 | 397735013 |
| gradual smoking reduction | 852111000006118 | 852111000006102 | 852111000006118 |
| current smoker | 854071000006119 | 854071000006103 | 854071000006119 |
| tobacco consumption nos | 250387019 | 266918002 | 397730015 |
| grade b light smoker (1-10/day) | 854961000006110 | 854961000006106 | 854961000006110 |
| rolls own cigarettes | 852981000006111 | 852981000006107 | 852981000006111 |
| refuses stop smoking monitor | 285792013 | 765001003 | 3656021018 |
| heavy smoker - 20-39 cigs/day | 819331000006110 | 160605003 | 250360018 |
| [x]mental and behav dis due to use of tobacco: harmful use | 397911000006114 | 724697004 | 3445165019 |
| trying to give up smoking | 250372012 | 160616005 | 250372012 |
| exposure to cigarette/cigar smoke | 981841000006115 | 981841000006104 | 981841000006115 |
| tobacco dependence nos | 295260011 | 89765005 | 2951656017 |
| **Past smoker** | | | |
| ex-smoker - amount unknown | 250371017 | 8517006 | 15047015 |
| ex smoker | 649841000006110 | 8517006 | 15047015 |
| current non-smoker | 250374013 | 160618006 | 250374013 |

| | | | |
|---|---|---|---|
| smoking cessation therapy nos | 489931000000114 | 710081004 | 3043296015 |
| ex-cigar smoker | 852991000006114 | 852991000006105 | 852991000006114 |
| ex-smoker nos | 853001000006110 | 853001000006106 | 853001000006110 |
| ex-trivial smoker (<1/day) | 250363016 | 266921000 | 397736014 |
| ex-light smoker (1-9/day) | 250364010 | 266922007 | 397737017 |
| ex-heavy smoker (20-39/day) | 250366012 | 266924008 | 397739019 |
| ex-very heavy smoker (40+/day) | 250367015 | 266925009 | 397740017 |
| past smoker | 854111000006110 | 854111000006106 | 854111000006110 |
| ex cigar smoker | 649821000006115 | 160621008 | 250377018 |
| ex- rolled tobacco smoker | 649851000006112 | 649851000006108 | 649851000006112 |
| smoking cessation drug therapy | 482771000000118 | 713700008 | 3297364011 |
| smoking cessation therapy | 492511000000117 | 710081004 | 3043296015 |
| ex-smoker annual review - enhanced services administration | 1123951000000110 | 505761000000105 | 1154471000000114 |
| ex-smoker nos | 903041000006110 | 266928006 | 903041000006110 |
| cigarette pack-years | 1780396011 | 401201003 | 1780396011 |
| ex-pipe smoker | 854051000006112 | 854051000006108 | 854051000006112 |
| date stopped smoking | 854151000006111 | 854151000006107 | 854151000006111 |
| tobacco dependence in remission | 295259018 | 191889006 | 295259018 |
| ex-cigarette smoker | 418914010 | 281018007 | 418914010 |
| smoking cessation-maintain abstinence | 852121000006114 | 852121000006105 | 852121000006114 |
| other specified smoking cessation therapy | 482871000000111 | 710081004 | 3043296015 |
| cessation of smoking | 2636041000006110 | 8517006 | 15048013 |
| stopped smoking | 250373019 | 160617001 | 250373019 |
| ex pipe smoker | 649831000006117 | 160620009 | 250376010 |
| ex-moderate smoker (10-19/day) | 250365011 | 266923002 | 397738010 |
| ex-cigarette smoker | 649861000006114 | 649861000006105 | 649861000006114 |

#### Table S47. Measurements Aurum codes

| Term | Medcode ID | SNOMED CT Concept ID | SNOMED CT Description ID |
|---|---|---|---|
| **Height** | | | |
| O/E - height | 923831000006115 | 923831000006104 | 923831000006115 |
| O/E - height | 253669010 | 248333004 | 370729019 |
| Estimated height | 1910921000006114 | 1910921000006105 | 1910921000006114 |
| O/E - height NOS | 253676017 | 248333004 | 370729019 |
| Reported height | 1910931000006112 | 1910931000006108 | 1910931000006112 |
| **Weight** | | | |
| O/E - weight NOS | 253688015 | 27113001 | 45352010 |
| Baseline weight | 1780175010 | 400967004 | 1780175010 |
| Weight monitoring | 451201014 | 307818003 | 451201014 |
| Weight static | 2536041016 | 271398006 | 2536041016 |
| Reported weight | 1910911000006118 | 1910911000006102 | 1910911000006118 |
| O/E - weight | 923851000006110 | 923851000006106 | 923851000006110 |
| Weight screen | 59281000006111 | 804271000000109 | 1798011000000116 |
| Estimated weight | 1910901000006116 | 1910901000006100 | 1910901000006116 |
| O/E - weight | 253677014 | 27113001 | 45352010 |
| **BMI** | | | |
| OBESITY MULTIDISCIPLINARY CASE REVIEW | 2310101000000115 | 898011000000103 | 2310101000000115 |
| INTENSIVE WEIGHT MANAGEMENT PROGRAMME DECLINED | 2326491000000114 | 904631000000104 | 2326491000000114 |
| OBESE CLASS I (BODY MASS INDEX 30.0 - 34.9) | 2350241000000116 | 914721000000105 | 2350241000000116 |
| OBESE CLASS II (BODY MASS INDEX 35.0 - 39.9) | 2350261000000115 | 914731000000107 | 2350261000000115 |
| BODY MASS INDEX | 100716012 | 60621009 | 100716012 |
| TREATMENT OF OBESITY STARTED | 264758010 | 170799008 | 264758010 |
| REASON FOR OBESITY THERAPY - OCCUPATIONAL | 264768017 | 170807005 | 264768017 |
| OBESITY MONITORING NOS | 264769013 | 268522006 | 401534016 |
| INTERVENTION FOR RISK TO HEALTH ASSOCIATED WITH OVERWEIGHT AND OBESITY, ADVICE ABOUT DIET AND PHYSICAL ACTIVITY | 1753321000006116 | 756021000000103 | 1663521000000115 |
| INTERVENTION FOR RISK TO HEALTH ASSOCIATED WITH OVERWEIGHT AND OBESITY, ADVICE ABOUT DIET AND PHYSICAL ACTIVITY, CONSIDER DRUGS | 1753331000006118 | 756041000000105 | 1663561000000111 |
| INTERVENTION FOR RISK TO HEALTH ASSOCIATED WITH OVERWEIGHT AND OBESITY, ADVICE ABOUT DIET AND PHYSICAL ACTIVITY, CONSIDER DRUGS, CONSIDER SURGERY | 1753341000006111 | 756061000000106 | 1663601000000111 |
| BODY MASS INDEX LESS THAN 20 | 453856012 | 310252000 | 453856012 |
| BODY MASS INDEX NORMAL K/M2 | 253844013 | 35425004 | 59104012 |
| INTENSIVE WEIGHT MANAGEMENT PROGRAMME COMMENCED | 2326311000000114 | 904551000000107 | 2326311000000114 |
| HAS SEEN DIETICIAN - OBESITY | 264756014 | 170797005 | 264756014 |

| | | | |
|---|---|---|---|
| BODY MASS INDEX 18.5-24.9 | 1808071000006119 | 1808071000006103 | 1808071000006119 |
| BODY MASS INDEX 30+ - OBESITY | 253848011 | 162864005 | 253848011 |
| OBESITY MONITORING | 401534016 | 268522006 | 401534016 |
| FOLLOW-UP OBESITY ASSESSMENT | 264753018 | 170795002 | 264753018 |
| TREATMENT OF OBESITY CHANGED | 264757017 | 170798000 | 264757017 |
| BODY MASS INDEX HIGH K/M2 | 253845014 | 48499001 | 80800016 |
| BODY MASS INDEX LOW K/M2 | 253846010 | 6497000 | 11777011 |
| INTENSIVE WEIGHT MANAGEMENT PROGRAMME ENDED | 2326451000000118 | 904611000000107 | 2326451000000118 |
| TELEHEALTH OBESITY MONITORING | 2423851000000119 | 715279006 | 3302073018 |
| TREATMENT OF OBESITY STOPPED | 264759019 | 170800007 | 264759019 |
| ATTENDS SLIMMING CLINIC | 264760012 | 170801006 | 264760012 |
| BODY MASS INDEX 20-24 - NORMAL | 2474325012 | 412768003 | 2474325012 |
| RISK TO HEALTH ASSOCIATED WITH OVERWEIGHT AND OBESITY, AT HIGH RISK | 1753291000006113 | 755921000000105 | 1663311000000118 |
| BASELINE BODY MASS INDEX | 2196071000000116 | 846931000000101 | 2196071000000116 |
| BODY MASS INDEX LESS THAN 18.5 | 1808061000006114 | 1808061000006105 | 1808061000006114 |
| OBESE CLASS III (BMI EQUAL TO OR GREATER THAN 40.0) | 1900331000006113 | 914741000000103 | 2350281000000112 |
| BODY MASS INDEX 40+ - SEVERELY OBESE | 2160062010 | 408512008 | 2160062010 |
| BODY MASS INDEX CENTILE | 1551651000000111 | 446974000 | 2882934011 |
| BODY MASS INDEX INDEX 25-29 - OVERWEIGHT | 253847018 | 162863004 | 253847018 |
| BODY MASS INDEX | 923861000006112 | 923861000006108 | 923861000006112 |
| INITIAL OBESITY ASSESSMENT | 264752011 | 170794003 | 264752011 |
| RISK TO HEALTH ASSOCIATED WITH OVERWEIGHT AND OBESITY, AT NO INCREASED RISK | 1753271000006112 | 755881000000104 | 1663231000000115 |
| RISK TO HEALTH ASSOCIATED WITH OVERWEIGHT AND OBESITY, AT INCREASED RISK | 1753281000006110 | 755901000000101 | 1663271000000118 |
| RISK TO HEALTH ASSOCIATED WITH OVERWEIGHT AND OBESITY, AT VERY HIGH RISK | 1753301000006114 | 755941000000103 | 1663351000000119 |
| INTERVENTION FOR RISK TO HEALTH ASSOCIATED WITH OVERWEIGHT AND OBESITY, GENERAL ADVICE ON HEALTHY WEIGHT AND LIFESTYLE | 1753311000006112 | 756001000000107 | 1663481000000115 |
| **Blood pressure and Heart rate** | | | |
| Systolic arterial pressure | 114311000006111 | 72313002 | 120159016 |
| Diastolic arterial pressure | 619931000006119 | 1091811000000102 | 2734671000000117 |
| Heart rate | 487210016 | 364075005 | 487210016 |
| Baseline heart rate | 2378591000000110 | 928001000000104 | 2378591000000110 |

#### Table S48. Blood test Aurum codes

| Term | Medcode ID | SNOMED CT Concept ID | SNOMED CT Description ID |
|---|---|---|---|
| **Creatinine** | | | |
| EstimatedCreatinineClearance | 2470380013 | 395680003 | 2470380013 |
| EstimatedCreatinineClearance(cockcroft-gaultFormula) | 2465711000000116 | 968191000000100 | 2465711000000116 |
| SerumCreatinine | 380389013 | 1000731000000107 | 2577271000000111 |
| **eGFR** | | | |
| GfrCalculatedAbbreviatedMdrd | 976481000006110 | 1020291000000106 | 2569781000000116 |
| EgfrUsingCreatinine(ckd-epi)Per1.73SquareMetres | 1942831000006114 | 1011481000000105 | 2579541000000113 |
| GlomerularFiltrationRate | 133205018 | 80274001 | 133205018 |
| **Hb** | | | |
| Haemoglobin estimation | 813551000006113 | 1022431000000105 | 2553471000000116 |
| **HbA1C** | | | |
| HbA1c (haemoglobin A1c) level (diagnostic reference range) - IFCC (International Federation of Clinical Chemistry and Laboratory Medicine) standardised | 1986181000006111 | 1049301000000100 | 2643101000000114 |
| HbA1c - Haemoglobin A1c level | 3199181000006110 | 43396009 | 493589010 |
| Hemoglobin A1c (HBA1c) target | 6764291000006117 | 408591000 | 2621921019 |
| HbA1c - Hemoglobin A1c level | 3199191000006113 | 43396009 | 493590018 |
| HbA1c (haemoglobin A1c) target level - IFCC (International Federation of Clinical Chemistry and Laboratory Medicine) standardised | 1753211000006115 | 446074002 | 1725291000000118 |
| HbA1c measurement (DCCT aligned) | 6036701000006116 | 313835008 | 457955014 |
| HBA1c (Hemoglobin A1c) level | 7285151000006113 | 443911005 | 2840604014 |
| HbA1c level (Diabetes Control and Complications Trial aligned) | 457954013 | 1019431000000105 | 2566161000000115 |
| HbA1c (haemoglobin A1c) molar concentration in blood | 12622041000006111 | 1107481000000106 | 2773291000000116 |
| HbA1c level (DCCT aligned) | 6036711000006118 | 313835008 | 457954013 |
| HBA1c target | 2160141012 | 408591000 | 1739591000000117 |
| HbA1c - Glycated hemoglobin-A1c | 7960911000006117 | 733830002 | 3474736019 |
| HBA1c (Haemoglobin A1c) level | 7285141000006111 | 443911005 | 2840603015 |
| Raised HbA1c level | 1927791000006119 | 1927791000006103 | 1927791000006119 |
| HBA1c target | 6764261000006113 | 408591000 | 2160141012 |
| HbA1c (haemoglobin A1c) level (monitoring ranges) - IFCC (International Federation of Clinical Chemistry and Laboratory Medicine) standardised | 1986191000006114 | 1049321000000109 | 2643141000000112 |
| Target HbA1c level | 6764271000006118 | 408591000 | 2163995019 |
| HbA1c - Glycated haemoglobin-A1c | 7960921000006113 | 733830002 | 3474737011 |
| HbA1c (haemoglobin A1c) target - IFCC (International Federation of Clinical Chemistry and Laboratory Medicine) standardised | 7318701000006114 | 446074002 | 1725281000000115 |
| **Fasting lipids** | | | |
| High cholesterol | 2716221000006111 | 13644009 | 3035252016 |
| Fasting serum cholesterol | 667191000006111 | 667191000006107 | 667191000006111 |
| Cholesterol/HDL ratio | 457911015 | 313811003 | 457911015 |

| | | | |
|---|---|---|---|
| Serum cholesterol very high | 259230019 | 166831007 | 259230019 |
| Non HDL cholesterol level | 8411411000006113 | 1030411000000101 | 2595221000000113 |
| Serum cholesterol borderline high | 7261561000006118 | 442234001 | 2820789017 |
| Serum HDL (high density lipoprotein):non-HDL (high density lipoprotein) cholesterol ratio | 8396091000006112 | 1015271000000109 | 2607241000000118 |
| Cholesterol screening | 5579571000006112 | 275972003 | 411949013 |
| Serum non HDL (high density lipoprotein) cholesterol level | 8386311000006116 | 1006191000000106 | 2607621000000111 |
| Serum cholesterol borderline | 12716491000006111 | 166829003 | 259228016 |
| Calculated LDL cholesterol level | 6550661000006112 | 395065005 | 1488764011 |
| Lipoprotein cholesterol ratio measurement | 6040921000006112 | 313988001 | 458247013 |
| HDL cholesterol | 4079571000006118 | 102737005 | 166116016 |
| Target serum low density lipoprotein cholesterol level | 8241901000006112 | 774851000000106 | 1730921000000119 |
| Estimated serum non-high density lipoprotein cholesterol level | 1924001000006115 | 920471000000100 | 2362791000000114 |
| Serum high density lipoprotein cholesterol level | 259232010 | 1005681000000107 | 2577901000000110 |
| Serum random HDL (high density lipoprotein) cholesterol level | 259243019 | 1026461000000104 | 2599971000000117 |
| Serum random LDL (low density lipoprotein) cholesterol level | 259247018 | 1026481000000108 | 2599981000000115 |
| Serum total cholesterol measurement | 6831591000006111 | 412808005 | 2474364010 |
| High density lipoprotein cholesterol measurement | 2949971000006113 | 28036006 | 46931018 |
| Total cholesterol level | 8292941000006114 | 853681000000104 | 2210431000000111 |
| Cholesterol level | 8290381000006115 | 850981000000101 | 2204831000000119 |
| Serum VLDL (very low density lipoprotein) cholesterol level | 259236013 | 1003441000000101 | 2607471000000113 |
| Serum fasting LDL (low density lipoprotein) cholesterol level | 259246010 | 1026471000000106 | 2599941000000111 |
| Total cholesterol:HDL (high density lipoprotein) ratio | 259250015 | 1028551000000102 | 2602701000000112 |
| Plasma fasting HDL (high density lipoprotein) cholesterol level | 259567011 | 1028841000000102 | 2600031000000117 |
| HDL/total cholesterol ratio measurement | 4108441000006119 | 104583003 | 168549011 |
| Total cholesterol:HDL ratio measurement | 4600421000006112 | 166842003 | 259249015 |
| LDL-C (low density lipoprotein cholesterol) substance concentration in serum | 12623411000006112 | 1108551000000102 | 2774161000000118 |
| Target serum LDL (low density lipoprotein) cholesterol level | 8241911000006110 | 774851000000106 | 1730931000000117 |
| Plasma cholesterol/HDL (high density lipoprotein) ratio | 458252015 | 1015691000000106 | 2557701000000111 |
| Serum cholesterol/low density lipoprotein ratio | 458253013 | 1015701000000106 | 2565681000000110 |
| Plasma cholesterol/LDL (low density lipoprotein) ratio | 458256017 | 1015711000000108 | 2580131000000114 |
| Serum cholesterol/very low density lipoprotein ratio | 458257014 | 1015721000000102 | 2557711000000113 |
| Plasma HDL (high density lipoprotein) cholesterol level | 458313013 | 1010581000000101 | 2557051000000110 |
| Calculated LDL (low density lipoprotein) cholesterol level | 1488764011 | 1014501000000104 | 2552501000000118 |
| HDL - High density lipoprotein cholesterol level | 2950001000006114 | 28036006 | 483733018 |
| Target serum high density lipoprotein cholesterol level | 8241871000006112 | 774821000000101 | 1730851000000119 |
| Non-HDL cholesterol | 6014411000006114 | 312260007 | 455948011 |

| | | | |
|---|---|---|---|
| Serum HDL cholesterol measurement | 4600201000006115 | 166832000 | 259231015 |
| Fasting cholesterol level | 6052931000006111 | 315017003 | 459479011 |
| HDL (high density lipoprotein) cholesterol/total cholesterol ratio in plasma | 12622411000006116 | 1107731000000104 | 2771891000000118 |
| Serum fasting HDL (high density lipoprotein) cholesterol level | 259242012 | 1026451000000102 | 2599931000000119 |
| LDL - Low density lipoprotein cholesterol level | 4217681000006112 | 113079009 | 380428016 |
| Serum HDL cholesterol level | 4600211000006117 | 166832000 | 259232010 |
| Cholesterol/HDL ratio measurement | 6036221000006115 | 313811003 | 457910019 |
| Serum LDL cholesterol level | 4600241000006118 | 166833005 | 259233017 |
| Serum cholesterol borderline low | 7263101000006113 | 442350007 | 2819746011 |
| Serum LDL cholesterol measurement | 4600231000006111 | 166833005 | 259234011 |
| Serum total cholesterol level | 6831601000006115 | 412808005 | 2478443019 |
| Serum cholesterol/high density lipoprotein ratio | 458249011 | 1015681000000109 | 2574851000000110 |
| Serum cholesterol level | 6279041000006119 | 365794002 | 489235016 |
| Serum cholesterol level | 150921000006118 | 1005671000000105 | 2551271000000118 |
| Total cholesterol:HDL ratio | 4600431000006110 | 166842003 | 259250015 |
| LDL cholesterol | 4079621000006115 | 102739008 | 166118015 |
| Serum cholesterol normal | 259227014 | 166828006 | 259227014 |
| Serum cholesterol raised | 259229012 | 166830008 | 259229012 |
| Hypercholesterolaemia | 475418015 | 13644009 | 475418015 |
| Serum triglyceride/HDL cholesterol ratio | 1916571000006117 | 1916571000006101 | 1916571000006117 |
| Target serum total cholesterol level | 8241111000006113 | 773271000000103 | 1726681000000112 |
| HDL (high density lipoprotein) cholesterol/total cholesterol ratio in serum | 12622421000006112 | 1107741000000108 | 2771911000000115 |
| Serum cholesterol/HDL ratio | 6040951000006115 | 313989009 | 458249011 |
| Fasting lipids | 854781000006119 | 854781000006103 | 854781000006119 |
| Serum non high density lipoprotein cholesterol level | 2344491000000115 | 1006191000000106 | 2585861000000117 |
| Plasma random HDL (high density lipoprotein) cholesterol level | 259564016 | 1028831000000106 | 2600041000000114 |
| Plasma random LDL (low density lipoprotein) cholesterol level | 259569014 | 1028851000000104 | 2600051000000112 |
| Plasma fasting LDL (low density lipoprotein) cholesterol level | 259570010 | 1028861000000101 | 2600061000000110 |
| Plasma cholesterol/VLDL (very low density lipoprotein) ratio | 458260019 | 1015731000000100 | 2580141000000117 |
| Serum HDL (high density lipoprotein) cholesterol level | 8385731000006118 | 1005681000000107 | 2607231000000110 |
| HDL (high density lipoprotein) cholesterol substance concentration in serum | 12622321000006118 | 1107681000000108 | 2771791000000114 |
| HDL : total cholesterol ratio | 4108451000006117 | 104583003 | 279462018 |
| HDL cholesterol level | 855771000006118 | 855771000006102 | 855771000006118 |
| Serum cholesterol borderline | 259228016 | 166829003 | 259228016 |
| Serum total cholesterol level | 2478443019 | 994351000000103 | 2581451000000115 |
| Cholesterol | 34521000033114 | 84698008 | 34521000033114 |
| Serum cholesterol NOS | 259252011 | 1005671000000105 | 2551271000000118 |
| Pre-treatment serum cholesterol level | 1488851015 | 395153009 | 1488851015 |
| Serum HDL/non-HDL cholesterol ratio | 5519791000006113 | 271059008 | 405628014 |

| | | | |
|---|---|---|---|
| Serum cholesterol/HDL (high density lipoprotein) ratio | 8396611000006112 | 1015681000000109 | 2607531000000112 |
| Plasma LDL (low density lipoprotein) cholesterol level | 458314019 | 1010591000000104 | 2586471000000117 |
| Serum high density lipoprotein:non-high density lipoprotein cholesterol ratio | 144141000006119 | 1015271000000109 | 2565581000000114 |
| Serum low density lipoprotein cholesterol level | 259233017 | 1022191000000100 | 2566581000000116 |
| Cholesterol | 3877651000006116 | 84698008 | 140417016 |
| LDL cholesterol level | 857141000006116 | 857141000006100 | 857141000006116 |
| Serum cholesterol/LDL ratio | 6041011000006113 | 313991001 | 458253013 |
| Low density lipoprotein cholesterol measurement | 4217651000006116 | 113079009 | 186163011 |
| Plasma total cholesterol level | 1484985012 | 1017161000000104 | 2552911000000113 |
| Target cholesterol level | 1484928011 | 390896004 | 1484928011 |
| Target serum cholesterol | 107301000006115 | 107301000006104 | 107301000006115 |
| Total cholesterol measurement | 186091018 | 121868005 | 186091018 |
| Target serum non high density lipoprotein cholesterol level | 13602641000006117 | | |

#### Table S49. Atrial ablation Aurum codes

| Term | Medcode ID | SNOMED CT Concept ID | SNOMED CT Description ID |
|---|---|---|---|
| Open ablation of atrioventricular node | 271176014 | 175095005 | 271176014 |
| Percutaneous transluminal ablation of atrioventricular node | 271209012 | 175125003 | 271209012 |
| Percutaneous transluminal ablation of accessory pathway | 2694913017 | 428581004 | 2694913017 |
| Percutaneous transluminal ablation of conducting system of heart | 238631000006110 | 448042001 | 2900001016 |
| Percutaneous transluminal chemical mediated septal ablation | 357821000000110 | 437746009 | 3037266010 |
| Percutaneous transluminal ablation of wall of atrium | 376621000000113 | 428290007 | 2693316014 |
| Percutaneous transluminal laser ablation of vein | 385021000000117 | 609165007 | 2958779019 |
| Percutaneous transluminal ablation of atrial wall NEC | 488161000000110 | 428290007 | 2693316014 |
| Transluminal radiofreq ablation heart conducting system NEC | 998521000006116 | 448042001 | 2900001016 |
| Percutaneous transluminal ablation of congenital heart malformation | 1547441000006113 | 428391009 | 2692484013 |
| Percutaneous transluminal ablation of pulmonary vein to left atrium conducting system | 1587601000006117 | 707831001 | 3032482017 |
| Percutaneous transluminal ablation of atrial wall for atrial flutter | 1587611000006119 | 707832008 | 3032484016 |
| Percutaneous radiofrequency ablation of epicardium | 1686211000000116 | 609233003 | 2958816014 |
| Catheter ablation of tissue of heart | 2790601000006119 | 18286008 | 30881015 |
| Catheter ablation of lesion of heart | 3076931000006116 | 35823001 | 59752012 |
| Ablation | 3550661000006116 | 64597002 | 199515017 |
| Rotablation | 3567581000006119 | 65659003 | 2478792014 |
| Radiofrequency ablation procedure phase | 4422211000006110 | 129089003 | 207797015 |
| Ablation operation for arrhythmia | 5046591000006116 | 233159005 | 349323018 |
| Diathermy ablation operation for arrhythmia | 5046601000006112 | 233160000 | 349324012 |
| Cryoablation operation for arrhythmia | 5046611000006110 | 233161001 | 349325013 |
| Laser ablation operation for arrhythmia | 5046621000006119 | 233162008 | 349326014 |
| Radiofrequency ablation operation for arrhythmia | 5046631000006116 | 233163003 | 349327017 |
| Ablation - action | 5386721000006118 | 257729003 | 383646017 |
| Ablation | 5386731000006115 | 257729003 | 383645018 |
| Cryoablation - action | 5653621000006118 | 281609006 | 419659014 |
| Cryoablation | 5653641000006113 | 281609006 | 419661017 |
| Radiofrequency ablation device | 6360191000006115 | 371791001 | 1210387010 |
| Ablation frequency | 6450421000006111 | 386132006 | 1480316015 |
| Percutaneous transluminal coronary angioplasty by rotoablation | 6583111000006116 | 397193006 | 1776881019 |
| History of cardiac radiofrequency ablation | 7081241000006112 | 427951003 | 2694709018 |
| Percutaneous transluminal ablation | 7087871000006111 | 428389001 | 2693563010 |
| Ablation of atrioventricular node | 7091821000006115 | 428663009 | 2693280013 |
| CT guided laser ablation | 7111561000006115 | 429929004 | 3027495010 |
| CT guided ablation | 7117191000006114 | 430269006 | 3027505015 |
| Endocardial pulmonary vein ablation | 7142291000006112 | 431626004 | 706461000000113 |

| | | | |
|---|---|---|---|
| Endocardial ablation of pulmonary vein using fluoroscopic guidance | 7142281000006114 | 431626004 | 2772721014 |
| Percutaneous transluminal septal myocardial ablation | 7199391000006119 | 437746009 | 2793908011 |
| Catheter ablation for cardiac arrhythmia | 7484761000006112 | 473229000 | 2956223014 |
| Cardiac ultrasound ablation system | 7556141000006117 | 702209009 | 3014237014 |
| Cardiac radiofrequency ablation system | 7594051000006116 | 704706000 | 3014227016 |
| Cardiac ablation system | 7603631000006111 | 705732002 | 3024696011 |

#### Table S50. Hyperthyoidism Aurum codes

| Term | Medcode ID | SNOMED CT Concept ID | SNOMED CT Description ID |
|---|---|---|---|
| H/O: hyperthyroidism | 251587014 | 161442007 | 251587014 |
| Toxic adenomatous goitre | 3438831000006118 | 57777000 | 497989019 |
| Toxic uninodular goitre with thyrotoxic crisis | 3627481000006117 | 69329005 | 501258013 |
| Graves' eye disease | 5581851000006111 | 276177000 | 2536088017 |
| Thyrotoxicosis without mention of goitre or cause no crisis | 101891000006113 | 90739004 | 150375010 |
| Thyrotoxicosis with toxic single thyroid nodule | 3700961000006117 | 73869005 | 502554013 |
| Toxic multinodular goitre with crisis | 292342015 | 62278002 | 103507011 |
| Toxic diffuse goitre NOS | 292331012 | 267374005 | 398780011 |
| Toxic uninodular goitre NOS | 292338018 | 73869005 | 200396013 |
| Toxic multinodular goitre NOS | 292343013 | 26389007 | 196525014 |
| Thyrotoxicosis of other specified origin with no crisis | 292361016 | 90739004 | 150375010 |
| Thyrotoxicosis without mention of goitre or other cause | 292366014 | 90739004 | 150375010 |
| Graves' disease | 472452010 | 353295004 | 472452010 |
| Thyrotoxicosis from ectopic thyroid tissue | 3918281000006110 | 87232008 | 507482011 |
| Thyrotoxicosis due to uninodular goitre | 3700971000006112 | 73869005 | 502555014 |
| Thyrotoxicosis with or without goiter | 5721001000006113 | 286909009 | 426394012 |
| Thyrotoxicosis from ectopic thyroid nodule | 507481016 | 87232008 | 507481016 |
| Thyrotoxic exophthalmos | 33469013 | 19885005 | 33469013 |
| Toxic multinodular goitre with thyrotoxic storm | 3511901000006114 | 62278002 | 499238017 |
| Toxic multinodular goitre with thyrotoxic crisis | 3511871000006114 | 62278002 | 499237010 |
| Hyperthyroidism | 57561015 | 34486009 | 57561015 |
| Hyperthyroidism due to ectopic thyroid tissue | 4757781000006116 | 190255006 | 3035757011 |
| Thyrotoxicosis due to multinodular goitre | 2922501000006118 | 26389007 | 483220013 |
| History of hyperthyroidism | 4539511000006111 | 161442007 | 2986456017 |
| Toxic multinodular thyroid goitre | 2922531000006114 | 26389007 | 3036943012 |
| [RFC] Hyperthyroidism | 905741000006112 | 905741000006108 | 905741000006112 |
| Toxic diffuse goitre with crisis | 292330013 | 190242005 | 292330013 |
| Toxic diffuse goitre | 398780011 | 267374005 | 398780011 |
| Graves' ophthalmopathy | 5581801000006112 | 276177000 | 2536083014 |
| Toxic multinodular thyroid goiter | 2922521000006111 | 26389007 | 3036654016 |
| Toxic uninodular goitre | 200396013 | 73869005 | 200396013 |
| Thyrotoxicosis of other specified origin | 398782015 | 90739004 | 150375010 |
| Thyrotoxicosis with diffuse goiter | 5500071000006116 | 267374005 | 398778017 |
| Thyrotoxicosis of other specified origin NOS | 292365013 | 90739004 | 150375010 |
| Toxic uninodular goitre with thyrotoxic storm | 3627511000006113 | 69329005 | 200068013 |
| Toxic nodular goitre unspecified | 292346017 | 57777000 | 497988010 |
| Toxic multinodular goiter with thyrotoxic crisis | 3511881000006112 | 62278002 | 103507011 |
| Thyrotoxicosis due to uninodular goiter | 3700981000006110 | 73869005 | 502556010 |
| Thyrotoxicosis due to multinodular goiter | 2922481000006111 | 26389007 | 483218010 |

| | | | |
|---|---|---|---|
| Toxic multinodular goiter with thyrotoxic storm | 3511891000006110 | 62278002 | 103508018 |
| Toxic nodular goitre unspecified with no crisis | 292347014 | 57777000 | 96075015 |
| Diffuse toxic goitre | 5500051000006114 | 267374005 | 398776018 |
| Thyrotoxicosis from ectopic thyroid nodule with no crisis | 292355019 | 190255006 | 292355019 |
| Thyrotoxicosis from ectopic thyroid nodule with crisis | 292356018 | 190256007 | 292356018 |
| Toxic uninodular goitre with crisis | 292335015 | 69329005 | 115161014 |
| Toxic diffuse goitre with no crisis | 292327018 | 190241003 | 292327018 |
| Thyrotoxicosis with toxic multinodular goitre | 2922511000006115 | 26389007 | 483221012 |
| Toxic nodular goiter | 3438791000006113 | 57777000 | 96075015 |
| Thyrotoxicosis without mention of goitre, cause with crisis | 101911000006110 | 90739004 | 150375010 |
| Toxic nodular goitre NOS | 292354015 | 57777000 | 497988010 |
| Thyrotoxicosis NOS | 292372014 | 90739004 | 150375010 |
| Toxic uninodular thyroid goitre | 3700991000006113 | 73869005 | 3035712019 |
| Toxic multinodular goiter | 2922461000006118 | 26389007 | 44198012 |
| Graves' dermopathy | 5112881000006118 | 237825005 | 356407011 |
| Toxic goitre | 355932011 | 237498007 | 355932011 |
| Toxic multinodular goitre | 196525014 | 26389007 | 196525014 |
| Thyrotoxicosis with toxic multinodular goiter | 2922491000006114 | 26389007 | 483219019 |
| Thyrotoxicosis +/- goitre | 881371000006111 | 286909009 | 881371000006111 |
| Toxic diffuse thyroid goitre | 5500101000006114 | 267374005 | 3037800012 |
| Thyrotoxic myopathy | 3613541000006112 | 68437005 | 113658014 |
| Thyrotoxicosis with or without goitre | 5720991000006112 | 286909009 | 426395013 |
| Thyrotoxic crisis | 2966801000006110 | 29028009 | 48598011 |
| Hyperthyroidism resolved | 2548990010 | 416477006 | 2548990010 |
| Thyrotoxicosis from ectopic thyroid nodule NOS | 292357010 | 87232008 | 144647016 |
| Thyrotoxicosis of other specified origin with crisis | 292362011 | 90739004 | 150375010 |
| Thyrotoxicosis with diffuse goitre | 5500041000006112 | 267374005 | 398775019 |
| Toxic multinodular goiter with no crisis | 4757741000006110 | 190247004 | 292340011 |
| Hyperthyroidism due to ectopic thyroid nodule | 3918261000006117 | 87232008 | 144647016 |
| Thyrotoxic heart disease | 100438013 | 60446003 | 100438013 |
| Thyrotoxicosis | 150375010 | 90739004 | 150375010 |
| Toxic uninodular goitre with no crisis | 292334016 | 190244006 | 292334016 |
| Toxic multinodular goitre with no crisis | 292339014 | 190247004 | 292339014 |
| Toxic nodular goitre unspecified with crisis | 292350012 | 57777000 | 96075015 |

#### Table S51. Hypothyroidism Aurum codes

| Term | Medcode ID | SNOMED CT Concept ID | SNOMED CT Description ID |
|---|---|---|---|
| Myasthenic syndrome due to hypothyroidism | 297588017 | 193212008 | 297588017 |
| Congenital hypothyroidism with diffuse goitre | 415432015 | 278503003 | 415432015 |
| H/O: hypothyroidism | 251589012 | 161443002 | 251589012 |
| Hypothyroidism annual review | 637341000000118 | 341861000000109 | 637341000000118 |
| Hypothyrotropic hypothyroidism | 3843711000006117 | 82598004 | 505170011 |
| Congenital hypothyroidism with diffuse goiter | 5614081000006117 | 278503003 | 415433013 |
| Postsurgical hypothyroidism | 2933341000006117 | 27059002 | 2536050019 |
| Primary hypothyroidism AND adrenocortical insufficiency | 3862381000006114 | 83728000 | 505751010 |
| Other postablative hypothyroidism | 292394018 | 237527007 | 355975017 |
| Postablative hypothyroidism NOS | 292396016 | 237527007 | 355975017 |
| Cerebral degeneration due to myxoedema | 542671000006117 | 192816007 | 297016016 |
| Diabetes mellitus, Addison's disease and myxoedema | 3862341000006115 | 83728000 | 505747013 |
| Myxoedema | 493538010 | 43153006 | 493538010 |
| Endemic cretinism - neurological type | 5108961000006115 | 237566004 | 356025012 |
| Total substernal thyroidectomy | 858351000006117 | 52826006 | 858351000006117 |
| Complete thyroidectomy | 40998019 | 24443003 | 40998019 |
| Other acquired hypothyroidism | 12729831000006113 | 654491000000108 | 1438601000000113 |
| Hypothyroidism with positive thyroid antibodies | 5108381000006112 | 237519003 | 355965015 |
| Hypothyroidism-congen.+ acqui. | 881381000006114 | 111566002 | 881381000006114 |
| Primary hypothyroidism | 4197341000006114 | 111566002 | 363808017 |
| Congenital hypothyroidism without goitre | 355957018 | 237515009 | 355957018 |
| Autoimmune myxoedema | 355964016 | 237519003 | 355964016 |
| Post-infectious hypothyroidism | 5108491000006117 | 237528002 | 355976016 |
| Cretinism | 600791000006112 | 217710005 | 329968011 |
| Irradiation hypothyroidism | 292395017 | 190277005 | 292395017 |
| Hypothyroidism resulting from phenylbutazone | 292402012 | 190283008 | 292402012 |
| Acquired hypothyroidism | 178809013 | 111566002 | 178809013 |
| Hypothyroidism monitoring verbal invite | 2548835019 | 416326003 | 2548835019 |
| Congenital hypothyroidism NOS | 398783013 | 190268003 | 292373016 |
| Postoperative hypothyroidism | 2933321000006112 | 27059002 | 2532363015 |
| Hypothyroidism monitoring administration | 940091000006115 | 940091000006104 | 940091000006115 |
| Hypothyroidism monitoring invite 2 | 940111000006112 | 940111000006108 | 940111000006112 |
| Hypothyroidism monitoring invite 3 | 940121000006116 | 940121000006100 | 940121000006116 |
| Hypothyroidism monitoring administration | 225971000000116 | 713741000000106 | 1564981000000116 |
| Hypothyroidism monitoring telephone invitation | 406721000000115 | 248341000000100 | 406721000000115 |
| Iatrogenic hypothyroidism | 292400016 | 88273006 | 146340011 |
| Iatrogenic hypothyroidism NOS | 292404013 | 88273006 | 146340011 |
| Myxoedema encephalopathy | 4768951000006118 | 192816007 | 297016016 |
| Other acquired hypothyroidism | 292408011 | 111566002 | 178809013 |

| | | | |
|---|---|---|---|
| Hypothyroidism clinical management plan | 2474336017 | 736287007 | 3517030018 |
| Post-surgical hypothyroidism | 2536050019 | 27059002 | 45271010 |
| Autoimmune hypothyroidism | 5108351000006116 | 237519003 | 355966019 |
| Acquired hypothyroidism NOS | 881391000006112 | 654491000000108 | 881391000006112 |
| Hypothyroidism resulting from para-aminosalicylic acid | 292401017 | 190282003 | 292401017 |
| History of hypothyroidism | 4539531000006117 | 161443002 | 2986407018 |
| Endemic cretinism - mixed type | 5108941000006119 | 237565000 | 356023017 |
| Hypothyroidism NOS | 12727341000006116 | 681051000000104 | 1492161000000113 |
| Hypothyroidism confirmed | 1823931000006116 | 1823931000006100 | 1823931000006116 |
| [X]Systemic atrophy affecting the CNS in myxoedema | 428591000006112 | 23853001 | 40063015 |
| Postinfectious hypothyroidism | 211321000006115 | 237528002 | 355976016 |
| Congenital hypothyroidism without goiter | 5108301000006115 | 237515009 | 355958011 |
| Congenital iodine-deficiency syndrome, myxoedematous type | 583291000006115 | 75065003 | 502897012 |
| Atrophy of thyroid - acquired | 5108811000006113 | 237558008 | 356013014 |
| Hypothyroidism resulting from resorcinol | 292403019 | 190284002 | 292403019 |
| Congenital thyroid insufficiency | 588661000006115 | 190268003 | 292373016 |
| Myopathy due to myxoedema | 682701000006117 | 87844004 | 507778013 |
| Complete parathyroidectomy | 2696261000006111 | 12330002 | 21229013 |
| Hypothyroidism monitoring second letter | 2533534011 | 414441001 | 2533534011 |
| Hypothyroidism medication review | 939981000006114 | 939981000006105 | 939981000006114 |
| Hypothyroidism monitoring invite 1 | 940101000006114 | 940101000006105 | 940101000006114 |
| Hypothyroidism with sensorineural deafness | 3644031000006116 | 70348004 | 116847011 |
| Pretibial myxoedema - hypothyroid | 208391000006113 | 40930008 | 68268011 |
| Thyroid insufficiency | 101581000006112 | 40930008 | 68268011 |
| Acquired hypothyroidism NOS | 988831000006119 | 111566002 | 988831000006119 |
| Goitrous cretin | 415431010 | 278503003 | 415431010 |
| Endemic cretinism | 12704331000006114 | 75065003 | 124678011 |
| Myxoedema coma | 35727012 | 21263006 | 35727012 |
| Acquired thyroid atrophy | 5108831000006119 | 237558008 | 3037562011 |
| Hypothyroidism NOS | 398784019 | 40930008 | 68268011 |
| Congenital hypothyroidism | 292373016 | 190268003 | 292373016 |
| Thyroid atrophy | 292448015 | 190309006 | 292448015 |
| Other specified congenital hypothyroidism | 292376012 | 190268003 | 292373016 |
| Myxoedema myopathy | 3927661000006113 | 87844004 | 507778013 |
| Total thyroidectomy | 40996015 | 24443003 | 40996015 |
| Cretinism | 329968011 | 217710005 | 329968011 |
| Hypothyroidism monitoring third letter | 2533420013 | 414442008 | 2533420013 |
| Hypothyroidism monitoring first letter | 2533422017 | 414439002 | 2533422017 |
| Hypothyroidism after surgery | 2933351000006115 | 27059002 | 3037380018 |
| Cerebellar ataxia due to myxoedema | 542301000006117 | 192876003 | 297115015 |
| [X]Other sp cified hypothyroidism | 413721000006119 | 40930008 | 68268011 |

| | | | |
|---|---|---|---|
| Myxoedema cerebellar degeneration | 4769091000006117 | 192876003 | 297115015 |
| Congenital hypothyroidism not due to iodine deficiency | 4904391000006111 | 217710005 | 329969015 |
| Hypothyroidism clinical management plan no longer in place | 1877411000006112 | 1877411000006108 | 1877411000006112 |
| Endemic cretinism - hypothyroid | 3720391000006119 | 75065003 | 502894017 |
| Acquired atrophy of thyroid | 356012016 | 237558008 | 356012016 |
| Aplasia of thyroid with myxoedema | 4012861000006111 | 92978002 | 510171015 |
| Thyroid deficiency | 101171000006116 | 40930008 | 68268011 |
| Postablative hypothyroidism | 216181000006112 | 237527007 | 355975017 |
| Hypothyroidism | 68268011 | 40930008 | 68268011 |
| Iodine hypothyroidism | 292397013 | 190279008 | 292397013 |

#### Table S52. Chronic obstructive pulmonary disease Aurum codes

| Term | Medcode ID | SNOMED CT Concept ID | SNOMED CT Description ID |
|---|---|---|---|
| COPD GOLD group C | 11932351000006110 | 11932351000006106 | 11932351000006110 |
| COPD GOLD group D | 11932361000006112 | 11932361000006108 | 11932361000006112 |
| Chronic asthmatic bronchitis | 301450011 | 195949008 | 301450011 |
| Chronic wheezy bronchitis | 301451010 | 195949008 | 301451010 |
| Chronic bullous emphysema | 301460019 | 195957006 | 301460019 |
| Segmental bullous emphysema | 301463017 | 195958001 | 301463017 |
| Zonal bullous emphysema | 301464011 | 195959009 | 301464011 |
| Access to online patient COPD education given | 1916531000006115 | 1916531000006104 | 1916531000006115 |
| Mild chronic obstructive pulmonary disease | 457168017 | 313296004 | 457168017 |
| Moderate chronic obstructive pulmonary disease | 457169013 | 313297008 | 457169013 |
| Severe chronic obstructive pulmonary disease | 457171013 | 313299006 | 457171013 |
| COPD (chronic obstructive pulmonary disease) 6 monthly review | 8235131000006118 | 760621000000103 | 1710191000000113 |
| COPD self-management plan review | 1811741000006117 | 810951000000101 | 2115731000000110 |
| On COPD (chr obstruc pulmonary disease) supportv cre pathway | 1813881000006119 | 826111000000109 | 2155811000000119 |
| Asthma-chronic obstructive pulmonary disease overlap syndrom | 1948051000006112 | 10692761000119107 | 3046456013 |
| Chron obstruct pulmonary dis wth acute exacerbation, unspec | 553211000006119 | 195951007 | 301453013 |
| Chr. airway obstruction NOS | 990651000006112 | 13645005 | 990651000006112 |
| Chronic obstructive pulmonary disease | 475431013 | 13645005 | 475431013 |
| Admit COPD emergency | 12489801000006116 | 408501008 | 2160051010 |
| Telehealth chronic obstructive pulmonary disease monitoring | 2423731000000115 | 716358000 | 3305683010 |
| COPD (chronic obstructive pulmonary disease) disturbs sleep | 8058301000006110 | 198401000000104 | 1677391000000113 |
| Chronic bronchitis, acute exac | 851261000006116 | 851261000006100 | 851261000006116 |
| Simple chronic bronchitis NOS | 301444018 | 61937009 | 102938011 |
| Other chronic bronchitis NOS | 301458016 | 63480004 | 105519017 |
| Chronic bullous emphysema NOS | 301468014 | 195957006 | 301460019 |
| COPD patient unsuitable for pulmonary rehabilitation | 1656601000006119 | 1656601000006103 | 1656601000006119 |
| Giant bullous emphysema | 27096010 | 16003001 | 27096010 |
| Shared care COPD (chronic obstructive pulmonary disease) monitoring | 8316481000006116 | 897311000000101 | 2312551000000112 |
| Panacinar emphysema | 2578881000006118 | 4981000 | 9336013 |
| Chronic obstructive pulmonary disease disturbs sleep | 299001000000116 | 198401000000104 | 299001000000116 |
| Obliterative bronchiolitis due to chemical fumes | 269761000006112 | 196027008 | 301573017 |
| Not suitable for step down change in COPD management plan | 2009511000006113 | 2009511000006109 | 2009511000006113 |
| Acute non-infective exacerbation of chronic obstructive pulmonary disease | 2010061000006113 | 847091000000104 | 2196421000000114 |
| COPD (chronic obstructive pulmonary disease) care pathway | 8288261000006119 | 848431000000106 | 2199291000000116 |
| ACOS - asthma-chronic obstructive pulmonary disease overlap syndrome | 9317341000006115 | 10692761000119107 | 2496071000000117 |

| | | | |
|---|---|---|---|
| Acute exacerbation of COPD | 4781421000006111 | 195951007 | 3012683010 |
| Obstructive chronic bronchitis | 4733021000006111 | 185086009 | 285104011 |
| MacLeod's unilateral emphysema | 1230190015 | 45145000 | 1230190015 |
| Seen in chronic obstructive pulmonary disease clinic | 1839331000006117 | 1839331000006101 | 1839331000006117 |
| QOF (Quality and Outcomes Framework) chronic obstructive pulmonary disease quality indicator-related care invitation | 12626281000006113 | 1110861000000102 | 2779701000000115 |
| Chronic obstructive pulmonary disease annual review | 1488424013 | 394703002 | 1488424013 |
| Chronic obstructive pulmonary disease leaflet given | 1488856013 | 395159008 | 1488856013 |
| Chronic obstructive pulmonary disease monitoring by nurse | 1780380013 | 401184000 | 1780380013 |
| Chronic obstructive pulmonary disease monitoring by doctor | 1780381012 | 401185004 | 1780381012 |
| Centrilobular emphysema | 113497011 | 68328006 | 113497011 |
| COB - Chronic obstructive bronchitis | 4733011000006115 | 185086009 | 285103017 |
| COPD (chronic obstructive pulmonary disease) management plan declined | 8312921000006111 | 892321000000109 | 2297721000000117 |
| Chronic obstructive pulmonary disease severity | 1856571000006116 | 1856571000006100 | 1856571000006116 |
| Chronic bronchitis with emphysema | 4732991000006119 | 185086009 | 285101015 |
| Chronic emphysema caused by chemical fumes | 4781821000006113 | 196026004 | 3296151011 |
| COPD follow-up | 845451000006118 | 394702007 | 1488423019 |
| COPD structured smoking assessment declined | 1704581000006119 | 375911000000102 | 739421000000116 |
| [RFC] Chronic obstructive pulmonary disease (COPD) | 909711000006111 | 909711000006107 | 909711000006111 |
| [RFC] Emphysema | 909721000006115 | 909721000006104 | 909721000006115 |
| Atrophic (senile) emphysema | 396109014 | 266356006 | 396109014 |
| 1 COPD exacerbation in past year | 1882421000006113 | 1882421000006109 | 1882421000006113 |
| 3+ COPD exacerbations in past year | 1882431000006111 | 1882431000006107 | 1882431000006111 |
| 2 COPD exacerbations in past year | 1882441000006118 | 1882441000006102 | 1882441000006118 |
| Interstitial emphysema | 12704691000006117 | 77690003 | 770361000006117 |
| COLD - Chronic obstructive lung disease | 2716351000006113 | 13645005 | 475430014 |
| COPD (Chronic obstructive pulmonary disease) patient unsuitable for pulmonary rehabilitation | 8120981000006112 | 371611000000107 | 730041000000110 |
| COPD (chronic obstructive pulmonary disease) written self management plan declined | 8294671000006116 | 857811000000102 | 2219931000000113 |
| COPD medication review | 939991000006112 | 939991000006108 | 939991000006112 |
| COPD monitoring administration | 940131000006118 | 940131000006102 | 940131000006118 |
| COPD monitoring invite 2 | 940151000006113 | 940151000006109 | 940151000006113 |
| COPD monitoring invite 3 | 940161000006110 | 940161000006106 | 940161000006110 |
| Chronic obstructive pulmonary disease monitoring verb invite | 967571000006116 | 716281000000103 | 1568831000000118 |
| Emergency COPD admission since last appointment | 977901000006111 | 414087000 | 2533539018 |
| Refer COPD structured smoking assessment - enhanc serv admin | 1704561000006112 | 375851000000108 | 739301000000115 |
| COPD structured smoking assessment declined - enh serv admin | 1704571000006117 | 375911000000102 | 739421000000116 |
| Mucopurulent chronic bronchitis NOS | 301448015 | 74417001 | 123588010 |
| End stage chronic obstructive pulmonary disease | 4510801000006114 | 135836000 | 3034045017 |

| | | | |
|---|---|---|---|
| Chronic obstructive pulmonary disease 3 monthly review | 1683181000000112 | 760601000000107 | 1683181000000112 |
| Bullous emphysema with collapse | 396108018 | 266355005 | 396108018 |
| Preferred place of care for next exacerbation of COPD | 1784071000006113 | 789661000000102 | 1765431000000114 |
| GP OOH service notified of COPD care plan | 1785171000006113 | 783631000000109 | 1752261000000117 |
| Health education - chronic obstructive pulmonary disease | 405121000000111 | 741056003 | 3543021016 |
| Chronic obstructive pulmonary disease care pathway | 2199261000000110 | 848431000000106 | 2199261000000110 |
| COPD (chronic obstructive pulmonary disease) rescue pack declined | 8294651000006114 | 857791000000103 | 2219871000000119 |
| Interstitial emphysema | 19421011 | 77690003 | 503646013 |
| Optimization of medication for chronic obstructive lung disease | 7484341000006112 | 473202005 | 2956280014 |
| Step down change in chronic obstructive pulmonary disease management plan | 3011135010 | 704123000 | 3011135010 |
| Suitable for step down change in COPD management plan | 2009501000006110 | 2009501000006106 | 2009501000006110 |
| Acute infective exacerbation of chronic obstructive airways disease | 424365019 | 285381006 | 424365019 |
| Chronic obstructive pulmonary disease post discharge review | 2009451000006113 | 2009451000006109 | 2009451000006113 |
| Acute non-infective exacerbation of COPD (chronic obstructive pulmonary disease) | 8287171000006115 | 847091000000104 | 2196431000000111 |
| COPD self-management plan given | 839001000006111 | 390891009 | 1484924013 |
| [X]Other emphysema | 301835010 | 87433001 | 144964015 |
| Other emphysema NOS | 396110016 | 87433001 | 144964015 |
| COPD GOLD group A | 11932331000006115 | 11932331000006104 | 11932331000006115 |
| Chr. airway obstruction NOS | 885301000006119 | 611541000000106 | 885301000006119 |
| Acute exacerbation of chronic obstructive airways disease | 301453013 | 195951007 | 301453013 |
| Chronic obstructive pulmonary disease monitoring 1st letter | 189761000000117 | 716241000000106 | 1568801000000112 |
| Issue of COPD (chronic obstructive pulmonary disease) rescue pack | 8219501000006114 | 718241000000107 | 1573141000000119 |
| Shared care chronic obstructive pulmonary disease monitoring | 1882371000006118 | 1882371000006102 | 1882371000006118 |
| COPD (chronic obstructive pulmonary disease) does not disturb sleep | 8058321000006117 | 198411000000102 | 1677401000000111 |
| GP (general practitioner) OOH (out of hours) service notified of COPD (chronic obstructive pulmonary disease) care plan | 8244871000006114 | 783631000000109 | 1752291000000111 |
| Fetid chronic bronchitis | 3873191000006110 | 84409004 | 139979010 |
| Chronic catarrhal bronchitis | 508562012 | 89549007 | 508562012 |
| COPD (chronic obstructive pulmonary disease) 3 monthly review | 8235101000006114 | 760601000000107 | 1709911000000112 |
| COPD clinical pathway protocol followed | 1824071000006115 | 1824071000006104 | 1824071000006115 |
| Chronic obstruct pulmonary dis with acute lower resp infectn | 555461000006119 | 196001008 | 301542016 |
| Vesicular emphysema | 2578901000006116 | 4981000 | 9338014 |
| Chronic obstructive pulmonary disease monitoring due | 1484971019 | 390941006 | 1484971019 |
| Chronic obstructive pulmonary disease follow-up | 1488423019 | 394702007 | 1488423019 |
| Interstitial pulmonary emphysema | 3764031000006119 | 77690003 | 3764031000006119 |
| Mixed simple and mucopurulent chronic bronchitis | 301456017 | 195953005 | 301456017 |

| | | | |
|---|---|---|---|
| Acute vesicular emphysema | 301470017 | 195963002 | 301470017 |
| Chronic obstructive bronchitis | 4733031000006114 | 185086009 | 2668815014 |
| Suspected chronic obstructive pulmonary disease | 314261000000118 | 204991000000107 | 314261000000118 |
| Optimisation of medication for chronic obstructive lung disease | 7484351000006114 | 473202005 | 3297724010 |
| Chronic obstructive airways disease | 555471000006114 | 13645005 | 475423015 |
| Emphysema | 640491000006111 | 87433001 | 144964015 |
| Chronic obstructive pulmonary disease monitoring admin | 967531000006119 | 713731000000102 | 1564971000000118 |
| Bullous emphysema | 4781471000006112 | 195957006 | 301462010 |
| Referral to COPD community nursing team | 1784371000006111 | 790301000000101 | 1766811000000114 |
| Chronic obstructive pulmonary disease clini management plan | 966841000006111 | 736283006 | 3517019016 |
| Chronic obst. pulm. dis. NOS | 885311000006116 | 611541000000106 | 885311000006116 |
| Emphysema of lung | 3921361000006112 | 87433001 | 144967010 |
| Other chronic bronchitis | 301457014 | 63480004 | 105519017 |
| Chronic bronchitis NOS | 301459012 | 63480004 | 105519017 |
| [X]Other specified chronic obstructive pulmonary disease | 301836011 | 13645005 | 475431013 |
| Seen in chronic obstructive pulmonary disease clinic | 2219551000000113 | 857661000000104 | 2219551000000113 |
| Very severe chronic obstructive pulmonary disease | 516801000000112 | 293991000000106 | 516801000000112 |
| Chronic emphysema due to chemical fumes | 301572010 | 196026004 | 301572010 |
| Chronic bronchitis | 105519017 | 63480004 | 105519017 |
| End stage chronic obstructive airways disease | 216596014 | 135836000 | 216596014 |
| Has chronic obstructive pulmonary disease care plan | 2152091000000112 | 827571000000106 | 2152091000000112 |
| Seen in COPD (chronic obstructive pulmonary disease) clinic | 8294561000006110 | 857661000000104 | 2219581000000119 |
| Centriacinar emphysema | 3611791000006110 | 68328006 | 113496019 |
| Chronic obstructive lung disease | 2716231000006114 | 13645005 | 23287019 |
| Emergency hospital admission for chronic obstructive pulmonary disease | 6763231000006119 | 408501008 | 6763231000006119 |
| Panlobular emphysema | 9337016 | 4981000 | 9337016 |
| Shared care chronic obstructive pulmonary disease monitoring | 2308511000000113 | 897311000000101 | 2308511000000113 |
| Chronic obstructive airways disease NOS | 301545019 | 13645005 | 475428012 |
| Obstructive chronic bronchitis NOS | 301455018 | 185086009 | 285100019 |
| Emphysema NOS | 301477019 | 87433001 | 144964015 |
| Chronic obstructive pulmonary disease rescue pack declined | 2219861000000114 | 857791000000103 | 2219861000000114 |
| Eosinophilic bronchitis | 2240631000000119 | 866901000000103 | 2240631000000119 |
| COPD monitoring invite 1 | 940141000006111 | 940141000006107 | 940141000006111 |
| COPD patient unsuitable for pulmonary rehabilitation | 1704541000006113 | 371611000000107 | 1152601000000114 |
| Other emphysema | 301469018 | 87433001 | 144964015 |
| Chronic tracheobronchitis | 87480013 | 52571006 | 87480013 |
| COPD - Chronic obstructive pulmonary disease | 2716321000006116 | 13645005 | 475427019 |
| Unilateral emphysema | 3228301000006115 | 45145000 | 75281018 |

| | | | |
|---|---|---|---|
| Chronic obstructive airways disease NOS | 12728161000006116 | 611541000000106 | 1352161000000113 |
| Fetid chronic bronchitis | 139979010 | 84409004 | 506053014 |
| Obstructive chronic bronchitis | 285104011 | 185086009 | 285100019 |
| Referred for COPD structured smoking assessment | 1704551000006110 | 375851000000108 | 739301000000115 |
| COPD GOLD group B | 11932341000006113 | 11932341000006109 | 11932341000006113 |
| Chronic obstructive pulmonary disease monitoring in primary care | 1856491000006119 | 1856491000006103 | 1856491000006119 |
| Chronic obstructive pulmonary disease monitoring secondary care | 1856501000006110 | 1856501000006106 | 1856501000006110 |
| Chronic obstructive pulmonary disease multidisciplinary review | 1771201000006116 | 1771201000006100 | 1771201000006116 |
| Referral to COPD (chronic obstructive pulmonary disease) community nursing team | 8249051000006111 | 790301000000101 | 1766841000000110 |
| At risk of COPD (chronic obstructive pulmonary disease) exacerbation | 8125721000006119 | 383611000000102 | 756791000000118 |
| Chronic obstructive pulmonary disease confirmed | 1823851000006119 | 1823851000006103 | 1823851000006119 |
| Acute exacerbation of chronic obstructive pulmonary disease | 4781431000006114 | 195951007 | 3012710012 |
| Issue of chronic obstructive pulmonary disease rescue pack | 1573111000000115 | 718241000000107 | 1573111000000115 |
| Emergency hospital admission for COPD (chronic obstructive pulmonary disease) | 6763251000006114 | 408501008 | 3082978011 |
| History of chronic obstructive airway disease | 5516841000006119 | 270473001 | 2986462010 |
| Interstitial emphysema of lung | 3764021000006117 | 77690003 | 128944012 |
| Chronic obstructive pulmonary disease follow-up assessment | 1856591000006115 | 1856591000006104 | 1856591000006115 |
| COPD accident and emergency attendance since last visit | 977891000006112 | 413845009 | 2533538014 |
| Multiple COPD emergency hospital admissions | 998281000006115 | 198901000000105 | 300201000000114 |
| Chronic obstructive pulmonary disease monitoring 3rd letter | 199481000000116 | 717521000000104 | 1570061000000113 |
| Chronic obst. pulm. dis. NOS | 990641000006110 | 13645005 | 990641000006110 |
| Number of COPD exacerbations in past year | 977911000006114 | 723245007 | 3335171010 |
| On chronic obstructive pulmonary disease supprtv cre pathway | 1813871000006117 | 826111000000109 | 2149041000000110 |
| Other specified chronic obstructive airways disease | 301539010 | 13645005 | 475428012 |
| Purulent chronic bronchitis | 506053014 | 84409004 | 506053014 |
| Chronic obstructive pulmonary disease monitoring | 1484924013 | 390891009 | 1484924013 |
| Chronic obstructive pulmonary disease 6 monthly review | 1683221000000119 | 760621000000103 | 1683221000000119 |
| Alveolar emphysema of lung | 2578911000006118 | 4981000 | 9339018 |
| COPD management plan declined | 1880061000006110 | 1880061000006106 | 1880061000006110 |
| COPD patient unsuitable for pulmonary rehab - enh serv admin | 1704531000006115 | 371611000000107 | 1152601000000114 |
| COPD self-management plan agreed | 1811661000006110 | 811961000000106 | 2117851000000118 |
| Admit COPD emergency | 2160051010 | 408501008 | 3082901016 |
| COPD (chronic obstructive pulmonary disease) self-management plan review | 8260631000006112 | 810951000000101 | 2115741000000118 |
| Asthma-COPD overlap syndrome (ACOS) | 9317331000006113 | 10692761000119107 | 3046475015 |
| Chronic obstructive pulmonary disease monitor phone invite | 967581000006118 | 716901000000101 | 1569451000000115 |
| Optimization of medication for chronic obstructive pulmonary disease | 7484331000006119 | 473202005 | 2956201010 |

| | | | |
|---|---|---|---|
| Simple chronic bronchitis | 508561017 | 61937009 | 102938011 |
| Other specified chronic obstructive pulmonary disease | 1222334016 | 13645005 | 475431013 |
| Chronic obstructive pulmonary disease NOS | 1222335015 | 13645005 | 475431013 |
| Chronic obstructive pulmonary disease monitoring 2nd letter | 198471000000111 | 717021000000106 | 1569571000000113 |
| Chronic obstructive pulmonary disease NOS | 12759361000006112 | 611541000000106 | 1352151000000110 |
| Emphysematous bulla | 4781461000006117 | 195957006 | 301461015 |
| Mucopurulent chronic bronchitis | 123588010 | 74417001 | 123588010 |
| GP out of hours notified of COPD care plan | 1763751000006117 | 1763751000006101 | 1763751000006117 |
| History of chronic obstructive pulmonary disease | 1765681000000110 | 270473001 | 1765681000000110 |
| Emphysematous bronchitis | 285100019 | 185086009 | 285100019 |

#### Table S53. Diabetic retinopathy Aurum codes

| Term | Medcode ID | SNOMED CT Concept ID | SNOMED CT Description ID |
|---|---|---|---|
| Moderate nonproliferative diabetic retinopathy of left eye | 12085211000006112 | 769186002 | 3689288014 |
| O/E - sight threatening diabetic retinopathy | 2549896013 | 417677008 | 2549896013 |
| Diabetic retinopathy clinic | 7567371000006117 | 702850009 | 3006342012 |
| Left proliferative diabetic retinopathy | 856621000006118 | 856621000006102 | 856621000006118 |
| Left non-proliferative diabetic retinopathy | 857031000006113 | 857031000006109 | 857031000006113 |
| Right proliferative diabetic retinopathy | 857411000006110 | 857411000006106 | 857411000006110 |
| Right non-proliferative diabetic retinopathy | 857971000006110 | 857971000006106 | 857971000006110 |
| On examination - right eye background diabetic retinopathy | 6761781000006114 | 408409007 | 2621799017 |
| Diabetic retinopathy screening offered | 12121381000006114 | 1103701000000102 | 2763011000000112 |
| Proliferative diabetic retinopathy - high risk | 6022721000006112 | 312907002 | 456707011 |
| On examination - right eye preproliferative diabetic retinopathy | 6761821000006115 | 408411003 | 2621610013 |
| DR - Diabetic retinopathy | 2576431000006117 | 4855003 | 1230610015 |
| Proliferative diabetic retinopathy | 98476015 | 59276001 | 98476015 |
| On examination - left eye stable treated proliferative diabetic retinopathy | 6866541000006117 | 414894003 | 2621811014 |
| Severe NPDR (nonproliferative diabetic retinopathy) | 12224761000006115 | 312905005 | 3691865016 |
| High risk proliferative diabetic retinopathy | 1785163015 | 312907002 | 1785163015 |
| O/E - right eye proliferative diabetic retinopathy | 2159977011 | 408413000 | 2159977011 |
| O/E - left eye proliferative diabetic retinopathy | 2159978018 | 408414006 | 2159978018 |
| Nonproliferative diabetic retinopathy | 6515171000006112 | 390834004 | 2643005014 |
| On examination - right eye stable treated proliferative diabetic retinopathy | 6866901000006111 | 414910007 | 2621810010 |
| Mild nonproliferative diabetic retinopathy | 6022611000006112 | 312903003 | 2921114017 |
| Mild nonproliferative diabetic retinopathy of right eye | 12085071000006114 | 769183005 | 3689271019 |
| Preproliferative diabetic retinopathy of left eye | 12085051000006116 | 769182000 | 3689267017 |
| Very severe proliferative diabetic retinopathy | 6630581000006113 | 399865004 | 1779161016 |
| O/E - left eye stable treated proliferative diabetic retinopathy | 975261000006113 | 414894003 | 2532964019 |
| Severe nonproliferative diabetic retinopathy of right eye | 12085251000006113 | 769187006 | 3689295017 |
| Moderate nonproliferative diabetic retinopathy | 6022621000006116 | 312904009 | 1775724010 |
| Proliferative diabetic retinopathy - non high risk | 6022671000006115 | 312906006 | 456706019 |
| Left laser treated diabetic retinopathy | 856631000006115 | 856631000006104 | 856631000006115 |
| Left preproliferative diabetic retinopathy | 857051000006118 | 857051000006102 | 857051000006118 |
| Right laser treated diabetic retinopathy | 857421000006119 | 857421000006103 | 857421000006119 |
| Moderate non-proliferative diabetic retinopathy of left eye | 12085231000006118 | 769186002 | 3689292019 |
| Mild nonproliferative diabetic retinopathy of left eye | 12085121000006114 | 769184004 | 3689282010 |
| Proliferative diabetic retinopathy - quiescent | 6022761000006118 | 312908007 | 456708018 |

| | | | |
|---|---|---|---|
| Mild non-proliferative diabetic retinopathy of left eye | 12085141000006119 | 769184004 | 3689278013 |
| Moderate nonproliferative diabetic retinopathy of right eye | 12085171000006110 | 769185003 | 3689286013 |
| Severe nonproliferative diabetic retinopathy with no macular oedema | 6630751000006114 | 399873008 | 1773465013 |
| Severe nonproliferative diabetic retinopathy of left eye | 12085291000006119 | 769188001 | 3689299011 |
| Background diabetic retinopathy | 1785332013 | 390834004 | 1785332013 |
| O/E - right eye background diabetic retinopathy | 2159973010 | 408409007 | 2159973010 |
| Diabetic retinopathy 6 month review | 2159949019 | 408385003 | 2159949019 |
| Very severe nonproliferative diabetic retinopathy | 6630851000006115 | 399876000 | 1779167017 |
| Impaired vision due to diabetic retinopathy | 733161000000116 | 373041000000101 | 733161000000116 |
| On examination - left eye preproliferative diabetic retinopathy | 6761841000006110 | 408412005 | 2621611012 |
| O/E - right eye stable treated proliferative diabetic retinopathy | 975251000006111 | 414910007 | 2532963013 |
| On examination - left eye proliferative diabetic retinopathy | 6761881000006116 | 408414006 | 2621613010 |
| On examination - left eye background diabetic retinopathy | 6761801000006113 | 408410002 | 2621800018 |
| PPDR - Preproliferative diabetic retinopathy | 4771421000006116 | 193349004 | 2579151010 |
| Diabetic retinopathy detected by national screening programme | 8242331000006114 | 775841000000109 | 1733071000000112 |
| Preproliferative diabetic retinopathy | 297754014 | 193349004 | 297754014 |
| Diabetic retinopathy NOS | 297758012 | 4855003 | 9093013 |
| Laser treated diabetic retinopathy | 856611000006114 | 856611000006105 | 856611000006114 |
| O/E - left eye background diabetic retinopathy | 2159974016 | 408410002 | 2159974016 |
| O/E - right eye preproliferative diabetic retinopathy | 2159975015 | 408411003 | 2159975015 |
| NPDR - Non proliferative diabetic retinopathy | 6515201000006111 | 390834004 | 2579521014 |
| BDR - Background diabetic retinopathy | 6515211000006114 | 390834004 | 2621399015 |
| Moderate non-proliferative diabetic retinopathy of right eye | 12085181000006113 | 769185003 | 3689284011 |
| Early proliferative diabetic retinopathy | 6022691000006119 | 312906006 | 1785158019 |
| Right preproliferative diabetic retinopathy | 857981000006113 | 857981000006109 | 857981000006113 |
| Non proliferative diabetic retinopathy | 1484867016 | 390834004 | 1484867016 |
| Diabetic retinopathy 12 month review | 2159948010 | 408384004 | 2159948010 |
| O/E - left eye preproliferative diabetic retinopathy | 2159976019 | 408412005 | 2159976019 |
| Mild non-proliferative diabetic retinopathy of right eye | 12085091000006110 | 769183005 | 3689273016 |
| Mild non-proliferative diabetic retinopathy | 6022581000006116 | 312903003 | 2642672017 |
| PDR - proliferative diabetic retinopathy | 12189891000006110 | 59276001 | 3699415016 |
| On examination - right eye proliferative diabetic retinopathy | 6761861000006114 | 408413000 | 2621612017 |
| Preproliferative diabetic retinopathy of right eye | 12085021000006113 | 769181007 | 3689265013 |
| Severe nonproliferative diabetic retinopathy | 841011000006112 | 312905005 | 1775725011 |
| Moderate non-proliferative diabetic retinopathy | 12224731000006112 | 312904009 | 3688869017 |
| Diabetic retinopathy | 9093013 | 4855003 | 9093013 |

#### Table S54. Retinal vascular occlusion Aurum codes

| Term | Medcode ID | SNOMED CT Concept ID | SNOMED CT Description ID |
|---|---|---|---|
| BRVO - Branch retinal vein occlusion | 2892671000006112 | 24596005 | 481962015 |
| Branch retinal vein occlusion | 481961010 | 24596005 | 481961010 |
| Central retinal vein occlusion with macular oedema | 5031001000006113 | 232039004 | 347681015 |
| Central retinal artery occlusion | 64469014 | 38742007 | 64469014 |
| Central retinal vein occlusion | 113737013 | 68478007 | 113737013 |
| BRAO - Branch retinal artery occlusion | 3323131000006114 | 50821009 | 1230907012 |
| Branch retinal vein occlusion with macular oedema | 5031181000006115 | 232048009 | 347698015 |
| Hemicentral retinal vein occlusion | 5031091000006118 | 232043000 | 2619565019 |
| Central retinal vein occlusion - juvenile with macular oedema | 5031061000006114 | 232042005 | 347686013 |
| Cilioretinal artery occlusion | 5030961000006116 | 232036006 | 347676012 |
| Hemispheric retinal vein occlusion with macular oedema | 5031131000006116 | 232045007 | 347692019 |
| Branch retinal artery occlusion | 1230906015 | 50821009 | 1230906015 |
| Central retinal vein occlusion - ischaemic | 6024101000006114 | 312997008 | 456819013 |
| Central retinal vein occlusion - non-ischaemic | 6024121000006116 | 312998003 | 456822010 |
| Central retinal vein occlusion with neovascularisation | 5030981000006114 | 232038007 | 347678013 |
| Macular branch retinal vein occlusion | 5239581000006115 | 247121001 | 369049012 |
| Branch retinal vein occlusion with no neovascularisation | 6041221000006114 | 314000002 | 458269018 |
| Central retinal vein occlusion - ischemic | 6024111000006112 | 312997008 | 456820019 |
| Retinal artery occlusion | 299476010 | 232035005 | 347674010 |
| Retinal vein occlusion | 3244281000006112 | 46085004 | 494432015 |
| Hemispheric retinal vein occlusion | 5031081000006116 | 232043000 | 347688014 |
| CRAO - Central retinal artery occlusion | 3123581000006112 | 38742007 | 1229456016 |
| CRVO - Central retinal vein occlusion | 3614321000006112 | 68478007 | 501027019 |

#### Table S55. Glaucoma Aurum codes

| Term | Medcode ID | SNOMED CT Concept ID | SNOMED CT Description ID |
|---|---|---|---|
| COAG - Chronic open-angle glaucoma | 3753261000006114 | 77075001 | 503455016 |
| Secondary angle-closure glaucoma with pupil block | 4772381000006115 | 193561006 | 298063016 |
| Primary angle-closure glaucoma residual stage | 298048019 | 193546005 | 298048019 |
| Steroid-induced glaucoma glaucomatous stage | 298050010 | 193548006 | 298050010 |
| Glaucoma due to ocular cyst | 411465018 | 275477002 | 411465018 |
| Chronic angle-closure glaucoma | 3041311000006118 | 33647009 | 485587011 |
| Secondary glaucoma | 298053012 | 95717004 | 158563019 |
| Family history: Glaucoma | 4530051000006112 | 160347007 | 2666122010 |
| Glaucoma associated with ocular trauma | 3610251000006115 | 68241007 | 113353013 |
| Borderline glaucoma NOS | 298033013 | 193531003 | 298024011 |
| Open-angle glaucoma NOS | 298042018 | 84494001 | 140102012 |
| Unspecified primary angle-closure glaucoma | 298047012 | 392288006 | 1486268015 |
| Primary angle-closure glaucoma NOS | 298049010 | 392288006 | 1486268015 |
| Glaucoma associated with other ocular disorders | 298061019 | 95717004 | 158563019 |
| Other specified forms of glaucoma | 298072012 | 23986001 | 40268016 |
| Other specified glaucoma NOS | 298074013 | 23986001 | 40268016 |
| [X]Other glaucoma | 299482013 | 23986001 | 40268016 |
| Congenital glaucoma | 582671000006113 | 204113001 | 312991013 |
| Glaucomatous atrophy of optic disc | 2517641000006119 | 1207009 | 3144014 |
| Simple chronic glaucoma | 141721000006112 | 77075001 | 503452018 |
| Corticosteroid-induced glaucoma | 2524871000006112 | 1654001 | 3874015 |
| Adverse reaction to Drugs Used For Glaucoma | 1010721000006116 | 1010721000006100 | 1010721000006116 |
| Secondary open-angle glaucoma with pigment dispersion | 3245761000006114 | 46168003 | 494450010 |
| Phacomorphic glaucoma | 6532921000006114 | 392300000 | 1486280011 |
| Neovascular glaucoma | 347755010 | 232086000 | 347755010 |
| Primary angle-closure glaucoma | 1486268015 | 392288006 | 1486268015 |
| Phacolytic glaucoma | 54899010 | 32893002 | 54899010 |
| Pigmentary glaucoma | 76978010 | 46168003 | 76978010 |
| Low tension glaucoma | 84105018 | 50485007 | 84105018 |
| Glaucoma due to ocular inflammation | 803011000006118 | 37155002 | 486637019 |
| Intermittent angle-closure glaucoma | 3564451000006117 | 65460003 | 108757016 |
| Needling of bleb following glaucoma surgery | 383421000000110 | 236891000000101 | 383421000000110 |
| Chronic angle closure glaucoma | 6050201000006111 | 314784002 | 1490667012 |
| [X]Glaucoma in endocrine,nutritional+metabolic diseases CE | 388101000006114 | 193556009 | 298058015 |
| Family history of glaucoma | 4530041000006110 | 160347007 | 249916014 |
| Phacogenic glaucoma | 3872051000006113 | 84333006 | 1490002015 |
| Chronic narrow angle glaucoma | 3041351000006117 | 33647009 | 485589014 |
| Revision of bleb NEC following glaucoma surgery | 378061000000112 | 236891000000101 | 383421000000110 |
| Other specified operations following glaucoma surgery | 378721000000118 | 792741000000100 | 1772301000000112 |

| | | | |
|---|---|---|---|
| [X]Glaucoma | 388091000006115 | 23986001 | 40268016 |
| Glaucoma due to ocular tumour or cyst | 399447011 | 267625001 | 399447011 |
| Pseudoexfoliation glaucoma | 178757012 | 111514006 | 178757012 |
| Narrow-angle glaucoma | 6532591000006118 | 392291006 | 2647495010 |
| Glaucoma screening | 803131000006113 | 171215009 | 265311011 |
| Steroid-induced glaucoma NOS | 298052019 | 1654001 | 3874015 |
| Glaucoma associated with other ocular disorders NOS | 298071017 | 95717004 | 158563019 |
| Glaucoma drainage surgery | 6353481000006112 | 371370007 | 2476699016 |
| Glaucoma due to combination of mechanisms | 4010061000006117 | 92829008 | 153471016 |
| Open angle glaucoma | 3874601000006113 | 84494001 | 2579462017 |
| Bilateral angle-closure glaucoma | 9846871000006112 | 15736721000119106 | 3373331019 |
| Injection of filtering bleb following glaucoma surgery | 7089401000006110 | 428494000 | 2694522012 |
| Glaucoma due to another disorder | 299484014 | 95717004 | 158563019 |
| Aphakic glaucoma | 2743971000006118 | 15374009 | 26072012 |
| Subcapsular glaucomatous flecks | 3601481000006119 | 67733005 | 112484015 |
| Subacute closed-angle glaucoma | 3564481000006113 | 65460003 | 1232678013 |
| Acute angle-closure glaucoma of right eye | 8107961000006110 | 336611000119109 | 3316703019 |
| Glaucoma associated with disorders of the lens | 802911000006110 | 84333006 | 1234928013 |
| Wide-angle glaucoma | 3874591000006117 | 84494001 | 140106010 |
| Glaucoma due to other anterior segment anomaly | 298056016 | 95717004 | 158563019 |
| Glaucoma NOS | 298075014 | 23986001 | 40268016 |
| Secondary open-angle glaucoma | 2848811000006113 | 21928008 | 36783018 |
| Glaucoma due to episode of increased venous pressure | 802981000006115 | 34623005 | 1228189019 |
| Glaucoma due to pupillary block | 803061000006115 | 193561006 | 298063016 |
| Steroid-induced glaucoma residual stage | 298051014 | 193549003 | 298051014 |
| Glaucoma with lens disorder | 3872071000006115 | 84333006 | 1234928013 |
| Uveitic glaucoma | 6041601000006118 | 314033007 | 458309018 |
| Glaucoma surgery | 3899381000006119 | 86077009 | 2621280010 |
| Glaucoma suspect | 5031661000006115 | 232079008 | 347743016 |
| Steroid-induced glaucoma | 1221368012 | 1654001 | 1221368012 |
| Glaucomatous subcapsular flecks | 1232965011 | 67733005 | 1232965011 |
| Suspected angle-closure glaucoma | 7629711000006119 | 708176002 | 3042911012 |
| Glaucoma following surgery | 5031801000006115 | 232090003 | 347762018 |
| Mixed mechanism glaucoma | 4010091000006113 | 92829008 | 2770265014 |
| Suspected glaucoma of bilateral eyes caused by steroids | 9351591000006115 | 11001071000119105 | 3326549013 |
| Chronic closed-angle glaucoma | 556201000006114 | 33647009 | 485588018 |
| Cupping of optic discs of bilateral eyes co-occurrent and due to open-angle glaucoma | 3526208016 | 737006004 | 3526208016 |
| Bilateral absolute glaucoma | 8111981000006112 | 347401000119106 | 3316706010 |
| No family history of glaucoma | 4528321000006118 | 160267000 | 249716018 |
| Borderline glaucoma steroid responder | 1490617017 | 302895007 | 1490617017 |

| | | | |
|---|---|---|---|
| ACG - Angle closure glaucoma | 6532581000006116 | 392291006 | 2621430019 |
| Residual stage of open angle glaucoma | 47081000006113 | 66990007 | 111276019 |
| Secondary/other glaucoma | 883301000006118 | 660261000000103 | 883301000006118 |
| Angle-closure glaucoma of both eyes | 9846911000006110 | 15736721000119106 | 3373334010 |
| Angle-closure glaucoma | 6532571000006119 | 392291006 | 1486271011 |
| Glaucoma right eye | 856591000006115 | 856591000006104 | 856591000006115 |
| Glaucoma left eye | 856601000006111 | 856601000006107 | 856601000006111 |
| Borderline glaucoma | 298024011 | 193531003 | 298024011 |
| Glaucoma due to chamber angle anomaly | 298054018 | 193552006 | 298054018 |
| Glaucoma due to iris anomaly | 298055017 | 193553001 | 298055017 |
| Operation following glaucoma surgery | 361511000000118 | 792741000000100 | 1772301000000112 |
| Glaucoma caused by silicone oil | 7727931000006113 | 715144004 | 3301594012 |
| Primary open angle glaucoma | 3753211000006111 | 77075001 | 127956010 |
| Secondary open-angle glaucoma with pseudoexfoliation | 4196741000006113 | 111514006 | 1219666018 |
| Secondary angle-closure glaucoma | 2843081000006117 | 21571006 | 36185019 |
| Closed angle glaucoma | 560611000006117 | 392288006 | 1486268015 |
| Newborn glaucoma | 674741000006115 | 204113001 | 312991013 |
| Narrow angle glaucoma | 6532601000006114 | 392291006 | 2647496011 |
| Glaucoma tube shunt | 5484711000006110 | 265291005 | 2551168012 |
| Absolute glaucoma | 802901000006112 | 19144002 | 32262013 |
| Glaucoma due to disease NOS | 298059011 | 95717004 | 158563019 |
| Operations following glaucoma surgery NOS | 383401000000118 | 792741000000100 | 1772301000000112 |
| Removal of releasable suture following glaucoma surgery | 2676361011 | 426877004 | 2676361011 |
| Glaucoma associated with disorders of the lens NOS | 298060018 | 84333006 | 139867011 |
| Needling of bleb following glaucoma surgery | 7089411000006113 | 428494000 | 3032787012 |
| Acute-on-chronic glaucoma | 6041451000006116 | 314017009 | 458290014 |
| Angle-closure glaucoma - borderline | 521031000006117 | 193534006 | 298027016 |
| Glaucoma monitoring | 264949013 | 170946006 | 264949013 |
| Unspecified open-angle glaucoma | 298034019 | 84494001 | 140102012 |
| Glaucoma due to unspecified ocular disorder | 298062014 | 95717004 | 158563019 |
| Glaucomatous optic atrophy | 2517661000006115 | 1207009 | 1220515013 |
| Primary closed-angle glaucoma | 883291000006119 | 392288006 | 883291000006119 |
| At risk of glaucoma | 1697781000006112 | 314018004 | 458291013 |
| Injection of bleb following glaucoma surgery | 383381000000118 | 428494000 | 1659491000000113 |
| Traumatic glaucoma | 3610261000006118 | 68241007 | 1233025018 |
| Chronic simple glaucoma | 3753221000006115 | 77075001 | 127957018 |
| Glaucomatocyclitic crisis | 2974881000006116 | 29538005 | 49421018 |
| Glaucoma due to systemic syndrome | 298057013 | 193555008 | 298057013 |
| Glaucoma in endocrine, nutritional and metabolic diseases | 298058015 | 193556009 | 298058015 |
| Open-angle glaucoma | 140102012 | 84494001 | 140102012 |
| Narrow angle glaucoma of bilateral eyes | 9846901000006112 | 15736721000119106 | 3373333016 |

| | | | |
|---|---|---|---|
| Glaucoma surgery | 6979851000006114 | 421885009 | 2618478010 |
| Glaucoma | 40268016 | 23986001 | 40268016 |
| Hypersecretion glaucoma | 49139016 | 29369005 | 49139016 |
| Primary congenital glaucoma | 6871531000006110 | 415176004 | 2535441016 |
| Laser suture lysis following glaucoma surgery | 378701000000110 | 234611000000100 | 378701000000110 |
| Suspected glaucoma | 558691000000111 | 232079008 | 2408401000000115 |
| Advanced open-angle glaucoma | 4196691000006111 | 111513000 | 178756015 |
| Panretinal photocoagulation for glaucoma | 249331000006112 | 172586003 | 267176015 |
| CSG - Chronic simple glaucoma | 3753251000006112 | 77075001 | 503454017 |
| POAG - Primary open-angle glaucoma | 3753241000006110 | 77075001 | 503453011 |
| Low tension glaucoma | 734071000006115 | 50485007 | 84105018 |
| Suspected glaucoma | 1629641000006111 | 1629641000006107 | 1629641000006111 |
| Glaucoma implant surgery | 5484721000006119 | 265291005 | 2883614019 |
| CNAG - Chronic narrow angle glaucoma | 3041361000006115 | 33647009 | 2620650018 |
| Referral to glaucoma clinic | 2533408017 | 415273001 | 2533408017 |
| [V]Screening for glaucoma | 461299019 | 171215009 | 265311011 |
| Bilateral narrow angle glaucoma | 9846881000006110 | 15736721000119106 | 3373330018 |
| Unspecified preglaucoma | 298025012 | 23986001 | 40268016 |
| Primary open-angle glaucoma | 503452018 | 77075001 | 503452018 |
| Optic disc glaucomatous atrophy | 1220514012 | 1207009 | 1220514012 |
| Glaucoma due to ocular trauma | 1233026017 | 68241007 | 1233026017 |
| Open-angle glaucoma - borderline | 264071000006115 | 193533000 | 298026013 |
| Acute closed-angle glaucoma | 459721000006110 | 30041005 | 1226810019 |
| No evidence of glaucoma | 12624091000006111 | 1109141000000102 | 2775491000000119 |
| Acute angle-closure glaucoma | 2983571000006113 | 30041005 | 50268012 |
| Normal pressure glaucoma | 495680018 | 50485007 | 495680018 |
| AACG - Acute angle closure glaucoma | 2983591000006114 | 30041005 | 2620418016 |
| Glaucoma micro-stent | 3532561018 | 738510009 | 3532561018 |
| Glaucomatocyclitic crises | 1226368013 | 29538005 | 1226368013 |
| Glaucoma with ocular inflammation | 3098461000006112 | 37155002 | 486637019 |
| Angle recession glaucoma | 6535011000006114 | 392352004 | 1486331015 |
| [RFC] Glaucoma | 907841000006110 | 907841000006106 | 907841000006110 |
| Intermittent primary angle-closure glaucoma | 770481000006118 | 65460003 | 1232679017 |
| Cupping of optic disc co-occurrent and due to open angle glaucoma | 3526206017 | 736838001 | 3526206017 |
| Glaucoma due to ocular vascular disorder | 298064010 | 193562004 | 298064010 |
| H/O: glaucoma | 251653013 | 161488001 | 251653013 |
| Normal tension glaucoma | 3317631000006118 | 50485007 | 495682014 |
| Open angle glaucoma suspect | 14154791000006118 | | |
| Angle closure glaucoma suspect | 14153851000006115 | | |
| Glaucoma suspect | 13933811000006119 | | |

#### Table S56. Age-related macular degeneration Aurum codes

| Term | Medcode ID | SNOMED CT Concept ID | SNOMED CT Description ID |
|---|---|---|---|
| Myopic macular degeneration | 6022521000006115 | 312898002 | 456698016 |
| Drusen stage macular degeneration | 5478331000006117 | 264633009 | 393115018 |
| Disciform macular degeneration | 6853451000006118 | 414173003 | 2538082010 |
| Atrophic age-related macular degeneration | 6866121000006116 | 414875008 | 2537363011 |
| Senile macular degeneration | 399676018 | 267718000 | 399676018 |
| EMD - Exudative macular degeneration | 6853541000006119 | 414173003 | 2621806012 |
| Age related macular degeneration | 78571000006118 | 267718000 | 399670012 |
| AAMD - Age related macular degeneration | 5501831000006117 | 267718000 | 2620215016 |
| Neovascular age-related macular degeneration | 6853501000006116 | 414173003 | 2538087016 |
| ARMD - Age-related macular degeneration | 5501781000006114 | 267718000 | 399672016 |
| Dry senile macular degeneration | 2537365016 | 414875008 | 2537365016 |
| Wet senile macular degeneration | 2538088014 | 414173003 | 2538088014 |
| AMD - Age-related macular degeneration | 5501811000006111 | 267718000 | 399675019 |
| Nonexudative age-related macular degeneration | 6866111000006112 | 414875008 | 2534426012 |
| Exudative age-related macular degeneration | 6853441000006115 | 414173003 | 2535500012 |
| Age-related macular degeneration | 5501771000006111 | 267718000 | 399671011 |
| Disciform macular degeneration | 14198731000006115 | | |

#### Table S57. Retinal Laser Aurum codes

| Term | Medcode ID | SNOMED CT Concept ID | SNOMED CT Description ID |
|---|---|---|---|
| Panretinal laser photocoagulation to lesion of retina NEC | 361631000000110 | 283851000000107 | 489321000000113 |
| Panretinal laser photocoagulation | 6020151000006115 | 312713003 | 1785150014 |
| Retinal laser photocoagulation | 6630611000006117 | 399867007 | 1787015019 |
| Focal retinal laser photocoagulation | 6588671000006112 | 397538008 | 1785845018 |
| Panretinal laser photocoagulation to lesion of retina | 489321000000113 | 283851000000107 | 489321000000113 |
| O/E - No retinal laser photocoagulation scars | 2534106015 | 414902006 | 2534106015 |
| Scatter retinal laser photocoagulation | 6020161000006118 | 312713003 | 2536171014 |
| Retinal laser therapy | 262909018 | 169425004 | 262911010 |

#### Table S58. Vitrectomy Aurum codes

| Term | Medcode ID | SNOMED CT Concept ID | SNOMED CT Description ID |
|---|---|---|---|
| Mechanical vitrectomy by pars plana approach | 3914871000006116 | 87021001 | 144315019 |
| Pars plana vitrectomy | 507373018 | 87021001 | 507373018 |
| Vitrectomy | 3731361000006116 | 75732000 | 125783018 |
| Anterior vitrectomy | 267129012 | 172555009 | 267129012 |
| Vitrectomy using pars plana approach | 465551000000114 | 87021001 | 465551000000114 |
| PPV - Pars plana vitrectomy | 3914891000006115 | 87021001 | 507374012 |
| Anterior vitrectomy | 486601000006113 | 172555009 | 267129012 |
| Vitrectomy by anterior approach | 1541981000006119 | 172555009 | 267130019 |
| Phacoemulsification/vitrectomy system | 7406901000006117 | 462940007 | 2944713011 |

#### Table S59. Scleral buckle Aurum codes

| Term | Medcode ID | SNOMED CT Concept ID | SNOMED CT Description ID |
|---|---|---|---|
| Scleral buckle | 6346771000006113 | 370944007 | 2673018011 |
| Removal of scleral buckle | 6353851000006118 | 371394007 | 1210056018 |
| Extrusion of scleral buckle | 6516171000006117 | 390909000 | 1484941010 |
| Scleral buckle procedure | 2929071000006111 | 26786002 | 2162369019 |

#### Table S60. Intravitreal injection Aurum codes

| Term | Medcode ID | SNOMED CT Concept ID | SNOMED CT Description ID |
|---|---|---|---|
| Ranibizumab 10mg/mL solution for intravitreal injection 0.05mL vial | 7021931000006119 | 424204000 | 2645710015 |
| Intravitreal injection | 8255441000006119 | 800481000000101 | 1789871000000114 |

#### Table S61. Antiplatelet Aurum product codes

| Term from EMIS | Prod code ID | dmd ID | Drug substance name |
|---|---|---|---|
| Angettes 75 tablets (Bristol-Myers Squibb Pharmaceuticals Ltd) | 71241000033119 | 800511000001106 | Aspirin |
| Aspirin 300mg dispersible tablets | 82841000033117 | 39695211000001102 | Aspirin |
| Aspirin 75mg dispersible tablets | 82941000033113 | 319773006 | Aspirin |
| Aspirin 75mg gastro-resistant tablets | 83041000033115 | 319781007 | Aspirin |
| Aspirin 300mg gastro-resistant tablets | 83141000033116 | 322225002 | Aspirin |
| Aspirin 300mg modified-release tablets | 84441000033114 | 4558011000001103 | Aspirin |
| Aspirin 300mg tablets | 86641000033119 | 329525004 | Aspirin |
| Aspav dispersible tablets (Actavis UK Ltd) | 86841000033118 | 3690911000001103 | Aspirin/ Papaveretum |
| Aspirin 75mg tablets | 87041000033110 | 319775004 | Aspirin |
| Caprin 75mg gastro-resistant tablets (Wockhardt UK Ltd) | 189841000033119 | 927411000001108 | Aspirin |
| Caprin 300mg gastro-resistant tablets (Pinewood Healthcare) | 212341000033117 | 901211000001107 | Aspirin |
| Co-codaprin 8mg/400mg tablets | 370941000033119 | 11762111000001100 | Aspirin/ Codeine phosphate |
| Co-codaprin 8mg/400mg dispersible tablets | 371041000033112 | 39695111000001108 | Aspirin/ Codeine phosphate |
| Disprin CV 300mg modified-release tablets (Reckitt Benckiser Healthcare (UK) Ltd) | 452541000033115 | 4532311000001104 | Aspirin |
| Nu-Seals 75 gastro-resistant tablets (Alliance Pharmaceuticals Ltd) | 991441000033115 | 414311000001102 | Aspirin |
| Nu-Seals 300 gastro-resistant tablets (Alliance Pharmaceuticals Ltd) | 993341000033113 | 183311000001101 | Aspirin |
| PostMI 75 dispersible tablets (Ashbourne Pharmaceuticals Ltd) | 1098041000033111 | 636011000001100 | Aspirin |
| PostMI 75 EC tablets (Ashbourne Pharmaceuticals Ltd) | 1101541000033119 | 38411000001101 | Aspirin |
| Asasantin Retard capsules (Boehringer Ingelheim Ltd) | 1571541000033117 | 3292511000001108 | Aspirin/ Dipyridamole |
| Enprin 75mg gastro-resistant tablets (Galpharm International Ltd) | 1729141000033110 | 473711000001109 | Aspirin |
| Imazin XL tablets (Napp Pharmaceuticals Ltd) | 1739541000033116 | 3460911000001104 | Aspirin/ Isosorbide mononitrate |
| Imazin XL forte tablets (Napp Pharmaceuticals Ltd) | 1739641000033115 | 3461511000001104 | Aspirin/ Isosorbide mononitrate |
| Aspirin 300mg suppositories | 2013341000033115 | 322232006 | Aspirin |
| Aspirin 150mg suppositories | 2013441000033114 | 322233001 | Aspirin |
| Aspirin powder | 2085241000033111 | 5145711000001107 | Aspirin |
| MigraMax oral powder sachets (Zentiva Pharma UK Ltd) | 2086841000033117 | 3637911000001108 | Aspirin DL-Lysine/ Metoclopramide hydrochloride |
| Micropirin 75mg gastro-resistant tablets (Dexcel-Pharma Ltd) | 2273741000033118 | 281311000001104 | Aspirin |
| Dipyridamole 200mg modified-release / Aspirin 25mg capsules | 3160741000033110 | 36069911000001109 | Aspirin/ Dipyridamole |
| Aspirin 75mg / Isosorbide mononitrate 60mg modified-release tablets | 3162041000033113 | 35903111000001101 | Aspirin/ Isosorbide mononitrate |

| | | | |
|---|---|---|---|
| Aspirin 150mg / Isosorbide mononitrate 60mg modified-release tablets | 3162141000033112 | 35903011000001102 | Aspirin/ Isosorbide mononitrate |
| Aspirin 900mg / Metoclopramide 10mg oral powder sachets sugar free | 3836841000033119 | 322784004 | Aspirin DL-Lysine/ Metoclopramide hydrochloride |
| Aspirin 500mg / Papaveretum 7.71mg dispersible tablets sugar free | 3849841000033110 | 39695311000001105 | Aspirin/ Papaveretum |
| Aspirin 500mg / Codeine 8mg dispersible tablets sugar free | 3934341000033112 | 4956111000001107 | Aspirin/ Codeine phosphate |
| Codis 500 dispersible tablets (Reckitt Benckiser Healthcare (UK) Ltd) | 3934441000033118 | 4951811000001105 | Aspirin/ Codeine phosphate |
| Aspro Clear 300mg effervescent tablets (Bayer Plc) | 4431841000033118 | 4649111000001101 | Aspirin |
| Maximum Strength Aspro Clear 500mg effervescent tablets (Bayer Plc) | 4431941000033114 | 4648711000001109 | Aspirin |
| Molita 200mg/25mg modified-release capsules (Dr Reddy's Laboratories (UK) Ltd) | 8619141000033116 | 23241511000001106 | Aspirin/ Dipyridamole |
| Bisoprolol 5mg / Aspirin 75mg capsules | 10024941000033116 | 28365711000001103 | Aspirin/ Bisoprolol fumarate |
| Bisoprolol 5mg / Aspirin 100mg capsules | 10025041000033116 | 28365611000001107 | Aspirin/ Bisoprolol fumarate |
| Bisoprolol 10mg / Aspirin 100mg capsules | 10025241000033112 | 28365311000001102 | Aspirin/ Bisoprolol fumarate |
| Danamep 75mg dispersible tablets (Ecogen Europe Ltd) | 11716941000033112 | 32968711000001107 | Aspirin |
| Clopidogrel 75mg tablets | 1583541000033113 | 39689111000001106 | Clopidogrel |
| Plavix 75mg tablets (Sanofi) | 1667041000033119 | 454611000001104 | Clopidogrel |
| Clopidogrel 300mg tablets | 4519641000033119 | 429540001 | Clopidogrel hydrogen sulfate |
| Plavix 300mg tablets (Sanofi) | 4519741000033111 | 13663511000001101 | Clopidogrel hydrogen sulfate |
| Grepid 75mg tablets (Kent Pharma (UK) Ltd) | 5242341000033112 | 15907411000001101 | Clopidogrel |
| Clopidogrel 75mg/5ml oral suspension | 6122841000033118 | 8426611000001103 | Clopidogrel hydrogen sulfate |
| Clopidogrel 1mg/ml oral suspension | 8031441000033112 | 18747911000001101 | Clopidogrel hydrogen sulfate |
| Clopidogrel 4mg oral powder sachets | 8300741000033118 | 22342711000001109 | Clopidogrel hydrogen sulfate |
| Clopidogrel 75mg/5ml oral solution | 8301241000033117 | 22402511000001104 | Clopidogrel hydrogen sulfate |
| Clopidogrel 25mg/5ml oral suspension | 13283841000033111 | 16072911000001108 | Clopidogrel hydrogen sulfate |
| Ticagrelor 90mg tablets | 6059341000033111 | 704465002 | Ticagrelor |
| Brilique 90mg tablets (AstraZeneca UK Ltd) | 6059441000033117 | 18290311000001104 | Ticagrelor |
| Ticagrelor 60mg tablets | 11422441000033119 | 32472211000001106 | Ticagrelor |
| Brilique 60mg tablets (AstraZeneca UK Ltd) | 11422741000033114 | 32447511000001106 | Ticagrelor |
| Ticagrelor 90mg orodispersible tablets sugar free | 12371041000033112 | 34713411000001108 | Ticagrelor |
| Brilique 90mg orodispersible tablets (AstraZeneca UK Ltd) | 12371141000033111 | 34672011000001105 | Ticagrelor |
| Dipyridamole 10mg/2ml solution for injection ampoules | 447141000033112 | 36069811000001104 | Dipyridamole |
| Dipyridamole 200mg modified-release capsules | 454141000033119 | 39020811000001108 | Dipyridamole |
| Dipyridamole 100mg tablets | 469641000033119 | 319759002 | Dipyridamole |

| | | | |
|---|---|---|---|
| Dipyridamole 25mg tablets | 469741000033111 | 319758005 | Dipyridamole |
| Persantin 10mg/2ml solution for injection ampoules (Boehringer Ingelheim Ltd) | 1052141000033119 | 642111000001104 | Dipyridamole |
| Persantin Retard 200mg capsules (Boehringer Ingelheim Ltd) | 1054941000033114 | 452911000001101 | Dipyridamole |
| Persantin 100mg tablets (Boehringer Ingelheim Ltd) | 1065441000033113 | 623011000001106 | Dipyridamole |
| Persantin 25mg tablets (Boehringer Ingelheim Ltd) | 1065541000033114 | 635611000001102 | Dipyridamole |
| Dipyridamole 50mg/5ml oral suspension sugar free | 1728341000033118 | 36070011000001108 | Dipyridamole |
| Dipyridamole 200mg/5ml oral suspension | 5020341000033117 | 8456311000001107 | Dipyridamole |
| Dipyridamole 100mg/5ml oral suspension | 5052341000033115 | 8456511000001101 | Dipyridamole |
| Dipyridamole 250mg/5ml oral solution | 5401541000033119 | 8456211000001104 | Dipyridamole |
| Dipyridamole 100mg/5ml oral solution | 5889941000033115 | 8456611000001102 | Dipyridamole |
| Dipyridamole 200mg/5ml oral solution | 5992041000033118 | 8456411000001100 | Dipyridamole |
| Attia 200mg modified-release capsules (Dr Reddy's Laboratories (UK) Ltd) | 9165341000033117 | 24435511000001109 | Dipyridamole |
| Ofcram PR 200mg capsules (Advanz Pharma) | 10333041000033118 | 26779011000001104 | Dipyridamole |
| Dipyridamole 200mg/5ml oral suspension sugar free | 11072641000033113 | 31948811000001109 | Dipyridamole |
| Trolactin 200mg modified-release capsules (Actavis UK Ltd) | 12195441000033115 | 34175911000001100 | Dipyridamole |
| Prasugrel 5mg tablets | 4956441000033110 | 39702911000001108 | Prasugrel |
| Prasugrel 10mg tablets | 4956541000033111 | 39702811000001103 | Prasugrel |
| Efient 5mg tablets (Daiichi Sankyo UK Ltd) | 4956641000033112 | 15241111000001105 | Prasugrel |
| Efient 10mg tablets (Daiichi Sankyo UK Ltd) | 4956741000033115 | 15240211000001105 | Prasugrel |

#### Table S62. Thiazide diuretics Aurum product codes

| Term from EMIS | Prod code ID | dmd ID | Drug substance name |
|---|---|---|---|
| Accuretic 10mg/12.5mg tablets (Pfizer Ltd) | 11241000033113 | 260211000001104 | Hydrochlorothiazide/ Quinapril hydrochloride |
| Acezide 25mg/50mg tablets (Bristol-Myers Squibb Pharmaceuticals Ltd) | 13141000033119 | 263311000001101 | Captopril/ Hydrochlorothiazide |
| Amilmaxco 5mg/50mg tablets (Ashbourne Pharmaceuticals Ltd) | 57341000033115 | 712211000001108 | Amiloride hydrochloride/ Hydrochlorothiazide |
| Amil-Co 5mg/50mg tablets (IVAX Pharmaceuticals UK Ltd) | 60341000033110 | 636611000001107 | Amiloride hydrochloride/ Hydrochlorothiazide |
| Capozide LS 12.5mg/25mg tablets (Bristol-Myers Squibb Pharmaceuticals Ltd) | 209241000033119 | 546711000001107 | Captopril/ Hydrochlorothiazide |
| Carace 10 Plus tablets (Organon Pharma (UK) Ltd) | 209841000033115 | 3144511000001107 | Hydrochlorothiazide/ Lisinopril |
| Capozide 25mg/50mg tablets (Bristol-Myers Squibb Pharmaceuticals Ltd) | 216441000033110 | 17311000001102 | Captopril/ Hydrochlorothiazide |
| Carace 20 Plus tablets (Organon Pharma (UK) Ltd) | 217041000033116 | 3143511000001109 | Hydrochlorothiazide/ Lisinopril |
| Co-amilozide 5mg/50mg/5ml oral solution | 351941000033111 | 8792311000001109 | Amiloride hydrochloride/ Hydrochlorothiazide |
| Co-amilozide 5mg/50mg tablets | 369141000033118 | 377566005 | Amiloride hydrochloride/ Hydrochlorothiazide |
| Co-triamterzide 50mg/25mg tablets | 370241000033111 | 410896007 | Hydrochlorothiazide/ Triamterene |
| Cozaar-Comp 50mg/12.5mg tablets (Organon Pharma (UK) Ltd) | 370841000033110 | 255911000001105 | Hydrochlorothiazide/ Losartan potassium |
| Co-Betaloc tablets (Pfizer Ltd) | 372741000033116 | 2977611000001106 | Hydrochlorothiazide/ Metoprolol tartrate |
| Co-Betaloc SA tablets (Pfizer Ltd) | 372841000033114 | 3853411000001104 | Hydrochlorothiazide/ Metoprolol tartrate |
| Co-amilozide 2.5mg/25mg tablets | 376941000033117 | 318121006 | Amiloride hydrochloride/ Hydrochlorothiazide |
| Dyazide 50mg/25mg tablets (Advanz Pharma) | 492341000033115 | 132811000001106 | Hydrochlorothiazide/ Triamterene |
| Hydrosaluric 25mg tablets (Organon Pharma (UK) Ltd) | 742541000033112 | 4544011000001109 | Hydrochlorothiazide |
| Hydrosaluric 50mg tablets (Organon Pharma (UK) Ltd) | 742641000033113 | 4546211000001109 | Hydrochlorothiazide |
| Hydrochlorothiazide 25mg tablets | 744941000033110 | 376209006 | Hydrochlorothiazide |
| Hydrochlorothiazide 50mg tablets | 745441000033118 | 376508004 | Hydrochlorothiazide |
| Innozide 20mg/12.5mg tablets (Organon Pharma (UK) Ltd) | 770141000033115 | 146811000001108 | Enalapril maleate/ Hydrochlorothiazide |
| Kalten capsules (M & A Pharmachem Ltd) | 793741000033117 | 237011000001100 | Amiloride hydrochloride/ Atenolol/ Hydrochlorothiazide |
| Moducren tablets (Organon Pharma (UK) Ltd) | 935341000033118 | 74311000001101 | Amiloride hydrochloride/ Hydrochlorothiazide/ Timolol maleate |
| Moduret 25 tablets (Organon Pharma (UK) Ltd) | 935441000033112 | 314211000001100 | Amiloride hydrochloride/ Hydrochlorothiazide |

| | | | |
|---|---|---|---|
| Moduretic 5mg/50mg tablets (Merck Sharp & Dohme (UK) Ltd) | 935541000033113 | 453811000001103 | Amiloride hydrochloride/ Hydrochlorothiazide |
| Monozide 10 tablets (Wyeth Pharmaceuticals) | 937341000033111 | 4542911000001104 | Bisoprolol fumarate/ Hydrochlorothiazide |
| Secadrex 200mg/12.5mg tablets (Sanofi) | 1274641000033111 | 878911000001106 | Acebutolol hydrochloride/ Hydrochlorothiazide |
| Triamaxco 50mg/25mg tablets (Ashbourne Pharmaceuticals Ltd) | 1468141000033118 | 721311000001104 | Hydrochlorothiazide/ Triamterene |
| Triam-Co 50mg/25mg tablets (IVAX Pharmaceuticals UK Ltd) | 1470341000033112 | 191211000001106 | Hydrochlorothiazide/ Triamterene |
| Zestoretic 10 tablets (AstraZeneca UK Ltd) | 1551841000033118 | 3144311000001101 | Hydrochlorothiazide/ Lisinopril |
| Zestoretic 20 tablets (AstraZeneca UK Ltd) | 1552341000033118 | 3143111000001100 | Hydrochlorothiazide/ Lisinopril |
| Co-zidocapt 12.5mg/25mg tablets | 1893141000033119 | 318806002 | Captopril/ Hydrochlorothiazide |
| Co-zidocapt 25mg/50mg tablets | 1893241000033114 | 318807006 | Captopril/ Hydrochlorothiazide |
| CoAprovel 150mg/12.5mg tablets (Sanofi) | 2216141000033115 | 792411000001108 | Hydrochlorothiazide/ Irbesartan |
| CoAprovel 300mg/12.5mg tablets (Sanofi) | 2216241000033110 | 682711000001109 | Hydrochlorothiazide/ Irbesartan |
| Hydrochlorothiazide 50mg/5ml oral solution | 2746441000033111 | 8529611000001101 | Hydrochlorothiazide |
| MicardisPlus 40mg/12.5mg tablets (Boehringer Ingelheim Ltd) | 2864241000033110 | 3806311000001109 | Hydrochlorothiazide/ Telmisartan |
| MicardisPlus 80mg/12.5mg tablets (Boehringer Ingelheim Ltd) | 2864341000033117 | 3806911000001105 | Hydrochlorothiazide/ Telmisartan |
| Caralpha 10mg/12.5mg tablets (Actavis UK Ltd) | 3136041000033117 | 7385911000001108 | Hydrochlorothiazide/ Lisinopril |
| Caralpha 20mg/12.5mg tablets (Actavis UK Ltd) | 3136141000033118 | 7385711000001106 | Hydrochlorothiazide/ Lisinopril |
| Co-Diovan 160mg/12.5mg tablets (Novartis Pharmaceuticals UK Ltd) | 3152541000033112 | 7668611000001104 | Hydrochlorothiazide/ Valsartan |
| Co-Diovan 160mg/25mg tablets (Novartis Pharmaceuticals UK Ltd) | 3152641000033113 | 7668911000001105 | Hydrochlorothiazide/ Valsartan |
| Quinapril 10mg / Hydrochlorothiazide 12.5mg tablets | 3160641000033118 | 39693411000001102 | Hydrochlorothiazide/ Quinapril hydrochloride |
| Lisinopril 10mg / Hydrochlorothiazide 12.5mg tablets | 3160841000033117 | 318884002 | Hydrochlorothiazide/ Lisinopril |
| Lisinopril 20mg / Hydrochlorothiazide 12.5mg tablets | 3160941000033113 | 318880006 | Hydrochlorothiazide/ Lisinopril |
| Irbesartan 150mg / Hydrochlorothiazide 12.5mg tablets | 3161141000033116 | 134461004 | Hydrochlorothiazide/ Irbesartan |
| Irbesartan 300mg / Hydrochlorothiazide 12.5mg tablets | 3161241000033111 | 134460003 | Hydrochlorothiazide/ Irbesartan |
| Metoprolol 100mg / Hydrochlorothiazide 12.5mg tablets | 3161341000033118 | 318546001 | Hydrochlorothiazide/ Metoprolol tartrate |
| Valsartan 160mg / Hydrochlorothiazide 12.5mg tablets | 3161441000033112 | 395521005 | Hydrochlorothiazide/ Valsartan |
| Valsartan 160mg / Hydrochlorothiazide 25mg tablets | 3161541000033113 | 409298002 | Hydrochlorothiazide/ Valsartan |
| Losartan 50mg / Hydrochlorothiazide 12.5mg tablets | 3161741000033117 | 318959004 | Hydrochlorothiazide/ Losartan potassium |
| Enalapril 20mg / Hydrochlorothiazide 12.5mg tablets | 3162241000033117 | 39691911000001109 | Enalapril maleate/ Hydrochlorothiazide |
| Telmisartan 40mg / Hydrochlorothiazide 12.5mg tablets | 3162641000033119 | 407855002 | Hydrochlorothiazide/ Telmisartan |
| Telmisartan 80mg / Hydrochlorothiazide 12.5mg tablets | 3162741000033111 | 407856001 | Hydrochlorothiazide/ Telmisartan |

| | | | |
|---|---|---|---|
| Acebutolol 200mg / Hydrochlorothiazide 12.5mg tablets | 3162941000033114 | 39725011000001105 | Acebutolol hydrochloride/ Hydrochlorothiazide |
| Valsartan 80mg / Hydrochlorothiazide 12.5mg tablets | 3190741000033112 | 377488008 | Hydrochlorothiazide/ Valsartan |
| Co-Diovan 80mg/12.5mg tablets (Novartis Pharmaceuticals UK Ltd) | 3190841000033119 | 8150111000001108 | Hydrochlorothiazide/ Valsartan |
| Lisicostad 10mg/12.5mg tablets (Genus Pharmaceuticals Ltd) | 3279541000033115 | 8145711000001102 | Hydrochlorothiazide/ Lisinopril |
| Lisicostad 20mg/12.5mg tablets (Genus Pharmaceuticals Ltd) | 3279641000033119 | 8145911000001100 | Hydrochlorothiazide/ Lisinopril |
| Losartan 100mg / Hydrochlorothiazide 25mg tablets | 3346841000033117 | 395497004 | Hydrochlorothiazide/ Losartan potassium |
| Cozaar-Comp 100mg/25mg tablets (Organon Pharma (UK) Ltd) | 3346941000033113 | 9566911000001105 | Hydrochlorothiazide/ Losartan potassium |
| Olmesartan medoxomil 20mg / Hydrochlorothiazide 12.5mg tablets | 3908941000033112 | 409184002 | Hydrochlorothiazide/ Olmesartan medoxomil |
| Olmesartan medoxomil 20mg / Hydrochlorothiazide 25mg tablets | 3909041000033115 | 10270711000001105 | Hydrochlorothiazide/ Olmesartan medoxomil |
| Olmetec Plus 20mg/12.5mg tablets (Daiichi Sankyo UK Ltd) | 3909141000033116 | 10261511000001103 | Hydrochlorothiazide/ Olmesartan medoxomil |
| Olmetec Plus 20mg/25mg tablets (Daiichi Sankyo UK Ltd) | 3909241000033111 | 10261811000001100 | Hydrochlorothiazide/ Olmesartan medoxomil |
| Irbesartan 300mg / Hydrochlorothiazide 25mg tablets | 3995241000033112 | 10970311000001105 | Hydrochlorothiazide/ Irbesartan |
| CoAprovel 300mg/25mg tablets (Sanofi) | 3995341000033119 | 10968611000001106 | Hydrochlorothiazide/ Irbesartan |
| Losartan 100mg / Hydrochlorothiazide 12.5mg tablets | 4424741000033110 | 13112711000001103 | Hydrochlorothiazide/ Losartan potassium |
| Cozaar-Comp 100mg/12.5mg tablets (Organon Pharma (UK) Ltd) | 4424841000033117 | 13094111000001102 | Hydrochlorothiazide/ Losartan potassium |
| Telmisartan 80mg / Hydrochlorothiazide 25mg tablets | 4548741000033111 | 13731911000001109 | Hydrochlorothiazide/ Telmisartan |
| MicardisPlus 80mg/25mg tablets (Boehringer Ingelheim Ltd) | 4548841000033118 | 13719711000001103 | Hydrochlorothiazide/ Telmisartan |
| Timolol 10mg / Amiloride 2.5mg / Hydrochlorothiazide 25mg tablets (Essential Generics Ltd) | 5376441000033114 | 16458411000001107 | Amiloride hydrochloride/ Hydrochlorothiazide/ Timolol maleate |
| Olmesartan medoxomil 40mg / Hydrochlorothiazide 12.5mg tablets | 5566341000033111 | 409185001 | Hydrochlorothiazide/ Olmesartan medoxomil |
| Olmetec Plus 40mg/12.5mg tablets (Daiichi Sankyo UK Ltd) | 5566441000033117 | 17220911000001102 | Hydrochlorothiazide/ Olmesartan medoxomil |
| Hydrochlorothiazide 5mg/5ml oral solution | 5969841000033119 | 12538711000001107 | Hydrochlorothiazide |
| Hydrochlorothiazide 5mg/5ml oral suspension | 5969941000033110 | 12538811000001104 | Hydrochlorothiazide |
| Sevikar HCT 20mg/5mg/12.5mg tablets (Daiichi Sankyo UK Ltd) | 6180641000033112 | 18986411000001108 | Amlodipine besilate/ Hydrochlorothiazide/ Olmesartan medoxomil |
| Sevikar HCT 40mg/5mg/12.5mg tablets (Daiichi Sankyo UK Ltd) | 6180741000033115 | 18986711000001102 | Amlodipine besilate/ Hydrochlorothiazide/ Olmesartan medoxomil |
| Sevikar HCT 40mg/10mg/12.5mg tablets (Daiichi Sankyo UK Ltd) | 6180841000033113 | 18987011000001101 | Amlodipine besilate/ Hydrochlorothiazide/ |

| | | | |
|---|---|---|---|
| | | | Olmesartan medoxomil |
| Sevikar HCT 40mg/5mg/25mg tablets (Daiichi Sankyo UK Ltd) | 6180941000033117 | 18987311000001103 | Amlodipine besilate/ Hydrochlorothiazide/ Olmesartan medoxomil |
| Sevikar HCT 40mg/10mg/25mg tablets (Daiichi Sankyo UK Ltd) | 6181041000033110 | 18987611000001108 | Amlodipine besilate/ Hydrochlorothiazide/ Olmesartan medoxomil |
| Actelsar HCT 40mg/12.5mg tablets (Actavis UK Ltd) | 9159341000033112 | 24411411000001100 | Hydrochlorothiazide/ Telmisartan |
| Actelsar HCT 80mg/12.5mg tablets (Actavis UK Ltd) | 9159441000033118 | 24412511000001109 | Hydrochlorothiazide/ Telmisartan |
| Actelsar HCT 80mg/25mg tablets (Accord Healthcare Ltd) | 9159541000033117 | 24413611000001103 | Hydrochlorothiazide/ Telmisartan |
| Tolucombi 40mg/12.5mg tablets (Consilient Health Ltd) | 9292641000033116 | 24573211000001101 | Hydrochlorothiazide/ Telmisartan |
| Tolucombi 80mg/12.5mg tablets (Consilient Health Ltd) | 9292741000033113 | 24573411000001102 | Hydrochlorothiazide/ Telmisartan |
| Lisoretic 10mg/12.5mg tablets (Bristol Laboratories Ltd) | 13443441000033115 | 38240211000001100 | Hydrochlorothiazide/ Lisinopril |
| Lisoretic 20mg/12.5mg tablets (Bristol Laboratories Ltd) | 13443541000033119 | 38240011000001105 | Hydrochlorothiazide/ Lisinopril |
| Chlorothiazide 250mg/5ml oral suspension | 246241000033116 | 408039005 | Chlorothiazide |
| Chlorothiazide 200mg/5ml oral suspension | 3924141000033111 | 12502311000001102 | Chlorothiazide |
| Chlorothiazide 250mg tablets | 5093641000033110 | 395516007 | Chlorothiazide |
| Chlorothiazide 150mg/5ml oral suspension | 5899341000033118 | 8358511000001105 | Chlorothiazide |
| Chlorothiazide 25mg/5ml oral suspension | 5899441000033112 | 8358711000001100 | Chlorothiazide |
| Chlorothiazide 250mg/5ml oral solution | 5991141000033110 | 12503011000001109 | Chlorothiazide |
| Spironolactone 4mg / Chlorothiazide 40mg capsules | 7681541000033117 | 18520511000001107 | Chlorothiazide/ Spironolactone |
| Spironolactone 3mg / Chlorothiazide 30mg capsules | 9278541000033112 | 16072811000001103 | Chlorothiazide/ Spironolactone |
| Aprinox 2.5mg tablets (Advanz Pharma) | 74541000033119 | 672111000001100 | Bendroflumethiazide |
| Aprinox 5mg tablets (Amdipharm Plc) | 74641000033118 | 120911000001103 | Bendroflumethiazide |
| Centyl K modified-release tablets (LEO Pharma) | 234841000033110 | 3932711000001105 | Bendroflumethiazide/ Potassium chloride |
| Corgaretic 40mg tablets (Sanofi-Synthelabo Ltd) | 375441000033118 | 3886211000001102 | Bendroflumethiazide/ Nadolol |
| Corgaretic 80mg tablets (Sanofi-Synthelabo Ltd) | 375541000033117 | 4057911000001102 | Bendroflumethiazide/ Nadolol |
| Inderetic 80mg/2.5mg capsules (AstraZeneca UK Ltd) | 756641000033114 | 333111000001104 | Bendroflumethiazide/ Propranolol hydrochloride |
| Inderex 160mg/5mg modified-release capsules (AstraZeneca UK Ltd) | 756741000033117 | 350811000001100 | Bendroflumethiazide/ Propranolol hydrochloride |
| Neo-Naclex 5mg tablets (Mercury Pharma Group Ltd) | 965341000033114 | 817911000001100 | Bendroflumethiazide |
| Neo-Naclex-K modified-release tablets (Mercury Pharma Group Ltd) | 967041000033118 | 3638211000001100 | Bendroflumethiazide/ Potassium chloride |
| Prestim tablets (Meda Pharmaceuticals Ltd) | 1132741000033111 | 98411000001109 | Bendroflumethiazide/ Timolol maleate |
| Tenben 25mg/1.25mg capsules (Galen Ltd) | 1414241000033117 | 721411000001106 | Atenolol/ Bendroflumethiazide |
| Urizide 5mg tablets (Dr Reddy's Laboratories (UK) Ltd) | 1495441000033116 | 898211000001109 | Bendroflumethiazide |

| | | | |
|---|---|---|---|
| Urizide 2.5mg tablets (Dr Reddy's Laboratories (UK) Ltd) | 2955941000033119 | 485311000001101 | Bendroflumethiazide |
| Bendroflumethiazide 2.5mg tablets | 3083141000033112 | 317919004 | Bendroflumethiazide |
| Bendroflumethiazide 5mg tablets | 3083241000033117 | 317920005 | Bendroflumethiazide |
| Bendroflumethiazide 2.5mg/5ml oral suspension | 3151841000033117 | 8306811000001101 | Bendroflumethiazide |
| Bendroflumethiazide 2.5mg / Potassium chloride 573mg (potassium 7.7mmol) modified-release tablets | 3161041000033115 | 35910011000001105 | Bendroflumethiazide/ Potassium chloride |
| Bendroflumethiazide 2.5mg / Potassium chloride 630mg (potassium 8.4mmol) modified-release tablets | 3162841000033118 | 35910111000001106 | Bendroflumethiazide/ Potassium chloride |
| Timolol 10mg / Bendroflumethiazide 2.5mg tablets | 3346641000033118 | 318556002 | Bendroflumethiazide/ Timolol maleate |
| Bendroflumethiazide 5mg/5ml oral suspension | 5401741000033110 | 8306711000001109 | Bendroflumethiazide |
| Bendroflumethiazide 1.25mg/5ml oral suspension | 5453941000033119 | 8307011000001105 | Bendroflumethiazide |
| Neo-Naclex 2.5mg tablets (Advanz Pharma) | 6045641000033115 | 18149011000001101 | Bendroflumethiazide |
| Nephril 1mg tablets (Pfizer Ltd) | 965441000033115 | 4549211000001100 | Polythiazide |
| Polythiazide 1mg tablets | 1110141000033119 | 317967009 | Polythiazide |

#### Table S63. Thiazide like diuretics Aurum product codes

| Term from EMIS | Prod code ID | dmd ID | Drug substance name |
|---|---|---|---|
| Indapamide 1.5mg modified-release tablets | 765741000033116 | 39020711000001100 | Indapamide |
| Indapamide 2.5mg tablets | 768641000033118 | 39696211000001108 | Indapamide hemihydrate |
| Nindaxa 2.5 tablets (Ashbourne Pharmaceuticals Ltd) | 769741000033118 | 424311000001108 | Indapamide hemihydrate |
| Natrilix SR 1.5mg tablets (Servier Laboratories Ltd) | 953141000033119 | 456611000001108 | Indapamide |
| Natrilix 2.5mg tablets (Servier Laboratories Ltd) | 955341000033117 | 321811000001109 | Indapamide hemihydrate |
| Coversyl Plus tablets (Servier Laboratories Ltd) | 2739541000033117 | 562511000001109 | Indapamide/ Perindopril erbumine |
| Perindopril erbumine 4mg / Indapamide 1.25mg tablets | 3161641000033114 | 3437611000001100 | Indapamide/ Perindopril erbumine |
| Perindopril arginine 5mg / Indapamide 1.25mg tablets | 4454541000033118 | 13454311000001101 | Indapamide/ Perindopril arginine |
| Coversyl Arginine Plus 5mg/1.25mg tablets (Servier Laboratories Ltd) | 4454641000033117 | 13444311000001104 | Indapamide/ Perindopril arginine |
| Ethibide XL 1.5mg tablets (Genus Pharmaceuticals Ltd) | 4571241000033110 | 13824811000001102 | Indapamide |
| Tensaid XL 1.5mg tablets (Mylan) | 4656741000033119 | 14242211000001100 | Indapamide |
| Indipam XL 1.5mg tablets (Accord Healthcare Ltd) | 5053141000033113 | 15436111000001104 | Indapamide |
| Mapemid XL 1.5mg tablets (Teva UK Ltd) | 5128741000033110 | 15600711000001107 | Indapamide |
| Rawel XL 1.5mg tablets (Consilient Health Ltd) | 5566041000033114 | 16737911000001105 | Indapamide |
| Varbim XL 1.5mg tablets (Teva UK Ltd) | 5816741000033112 | 14693611000001102 | Indapamide |
| Indapamide 2.5mg/5ml oral suspension | 8197041000033112 | 21578911000001105 | Indapamide |
| Perindopril tosilate 5mg / Indapamide 1.25mg tablets | 8263641000033112 | 21940011000001101 | Indapamide/ Perindopril tosilate |
| Cardide SR 1.5mg tablets (Teva UK Ltd) | 9121841000033115 | 24331611000001104 | Indapamide |
| Alkapamid XL 1.5mg tablets (HBS Healthcare Ltd) | 12312641000033118 | 34494911000001106 | Indapamide |
| Lorvacs XL 1.5mg tablets (Torrent Pharma (UK) Ltd) | 12988441000033113 | 36812711000001103 | Indapamide |
| Diurexan 20mg tablets (Mylan) | 470741000033113 | 348911000001105 | Xipamide |
| Xipamide 20mg tablets | 1543941000033111 | 317970008 | Xipamide |
| Viskaldix tablets (Advanz Pharma) | 1523941000033114 | 3638411000001101 | Clopamide/ Pindolol |
| Pindolol 10mg / Clopamide 5mg tablets | 3163141000033117 | 318552000 | Clopamide/ Pindolol |

#### Table S64. Potassium sparing diuretics Aurum product codes

| Term from EMIS | Prod code ID | dmd ID | Drug substance name |
|---|---|---|---|
| Amiloride 5mg/5ml oral solution sugar free | 53841000033117 | 35900111000001108 | Amiloride hydrochloride |
| Amiloride 5mg tablets | 57141000033118 | 318052005 | Amiloride hydrochloride |
| Amilmaxco 5mg/50mg tablets (Ashbourne Pharmaceuticals Ltd) | 57341000033115 | 712211000001108 | Amiloride hydrochloride/ Hydrochlorothiazide |
| Amil-Co 5mg/50mg tablets (IVAX Pharmaceuticals UK Ltd) | 60341000033110 | 636611000001107 | Amiloride hydrochloride/ Hydrochlorothiazide |
| Burinex A 5mg/1mg tablets (LEO Pharma) | 172341000033112 | 33911000001104 | Amiloride hydrochloride/ Bumetanide |
| Co-amilozide 5mg/50mg/5ml oral solution | 351941000033111 | 8792311000001109 | Amiloride hydrochloride/ Hydrochlorothiazide |
| Co-amilozide 5mg/50mg tablets | 369141000033118 | 377566005 | Amiloride hydrochloride/ Hydrochlorothiazide |
| Co-amilofruse 5mg/40mg tablets | 376641000033112 | 318136009 | Amiloride hydrochloride/ Furosemide |
| Co-amilofruse 10mg/80mg tablets | 376741000033115 | 318137000 | Amiloride hydrochloride/ Furosemide |
| Co-amilofruse 2.5mg/20mg tablets | 376841000033113 | 318135008 | Amiloride hydrochloride/ Furosemide |
| Co-amilozide 2.5mg/25mg tablets | 376941000033117 | 318121006 | Amiloride hydrochloride/ Hydrochlorothiazide |
| Frumil LS 20mg/2.5mg tablets (Sanofi) | 614341000033113 | 550711000001106 | Amiloride hydrochloride/ Furosemide |
| Frumil 40mg/5mg tablets (Sanofi) | 614641000033117 | 427411000001106 | Amiloride hydrochloride/ Furosemide |
| Fru-Co 5mg/40mg tablets (Teva UK Ltd) | 615241000033116 | 818511000001106 | Amiloride hydrochloride/ Furosemide |
| Frumil Forte 10mg/80mg tablets (Sanofi) | 615641000033118 | 82611000001106 | Amiloride hydrochloride/ Furosemide |
| Kalten capsules (M & A Pharmachem Ltd) | 793741000033117 | 237011000001100 | Amiloride hydrochloride/ Atenolol/ Hydrochlorothiazide |
| Lasoride 5mg/40mg tablets (Sanofi) | 818941000033112 | 678511000001106 | Amiloride hydrochloride/ Furosemide |
| Moducren tablets (Organon Pharma (UK) Ltd) | 935341000033118 | 74311000001101 | Amiloride hydrochloride/ Hydrochlorothiazide/ Timolol maleate |
| Moduret 25 tablets (Organon Pharma (UK) Ltd) | 935441000033112 | 314211000001100 | Amiloride hydrochloride/ Hydrochlorothiazide |
| Moduretic 5mg/50mg tablets (Merck Sharp & Dohme (UK) Ltd) | 935541000033113 | 453811000001103 | Amiloride hydrochloride/ Hydrochlorothiazide |

| | | | |
|---|---|---|---|
| Navispare 2.5mg/250microgram tablets (Advanz Pharma) | 956741000033114 | 535711000001100 | Amiloride hydrochloride/ Cyclopenthiazide |
| Froop Co 5mg/40mg tablets (Ashbourne Pharmaceuticals Ltd) | 1611441000033117 | 331311000001103 | Amiloride hydrochloride/ Furosemide |
| Amiloride 5mg / Bumetanide 1mg tablets | 1845641000033119 | 318097001 | Amiloride hydrochloride/ Bumetanide |
| Amiloride 2.5mg / Cyclopenthiazide 250microgram tablets | 1903541000033116 | 318096005 | Amiloride hydrochloride/ Cyclopenthiazide |
| Amilamont 5mg/5ml oral solution sugar free (Rosemont Pharmaceuticals Ltd) | 2101641000033119 | 799711000001108 | Amiloride hydrochloride |
| Amoride 5mg tablets (Dr Reddy's Laboratories (UK) Ltd) | 2956041000033112 | 220711000001102 | Amiloride hydrochloride |
| Timolol 10mg / Amiloride 2.5mg / Hydrochlorothiazide 25mg tablets (Essential Generics Ltd) | 5376441000033114 | 16458411000001107 | Amiloride hydrochloride/ Hydrochlorothiazide/ Timolol maleate |
| Co-amilofruse 5mg/40mg/5ml oral suspension | 5710441000033117 | 8427011000001108 | Amiloride hydrochloride/ Furosemide |
| Co-triamterzide 50mg/25mg tablets | 370241000033111 | 410896007 | Hydrochlorothiazide/ Triamterene |
| Dytac 50mg capsules (Advanz Pharma) | 491641000033116 | 3907911000001100 | Triamterene |
| Dytide capsules (Mercury Pharma Group Ltd) | 491741000033113 | 714911000001108 | Benzthiazide/ Triamterene |
| Dyazide 50mg/25mg tablets (Advanz Pharma) | 492341000033115 | 132811000001106 | Hydrochlorothiazide/ Triamterene |
| Frusene 50mg/40mg tablets (Orion Pharma (UK) Ltd) | 615041000033112 | 25411000001108 | Furosemide/ Triamterene |
| Kalspare tablets (DHP Healthcare Ltd) | 796541000033115 | 3252011000001105 | Chlortalidone/ Triamterene |
| Triamterene 50mg capsules | 1455341000033115 | 318082004 | Triamterene |
| Triamaxco 50mg/25mg tablets (Ashbourne Pharmaceuticals Ltd) | 1468141000033118 | 721311000001104 | Hydrochlorothiazide/ Triamterene |
| Triam-Co 50mg/25mg tablets (IVAX Pharmaceuticals UK Ltd) | 1470341000033112 | 191211000001106 | Hydrochlorothiazide/ Triamterene |
| Triamterene 50mg / Furosemide 40mg tablets | 3092741000033119 | 318101005 | Furosemide/ Triamterene |
| Triamterene 50mg / Benzthiazide 25mg capsules | 3161941000033119 | 318098006 | Benzthiazide/ Triamterene |
| Triamterene 50mg / Chlortalidone 50mg tablets | 3162341000033110 | 318100006 | Chlortalidone/ Triamterene |

#### Table S65. Dihydropyridines Aurum product codes

| Term from EMIS | Prod code ID | dmd ID | Drug substance name |
|---|---|---|---|
| Istin 5mg tablets (Upjohn UK Ltd) | 787941000033112 | 172711000001100 | Amlodipine |
| Istin 10mg tablets (Upjohn UK Ltd) | 788041000033110 | 408111000001107 | Amlodipine |
| Amlodipine 5mg tablets | 3038041000033111 | 39732011000001102 | Amlodipine |
| Amlodipine 10mg tablets | 3038141000033110 | 39731911000001109 | Amlodipine |
| Amlostin 5mg tablets (Discovery Pharmaceuticals) | 3188741000033111 | 8046211000001107 | Amlodipine |
| Amlostin 10mg tablets (Discovery Pharmaceuticals) | 3188841000033118 | 8046411000001106 | Amlodipine |
| Amlodipine 10mg/5ml oral suspension | 3963741000033115 | 8278111000001105 | Amlodipine |
| Amlodipine 5mg / Valsartan 80mg tablets | 4021441000033114 | 11160311000001109 | Amlodipine besilate/ Valsartan |
| Amlodipine 5mg / Valsartan 160mg tablets | 4021541000033110 | 11160211000001101 | Amlodipine besilate/ Valsartan |
| Amlodipine 10mg / Valsartan 160mg tablets | 4021641000033111 | 11160111000001107 | Amlodipine besilate/ Valsartan |
| Exforge 5mg/80mg tablets (Novartis Pharmaceuticals UK Ltd) | 4021741000033119 | 11161811000001108 | Amlodipine besilate/ Valsartan |
| Exforge 5mg/160mg tablets (Novartis Pharmaceuticals UK Ltd) | 4021841000033112 | 11161511000001105 | Amlodipine besilate/ Valsartan |
| Exforge 10mg/160mg tablets (Novartis Pharmaceuticals UK Ltd) | 4021941000033116 | 11160711000001108 | Amlodipine besilate/ Valsartan |
| Amlodipine 5mg/5ml oral solution sugar free | 4508041000033119 | 29826311000001101 | Amlodipine |
| Amlodipine 1.5mg/5ml oral suspension | 4897741000033118 | 15773511000001108 | Amlodipine |
| Olmesartan medoxomil 20mg / Amlodipine 5mg tablets | 5005841000033114 | 429502004 | Amlodipine besilate/ Olmesartan medoxomil |
| Olmesartan medoxomil 40mg / Amlodipine 5mg tablets | 5005941000033118 | 429503009 | Amlodipine besilate/ Olmesartan medoxomil |
| Olmesartan medoxomil 40mg / Amlodipine 10mg tablets | 5006041000033111 | 429678006 | Amlodipine besilate/ Olmesartan medoxomil |
| Sevikar 20mg/5mg tablets (Daiichi Sankyo UK Ltd) | 5006141000033110 | 15773211000001105 | Amlodipine besilate/ Olmesartan medoxomil |
| Sevikar 40mg/5mg tablets (Daiichi Sankyo UK Ltd) | 5006241000033115 | 15772911000001108 | Amlodipine besilate/ Olmesartan medoxomil |
| Sevikar 40mg/10mg tablets (Daiichi Sankyo UK Ltd) | 5006341000033113 | 15772611000001102 | Amlodipine besilate/ Olmesartan medoxomil |
| Amlodipine 5mg/5ml oral solution | 5490241000033113 | 13892511000001100 | Amlodipine |
| Amlodipine 5mg/5ml oral suspension | 5888741000033110 | 8278311000001107 | Amlodipine |
| Sevikar HCT 20mg/5mg/12.5mg tablets (Daiichi Sankyo UK Ltd) | 6180641000033112 | 18986411000001108 | Amlodipine besilate/ Hydrochlorothiazide/ Olmesartan medoxomil |
| Sevikar HCT 40mg/5mg/12.5mg tablets (Daiichi Sankyo UK Ltd) | 6180741000033115 | 18986711000001102 | Amlodipine besilate/ Hydrochlorothiazide/ Olmesartan medoxomil |
| Sevikar HCT 40mg/10mg/12.5mg tablets (Daiichi Sankyo UK Ltd) | 6180841000033113 | 18987011000001101 | Amlodipine besilate/ Hydrochlorothiazide/ Olmesartan medoxomil |

| | | | |
|---|---|---|---|
| Sevikar HCT 40mg/5mg/25mg tablets (Daiichi Sankyo UK Ltd) | 6180941000033117 | 18987311000001103 | Amlodipine besilate/ Hydrochlorothiazide/ Olmesartan medoxomil |
| Sevikar HCT 40mg/10mg/25mg tablets (Daiichi Sankyo UK Ltd) | 6181041000033110 | 18987611000001108 | Amlodipine besilate/ Hydrochlorothiazide/ Olmesartan medoxomil |
| Amlodipine 10mg/5ml oral solution | 7740641000033112 | 20478011000001105 | Amlodipine |
| Perindopril erbumine 4mg / Amlodipine 5mg tablets | 9105241000033114 | 23985011000001108 | Amlodipine besilate/ Perindopril erbumine |
| Perindopril erbumine 4mg / Amlodipine 10mg tablets | 9105341000033116 | 23984911000001108 | Amlodipine besilate/ Perindopril erbumine |
| Perindopril erbumine 8mg / Amlodipine 5mg tablets | 9105441000033110 | 23985211000001103 | Amlodipine besilate/ Perindopril erbumine |
| Perindopril erbumine 8mg / Amlodipine 10mg tablets | 9105541000033111 | 23985111000001109 | Amlodipine besilate/ Perindopril erbumine |
| Amlodipine 10mg/5ml oral solution sugar free | 10387441000033111 | 29826211000001109 | Amlodipine |
| Amlodipine 5mg/5ml oral suspension sugar free | 12682241000033110 | 36237311000001101 | Amlodipine |
| Amlodipine 2.5mg tablets | 13422241000033114 | 429828006 | Amlodipine |
| Felodipine 2.5mg modified-release tablets | 568041000033111 | 39020311000001104 | Felodipine |
| Felodipine 10mg modified-release tablets | 569041000033115 | 39020511000001105 | Felodipine |
| Felodipine 5mg modified-release tablets | 569141000033116 | 39020611000001109 | Felodipine |
| Plendil 2.5mg modified-release tablets (AstraZeneca UK Ltd) | 1093941000033116 | 562711000001104 | Felodipine |
| Plendil 10mg modified-release tablets (AstraZeneca UK Ltd) | 1094041000033119 | 48511000001101 | Felodipine |
| Plendil 5mg modified-release tablets (AstraZeneca UK Ltd) | 1094141000033115 | 490211000001101 | Felodipine |
| Triapin 5mg/5mg modified-release tablets (Sanofi) | 1834341000033110 | 3887911000001109 | Felodipine/ Ramipril |
| Cabren 2.5mg modified-release tablets (Teva UK Ltd) | 2928841000033117 | 3800311000001106 | Felodipine |
| Cabren 5mg modified-release tablets (Teva UK Ltd) | 2928941000033113 | 3800511000001100 | Felodipine |
| Cabren 10mg modified-release tablets (Teva UK Ltd) | 2929041000033116 | 3800711000001105 | Felodipine |
| Felotens XL 5mg tablets (Thornton & Ross Ltd) | 2979941000033119 | 4785111000001103 | Felodipine |
| Felotens XL 10mg tablets (Thornton & Ross Ltd) | 2980041000033115 | 4785511000001107 | Felodipine |
| Vascalpha 5mg modified-release tablets (Accord Healthcare Ltd) | 3034341000033117 | 5638311000001102 | Felodipine |
| Vascalpha 10mg modified-release tablets (Accord Healthcare Ltd) | 3034441000033111 | 5638811000001106 | Felodipine |
| Cardioplen XL 5mg tablets (Chiesi Ltd) | 3154841000033119 | 7887011000001104 | Felodipine |
| Cardioplen XL 10mg tablets (Chiesi Ltd) | 3154941000033110 | 7887511000001107 | Felodipine |
| Felodipine 5mg modified-release / Ramipril 5mg tablets | 3163241000033112 | 318177008 | Felodipine/ Ramipril |
| Felodipine 2.5mg modified-release / Ramipril 2.5mg tablets | 3163341000033119 | 318176004 | Felodipine/ Ramipril |
| Felogen XL 5mg tablets (Mylan) | 3177441000033110 | 4972811000001103 | Felodipine |
| Felogen XL 10mg tablets (Mylan) | 3177541000033111 | 4973011000001100 | Felodipine |
| Cardioplen XL 2.5mg tablets (Chiesi Ltd) | 4152841000033112 | 11506711000001103 | Felodipine |
| Felotens XL 2.5mg tablets (Thornton & Ross Ltd) | 4429241000033115 | 13127311000001107 | Felodipine |
| Neofel XL 2.5mg tablets (Actavis UK Ltd) | 4521541000033113 | 18167311000001103 | Felodipine |
| Neofel XL 5mg tablets (Kent Pharma (UK) Ltd) | 4521641000033114 | 8090111000001106 | Felodipine |

| | | | |
|---|---|---|---|
| Neofel XL 10mg tablets (Kent Pharma (UK) Ltd) | 4521741000033117 | 8089811000001107 | Felodipine |
| Folpik XL 10mg tablets (Teva UK Ltd) | 5576341000033118 | 5008911000001100 | Felodipine |
| Folpik XL 2.5mg tablets (Teva UK Ltd) | 5576441000033112 | 13565311000001101 | Felodipine |
| Folpik XL 5mg tablets (Teva UK Ltd) | 5576541000033113 | 5008511000001107 | Felodipine |
| Triapin 2.5mg/2.5mg modified-release tablets (Sanofi) | 8264941000033119 | 4093211000001109 | Felodipine/ Ramipril |
| Parmid XL 2.5mg tablets (Sandoz Ltd) | 9121541000033117 | 24221811000001105 | Felodipine |
| Parmid XL 5mg tablets (Sandoz Ltd) | 9121641000033116 | 7387911000001103 | Felodipine |
| Parmid XL 10mg tablets (Sandoz Ltd) | 9121741000033113 | 7388311000001103 | Felodipine |
| Isradipine 2.5mg tablets | 784641000033112 | 319280009 | Isradipine |
| Prescal 2.5mg tablets (Novartis Pharmaceuticals UK Ltd) | 1140241000033117 | 3689711000001108 | Isradipine |
| Lercanidipine 10mg tablets | 825941000033112 | 319316005 | Lercanidipine hydrochloride |
| Zanidip 10mg tablets (Recordati Pharmaceuticals Ltd) | 1550841000033111 | 20011000001105 | Lercanidipine hydrochloride |
| Lercanidipine 20mg tablets | 3908741000033114 | 10225911000001102 | Lercanidipine hydrochloride |
| Zanidip 20mg tablets (Recordati Pharmaceuticals Ltd) | 3908841000033116 | 10198711000001102 | Lercanidipine hydrochloride |
| Cardene 20mg capsules (Astellas Pharma Ltd) | 176741000033114 | 344811000001108 | Nicardipine hydrochloride |
| Cardene 30mg capsules (Astellas Pharma Ltd) | 176841000033116 | 291111000001102 | Nicardipine hydrochloride |
| Cardene SR 30mg capsules (Astellas Pharma Ltd) | 198941000033118 | 540311000001105 | Nicardipine hydrochloride |
| Cardene SR 45mg capsules (Astellas Pharma Ltd) | 199041000033110 | 118811000001102 | Nicardipine hydrochloride |
| Nicardipine 30mg capsules | 967141000033119 | 319218009 | Nicardipine hydrochloride |
| Nicardipine 20mg capsules | 967741000033115 | 319217004 | Nicardipine hydrochloride |
| Nicardipine 30mg modified-release capsules | 971441000033112 | 39021711000001108 | Nicardipine hydrochloride |
| Nicardipine 45mg modified-release capsules | 971541000033113 | 39021811000001100 | Nicardipine hydrochloride |
| Adalat 10mg capsules (Bayer Plc) | 14041000033115 | 782511000001108 | Nifedipine |
| Adalat 5mg capsules (Bayer Plc) | 14141000033116 | 27111000001107 | Nifedipine |
| Adalat LA 30mg tablets (Bayer Plc) | 17141000033112 | 2881311000001105 | Nifedipine |
| Adalat LA 60mg tablets (Bayer Plc) | 17241000033117 | 235511000001104 | Nifedipine |
| Adipine MR 20 tablets (Chiesi Ltd) | 18241000033116 | 74111000001103 | Nifedipine |
| Adipine MR 10 tablets (Chiesi Ltd) | 18341000033114 | 833611000001109 | Nifedipine |
| Adalat retard 20mg tablets (Bayer Plc) | 19341000033119 | 5011000001109 | Nifedipine |
| Adalat retard 10mg tablets (Bayer Plc) | 19441000033113 | 569011000001108 | Nifedipine |
| Angiopine 10 capsules (Ashbourne Pharmaceuticals Ltd) | 61841000033111 | 811311000001107 | Nifedipine |
| Angiopine MR 20mg tablets (Ashbourne Pharmaceuticals Ltd) | 65241000033114 | 865011000001105 | Nifedipine |
| Angiopine MR 10mg tablets (Ashbourne Pharmaceuticals Ltd) | 66141000033114 | 568911000001104 | Nifedipine |
| Beta-Adalat modified-release capsules (Bayer Plc) | 124341000033119 | 3142711000001107 | Atenolol/ Nifedipine |
| Cardilate MR 20mg tablets (IVAX Pharmaceuticals UK Ltd) | 199141000033114 | 905711000001103 | Nifedipine |
| Coracten SR 10mg capsules (UCB Pharma Ltd) | 336641000033114 | 126411000001108 | Nifedipine |

| | | | |
|---|---|---|---|
| Coracten SR 20mg capsules (UCB Pharma Ltd) | 364241000033112 | 389611000001101 | Nifedipine |
| Fortipine LA 40 tablets (Advanz Pharma) | 605941000033110 | 188711000001108 | Nifedipine |
| Hypolar Retard 20 tablets (Sandoz Ltd) | 738141000033119 | 677411000001108 | Nifedipine |
| Nifedipine 10mg capsules | 967241000033114 | 319223009 | Nifedipine |
| Nifedipine 5mg capsules | 967341000033116 | 319222004 | Nifedipine |
| Nifedipine 20mg modified-release tablets | 970941000033113 | 39022811000001109 | Nifedipine |
| Nifedipine 10mg modified-release capsules | 971141000033116 | 39022911000001104 | Nifedipine |
| Nifedipine 30mg modified-release tablets | 971241000033111 | 39111711000001105 | Nifedipine |
| Nifedipine 60mg modified-release tablets | 971341000033118 | 39022611000001105 | Nifedipine |
| Nifedipine 10mg modified-release tablets | 971841000033110 | 39022711000001101 | Nifedipine |
| Nifedipine 40mg modified-release tablets | 972441000033116 | 39107611000001108 | Nifedipine |
| Nifedipine 20mg modified-release capsules | 974141000033119 | 39022511000001106 | Nifedipine |
| Tenif 50mg/20mg modified-release capsules (AstraZeneca UK Ltd) | 1414941000033114 | 3142511000001102 | Atenolol/ Nifedipine |
| Tensipine MR 10 tablets (Genus Pharmaceuticals Ltd) | 1421241000033110 | 413111000001101 | Nifedipine |
| Tensipine MR 20 tablets (Genus Pharmaceuticals Ltd) | 1421341000033117 | 385611000001104 | Nifedipine |
| Cardilate MR 10mg tablets (Teva UK Ltd) | 1580841000033116 | 25911000001100 | Nifedipine |
| Slofedipine 20mg tablets (Sterwin Medicines) | 1697641000033111 | 843411000001105 | Nifedipine |
| Coracten XL 30mg capsules (UCB Pharma Ltd) | 1724941000033113 | 162811000001100 | Nifedipine |
| Coracten XL 60mg capsules (UCB Pharma Ltd) | 1725041000033113 | 3381511000001105 | Nifedipine |
| Nifedipress MR 10 tablets (Dexcel-Pharma Ltd) | 1745041000033110 | 904011000001104 | Nifedipine |
| Nifedipress MR 20 tablets (Dexcel-Pharma Ltd) | 1745141000033114 | 619111000001101 | Nifedipine |
| Slofedipine XL 30mg tablets (Zentiva Pharma UK Ltd) | 1752941000033115 | 2882011000001104 | Nifedipine |
| Slofedipine XL 60 tablets (Zentiva Pharma UK Ltd) | 1753041000033113 | 630411000001107 | Nifedipine |
| Adalat LA 20mg tablets (Bayer Plc) | 1766341000033113 | 881811000001100 | Nifedipine |
| Nifedipine 30mg modified-release capsules | 1778741000033118 | 39107511000001109 | Nifedipine |
| Nifedipine 60mg modified-release capsules | 1778841000033111 | 38896511000001101 | Nifedipine |
| Coroday MR 20mg tablets (Mylan) | 2051241000033116 | 309611000001102 | Nifedipine |
| Nifopress Retard 20mg tablets (Advanz Pharma) | 2189541000033117 | 280811000001100 | Nifedipine |
| Calchan MR 10 tablets (Ranbaxy (UK) Ltd) | 2295641000033116 | 627111000001104 | Nifedipine |
| Calchan MR 20 tablets (Ranbaxy (UK) Ltd) | 2295741000033113 | 17011000001100 | Nifedipine |
| Nifedipine 20mg/ml oral drops | 2639341000033117 | 9096811000001100 | Nifedipine |
| Hypolar XL 30 tablets (Sandoz Ltd) | 2776541000033111 | 2881811000001101 | Nifedipine |
| Valni 20 Retard tablets (Tillomed Laboratories Ltd) | 2955041000033115 | 693311000001101 | Nifedipine |
| Adipine XL 30mg tablets (Chiesi Ltd) | 3225441000033117 | 9049711000001108 | Nifedipine |
| Adipine XL 60mg tablets (Chiesi Ltd) | 3225541000033116 | 9049911000001105 | Nifedipine |
| Atenolol 50mg / Nifedipine 20mg modified-release capsules | 3246941000033118 | 35903311000001104 | Atenolol/ Nifedipine |
| Nimodrel XL 30mg tablets (Zurich Pharmaceuticals) | 3952641000033110 | 10189111000001106 | Nifedipine |
| Nimodrel XL 60mg tablets (Zurich Pharmaceuticals) | 3952741000033118 | 10189311000001108 | Nifedipine |
| Valni XL 30mg tablets (Zentiva Pharma UK Ltd) | 4451941000033110 | 13401911000001109 | Nifedipine |

| | | | |
|---|---|---|---|
| Valni XL 60mg tablets (Zentiva Pharma UK Ltd) | 4452041000033116 | 13402111000001101 | Nifedipine |
| Nifedipine 5mg/5ml oral suspension | 5056241000033112 | 8670311000001108 | Nifedipine |
| Nifedipine 2.5mg/5ml oral suspension | 5402541000033112 | 12303311000001109 | Nifedipine |
| Neozipine XL 30mg tablets (Kent Pharma (UK) Ltd) | 5817441000033119 | 10751211000001100 | Nifedipine |
| Neozipine XL 60mg tablets (Kent Pharma (UK) Ltd) | 5817541000033118 | 10751411000001101 | Nifedipine |
| Adanif XL 30mg tablets (Advanz Pharma) | 5891941000033115 | 17666011000001106 | Nifedipine |
| Adanif XL 60mg tablets (Advanz Pharma) | 5892041000033114 | 17666211000001101 | Nifedipine |
| Nifedipine 10mg/5ml oral suspension | 5973741000033117 | 8670111000001106 | Nifedipine |
| Nidef 30mg modified-release tablets (Morningside Healthcare Ltd) | 12372341000033117 | 34685211000001104 | Nifedipine |
| Nidef 60mg modified-release tablets (Morningside Healthcare Ltd) | 12372441000033111 | 34685811000001103 | Nifedipine |
| Nimodipine 10mg/50ml solution for infusion vials | 970741000033110 | 36031711000001104 | Nimodipine |
| Nimodipine 30mg tablets | 977441000033119 | 323273000 | Nimodipine |
| Nimotop 30mg tablets (Bayer Plc) | 977741000033114 | 3879211000001104 | Nimodipine |
| Nisoldipine 10mg modified-release tablets | 972141000033112 | 36031811000001107 | Nisoldipine |
| Nisoldipine 20mg modified-release tablets | 972241000033117 | 36031911000001102 | Nisoldipine |
| Nisoldipine 30mg modified-release tablets | 972341000033110 | 36032011000001109 | Nisoldipine |
| Syscor MR 10 tablets (Forest Laboratories UK Ltd) | 1401341000033110 | 3877011000001100 | Nisoldipine |
| Syscor MR 20 tablets (Forest Laboratories UK Ltd) | 1401441000033116 | 4070011000001100 | Nisoldipine |

#### Table S66. Non Dihydropyridines Aurum product codes

| Term from EMIS | Prod code ID | dmd ID | Drug substance name |
|---|---|---|---|
| Adizem-SR 120mg capsules (Napp Pharmaceuticals Ltd) | 17341000033110 | 2887311000001104 | Diltiazem hydrochloride |
| Adizem-SR 180mg capsules (Napp Pharmaceuticals Ltd) | 17441000033116 | 2886711000001103 | Diltiazem hydrochloride |
| Adizem-SR 90mg capsules (Napp Pharmaceuticals Ltd) | 17641000033119 | 2887011000001102 | Diltiazem hydrochloride |
| Adizem-XL 120mg capsules (Napp Pharmaceuticals Ltd) | 17741000033111 | 2937811000001108 | Diltiazem hydrochloride |
| Adizem-XL 180mg capsules (Napp Pharmaceuticals Ltd) | 17841000033118 | 2938011000001101 | Diltiazem hydrochloride |
| Adizem-XL 240mg capsules (Napp Pharmaceuticals Ltd) | 17941000033114 | 2886311000001102 | Diltiazem hydrochloride |
| Adizem-XL 300mg capsules (Napp Pharmaceuticals Ltd) | 18041000033112 | 2886111000001104 | Diltiazem hydrochloride |
| Adizem-SR 120mg tablets (Napp Pharmaceuticals Ltd) | 19641000033110 | 2885611000001102 | Diltiazem hydrochloride |
| Angitil SR 90 capsules (Ethypharm UK Ltd) | 65341000033116 | 857011000001109 | Diltiazem hydrochloride |
| Angitil SR 120 capsules (Ethypharm UK Ltd) | 65441000033110 | 298811000001103 | Diltiazem hydrochloride |
| Angitil SR 180 capsules (Ethypharm UK Ltd) | 65541000033111 | 336611000001101 | Diltiazem hydrochloride |
| Angiozem CR 90mg tablets (Ashbourne Pharmaceuticals Ltd) | 65941000033117 | 440711000001104 | Diltiazem hydrochloride |
| Angiozem CR 120mg tablets (Ashbourne Pharmaceuticals Ltd) | 66041000033110 | 144811000001101 | Diltiazem hydrochloride |
| Angiozem 60mg modified-release tablets (Ashbourne Pharmaceuticals Ltd) | 71141000033114 | 672311000001103 | Diltiazem hydrochloride |
| Calcicard CR 120mg tablets (Teva UK Ltd) | 199241000033119 | 104111000001100 | Diltiazem hydrochloride |
| Calcicard CR 90mg tablets (Teva UK Ltd) | 199341000033112 | 219611000001107 | Diltiazem hydrochloride |
| Diltiazem 120mg modified-release capsules | 451841000033110 | 39023211000001102 | Diltiazem hydrochloride |
| Diltiazem 180mg modified-release capsules | 451941000033119 | 39023311000001105 | Diltiazem hydrochloride |
| Diltiazem 300mg modified-release capsules | 452041000033113 | 39023611000001100 | Diltiazem hydrochloride |
| Diltiazem 90mg modified-release capsules | 452141000033112 | 39023111000001108 | Diltiazem hydrochloride |
| Dilzem SR 60 capsules (Teva UK Ltd) | 452641000033119 | 417111000001109 | Diltiazem hydrochloride |
| Dilzem SR 90 capsules (Teva UK Ltd) | 452741000033111 | 682311000001105 | Diltiazem hydrochloride |
| Dilzem SR 120 capsules (Teva UK Ltd) | 452841000033118 | 5711000001106 | Diltiazem hydrochloride |
| Diltiazem 60mg modified-release capsules | 452941000033114 | 39023011000001107 | Diltiazem hydrochloride |
| Dilzem XL 120 capsules (Teva UK Ltd) | 453041000033116 | 243111000001108 | Diltiazem hydrochloride |
| Dilzem XL 180 capsules (Teva UK Ltd) | 453141000033117 | 254911000001106 | Diltiazem hydrochloride |
| Dilzem XL 240 capsules (Teva UK Ltd) | 453241000033112 | 733511000001107 | Diltiazem hydrochloride |
| Diltiazem 240mg modified-release capsules | 453741000033118 | 39023511000001104 | Diltiazem hydrochloride |
| Diltiazem 200mg modified-release capsules | 453941000033115 | 39023411000001103 | Diltiazem hydrochloride |

| | | | |
|---|---|---|---|
| Diltiazem 60mg modified-release tablets | 454041000033118 | 39023811000001101 | Diltiazem hydrochloride |
| Diltiazem 90mg modified-release tablets | 458841000033115 | 39023911000001106 | Diltiazem hydrochloride |
| Diltiazem 120mg modified-release tablets | 459641000033113 | 39024011000001109 | Diltiazem hydrochloride |
| Slozem 120mg capsules (Zentiva Pharma UK Ltd) | 1346441000033111 | 130211000001109 | Diltiazem hydrochloride |
| Slozem 180mg capsules (Zentiva Pharma UK Ltd) | 1346541000033112 | 119211000001108 | Diltiazem hydrochloride |
| Slozem 240mg capsules (Zentiva Pharma UK Ltd) | 1346641000033113 | 599811000001104 | Diltiazem hydrochloride |
| Tildiem LA 300 capsules (Sanofi) | 1441741000033119 | 893111000001107 | Diltiazem hydrochloride |
| Tildiem LA 200 capsules (Sanofi) | 1441841000033112 | 261611000001107 | Diltiazem hydrochloride |
| Tildiem Retard 120mg tablets (Sanofi) | 1442741000033113 | 383911000001109 | Diltiazem hydrochloride |
| Tildiem Retard 90mg tablets (Sanofi) | 1442841000033115 | 103611000001105 | Diltiazem hydrochloride |
| Tildiem 60mg modified-release tablets (Sanofi) | 1443841000033113 | 762011000001102 | Diltiazem hydrochloride |
| Viazem XL 120mg capsules (Thornton & Ross Ltd) | 1520141000033118 | 32311000001105 | Diltiazem hydrochloride |
| Viazem XL 180mg capsules (Thornton & Ross Ltd) | 1520241000033113 | 33211000001108 | Diltiazem hydrochloride |
| Viazem XL 240mg capsules (Thornton & Ross Ltd) | 1520341000033115 | 886511000001107 | Diltiazem hydrochloride |
| Viazem XL 300mg capsules (Thornton & Ross Ltd) | 1520441000033114 | 407011000001105 | Diltiazem hydrochloride |
| Viazem XL 360mg capsules (Thornton & Ross Ltd) | 1520541000033110 | 648211000001107 | Diltiazem hydrochloride |
| Angitil XL 240 capsules (Ethypharm UK Ltd) | 1570441000033110 | 75111000001104 | Diltiazem hydrochloride |
| Angitil XL 300 capsules (Ethypharm UK Ltd) | 1570541000033111 | 467111000001105 | Diltiazem hydrochloride |
| Dilcardia SR 120mg capsules (Mylan) | 1602341000033116 | 723811000001107 | Diltiazem hydrochloride |
| Dilcardia SR 60mg capsules (Mylan) | 1602441000033110 | 937011000001101 | Diltiazem hydrochloride |
| Dilcardia SR 90mg capsules (Mylan) | 1602541000033111 | 353011000001101 | Diltiazem hydrochloride |
| Optil 60mg modified-release tablets (Opus Pharmaceuticals Ltd) | 1661641000033116 | 527711000001108 | Diltiazem hydrochloride |
| Diltiazem 360mg modified-release capsules | 1800341000033113 | 39023711000001109 | Diltiazem hydrochloride |
| Zemtard 120 XL capsules (Galen Ltd) | 1833041000033110 | 105211000001107 | Diltiazem hydrochloride |
| Zemtard 180 XL capsules (Galen Ltd) | 1833141000033114 | 813611000001103 | Diltiazem hydrochloride |
| Zemtard 240 XL capsules (Galen Ltd) | 1833241000033119 | 345411000001107 | Diltiazem hydrochloride |
| Zemtard 300 XL capsules (Galen Ltd) | 1833341000033112 | 866811000001107 | Diltiazem hydrochloride |
| Slozem 300mg capsules (Zentiva Pharma UK Ltd) | 2261241000033118 | 550211000001104 | Diltiazem hydrochloride |
| Adizem-XL 200mg capsules (Napp Pharmaceuticals Ltd) | 2653341000033118 | 2886511000001108 | Diltiazem hydrochloride |
| Disogram SR 60mg capsules (Ranbaxy (UK) Ltd) | 2674841000033110 | 469811000001106 | Diltiazem hydrochloride |
| Disogram SR 90mg capsules (Ranbaxy (UK) Ltd) | 2674941000033119 | 256611000001109 | Diltiazem hydrochloride |

| | | | |
|---|---|---|---|
| Disogram SR 120mg capsules (Ranbaxy (UK) Ltd) | 2675041000033119 | 116711000001106 | Diltiazem hydrochloride |
| Disogram SR 180mg capsules (Ranbaxy (UK) Ltd) | 2675341000033117 | 713411000001103 | Diltiazem hydrochloride |
| Disogram SR 240mg capsules (Ranbaxy (UK) Ltd) | 2675641000033113 | 108711000001107 | Diltiazem hydrochloride |
| Disogram SR 300mg capsules (Ranbaxy (UK) Ltd) | 2676041000033111 | 34711000001104 | Diltiazem hydrochloride |
| Bi-Carzem SR 60mg capsules (Tillomed Laboratories Ltd) | 2779141000033112 | 540011000001107 | Diltiazem hydrochloride |
| Bi-Carzem SR 90mg capsules (Tillomed Laboratories Ltd) | 2779241000033117 | 764511000001100 | Diltiazem hydrochloride |
| Bi-Carzem SR 120mg capsules (Tillomed Laboratories Ltd) | 2779341000033110 | 580711000001101 | Diltiazem hydrochloride |
| Diltiazem 2% cream | 2920141000033113 | 8793111000001101 | Diltiazem hydrochloride |
| Zildil 90mg modified-release capsules (Healthcare Pharma Ltd) | 2952341000033113 | 200811000001104 | Diltiazem hydrochloride |
| Zildil SR 120mg capsules (Healthcare Pharma Ltd) | 2952441000033119 | 655411000001104 | Diltiazem hydrochloride |
| Zemret 180 XL capsules (Tillomed Laboratories Ltd) | 2955141000033116 | 924711000001109 | Diltiazem hydrochloride |
| Zemret 240 XL capsules (Tillomed Laboratories Ltd) | 2955241000033111 | 823511000001107 | Diltiazem hydrochloride |
| Zemret 300 XL capsules (Tillomed Laboratories Ltd) | 2955341000033118 | 640311000001101 | Diltiazem hydrochloride |
| Retalzem 60 modified-release tablets (Kent Pharma (UK) Ltd) | 4590741000033116 | 865711000001107 | Diltiazem hydrochloride |
| Diltiazem 60mg/5ml oral suspension | 4953741000033114 | 8456911000001108 | Diltiazem hydrochloride |
| Bi-Carzem XL 240mg capsules (Tillomed Laboratories Ltd) | 5816841000033119 | 884811000001103 | Diltiazem hydrochloride |
| Bi-Carzem XL 300mg capsules (Tillomed Laboratories Ltd) | 5816941000033110 | 158411000001105 | Diltiazem hydrochloride |
| Diltiazem 2% ointment | 5991841000033116 | 8793311000001104 | Diltiazem hydrochloride |
| Diltiazem 60mg/5ml oral solution | 5991941000033112 | 8457011000001107 | Diltiazem hydrochloride |
| Kenzem SR 60mg capsules (Kent Pharma (UK) Ltd) | 9118241000033117 | 8885711000001100 | Diltiazem hydrochloride |
| Kenzem SR 90mg capsules (Kent Pharma (UK) Ltd) | 9118341000033110 | 8886211000001101 | Diltiazem hydrochloride |
| Kenzem SR 120mg capsules (Kent Pharma (UK) Ltd) | 9118441000033116 | 8886511000001103 | Diltiazem hydrochloride |
| Diltiazem 0.2% cream | 13771541000033112 | 18519711000001103 | Diltiazem hydrochloride |
| Cordilox 120mg tablets (IVAX Pharmaceuticals UK Ltd) | 374841000033119 | 867011000001103 | Verapamil hydrochloride |
| Cordilox 160mg tablets (IVAX Pharmaceuticals UK Ltd) | 374941000033110 | 783511000001101 | Verapamil hydrochloride |
| Cordilox 40mg tablets (IVAX Pharmaceuticals UK Ltd) | 375041000033110 | 557111000001106 | Verapamil hydrochloride |
| Cordilox 80mg tablets (IVAX Pharmaceuticals UK Ltd) | 375141000033114 | 258911000001104 | Verapamil hydrochloride |
| Half Securon SR 120mg tablets (Mylan) | 660641000033112 | 846611000001102 | Verapamil hydrochloride |
| Securon IV 5mg/2ml solution for injection ampoules (Mylan) | 1271741000033116 | 4399811000001108 | Verapamil hydrochloride |
| Securon 120mg tablets (Abbott Laboratories Ltd) | 1274741000033119 | 140411000001109 | Verapamil hydrochloride |
| Securon SR 240mg tablets (Mylan) | 1275141000033117 | 540111000001108 | Verapamil hydrochloride |

| | | | |
|---|---|---|---|
| Tarka modified-release capsules (Abbott Laboratories Ltd) | 1406341000033117 | 3691211000001101 | Trandolapril/ Verapamil hydrochloride |
| Univer 120mg modified-release capsules (Teva UK Ltd) | 1489541000033117 | 219711000001103 | Verapamil hydrochloride |
| Univer 180mg modified-release capsules (Teva UK Ltd) | 1489641000033116 | 391811000001102 | Verapamil hydrochloride |
| Univer 240mg modified-release capsules (Teva UK Ltd) | 1489741000033113 | 65411000001102 | Verapamil hydrochloride |
| Verapamil 120mg modified-release tablets | 1509041000033119 | 35367911000001103 | Verapamil hydrochloride |
| Verapamil 240mg modified-release tablets | 1509141000033115 | 38750211000001107 | Verapamil hydrochloride |
| Verapamil 40mg/5ml oral solution sugar free | 1509841000033114 | 4139411000001106 | Verapamil hydrochloride |
| Verapamil 120mg modified-release capsules | 1511141000033118 | 36565011000001105 | Verapamil hydrochloride |
| Verapamil 180mg modified-release capsules | 1511241000033113 | 36149411000001103 | Verapamil hydrochloride |
| Verapamil 240mg modified-release capsules | 1511341000033115 | 39021011000001106 | Verapamil hydrochloride |
| Verapamil 120mg tablets | 1512941000033111 | 318206008 | Verapamil hydrochloride |
| Verapamil 160mg tablets | 1513141000033119 | 318248001 | Verapamil hydrochloride |
| Verapamil 40mg tablets | 1513241000033114 | 318204006 | Verapamil hydrochloride |
| Verapamil 80mg tablets | 1513341000033116 | 318205007 | Verapamil hydrochloride |
| Vertab SR 240 tablets (Chiesi Ltd) | 1754841000033116 | 675611000001104 | Verapamil hydrochloride |
| Verapress MR 240mg tablets (Dexcel-Pharma Ltd) | 1833541000033117 | 525311000001101 | Verapamil hydrochloride |
| Cordilox MR 240mg tablets (Teva UK Ltd) | 2129041000033118 | 740811000001102 | Verapamil hydrochloride |
| Zolvera 40mg/5ml oral solution (Rosemont Pharmaceuticals Ltd) | 2641041000033116 | 4129511000001107 | Verapamil hydrochloride |
| Ranvera MR 240mg tablets (Ranbaxy (UK) Ltd) | 2845841000033119 | 3634111000001100 | Verapamil hydrochloride |
| Vera-Til SR 120mg tablets (Tillomed Laboratories Ltd) | 2890641000033116 | 7418511000001104 | Verapamil hydrochloride |
| Vera-Til SR 240mg tablets (Tillomed Laboratories Ltd) | 2890741000033113 | 7418711000001109 | Verapamil hydrochloride |
| Verapamil 180mg modified-release / Trandolapril 2mg capsules | 3163041000033116 | 36149211000001102 | Trandolapril/ Verapamil hydrochloride |
| Verapamil 5mg/2ml solution for injection ampoules | 4260341000033110 | 36149611000001100 | Verapamil hydrochloride |

#### Table S67. Angiotensin converting enzyme inhibitors Aurum product codes

| Term from EMIS | Prod code ID | dmd ID | Drug substance name |
|---|---|---|---|
| Ramipril 1.25mg capsules | 1151341000033117 | 318900007 | Ramipril |
| Ramipril 2.5mg capsules | 1151441000033111 | 318901006 | Ramipril |
| Ramipril 5mg capsules | 1151541000033112 | 318902004 | Ramipril |
| Tritace 1.25mg capsules (Aventis Pharma) | 1455541000033110 | 111611000001109 | Ramipril |
| Tritace 2.5mg capsules (Sanofi) | 1455641000033111 | 835411000001105 | Ramipril |
| Tritace 5mg capsules (Sanofi) | 1455741000033119 | 802311000001101 | Ramipril |
| Ramipril 10mg capsules | 1769841000033111 | 318906001 | Ramipril |
| Tritace 10mg capsules (Sanofi) | 1769941000033115 | 43711000001100 | Ramipril |
| Triapin 5mg/5mg modified-release tablets (Sanofi) | 1834341000033110 | 3887911000001109 | Felodipine/ Ramipril |
| Ramipril 5mg/5ml oral suspension | 2883341000033118 | 8720911000001100 | Ramipril |
| Ramipril 1.25mg tablets | 2989541000033115 | 408040007 | Ramipril |
| Ramipril 2.5mg tablets | 2989641000033119 | 408050008 | Ramipril |
| Ramipril 5mg tablets | 2989741000033111 | 408051007 | Ramipril |
| Ramipril 10mg tablets | 2989841000033118 | 408052000 | Ramipril |
| Tritace 1.25mg tablets (Sanofi) | 2989941000033114 | 5010511000001106 | Ramipril |
| Tritace 2.5mg tablets (Sanofi) | 2990041000033116 | 5010811000001109 | Ramipril |
| Tritace 5mg tablets (Sanofi) | 2990141000033117 | 5011111000001108 | Ramipril |
| Tritace 10mg tablets (Sanofi) | 2990241000033112 | 5011411000001103 | Ramipril |
| Lopace 2.5mg capsules (Discovery Pharmaceuticals) | 3159741000033111 | 7948711000001102 | Ramipril |
| Lopace 5mg capsules (Discovery Pharmaceuticals) | 3159841000033118 | 7948911000001100 | Ramipril |
| Lopace 10mg capsules (Discovery Pharmaceuticals) | 3159941000033114 | 7949111000001105 | Ramipril |
| Felodipine 5mg modified-release / Ramipril 5mg tablets | 3163241000033112 | 318177008 | Felodipine/ Ramipril |
| Felodipine 2.5mg modified-release / Ramipril 2.5mg tablets | 3163341000033119 | 318176004 | Felodipine/ Ramipril |
| Ramipril 1.25mg/5ml oral suspension | 4152241000033113 | 8720311000001101 | Ramipril |
| Ramipril 2.5mg/5ml oral suspension | 4805941000033114 | 8720711000001102 | Ramipril |
| Ramipril 10mg/5ml oral solution | 5887841000033118 | 8720411000001108 | Ramipril |
| Ramipril 10mg/5ml oral suspension | 5887941000033114 | 8720511000001107 | Ramipril |
| Ramipril 5mg/5ml oral solution | 5890341000033110 | 8720811000001105 | Ramipril |
| Ramipril 2.5mg/5ml oral solution | 5890541000033115 | 8720611000001106 | Ramipril |
| Ramipril 1.25mg/5ml oral solution | 5998241000033112 | 8720211000001109 | Ramipril |
| Ramipril 2.5mg/5ml oral solution sugar free | 6517041000033110 | 19877111000001100 | Ramipril |
| Triapin 2.5mg/2.5mg modified-release tablets (Sanofi) | 8264941000033119 | 4093211000001109 | Felodipine/ Ramipril |
| Enalapril 10mg tablets | 522641000033110 | 318853004 | Enalapril maleate |
| Enalapril 20mg tablets | 522741000033118 | 318855006 | Enalapril maleate |
| Enalapril 5mg tablets | 522841000033111 | 318851002 | Enalapril maleate |
| Enalapril 2.5mg tablets | 523541000033115 | 318850001 | Enalapril maleate |
| Innovace 10mg tablets (Organon Pharma (UK) Ltd) | 769041000033116 | 316111000001104 | Enalapril maleate |

| | | | |
|---|---|---|---|
| Innovace 2.5mg tablets (Organon Pharma (UK) Ltd) | 769141000033117 | 749611000001107 | Enalapril maleate |
| Innovace 20mg tablets (Organon Pharma (UK) Ltd) | 769241000033112 | 302311000001104 | Enalapril maleate |
| Innovace 5mg tablets (Organon Pharma (UK) Ltd) | 769341000033119 | 730211000001104 | Enalapril maleate |
| Innozide 20mg/12.5mg tablets (Organon Pharma (UK) Ltd) | 770141000033115 | 146811000001108 | Enalapril maleate/ Hydrochlorothiazide |
| Pralenal 2.5 tablets (Opus Pharmaceuticals Ltd) | 1918641000033112 | 448611000001106 | Enalapril maleate |
| Pralenal 5 tablets (Opus Pharmaceuticals Ltd) | 1918741000033115 | 578511000001100 | Enalapril maleate |
| Pralenal 10 tablets (Opus Pharmaceuticals Ltd) | 1918841000033113 | 761411000001101 | Enalapril maleate |
| Pralenal 20 tablets (Opus Pharmaceuticals Ltd) | 1918941000033117 | 508511000001105 | Enalapril maleate |
| Enalapril 20mg / Hydrochlorothiazide 12.5mg tablets | 3162241000033117 | 39691911000001109 | Enalapril maleate/ Hydrochlorothiazide |
| Enalapril 5mg/5ml oral suspension sugar free | 3279241000033117 | 20092911000001108 | Enalapril maleate |
| Enalapril 10mg/5ml oral solution | 5967041000033117 | 8485711000001108 | Enalapril maleate |
| Enalapril 10mg/5ml oral suspension | 5967141000033118 | 8485811000001100 | Enalapril maleate |
| Enalapril 5mg/5ml oral solution | 5992541000033111 | 8486911000001103 | Enalapril maleate |
| Enalapril 5mg/5ml oral suspension | 5992641000033112 | 8487011000001104 | Enalapril maleate |
| Enalapril 1.25mg/5ml oral solution | 6012441000033118 | 8485511000001103 | Enalapril maleate |
| Enalapril 1.25mg/5ml oral suspension | 6012541000033117 | 8485611000001104 | Enalapril maleate |
| Enalapril 20mg tablets (Imported) | 13740241000033119 | 38955411000001109 | Enalapril maleate |

#### Table S68. Angiotensin II receptor blocker Aurum product codes

| Term from EMIS | Prod code ID | dmd ID | Drug substance name |
|---|---|---|---|
| Azilsartan medoxomil 20mg tablets | 7754841000033119 | 20418711000001104 | Azilsartan medoxomil |
| Azilsartan medoxomil 40mg tablets | 7754941000033110 | 449109006 | Azilsartan medoxomil |
| Azilsartan medoxomil 80mg tablets | 7755041000033110 | 449333009 | Azilsartan medoxomil |
| Edarbi 20mg tablets (Takeda UK Ltd) | 7755141000033114 | 20350911000001100 | Azilsartan medoxomil |
| Edarbi 40mg tablets (Takeda UK Ltd) | 7755241000033119 | 20351211000001103 | Azilsartan medoxomil |
| Edarbi 80mg tablets (Takeda UK Ltd) | 7755341000033112 | 20351811000001102 | Azilsartan medoxomil |
| Amias 16mg tablets (Takeda UK Ltd) | 59141000033113 | 908511000001100 | Candesartan cilexetil |
| Amias 2mg tablets (Takeda UK Ltd) | 59241000033118 | 97311000001103 | Candesartan cilexetil |
| Amias 4mg tablets (Takeda UK Ltd) | 59341000033111 | 857411000001100 | Candesartan cilexetil |
| Amias 8mg tablets (Takeda UK Ltd) | 59441000033117 | 36011000001106 | Candesartan cilexetil |
| Candesartan 16mg tablets | 211541000033119 | 318980005 | Candesartan cilexetil |
| Candesartan 2mg tablets | 211641000033118 | 318977009 | Candesartan cilexetil |
| Candesartan 4mg tablets | 211741000033110 | 318978004 | Candesartan cilexetil |
| Candesartan 8mg tablets | 211841000033117 | 318979007 | Candesartan cilexetil |
| Candesartan 32mg tablets | 3227241000033111 | 376998003 | Candesartan cilexetil |
| Amias 32mg tablets (Takeda UK Ltd) | 3227341000033118 | 8983911000001107 | Candesartan cilexetil |
| Eprosartan 300mg tablets | 2036341000033115 | 318994006 | Eprosartan mesilate |
| Eprosartan 400mg tablets | 2036441000033114 | 318995007 | Eprosartan mesilate |
| Eprosartan 600mg tablets | 2036541000033110 | 318996008 | Eprosartan mesilate |
| Teveten 300mg tablets (Mylan) | 2040841000033113 | 401211000001105 | Eprosartan mesilate |
| Teveten 400mg tablets (Abbott Healthcare Products Ltd) | 2040941000033117 | 151411000001103 | Eprosartan mesilate |
| Teveten 600mg tablets (Mylan) | 2041041000033110 | 872011000001109 | Eprosartan mesilate |
| Aprovel 150mg tablets (Sanofi) | 75941000033114 | 859711000001103 | Irbesartan |
| Aprovel 300mg tablets (Sanofi) | 76041000033116 | 323211000001107 | Irbesartan |
| Aprovel 75mg tablets (Sanofi) | 76141000033117 | 434511000001104 | Irbesartan |
| Irbesartan 150mg tablets | 775441000033117 | 318969005 | Irbesartan |
| Irbesartan 300mg tablets | 775541000033116 | 318970006 | Irbesartan |
| Irbesartan 75mg tablets | 775641000033115 | 318968002 | Irbesartan |
| CoAprovel 150mg/12.5mg tablets (Sanofi) | 2216141000033115 | 792411000001108 | Hydrochlorothiazide/ Irbesartan |
| CoAprovel 300mg/12.5mg tablets (Sanofi) | 2216241000033110 | 682711000001109 | Hydrochlorothiazide/ Irbesartan |
| Irbesartan 150mg / Hydrochlorothiazide 12.5mg tablets | 3161141000033116 | 134461004 | Hydrochlorothiazide/ Irbesartan |
| Irbesartan 300mg / Hydrochlorothiazide 12.5mg tablets | 3161241000033111 | 134460003 | Hydrochlorothiazide/ Irbesartan |
| Irbesartan 300mg / Hydrochlorothiazide 25mg tablets | 3995241000033112 | 10970311000001105 | Hydrochlorothiazide/ Irbesartan |
| CoAprovel 300mg/25mg tablets (Sanofi) | 3995341000033119 | 10968611000001106 | Hydrochlorothiazide/ Irbesartan |
| Irbesartan 300mg/5ml oral suspension | 4954341000033111 | 12639511000001103 | Irbesartan |

| | | | |
|---|---|---|---|
| Irbesartan 150mg/5ml oral suspension | 5970641000033117 | 8580811000001101 | Irbesartan |
| Irbesartan 37.5mg oral powder sachets | 6435741000033117 | 19481911000001105 | Irbesartan |
| Ifirmasta 75mg tablets (Consilient Health Ltd) | 8554141000033116 | 22720311000001108 | Irbesartan |
| Ifirmasta 150mg tablets (Consilient Health Ltd) | 8554241000033111 | 22720611000001103 | Irbesartan |
| Ifirmasta 300mg tablets (Consilient Health Ltd) | 8554341000033118 | 22720811000001104 | Irbesartan |
| Cozaar 25mg tablets (Organon Pharma (UK) Ltd) | 370441000033112 | 266511000001104 | Losartan potassium |
| Cozaar 50mg tablets (Organon Pharma (UK) Ltd) | 370541000033113 | 53611000001106 | Losartan potassium |
| Cozaar-Comp 50mg/12.5mg tablets (Organon Pharma (UK) Ltd) | 370841000033110 | 255911000001105 | Hydrochlorothiazide/ Losartan potassium |
| Losartan 25mg tablets | 851841000033117 | 318955005 | Losartan potassium |
| Losartan 50mg tablets | 851941000033113 | 318956006 | Losartan potassium |
| Losartan 100mg tablets | 2720141000033115 | 407784004 | Losartan potassium |
| Cozaar 100mg tablets (Organon Pharma (UK) Ltd) | 2720241000033110 | 245811000001102 | Losartan potassium |
| Losartan 50mg / Hydrochlorothiazide 12.5mg tablets | 3161741000033117 | 318959004 | Hydrochlorothiazide/ Losartan potassium |
| Losartan 100mg / Hydrochlorothiazide 25mg tablets | 3346841000033117 | 395497004 | Hydrochlorothiazide/ Losartan potassium |
| Cozaar-Comp 100mg/25mg tablets (Organon Pharma (UK) Ltd) | 3346941000033113 | 9566911000001105 | Hydrochlorothiazide/ Losartan potassium |
| Losartan 100mg / Hydrochlorothiazide 12.5mg tablets | 4424741000033110 | 13112711000001103 | Hydrochlorothiazide/ Losartan potassium |
| Cozaar-Comp 100mg/12.5mg tablets (Organon Pharma (UK) Ltd) | 4424841000033117 | 13094111000001102 | Hydrochlorothiazide/ Losartan potassium |
| Losartan 12.5mg tablets | 4957541000033114 | 15148111000001100 | Losartan potassium |
| Cozaar 12.5mg tablets (Organon Pharma (UK) Ltd) | 4957641000033110 | 15138911000001101 | Losartan potassium |
| Losartan 2.5mg/ml oral suspension sugar free | 5149941000033118 | 15507411000001105 | Losartan potassium |
| Cozaar 2.5mg/ml oral suspension (Organon Pharma (UK) Ltd) | 5150041000033110 | 15506811000001105 | Losartan potassium |
| Losartan 100mg/5ml oral solution | 5971741000033116 | 15451111000001107 | Losartan potassium |
| Losartan 100mg/5ml oral suspension | 5971841000033114 | 15451211000001101 | Losartan potassium |
| Losartan 50mg/5ml oral solution | 5971941000033118 | 14159411000001106 | Losartan potassium |
| Losartan 50mg/5ml oral suspension | 5972041000033112 | 14159511000001105 | Losartan potassium |
| Olmesartan medoxomil 10mg tablets | 2944541000033111 | 408055003 | Olmesartan medoxomil |
| Olmesartan medoxomil 20mg tablets | 2944641000033112 | 385542009 | Olmesartan medoxomil |
| Olmesartan medoxomil 40mg tablets | 2944741000033115 | 385543004 | Olmesartan medoxomil |
| Olmetec 10mg tablets (Daiichi Sankyo UK Ltd) | 2944841000033113 | 4624011000001101 | Olmesartan medoxomil |
| Olmetec 20mg tablets (Daiichi Sankyo UK Ltd) | 2944941000033117 | 4624311000001103 | Olmesartan medoxomil |
| Olmetec 40mg tablets (Daiichi Sankyo UK Ltd) | 2945041000033117 | 4624611000001108 | Olmesartan medoxomil |
| Olmesartan medoxomil 20mg / Hydrochlorothiazide 12.5mg tablets | 3908941000033112 | 409184002 | Hydrochlorothiazide/ Olmesartan medoxomil |
| Olmesartan medoxomil 20mg / Hydrochlorothiazide 25mg tablets | 3909041000033115 | 10270711000001105 | Hydrochlorothiazide/ Olmesartan medoxomil |

| | | | |
|---|---|---|---|
| Olmetec Plus 20mg/12.5mg tablets (Daiichi Sankyo UK Ltd) | 3909141000033116 | 10261511000001103 | Hydrochlorothiazide/ Olmesartan medoxomil |
| Olmetec Plus 20mg/25mg tablets (Daiichi Sankyo UK Ltd) | 3909241000033111 | 10261811000001100 | Hydrochlorothiazide/ Olmesartan medoxomil |
| Olmesartan medoxomil 10mg/5ml oral suspension | 4273241000033118 | 14680711000001103 | Olmesartan medoxomil |
| Olmesartan medoxomil 20mg / Amlodipine 5mg tablets | 5005841000033114 | 429502004 | Amlodipine besilate/ Olmesartan medoxomil |
| Olmesartan medoxomil 40mg / Amlodipine 5mg tablets | 5005941000033118 | 429503009 | Amlodipine besilate/ Olmesartan medoxomil |
| Olmesartan medoxomil 40mg / Amlodipine 10mg tablets | 5006041000033111 | 429678006 | Amlodipine besilate/ Olmesartan medoxomil |
| Sevikar 20mg/5mg tablets (Daiichi Sankyo UK Ltd) | 5006141000033110 | 15773211000001105 | Amlodipine besilate/ Olmesartan medoxomil |
| Sevikar 40mg/5mg tablets (Daiichi Sankyo UK Ltd) | 5006241000033115 | 15772911000001108 | Amlodipine besilate/ Olmesartan medoxomil |
| Sevikar 40mg/10mg tablets (Daiichi Sankyo UK Ltd) | 5006341000033113 | 15772611000001102 | Amlodipine besilate/ Olmesartan medoxomil |
| Olmesartan medoxomil 40mg / Hydrochlorothiazide 12.5mg tablets | 5566341000033111 | 409185001 | Hydrochlorothiazide/ Olmesartan medoxomil |
| Olmetec Plus 40mg/12.5mg tablets (Daiichi Sankyo UK Ltd) | 5566441000033117 | 17220911000001102 | Hydrochlorothiazide/ Olmesartan medoxomil |
| Sevikar HCT 20mg/5mg/12.5mg tablets (Daiichi Sankyo UK Ltd) | 6180641000033112 | 18986411000001108 | Amlodipine besilate/ Hydrochlorothiazide/ Olmesartan medoxomil |
| Sevikar HCT 40mg/5mg/12.5mg tablets (Daiichi Sankyo UK Ltd) | 6180741000033115 | 18986711000001102 | Amlodipine besilate/ Hydrochlorothiazide/ Olmesartan medoxomil |
| Sevikar HCT 40mg/10mg/12.5mg tablets (Daiichi Sankyo UK Ltd) | 6180841000033113 | 18987011000001101 | Amlodipine besilate/ Hydrochlorothiazide/ Olmesartan medoxomil |
| Sevikar HCT 40mg/5mg/25mg tablets (Daiichi Sankyo UK Ltd) | 6180941000033117 | 18987311000001103 | Amlodipine besilate/ Hydrochlorothiazide/ Olmesartan medoxomil |
| Sevikar HCT 40mg/10mg/25mg tablets (Daiichi Sankyo UK Ltd) | 6181041000033110 | 18987611000001108 | Amlodipine besilate/ Hydrochlorothiazide/ Olmesartan medoxomil |
| Diovan 160mg capsules (Novartis Pharmaceuticals UK Ltd) | 437641000033118 | 117011000001107 | Valsartan |
| Diovan 40mg capsules (Novartis Pharmaceuticals UK Ltd) | 437741000033110 | 777611000001101 | Valsartan |
| Diovan 80mg capsules (Novartis Pharmaceuticals UK Ltd) | 437841000033117 | 554511000001105 | Valsartan |
| Valsartan 160mg capsules | 1498241000033111 | 318963006 | Valsartan |
| Valsartan 40mg capsules | 1498341000033118 | 318961008 | Valsartan |
| Valsartan 80mg capsules | 1498441000033112 | 318962001 | Valsartan |

| | | | |
|---|---|---|---|
| Co-Diovan 160mg/12.5mg tablets (Novartis Pharmaceuticals UK Ltd) | 3152541000033112 | 7668611000001104 | Hydrochlorothiazide/ Valsartan |
| Co-Diovan 160mg/25mg tablets (Novartis Pharmaceuticals UK Ltd) | 3152641000033113 | 7668911000001105 | Hydrochlorothiazide/ Valsartan |
| Valsartan 160mg / Hydrochlorothiazide 12.5mg tablets | 3161441000033112 | 395521005 | Hydrochlorothiazide/ Valsartan |
| Valsartan 160mg / Hydrochlorothiazide 25mg tablets | 3161541000033113 | 409298002 | Hydrochlorothiazide/ Valsartan |
| Valsartan 80mg / Hydrochlorothiazide 12.5mg tablets | 3190741000033112 | 377488008 | Hydrochlorothiazide/ Valsartan |
| Co-Diovan 80mg/12.5mg tablets (Novartis Pharmaceuticals UK Ltd) | 3190841000033119 | 8150111000001108 | Hydrochlorothiazide/ Valsartan |
| Valsartan 40mg tablets | 3201341000033115 | 416515008 | Valsartan |
| Diovan 40mg tablets (Novartis Pharmaceuticals UK Ltd) | 3201441000033114 | 8263211000001101 | Valsartan |
| Amlodipine 5mg / Valsartan 80mg tablets | 4021441000033114 | 11160311000001109 | Amlodipine besilate/ Valsartan |
| Amlodipine 5mg / Valsartan 160mg tablets | 4021541000033110 | 11160211000001101 | Amlodipine besilate/ Valsartan |
| Amlodipine 10mg / Valsartan 160mg tablets | 4021641000033111 | 11160111000001107 | Amlodipine besilate/ Valsartan |
| Exforge 5mg/80mg tablets (Novartis Pharmaceuticals UK Ltd) | 4021741000033119 | 11161811000001108 | Amlodipine besilate/ Valsartan |
| Exforge 5mg/160mg tablets (Novartis Pharmaceuticals UK Ltd) | 4021841000033112 | 11161511000001105 | Amlodipine besilate/ Valsartan |
| Exforge 10mg/160mg tablets (Novartis Pharmaceuticals UK Ltd) | 4021941000033116 | 11160711000001108 | Amlodipine besilate/ Valsartan |
| Valsartan 320mg tablets | 4424941000033113 | 376487009 | Valsartan |
| Diovan 320mg tablets (Novartis Pharmaceuticals UK Ltd) | 4425041000033113 | 13143311000001102 | Valsartan |
| Valsartan 80mg tablets | 6515441000033113 | 375034009 | Valsartan |
| Valsartan 160mg tablets | 6515541000033114 | 375035005 | Valsartan |
| Valsartan 3mg/ml oral solution | 6528441000033113 | 20007411000001100 | Valsartan |
| Diovan 3mg/1ml oral solution (Novartis Pharmaceuticals UK Ltd) | 6528541000033114 | 20001711000001102 | Valsartan |
| Sacubitril 24mg / Valsartan 26mg tablets | 10943641000033117 | 31142011000001103 | Sacubitril/ Valsartan |
| Sacubitril 49mg / Valsartan 51mg tablets | 10943741000033114 | 31142111000001102 | Sacubitril/ Valsartan |
| Sacubitril 97mg / Valsartan 103mg tablets | 10943841000033116 | 31142211000001108 | Sacubitril/ Valsartan |
| Entresto 24mg/26mg tablets (Novartis Pharmaceuticals UK Ltd) | 10943941000033112 | 31136811000001109 | Sacubitril/ Valsartan |
| Entresto 49mg/51mg tablets (Novartis Pharmaceuticals UK Ltd) | 10944041000033114 | 31136411000001107 | Sacubitril/ Valsartan |
| Entresto 97mg/103mg tablets (Novartis Pharmaceuticals UK Ltd) | 10944141000033113 | 31138011000001104 | Sacubitril/ Valsartan |

#### Table S69. Beta blockers Aurum product codes

| Term from EMIS | Prod code ID | dmd ID | Drug substance name |
|---|---|---|---|
| Sotalol 25mg/5ml oral solution | 3953441000033116 | 8726811000001107 | Sotalol hydrochloride |
| Sotalol 25mg/5ml oral suspension | 9776441000033119 | 8726911000001102 | Sotalol hydrochloride |
| Acebutolol 100mg capsules | 3041000033115 | 318412000 | Acebutolol hydrochloride |
| Sectral 100mg capsules (Sanofi) | 1268141000033119 | 632811000001105 | Acebutolol hydrochloride |
| Sectral 200mg capsules (Sanofi) | 1268241000033114 | 925711000001108 | Acebutolol hydrochloride |
| Acebutolol 200mg capsules | 3141000033116 | 318413005 | Acebutolol hydrochloride |
| Acebutolol 400mg tablets | 10741000033112 | 318414004 | Acebutolol hydrochloride |
| Sectral 400mg tablets (Sanofi) | 1278241000033118 | 298111000001105 | Acebutolol hydrochloride |
| Atenix 50 tablets (Ashbourne Pharmaceuticals Ltd) | 92541000033118 | 271911000001106 | Atenolol |
| Tenormin LS 50mg tablets (AstraZeneca UK Ltd) | 1426541000033115 | 423911000001107 | Atenolol |
| Antipressan 50mg tablets (Teva UK Ltd) | 71041000033110 | 874711000001101 | Atenolol |
| Atenolol 50mg tablets | 91141000033119 | 318420003 | Atenolol |
| Atenolol 100mg tablets | 91041000033118 | 318421004 | Atenolol |
| Antipressan 100mg tablets (Teva UK Ltd) | 70941000033117 | 734011000001102 | Atenolol |
| Atenix 100 tablets (Ashbourne Pharmaceuticals Ltd) | 90541000033117 | 181611000001107 | Atenolol |
| Tenormin 100mg tablets (AstraZeneca UK Ltd) | 1426441000033116 | 162411000001102 | Atenolol |
| Tenormin 25mg tablets (AstraZeneca UK Ltd) | 1428741000033112 | 317111000001101 | Atenolol |
| Atenix 25 tablets (Ashbourne Pharmaceuticals Ltd) | 91841000033113 | 482511000001101 | Atenolol |
| Antipressan 25mg tablets (Teva UK Ltd) | 69441000033112 | 877011000001106 | Atenolol |
| Atenolol 25mg tablets | 90641000033116 | 318434003 | Atenolol |
| Betaloc 100mg tablets (AstraZeneca UK Ltd) | 144341000033117 | 193911000001101 | Metoprolol tartrate |
| Lopresor 100mg tablets (Recordati Pharmaceuticals Ltd) | 850841000033110 | 756911000001107 | Metoprolol tartrate |
| Metoprolol 100mg tablets | 897741000033111 | 318474009 | Metoprolol tartrate |
| Metoprolol 50mg tablets | 897841000033118 | 318475005 | Metoprolol tartrate |
| Betaloc 50mg tablets (AstraZeneca UK Ltd) | 144441000033111 | 31811000001105 | Metoprolol tartrate |
| Lopresor 50mg tablets (Recordati Pharmaceuticals Ltd) | 850941000033119 | 113511000001109 | Metoprolol tartrate |
| Corgard 40mg tablets (Sanofi-Synthelabo Ltd) | 375241000033119 | 3686511000001108 | Nadolol |
| Nadolol 40mg tablets | 956341000033113 | 318480001 | Nadolol |
| Nadolol 80mg tablets | 956541000033118 | 318481002 | Nadolol |
| Corgard 80mg tablets (Sanofi) | 375341000033112 | 3687411000001106 | Nadolol |
| Trasicor 20mg tablets (Amdipharm Plc) | 1469641000033119 | 3379011000001106 | Oxprenolol hydrochloride |
| Oxprenolol 20mg tablets | 1026141000033119 | 318483004 | Oxprenolol hydrochloride |
| Oxprenolol 40mg tablets | 1026241000033114 | 318484005 | Oxprenolol hydrochloride |
| Trasicor 40mg tablets (Amdipharm Plc) | 1469741000033111 | 592911000001104 | Oxprenolol hydrochloride |

| | | | |
|---|---|---|---|
| Trasicor 80mg tablets (Amdipharm Plc) | 1469841000033118 | 568511000001106 | Oxprenolol hydrochloride |
| Oxprenolol 80mg tablets | 1026341000033116 | 318485006 | Oxprenolol hydrochloride |
| Pindolol 5mg tablets | 1091241000033112 | 318512002 | Pindolol |
| Visken 5mg tablets (Advanz Pharma) | 1524041000033111 | 3706111000001105 | Pindolol |
| Visken 15mg tablets (Advanz Pharma) | 1525141000033111 | 3887411000001101 | Pindolol |
| Pindolol 15mg tablets | 1091141000033117 | 318513007 | Pindolol |
| Sotalol 40mg tablets | 1367441000033117 | 318525005 | Sotalol hydrochloride |
| Beta-Cardone 40mg tablets (Focus Pharmaceuticals Ltd) | 144041000033119 | 900511000001103 | Sotalol hydrochloride |
| Sotacor 80mg tablets (Cheplapharm Arzneimittel GmbH) | 1366141000033111 | 463811000001109 | Sotalol hydrochloride |
| Beta-Cardone 80mg tablets (Focus Pharmaceuticals Ltd) | 144141000033115 | 104011000001101 | Sotalol hydrochloride |
| Sotalol 80mg tablets | 1367541000033116 | 318526006 | Sotalol hydrochloride |
| Sotalol 200mg tablets | 1367241000033118 | 318527002 | Sotalol hydrochloride |
| Beta-Cardone 200mg tablets (Advanz Pharma) | 143941000033116 | 447111000001104 | Sotalol hydrochloride |
| Sotacor 160mg tablets (Bristol-Myers Squibb Pharmaceuticals Ltd) | 1366041000033112 | 446211000001108 | Sotalol hydrochloride |
| Sotalol 160mg tablets | 1367141000033113 | 318528007 | Sotalol hydrochloride |
| Timolol 10mg tablets | 1444541000033113 | 318534000 | Timolol maleate |
| Betim 10mg tablets (Meda Pharmaceuticals Ltd) | 144741000033116 | 72811000001104 | Timolol maleate |
| Co-Betaloc tablets (Pfizer Ltd) | 372741000033116 | 2977611000001106 | Hydrochlorothiazide/ Metoprolol tartrate |
| Metoprolol 100mg / Hydrochlorothiazide 12.5mg tablets | 3161341000033118 | 318546001 | Hydrochlorothiazide/ Metoprolol tartrate |
| Corgaretic 40mg tablets (Sanofi-Synthelabo Ltd) | 375441000033118 | 3886211000001102 | Bendroflumethiazide/ Nadolol |
| Corgaretic 80mg tablets (Sanofi-Synthelabo Ltd) | 375541000033117 | 4057911000001102 | Bendroflumethiazide/ Nadolol |
| Viskaldix tablets (Advanz Pharma) | 1523941000033114 | 3638411000001101 | Clopamide/ Pindolol |
| Pindolol 10mg / Clopamide 5mg tablets | 3163141000033117 | 318552000 | Clopamide/ Pindolol |
| Moducren tablets (Merck Sharp & Dohme Ltd) | 935341000033118 | 74311000001101 | Amiloride hydrochloride/ Hydrochlorothiazide/ Timolol maleate |
| Prestim tablets (Meda Pharmaceuticals Ltd) | 1132741000033111 | 98411000001109 | Bendroflumethiazide/ Timolol maleate |
| Timolol 10mg / Bendroflumethiazide 2.5mg tablets | 3346641000033118 | 318556002 | Bendroflumethiazide/ Timolol maleate |
| Tenben 25mg/1.25mg capsules (Galen Ltd) | 1414241000033117 | 721411000001106 | Atenolol/ Bendroflumethiazide |
| Tenoret 50mg/12.5mg tablets (AstraZeneca UK Ltd) | 1426241000033117 | 825011000001103 | Atenolol/ Chlortalidone |
| Tenchlor 50mg/12.5mg tablets (Teva UK Ltd) | 1425441000033113 | 439411000001104 | Atenolol/ Chlortalidone |
| Atenix Co 50 tablets (Ashbourne Pharmaceuticals Ltd) | 92441000033119 | 629011000001109 | Atenolol/ Chlortalidone |
| Co-tenidone 50mg/12.5mg tablets | 376441000033110 | 318575007 | Atenolol/ Chlortalidone |
| Secadrex 200mg/12.5mg tablets (Sanofi) | 1274641000033111 | 878911000001106 | Acebutolol hydrochloride/ Hydrochlorothiazide |
| Acebutolol 200mg / Hydrochlorothiazide 12.5mg tablets | 3162941000033114 | 318586009 | Acebutolol hydrochloride/ Hydrochlorothiazide |

| | | | |
|---|---|---|---|
| Kalten capsules (M & A Pharmachem Ltd) | 793741000033117 | 237011000001100 | Amiloride hydrochloride/ Atenolol/ Hydrochlorothiazide |
| Celectol 200mg tablets (Cheplapharm Arzneimittel GmbH) | 236141000033110 | 864811000001100 | Celiprolol hydrochloride |
| Celiprolol 200mg tablets | 236041000033111 | 318619000 | Celiprolol hydrochloride |
| Celiprolol 400mg tablets | 233341000033116 | 318622003 | Celiprolol hydrochloride |
| Celectol 400mg tablets (Cheplapharm Arzneimittel GmbH) | 233241000033114 | 479211000001100 | Celiprolol hydrochloride |
| Eucardic 12.5mg tablets (Roche Products Ltd) | 556941000033118 | 334711000001101 | Carvedilol |
| Carvedilol 12.5mg tablets | 209941000033111 | 318631003 | Carvedilol |
| Carvedilol 25mg tablets | 210041000033115 | 318632005 | Carvedilol |
| Eucardic 25mg tablets (Roche Products Ltd) | 557041000033117 | 709911000001107 | Carvedilol |
| Eucardic 3.125mg tablets (Roche Products Ltd) | 1608941000033115 | 873111000001104 | Carvedilol |
| Carvedilol 3.125mg tablets | 1581241000033110 | 318633000 | Carvedilol |
| Carvedilol 6.25mg tablets | 1581341000033117 | 318635007 | Carvedilol |
| Eucardic 6.25mg tablets (Roche Products Ltd) | 1609041000033112 | 690511000001101 | Carvedilol |
| Nadolol 40mg/5ml oral suspension | 5378241000033112 | 12300811000001107 | Nadolol |
| Tenormin 25mg/5ml syrup (AstraZeneca UK Ltd) | 1424541000033110 | 373311000001100 | Atenolol |
| Atenolol 25mg/5ml oral solution sugar free | 90441000033118 | 35903211000001107 | Atenolol |
| Tenif 50mg/20mg modified-release capsules (AstraZeneca UK Ltd) | 1414941000033114 | 3142511000001102 | Atenolol/ Nifedipine |
| Beta-Adalat modified-release capsules (Bayer Plc) | 124341000033119 | 3142711000001107 | Atenolol/ Nifedipine |
| Atenolol 50mg / Nifedipine 20mg modified-release capsules | 3246941000033118 | 35903311000001104 | Atenolol/ Nifedipine |
| Tenormin 5mg/10ml solution for injection ampoules (AstraZeneca UK Ltd) | 1420741000033114 | 9111000001107 | Atenolol |
| Atenolol 5mg/10ml solution for injection ampoules | 89541000033111 | 35903411000001106 | Atenolol |
| Metoprolol 5mg/5ml solution for injection ampoules | 887141000033113 | 3631211000001109 | Metoprolol tartrate |
| Betaloc I.V. 5mg/5ml solution for injection ampoules (Recordati Pharmaceuticals Ltd) | 131141000033116 | 3615711000001109 | Metoprolol tartrate |
| Sotacor 40mg/4ml solution for injection ampoules (Bristol-Myers Squibb Pharmaceuticals Ltd) | 1358541000033113 | 694711000001100 | Sotalol hydrochloride |
| Sotalol 40mg/4ml solution for injection ampoules | 2908541000033114 | 35930811000001109 | Sotalol hydrochloride |
| Slow-Trasicor 160mg tablets (Advanz Pharma) | 1347441000033114 | 473011000001107 | Oxprenolol hydrochloride |
| Oxprenolol 160mg modified-release tablets | 1026041000033118 | 36023011000001102 | Oxprenolol hydrochloride |
| Co-Betaloc SA tablets (Pfizer Ltd) | 372841000033114 | 3853411000001104 | Hydrochlorothiazide/ Metoprolol tartrate |
| Lopresor SR 200mg tablets (Recordati Pharmaceuticals Ltd) | 851041000033112 | 916111000001108 | Metoprolol tartrate |
| Betaloc-SA 200mg tablets (AstraZeneca UK Ltd) | 144541000033112 | 185111000001102 | Metoprolol tartrate |
| Metoprolol 200mg modified-release tablets | 891641000033111 | 36035411000001108 | Metoprolol tartrate |
| Nadolol 30mg/5ml oral solution | 7871341000033114 | 16109911000001102 | Nadolol |
| Nadolol 30mg/5ml oral suspension | 10650141000033117 | 16110011000001107 | Nadolol |

| | | | |
|---|---|---|---|
| Nadolol 20mg/5ml oral solution | 5814141000033119 | 16130911000001101 | Nadolol |
| Nadolol 20mg/5ml oral suspension | 10650041000033116 | 16131011000001109 | Nadolol |
| Timolol 10mg / Amiloride 2.5mg / Hydrochlorothiazide 25mg tablets (Essential Generics Ltd) | 5376441000033114 | 16458411000001107 | Amiloride hydrochloride/ Hydrochlorothiazide/ Timolol maleate |
| Co-tenidone 100mg/25mg tablets | 376541000033111 | 377211005 | Atenolol/ Chlortalidone |
| Atenix Co 100 tablets (Ashbourne Pharmaceuticals Ltd) | 90741000033113 | 101411000001108 | Atenolol/ Chlortalidone |
| Tenoretic 100mg/25mg tablets (AstraZeneca UK Ltd) | 1426341000033110 | 288711000001105 | Atenolol/ Chlortalidone |
| Tenchlor 100mg/25mg tablets (Teva UK Ltd) | 1425541000033114 | 298311000001107 | Atenolol/ Chlortalidone |
| Trasidrex modified-release tablets (Mercury Pharma Group Ltd) | 1469941000033114 | 3444411000001107 | Cyclopenthiazide/ Oxprenolol hydrochloride |
| Co-prenozide 160mg/0.25mg modified-release tablets | 377341000033119 | 36091611000001101 | Cyclopenthiazide/ Oxprenolol hydrochloride |
| Carvedilol 5mg/5ml oral suspension | 5288741000033116 | 8356811000001107 | Carvedilol |
| Metoprolol 12.5mg/5ml oral solution | 2616041000033116 | 8668211000001107 | Metoprolol tartrate |
| Metoprolol 12.5mg/5ml oral suspension | 4815641000033117 | 8668411000001106 | Metoprolol tartrate |
| Metoprolol 25mg/5ml oral solution | 5575541000033119 | 8668711000001100 | Metoprolol tartrate |
| Metoprolol 50mg/5ml oral solution | 5972841000033117 | 8668911000001103 | Metoprolol tartrate |
| Metoprolol 50mg/5ml oral suspension | 5972941000033113 | 8669111000001108 | Metoprolol tartrate |

#### Table S70. Alpha blockers Aurum product codes

| Term from EMIS | Prod code ID | dmd ID | Drug substance name |
|---|---|---|---|
| Cardura 1mg tablets (Upjohn UK Ltd) | 216741000033115 | 907711000001109 | Doxazosin mesilate |
| Cardura 2mg tablets (Upjohn UK Ltd) | 216841000033113 | 41811000001109 | Doxazosin mesilate |
| Doxazosin 1mg tablets | 480641000033111 | 318781001 | Doxazosin mesilate |
| Doxazosin 2mg tablets | 480741000033119 | 318782008 | Doxazosin mesilate |
| Doxazosin 4mg tablets | 480841000033112 | 318783003 | Doxazosin mesilate |
| Doxazosin 4mg modified-release tablets | 2273841000033111 | 39020411000001106 | Doxazosin mesilate |
| Doxazosin 8mg modified-release tablets | 2273941000033115 | 39021111000001107 | Doxazosin mesilate |
| Cardura XL 4mg tablets (Upjohn UK Ltd) | 2274041000033118 | 123911000001106 | Doxazosin mesilate |
| Cardura XL 8mg tablets (Upjohn UK Ltd) | 2274141000033119 | 873411000001109 | Doxazosin mesilate |
| Cascor 2mg tablets (Ranbaxy (UK) Ltd) | 2760041000033110 | 904511000001107 | Doxazosin mesilate |
| Cascor 4mg tablets (Ranbaxy (UK) Ltd) | 2760141000033114 | 179311000001107 | Doxazosin mesilate |
| Doxadura 1mg tablets (Dexcel-Pharma Ltd) | 2957941000033110 | 4857511000001108 | Doxazosin mesilate |
| Doxadura 2mg tablets (Dexcel-Pharma Ltd) | 2958041000033113 | 4857711000001103 | Doxazosin mesilate |
| Doxadura 4mg tablets (Dexcel-Pharma Ltd) | 2958141000033112 | 4858111000001103 | Doxazosin mesilate |
| Slocinx XL 4mg tablets (Zentiva Pharma UK Ltd) | 4021141000033118 | 11098311000001104 | Doxazosin mesilate |
| Doxadura XL 4mg tablets (Dexcel-Pharma Ltd) | 4063741000033112 | 11269911000001101 | Doxazosin mesilate |
| Oxandosin XL 4mg tablets (Ratiopharm UK Ltd) | 4288341000033111 | 11812211000001105 | Doxazosin mesilate |
| Doxazosin 1mg/5ml oral suspension | 4432941000033119 | 8483411000001101 | Doxazosin mesilate |
| Doxzogen XL 4mg tablets (Mylan) | 4575941000033115 | 13811611000001108 | Doxazosin mesilate |
| Cardozin XL 4mg tablets (Almus Pharmaceuticals Ltd) | 4936841000033112 | 18197411000001107 | Doxazosin mesilate |
| Doxazosin 4mg/5ml oral suspension | 5138341000033111 | 8483511000001102 | Doxazosin mesilate |
| Colixil XL 4mg tablets (Sandoz Ltd) | 5403641000033117 | 11757711000001107 | Doxazosin mesilate |
| Larbex XL 4mg tablets (Teva UK Ltd) | 5809641000033111 | 17338211000001104 | Doxazosin mesilate |
| Raporsin XL 4mg tablets (Accord Healthcare Ltd) | 6036141000033119 | 18164911000001108 | Doxazosin mesilate |
| Doxazosin 4mg/5ml oral solution | 8870641000033119 | 23466611000001105 | Doxazosin mesilate |
| Doxazosin 1mg/5ml oral solution | 9173941000033111 | 24509711000001103 | Doxazosin mesilate |
| Doxazosin 8mg tablets | 12682141000033115 | 421069003 | Doxazosin mesilate |
| Rogitine 10mg/1ml solution for injection ampoules (Alliance Pharmaceuticals Ltd) | 1180941000033115 | 4078811000001109 | Phentolamine mesilate |
| Invicorp 1 solution for injection (Senetek Plc) | 1619941000033111 | 5048711000001108 | Aviptadil/ Phentolamine mesilate |

| | | | |
|---|---|---|---|
| Invicorp 2 solution for injection (Senetek Plc) | 1620041000033114 | 5049011000001101 | Aviptadil/ Phentolamine mesilate |
| Phentolamine 10mg/1ml solution for injection ampoules | 3108541000033116 | 36015411000001100 | Phentolamine mesilate |
| Phentolamine 2mg/0.35ml / Aviptadil 25micrograms/0.35ml solution for injection ampoules | 10617641000033119 | 5049911000001102 | Aviptadil/ Phentolamine mesilate |
| Invicorp 25micrograms/2mg/0.35ml solution for injection ampoules (Evolan Pharma AB) | 10617741000033111 | 30737111000001103 | Aviptadil/ Phentolamine mesilate |
| Baratol 25mg tablets (Amdipharm Plc) | 116741000033118 | 3689111000001107 | Indoramin hydrochloride |
| Doralese Tiltab 20mg tablets (Chemidex Pharma Ltd) | 480541000033110 | 3354611000001100 | Indoramin hydrochloride |
| Indoramin 20mg tablets | 768541000033119 | 39696511000001106 | Indoramin hydrochloride |
| Indoramin 25mg tablets | 769841000033111 | 39696711000001101 | Indoramin hydrochloride |
| Dibenyline 10mg capsules (Mercury Pharma Group Ltd) | 438441000033119 | 3895211000001109 | Phenoxybenzamine hydrochloride |
| Phenoxybenzamine 10mg capsules | 1070341000033119 | 318745004 | Phenoxybenzamine hydrochloride |
| Phenoxybenzamine 25mg/5ml oral suspension | 2757941000033118 | 8671511000001109 | Phenoxybenzamine hydrochloride |
| Alphavase 1 tablets (Ashbourne Pharmaceuticals Ltd) | 37741000033118 | 934411000001100 | Prazosin hydrochloride |
| Alphavase 2 tablets (Ashbourne Pharmaceuticals Ltd) | 37841000033111 | 570011000001106 | Prazosin hydrochloride |
| Alphavase 5 tablets (Ashbourne Pharmaceuticals Ltd) | 37941000033115 | 74711000001102 | Prazosin hydrochloride |
| Hypovase 500microgram tablets (Pfizer Ltd) | 743541000033118 | 321311000001100 | Prazosin hydrochloride |
| Hypovase 1mg tablets (Pfizer Ltd) | 743641000033117 | 347411000001101 | Prazosin hydrochloride |
| Hypovase 2mg tablets (Pfizer Ltd) | 743741000033114 | 150911000001104 | Prazosin hydrochloride |
| Prazosin 500microgram tablets | 1131341000033118 | 318767003 | Prazosin hydrochloride |
| Prazosin 1mg tablets | 1131441000033112 | 318768008 | Prazosin hydrochloride |
| Prazosin 2mg tablets | 1131541000033113 | 318769000 | Prazosin hydrochloride |
| Prazosin 5mg tablets | 1131641000033114 | 318770004 | Prazosin hydrochloride |
| Prazosin 500micrograms/5ml oral solution | 11567441000033119 | 13078011000001104 | Prazosin hydrochloride |
| Hytrin 10mg tablets (Advanz Pharma) | 745641000033116 | 3150911000001107 | Terazosin hydrochloride |
| Hytrin 2mg tablets (Advanz Pharma) | 745741000033113 | 3147611000001100 | Terazosin hydrochloride |
| Hytrin 5mg tablets (Advanz Pharma) | 745841000033115 | 3154311000001103 | Terazosin hydrochloride |
| Terazosin 2mg tablets | 1425341000033119 | 318779003 | Terazosin hydrochloride |
| Terazosin 10mg tablets | 1429041000033118 | 318777001 | Terazosin hydrochloride |
| Terazosin 5mg tablets | 1429141000033119 | 318776005 | Terazosin hydrochloride |
| Benph 5mg tablets (Mylan) | 11807841000033119 | 33628611000001102 | Terazosin hydrochloride |

#### Table S71. Aldosterone Aurum product codes

| Term from EMIS | Prod code ID | dmd ID | Drug substance name |
|---|---|---|---|
| Spironolactone 50mg/5ml oral suspension | 1371341000033114 | 8726411000001105 | Spironolactone |
| Spironolactone 25mg/5ml oral suspension | 1371041000033112 | 8726511000001109 | Spironolactone |
| Spironolactone 15mg/5ml oral suspension | 5975441000033113 | 8727011000001103 | Spironolactone |
| Spironolactone 10mg/5ml oral suspension | 1371241000033116 | 8727311000001100 | Spironolactone |
| Spironolactone 100mg/5ml oral suspension | 2011541000033114 | 8727611000001105 | Spironolactone |
| Spironolactone 25mg tablets | 1374241000033118 | 318056008 | Spironolactone |
| Aldactone 25mg tablets (Pfizer Ltd) | 40541000033112 | 930511000001105 | Spironolactone |
| Aldactone 50mg tablets (Pfizer Ltd) | 40641000033113 | 921811000001103 | Spironolactone |
| Spironolactone 50mg tablets | 1374341000033111 | 318057004 | Spironolactone |
| Spironolactone 100mg tablets | 1374141000033113 | 318058009 | Spironolactone |
| Spirospare 100 tablets (Ashbourne Pharmaceuticals Ltd) | 1374541000033116 | 3411000001104 | Spironolactone |
| Aldactone 100mg tablets (Pfizer Ltd) | 40441000033111 | 421611000001100 | Spironolactone |
| Lasilactone 20mg/50mg capsules (Sanofi) | 808841000033119 | 3645811000001107 | Furosemide/ Spironolactone |
| Spironolactone 50mg / Furosemide 20mg capsules | 3162541000033115 | 318102003 | Furosemide/ Spironolactone |
| Aldactide 25 tablets (Pfizer Ltd) | 40241000033110 | 762511000001105 | Hydroflumethiazide/ Spironolactone |
| Co-flumactone 25mg/25mg tablets | 377041000033116 | 318127005 | Hydroflumethiazide/ Spironolactone |
| Co-flumactone 50mg/50mg tablets | 377141000033117 | 318128000 | Hydroflumethiazide/ Spironolactone |
| Aldactide 50 tablets (Pfizer Ltd) | 40341000033117 | 4669111000001107 | Hydroflumethiazide/ Spironolactone |
| Spironolactone 250mg/5ml oral solution | 5975541000033114 | 13353511000001101 | Spironolactone |
| Spironolactone 250mg/5ml oral suspension | 5975641000033110 | 13353611000001102 | Spironolactone |
| Spironolactone 100mg/5ml oral solution | 5999141000033111 | 13894611000001109 | Spironolactone |
| Spironolactone 10mg/5ml oral solution | 5999241000033116 | 13894711000001100 | Spironolactone |
| Spironolactone 25mg/5ml oral solution | 5890141000033112 | 13894811000001108 | Spironolactone |
| Spironolactone 50mg/5ml oral solution | 5890441000033116 | 13894911000001103 | Spironolactone |
| Spironolactone 5mg/5ml oral solution | 5999341000033114 | 13895011000001103 | Spironolactone |
| Spironolactone 3mg / Chlorothiazide 30mg capsules | 9278541000033112 | 16072811000001103 | Chlorothiazide/ Spironolactone |
| Inspra 25mg tablets (Upjohn UK Ltd) | 3199041000033116 | 8477211000001104 | Eplerenone |
| Eplerenone 25mg tablets | 3198841000033117 | 407011007 | Eplerenone |
| Eplerenone 50mg tablets | 3198941000033113 | 407012000 | Eplerenone |
| Inspra 50mg tablets (Upjohn UK Ltd) | 3199141000033117 | 8479811000001106 | Eplerenone |
| Spironolactone 4mg / Chlorothiazide 40mg capsules | 7681541000033117 | 18520511000001107 | Chlorothiazide/ Spironolactone |
| Spironolactone 5mg/5ml oral suspension | 1371141000033111 | 8726311000001103 | Spironolactone |

#### Table S72. Other antihypertensive Aurum product codes

| Term from EMIS | Prod code ID | dmd ID | Drug substance name |
|---|---|---|---|
| Apraclonidine 1% eye drops 0.25ml unit dose preservative free | 73541000033116 | 3958011000001107 | Apraclonidine hydrochloride |
| Apraclonidine 5mg/ml eye drops | 73641000033115 | 330873002 | Apraclonidine hydrochloride |
| Catapres 150micrograms/1ml solution for injection ampoules (Glenwood GmbH) | 196241000033111 | 364911000001108 | Clonidine hydrochloride |
| Catapres 100microgram tablets (Glenwood GmbH) | 215341000033112 | 215111000001101 | Clonidine hydrochloride |
| Catapres 300microgram tablets (Boehringer Ingelheim Ltd) | 215441000033118 | 368711000001103 | Clonidine hydrochloride |
| Clonidine 250microgram modified-release capsules | 261841000033118 | 36089511000001100 | Clonidine hydrochloride |
| Clonidine 150micrograms/1ml solution for injection ampoules | 275641000033110 | 36089211000001103 | Clonidine hydrochloride |
| Clonidine 100microgram tablets | 285841000033118 | 318667005 | Clonidine hydrochloride |
| Clonidine 25microgram tablets | 285941000033114 | 322840006 | Clonidine hydrochloride |
| Clonidine 300microgram tablets | 287241000033110 | 318668000 | Clonidine hydrochloride |
| Dixarit 25microgram tablets (Boehringer Ingelheim Ltd) | 464341000033111 | 344511000001105 | Clonidine hydrochloride |
| Iopidine 1% eye drops 0.25ml unit dose (Novartis Pharmaceuticals UK Ltd) | 771741000033114 | 3956211000001103 | Apraclonidine hydrochloride |
| Iopidine 5mg/ml eye drops (Novartis Pharmaceuticals UK Ltd) | 771841000033116 | 3799011000001100 | Apraclonidine hydrochloride |
| Clonidine 200micrograms/24hours transdermal patches | 5289041000033110 | 7660211000001104 | Clonidine |
| Clonidine 50micrograms/5ml oral solution | 5900141000033118 | 8398511000001100 | Clonidine hydrochloride |
| Clonidine 50micrograms/5ml oral suspension | 5900241000033113 | 8398611000001101 | Clonidine hydrochloride |
| Clonidine 100micrograms/24hours transdermal patches | 12106941000033117 | 10448511000001104 | Clonidine |
| Clonidine 300micrograms/24hours transdermal patches | 12107041000033116 | 10449111000001101 | Clonidine |
| Catapres TTS 1 patches (Imported (Germany)) | 12107141000033117 | 10448811000001101 | Clonidine |
| Catapres TTS 2 patches (Imported (Germany)) | 12117941000033118 | 7657311000001101 | Clonidine |
| Catapres TTS 3 patches (Imported (Germany)) | 12118541000033113 | 10449311000001104 | Clonidine |
| Clonidine 100micrograms/5ml oral suspension | 12569041000033115 | 11813011000001109 | Clonidine hydrochloride |
| Clonidine 50micrograms/5ml oral solution sugar free | 12684041000033114 | 36392711000001102 | Clonidine hydrochloride |
| Guanfacine 1mg modified-release tablets | 10740841000033116 | 31087611000001109 | Guanfacine hydrochloride |
| Guanfacine 2mg modified-release tablets | 10740941000033112 | 31087711000001100 | Guanfacine hydrochloride |
| Guanfacine 3mg modified-release tablets | 10741041000033119 | 31087811000001108 | Guanfacine hydrochloride |

| | | | |
|---|---|---|---|
| Guanfacine 4mg modified-release tablets | 10741141000033115 | 31087911000001103 | Guanfacine hydrochloride |
| Intuniv 1mg modified-release tablets (Takeda UK Ltd) | 10741241000033110 | 31074011000001104 | Guanfacine hydrochloride |
| Intuniv 2mg modified-release tablets (Takeda UK Ltd) | 10741341000033117 | 31074311000001101 | Guanfacine hydrochloride |
| Intuniv 3mg modified-release tablets (Takeda UK Ltd) | 10741441000033111 | 31074611000001106 | Guanfacine hydrochloride |
| Intuniv 4mg modified-release tablets (Takeda UK Ltd) | 10741541000033112 | 31074911000001100 | Guanfacine hydrochloride |
| Aldomet 250mg tablets (Aspen Pharma Trading Ltd) | 40941000033118 | 253711000001107 | Methyldopa anhydrous |
| Aldomet 500mg tablets (Aspen Pharma Trading Ltd) | 41041000033111 | 73611000001108 | Methyldopa anhydrous |
| Methyldopa 250mg/5ml oral suspension | 892341000033112 | 8667311000001100 | Methyldopa anhydrous |
| Methyldopa 125mg tablets | 897341000033110 | 318671008 | Methyldopa anhydrous |
| Methyldopa 250mg tablets | 897441000033116 | 318672001 | Methyldopa anhydrous |
| Methyldopa 500mg tablets | 897541000033115 | 318673006 | Methyldopa anhydrous |
| Moxonidine 200microgram tablets | 938541000033115 | 318707000 | Moxonidine |
| Moxonidine 400microgram tablets | 938641000033119 | 318708005 | Moxonidine |
| Physiotens 200microgram tablets (Mylan) | 1080841000033119 | 41111000001102 | Moxonidine |
| Physiotens 400microgram tablets (Mylan) | 1080941000033110 | 142811000001107 | Moxonidine |
| Moxonidine 300microgram tablets | 2635641000033111 | 408604009 | Moxonidine |
| Physiotens 300microgram tablets (Mylan) | 2635741000033119 | 522011000001109 | Moxonidine |
| Ganda 3 + 0.5 eye drops (Bausch & Lomb UK Ltd) | 622441000033118 | 4668411000001105 | Adrenaline/ Guanethidine monosulfate |
| Ganda 1 + 0.2 eye drops (Bausch & Lomb UK Ltd) | 622641000033116 | 4668111000001100 | Adrenaline/ Guanethidine monosulfate |
| Guanethidine 10mg/1ml solution for injection ampoules | 655241000033111 | 36053311000001103 | Guanethidine monosulfate |
| Ismelin 10mg/1ml solution for injection ampoules (Amdipharm Plc) | 780741000033110 | 4369611000001106 | Guanethidine monosulfate |
| Loniten 10mg tablets (Pfizer Ltd) | 850441000033112 | 3666711000001100 | Minoxidil |
| Loniten 2.5mg tablets (Pfizer Ltd) | 850541000033113 | 3667011000001104 | Minoxidil |
| Loniten 5mg tablets (Pfizer Ltd) | 850641000033114 | 3666411000001106 | Minoxidil |
| Minoxidil 10mg tablets | 920641000033119 | 318657000 | Minoxidil |
| Minoxidil 2.5mg tablets | 920841000033118 | 318655008 | Minoxidil |
| Minoxidil 5mg tablets | 920941000033114 | 318656009 | Minoxidil |
| Minoxidil 5% solution | 921841000033111 | 36036311000001106 | Minoxidil |
| Minoxidil 2% solution | 921941000033115 | 36036211000001103 | Minoxidil |
| Regaine for Women Regular Strength 2% solution (McNeil Products Ltd) | 4026341000033113 | 9524011000001105 | Minoxidil |
| Regaine for Men Regular Strength 2% solution (McNeil Products Ltd) | 4026741000033114 | 9524811000001104 | Minoxidil |
| Regaine for Men Extra Strength 5% scalp solution (Johnson & Johnson Ltd) | 4026941000033112 | 9525311000001107 | Minoxidil |

| | | | |
|---|---|---|---|
| Minoxidil 2% gel | 4125241000033118 | 11472511000001105 | Minoxidil |
| Regaine for Men 2% gel (McNeil Products Ltd) | 4125341000033111 | 11465411000001105 | Minoxidil |
| Regaine for Men Extra Strength 5% scalp foam (McNeil Products Ltd) | 6448241000033118 | 18550611000001107 | Minoxidil |
| Minoxidil 5% foam | 6448341000033111 | 421143007 | Minoxidil |
| Regaine for Women Once a Day 5% scalp foam (McNeil Products Ltd) | 11803241000033110 | 32066411000001107 | Minoxidil |
| Sodium nitroprusside 50mg powder for solution for infusion vials | 1357141000033115 | 375899001 | Sodium nitroprusside dihydrate |
| Apresoline 20mg powder for solution for injection ampoules (Advanz Pharma) | 73441000033117 | 3925011000001100 | Hydralazine hydrochloride |
| Apresoline 25mg tablets (Advanz Pharma) | 74341000033114 | 657011000001101 | Hydralazine hydrochloride |
| Hydralazine 20mg powder for solution for injection ampoules | 734141000033114 | 34193811000001100 | Hydralazine hydrochloride |
| Hydralazine 25mg tablets | 741941000033113 | 318649003 | Hydralazine hydrochloride |
| Hydralazine 50mg tablets | 742041000033119 | 318650003 | Hydralazine hydrochloride |
| Hydralazine 10mg/5ml oral suspension | 4456241000033111 | 8528611000001105 | Hydralazine hydrochloride |
| Hydralazine 25mg/5ml oral solution | 11072741000033116 | 8580911000001106 | Hydralazine hydrochloride |
| Hydralazine 10mg tablets | 12877141000033119 | 414426001 | Hydralazine hydrochloride |
| Aliskiren 150mg tablets | 4213641000033112 | 39731411000001101 | Aliskiren hemifumarate |
| Aliskiren 300mg tablets | 4213741000033115 | 39731511000001102 | Aliskiren hemifumarate |
| Rasilez 150mg tablets (Noden Pharma DAC) | 4213841000033113 | 11960911000001108 | Aliskiren hemifumarate |
| Rasilez 300mg tablets (Noden Pharma DAC) | 4213941000033117 | 11961711000001103 | Aliskiren hemifumarate |

#### Table S73. Antiarrhythmic drugs class 1 Aurum product codes

| Term from EMIS | Prod code ID | dmd ID | Drug substance name |
|---|---|---|---|
| Emla 5% cream (Aspen Pharma Trading Ltd) | 511941000033116 | 2936311000001102 | Lidocaine/ Prilocaine |
| Xylotox 2% E80 Dental injection 2.2ml cartridges (Dentsply Ltd) | 1544441000033116 | 4207211000001108 | Adrenaline acid tartrate/ Lidocaine hydrochloride |
| Lidoderm 5% patches (Teikoku Seiyaku) | 2942141000033112 | 10570211000001105 | Lidocaine |
| Solarcaine 0.5% gel (Schering-Plough Ltd) | 2971941000033115 | 4011311000001103 | Lidocaine hydrochloride |
| Glucose 5% / Lidocaine 500mg/500ml (0.1%) infusion 500ml bags | 3096041000033116 | 5042311000001106 | Glucose anhydrous/ Lidocaine hydrochloride |
| Glucose 5% / Lidocaine 1g/500ml (0.2%) infusion 500ml bags | 3096141000033117 | 5042411000001104 | Glucose anhydrous/ Lidocaine hydrochloride |
| Lidocaine 100mg/10ml (1%) solution for injection Minijet pre-filled syringes (UCB Pharma Ltd) | 3096241000033112 | 4716611000001103 | Lidocaine hydrochloride |
| Lidocaine 100mg/5ml (2%) solution for injection Minijet pre-filled syringes (UCB Pharma Ltd) | 3096341000033119 | 9153311000001102 | Lidocaine hydrochloride |
| Lignospan Special 20mg/ml / 12.5micrograms/ml solution for injection 2.2ml cartridges (Septodont Ltd) | 4292941000033112 | 21313511000001103 | Adrenaline acid tartrate/ Lidocaine hydrochloride |
| Lignospan Special 20mg/ml / 12.5micrograms/ml solution for injection 1.8ml cartridges (Septodont Ltd) | 4418841000033118 | 35061911000001107 | Adrenaline acid tartrate/ Lidocaine hydrochloride |
| Lidocaine 70mg / Tetracaine 70mg medicated plasters | 4434341000033112 | 13210811000001108 | Lidocaine/ Tetracaine |
| Rapydan 70mg/70mg medicated plasters (EUSA Pharma (UK) Ltd) | 4434441000033118 | 13205811000001100 | Lidocaine/ Tetracaine |
| Lidocaine 4% cream | 4657441000033112 | 14203911000001107 | Lidocaine |
| Denela 5% cream (Teva UK Ltd) | 8840141000033111 | 23364211000001105 | Lidocaine/ Prilocaine |
| Ralvo 700mg medicated plasters (Grunenthal Ltd) | 12193941000033114 | 34178211000001102 | Lidocaine |
| Lidocaine 200mg/20ml (1%) solution for injection vials | 6132841000033115 | 3602411000001109 | Lidocaine hydrochloride |
| Lidocaine 400mg/20ml (2%) solution for injection vials | 6132641000033116 | 36039711000001100 | Lidocaine hydrochloride |
| Nulbia 5% cream (Glenmark Pharmaceuticals Europe Ltd) | 12421141000033115 | 34881811000001106 | Lidocaine/ Prilocaine |
| Lidocaine 50mg/5ml (1%) solution for injection Mini-Plasco ampoules (B.Braun Medical Ltd) | 12685841000033115 | 35863311000001108 | Lidocaine hydrochloride |
| Lidocaine 200mg/20ml (1%) solution for injection Mini-Plasco ampoules (B.Braun Medical Ltd) | 12685941000033111 | 35863611000001103 | Lidocaine hydrochloride |
| Lidocaine 100mg/5ml (2%) solution for injection Mini-Plasco ampoules (B.Braun Medical Ltd) | 12686041000033118 | 35863911000001109 | Lidocaine hydrochloride |
| Lidocaine 400mg/20ml (2%) solution for injection Mini-Plasco ampoules (B.Braun Medical Ltd) | 12686141000033119 | 35864311000001105 | Lidocaine hydrochloride |
| Lidocaine 100mg/10ml (1%) solution for injection Mini-Plasco ampoules (B.Braun Medical Ltd) | 12686341000033116 | 3768311000001105 | Lidocaine hydrochloride |
| Lidocaine 200mg/10ml (2%) solution for injection Mini-Plasco ampoules (B.Braun Medical Ltd) | 12686441000033110 | 4881511000001106 | Lidocaine hydrochloride |
| Kinidin 250mg Durules (AstraZeneca UK Ltd) | 804541000033110 | 3848511000001106 | Quinidine bisulfate |
| Quinidine sulfate 200mg tablets | 1149641000033114 | 318317007 | Quinidine sulfate |
| Quinidine bisulfate 250mg modified-release tablets | 1150941000033112 | 35933811000001102 | Quinidine bisulfate |

| | | | |
|---|---|---|---|
| Procainamide 1g/10ml solution for injection vials | 1119441000033119 | 34194111000001109 | Procainamide hydrochloride |
| Procainamide 250mg capsules | 5265241000033119 | 375342002 | Procainamide hydrochloride |
| Disopyramide 100mg capsules | 438841000033116 | 39690511000001109 | Disopyramide |
| Disopyramide 150mg capsules | 438941000033112 | 39690611000001108 | Disopyramide |
| Disopyramide 50mg/5ml solution for injection ampoules | 447241000033117 | 36070411000001104 | Disopyramide phosphate |
| Disopyramide 150mg modified-release tablets | 465241000033119 | 36070111000001109 | Disopyramide phosphate |
| Disopyramide 250mg modified-release tablets | 465341000033112 | 36070311000001106 | Disopyramide phosphate |
| Dirythmin SA 150mg tablets (AstraZeneca UK Ltd) | 469841000033118 | 3691511000001103 | Disopyramide phosphate |
| Rythmodan 100mg capsules (Neon Healthcare Ltd) | 1203941000033114 | 3465211000001104 | Disopyramide |
| Rythmodan 150mg capsules (Cheplapharm Arzneimittel GmbH) | 1204041000033111 | 3734711000001106 | Disopyramide |
| Rythmodan 50mg/5ml solution for injection ampoules (Sanofi) | 1204241000033115 | 4867311000001109 | Disopyramide phosphate |
| Rythmodan Retard 250mg tablets (Neon Healthcare Ltd) | 1205041000033112 | 3692111000001102 | Disopyramide phosphate |
| Disopyramide 250mg modified-release capsules | 2907741000033117 | 36070211000001103 | Disopyramide phosphate |
| Isomide CR 250mg capsules (Tillomed Laboratories Ltd) | 2907841000033110 | 3982011000001100 | Disopyramide phosphate |
| Disopyramide 50mg/5ml oral solution | 8770141000033110 | 8454511000001107 | Disopyramide phosphate |
| Mexiletine 50mg capsules | 872841000033111 | 318299008 | Mexiletine hydrochloride |
| Mexitil 50mg capsules (Boehringer Ingelheim Ltd) | 873041000033113 | 3877911000001101 | Mexiletine hydrochloride |
| Mexitil PL Perlongets 360mg capsules (Boehringer Ingelheim Ltd) | 873141000033112 | 4543311000001105 | Mexiletine hydrochloride |
| Mexiletine 250mg/10ml solution for injection ampoules | 879641000033116 | 36035711000001102 | Mexiletine hydrochloride |
| Mexitil 250mg/10ml solution for injection ampoules (Boehringer Ingelheim Ltd) | 879741000033113 | 4378311000001101 | Mexiletine hydrochloride |
| Mexiletine 100mg capsules | 8129541000033115 | 21394911000001108 | Mexiletine hydrochloride |
| Mexiletine hydrochloride 200mg (Mexiletine 167mg) capsules | 12888441000033116 | 36595811000001101 | Mexiletine hydrochloride |
| Namuscla 167mg capsules (Lupin Healthcare (UK) Ltd) | 12888541000033115 | 36538811000001103 | Mexiletine hydrochloride |
| Flecainide 150mg/15ml solution for injection ampoules | 587241000033119 | 36067511000001109 | Flecainide acetate |
| Flecainide 50mg tablets | 594541000033114 | 318276009 | Flecainide acetate |
| Flecainide 100mg tablets | 595141000033116 | 318277000 | Flecainide acetate |
| Tambocor 150mg/15ml solution for injection ampoules (Teva UK Ltd) | 1405841000033116 | 638511000001109 | Flecainide acetate |
| Tambocor 50mg tablets (Teva UK Ltd) | 1407441000033114 | 77511000001104 | Flecainide acetate |
| Tambocor 100mg tablets (Teva UK Ltd) | 1408041000033118 | 516411000001107 | Flecainide acetate |
| Flecainide 25mg/5ml oral solution | 2780641000033119 | 8514511000001105 | Flecainide acetate |
| Flecainide 200mg modified-release capsules | 4043141000033117 | 11169611000001105 | Flecainide acetate |
| Tambocor XL 200mg capsules (Teva UK Ltd) | 4043241000033112 | 11158611000001107 | Flecainide acetate |
| Flecainide 10mg/5ml oral suspension | 4432741000033117 | 8514411000001106 | Flecainide acetate |
| Flecainide 25mg/5ml oral suspension | 5993141000033114 | 8514611000001109 | Flecainide acetate |

| | | | |
|---|---|---|---|
| Arythmol 150mg tablets (Mylan) | 81441000033111 | 273811000001102 | Propafenone hydrochloride |
| Arythmol 300mg tablets (Mylan) | 81541000033112 | 69011000001108 | Propafenone hydrochloride |
| Propafenone 150mg tablets | 1140341000033110 | 318334004 | Propafenone hydrochloride |
| Propafenone 300mg tablets | 1140441000033116 | 318335003 | Propafenone hydrochloride |

#### Table S74. Antiarrhythmic drugs class 3 Aurum product codes

| Term from EMIS | Prod code ID | dmd ID | Drug substance name |
|---|---|---|---|
| Amiodarone 150mg/3ml solution for injection ampoules | 48341000033119 | 35900911000001106 | Amiodarone hydrochloride |
| Amiodarone 100mg tablets | 57641000033111 | 318186003 | Amiodarone hydrochloride |
| Amiodarone 200mg tablets | 57741000033119 | 318187007 | Amiodarone hydrochloride |
| Cordarone X 100 tablets (Zentiva Pharma UK Ltd) | 374641000033115 | 294711000001103 | Amiodarone hydrochloride |
| Cordarone X 200 tablets (Zentiva Pharma UK Ltd) | 374741000033112 | 269511000001109 | Amiodarone hydrochloride |
| Amiodarone 300mg/10ml solution for injection pre-filled syringes | 2644341000033117 | 35901011000001103 | Amiodarone hydrochloride |
| Amyben 100mg tablets (Lexon (UK) Ltd) | 2866241000033119 | 9108411000001107 | Amiodarone hydrochloride |
| Amyben 200mg tablets (Lexon (UK) Ltd) | 2866341000033112 | 9108711000001101 | Amiodarone hydrochloride |
| Amiodarone 25mg/5ml oral suspension | 3980241000033116 | 8275411000001105 | Amiodarone hydrochloride |
| Amiodarone 100mg/5ml oral solution | 3984941000033117 | 8274311000001102 | Amiodarone hydrochloride |
| Amiodarone 200mg/5ml oral suspension | 4505441000033115 | 8274611000001107 | Amiodarone hydrochloride |
| Amiodarone 50mg/5ml oral solution | 5897541000033119 | 8276111000001106 | Amiodarone hydrochloride |
| Amiodarone 50mg/5ml oral suspension | 5897641000033118 | 8276211000001100 | Amiodarone hydrochloride |
| Amiodarone 100mg/5ml oral suspension | 9774741000033117 | 8274411000001109 | Amiodarone hydrochloride |
| Beta-Cardone 200mg tablets (Advanz Pharma) | 143941000033116 | 447111000001104 | Sotalol hydrochloride |
| Beta-Cardone 40mg tablets (Focus Pharmaceuticals Ltd) | 144041000033119 | 900511000001103 | Sotalol hydrochloride |
| Beta-Cardone 80mg tablets (Focus Pharmaceuticals Ltd) | 144141000033115 | 104011000001101 | Sotalol hydrochloride |
| Sotacor 40mg/4ml solution for injection ampoules (Bristol-Myers Squibb Pharmaceuticals Ltd) | 1358541000033113 | 694711000001100 | Sotalol hydrochloride |
| Sotacor 160mg tablets (Bristol-Myers Squibb Pharmaceuticals Ltd) | 1366041000033112 | 446211000001108 | Sotalol hydrochloride |
| Sotacor 80mg tablets (Neon Healthcare Ltd) | 1366141000033111 | 463811000001109 | Sotalol hydrochloride |
| Sotalol 160mg tablets | 1367141000033113 | 318528007 | Sotalol hydrochloride |
| Sotalol 200mg tablets | 1367241000033118 | 318527002 | Sotalol hydrochloride |
| Sotalol 40mg tablets | 1367441000033117 | 318525005 | Sotalol hydrochloride |
| Sotalol 80mg tablets | 1367541000033116 | 318526006 | Sotalol hydrochloride |
| Sotalol 40mg/4ml solution for injection ampoules | 2908541000033114 | 35930811000001109 | Sotalol hydrochloride |
| Sotalol 25mg/5ml oral solution | 3953441000033116 | 8726811000001107 | Sotalol hydrochloride |
| Sotalol 25mg/5ml oral suspension | 9776441000033119 | 8726911000001102 | Sotalol hydrochloride |
| Dronedarone 400mg tablets | 5566541000033116 | 39691711000001107 | Dronedarone hydrochloride |
| Multaq 400mg tablets (Sanofi) | 5566641000033115 | 17066411000001106 | Dronedarone hydrochloride |

#### Table S75. Digoxin Aurum product codes

| Term from EMIS | Prod code ID | dmd ID | Drug substance name |
|---|---|---|---|
| Azilsartan medoxomil 20mg tablets | 7754841000033119 | 20418711000001104 | Azilsartan medoxomil |
| Azilsartan medoxomil 40mg tablets | 7754941000033110 | 449109006 | Azilsartan medoxomil |
| Azilsartan medoxomil 80mg tablets | 7755041000033110 | 449333009 | Azilsartan medoxomil |
| Edarbi 20mg tablets (Takeda UK Ltd) | 7755141000033114 | 20350911000001100 | Azilsartan medoxomil |
| Edarbi 40mg tablets (Takeda UK Ltd) | 7755241000033119 | 20351211000001103 | Azilsartan medoxomil |
| Edarbi 80mg tablets (Takeda UK Ltd) | 7755341000033112 | 20351811000001102 | Azilsartan medoxomil |
| Amias 16mg tablets (Takeda UK Ltd) | 59141000033113 | 908511000001100 | Candesartan cilexetil |
| Amias 2mg tablets (Takeda UK Ltd) | 59241000033118 | 97311000001103 | Candesartan cilexetil |
| Amias 4mg tablets (Takeda UK Ltd) | 59341000033111 | 857411000001100 | Candesartan cilexetil |
| Amias 8mg tablets (Takeda UK Ltd) | 59441000033117 | 36011000001106 | Candesartan cilexetil |
| Candesartan 16mg tablets | 211541000033119 | 318980005 | Candesartan cilexetil |
| Candesartan 2mg tablets | 211641000033118 | 318977009 | Candesartan cilexetil |
| Candesartan 4mg tablets | 211741000033110 | 318978004 | Candesartan cilexetil |
| Candesartan 8mg tablets | 211841000033117 | 318979007 | Candesartan cilexetil |
| Candesartan 32mg tablets | 3227241000033111 | 376998003 | Candesartan cilexetil |
| Amias 32mg tablets (Takeda UK Ltd) | 3227341000033118 | 8983911000001107 | Candesartan cilexetil |
| Eprosartan 300mg tablets | 2036341000033115 | 318994006 | Eprosartan mesilate |
| Eprosartan 400mg tablets | 2036441000033114 | 318995007 | Eprosartan mesilate |
| Eprosartan 600mg tablets | 2036541000033110 | 318996008 | Eprosartan mesilate |
| Teveten 300mg tablets (Mylan) | 2040841000033113 | 401211000001105 | Eprosartan mesilate |
| Teveten 400mg tablets (Abbott Healthcare Products Ltd) | 2040941000033117 | 151411000001103 | Eprosartan mesilate |
| Teveten 600mg tablets (Mylan) | 2041041000033110 | 872011000001109 | Eprosartan mesilate |
| Aprovel 150mg tablets (Sanofi) | 75941000033114 | 859711000001103 | Irbesartan |
| Aprovel 300mg tablets (Sanofi) | 76041000033116 | 323211000001107 | Irbesartan |
| Aprovel 75mg tablets (Sanofi) | 76141000033117 | 434511000001104 | Irbesartan |
| Irbesartan 150mg tablets | 775441000033117 | 318969005 | Irbesartan |
| Irbesartan 300mg tablets | 775541000033116 | 318970006 | Irbesartan |
| Irbesartan 75mg tablets | 775641000033115 | 318968002 | Irbesartan |
| CoAprovel 150mg/12.5mg tablets (Sanofi) | 2216141000033115 | 792411000001108 | Hydrochlorothiazide/ Irbesartan |
| CoAprovel 300mg/12.5mg tablets (Sanofi) | 2216241000033110 | 682711000001109 | Hydrochlorothiazide/ Irbesartan |
| Irbesartan 150mg / Hydrochlorothiazide 12.5mg tablets | 3161141000033116 | 134461004 | Hydrochlorothiazide/ Irbesartan |
| Irbesartan 300mg / Hydrochlorothiazide 12.5mg tablets | 3161241000033111 | 134460003 | Hydrochlorothiazide/ Irbesartan |
| Irbesartan 300mg / Hydrochlorothiazide 25mg tablets | 3995241000033112 | 10970311000001105 | Hydrochlorothiazide/ Irbesartan |
| CoAprovel 300mg/25mg tablets (Sanofi) | 3995341000033119 | 10968611000001106 | Hydrochlorothiazide/ Irbesartan |
| Irbesartan 300mg/5ml oral suspension | 4954341000033111 | 12639511000001103 | Irbesartan |

| | | | |
|---|---|---|---|
| Irbesartan 150mg/5ml oral suspension | 5970641000033117 | 8580811000001101 | Irbesartan |
| Irbesartan 37.5mg oral powder sachets | 6435741000033117 | 19481911000001105 | Irbesartan |
| Ifirmasta 75mg tablets (Consilient Health Ltd) | 8554141000033116 | 22720311000001108 | Irbesartan |
| Ifirmasta 150mg tablets (Consilient Health Ltd) | 8554241000033111 | 22720611000001103 | Irbesartan |
| Ifirmasta 300mg tablets (Consilient Health Ltd) | 8554341000033118 | 22720811000001104 | Irbesartan |
| Cozaar 25mg tablets (Organon Pharma (UK) Ltd) | 370441000033112 | 266511000001104 | Losartan potassium |
| Cozaar 50mg tablets (Organon Pharma (UK) Ltd) | 370541000033113 | 53611000001106 | Losartan potassium |
| Cozaar-Comp 50mg/12.5mg tablets (Organon Pharma (UK) Ltd) | 370841000033110 | 255911000001105 | Hydrochlorothiazide/ Losartan potassium |
| Losartan 25mg tablets | 851841000033117 | 318955005 | Losartan potassium |
| Losartan 50mg tablets | 851941000033113 | 318956006 | Losartan potassium |
| Losartan 100mg tablets | 2720141000033115 | 407784004 | Losartan potassium |
| Cozaar 100mg tablets (Organon Pharma (UK) Ltd) | 2720241000033110 | 245811000001102 | Losartan potassium |
| Losartan 50mg / Hydrochlorothiazide 12.5mg tablets | 3161741000033117 | 318959004 | Hydrochlorothiazide/ Losartan potassium |
| Losartan 100mg / Hydrochlorothiazide 25mg tablets | 3346841000033117 | 395497004 | Hydrochlorothiazide/ Losartan potassium |
| Cozaar-Comp 100mg/25mg tablets (Organon Pharma (UK) Ltd) | 3346941000033113 | 9566911000001105 | Hydrochlorothiazide/ Losartan potassium |
| Losartan 100mg / Hydrochlorothiazide 12.5mg tablets | 4424741000033110 | 13112711000001103 | Hydrochlorothiazide/ Losartan potassium |
| Cozaar-Comp 100mg/12.5mg tablets (Organon Pharma (UK) Ltd) | 4424841000033117 | 13094111000001102 | Hydrochlorothiazide/ Losartan potassium |
| Losartan 12.5mg tablets | 4957541000033114 | 15148111000001100 | Losartan potassium |
| Cozaar 12.5mg tablets (Organon Pharma (UK) Ltd) | 4957641000033110 | 15138911000001101 | Losartan potassium |
| Losartan 2.5mg/ml oral suspension sugar free | 5149941000033118 | 15507411000001105 | Losartan potassium |
| Cozaar 2.5mg/ml oral suspension (Organon Pharma (UK) Ltd) | 5150041000033110 | 15506811000001105 | Losartan potassium |
| Losartan 100mg/5ml oral solution | 5971741000033116 | 15451111000001107 | Losartan potassium |
| Losartan 100mg/5ml oral suspension | 5971841000033114 | 15451211000001101 | Losartan potassium |
| Losartan 50mg/5ml oral solution | 5971941000033118 | 14159411000001106 | Losartan potassium |
| Losartan 50mg/5ml oral suspension | 5972041000033112 | 14159511000001105 | Losartan potassium |
| Olmesartan medoxomil 10mg tablets | 2944541000033111 | 408055003 | Olmesartan medoxomil |
| Olmesartan medoxomil 20mg tablets | 2944641000033112 | 385542009 | Olmesartan medoxomil |
| Olmesartan medoxomil 40mg tablets | 2944741000033115 | 385543004 | Olmesartan medoxomil |
| Olmetec 10mg tablets (Daiichi Sankyo UK Ltd) | 2944841000033113 | 4624011000001101 | Olmesartan medoxomil |
| Olmetec 20mg tablets (Daiichi Sankyo UK Ltd) | 2944941000033117 | 4624311000001103 | Olmesartan medoxomil |
| Olmetec 40mg tablets (Daiichi Sankyo UK Ltd) | 2945041000033117 | 4624611000001108 | Olmesartan medoxomil |
| Olmesartan medoxomil 20mg / Hydrochlorothiazide 12.5mg tablets | 3908941000033112 | 409184002 | Hydrochlorothiazide/ Olmesartan medoxomil |
| Olmesartan medoxomil 20mg / Hydrochlorothiazide 25mg tablets | 3909041000033115 | 10270711000001105 | Hydrochlorothiazide/ Olmesartan medoxomil |
| Olmetec Plus 20mg/12.5mg tablets (Daiichi Sankyo UK Ltd) | 3909141000033116 | 10261511000001103 | Hydrochlorothiazide/ Olmesartan medoxomil |

| | | | |
|---|---|---|---|
| Olmetec Plus 20mg/25mg tablets (Daiichi Sankyo UK Ltd) | 3909241000033111 | 10261811000001100 | Hydrochlorothiazide/ Olmesartan medoxomil |
| Olmesartan medoxomil 10mg/5ml oral suspension | 4273241000033118 | 14680711000001103 | Olmesartan medoxomil |
| Olmesartan medoxomil 20mg / Amlodipine 5mg tablets | 5005841000033114 | 429502004 | Amlodipine besilate/ Olmesartan medoxomil |
| Olmesartan medoxomil 40mg / Amlodipine 5mg tablets | 5005941000033118 | 429503009 | Amlodipine besilate/ Olmesartan medoxomil |
| Olmesartan medoxomil 40mg / Amlodipine 10mg tablets | 5006041000033111 | 429678006 | Amlodipine besilate/ Olmesartan medoxomil |
| Sevikar 20mg/5mg tablets (Daiichi Sankyo UK Ltd) | 5006141000033110 | 15773211000001105 | Amlodipine besilate/ Olmesartan medoxomil |
| Sevikar 40mg/5mg tablets (Daiichi Sankyo UK Ltd) | 5006241000033115 | 15772911000001108 | Amlodipine besilate/ Olmesartan medoxomil |
| Sevikar 40mg/10mg tablets (Daiichi Sankyo UK Ltd) | 5006341000033113 | 15772611000001102 | Amlodipine besilate/ Olmesartan medoxomil |
| Olmesartan medoxomil 40mg / Hydrochlorothiazide 12.5mg tablets | 5566341000033111 | 409185001 | Hydrochlorothiazide/ Olmesartan medoxomil |
| Olmetec Plus 40mg/12.5mg tablets (Daiichi Sankyo UK Ltd) | 5566441000033117 | 17220911000001102 | Hydrochlorothiazide/ Olmesartan medoxomil |
| Sevikar HCT 20mg/5mg/12.5mg tablets (Daiichi Sankyo UK Ltd) | 6180641000033112 | 18986411000001108 | Amlodipine besilate/ Hydrochlorothiazide/ Olmesartan medoxomil |
| Sevikar HCT 40mg/5mg/12.5mg tablets (Daiichi Sankyo UK Ltd) | 6180741000033115 | 18986711000001102 | Amlodipine besilate/ Hydrochlorothiazide/ Olmesartan medoxomil |
| Sevikar HCT 40mg/10mg/12.5mg tablets (Daiichi Sankyo UK Ltd) | 6180841000033113 | 18987011000001101 | Amlodipine besilate/ Hydrochlorothiazide/ Olmesartan medoxomil |
| Sevikar HCT 40mg/5mg/25mg tablets (Daiichi Sankyo UK Ltd) | 6180941000033117 | 18987311000001103 | Amlodipine besilate/ Hydrochlorothiazide/ Olmesartan medoxomil |
| Sevikar HCT 40mg/10mg/25mg tablets (Daiichi Sankyo UK Ltd) | 6181041000033110 | 18987611000001108 | Amlodipine besilate/ Hydrochlorothiazide/ Olmesartan medoxomil |
| Diovan 160mg capsules (Novartis Pharmaceuticals UK Ltd) | 437641000033118 | 117011000001107 | Valsartan |
| Diovan 40mg capsules (Novartis Pharmaceuticals UK Ltd) | 437741000033110 | 777611000001101 | Valsartan |
| Diovan 80mg capsules (Novartis Pharmaceuticals UK Ltd) | 437841000033117 | 554511000001105 | Valsartan |
| Valsartan 160mg capsules | 1498241000033111 | 318963006 | Valsartan |
| Valsartan 40mg capsules | 1498341000033118 | 318961008 | Valsartan |
| Valsartan 80mg capsules | 1498441000033112 | 318962001 | Valsartan |
| Co-Diovan 160mg/12.5mg tablets (Novartis Pharmaceuticals UK Ltd) | 3152541000033112 | 7668611000001104 | Hydrochlorothiazide/ Valsartan |
| Co-Diovan 160mg/25mg tablets (Novartis Pharmaceuticals UK Ltd) | 3152641000033113 | 7668911000001105 | Hydrochlorothiazide/ Valsartan |

| | | | |
|---|---|---|---|
| Valsartan 160mg / Hydrochlorothiazide 12.5mg tablets | 3161441000033112 | 395521005 | Hydrochlorothiazide/ Valsartan |
| Valsartan 160mg / Hydrochlorothiazide 25mg tablets | 3161541000033113 | 409298002 | Hydrochlorothiazide/ Valsartan |
| Valsartan 80mg / Hydrochlorothiazide 12.5mg tablets | 3190741000033112 | 377488008 | Hydrochlorothiazide/ Valsartan |
| Co-Diovan 80mg/12.5mg tablets (Novartis Pharmaceuticals UK Ltd) | 3190841000033119 | 8150111000001108 | Hydrochlorothiazide/ Valsartan |
| Valsartan 40mg tablets | 3201341000033115 | 416515008 | Valsartan |
| Diovan 40mg tablets (Novartis Pharmaceuticals UK Ltd) | 3201441000033114 | 8263211000001101 | Valsartan |
| Amlodipine 5mg / Valsartan 80mg tablets | 4021441000033114 | 11160311000001109 | Amlodipine besilate/ Valsartan |
| Amlodipine 5mg / Valsartan 160mg tablets | 4021541000033110 | 11160211000001101 | Amlodipine besilate/ Valsartan |
| Amlodipine 10mg / Valsartan 160mg tablets | 4021641000033111 | 11160111000001107 | Amlodipine besilate/ Valsartan |
| Exforge 5mg/80mg tablets (Novartis Pharmaceuticals UK Ltd) | 4021741000033119 | 11161811000001108 | Amlodipine besilate/ Valsartan |
| Exforge 5mg/160mg tablets (Novartis Pharmaceuticals UK Ltd) | 4021841000033112 | 11161511000001105 | Amlodipine besilate/ Valsartan |
| Exforge 10mg/160mg tablets (Novartis Pharmaceuticals UK Ltd) | 4021941000033116 | 11160711000001108 | Amlodipine besilate/ Valsartan |
| Valsartan 320mg tablets | 4424941000033113 | 376487009 | Valsartan |
| Diovan 320mg tablets (Novartis Pharmaceuticals UK Ltd) | 4425041000033113 | 13143311000001102 | Valsartan |
| Valsartan 80mg tablets | 6515441000033113 | 375034009 | Valsartan |
| Valsartan 160mg tablets | 6515541000033114 | 375035005 | Valsartan |
| Valsartan 3mg/ml oral solution | 6528441000033113 | 20007411000001100 | Valsartan |
| Diovan 3mg/1ml oral solution (Novartis Pharmaceuticals UK Ltd) | 6528541000033114 | 20001711000001102 | Valsartan |
| Sacubitril 24mg / Valsartan 26mg tablets | 10943641000033117 | 31142011000001103 | Sacubitril/ Valsartan |
| Sacubitril 49mg / Valsartan 51mg tablets | 10943741000033114 | 31142111000001102 | Sacubitril/ Valsartan |
| Sacubitril 97mg / Valsartan 103mg tablets | 10943841000033116 | 31142211000001108 | Sacubitril/ Valsartan |
| Entresto 24mg/26mg tablets (Novartis Pharmaceuticals UK Ltd) | 10943941000033112 | 31136811000001109 | Sacubitril/ Valsartan |
| Entresto 49mg/51mg tablets (Novartis Pharmaceuticals UK Ltd) | 10944041000033114 | 31136411000001107 | Sacubitril/ Valsartan |
| Entresto 97mg/103mg tablets (Novartis Pharmaceuticals UK Ltd) | 10944141000033113 | 31138011000001104 | Sacubitril/ Valsartan |

#### Table S76. Sodium-glucose co-transporter-2 inhibitors Aurum product codes

| Term from EMIS | Prod code ID | dmd ID | Drug substance name |
|---|---|---|---|
| Forxiga 5mg tablets (AstraZeneca UK Ltd) | 8199541000033113 | 21609511000001105 | Dapagliflozin propanediol monohydrate |
| Dapagliflozin 5mg tablets | 8199341000033118 | 703679006 | Dapagliflozin propanediol monohydrate |
| Dapagliflozin 10mg tablets | 8199441000033112 | 703680009 | Dapagliflozin propanediol monohydrate |
| Forxiga 10mg tablets (AstraZeneca UK Ltd) | 8199641000033114 | 21609811000001108 | Dapagliflozin propanediol monohydrate |
| Xigduo 5mg/1000mg tablets (AstraZeneca UK Ltd) | 9106241000033119 | 24018511000001102 | Dapagliflozin propanediol monohydrate/ Metformin hydrochloride |
| Dapagliflozin 5mg / Metformin 1g tablets | 9106041000033110 | 24054611000001100 | Dapagliflozin propanediol monohydrate/ Metformin hydrochloride |
| Dapagliflozin 5mg / Metformin 850mg tablets | 9106141000033114 | 24054711000001109 | Dapagliflozin propanediol monohydrate/ Metformin hydrochloride |
| Xigduo 5mg/850mg tablets (AstraZeneca UK Ltd) | 9106341000033112 | 24018111000001106 | Dapagliflozin propanediol monohydrate/ Metformin hydrochloride |
| Saxagliptin 5mg / Dapagliflozin 10mg tablets | 11898041000033111 | 33745311000001109 | Dapagliflozin propanediol monohydrate/ Saxagliptin hydrochloride |
| Qtern 5mg/10mg tablets (AstraZeneca UK Ltd) | 11898141000033110 | 33682311000001103 | Dapagliflozin propanediol monohydrate/ Saxagliptin hydrochloride |
| Jardiance 10mg tablets (Boehringer Ingelheim Ltd) | 9337141000033114 | 25238811000001107 | Empagliflozin |
| Empagliflozin 10mg tablets | 9336641000033119 | 25290511000001101 | Empagliflozin |
| Empagliflozin 25mg tablets | 9336841000033118 | 25290611000001102 | Empagliflozin |
| Jardiance 25mg tablets (Boehringer Ingelheim Ltd) | 9337241000033119 | 25239711000001108 | Empagliflozin |
| Empagliflozin 12.5mg / Metformin 1g tablets | 10614441000033112 | 30318111000001109 | Empagliflozin/ Metformin hydrochloride |
| Synjardy 12.5mg/1000mg tablets (Boehringer Ingelheim Ltd) | 10614841000033110 | 30175711000001100 | Empagliflozin/ Metformin hydrochloride |
| Synjardy 12.5mg/850mg tablets (Boehringer Ingelheim Ltd) | 10614741000033117 | 30175011000001102 | Empagliflozin/ Metformin hydrochloride |
| Empagliflozin 12.5mg / Metformin 850mg tablets | 10614341000033118 | 30318211000001103 | Empagliflozin/ Metformin hydrochloride |

| | | | |
|---|---|---|---|
| Empagliflozin 5mg / Metformin 1g tablets | 10614241000033111 | 30318311000001106 | Empagliflozin/ Metformin hydrochloride |
| Synjardy 5mg/1000mg tablets (Boehringer Ingelheim Ltd) | 10614641000033114 | 30174111000001100 | Empagliflozin/ Metformin hydrochloride |
| Synjardy 5mg/850mg tablets (Boehringer Ingelheim Ltd) | 10614541000033113 | 30173411000001107 | Empagliflozin/ Metformin hydrochloride |
| Empagliflozin 5mg / Metformin 850mg tablets | 10614141000033116 | 30318411000001104 | Empagliflozin/ Metformin hydrochloride |
| Empagliflozin 10mg / Linagliptin 5mg tablets | 13116741000033112 | 37280311000001109 | Empagliflozin/ Linagliptin |
| Glyxambi 10mg/5mg tablets (Boehringer Ingelheim Ltd) | 13116941000033110 | 37225211000001100 | Empagliflozin/ Linagliptin |
| Glyxambi 25mg/5mg tablets (Boehringer Ingelheim Ltd) | 13117041000033111 | 37225511000001102 | Empagliflozin/ Linagliptin |
| Empagliflozin 25mg / Linagliptin 5mg tablets | 13116841000033119 | 37280511000001103 | Empagliflozin/ Linagliptin |
| Canagliflozin 100mg tablets | 9110341000033112 | 703682001 | Canagliflozin hemihydrate |
| Invokana 100mg tablets (Napp Pharmaceuticals Ltd) | 9110541000033117 | 24088611000001101 | Canagliflozin hemihydrate |
| Canagliflozin 300mg tablets | 9110441000033118 | 24104511000001103 | Canagliflozin hemihydrate |
| Invokana 300mg tablets (Napp Pharmaceuticals Ltd) | 9110641000033116 | 24088311000001106 | Canagliflozin hemihydrate |
| Canagliflozin 50mg / Metformin 1g tablets | 9851541000033114 | 28049211000001101 | Canagliflozin hemihydrate/ Metformin hydrochloride |
| Vokanamet 50mg/1000mg tablets (Napp Pharmaceuticals Ltd) | 9851841000033111 | 28024411000001103 | Canagliflozin hemihydrate/ Metformin hydrochloride |
| Vokanamet 50mg/850mg tablets (Napp Pharmaceuticals Ltd) | 9851741000033118 | 28022511000001109 | Canagliflozin hemihydrate/ Metformin hydrochloride |
| Canagliflozin 50mg / Metformin 850mg tablets | 9851641000033110 | 28049311000001109 | Canagliflozin hemihydrate/ Metformin hydrochloride |

#### Table S77. Nonsteroidal anti-inflammatory drug Aurum product codes

| Term from EMIS | Prod code ID | dmd ID | Drug substance name |
|---|---|---|---|
| Aspirin 75mg dispersible tablets | 82941000033113 | 319773006 | Aspirin |
| PostMI 75 dispersible tablets (Ashbourne Pharmaceuticals Ltd) | 1098041000033111 | 636011000001100 | Aspirin |
| Danamep 75mg dispersible tablets (Ecogen Europe Ltd) | 11716941000033112 | 32968711000001107 | Aspirin |
| Mandaprin 75mg dispersible tablets (M & A Pharmachem Ltd) | 12684641000033115 | 35581511000001100 | Aspirin |
| Angettes 75 tablets (Bristol-Myers Squibb Pharmaceuticals Ltd) | 71241000033119 | 800511000001106 | Aspirin |
| Aspirin 75mg tablets | 87041000033110 | 319775004 | Aspirin |
| Aspirin 75mg gastro-resistant tablets | 83041000033115 | 319781007 | Aspirin |
| Caprin 75mg gastro-resistant tablets (Wockhardt UK Ltd) | 189841000033119 | 927411000001108 | Aspirin |
| Enprin 75mg gastro-resistant tablets (Galpharm International Ltd) | 1729141000033110 | 473711000001109 | Aspirin |
| Nu-Seals 75 gastro-resistant tablets (Alliance Pharmaceuticals Ltd) | 991441000033115 | 414311000001102 | Aspirin |
| Micropirin 75mg gastro-resistant tablets (Dexcel-Pharma Ltd) | 2273741000033118 | 281311000001104 | Aspirin |
| PostMI 75 EC tablets (Ashbourne Pharmaceuticals Ltd) | 1101541000033119 | 38411000001101 | Aspirin |
| Combogesic 500mg/150mg tablets (Thornton & Ross Ltd) | 11781641000033113 | 33563011000001103 | Ibuprofen/ Paracetamol |
| Paracetamol 500mg / Ibuprofen 150mg tablets | 11781541000033112 | 33568211000001100 | Ibuprofen/ Paracetamol |
| Ibuprofen lysine 400mg tablets | 5376641000033111 | 10245111000001102 | Ibuprofen lysine |
| Nurofen Maximum Strength Migraine Pain 684mg caplets (Reckitt Benckiser Healthcare (UK) Ltd) | 5376741000033119 | 10239611000001109 | Ibuprofen lysine |
| Caprin 300mg gastro-resistant tablets (Pinewood Healthcare) | 212341000033117 | 901211000001107 | Aspirin |
| Nu-Seals 300 gastro-resistant tablets (Alliance Pharmaceuticals Ltd) | 993341000033113 | 183311000001101 | Aspirin |
| Aspirin 300mg gastro-resistant tablets | 83141000033116 | 322225002 | Aspirin |
| Aspirin 300mg suppositories | 2013341000033115 | 322232006 | Aspirin |
| Aspirin 150mg suppositories | 2013441000033114 | 322233001 | Aspirin |
| Co-codaprin 8mg/400mg dispersible tablets | 371041000033112 | 322310004 | Aspirin/ Codeine phosphate |
| Aspav dispersible tablets (Actavis UK Ltd) | 86841000033118 | 3690911000001103 | Aspirin/ Papaveretum |
| Aspirin 500mg / Papaveretum 7.71mg dispersible tablets sugar free | 3849841000033110 | 322321008 | Aspirin/ Papaveretum |
| MigraMax oral powder sachets (Zentiva) | 2086841000033117 | 3637911000001108 | Aspirin DL-Lysine/ Metoclopramide hydrochloride |
| Aspirin 900mg / Metoclopramide 10mg oral powder sachets sugar free | 3836841000033119 | 322784004 | Aspirin DL-Lysine/ Metoclopramide hydrochloride |
| Brufen Retard 800mg tablets (Mylan) | 159441000033110 | 368011000001100 | Ibuprofen |
| Ibuprofen 800mg modified-release tablets | 746941000033117 | 39110311000001101 | Ibuprofen |
| Ibuprofen 200mg / Phenylephrine 6.1mg tablets | 12496141000033117 | 34878411000001108 | Ibuprofen/ Phenylephrine |

| | | | |
|---|---|---|---|
| Bisoprolol 10mg / Aspirin 100mg capsules | 10025241000033112 | 28365311000001102 | Aspirin/ Bisoprolol fumarate |
| Bisoprolol 10mg / Aspirin 75mg capsules | 10025141000033117 | 28365411000001109 | Aspirin/ Bisoprolol fumarate |
| Bisoprolol 5mg / Aspirin 100mg capsules | 10025041000033116 | 28365611000001107 | Aspirin/ Bisoprolol fumarate |
| Bisoprolol 5mg / Aspirin 75mg capsules | 10024941000033116 | 28365711000001103 | Aspirin/ Bisoprolol fumarate |
| Co-codaprin 8mg/400mg tablets | 370941000033119 | 11762111000001100 | Aspirin/ Codeine phosphate |
| Imazin XL forte tablets (Napp Pharmaceuticals Ltd) | 1739641000033115 | 3461511000001104 | Aspirin/ Isosorbide mononitrate |
| Aspirin 150mg / Isosorbide mononitrate 60mg modified-release tablets | 3162141000033112 | 35903011000001102 | Aspirin/ Isosorbide mononitrate |
| Imazin XL tablets (Napp Pharmaceuticals Ltd) | 1739541000033116 | 3460911000001104 | Aspirin/ Isosorbide mononitrate |
| Aspirin 75mg / Isosorbide mononitrate 60mg modified-release tablets | 3162041000033113 | 35903111000001101 | Aspirin/ Isosorbide mononitrate |
| Nurofen Meltlets 200mg tablets (Reckitt Benckiser Healthcare (UK) Ltd) | 2839541000033115 | 3869111000001103 | Ibuprofen |
| Ibuprofen 200mg orodispersible tablets sugar free | 2839441000033116 | 3875511000001104 | Ibuprofen |
| Aspirin 300mg tablets | 86641000033119 | 329525004 | Aspirin |
| Aspirin 300mg dispersible tablets | 82841000033117 | 329526003 | Aspirin |
| Ibuprofen 200mg tablets | 747341000033119 | 329652003 | Ibuprofen |
| Cuprofen 200mg tablets (SSL International Plc) | 4432641000033114 | 640911000001100 | Ibuprofen |
| Brufen 200mg tablets (Abbott Laboratories Ltd) | 162841000033113 | 643711000001106 | Ibuprofen |
| Nurofen 200mg caplets (Reckitt Benckiser Healthcare (UK) Ltd) | 2970741000033111 | 621911000001109 | Ibuprofen |
| Arthrofen 200 tablets (Ashbourne Pharmaceuticals Ltd) | 81641000033113 | 389511000001100 | Ibuprofen |
| Librofem 200mg tablets (LPC Medical (UK) Ltd) | 1627241000033117 | 901911000001103 | Ibuprofen |
| Advil 200mg tablets (Wyeth Consumer Healthcare) | 2968941000033119 | 8970311000001109 | Ibuprofen |
| Hedex Ibuprofen 200mg tablets (Omega Pharma Ltd) | 2968541000033113 | 17042011000001106 | Ibuprofen |
| Motrin 400mg tablets (Pfizer Ltd) | 939741000033119 | 868911000001103 | Ibuprofen |
| Arthrofen 400 tablets (Ashbourne Pharmaceuticals Ltd) | 81741000033116 | 933011000001107 | Ibuprofen |
| Brufen 400mg tablets (Mylan) | 162941000033117 | 263211000001109 | Ibuprofen |
| Ibuprofen 400mg tablets | 747441000033113 | 329653008 | Ibuprofen |
| Ibuprofen 600mg tablets | 747541000033114 | 329654002 | Ibuprofen |
| Brufen 600mg tablets (Mylan) | 163041000033110 | 230111000001100 | Ibuprofen |
| Arthrofen 600 tablets (Ashbourne Pharmaceuticals Ltd) | 81841000033114 | 415611000001108 | Ibuprofen |
| Motrin 600mg tablets (Pfizer Ltd) | 939841000033112 | 550011000001109 | Ibuprofen |
| Ibuprofen 200mg / Codeine 12.8mg tablets | 12708841000033116 | 329683004 | Codeine phosphate/ Ibuprofen |

| | | | |
|---|---|---|---|
| Nurofen Plus tablets (Reckitt Benckiser Healthcare (UK) Ltd) | 2228341000033113 | 3915411000001104 | Codeine phosphate/ Ibuprofen |
| Solpaflex tablets (GlaxoSmithKline Consumer Healthcare) | 2968641000033114 | 3916211000001109 | Codeine phosphate/ Ibuprofen |
| Cuprofen PLUS tablets (SSL International Plc) | 3077941000033110 | 8092711000001106 | Codeine phosphate/ Ibuprofen |
| Motrin 800mg tablets (Pfizer Ltd) | 939941000033116 | 62911000001102 | Ibuprofen |
| Ibuprofen 800mg tablets | 746541000033111 | 329708004 | Ibuprofen |
| Ibuprofen lysine 200mg tablets | 4661241000033110 | 4557011000001104 | Ibuprofen lysine |
| Disprin CV 300mg modified-release tablets (Reckitt Benckiser Healthcare (UK) Ltd) | 452541000033115 | 4532311000001104 | Aspirin |
| Aspirin 300mg modified-release tablets | 84441000033114 | 4558011000001103 | Aspirin |
| Maximum Strength Aspro Clear 500mg effervescent tablets (Bayer Plc) | 4431941000033114 | 4648711000001109 | Aspirin |
| Aspro Clear 300mg effervescent tablets (Bayer Plc) | 4431841000033118 | 4649111000001101 | Aspirin |
| Codafen Continus tablets (Napp Pharmaceuticals Ltd) | 336741000033117 | 3420111000001109 | Codeine phosphate/ Ibuprofen |
| Ibuprofen 300mg modified-release / Codeine 20mg tablets | 3229141000033111 | 36045211000001108 | Codeine phosphate/ Ibuprofen |
| Aspirin 500mg / Codeine 8mg dispersible tablets sugar free | 3934341000033112 | 4956111000001107 | Aspirin/ Codeine phosphate |
| Codis 500 dispersible tablets (Reckitt Benckiser Healthcare (UK) Ltd) | 3934441000033118 | 4951811000001105 | Aspirin/ Codeine phosphate |
| Aspirin powder | 2085241000033111 | 5145711000001107 | Aspirin |
| Asasantin Retard capsules (Boehringer Ingelheim Ltd) | 1571541000033117 | 3292511000001108 | Aspirin/ Dipyridamole |
| Dipyridamole 200mg modified-release / Aspirin 25mg capsules | 3160741000033110 | 36069911000001109 | Aspirin/ Dipyridamole |
| Atransipar 200mg/25mg modified-release capsules (Par Laboratories Europe Ltd) | 12352041000033117 | 34609311000001105 | Aspirin/ Dipyridamole |
| Molita 200mg/25mg modified-release capsules (Dr Reddy's Laboratories (UK) Ltd) | 8619141000033116 | 23241511000001106 | Aspirin/ Dipyridamole |
| Ibuprofen 200mg / Pseudoephedrine hydrochloride 30mg tablets | 4123241000033119 | 407904005 | Ibuprofen/ Pseudoephedrine hydrochloride |
| Nurofen Cold and Flu tablets (Reckitt Benckiser Healthcare (UK) Ltd) | 4123741000033113 | 4606211000001109 | Ibuprofen/ Pseudoephedrine hydrochloride |
| Dexibuprofen 300mg tablets | 3334141000033116 | 418352003 | Dexibuprofen |
| Seractil 300mg tablets (Gebro Pharma GmbH) | 3334341000033118 | 35096411000001107 | Dexibuprofen |
| Dexibuprofen 400mg tablets | 3334241000033111 | 418855005 | Dexibuprofen |
| Seractil 400mg tablets (Thornton & Ross Ltd) | 3334441000033112 | 9447611000001101 | Dexibuprofen |
| Nuromol 200mg/500mg tablets (Reckitt Benckiser Healthcare (UK) Ltd) | 6386841000033117 | 18594011000001100 | Ibuprofen/ Paracetamol |
| Paracetamol 500mg / Ibuprofen 200mg tablets | 6386741000033110 | 18595211000001106 | Ibuprofen/ Paracetamol |
